# Integrating GWAS meta-analysis with human brain cell mapping implicates the amygdala and cortical inhibitory interneurons in tinnitus

**DOI:** 10.1101/2025.11.28.25340151

**Authors:** Nick M.A. Schubert, Shuyang Yao, Maryam Kazemi Naeini, Natalia Trpchevska, Charles Bishop, Argyro Bizaki-Vallaskangas, Linda Broer, Fabio Castellana, Kaare Christensen, Jacob v. B. Hjelmborg, Maria Pina Concas, Aurora Santin, Giorgia Girotto, Giuseppe Giovanni Nardone, Licia Iacoviello, Estonian Biobank Research Team, Jonas Mengel-From, Scott Gallagher, Alexander Gudjonsson, Vilmundur Gudnason, Nancy L. Heard-Costa, Sudha Seshadri, Howard J. Hoffman, Chuan-Ming Li, Jaakko Kaprio, Joel Rämö, Elmo Saarentaus, Johannes Kettunen, Kristi Krebs, Anna K. Kähler, Lenore J. Launer, Hampton Leonard, Mike A. Nalls, Patrik K.E. Magnusson, Joyce van Meurs, Lili Milani, Antti Mäkitie, Berthe C. Oosterloo, Ilja M. Nolte, Marianne Nygaard, Teemu Palviainen, Sheila Pratt, Nicola Quaranta, Rodolfo Sardone, Claudia L. Satizabal, John M. Schweinfurth, Eric Shiroma, Eleanor Simonsick, Lifelines Cohort Study, Harold Snieder, Christopher Spankovich, Patrick F. Sullivan, Andre Goedegebure, Andries Paul Nagtegaal, Sonja J. Pyott, Christopher R. Cederroth, Maxim B. Freidin, Frances MK Williams

**Affiliations:** Department of Otorhinolaryngology / Head and Neck Surgery, University of Groningen, University Medical Center Groningen, The Netherlands, RB Groningen, The Netherlands; Department of Medical Epidemiology and Biostatistics, Karolinska Institutet, Stockholm, Sweden; Department of Twin Research and Genetic Epidemiology, School of Life Course and Population Sciences, King’s College London, London, UK; Department of Physiology and Pharmacology, Karolinska Institutet, Stockholm, Sweden; Department of Otolaryngology and Communicative Sciences, The University of Mississippi Medical Center, Jackson, MS, USA; Tampere University, Faculty of Medicine and Health Technology, Department of Otolaryngology, Tampere, Finland; Pirkanmaan hyvinvointialue, Department of Ear and Oral Diseases, Tampere, Finland; Department of Internal Medicine, Erasmus Medical Center, GD Rotterdam, The Netherlands; Unit of Epidemiology and Statistics, Local Health Authority of Taranto, Taranto, Italy; The Danish Twin Registry, Department of Public Health, University of Southern Denmark, Odense C, Denmark; Department of Clinical Genetics, Odense University Hospital, Odense C, Denmark; Department of Clinical Biochemistry and Pharmacology, Odense University Hospital, Odense C, Denmark; Institute for Maternal and Child Health — IRCCS, Burlo Garofolo, Trieste, Italy; Department of Medicine Surgery and Health Sciences, University of Trieste, Trieste Italy; Department of Epidemiology and Prevention, I.R.C.C.S. Neuromed, Pozzilli, Italy; Department of Medicine and Surgery, LUM University, Casamassima, Italy; Estonian Genome Centre, Institute of Genomics, University of Tartu, Tartu, Estonia; Decibel Therapeutics Inc; Icelandic Heart Association, 201 Kopavogur, Iceland; Faculty of Medicine, University of Iceland, 101 Reykjavik, Iceland; Department of Neurology, Boston University Chobanian & Avedisian School of Medicine, Boston, MA, USA; Framingham Heart Study, Framingham, MA, USA; Glenn Biggs Institute for Alzheimer’s & Neurodegenerative Diseases and Department of Population Health Sciences, University of Texas Health Sciences Center, San Antonio, TX, USA; Epidemiology, Statistics, and Population Sciences, National Institute on Deafness and Other Communications Disorders (NIDCD), NIH, Bethesda, MD, USA; Institute for Molecular Medicine Finland (FIMM), University of Helsinki, Helsinki, Finland; Systems medicine, Research unit of Population Health, University of Oulu, Oulu, Finland; Biocenter Oulu, University of Oulu, Oulu, Finland; Finnish Institute for Health and Welfare, Helsinki, Finland; Laboratory of Epidemiology and Population Sciences, Intramural Research Program National Institute on Aging, Bethesda, MD, USA; Laboratory of Neurogenetics, National Institute on Aging, National Institutes of Health, Bethesda, MD, USA; Center for Alzheimer’s and Related Dementias, Bethesda, MD, USA; Data Tecnica, Washington DC, 20037, USA; Department of Otorhinolaryngology, Head and Neck Surgery, University of Helsinki and Helsinki University Hospital, HUS, Helsinki, Finland; Department of Otorhinolaryngology, Erasmus Medical Center, GD Rotterdam, The Netherlands; Department of Epidemiology, University of Groningen, University Medical Center Groningen, University of Groningen, RB Groningen, The Netherlands; Department of Communication Science & Disorders, University of Pittsburgh, Pittsburgh, Pa, USA; Otolaryngology Unit, Department of Translational Biomedicine and Neuroscience (DiBrain), University of Bari, Policlinico di Bari, Italy; Laboratory of Epidemiology and Population Sciences, National Institute on Aging, Baltimore, MD, USA; Longitudinal Studies Section, Translational Gerontology Branch, National Institute on Aging, Baltimore, MD, USA; Department of Genetics, University of North Carolina, Chapel Hill, NC, USA; Department of Neuroscience, School of Medicine and Health Science, Carl von Ossietzky Universität Oldenburg, Oldenburg, Germany; Department of Otolaryngology, Head & Neck Surgery, University of Tübingen Medical Center, Tübingen, Germany

**Author notes:** equal contribution (Shuyang Yao, Maryam Kazemi Naeini). equal contribution (Sonja J. Pyott, Christopher R. Cederroth, Maxim B. Freidin, Frances MK Williams). Correspondence: Christopher R. Cederroth.

**Keywords:** Tinnitus, Genome-wide association study, Heritability, Brain cell types, Amygdala, Phenotype heterogeneity

## Abstract

**Purpose:** Tinnitus is a common and heterogeneous condition whose genetic architecture remains poorly defined and whose definition varies across studies. We asked which common genetic variants are associated with tinnitus, to what extent genetic associations are shared across heterogeneous tinnitus definitions, and in which brain cell populations tinnitus heritability is concentrated.

**Methods:** Genome-wide association meta-analysis of 19 independent population-based cohorts from Europe and the United States (hereby named EARGEN), comprising 961,085 adults of European ancestry (75,584 tinnitus cases and 885,501 controls). Tinnitus was defined by self-report in 17 cohorts and by diagnostic codes in two. Single-nucleotide polymorphism (SNP) based heritability and genetic correlations were estimated by linkage disequilibrium score regression, genes were mapped by gene-based association together with positional and expression quantitative trait locus mapping. Heritability enrichment was tested by stratified linkage disequilibrium score regression across 31 brain cell superclusters derived from post-mortem human brain donors, and across the 461 cell clusters that compose them. Enrichment was tested for replication in the Million Veteran Program (MVP) under a self-report definition (114,325 cases, 201,343 controls) and a diagnostic-code definition (77,601 cases, 310,953 controls).

**Results:** Five independent loci reached genome-wide significance (p < 5 x 10^-8^) on chromosomes 2, 8, 12, 13 and 16, two of which (rs10877884 and rs2147632) have not been reported previously. SNP-based heritability was 0.0137 (SE = 0.0009) on the observed scale and 0.0535 (0.0036) on the liability scale. Genetic correlations were significant with hearing loss, anxiety disorder, major depressive disorder, chronic pain, and four further psychiatric, sleep and vestibular traits. Three superclusters showed a suggestive enrichment in the midbrain-derived inhibitory neurons, amygdala excitatory neurons, and medial ganglionic eminence interneurons at a false discovery rate (FRD) of 0.067, the two latter of which reached significance in the Million Veteran Program (MVP) under the diagnostic-code definition (0.030 and 0.006). At cluster resolution, the amygdala excitatory cluster was found significant (z = 4.82, false discovery rate = 3.3 x 10^-4^), whereas none of the ascending auditory relay cluster were found enriched (z ≤ 1.71, false discovery rate ≥ 0.21).

**Conclusion:** Tinnitus-associated common variation concentrates in limbic and cortical neuronal populations rather than in the ascending auditory pathway. The predisposition to tinnitus therefore appears to be central and largely non-auditory in origin. The small number of genome-wide significant associations obtained despite a large sample emphasizes the need for more detailed and standardized phenotyping focusing on the tinnitus disorder.

## Introduction

Tinnitus is the phantom perception of sounds in their physical absence. Its global prevalence is 14.2% of adults, with 1.4% of these individuals experiencing severe tinnitus (either self-reported or medically evaluated/diagnosed) [1]. It can be incapacitating and associated with insomnia, depression, anxiety, stress, and increased suicide risk [2–4]. Despite this burden, no disease-modifying treatment exists, reflecting our incomplete understanding of the pathophysiology underlying tinnitus. Investigating this pathophysiology is complicated by the heterogeneity of tinnitus, which varies in severity, associated hearing status and comorbidities, as well as in how it is characterized across clinical and population-based studies. This heterogeneity poses a challenge for identifying genetic variants shared across tinnitus phenotypes and for determining how these associations relate to other conditions and relevant neuronal populations.

Evidence from twin and adoption studies supports a genetic contribution to tinnitus, although heritability differs across phenotypes. Bilateral tinnitus is more heritable than unilateral tinnitus [5, 6], and familial aggregation studies [7] indicate greater genetic transmission in severe tinnitus. The first large, published genome-wide association study (GWAS), conducted in >200,000 individuals with self-reported tinnitus from the UK Biobank (UKB) and replicated in the US Million Veteran Program (MVP) cohort, found three loci (lead SNPs: rs143424888, rs553448379, rs11249981) with small effect sizes and estimated SNP-based heritability of approximately 6% [8]. A subsequent meta-analysis of these two datasets identified 29 genome-wide significant loci, and a comparison with hearing loss identified two new loci for tinnitus and suggested that tinnitus and hearing difficulties have distinct genetic contributions [9]. Studies of rare variants in severe or catastrophic tinnitus have identified synaptic and ion-channel genes [10, 11].

Although these studies establish a genetic contribution to tinnitus, they do not by themselves identify the biological context in which tinnitus-associated genetic variation acts. Such insights require integration of GWAS results with complementary genomic data. Genetic correlation analyses can assess shared genetic associations between tinnitus and related auditory, psychiatric, sleep, and pain phenotypes [12], while cell-type-resolved transcriptomic datasets can identify neuronal populations enriched for tinnitus-associated genetic variation [13]. Together, these approaches link tinnitus-associated genetic variation with other related phenotypes as well as specific neuronal phenotypes.

In this study, we assembled 19 population-based cohorts, the majority of which had not been included in previous genetic analyses of tinnitus. We then conducted a large-scale tinnitus GWAS meta-analysis and integrated these findings with genetic correlation and single-nucleus transcriptomic analyses. We sought to identify common genetic variants associated with tinnitus, determine the extent to which genetic associations with tinnitus were shared across cohorts with heterogeneous tinnitus definitions, assess genetic correlations between tinnitus and related phenotypes, and identify neuronal populations enriched for tinnitus-associated genetic variation.

## Materials and Methods

### Ethics Approval

Each contributing cohort obtained approval from its responsible research ethics committee or institutional review board and written informed consent from all participants, in accordance with the Declaration of Helsinki. Approving committees and permit numbers for each cohort are listed in the Supplementary Information. No new data were collected for the present analysis, which used summary-level results only. Ethical approval was obtained locally, and under an umbrella license in Stockholm (*Regionala etikprövningsnämnden, #*2015/2129-31/1).

### Study design and population cohorts

EARGEN comprises a total of 961,085 adult male and female participants of European ancestry that were included from the following 19 cohorts: Age, Genes/Environment Susceptibility – Reykjavik (AGES; n = 3,134), the Danish Twin Registry (DTR; n = 1,314), the Estonian Genome Center at the University of Tartu (EstBB; n = 195,394), FinnGen R12 (n = 520,210), Framingham Heart Study (FHS; n = 3,041), the Lifelines (LL) cohort study (original genotyping cohort (LL_cytosnp), UMCG Genetics Lifelines Initiative (UGLI) release 1, UGLI release 2; n = 54,184), Moli Sani cohort study (n = 1,198), the Rotterdam Study (RS, cohorts 1 – 3; n = 6,146), the Great Age Study (SA; n = 1,780; formerly known as Salus in Apulia study), Health ABC (HABC; n = 1288), three sub-studies of the Swedish Twin Registry: Screening Across the Lifespan Twin (SALT; n = 10,693), SALTY – young (n = 4,436), Screening Twin Adults: Genes and Environment (STAGE; n = 8,310), TwinsUK (n = 2,197), and UK Biobank (UKB; n = 147,760). For most cohorts, tinnitus was defined using cohort-specific self-report questionnaires. Self-report definitions varied across cohorts in question wording and criteria used to define cases and controls. For the EstBB and FinnGen cohorts, tinnitus cases and controls were defined using the International Classification of Diseases (ICD9: 3891 and ICD10: H93.1) diagnostic codes. A detailed description of phenotype definitions, case control distribution, and age ranges for each study is provided **Supplementary Table S1** and **Supplementary Information**.

### Genome-wide association studies and meta-analysis

GWAS was performed locally for each cohort, with association analyses adjusted for age, sex, and genetic principal components, after which summary statistics were collected for each study (more information in **Supplementary Table S1** and **Supplementary Information**). If necessary, cohort-level GWAS summary statistics were harmonized to the hg19 genome build prior to further analyses. Standardized quality control was performed using EasyQC software [14], followed by meta-analysis using GWAMA software (fixed-effect model, without using genomic control) [15]. Briefly, quality control was performed using EasyQC and excluded monomorphic SNPs, minor allele count ≤20, SNPs only in <100 individuals, duplicate SNPs, and SNPs with an imputation score < 0.5. Analyses were restricted to autosomal variants. After harmonizing allele coding and marker names, harmonized summary statistics were produced. LD score regression was applied to the quality-controlled and harmonized GWAS summary statistics from cohorts with >5,000 participants to assess the contribution of population stratification and other confounders to test-statistic inflation [12]. A genome-wide significance threshold was defined as P < 5 x 10^-8^. To identify lead SNPs representing independent association signals, conditional and joint association (COJO) analysis was performed [16].

### SNP-heritability and genetic correlation

LD score regression was used to estimate SNP-based heritability and genetic correlations. SNP heritability was calculated from the meta-analysis summary statistics and for individual cohorts with >5,000 participants. SNP-based heritability calculation from the meta-analysis summary statistics was performed on the observed and liability scale (assumed population prevalence 0.144). Genetic correlations were calculated between the tinnitus meta-analysis 13 selected disorders and traits (Hearing loss, Major Depressive Disorder, Neuroticism, Anxiety Disorder, Insomnia, Vertigo, Chronic Pain, Post-traumatic stress syndrome, Schizophrenia, Bipolar disorder, Alzheimer’s disease, Type 2 Diabetes, Cerebellar volume) [12]. Traits were selected based on previous known associations with tinnitus or hearing loss, type 2 diabetes mellitus was added as a negative control. To address multiple testing across 13 traits, statistical significance was set at FDR < 0.05. Pairwise genetic correlations were additionally estimated between all individual cohorts with >5,000 participants, multiple testing correction via FDR < 0.05 was applied.

### Gene mapping

Gene-based association analyses were performed using VEGAS2 [17] and MAGMA [18] v1.08 (both default settings) to identify genes associated with tinnitus based on the combined association signals of variants within each gene. In parallel, tinnitus-associated genes were mapped to genes using positional mapping and expression quantitative trait locus (eQTL) mapping using FUMA v1.5.2 (posMap, eqtlMap) [19].

### Brain cell-type and cross-trait enrichment analysis

Brain cell-type enrichment analysis was performed using a recently published methodology integrating GWAS summary statistics with cell-type resolved gene-expression data [13]. Brain cell populations were defined using single-nucleus RNA-sequencing gene expression data from a published human brain atlas [20], comprising approximately 100 dissections and three million nuclei from three postmortem donors. Cells were classified into 461 clusters and 31 superclusters. For each cell population, genes in the top decile of expression proportion (TDEP) [13] were used to define cell-type-specific annotations for enrichment testing. Stratified LD score regression was then used to test whether SNP-based heritability for tinnitus was enriched in each of these cell-type-specific annotations. To compare the tinnitus enrichment profile in the 31 superclusters with other phenotypes, results from Yao et al., were compared with the tinnitus enrichment results from our analysis, including bipolar disorder, hearing loss, insomnia, major depressive disorder, neuroticism, schizophrenia. Enrichment was considered significant at FDR < 0.05 and suggestive at FDR < 0.1.

### Replication of brain cell-type enrichment in an independent cohort

To test whether the brain cell-type enrichment observed here is reproducible in data that did not contribute to the meta-analysis, we obtained summary statistics for tinnitus in the 4^th^ release of the Million Veteran Program (MVP) from the MVP genome-wide and phenome-wide association resource, in which 2,068 traits were analyzed under a single uniform pipeline and deposited publicly (database of Genotypes and Phenotypes accession phs002453) [21]. The MVP shares no participants with any of the EARGEN cohorts analyzed here. Two definitions of tinnitus available within the same cohort of European ancestry were analyzed separately: self-reported tinnitus (114,325 cases, 201,343 controls) and tinnitus recorded by diagnostic code (phecode 389.4; 77,601 cases, 310,953 controls). Effect sizes were converted to the log-odds scale and standard errors derived from the reported 95% confidence intervals. Variants were restricted to HapMap3 outside the major histocompatibility complex. The reference panel, cell-type annotations, regression model and multiple-testing correction were identical to those applied to the EARGEN meta-analysis, so that coefficient z-scores are directly comparable between datasets. It is this comparability of the enrichment analysis, and not of the upstream association analysis, on which the replication depends. Before the MVP data were analyzed, the cell-type results of the EARGEN meta-analysis were reproduced from the deposited summary statistics as a calibration step. Tinnitus in the MVP has been previously examined by genome-wide association [9]. Because that study combined the MVP with the UK Biobank, which is one of the 19 cohorts analyzed here, it is not independent of either dataset. The replication reported here rests only on the absence of shared participants between the MVP and EARGEN.

## Results

### 2 new GWA-significant loci are associated with tinnitus

Among 961,085 individuals of European ancestry, including 75,584 tinnitus cases and 885,501 controls (**Table 1**, **Supplementary Table S1**, S3), corresponding to a prevalence of 7.86%, the EARGEN genome-wide meta-analysis identified five independent loci based on the COJO analysis reaching genome-wide significance on chromosomes 2, 8, 12, 13, and 16 (**Figure 1a**, b). The quantile-quantile plot of the meta-analysis showed a departure from the null expectation across the distribution (**Supplementary Figure S1**), with a genomic inflation factor of 1.21 and a mean chi-square of 1.26. The LD score regression intercept was 1.010 (SE 0.0085) and the ratio statistic, which estimates the share of that inflation not attributable to polygenicity, was 0.039 (SE 0.033), meaning the departure reflects polygenic signal rather than population stratification (**Supplementary Table S2**). Significant associations were observed at rs35098301, rs35009431, rs10877884, rs2147632, and rs12922637 (**Figure 1c**-g, **Supplementary Figure S3**). Three of these loci (i.e., rs35098301, rs35009431 and rs12922637) were reported in a previous tinnitus GWAS [9]. Two loci on chromosomes 12 and 13, represented by rs10877884 and rs2147632, respectively, have not been previously associated with tinnitus (**Supplementary Table S4**). Next, to estimate the proportion of phenotypic variation attributable to common genetic variation, SNP-based heritability was estimated at 0.0137 (SE = 0.0009) on the observed scale and 0.0535 (SE = 0.0036) on the liability scale. Finally, to assess the robustness of the meta-analysis to variation across cohorts, inspection of forest plots showed consistent differences in effect sizes, standard error and/or effect direction in the Lifelines subcohort genotyped on the Illumina HumanCytoSNP-12 array (LL_cytosnp; **Figure 1c**, **Supplementary Table S1**B), which prompted a leave-one-out meta-analysis excluding that subcohort. Exclusion of this cohort did not alter the meta-analysis effect estimates or I2 values (**Supplementary Table S5**), indicating that the principal associations were not driven by this cohort. Per-locus power ranged 0.52 to 0.80 with a mean of 0.66, and the minimum detectable effect at 80 % power was β 0.0081 at MAF 0.50 rising to 0.0185 at MAF 0.05 (**Supplementary Figure S2**).

**Figure 1.**
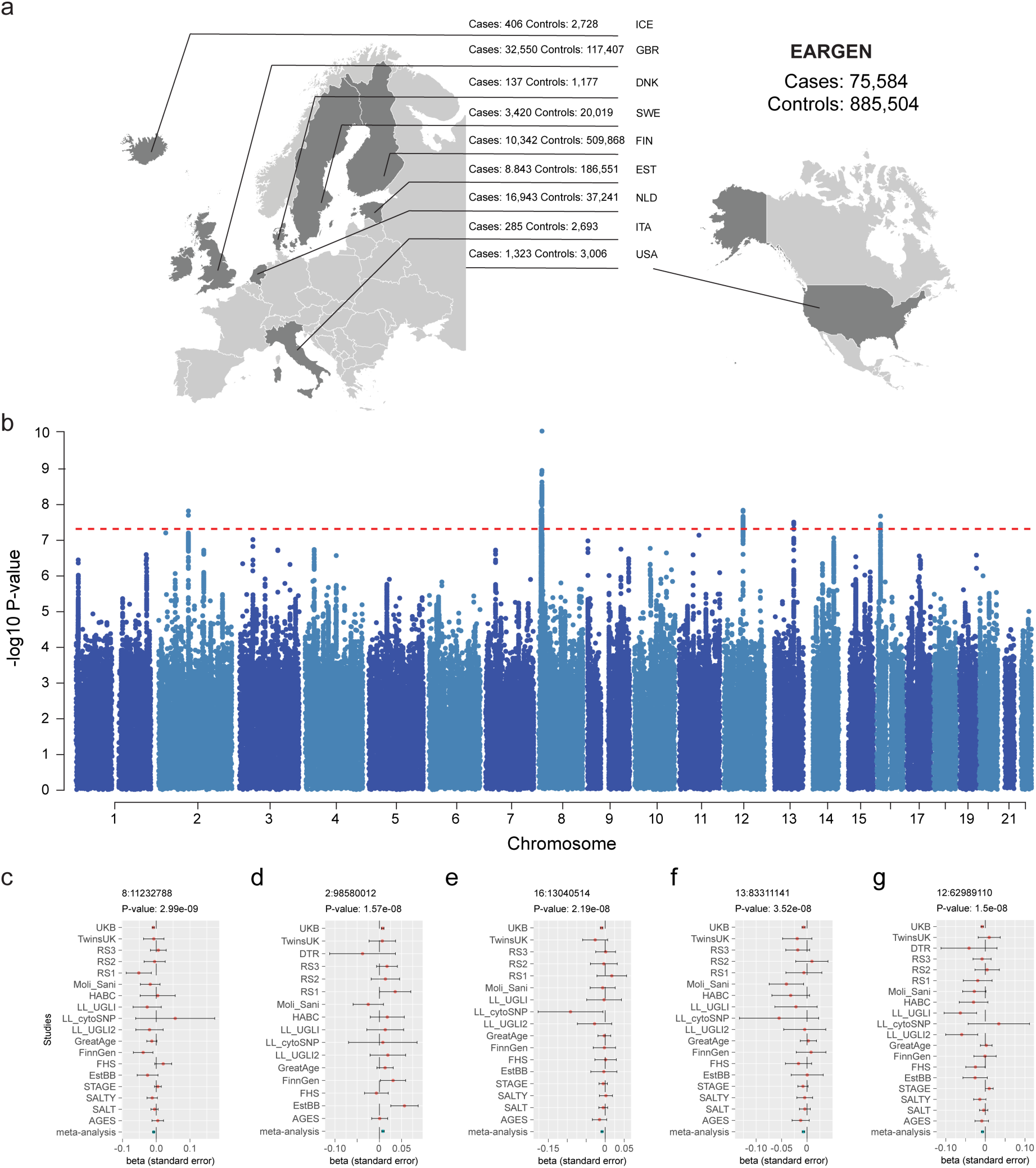
Genome-wide association meta-analysis for tinnitus. (a) Summary of contributing cohorts. More detailed information for each cohort can be found in **Supplementary Table 1**. (b) Manhattan plot of the tinnitus GWAS meta-analysis showing five genome-wide significant independent loci. The x-axis represents genomic position by chromosome, and the y-axis represents –log_10_ p-values. The red dotted line indicates the threshold for genome-wide significance (p-value 5 x 10-8). (c) Forest plots for the lead SNPs at the five genome-wide significant loci: rs35098301, rs35009431, rs10877884, rs2147632, and rs12922637. The x-axis represents the beta coefficient, and the y-axis represents the cohorts included in the analysis. Points (red) indicate the beta coefficients, and error bars (green) represent the standard error.

**Table 1.**
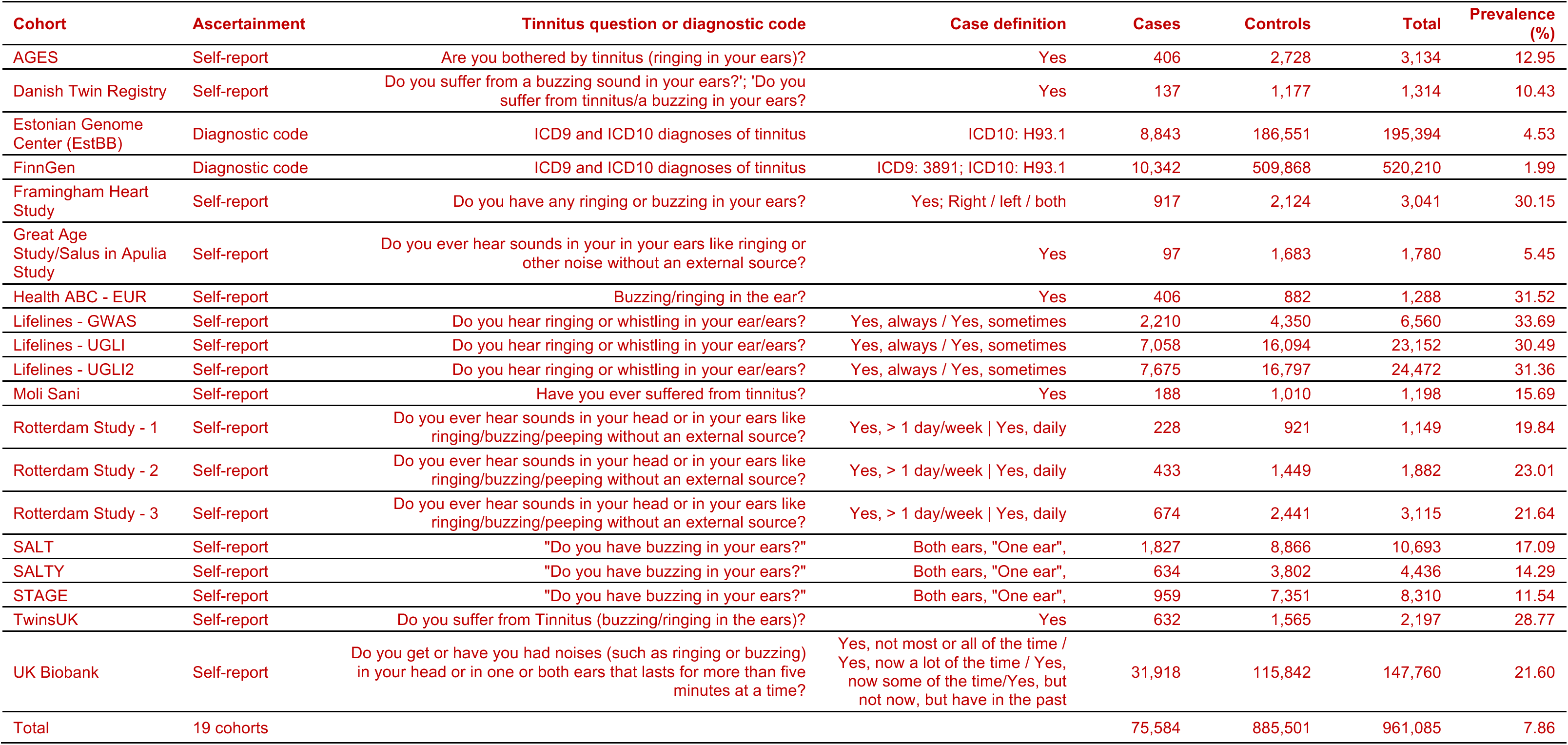
Tinnitus definition, ascertainment and prevalence in the 19 contributing cohorts. Prevalence is the proportion of cases among all participants of that cohort. Full cohort descriptions, genotyping and quality control are given in Supplementary Table 1.

### Coherent genetic associations across tinnitus phenotype definitions

Because tinnitus phenotype definitions differed across cohorts, with most cohorts using self-report and two cohorts using ICD-based definitions, h^2^ and genetic correlations across cohorts with >5000 individuals were estimated to test shared genetic associations across cohorts (**Supplementary Table S2**). No distinct pattern of genetic correlation distinguished self-report from ICD-based cohorts and most pairwise estimates indicated shared genetic associations across cohorts. Only the genetic correlations between EstBB and the Lifelines UGLI subcohort (LL_gsa), and between FinnGen and the Lifelines UGLI2 subcohort (LLaffy), were not statistically significant (**Supplementary Table S6**). To further investigate differences associated with tinnitus phenotype definition, separate meta-analyses were performed for self-report and ICD-based cohorts, and effect sizes and effect directions were compared with the full meta-analysis. Most genome-wide significant loci showed the same direction of effect in all three meta-analyses. Only SNP 12:62993793 showed a difference in direction between the self-report and ICD-based meta-analyses (**Supplementary Table S7**).

### Gene-based analyses and gene mapping

Gene-based association analyses were performed using VEGAS2 and MAGMA. These analyses identified the same 17 identical genes associated with tinnitus. A complementary variant-to-gene mapping using FUMA identified 30 genes by positional mapping and 37 genes by eQTL mapping (**Supplementary Table S8-S10**). After accounting for overlap among these gene lists, a total of 51 unique genes were identified. Seven genes were commonly identified by all four methods: *MFHAS1*, *TMEM131*, *MON2*, *C8orf12*, *XKR6*, *SLC35G5*, *C12orf61* (**Supplementary Figure S4**). Two of these genes, *TMEM131* and *SLC35G5*, were previously reported [9].

### Genetic correlations and brain cell-type enrichment

To examine shared genetic associations between tinnitus and other disorders, LD score regression was used to estimate genetic correlations between tinnitus and 13 selected disorders and traits. Significant positive genetic correlations were observed with major depressive disorder (Rg = 0.32), neuroticism (Rg = 0.26), anxiety disorder (Rg = 0.37), insomnia (Rg = 0.19), post-traumatic stress disorder (Rg = 0.24), chronic pain (Rg = 0.30), hearing loss (Rg = 0.50) and vertigo (Rg = 0.22); **Figure 2a**, **Supplementary Table S13**).

**Figure 2.**
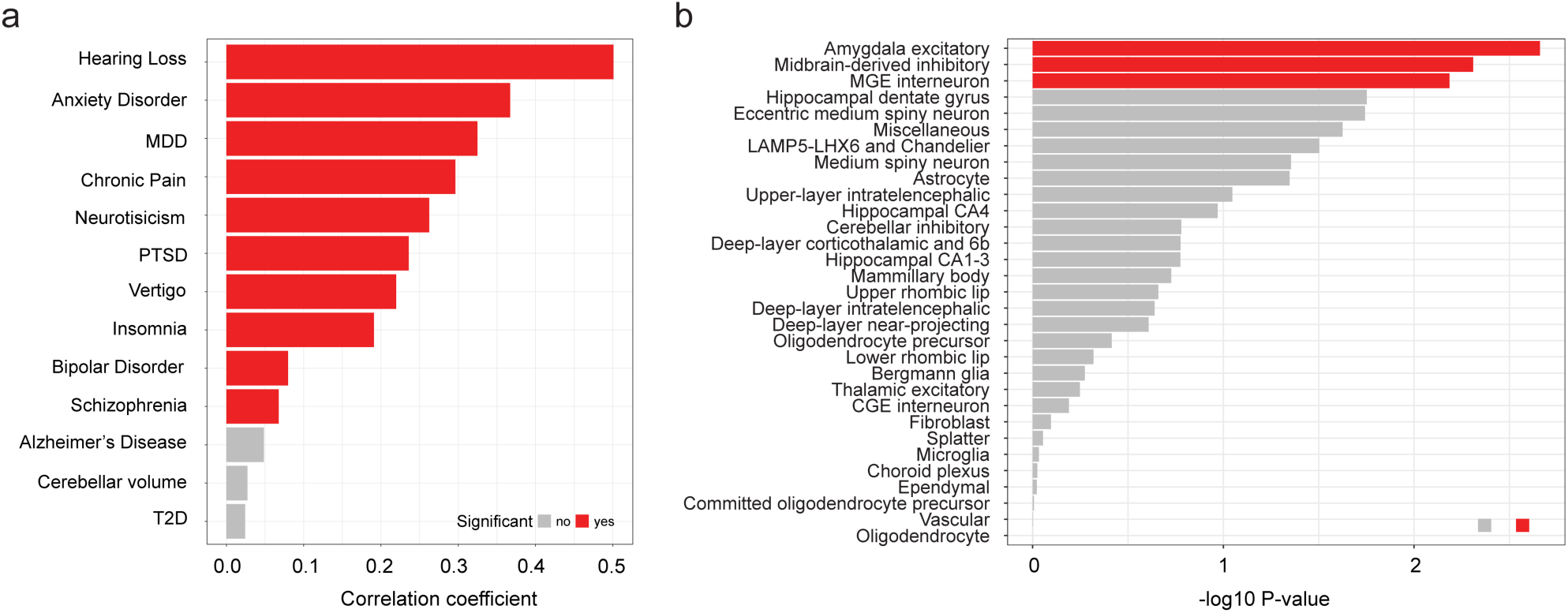
Brain cell-type enrichment and genetic correlations of tinnitus with other traits. (a) Brain cell supercluster enrichment analysis for tinnitus. The x-axis shows –log10 p-values. Superclusters with suggestive enrichment (FDR < 0.1) are shown in red; all others are shown in grey. (b) Genetic correlations between tinnitus and selected disorders and traits estimated using LD score regression. Correlations with FDR < 0.05 were considered significant. Abbreviations: MGE, medial ganglionic eminence; CGE, caudal ganglionic eminence; MDD, major depressive disorder; PTSD, post-traumatic stress disorder; T2D = type 2 diabetes mellitus.

We next used stratified LD score regression and cell-type specific gene-expression analyses derived from a single-nucleus RNA-sequencing atlas to test whether SNP-based heritability for tinnitus was enriched in genomic annotations for specific brain-cell populations[13]. After correction for multiple testing, three superclusters showed suggestive enrichment: amygdala excitatory neurons (FDR = 0.067), midbrain-derived inhibitory neurons (FDR = 0.067), and medial ganglionic eminence interneurons (FDR = 0.067; **Figure 2b**, Supplementary Tables S11 and S12).[13]

This approach also revealed that enrichment in these superclusters was also observed for several other disorders and traits associated previously with tinnitus in epidemiological studies, including major depressive disorder, insomnia, schizophrenia, bipolar disorder, and neuroticism **(Supplementary Figure S5, Supplementary Table S12)** [2, 22, 23].

### Replication of brain cell-type enrichment in an independent cohort

Because none of the enrichments described above reached the prespecified FDR < 0.05 threshold, we sought replication in the MVP, which contributes no participants to the present meta-analysis. SNP-based heritability in the MVP was higher than in our meta-analysis under both definitions (self-report 0.0440, SE = 0.0023; diagnostic code 0.0286, SE = 0.0017; observed scale). Of the three superclusters showing suggestive enrichment here, two were also enriched in the MVP. Amygdala excitatory neurons and medial ganglionic eminence interneurons remained suggestive under the self-report definition (z = 2.54 and z = 2.56, FDR = 0.053 for both), but passed below FDR < 0.05 under the diagnostic-code definition of the same cohort (coefficient z = 2.52, FDR = 0.030 and z = 3.41, FDR = 0.006, respectively). Midbrain-derived inhibitory neurons were not enriched in either definition (z = 1.64, FDR = 0.195; z = 1.52, FDR = 0.159; **Figure 3a**, **Table 2, Supplementary Table S14**).

**Figure 3.**
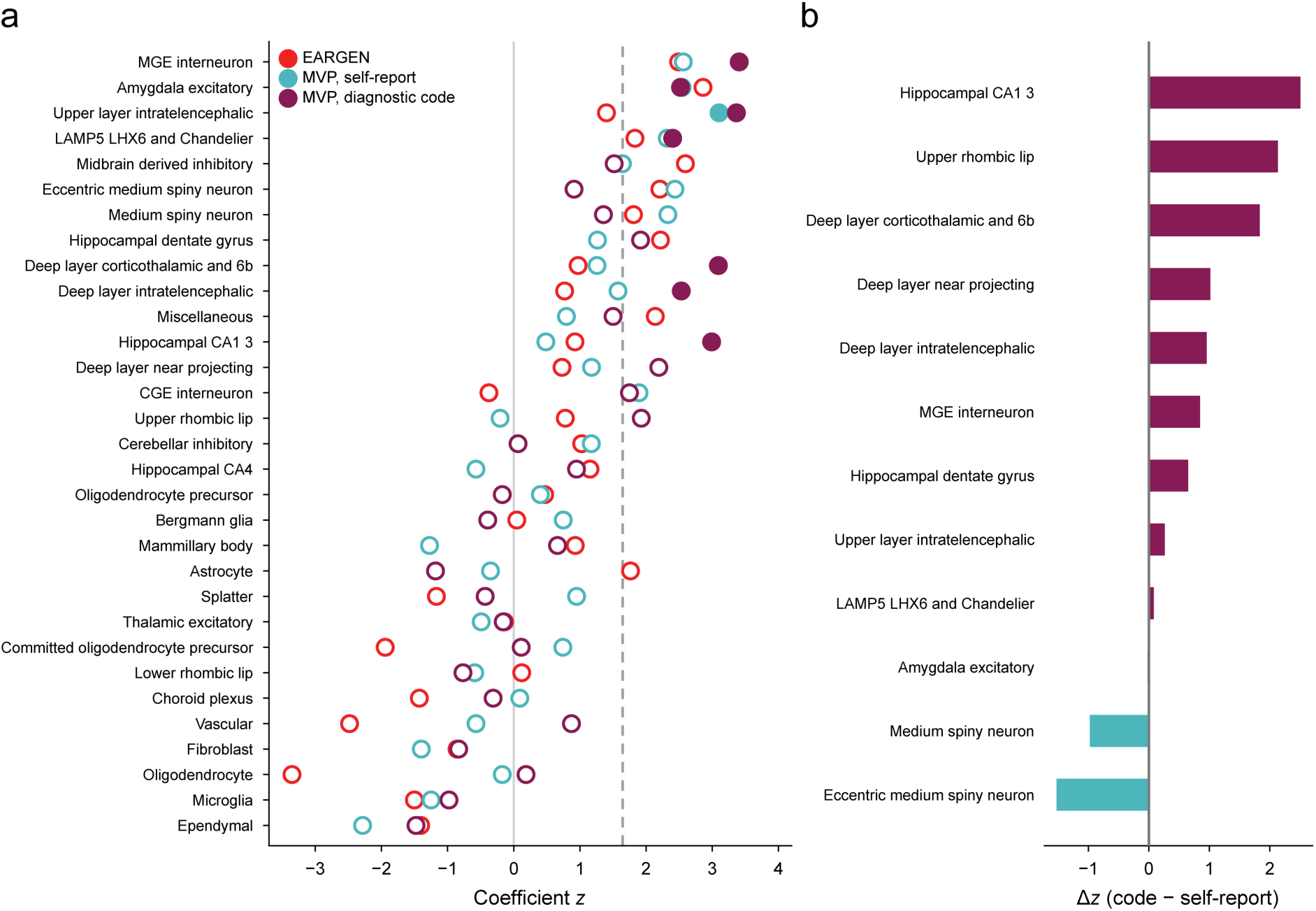
Replication of brain cell-type enrichment in the Million Veteran Program. (a) Coefficient z-scores for the 31 superclusters in the tinnitus meta-analysis (EARGEN, red circles) and in the two MVP tinnitus definitions, self-report (cyan circles) and diagnostic code (purple circles). Filled symbols indicate FDR < 0.05, open symbols FDR ≥ 0.05; the dashed line marks a one-sided z of 1.645. Superclusters are ordered by the mean z across the three analyses. (b) Change in coefficient z between the self-report and diagnostic-code definitions within the MVP, for the superclusters reaching FDR < 0.10 in either. Positive values indicate stronger enrichment under the diagnostic-code definition.

**Table 2.**
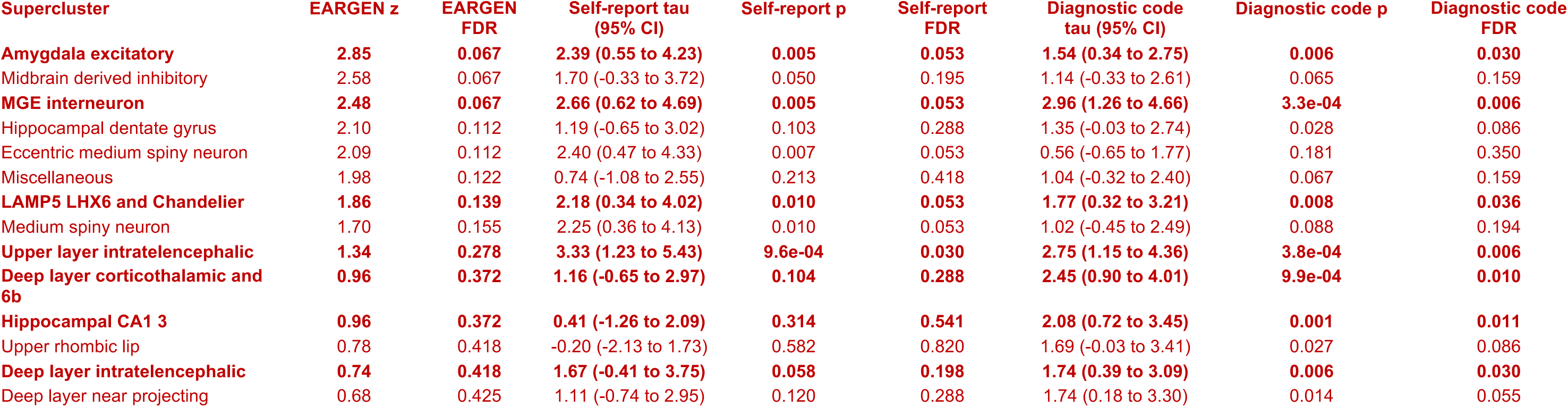
Replication of brain cell supercluster enrichment in the Million Veteran Program. Superclusters reaching a false discovery rate below 0.10 in EARGEN or in either Million Veteran Program phenotype definition are listed, ordered by their EARGEN coefficient z. Regression coefficients are shown multiplied by 109 with 95% confidence intervals. p values are one-sided and false discovery rates are corrected across the 31 superclusters within each analysis.

Models of thalamocortical dysrhythmia propose that tinnitus arises from altered firing of thalamic relay neurons following deafferentation [24–27]. This was not supported by the present analysis whereby thalamic excitatory neurons, the supercluster containing the medial geniculate body, showed no enrichment in any analysis (z = −0.14 in the EARGEN meta-analysis, −0.49 and −0.16 in the MVP). Because this supercluster spans the whole thalamus, it is possible that the heritability concentrated specifically in the medial geniculate body would be diluted. As a consequence, the result constrains the cell class, not the nucleus. No non-neuronal supercluster reached FDR < 0.10 in any dataset (**Figure 3a**).

Enrichment was more pronounced under the diagnostic-code definition than under self-report, in spite of 36,724 fewer cases and a lower heritability z-score (16.9 versus 18.9). Seven superclusters reached FDR < 0.05 with diagnostic codes, against one with self-report. Five of these were not detected in the EARGEN-cohort analysis reported above: upper-layer intratelencephalic neurons (z = 3.36, FDR = 0.006), deep-layer corticothalamic and 6b neurons (z = 3.09, FDR = 0.010), hippocampal CA1–3 neurons (z = 2.99, FDR = 0.011), deep-layer intratelencephalic neurons (z = 2.53, FDR = 0.030) and LAMP5 LHX6 and chandelier cells (z = 2.40, FDR = 0.036). Coefficient z-scores for the two definitions were correlated across the 31 superclusters (r = 0.72). Striatal populations moved in the opposite direction, medium spiny neurons falling from z = 2.33 (FDR = 0.053) under self-report to z = 1.35 (FDR = 0.194) under diagnostic codes, and eccentric medium spiny neurons from z = 2.44 (FDR = 0.053) to z = 0.91 (FDR = 0.350; **Figure 3b**).

### The ascending auditory pathway is not enriched in tinnitus GWA signals

The 31 superclusters analyzed here are defined by cell class rather than by anatomy, and none of them corresponds to a structure of the ascending auditory pathway. To assess whether the latter is enriched in tinnitus GWA signals, we repeated the analysis across the 461 clusters that compose these superclusters (**Figure 4, Supplementary Table S15**). Nomenclature for these clusters can be found in Siletti et al. [20]. Several of these clusters were drawn almost entirely from a single dissection, so the auditory relays could be tested directly. Two clusters reached a false discovery rate below 0.05 across the 461 tests (**Figure 4a**). The strongest was Amyg-Exc 161 (z = 4.82, FDR = 3.3 × 10⁻⁴), a glutamatergic population whose cells derive from dissections of the corticomedial nucleus of the amygdala (57.7%), the piriform cortex (21.2%) and the perirhinal gyrus (10.4%). The second was Misc 164 (z = 3.59, FDR = 0.039), a glutamatergic population of 659 cells also sharing the piriform cortex (23.8%), the lateral entorhinal cortex (42.3%) and the subiculum (15.0%).[20]

**Figure 4.**
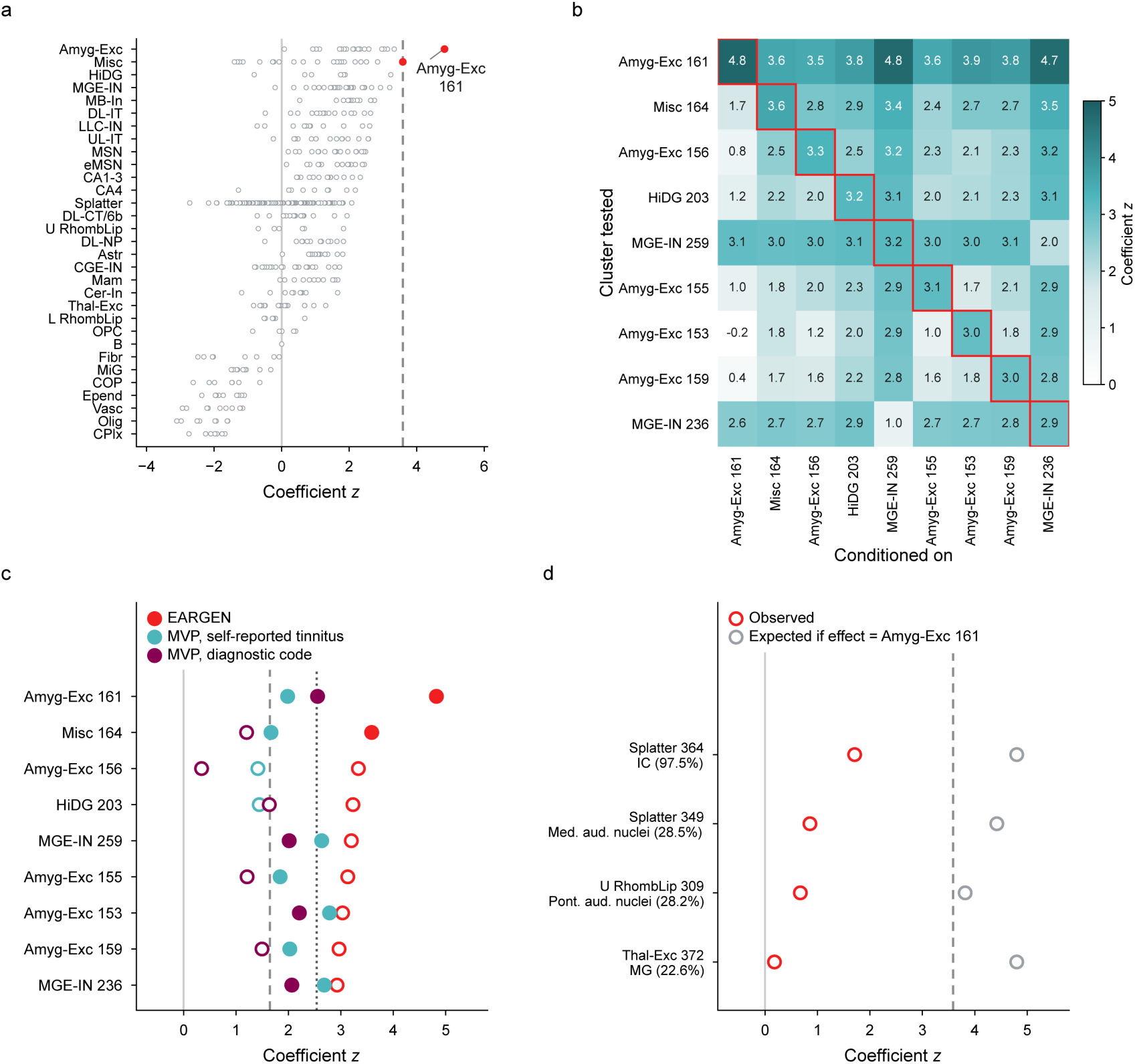
Cell-type enrichment of tinnitus SNP-heritability at the resolution of the 461 Siletti clusters. (a) Stratified LD score regression coefficient z for each of the 461 clusters of the adult human brain atlas in the EARGEN meta-analysis, grouped by the supercluster to which each cluster belongs. Superclusters are ordered by the largest coefficient z among their clusters and abbreviated as in Yao et al. [13]. The abbreviations are defined in **Supplementary Table S15**. Filled red circles denote clusters reaching a false discovery rate below 0.05 across the 461 tests, open circles the remainder, and the dashed line the corresponding threshold of z = 3.59. (b) Pairwise conditional analysis of the nine clusters reaching a false discovery rate below 0.10 in panel (a). Each cell gives the coefficient z of the cluster named on the vertical axis when the top decile expressed genes of the cluster named on the horizontal axis are added to the baseline model. Cells outlined in purple on the diagonal give the unconditional value. (c) Coefficient z for the same nine clusters in EARGEN and in the two Million Veteran Program phenotype definitions. Filled circles denote a false discovery rate below 0.05 for EARGEN (red circles) and a one-sided p below 0.05 for the two Million Veteran Program analyses [self-report (cyan circles) and diagnostic code (purple circles)], the latter being targeted tests of clusters specified in advance by EARGEN. The dashed line marks a one-sided p of 0.05 and the dotted line the Bonferroni threshold of 0.05/9 for the nine clusters tested. (d) Coefficient z of the four clusters that sample the subcortical relays of the ascending auditory pathway, with the percentage of each cluster drawn from the relevant dissection given in parentheses. Only Splatter 364 is dominated by a single structure, so the three remaining clusters constrain their relay less tightly. Grey circles give the observed value and purple circles the value that the same test would have returned had the cluster carried a coefficient equal to that of Amyg-Exc 161. The dashed line marks z = 3.59 as in panel (a). Primary auditory cortex has no cluster of its own, since its cells distribute across the general neocortical clusters, and is therefore absent from this panel.

Amyg-Exc and Misc belong to a set of nine clusters that all reached FDR < 0.1. Because neighbouring cell types share many of the genes that most specifically mark them, we evaluated how many distinct signals these nine represent using an “add-one-in” methodology as in Yao et al. [13] by repeating each test with the marker genes of one of the other clusters added to the model (**Figure 4b, Supplementary Table S16**). The amygdala clusters behaved as a single finding, since each of them fell below z = 1.1 once the marker genes of Amyg-Exc 161 were added. Misc 164 and HiDG 203 lost their enrichment in the same way, falling to z = 1.7 and 1.2. In contrast, Amyg-Exc 161 itself held between z = 3.5 and 4.8 whichever other cluster was added. The two interneuron clusters likewise reduced to one, and the amygdala and the interneuron signals proved independent of each other, as each kept its enrichment when the other was added (z = 3.1 and 2.6). The nine clusters thus represent two distinct signals, one carried by excitatory neurons of the amygdala and the neighboring paleocortex and one by cortical MGE interneurons. Which amygdala population carries the first could not resolved here, as Amyg-Exc 161 shares most of its marker genes with its neighbours and is the strongest of twenty estimates of what appears to be one shared enrichment.[13]

We then tested these nine clusters in the Million Veteran Program, treating them as hypotheses specified in advance in EARGEN (**Figure 4c**). All nine carried a positive coefficient under both phenotype definitions. Four reached a one-sided p below 0.05 under the diagnostic code and seven under self-report, where 0.45 of nine would be expected by chance (binomial p = 6.4 × 10⁻⁴ and 2.6 × 10⁻⁸). Amyg-Exc 161 was the only cluster to satisfy a Bonferroni threshold of 0.05/9 under the diagnostic code (p = 0.0053).

Several of the 461 clusters are drawn almost entirely from a single dissection, so the auditory relays can be tested directly (**Figure 4d**). One cluster, Splatter 364, is composed of 97.5% inferior colliculus and reaches a coefficient z of 1.71 (FDR = 0.21); the cluster that samples the medial geniculate, Thal-Exc 372, reaches 0.18 (FDR = 0.68), and the clusters that sample the pontine and medullary auditory nuclei, U RhombLip 309 and Splatter 349, reach 0.67 (FDR = 0.52) and 0.86 (FDR = 0.46). Only Splatter 364 is dominated by a single structure, as the three remaining clusters draw between 22% and 28% of their cells from the relevant nucleus and therefore constrain it less tightly. Had any of these populations carried a coefficient equal to that of Amyg-Exc 161, each of these tests would have returned a z between 3.8 and 4.8. The absence of enrichment in the subcortical auditory relays is therefore a result and not a limit of resolution. Primary auditory cortex is the exception, as its cells distribute across the general neocortical clusters and no cluster is specific to it.

## Discussion

Here we report the EARGEN genome-wide meta-analysis of tinnitus comprising 961,085 individuals from 19 cohorts, including 75,584 tinnitus cases. We identified associations with tinnitus at five independent loci, two of which have not been reported previously. Despite differences in tinnitus phenotype definitions, genetic correlation analyses indicated substantial shared genetic associations across cohorts. Tinnitus was also genetically correlated with hearing loss and several psychiatric, sleep, and pain-related phenotypes. The integration of GWAS results with human brain cell-type expression data showed suggestive enrichment in amygdala excitatory neurons, midbrain-derived inhibitory neurons and medial ganglionic eminence-derived interneurons. Whereas none of these reached the pre-specified significance threshold (FDR < 0.05) in the EARGEN meta-analysis itself, the first and the third did so in an independent cohort, whereas the midbrain-derived population did not.

### Genetic associations across heterogeneous tinnitus phenotypes

Despite differences in tinnitus phenotype definition across cohorts, our genetic correlation analyses showed substantial shared genetic associations across cohorts, with most genome-wide significant loci showing the same direction of effect in meta-analyses separately examining self-report and ICD-based definitions of tinnitus. Nevertheless, despite the large overall sample size included in our study, we identified fewer loci than a recent GWAS meta-analysis of the UKB and the MVP, which reported 29 loci in its European-ancestry meta-analysis of 481,874 participants, and 39 non-overlapping loci across its European, African, Latin American and cross-ancestry analyses [9].

Several differences between the studies may have contributed to the greater number of loci identified in the previous study. First, the substantially larger number of tinnitus cases in that study likely contributed to greater statistical power for locus discovery. Second, the MVP comprises military veterans, who may share relevant environmental exposures, including noise exposure, potentially reducing some sources of heterogeneity. Third, differences in tinnitus phenotype definition may also have contributed. In the MVP cohort, almost 72% of tinnitus cases were identified using ICD codes, whereas two of our 19 cohorts used ICD codes for classifying tinnitus, accounting for only 25% of tinnitus cases in our study. Although ICD codes do not directly measure tinnitus severity, ICD-based definitions of tinnitus may enrich for individuals with tinnitus sufficiently bothersome to seek healthcare and potentially for more severe tinnitus. Because previous studies suggest greater heritability of severe tinnitus [6, 7], such enrichment could also increase the power for locus discovery. Thus, while our findings indicate substantial genetic sharing despite heterogenous definitions of tinnitus, better characterization of tinnitus heterogeneity, including severity and relevant exposure factors, will be important for determining which genetic contributions are broadly shared and which vary with specific tinnitus characteristics [28].

### Gene-based associations and candidate gene mapping in tinnitus

Gene-based association and variant-to-gene mapping analyses identified 51 unique genes, with seven genes identified across all four approaches. Among these, *TMEM131* and *SLC35G5* have previously been associated with tinnitus [29], while *MON2* appears as a new candidate. *MON2* is expressed in the mouse cortex, hippocampus, thalamus and striatum (Allen Brain Atlas; https://atlas.brain-map.org/) and functions as a membrane traffic regulator between the Golgi and endosomes [30], potentially regulating neuronal communication. Nevertheless, the role of *MON2* in neuronal function remains largely unknown, underscoring the need for further research to determine whether and how it may contribute to tinnitus.

### Genetic correlations and brain cell-type enrichment, and potential neuronal involvement in tinnitus

The genetic correlations observed with hearing loss and several psychiatric, sleep, and pain-related disorders indicate that tinnitus shares genetic associations with conditions extending beyond the auditory system. Subsequent integration with human brain cell-type expression data provided further, although suggestive, evidence for involvement of specific neuronal populations, notably amygdala excitatory neurons, midbrain-derived inhibitory neurons, and medial ganglionic eminence-derived interneurons. Because these cell-type enrichments are suggestive (not reaching significance at the prespecified FDR < 0.05 threshold) and cannot establish causal involvement of these neuronal populations in tinnitus, they should be interpreted cautiously, while remaining consistent with proposed neuronal mechanisms of tinnitus. Two of the three enrichments were also observed in an independent cohort, as described below.

The amygdala comprises GABAergic neurons, predominantly in its central and medial nuclei, and glutamatergic pyramidal-like neurons in the basolateral complex and has been associated with tinnitus in a range of studies. Work on brain imaging has in general pointed to the involvement of the limbic system [31], and a previous tinnitus GWAS also reported enrichment of the amygdala [9]. That analysis tested bulk tissue expression from GTEx, where the unit of analysis is the dissected region: eleven of the thirteen brain tissues examined by Clifford et al. passed the Bonferroni threshold, the strongest signals arising in cerebellum and cortex. However, bulk tissue cannot separate the cell populations that a region contains, which is what our analysis aimed to address. Partitioning heritability across 31 transcriptomically defined cell populations rather than across dissected regions places the signal in an excitatory neuronal population whose cells derive principally from amygdala dissections, while 28 of the 31 populations tested showed no suggestive enrichment, among them every non-neuronal population examined. Heritability concentrated in a specific excitatory neuronal population rather than being distributed across the cells a region contains, an observation that was found on two independent datasets. The amygdala connects to both the ventromedial prefrontal cortex and the nucleus accumbens, which together form a frontostriatal gating system, and failure of this gatekeeping mechanism has been proposed to contribute to tinnitus [32]. Our cell-type enrichment could not localize signal within that circuit nor establish its direction, thus further work is needed to determine how enrichment in these neurons might contribute to tinnitus or influence sensory salience.

Suggestive enrichment was also observed in medial ganglionic eminence-derived interneurons and in midbrain-derived inhibitory neurons, the second and third most enriched superclusters in the cell-type analysis. Only the former was also enriched in the MVP (FDR = 0.006 under its diagnostic-code definition). Medial ganglionic eminence-derived interneurons are the parvalbumin– and somatostatin-expressing GABAergic populations of the cortex and the hippocampus, where they constrain the excitability of principal neurons. Disrupted GABAergic function in animal models of tinnitus is well established [33], and GABAergic enhancement in mice abolishes tinnitus [34]. These findings are consistent with models in which reduced inhibition contributes to increased compensatory central activity following peripheral deafferentation [35]. In humans, evidence for altered GABAergic signaling remains inconclusive, largely due to conflicting results in clinical trials repurposing GABA-enhancing drugs for tinnitus [36]. Thus, our enrichment findings are consistent with a contribution of forebrain inhibitory populations to tinnitus, although future work is needed to determine the cell-type specific contributions to alterations in inhibition. In contrast, the midbrain-derived population was not found enriched in either MVP analysis, suggesting the absence of contribution from midbrain inhibitory circuits on tinnitus.

Two consequences derive from these observations. First, amygdala excitatory neurons and medial ganglionic eminence interneurons only reached significance under a diagnostic-code definition, which underlines the principal weakness of the EARGEN analysis (diagnostic-code cases being 7.4-fold rarer in EARGEN than in MVP). Second, neither of the two cell types belongs to the ascending auditory pathway. Amygdala excitatory neurons are the glutamatergic projection neurons of the basolateral complex, and medial ganglionic eminence interneurons give rise to the parvalbumin– and somatostatin-expressing inhibitory populations distributed throughout cortex and hippocampus. Common genetic variation associated with tinnitus therefore concentrates in genes specifically expressed by limbic projection neurons and by cortical inhibitory interneurons. This distribution is informative when compared against the genetic correlation we observe with hearing loss (Rg = 0.50). Tinnitus shares a substantial part of its common-variant architecture with the hearing loss [9], yet the cells in which that heritability is concentrated are limbic and cortical rather than auditory. Whether the inhibitory contribution reflects an altered balance between excitation and inhibition, as proposed from animal models of tinnitus in which enhancing inhibition on GABAergic neurons prevents noise-induced tinnitus [37–40], remains to be investigated.

The cell types that emerge only under the diagnostic-code definition are very informative. Layer 5 extratelencephalic and layer 6 corticothalamic neurons show cell-type-specific plasticity after noise-induced hearing loss [41] and become overamplified after cochlear synaptopathy [42]. Cortical parvalbumin interneurons, of which chandelier cells are one subtype, causally control noise-impaired gap detection and loudness perception [43, 44], and hippocampal excitability and synaptic connectivity are altered after noise trauma [45]. The contribution of intratelencephalic neurons to tinnitus remains to be determined. Together with the amygdala and interneuron populations already identified, what these sup erclusters have in common is a position within cortico-cortical, corticothalamic and cortico-limbic loops (**Figure 5**). In contrast, no enrichment was found in the ascending auditory relays, where the precision at this resolution would have revealed an effect of the size seen in the amygdala, making it difficult to reconcile with the large animal literature in which these same structures become hyperactive after acoustic trauma or salicylate [25, 33, 46–49]. We note however that the auditory cortex could not be tested here, as its cells distribute across the general neocortical clusters and no cluster of this atlas is specific to it. Tinnitus and tinnitus disorder are now distinguished on clinical, neurobiological and genetic grounds [50, 51], and the non-auditory contribution is what correlates with suffering [52–55]. The same framework assigned common variants of small effect to tinnitus and rarer variants of larger effect to tinnitus disorder. Instead, our results suggest that common variation also contributes to the disorder dimension through the circuitry that gives a percept its aversive value, consistent with the sharper enrichment obtained under the diagnostic-code definition within the MVP. Thus, the heritability captured here, read through this atlas, is a property of distributed forebrain circuitry and its inhibitory cells, rather than of the ascending auditory pathway.

**Figure 5.**
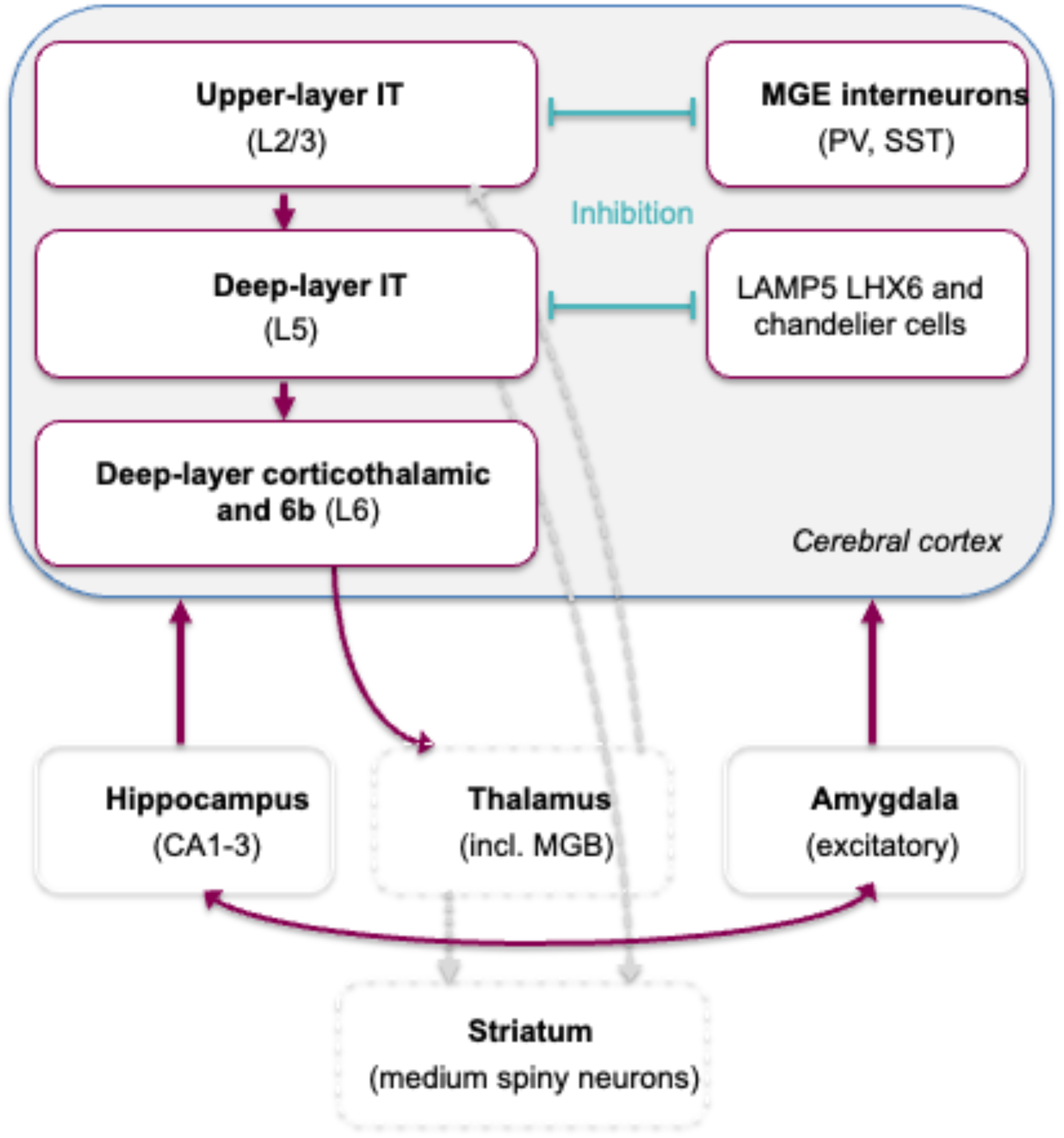
Forebrain cell populations enriched for tinnitus heritability under a diagnostic-code definition. Schematic of the cell classes reaching FDR < 0.05 in the MVP diagnostic-code analysis and of the projections that connect them. The diagram shows cell classes and canonical connectivity between the structures in which they reside. It does not depict directionality inferred from the genetic data, which cell-type enrichment cannot provide.

### Strengths and limitations

Strengths of this study include the integration of 19 cohorts with heterogeneous tinnitus definitions, which allowed genetic sharing across phenotypes to be measured rather than assumed, together with complementary gene-based, genetic correlation and human brain cell-type analyses. The cell-type analysis was performed at two resolutions, and the clusters that reached significance were tested against one another so that the number of distinct signals could be counted. It was then repeated in a cohort contributing no participants, under two phenotype definitions, with the same reference panel, annotations and regression model, which makes the coefficients directly comparable between datasets. This study also has several limitations. This study also has several limitations. The cohorts investigated included only individuals of European ancestry, limiting generalizability to other populations. The study operated at its own detection boundary, so a comparable number of loci of similar effect remains undetected, and this rather than any difference in method is why fewer loci were identified here than in the larger case series of Clifford et al. [9]. Genetic sharing across definitions, although substantial, was incomplete, so each case of a coarsely defined phenotype carries less information than a case of a sharply defined one. Only two cohorts used ICD-based definitions, and they contribute most of the participants but a minority of the cases, which limits the direct comparison between self-report and ICD-based definitions; the comparison within the MVP carries that contrast instead. Tinnitus severity, persistence and laterality were only partly characterized, environmental exposures were not, and hyperacusis, which commonly co-occurs with tinnitus [56] and may contribute to tinnitus phenotype heterogeneity, was not evaluated in our cohorts. Sex– and gender-based analyses were not performed although association analyses were adjusted for sex within each cohort. In addition, the analyses were restricted to the autosomes, and the X chromosome was not examined. This gap is shared across all auditory GWAS, none of which report X-chromosome associations [9, 57–64], something that will have to be addressed in future studies. Finally, cell-type enrichment identifies the populations whose specifically expressed genes carry heritability, but it does not establish directionality, does not indicate whether those genes are over– or under-active, and does not localize the signal within a cell class; the annotation derives from a small number of post-mortem donors, no cluster is specific to the auditory cortex, and the two MVP definitions are not independent samples, so their agreement speaks to the robustness of the phenotype definition rather than to a second replication.

## Conclusions

Overall, EARGEN, a meta-analysis of 19 independent cohorts and 75,584 cases, identified five loci associated with tinnitus, two of them not previously reported, and showed that genetic associations are substantially shared across heterogeneous phenotype definitions. Tinnitus was genetically correlated with hearing loss and with psychiatric, sleep and pain-related phenotypes, and its heritability was enriched in defined neuronal populations. Resolving that enrichment to individual cell clusters placed it in excitatory neurons of the amygdala and the neighboring paleocortex and in cortical interneurons, with no enrichment in the clusters representing the ascending auditory relays, where an effect of the same size would have been detected. Both enriched populations were recovered in the Million Veteran Program, which shares no participants with our cohorts, and more strongly under a diagnostic-code definition of tinnitus, so the predisposition to tinnitus appears to be central and largely non-auditory. The yield of genome-wide significant associations nevertheless remained small relative to the sample size, and sharing across definitions, while substantial, was incomplete. Phenotyping that resolves severity, persistence and laterality is therefore likely to advance tinnitus genetics further than additional cases of a binary phenotype.

## Additional information

### Data Availability

The EARGEN meta-analysis summary statistics are deposited in Zenodo (https://doi.org/10.5281/zenodo.17639792) and will be made publicly available upon publication. The Million Veteran Program summary statistics analysed here are the open-access release of the MVP genome-wide and phenome-wide association resource (database of Genotypes and Phenotypes accession phs002453) and were downloaded from https://ftp.ncbi.nlm.nih.gov/dbgap/studies/phs002453/analyses/. The single-nucleus RNA-sequencing data of the adult human brain are available as the Human Brain Cell Atlas v1.0 through CELLxGENE (https://cellxgene.cziscience.com/collections/283d65eb-dd53-496d-adb7-7570c7caa443), and the cluster annotation and dissection tables used here are those distributed at https://github.com/linnarsson-lab/adult-human-brain. The LD scores for the top decile expressed genes of each supercluster and cluster, together with the LDSC baseline model version 1.2, are deposited in Zenodo (https://doi.org/10.5281/zenodo.15194124).

### Code Availability

Analysis code for the meta-analysis and post-GWAS analyses, including the stratified LD score regression scripts for the brain cell-type enrichment, its conditional analysis and the Million Veteran Program replication, is available at https://github.com/translational-audiology-lab/GWAS_tinnitus. The cell-type enrichment follows the top decile expression proportion framework of Yao et al. [13], implemented at https://github.com/Hjerling-Leffler-Lab/TDEP-sLDSC. Stratified LD score regression was run with LDSC (https://github.com/bulik/ldsc) at the supercluster level and with the ldsR implementation (https://github.com/ararder/ldsR) at the cluster level.

### Artificial Intelligence

A large language model [Perplexity, Version 2.260428.0 (670)] was used during the drafting phase to assist in proofreading and revising author-written text, and verification that all cited numbers in the main text were consistent with the ones provided in the Supplementary materials. No AI tools were used to generate data, produce figures, or create artwork. All content was subsequently written, revised, and verified by the authors. The authors take full responsibility for the integrity, accuracy, and originality of all manuscript content.

### Ethics Approval

Each contributing cohort obtained approval from its responsible research ethics committee or institutional review board, and written informed consent from all participants, in accordance with the Declaration of Helsinki. Approving committees and permit numbers for each cohort are listed in the Supplementary Information.

## Acknowledgements

We are grateful for the participation of all individuals across the various cohorts of this study.

## Author contributions

Cohort specific data collection: I.M.L., L.M., P.K.E.M., E.S., T.P., S.J.P., H.S., I.M.N., C.C.M., A.B.S.G., H.L., A.T., C.L.S., D.I.C. F.M.K.W, S.P., K.C., H.J.H., C.M.L., V.G., A.S., G.G.N., M.P.C. Cohort specific analysis: N.M.A.S., N.T., M.B.F., L.B., E.B.R.T., K.K., Y.Z., M.N., D.I.C., F.M.K.W., H.S., I.M.N., A.G., J.K., E.S., R.E., M.P.C., S.C., G.G.N., L.I., H.L., N.L.H.C., F.C. Meta-analysis and post-GWA analysis: N.M.A.S., S.Y., M.K.N., N.T., M.B.F, C.R.C. Critical revision of manuscript: C.R.C., S.J.P., M.B.F., F.M.K.W.. A.P.N., M.N., J.M.F., J.V.B.H., K.C., P.K.E.M., A.B-V. Writing editing: C.R.C., S.J.P., N.M.A.S., A.B-V., H.J.H. Overseeing project: C.R.C., F.M.K.W., M.B.F., A.G., A.P.N., S.J.P., S.G., J.M.F., J.V.B.H., K.C. Study concept and design: C.R.C., F.M.K.W., M.B.F., A.G., A.P.N., S.J.P., S.G., P.S.

## Group Name

The Estonian Biobank Research Team is composed of Andres Metspalu, Tõnu Esko, Reedik Mägi, Mait Metspalu, Mari Nelis and Georgi Hudjashov. Contact

Lifelines Cohort Study is composed of Raul Aguirre-Gamboa, Patrick Deelen, Lude Franke, Jan A Kuivenhoven, Esteban A Lopera Maya, Ilja M Nolte, Serena Sanna, Harold Snieder, Morris A Swertz, Peter M. Visscher, Judith M Vonk, Cisca Wijmenga, Naomi Wray

