## Supplementary material for "Integrating GWAS meta-analysis with human brain cell mapping implicates the amygdala and cortical inhibitory interneurons in tinnitus"

Supplementary information

Contents:

1. **Cohort descriptions**
2. **Supplementary figures**
3. **References**
4. **Study specific acknowledgements**

#### 1. Cohort descriptions

##### Age, Gene/Environment Susceptibility Reykjavik Study: AGES-Reykjavik Study

The Reykjavik Study cohort originally comprised a random sample of 30,795 men and women born in 1907-1935 and living in Reykjavik in 1967[1]. A total of 19,381 people attended, resulting in 71% recruitment rate. The study sample was divided into six groups by birth year and birth date within month. One group was designated for longitudinal follow up and was examined in all stages. One group was designated a control group and was not included in examinations until 1991. Other groups were invited to participate in specific stages of the study. Between 2002 and 2006, the AGES-Reykjavik study re-examined 5764 survivors of the original cohort who had participated before in the Reykjavik Study. The study is approved by the Icelandic National Bioethics Committee, VSN: 00-063.

##### The Danish Twin Registry

The Danish Twin Registry (DTR) sample included 1,314 individuals (137 cases and 1177 controls) collected as part of the study of Middle Age Danish Twins (MADT, N=1,055) and the Longitudinal Study of Aging Danish Twins (LSADT, N=259)[2]. MADT was initiated in 1998 and includes 4,314 twins randomly chosen from the birth years 1931-1952. Surviving participants were revisited from 2008 to 2011, where the blood samples used in the present study were collected. The survey data used in the present study was obtained from the Omnibus 2 survey undertaken in 2002[2]. LSADT was initiated in 1995 and includes twins aged 70 years and older. Follow-up assessments were conducted every second year through 2005. The individuals included here all participated in the 1997 assessment, where blood samples were collected from same sex twin pairs, and in the 2001 assessment where the survey data used in the present study was collected[2].

Written informed consents were obtained from all participants. Collection and use of biological material, and survey and registry information were approved by the Regional Scientific Ethical Committees for Southern Denmark (MADT: S-VF-19980072, LSADT: S-VF-20040241). The study is registered in SDU’s internal list (notification no. 10.903) and complies with the rules in the General Data Protection Regulation.

##### EstBB

The Estonian Biobank (EstBB) is a large data-rich population-based biobank, covering approximately 20% of the adult population in Estonia (N=~210,000). All EstBB participants have signed an informed consent form and provided blood samples for genotyping. Electronic health records are regularly retrieved by linking to the national health databases and registries, such as the National Health Insurance Funds (NHIF) database, cause of death register, and hospital records[3].

For the current study, we identified 8,843 tinnitus cases from participants' EHRs, defined as individuals with at least two occurrences of the ICD-10 diagnosis code H93.1. Controls (n= 186,551) were defined as undiagnosed participants. Individuals with H93.2 (Other abnormal auditory perceptions), Z96.2 (Presence of otological and audiological implants), Z97.4 (Presence of external hearing-aid) ICD10 diagnoses were excluded from all analyses. The analysis was performed using REGENIE version 2.2.4[4], with gender, birth year, and the first 10 principal components included as covariates.

The activities of the EstBB are regulated by the Human Genes Research Act, which was adopted in 2000 specifically for the operations of the EstBB. Individual level data analysis in the EstBB was carried out under ethical approval 1.1-12/624 from the Estonian Committee on Bioethics and Human Research (Estonian Ministry of Social Affairs), using data according to release application 6-1/GI/131 from the Estonian Biobank.

**FinnGen**

The FinnGen research project (www.finngen.fi) was launched in 2017 with an aim to improve human health through genetic research. The project combines genome information with digital health care data from national registries. The genotype data are linked to national hospital discharge, death, cancer, and medication reimbursement registries using the national personal identification numbers. The FinnGen study aims to combine approximately 200,000 existing samples from Finnish biobanks with approximately 300,000 samples from ongoing collections. Once final, the data resource will cover roughly 10% of the Finnish population. The present study comprised data of 447,339 Finnish adults (10,008 cases and 437,331 controls) from FinnGen Preparatory Phase Data Freeze 10.

Patients and control subjects in FinnGen provided informed consent for biobank research, based on the Finnish Biobank Act. Alternatively, separate research cohorts, collected prior the Finnish Biobank Act came into effect (in September 2013) and start of FinnGen (August 2017), were collected based on study-specific consents and later transferred to the Finnish biobanks after approval by Fimea (Finnish Medicines Agency), the National Supervisory Authority for Welfare and Health. Recruitment protocols followed the biobank protocols approved by Fimea. The Coordinating Ethics Committee of the Hospital District of Helsinki and Uusimaa (HUS) statement number for the FinnGen study is Nr HUS/990/2017.

The FinnGen study is approved by Finnish Institute for Health and Welfare (permit numbers: THL/2031/6.02.00/2017, THL/1101/5.05.00/2017, THL/341/6.02.00/2018, THL/2222/6.02.00/2018, THL/283/6.02.00/2019, THL/1721/5.05.00/2019 and THL/1524/5.05.00/2020), Digital and population data service agency (permit numbers: VRK43431/2017-3, VRK/6909/2018-3, VRK/4415/2019-3), the Social Insurance Institution (permit numbers: KELA 58/522/2017, KELA 131/522/2018, KELA 70/522/2019, KELA 98/522/2019, KELA 134/522/2019, KELA 138/522/2019, KELA 2/522/2020, KELA 16/522/2020), Findata permit numbers THL/2364/14.02/2020, THL/4055/14.06.00/2020, THL/3433/14.06.00/2020, THL/4432/14.06/2020, THL/5189/14.06/2020, THL/5894/14.06.00/2020, THL/6619/14.06.00/2020, THL/209/14.06.00/2021, THL/688/14.06.00/2021, THL/1284/14.06.00/2021,

THL/1965/14.06.00/2021, THL/5546/14.02.00/2020, THL/2658/14.06.00/2021, THL/4235/14.06.00/2021, Statistics Finland (permit numbers: TK-53-1041-17 and TK/143/07.03.00/2020 (earlier TK-53-90-20) TK/1735/07.03.00/2021, TK/3112/07.03.00/2021) and Finnish Registry for Kidney Diseases permission/extract from the meeting minutes on 4th July 2019.

The Biobank Access Decisions for FinnGen samples and data utilized in FinnGen Data Freeze 10 include: THL Biobank BB2017_55, BB2017_111, BB2018_19, BB_2018_34, BB_2018_67, BB2018_71, BB2019_7, BB2019_8, BB2019_26, BB2020_1, BB2021_65, Finnish Red Cross Blood Service Biobank 7.12.2017, Helsinki Biobank HUS/359/2017, HUS/248/2020, HUS/150/2022 § 12, §13, §14, §15, §16, §17, §18, and §23, Auria Biobank AB17-5154 and amendment #1 (August 17 2020) and amendments BB_2021-0140, BB_2021-0156 (August 26 2021, Feb 2 2022), BB_2021-0169, BB_2021-0179, BB_2021-0161,  AB20-5926 and amendment #1 (April 23 2020)and it´s modification (Sep 22 2021), Biobank Borealis of Northern Finland_2017_1013, 2021_5010, 2021_5018, 2021_5015, 2021_5023, 2021_5017, 2022_6001,  Biobank of Eastern Finland 1186/2018 and amendment 22 § /2020, 53§/2021, 13§/2022, 14§/2022, 15§/2022, Finnish Clinical Biobank Tampere MH0004 and amendments (21.02.2020 & 06.10.2020), §8/2021, §9/2022, §10/2022, §12/2022, §20/2022, §21/2022, §22/2022, §23/2022, Central Finland Biobank 1-2017, and Terveystalo Biobank STB 2018001 and amendment 25th Aug 2020, Finnish Hematological Registry and Clinical Biobank decision 18th June 2021, Arctic biobank P0844: ARC_2021_1001.

**Framingham Heart Study**

The Framingham Heart Study is a prospective longitudinal investigation of the development of atherosclerosis and its clinical sequelae. Study participants were recruited across three time periods. The study was initiated between 1948 and 1950 with the recruitment of 5,209 individuals ages 28 to 62 (including some spouse pairs, parent-offspring pairs and siblings) for the purpose of investigating the multiple factors involved in the development of cardiovascular disease[5]. This group, known as the "original cohort", has been examined every two years with a total of 32 examinations to date. Between 1971 and 1975, "offspring" of the original cohort and the offspring of spouses of the original cohort were recruited to examine among other goals the familial components of cardiovascular disease and its risk factors[6]. Between 2002 and 2005, the third generation (children of the offspring and grandchildren of the original cohort) was recruited[7]. The offspring cohort totalled 5,124 and the third generation totalled 4,095 at recruitment and have been examined every 4 to 8 years. The offspring cohort now has nice examinations completed and the third generation has two examinations completed.

**G-EAR**

Within the International consortium called G-EAR, we used individuals coming from the **Salus in Apulia Study** (formerly known as Great Age Study).

The “Salus in Apulia Study” is an ongoing population-based prospective cohort comprising 2,472 individuals aged 65 years and older and residing in Castellana Grotte, a town located near Bari, Puglia, in the Southeast of Italy. It focuses on the analysis of the impact of lifestyle habits on frailty, other age-related impairments and age-related disease outcomes. Salus is a public health initiative funded by the Italian Ministry of Health and Apulia Regional Government and carried on at IRCCS “S. De Bellis” that combines data from two previous populations: the baseline data (MICOL3, M3) were recorded from 2003 to 2005 and the follow-up data collected from 2013 to 2015 (GreatAGE Study - MICOL4, M4). The GreatAGE-M4 study has been described elsewhere[8]. The invitation also included MICOL participants who were in the correct age range (age 64 years and older). In the GreatAge-M4 examination, in addition to the assessment of clinical and lifestyle aspects, sensory-related outcomes were also evaluated together with neuropsychological features and genetic components. The study adhered to the “Standards for Reporting Diagnostic Accuracy Studies” (STARD) guidelines (http://www.stard-statement.org/), the “Strengthening the Reporting of Observational Studies in Epidemiology” (STROBE) guidelines (https://www.strobe-statement.org/).

**Health, Aging, and Body Composition (HABC) Study**

The HABC Study is a NIA-sponsored cohort study of the factors that contribute to incident disability and the decline in function of healthier older persons, with a particular emphasis on changes in body composition in old age. Between March 1997 and July 1998, 3,075 community-dwelling adults between the ages of 70 and 79 years (41% African American) were recruited to participate. Medicare beneficiary listings were used to recruit in metropolitan areas surrounding Pittsburgh, Pennsylvania, and Memphis, Tennessee. Eligibility criteria included having no difficulty walking one-quarter of a mile, climbing ten steps, or performing activities of daily living (transferring, bathing, dressing, and eating); no history of active treatment for cancer in the prior 3 years; and no plans to move from the area within 3 years.

**The Lifelines Cohort Study**

Lifelines[9,10] is a multi-disciplinary prospective population-based cohort study examining in a unique three-generation design the health and health-related behaviors of 167,729 persons living in the northern Netherlands. It employs a broad range of investigative procedures in assessing the biomedical, socio-demographic, behavioral, physical and psychological factors which contribute to the health and disease of the general population, with a special focus on multi-morbidity and complex genetics. Genotyping data was obtained in the original GWAS[11] (including approximately 15,000 individuals) and via the UMCG Genetics Lifelines Initiative (UGLI), which genotyped approximately 66,500 individuals. Additional information on phenotyping and genotyping in Lifelines can be found here: https://wiki.lifelines.nl/.

The general Lifelines protocol has been approved by the UMCG Medical ethical committee under number 2007/152.

**Moli Sani**

"Moli-sani" (www.moli-sani.org) is a cohort study aiming at evaluating the risk factors (environmental, genetics, biomolecular) linked to chronic-degenerative disease, especially cancer, cardiovascular disease and their intermediate phenotypes hypertension, diabetes, dyslipidemia, obesity and metabolic syndrome. Between March 2005 and April 2010, the study recruited participants 35-year-old residents of the Molise region through multistage sampling from city hall registries. First, townships in major areas were selected by cluster sampling. Within each township, participants aged 35 years and older were selected by simple random sampling. For each selected person, all nuclear family members aged 35 years and older were also invited to participate. This approach allowed construction of a large family pedigree. Exclusion criteria were pregnancy at the time of recruitment, disturbances in understanding or willingness, current poly-traumas or coma, or refusal to sign the informed consent. The Moli-sani study was approved by the ethics committee of the Catholic University of Rome and is conducted under the supervision of both the Bioethics Institute of the Catholic University of Romethe and the Istituto Superiore di Sanità, Rome. All participants signed an informed consent before taking part in the study. This study was conducted in accordance with the Declaration of Helsinki.

**The Rotterdam Study**

The Rotterdam study is a prospective, population-based cohort study among inhabitants of Ommoord, a district of Rotterdam, The Netherlands[12]. As of 2008, the cohort is comprised of 14,926 subjects aged 45 years and older. Since 2016, it has been expanded to include particpatns aged 40 years and older. The Rotterdam study targets cardiovascular, endocrine, hepatic, neurological, ophthalmic, psychiatric, dermatological, otolaryngological, locomotor, and respiratory diseases. The participants were all examined in detail at baseline. They were interviewed at home (2 h) and then had an extensive set of examinations (totaling 5 h) in a specially built research facility in the center of the district. Written informed consent was obtained from all participants and the Medical Ethics Committee of the Erasmus Medical Center, Rotterdam, approved the study.

**The Swedish Twin Registry**

The Swedish Twin Registry (STR) included in this study was pulled from two national cohorts: 1) the Screening Across the Lifespan Twin Study (SALT), a telephone interview study conducted between 1998 and 2002 and including twins born before 1959 and 2) the Study of Twin Adults: Genes and Environment (STAGE), an online questionnaire study conducted between 2005 and –2006 and including twins born between 1959 and 1985[13]. Phenotypic information about hearing was collected though self-reports in these studies. Additional phenotypic information was available from a paper questionnaire (called SALTY) administered in 2009 to SALT participants born between 1944 and 1985. DNA was collected from blood or saliva and extracted by standard procedures. All genotyping was performed by SNP&SEQ genotyping facility in Uppsala, in three waves using three different Illumina chip arrays, OmniExpress for the blood DNA, Psychchip and Global screening array for the saliva DNA samples. Written informed consents were obtained from all participants.

**TwinsUK**

TwinsUK is the only adult twin registry in the UK, comprising of over 12,000 healthy twin volunteers between 16 and 98 years of age[14]. Collection of data and biologic materials commenced in 1992 and is ongoing. Twins have completed detailed health and lifestyle questionnaires and attended clinical evaluations. Pure tone audiometry data was collected in a subset of the cohort (N=1,242) between April 2010 and November 2012. Participants were recruited with an aim to study aging in females. An air-conduction pure-tone audiogram was conducted by trained personnel using a Madsen XETA audiometer including TDH39 headphones. All research was conducted according to the ethical standards as defined by the Helsinki declaration. Ethical approval for this study was obtained from the National Research Ethics service London-Westminster (REC reference number: 07/H0802/84). Written informed consent was obtained from all participants prior to study conduction. Participants were excluded from analysis based on missing data, male, age less than 45 years. There were 819 female participants aged 45 years and older remaining for analysis.

**UK Biobank**

The UK Biobank[15] is a large prospective cohort study with over 500,000 participants aged 40 to 69 years old. Recruitment occurred between 2006 and 2010. The study collected extensive phenotype and genotype data about the participants. Data includes questionnaires, physical measures, multimodal imaging and genome-wide genotyping. Follow-up data is also available for a wide range of health-related outcomes. The UK Biobank is available for researchers worldwide. This research has been conducted using the UK Biobank Resource under application number #74463.

#### 2. Supplementary figures

**Supplementary Figure 1**: Quantile-Quantile plot of association test results from the meta-analysis of tinnitus.

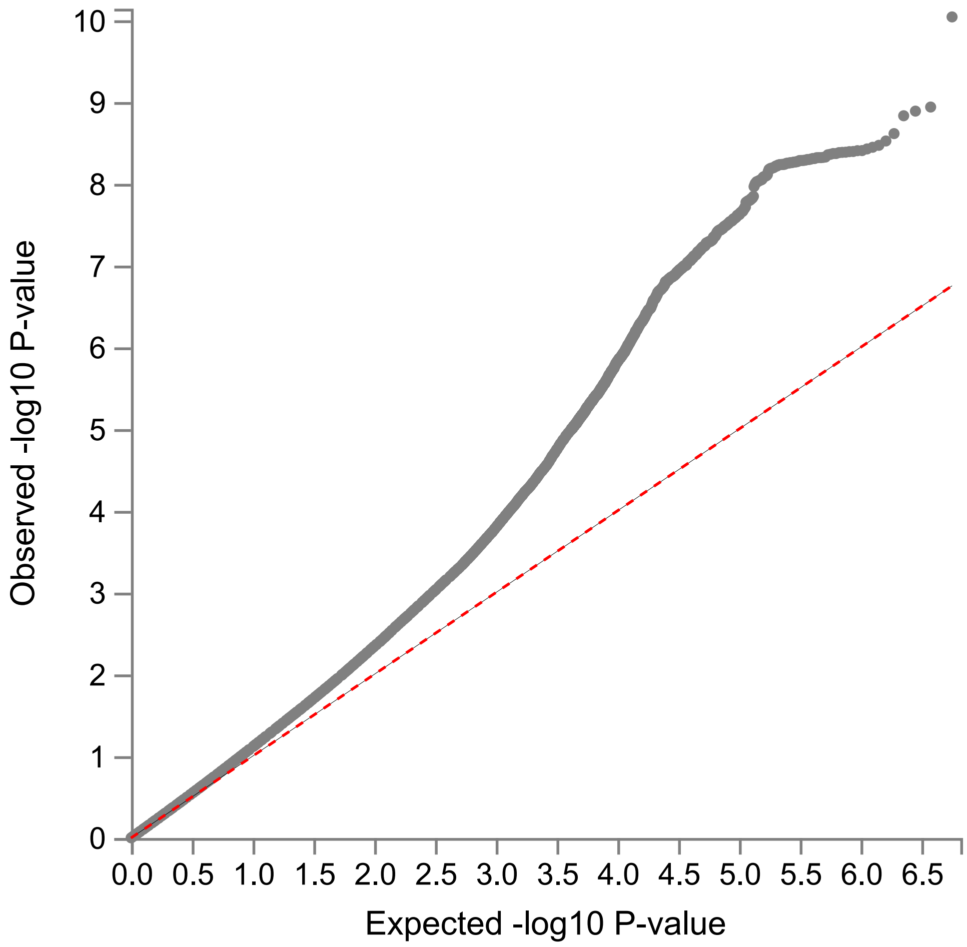

**Supplementary Figure 2**: Quantile-Quantile plot of association test results from the meta-analysis of tinnitus.

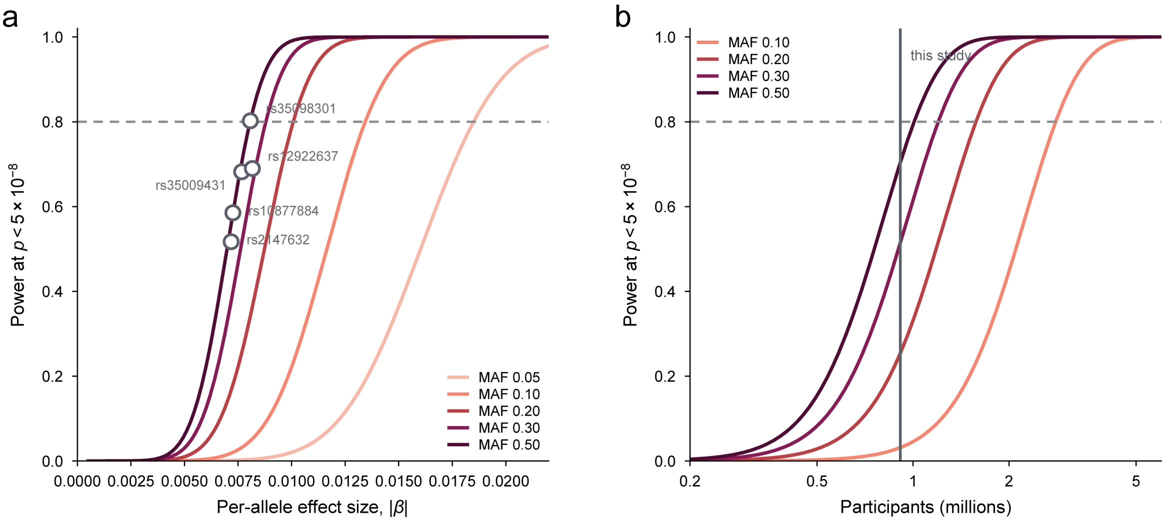

**Supplementary Figure 3**: Regional plots of significant and independent loci (A-E). Each regional plot shows chromosome position on the x-axis and -log10 p-value on the y-axis (from the meta-analysis). Colors and legend correspond to LD information. Gene annotations are shown just above the x-axis. Regional plots are generated using LocusZoom. Detailed information about the independent loci can be found in **Supplementary Table S3**.

a

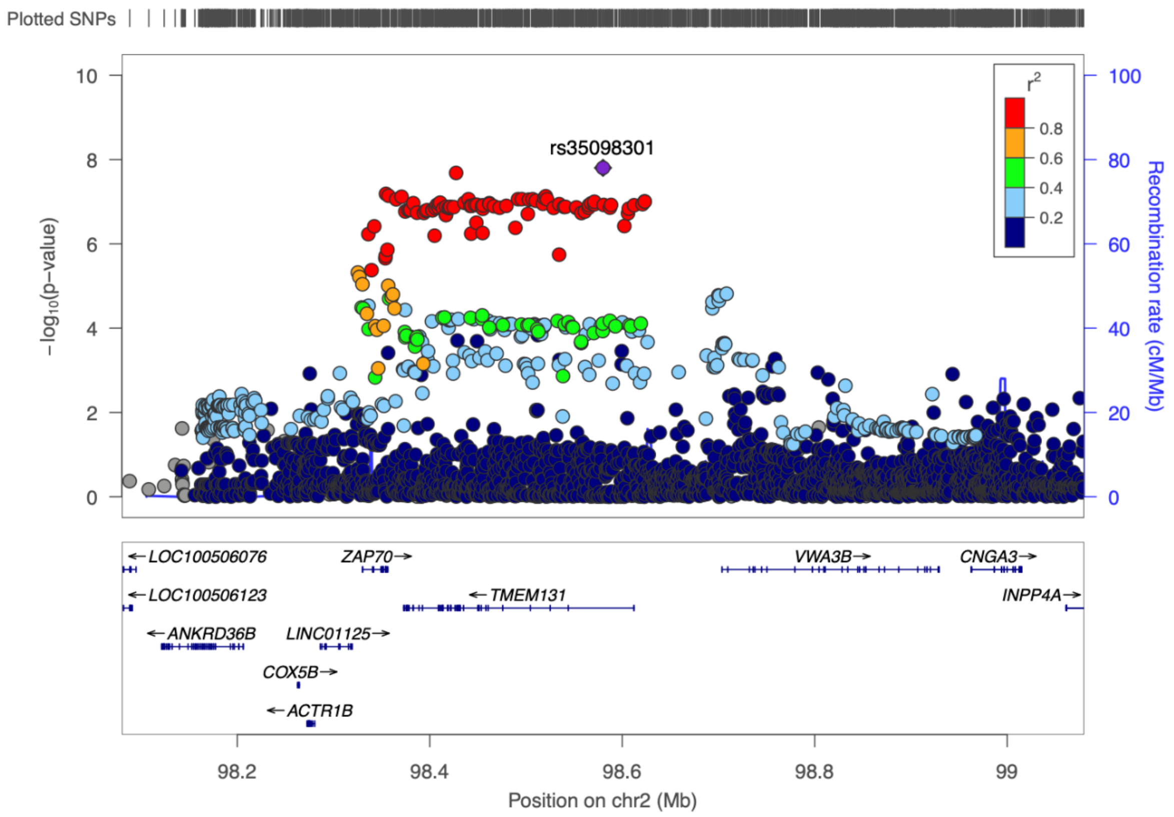

**b**

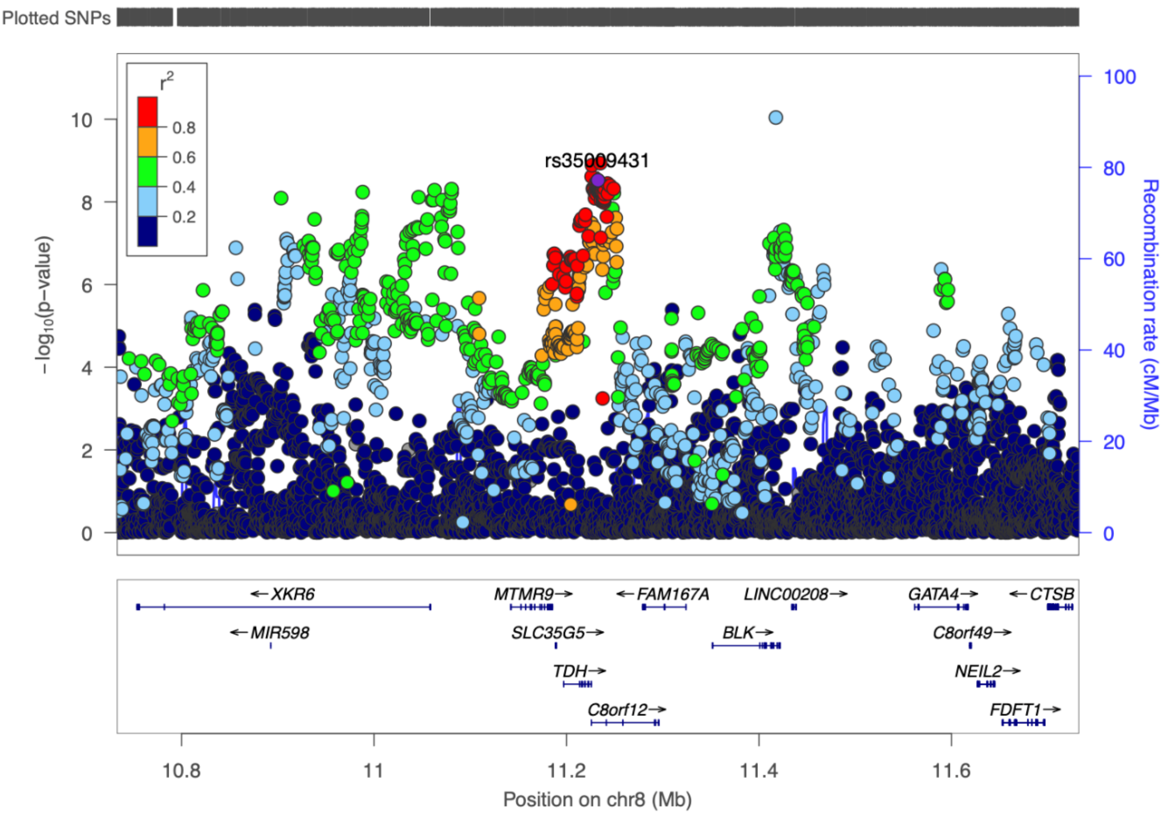
Plotted SNPs
r²
0.8
0.6
0.4
0.2
rs35009431
−log₁₀(p−value)
Recombination rate (cM/Mb)
←XKR6
←MIR598
MTMR9→
SLC35G5→
TDH→
C8orf12→
←FAM167A
BLK→
LINC00208→
GATA4→
C8orf49→
NEIL2→
FDFT1→
←CTSB
10.8
11
11.2
11.4
11.6
Position on chr8 (Mb)

**c**

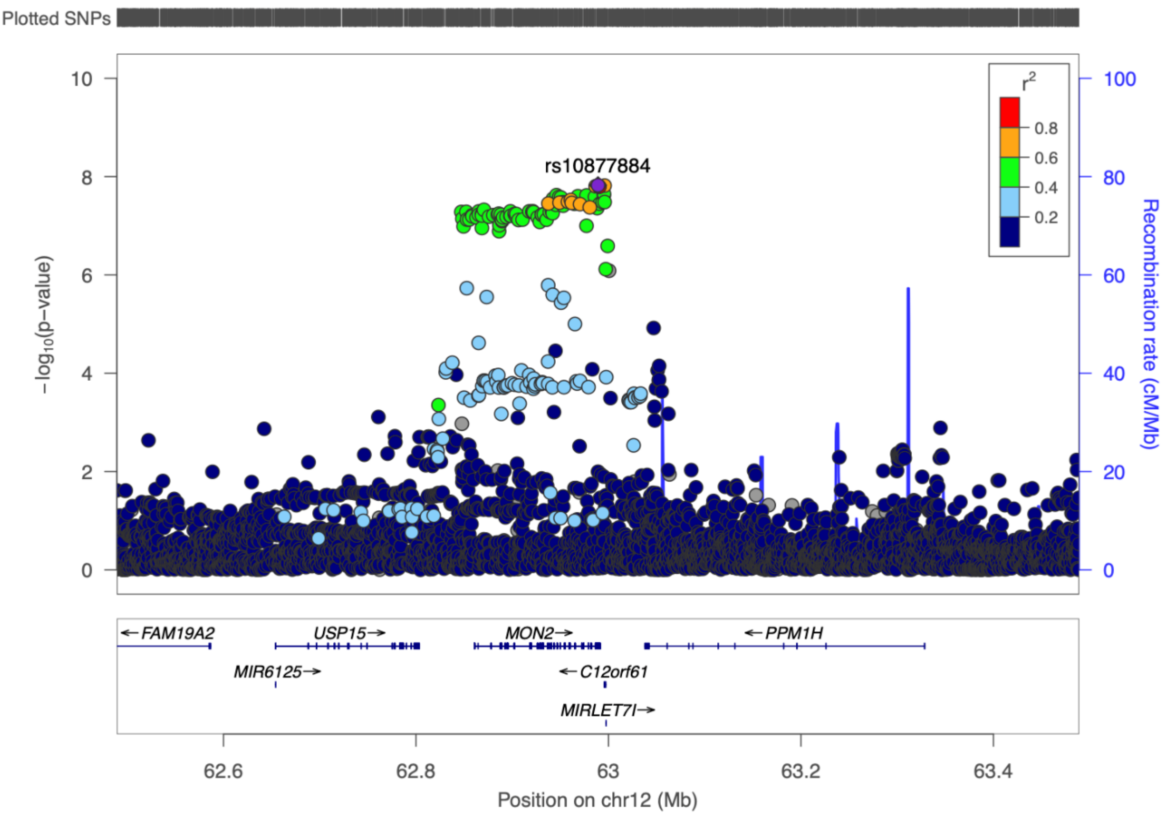
Plotted SNPs
r²
0.8
0.6
0.4
0.2
rs10877884
−log₁₀(p−value)
Recombination rate (cM/Mb)
←FAM19A2
MIR6125→
USP15→
MON2→
←C12orf61
MIRLET7I→
←PPM1H
62.6
62.8
63
63.2
63.4
Position on chr12 (Mb)

d

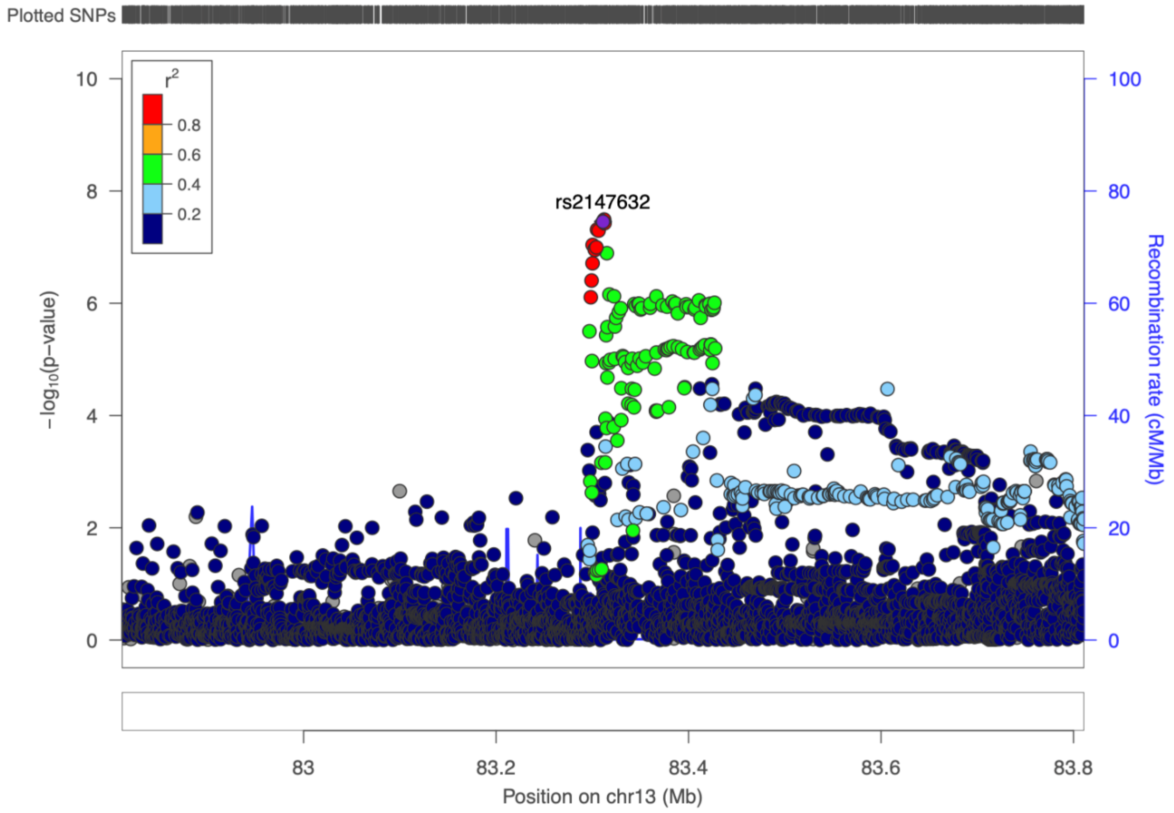
Plotted SNPs
r2
0.8
0.6
0.4
0.2
rs2147632
−log10(p−value)
Recombination rate (cM/Mb)
0
2
4
6
8
10
0
20
40
60
80
100
83
83.2
83.4
83.6
83.8
Position on chr13 (Mb)

e

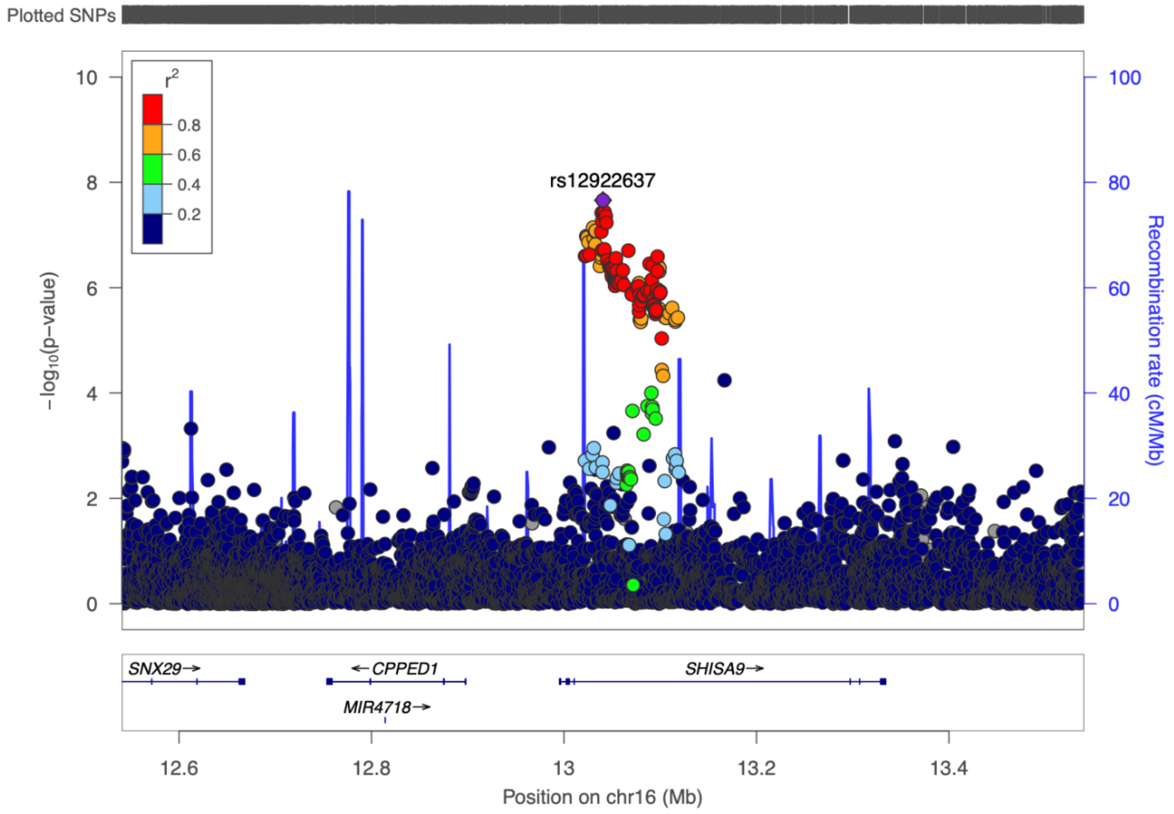
Plotted SNPs
r2
0.8
0.6
0.4
0.2
rs12922637
−log10(p−value)
Recombination rate (cM/Mb)
0
2
4
6
8
10
0
20
40
60
80
100
SNX29→
←CPPED1
MIR4718→
SHISA9→
12.6
12.8
13
13.2
13.4
Position on chr16 (Mb)

**Supplementary Figure 4**: Venn diagram of the different gene mapping methods. Detailed information about the mapped genes is available in **Supplementary tables S5, S6 and S7**.

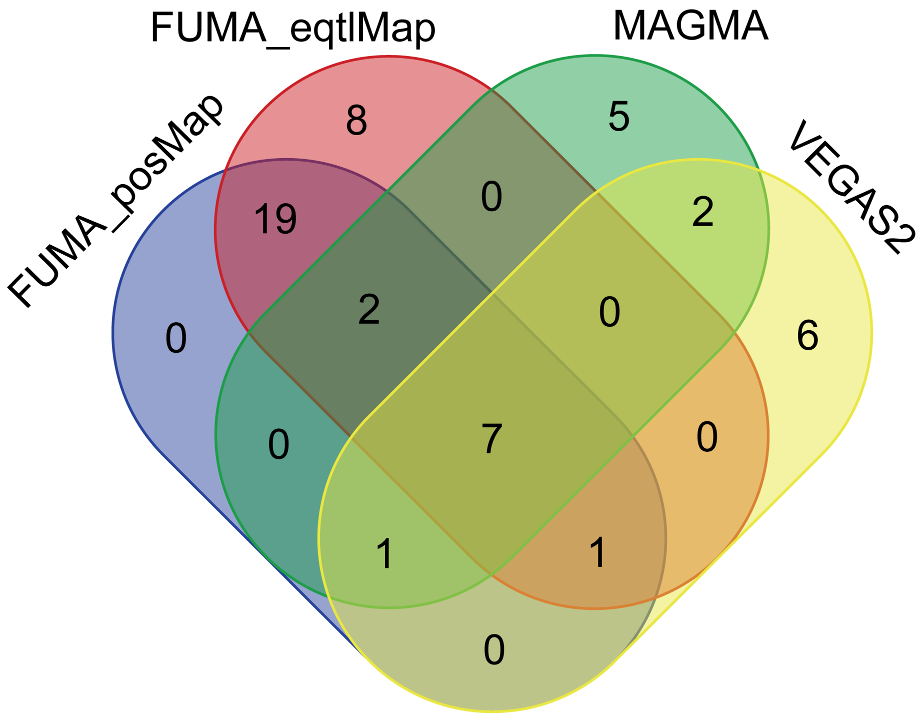

**Supplementary Figure 5**: **Enrichment of brain cell superclusters in other traits.** Each panel includes results on a single supercluster. On the y-axis are the following traits: hearing loss, major depressive disorder, insomnia, neuroticism, schizophrenia and bipolar disorder (from Yao et al., 2025). On the x-axis are the –log10 p-values. An FDR < 0.1 was considered suggestive significant (red). Detailed information in **Supplementary table S13**. Abbreviations: CGE = caudal ganglionic eminence, COP = committed oligodendrocyte precursor, CT = corticothalamic, MSN = medium spiny neuron, LLC = LAMP5-LHX6 and Chandelier, MGE = medial ganglionic eminence, ITC = intratelencephalic.

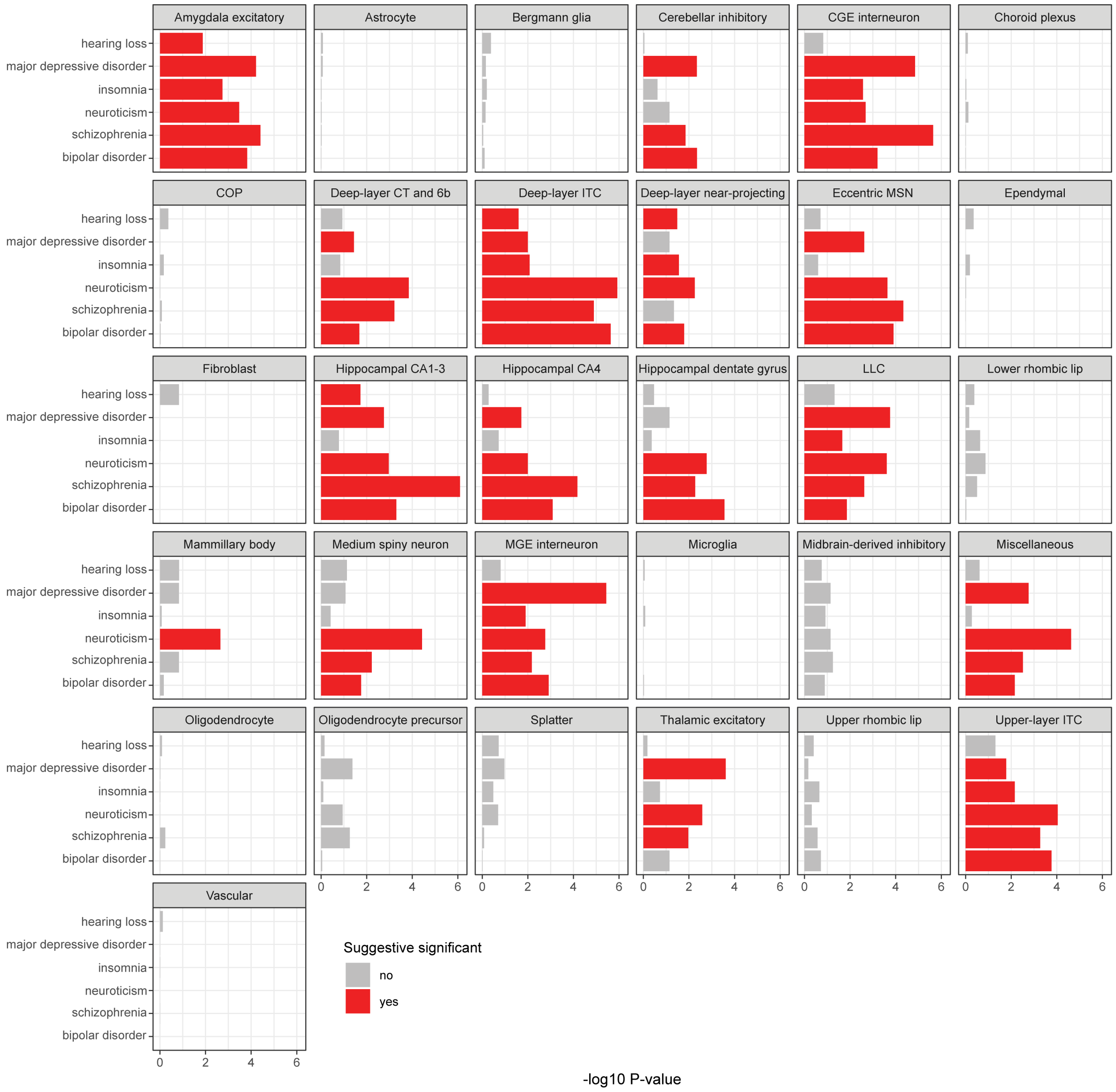

#### 4. Study specific acknowledgements

The **AGES-Reykjavik** study has been funded by NIH contract N01-AG012100, the NIA Intramural Research Program, an Intramural Research Program Award (ZIAEY000401) from the National Eye Institute, an award from the National Institute on Deafness and Other Communication Disorders (NIDCD) Division of Scientific Programs (IAA Y2-DC_1004-02), Hjartavernd (the Icelandic Heart Association), and the Althingi (the Icelandic Parliament). The researchers are indebted to the participants for their willingness to participate in the study.

**DTR** has been supported by grants from The National Program for Research Infrastructure 2007 from the Danish Agency for Science, Technology and Innovation (09-063256) and the US National Institutes of Health (P01 AG08761). Genotyping was supported by NIH R01 AG037985 (Pedersen).

The **FinnGen** project is funded by two grants from Business Finland (HUS 4685/31/2016 and UH 4386/31/2016) and the following industry partners: AbbVie Inc., AstraZeneca UK Ltd, Biogen MA Inc., Bristol Myers Squibb (and Celgene Corporation & Celgene International II Sàrl), Genentech Inc., Merck Sharp & Dohme Corp, Pfizer Inc., GlaxoSmithKline Intellectual Property Development Ltd., Sanofi US Services Inc., Maze Therapeutics Inc., Janssen Biotech Inc, Novartis AG, and Boehringer Ingelheim. The following biobanks are acknowledged for delivering biobank samples to FinnGen: Auria Biobank (www.auria.fi/biopankki), THL Biobank (www.thl.fi/biobank), Helsinki Biobank (www.helsinginbiopankki.fi), Biobank Borealis of Northern Finland (https://www.ppshp.fi/Tutkimus-ja-opetus/Biopankki/Pages/Biobank-Borealis-briefly-in-English.aspx), Finnish Clinical Biobank Tampere (www.tays.fi/en-US/Research_and_development/Finnish_Clinical_Biobank_Tampere), Biobank of Eastern Finland (www.ita-suomenbiopankki.fi/en), Central Finland Biobank (www.ksshp.fi/fi-FI/Potilaalle/Biopankki), Finnish Red Cross Blood Service Biobank (www.veripalvelu.fi/verenluovutus/biopankkitoiminta) and Terveystalo Biobank (www.terveystalo.com/fi/Yritystietoa/Terveystalo-Biopankki/Biopankki/). All Finnish Biobanks are members of BBMRI.fi infrastructure (www.bbmri.fi). Finnish Biobank Cooperative -FINBB (https://finbb.fi/) is the coordinator of BBMRI-ERIC operations in Finland. The Finnish biobank data can be accessed through the Fingenious® services (https://site.fingenious.fi/en/) managed by FINBB.The researchers would like to acknowledge all participants and investigators of the FinnGen study.

The **Framingham Heart Study** is conducted and supported by the National Heart, Lung, and Blood Institute (NHLBI) in collaboration with Boston University (Contract No. N01-HC-25195 and HHSN268201500001). This manuscript was not prepared in collaboration with investigators of the

Framingham Heart Study and does not necessarily reflect the opinions or views of the Framingham Heart Study, Boston University, or NHLBI. Funding for SHARe Affymetrix genotyping was provided by NHLBI Contract N02-HL- 64278. Nancy L. Heard-Costa is supported by NIH/NHLBI HHSN268201500001 (Ramachandran). Further support was provided by P30AG066546, funded by the National Institute on Aging (HHS - NIH), awarded to University of Texas Health Science Center of San Antonio.

**The Great Age study (Salus in Apulia study)** were funded by Apulia Government and Italian Ministry of Health, under the Studies on Aging Network, at Italian Research Hospitals (IRCCS). The authors thank the MICOL Study group, the Salus in Apulia Research Team, and the General Practitioners of Castellana Grotte for the fundamental role in recruiting participants to this study.

The **Health, Aging and Body Composition Study** was supported by the Intramural Research Program, National Institute on Aging, National Institutes of Health, Department of Health and Human Services, project ZO1 AG000949. This work utilized the computational resources of the NIH STRIDES Initiative (https://cloud.nih.gov) through the Other Transaction agreement - Azure: OT2OD032100, Google Cloud Platform: OT2OD027060, Amazon Web Services: OT2OD027852. This work utilized the computational resources of the NIH HPC Biowulf cluster (https://hpc.nih.gov). The Health, Aging, and Body Composition Study (Health ABC) was supported by National Institute on Aging (NIA) Contracts N01-AG-6-2101; N01-AG-6-2103; N01-AG-6-2106; NIA grant R01-AG028050, and NINR grant R01-NR012459. This research was funded in part by the Intramural Research Program of the NIH, National Institute on Aging.

The **Lifelines** initiative has been made possible by subsidy from the Dutch Ministry of Health, Welfare and Sport, the Dutch Ministry of Economic Affairs, the University Medical Center Groningen (UMCG), Groningen University and the Provinces in the North of the Netherlands (Drenthe, Friesland, Groningen). The generation and management of GWAS genotype data for the Lifelines Cohort Study is supported by the UMCG Genetics Lifelines Initiative (UGLI). UGLI is partly supported by a Spinoza Grant from NWO, awarded to Cisca Wijmenga. The authors wish to acknowledge the services of the Lifelines Cohort Study, the contributing research centers delivering data to Lifelines, and all the study participants.

The **Rotterdam Study** is funded by Erasmus Medical Center and Erasmus University, Rotterdam, Netherlands Organisation for the Health Research and Development (ZonMw), the Research Institute for Diseases in the Elderly (RIDE), the Ministry of Education, Culture and Science, the Ministry for Health, Welfare and Sports, the European Commission (DG XII), and the Municipality of Rotterdam. The authors are grateful to the study participants, the staff from the Rotterdam Study and the participating general practitioners and pharmacists. The generation and management of GWAS genotype data for the Rotterdam Study (RS I, RS II, RS III) was executed by the Human Genotyping Facility of the Genetic Laboratory of the Department of Internal Medicine, Erasmus MC, Rotterdam, The Netherlands. The GWAS datasets are supported by the Netherlands Organization of Scientific Research NWO Investments (nr. 175.010.2005.011, 911-03-012), the Genetic Laboratory of the Department of Internal Medicine, Erasmus MC, the Research Institute for Diseases in the Elderly (014-93-015; RIDE2), the Netherlands Genomics Initiative (NGI)/Netherlands Organization for Scientific Research (NWO) Netherlands Consortium for Healthy Aging (NCHA), project nr. 050-060-810. We thank Pascal Arp, Mila Jhamai, Marijn Verkerk, Lizbeth Herrera and Marjolein Peters, MSc, and Carolina Medina-Gomez, MSc, for their help in creating the GWAS database, and Karol Estrada, PhD, Yurii Aulchenko, PhD, and Carolina Medina-Gomez, MSc, for the creation and analysis of imputed data.

For the **TwinsUK** study, the authors of this paper wish to express their appreciation to all study participants.. TwinsUK is funded by the Wellcome Trust, Medical Research Council, European Union, the National Institute for Health Research (NIHR)-funded BioResource, Clinical Research Facility and Biomedical Research Centre based at Guy’s and St Thomas’ NHS Foundation Trust in partnership with King’s College London. HRRW is funded by Action on Hearing Loss, CJS is funded under a grant from the Chronic Disease Research Foundation (CDRF). FMKW is supported by Arthritis Research UK grant number 20682.

### Supplementary Tables

**Table S1** Summary of samples included in tinnitus meta-analysis

**Table S2** SNP-based h2 by LDSC across cohorts (N > 5000 individuals)

**Table S3** Genome-wide significant loci for tinnitus

**Table S4** Independent loci based on COJO analysis

**Table S5** Leave-one-out meta-analysis (LL_cytosnp)

**Table S6** Genetic correlation across cohorts (N > 5000 individuals)

**Table S7** Comparison of tinnitus phenotypes (meta-analysis vs self-report vs icd coded)

**Table S8** VEGAS2 gene mapping

**Table S9** Gene mapping FUMA

**Table S10** MAGMA gene mapping

**Table S11** Single-nucleus enrichment analysis for tinnitus.

**Table S12** Single nucleus enrichment analysis on hearing loss, major depressive disorder, insomnia, neuroticism, schizophrenia and bipolar disorder

**Table S13** LD score regression genetic correlation analysis

**Table S14** Brain cell-type enrichment in the meta-analysis and in the Million Veteran Program replication

**Table S15** Cluster-level enrichment of tinnitus heritability across the 461 Siletti clusters, with cluster annotation

**Table S16** Pairwise conditional analysis of the nine clusters reaching FDR < 0.10

#### Supplementary Table S1

Summary of samples included in tinnitus meta-analysis

*part 1 of 2 — first column repeated in every part*

| Cohort study | Phenotype | Cases |
|---|---|---|
| AGES | Are you bothered by tinnitus (ringing in your ears)? | Yes |
| Danish Twin Registry | Do you suffer from a buzzing sound in your ears?'; 'Do you suffer from tinnitus/a buzzing in your ears?' | Yes |
| Estonian Genome Center (EstBB) | ICD9 and ICD10 diagnoses of tinnitus | ICD10: H93.1 |
| FinnGen | ICD9 and ICD10 diagnoses oftinnitus | ICD9: 3891; ICD10: H93.1 |
| Framingham Heart Study | Do you have any ringing or buzzing in your ears? | Yes; Right / left / both |
| Great Age Study/Salus in Apulia Study | Do you ever hear sounds in your in your ears like ringing or other noise without an external source? | Yes |
| Health ABC - EUR | Buzzing/ringing in the ear? | Yes |
| Lifelines - GWAS | Do you hear ringing or whistling in your ear/ears? | Yes, always / Yes, sometimes |
| Lifelines - UGLI | Do you hear ringing or whistling in your ear/ears? | Yes, always / Yes, sometimes |
| Lifelines - UGLI2 | Do you hear ringing or whistling in your ear/ears? | Yes, always / Yes, sometimes |
| Moli Sani | Have you ever suffered from tinnitus? | Yes |
| Rotterdam Study - 1 | Do you ever hear sounds in your head or in your ears like ringing/buzzing/peeping without an external source? | Yes, > 1 day/week \| Yes, daily |
| Rotterdam Study - 2 | Do you ever hear sounds in your head or in your ears like ringing/buzzing/peeping without an external source? | Yes, > 1 day/week \| Yes, daily |
| Rotterdam Study - 3 | Do you ever hear sounds in your head or in your ears like ringing/buzzing/peeping without an external source? | Yes, > 1 day/week \| Yes, daily |
| SALT | "Do you have buzzing in your ears?" | Both ears, "One ear", |
| SALTY | "Do you have buzzing in your ears?" | Both ears, "One ear", |
| STAGE | "Do you have buzzing in your ears?" | Both ears, "One ear", |
| TwinsUK | Do you suffer from Tinnitus (buzzing/ringing in the ears)? | Yes |
| UK Biobank | Do you get or have you had noises (such as ringing or buzzing) in your head or in one or both ears that lasts for more than five minutes at a time? | Yes, not most or all of the time / Yes, now a lot of the time / Yes, now some of the time/Yes, but not now, but have in the past |

*part 2 of 2 — first column repeated in every part*

| Cohort study | Controls | N cases | N controls | Total N participants | Prevalence (%) |
|---|---|---|---|---|---|
| AGES | No | 406 | 2728 | 3134 | 12.95 |
| Danish Twin Registry | No | 137 | 1177 | 1314 | 10.43 |
| Estonian Genome Center (EstBB) | Other | 8843 | 186551 | 195394 | 4.526 |
| FinnGen | Other | 10342 | 509868 | 520210 | 1.988 |
| Framingham Heart Study | No | 917 | 2124 | 3041 | 30.15 |
| Great Age Study/Salus in Apulia Study | No | 97 | 1683 | 1780 | 5.449 |
| Health ABC - EUR | No | 406 | 882 | 1288 | 31.52 |
| Lifelines - GWAS | No, never | 2210 | 4350 | 6560 | 33.69 |
| Lifelines - UGLI | No, never | 7058 | 16094 | 23152 | 30.49 |
| Lifelines - UGLI2 | No, never | 7675 | 16797 | 24472 | 31.36 |
| Moli Sani | No | 188 | 1010 | 1198 | 15.69 |
| Rotterdam Study - 1 | Yes, < 1 day/week \| No never | 228 | 921 | 1149 | 19.84 |
| Rotterdam Study - 2 | Yes, < 1 day/week \| No never | 433 | 1449 | 1882 | 23.01 |
| Rotterdam Study - 3 | Yes, < 1 day/week \| No never | 674 | 2441 | 3115 | 21.64 |
| SALT | No | 1827 | 8866 | 10693 | 17.09 |
| SALTY | No | 634 | 3802 | 4436 | 14.29 |
| STAGE | No | 959 | 7351 | 8310 | 11.54 |
| TwinsUK | No | 632 | 1565 | 2197 | 28.77 |
| UK Biobank | No, never | 31918 | 115842 | 147760 | 21.6 |
| | Total | 75584 | 885501 | 961085 | 7.864 |

#### Table S1b

*part 1 of 2 — first column repeated in every part*

| Cohort study | Array Type | Genotyping center |
|---|---|---|
| AGES | Illumina hu370CNV | National Institute of Aging genotyping center |
| Danish Twin Registry | Illumina Infinium PsychArray | SNP&SEQ Technology Platform, Science for Life Laboratory, Uppsala, Sweden (http://snpseq.medsci.uu.se/genotyping/snp-services/) |
| Estonian Genome Center (EstBB) | Illumina Global Screening Array | Genotyping Core Facility of the Institute of Genomics, University of Tartu |
| FinnGen | Several Illumina and Affymetrix FinnGen Axiom arrays | Thermo Fisher Scientific for FinnGen Axiom array, several other places for Illumina arrays |
| Framingham Heart Study | Affymetrix GeneChip Human Mapping 500K Array & 50K supplemental Array | Affymetrix, Inc. |
| Great Age Study/Salus in Apulia Study | Illumina Infinium Global Screening Array (GSA) v1 | IRCCS S. De Bellis, Castellana Grotte, Bari, Italy |
| Health ABC - EUR | Illumina Human1M-Duov3_B | Johns Hopkins University Center for Inherited Disease Research (CIDR), Baltimore, MD, USA |
| Lifelines - GWAS | Illumina HumanCytoSNP-12 v2.1 | Human Genomics facility HuGeF (www.glimdna.org), Erasmus MC / Department of Genetics, UMCG |

| Cohort study | Array Type | Genotyping center |
|---|---|---|
| Lifelines - UGLI | Infinium Global Screening Array® (GSA) MultiEthnic Disease Version 1.0 | Human Genomics facility HuGeF (www.glimdna.org), Erasmus MC / Department of Genetics, UMCG |
| Lifelines - UGLI2 | FinnGen Thermo Fisher Axiom® custom array | Human Genomics facility HuGeF (www.glimdna.org), Erasmus MC / Department of Genetics, UMCG |
| Moli Sani | GSAMD-24v1-0_20011747_A1 (642K)/GSAMD- 24v3-0-EA_20034606_A1 (654K) | IRCSS Burlo Garofolo, Trieste, Italy |
| Rotterdam Study - 1 | Version 3 Illumina Infinium II HumanHap550 SNP chip array | Human Genomics facility HuGeF (www.glimdna.org), Erasmus MC |
| Rotterdam Study - 2 | Version 3 Illumina Infinium II HumanHap550 SNP chip array | Human Genomics facility HuGeF (www.glimdna.org), Erasmus MC |
| Rotterdam Study - 3 | Version 3 Illumina Infinium II HumanHap550 SNP chip array | Human Genomics facility HuGeF (www.glimdna.org), Erasmus MC |
| SALT | Illumina OmniExpress (HumanOmniExpress-12v1_A) | SNP&SEQ Technology Platform, Uppsala, Sweden |
| SALTY | Illumina PsychChip (PsychChip_15048346_A) | SNP&SEQ Technology Platform, Uppsala, Sweden |
| STAGE | Illumina GSA (GSAMD-24v1-0_20011747_A1) | SNP&SEQ Technology Platform, Uppsala, Sweden |
| TwinsUK | Several Illumina arrays (HumanHap3001,2, HumanHap610Q, 1M■Duo and 1.2MDuo 1M) | |
| UK Biobank | UK Biobank Affymetrix Axiom and UK BiLEVE Affymetrix Axiom array | Affymetrix Research Services Laboratory, Santa Clara, California, USA |

*part 2 of 2 — first column repeated in every part*

| Cohort study | Genotype calling | Exclusion on SNPs used for imputation | Exclusion on a per sample bias |
|---|---|---|---|
| AGES | Illumina BeadStudio | Callrate < 97%, HWE p-value < 1e-6, MAF < 1% | Callrate < 95%, sex discrepancy |
| Danish Twin Registry | Illumina GenomeStudio | Call rate <98%, HWE P<10-6, and MAF=0 | Call rate <99%, relatedness and gender mismatch |
| Estonian Genome Center (EstBB) | Illumina GenomeStudio | call-rate < 95%, HWE p-value < 1e-4 | call-rate < 95%, mismatching sex |
| FinnGen | GenCall or GenCall+zCall for Illumina and AxiomGT1 for Affymetrix chip genotypes | Call rate below 98%, minor allele count below 3 and Hardy-Weinberg Equilibrium p-value lower than 1e-06 | Call rate below 95%, heterozygosity exceed ±4SD from the mean, failed sex check and MDS outliers. |
| Framingham Heart Study | Affymetrix BRLMM | MAF <1%, Call rate <97%, HWE < 10-6, Mendelian errors > 1000, not in Hapmap and/or strandedness issues merging with Hapmap | Call rate <97%, autosomal heterozygosity > ± 5SD, excessive Mendelian errors |
| Great Age Study/Salus in Apulia Study | Illumina GenomeStudio | Call rate < 0.97; Hardy-Weinberg P < 1*e-06; MAF < 0.01 | Sample call rate < 0.97, gender check |
| Health ABC - EUR | Illumina GenomeStudio | Call rate <95%, no MAF/HWE filter | Call rate <95%, sex mismatch, excess heterozygosity F cut-off of -0.15 and <- 0.15 for inclusion, ancestry outliers removed, no relatives closer than cousin |
| Lifelines - GWAS | Opticall | Callrate < 99%, HWE p-value < 1e-6, MAF = 0 / Callrate < 95%, HWE p-value < 1e-4 , MAF < 1% | Sex mismatch, pedigree analysis, non-European ancestry |
| Lifelines - UGLI | BeagleCall | Callrate < 99%, HWE p-value < 1e-6, MAF = 0 / Callrate < 95%, HWE p-value < 1e-4 , MAF < 1% | Sex mismatch, pedigree analysis, non-European ancestry |
| Lifelines - UGLI2 | Opticall | Callrate < 99%, HWE p-value < 1e-6, MAF = 0 / Callrate < 95%, HWE p-value < 1e-4 , MAF < 1% | Sex mismatch, pedigree analysis, non-European ancestry |
| Moli Sani | Illumina BeadStudio | Call rate <95%, HWE p-value < 1e-6, MAF>0.01 | Individuals with sex discrepancy, aged less than 18 years, affected by any forms of inherited hearing loss or other diseases potentially leading to hearing defects, as well as subjects exposed to noise or ototoxic medications, were excluded from the analysis. Moreover, considering that in the study cohort some individuals are related, a as a linear mixed model was applied with GEMMA software (doi:10.1038/ng.2310) |
| Rotterdam Study - 1 | Illumina BeadStudio | Call rate <90%, no MAF/HWE filter | Call rate <97.5%, sex mismatch, excess autosomal heterozygosity > 0.336, outliers identified by the IBS clustering analysis |
| Rotterdam Study - 2 | Illumina BeadStudio | Call rate <90%, no MAF/HWE filter | Call rate <97.5%, sex mismatch, excess autosomal heterozygosity > 0.336, outliers identified by the IBS clustering analysis |
| Rotterdam Study - 3 | Illumina BeadStudio | Call rate <90%, no MAF/HWE filter | Call rate <97.5%, sex mismatch, excess autosomal heterozygosity > 0.336, outliers identified by the IBS clustering analysis |
| SALT | Illumina GenomeStudio | Call rate < 0.98, MAF=0, Hardy-Weinberg P < 1.0*10-10 | Call rate < 0.98, excessive heterozygosity (FHET outside +/- 0.2); sex mismatch; non-European ancestry |

| Cohort study | Genotype calling | Exclusion on SNPs used for imputation | Exclusion on a per sample bias |
|---|---|---|---|
| SALTY | Illumina GenomeStudio | Call rate < 0.98, MAF<1%, Hardy-Weinberg P < 1.0*10-10 | Call rate < 0.98, excessive heterozygosity (FHET outside +/- 0.2); sex mismatch; non-European ancestry |
| STAGE | Illumina GenomeStudio | Call rate < 0.98, MAF<1%, Hardy-Weinberg P < 1.0*10-10 | Call rate < 0.98, excessive heterozygosity (FHET outside +/- 0.2); sex mismatch; non-European ancestry |
| TwinsUK | Illumina BeadStudio | Call rate <98% | Mismatch between MZ twins and self-reported sex; non-European ancestry |
| UK Biobank | Affymetrix BRLMM | Multiple criteria as outlined here https://biobank.ctsu.ox.ac.uk/crystal/crystal/docs/genotyping_qc.pdf | Multiple criteria as outlined here https://biobank.ctsu.ox.ac.uk/crystal/crystal/docs/genotyping_qc.pdf |

#### Table S1c

*part 1 of 2 — first column repeated in every part*

| Cohort study | Imputation software | Imputation backbone (NBCI build) | Genome Build |
|---|---|---|---|
| AGES | MaCH | 1000 Genomes phase 1 v3 | GRCh37/Hg19 |
| Danish Twin Registry | IMPUTE2 | 1000G, Phase 3 | GRCh37/Hg19 |
| Estonian Genome Center (EstBB) | Beagle | Estonian population specific imputation reference | GRCh37/Hg19 |
| FinnGen | Eagle/Beagle | SISu v4 all-Finns reference panel | GRCh38/Hg38 |
| Framingham Heart Study | Michigan Server | HRCr1.1 | GRCh37/Hg19 |
| Great Age Study/Salus in Apulia Study | MINIMAC v4 | HRC (Build 37) | GRCh37/Hg19 |
| Health ABC - EUR | TOPMED Imputation Server | TOPMED | GRCh38/Hg38 |
| Lifelines - GWAS | IMPUTE2 | 1000Genomes & GoNL | GRCh37/Hg19 |
| Lifelines - UGLI | Sanger imputation service | HRC | GRCh37/Hg19 |
| Lifelines - UGLI2 | Sanger imputation service | HRCr1.1 | GRCh37/Hg19 |
| Moli Sani | IMPUTE2 | Italian Genome Reference Panel (https://doi.org/10.1038/s41431- 019-0551-x) | GRCh37/Hg19 |
| Rotterdam Study - 1 | MACH/minimac | HRCr1.1 | GRCh37/Hg19 |
| Rotterdam Study - 2 | MACH/minimac | HRCr1.1 | GRCh37/Hg19 |
| Rotterdam Study - 3 | MACH/minimac | HRCr1.1 | GRCh37/Hg19 |
| SALT | Sanger imputation service (EAGLE2, PBWT) | HRCr1.1 | GRCh37/Hg19 |
| SALTY | Sanger imputation service (EAGLE2, PBWT) | HRCr1.1 | GRCh37/Hg19 |
| STAGE | Sanger imputation service (EAGLE2, PBWT) | HRCr1.1 | GRCh37/Hg19 |
| TwinsUK | IMPUTE2 | HRCr1.1 | GRCh37/Hg19 |
| UK Biobank | IMPUTE2 | HRC/1000Genomes | GRCh37/Hg19 |

*part 2 of 2 — first column repeated in every part*

| Cohort study | Data handling and statistical tests software | Lambda GC | Age (mean) | Age (sd) |
|---|---|---|---|---|
| AGES | PLINKv1.9, ProbAbel | 1.018 | 76.35 | 5.4 |
| Danish Twin Registry | Plink, MLMA | 0.9888 | 63.7 | 10.7 |
| Estonian Genome Center (EstBB) | regenie | 1.047 | 53.96 | 16.91 |
| FinnGen | REGENIE | 1.052 | 59.37 | 17.8 |
| Framingham Heart Study | R, LME | 0.9953 | 63.64 | 10.96 |
| Great Age Study/Salus in Apulia Study | GEMMA | 0.9916 | 73.65 | 6.8 |
| Health ABC - EUR | PLINK, Rvtests | 1.02 | 77.59 | 2.83 |
| Lifelines - GWAS | regenie | 1.036 | 45.89 | 10.87 |
| Lifelines - UGLI | regenie | 1.041 | 42.85 | 14.3 |
| Lifelines - UGLI2 | regenie | 1.046 | 43.62 | 13.56 |
| Moli Sani | Gemma 0.98.4 | 1.028 | 64.32 | 9.34 |
| Rotterdam Study - 1 | PLINK, Mach2DAT | 1.046 | 83.37 | 4.42 |
| Rotterdam Study - 2 | PLINK, Mach2DAT | 1.003 | 76.15 | 5.38 |
| Rotterdam Study - 3 | PLINK, Mach2DAT | 1.005 | 62.08 | 6.22 |
| SALT | R, BOLT-LMM | 1 | 58.6 | 8.01 |
| SALTY | R, BOLT-LMM | 1 | 49 | 4.2 |
| STAGE | R, BOLT-LMM | 1.047 | 37.39 | 7.7 |
| TwinsUK | GEMMA | 1 | 60.59 | 13.51 |
| UK Biobank | R, BOLT-LMM | 1.047 | 59.59 | 8.03 |

Abbreviations: ICD = International Classification of Diseases; MAF = minor allele frequency; HWE = Hardy-Weinberg equilibrium

#### Supplementary Table S2

SNP-based h2 by LDSC across cohorts (N > 5000 individuals)

| trait | h2 | h2_se | lambda_gc | mean_chi2 | intercept | intercept_se | ratio | ratio_se | meta |
|---|---|---|---|---|---|---|---|---|---|
| UKB | 0.1199 | 0.0088 | 1.2 | 1.213 | 1.008 | 0.0076 | 0.0382 | 0.036 | no |
| FinnGen | 0.0933 | 0.0171 | 1.093 | 1.097 | 1.032 | 0.0088 | 0.3334 | 0.0902 | no |
| LLgsa | 0.0826 | 0.0285 | 1.059 | 1.045 | 1.017 | 0.0071 | 0.379 | 0.1586 | no |
| LLaffy | 0.0667 | 0.0281 | 1.053 | 1.059 | 1.035 | 0.0073 | 0.5936 | 0.124 | no |
| EBB | 0.0629 | 0.018 | 1.062 | 1.067 | 1.03 | 0.0071 | 0.4538 | 0.1063 | no |
| meta | 0.0535 | 0.0036 | 1.21 | 1.261 | 1.01 | 0.0085 | 0.0394 | 0.0325 | yes |

#### Supplementary Table S3

Genome-wide significant loci for tinnitus ordered from most significant to less significant per chromosome

| chromosome | position | other_allele | reference_allele | eaf | beta | se | p-value | i2 | n_studies | n_samples |
|---|---|---|---|---|---|---|---|---|---|---|
| 2 | 98580012 | T | C | 0.4898 | 0.008074 | 0.001427 | 1.57e-08 | 0.3447 | 16 | 888525 |
| 2 | 98427382 | T | C | 0.4907 | 0.008 | 0.001426 | 2.07e-08 | 0.413 | 15 | 887234 |
| 8 | 11417582 | T | C | 0.625 | 0.01009 | 0.001555 | 9.09e-11 | 0.4954 | 6 | 821622 |
| 8 | 11235150 | C | A | 0.5716 | -0.00851 | 0.001397 | 1.15e-09 | 0.3717 | 15 | 887212 |
| 8 | 11226456 | C | A | 0.5726 | -0.007853 | 0.001293 | 1.29e-09 | 0.4095 | 18 | 911943 |
| 8 | 10147398 | C | T | 0.4776 | -0.007824 | 0.001293 | 1.47e-09 | 0.2429 | 18 | 911943 |
| 8 | 11226071 | A | G | 0.5664 | -0.007704 | 0.00129 | 2.43e-09 | 0.4171 | 18 | 911943 |
| 8 | 11232788 | G | A | 0.5716 | -0.007669 | 0.001292 | 2.99e-09 | 0.4309 | 18 | 911943 |
| 8 | 11231249 | T | C | 0.4387 | 0.007613 | 0.001287 | 3.38e-09 | 0.402 | 18 | 911943 |
| 8 | 11242632 | G | A | 0.5709 | -0.007632 | 0.001292 | 3.57e-09 | 0.441 | 18 | 911943 |
| 8 | 11233318 | A | C | 0.5659 | -0.007603 | 0.001289 | 3.74e-09 | 0.4 | 18 | 911943 |
| 8 | 11236681 | T | G | 0.5705 | -0.007595 | 0.001289 | 3.92e-09 | 0.3885 | 19 | 913247 |
| 8 | 11236685 | C | T | 0.5705 | -0.007594 | 0.001289 | 3.93e-09 | 0.3889 | 19 | 913247 |
| 8 | 11236419 | T | C | 0.5711 | -0.007605 | 0.001292 | 4.02e-09 | 0.4042 | 19 | 913246 |
| 8 | 11245303 | C | G | 0.4339 | 0.007598 | 0.001291 | 4.04e-09 | 0.3975 | 18 | 911943 |
| 8 | 11236413 | C | T | 0.5711 | -0.007601 | 0.001292 | 4.1e-09 | 0.4063 | 19 | 913246 |
| 8 | 11236572 | T | A | 0.5704 | -0.00758 | 0.001288 | 4.12e-09 | 0.3963 | 19 | 913247 |
| 8 | 11230574 | G | A | 0.5712 | -0.007598 | 0.001292 | 4.15e-09 | 0.415 | 18 | 911943 |
| 8 | 11237480 | A | G | 0.5708 | -0.007577 | 0.001289 | 4.26e-09 | 0.3781 | 19 | 913247 |
| 8 | 11237477 | A | G | 0.5708 | -0.007577 | 0.001289 | 4.26e-09 | 0.3769 | 19 | 913247 |
| 8 | 11233419 | T | G | 0.5659 | -0.007571 | 0.001289 | 4.35e-09 | 0.4006 | 18 | 911943 |
| 8 | 11231886 | A | G | 0.5657 | -0.007569 | 0.001289 | 4.41e-09 | 0.4239 | 18 | 911943 |
| 8 | 11236392 | T | C | 0.5711 | -0.007572 | 0.001292 | 4.7e-09 | 0.402 | 19 | 913246 |
| 8 | 11227406 | G | A | 0.5709 | -0.007569 | 0.001292 | 4.75e-09 | 0.4118 | 18 | 911943 |
| 8 | 11249010 | G | T | 0.4338 | 0.007569 | 0.001292 | 4.78e-09 | 0.4165 | 18 | 911943 |
| 8 | 11239762 | A | T | 0.5659 | -0.007547 | 0.001288 | 4.78e-09 | 0.3914 | 18 | 911943 |
| 8 | 11232860 | G | A | 0.5714 | -0.007565 | 0.001291 | 4.79e-09 | 0.4451 | 18 | 911943 |
| 8 | 11080675 | C | T | 0.5403 | -0.007576 | 0.001294 | 4.9e-09 | 0.4587 | 18 | 911943 |
| 8 | 11227885 | C | T | 0.5714 | -0.007563 | 0.001292 | 4.9e-09 | 0.4246 | 18 | 911943 |
| 8 | 11228100 | G | A | 0.5709 | -0.007556 | 0.001291 | 4.98e-09 | 0.4132 | 18 | 911943 |
| 8 | 11234520 | A | G | 0.5706 | -0.007548 | 0.00129 | 5.04e-09 | 0.426 | 18 | 911943 |
| 8 | 11234500 | A | T | 0.5656 | -0.007536 | 0.001288 | 5.04e-09 | 0.41 | 18 | 911943 |
| 8 | 11234885 | C | G | 0.5705 | -0.007541 | 0.00129 | 5.12e-09 | 0.4243 | 18 | 911943 |
| 8 | 11233582 | A | G | 0.5659 | -0.007535 | 0.001289 | 5.16e-09 | 0.3987 | 18 | 911943 |
| 8 | 11234298 | C | T | 0.5706 | -0.007542 | 0.00129 | 5.18e-09 | 0.4195 | 18 | 911943 |
| 8 | 11233659 | A | G | 0.5659 | -0.007534 | 0.001289 | 5.18e-09 | 0.3984 | 18 | 911943 |
| 8 | 11062882 | T | C | 0.5367 | -0.007525 | 0.001287 | 5.19e-09 | 0.4822 | 19 | 913242 |
| 8 | 11234844 | G | A | 0.5706 | -0.007537 | 0.00129 | 5.31e-09 | 0.4234 | 18 | 911943 |
| 8 | 11233958 | A | G | 0.5707 | -0.007539 | 0.001291 | 5.37e-09 | 0.4345 | 18 | 911943 |
| 8 | 11236975 | A | G | 0.5705 | -0.007528 | 0.001289 | 5.4e-09 | 0.4179 | 18 | 911969 |
| 8 | 11232343 | C | G | 0.5706 | -0.007538 | 0.001291 | 5.42e-09 | 0.4423 | 18 | 911943 |
| 8 | 11234780 | C | G | 0.5706 | -0.007531 | 0.00129 | 5.47e-09 | 0.4236 | 18 | 911943 |
| 8 | 11233917 | C | T | 0.5707 | -0.007534 | 0.001291 | 5.51e-09 | 0.435 | 18 | 911943 |
| 8 | 11230206 | G | T | 0.5712 | -0.007537 | 0.001292 | 5.51e-09 | 0.4123 | 18 | 911943 |
| 8 | 11234367 | T | G | 0.57 | -0.007527 | 0.00129 | 5.57e-09 | 0.4166 | 18 | 911943 |
| 8 | 11235360 | A | C | 0.5706 | -0.007524 | 0.00129 | 5.59e-09 | 0.4291 | 18 | 911943 |
| 8 | 11234626 | T | C | 0.5707 | -0.00753 | 0.001291 | 5.61e-09 | 0.4236 | 18 | 911943 |
| 8 | 11235497 | G | A | 0.5705 | -0.007514 | 0.001289 | 5.7e-09 | 0.3965 | 19 | 913238 |
| 8 | 11234613 | T | A | 0.5707 | -0.007523 | 0.001291 | 5.78e-09 | 0.4237 | 18 | 911943 |
| 8 | 10988275 | A | G | 0.5736 | -0.008199 | 0.001407 | 5.79e-09 | 0.5068 | 14 | 885924 |
| 8 | 11230259 | A | T | 0.5711 | -0.007527 | 0.001292 | 5.79e-09 | 0.4141 | 18 | 911943 |
| 8 | 11236850 | C | T | 0.5705 | -0.007503 | 0.001288 | 5.81e-09 | 0.3909 | 19 | 913247 |
| 8 | 11236964 | C | T | 0.5707 | -0.007504 | 0.001288 | 5.81e-09 | 0.3878 | 19 | 913243 |
| 8 | 11235735 | T | C | 0.5705 | -0.007512 | 0.00129 | 5.85e-09 | 0.4263 | 18 | 911943 |
| 8 | 11229319 | C | G | 0.5714 | -0.007531 | 0.001293 | 5.9e-09 | 0.4259 | 18 | 911943 |
| 8 | 11080665 | T | A | 0.5402 | -0.00753 | 0.001294 | 6.04e-09 | 0.4613 | 18 | 911943 |
| 8 | 11235136 | C | G | 0.5707 | -0.00751 | 0.00129 | 6.04e-09 | 0.4269 | 18 | 911943 |
| 8 | 11249261 | T | C | 0.3757 | 0.008415 | 0.001447 | 6.14e-09 | 0.2665 | 15 | 887212 |
| 8 | 11243126 | C | G | 0.5755 | -0.007524 | 0.001294 | 6.27e-09 | 0.4757 | 18 | 911943 |
| 8 | 11236809 | T | C | 0.5703 | -0.007484 | 0.001288 | 6.35e-09 | 0.3937 | 19 | 913246 |
| 8 | 11238029 | C | T | 0.5754 | -0.007501 | 0.001291 | 6.4e-09 | 0.4699 | 18 | 911943 |
| 8 | 11239640 | C | T | 0.5752 | -0.00751 | 0.001293 | 6.44e-09 | 0.4631 | 18 | 911943 |
| 8 | 11231354 | G | T | 0.4393 | 0.007478 | 0.001287 | 6.46e-09 | 0.4068 | 18 | 911943 |
| 8 | 11235605 | T | G | 0.5706 | -0.007492 | 0.00129 | 6.55e-09 | 0.3969 | 19 | 913240 |

| chromosome | position | other_allele | reference_allele | eaf | beta | se | p-value | i2 | n_studies | n_samples |
|---|---|---|---|---|---|---|---|---|---|---|
| 8 | 11235393 | A | G | 0.5705 | -0.007485 | 0.00129 | 6.65e-09 | 0.4288 | 18 | 911943 |
| 8 | 11235579 | T | G | 0.5701 | -0.007475 | 0.00129 | 7.06e-09 | 0.3914 | 19 | 913239 |
| 8 | 11046394 | C | G | 0.6006 | -0.008162 | 0.001412 | 7.69e-09 | 0.4063 | 15 | 887212 |
| 8 | 11239510 | G | A | 0.575 | -0.00746 | 0.001292 | 7.93e-09 | 0.4666 | 18 | 911943 |
| 8 | 11239565 | C | T | 0.575 | -0.007459 | 0.001292 | 7.99e-09 | 0.4658 | 18 | 911943 |
| 8 | 10903475 | T | A | 0.5141 | -0.007448 | 0.001291 | 8.09e-09 | 0.5474 | 17 | 910655 |
| 8 | 11239352 | G | A | 0.575 | -0.00745 | 0.001291 | 8.18e-09 | 0.4677 | 18 | 911943 |
| 8 | 11229638 | C | T | 0.5705 | -0.007456 | 0.001293 | 8.2e-09 | 0.4441 | 17 | 910655 |
| 8 | 8700851 | C | T | 0.5142 | 0.007429 | 0.001289 | 8.41e-09 | 0.51 | 18 | 911943 |
| 8 | 11239137 | G | A | 0.5749 | -0.007434 | 0.001291 | 8.81e-09 | 0.4672 | 18 | 911943 |
| 8 | 11238597 | C | T | 0.5751 | -0.007428 | 0.001291 | 8.92e-09 | 0.4628 | 18 | 911943 |
| 8 | 11061792 | C | T | 0.5377 | -0.007402 | 0.001287 | 8.98e-09 | 0.4862 | 19 | 913246 |
| 8 | 11060311 | A | C | 0.5375 | -0.007402 | 0.001287 | 9.05e-09 | 0.477 | 19 | 913243 |
| 8 | 11071057 | A | G | 0.5908 | -0.007484 | 0.001301 | 9.1e-09 | 0.5376 | 17 | 910655 |
| 8 | 11237773 | T | C | 0.5746 | -0.007413 | 0.00129 | 9.23e-09 | 0.4664 | 18 | 911943 |
| 8 | 11235910 | G | A | 0.5744 | -0.007423 | 0.001292 | 9.37e-09 | 0.4585 | 18 | 911943 |
| 8 | 8646246 | T | C | 0.5115 | 0.007416 | 0.001291 | 9.37e-09 | 0.4478 | 17 | 910655 |
| 8 | 11239078 | T | G | 0.5751 | -0.007419 | 0.001291 | 9.43e-09 | 0.4687 | 18 | 911943 |
| 8 | 11237756 | C | A | 0.5744 | -0.007376 | 0.001285 | 9.75e-09 | 0.4688 | 18 | 911943 |
| 8 | 11239017 | T | A | 0.5751 | -0.007409 | 0.001291 | 9.87e-09 | 0.4648 | 18 | 911943 |
| 8 | 11239054 | C | T | 0.5752 | -0.007403 | 0.001292 | 1.03e-08 | 0.4671 | 18 | 911943 |
| 8 | 11079367 | G | A | 0.5331 | -0.007379 | 0.00129 | 1.08e-08 | 0.4981 | 18 | 911943 |
| 8 | 11078949 | G | A | 0.5332 | -0.007319 | 0.00129 | 1.42e-08 | 0.5077 | 18 | 911943 |
| 8 | 11247814 | G | A | 0.3784 | 0.007548 | 0.001331 | 1.46e-08 | 0.3576 | 18 | 911943 |
| 8 | 8633548 | T | G | 0.4989 | -0.007318 | 0.001292 | 1.51e-08 | 0.4658 | 18 | 911943 |
| 8 | 8664622 | G | A | 0.5241 | 0.007302 | 0.001292 | 1.61e-08 | 0.3344 | 17 | 910655 |
| 8 | 11076635 | T | G | 0.5317 | -0.007286 | 0.001289 | 1.63e-08 | 0.5173 | 17 | 910655 |
| 8 | 8661534 | T | C | 0.5086 | 0.007262 | 0.001286 | 1.65e-08 | 0.4618 | 19 | 913244 |
| 8 | 8652889 | G | A | 0.5085 | 0.007269 | 0.001287 | 1.67e-08 | 0.4619 | 19 | 913236 |
| 8 | 11078781 | A | C | 0.5317 | -0.007282 | 0.00129 | 1.68e-08 | 0.5045 | 18 | 911943 |
| 8 | 8685854 | A | G | 0.5105 | 0.007265 | 0.001292 | 1.92e-08 | 0.4535 | 17 | 910655 |
| 8 | 8661681 | C | G | 0.5125 | 0.007243 | 0.001288 | 1.92e-08 | 0.4413 | 18 | 911943 |
| 8 | 8641145 | C | T | 0.5003 | 0.007248 | 0.001291 | 2.01e-08 | 0.439 | 17 | 910655 |
| 8 | 11219781 | C | T | 0.4314 | 0.007242 | 0.00129 | 2.03e-08 | 0.4003 | 18 | 911943 |
| 8 | 8637429 | G | A | 0.4998 | -0.007226 | 0.00129 | 2.15e-08 | 0.4501 | 18 | 911943 |
| 8 | 8721473 | G | A | 0.5303 | 0.007253 | 0.001295 | 2.18e-08 | 0.5224 | 18 | 911943 |
| 8 | 10987553 | T | C | 0.5731 | -0.007267 | 0.001298 | 2.2e-08 | 0.4775 | 17 | 910655 |
| 8 | 8770512 | C | T | 0.5757 | 0.007282 | 0.001302 | 2.27e-08 | 0.3497 | 18 | 911943 |
| 8 | 11242025 | T | G | 0.5379 | -0.007269 | 0.001299 | 2.28e-08 | 0.4195 | 17 | 438262 |
| 8 | 8678530 | G | A | 0.5217 | 0.007207 | 0.00129 | 2.37e-08 | 0.3986 | 19 | 913239 |
| 8 | 11069960 | C | T | 0.5505 | -0.007195 | 0.001288 | 2.4e-08 | 0.5208 | 18 | 911969 |
| 8 | 11251175 | A | G | 0.4415 | 0.007353 | 0.001317 | 2.43e-08 | 0.4007 | 16 | 715261 |
| 8 | 11215617 | C | G | 0.4304 | 0.007177 | 0.001288 | 2.55e-08 | 0.3867 | 18 | 911943 |
| 8 | 10938260 | G | A | 0.5198 | -0.007176 | 0.001289 | 2.63e-08 | 0.5288 | 17 | 910655 |
| 8 | 10987651 | T | G | 0.573 | -0.007225 | 0.001298 | 2.65e-08 | 0.4855 | 17 | 910655 |
| 8 | 11055597 | C | A | 0.5451 | -0.007165 | 0.001287 | 2.67e-08 | 0.4161 | 18 | 911943 |
| 8 | 8685646 | T | C | 0.5102 | 0.007186 | 0.001291 | 2.69e-08 | 0.4889 | 18 | 911943 |
| 8 | 8673601 | A | C | 0.5173 | 0.007168 | 0.001289 | 2.76e-08 | 0.4025 | 18 | 911969 |
| 8 | 11214972 | A | G | 0.4697 | 0.007146 | 0.001286 | 2.8e-08 | 0.3389 | 18 | 911943 |
| 8 | 8639740 | A | G | 0.4998 | -0.007166 | 0.00129 | 2.85e-08 | 0.4663 | 18 | 911943 |
| 8 | 8704330 | G | C | 0.519 | 0.007156 | 0.001289 | 2.87e-08 | 0.5078 | 18 | 911943 |
| 8 | 11219334 | G | A | 0.4312 | 0.007168 | 0.001291 | 2.87e-08 | 0.4405 | 17 | 910655 |
| 8 | 11214455 | A | G | 0.47 | 0.00714 | 0.001286 | 2.89e-08 | 0.3137 | 18 | 911943 |
| 8 | 8682192 | A | T | 0.5216 | 0.007164 | 0.001291 | 2.91e-08 | 0.3828 | 19 | 913236 |
| 8 | 8679614 | T | C | 0.5215 | 0.00716 | 0.00129 | 2.92e-08 | 0.3824 | 19 | 913237 |
| 8 | 8675325 | A | T | 0.5129 | 0.007152 | 0.00129 | 3e-08 | 0.4612 | 18 | 911943 |
| 8 | 10987199 | T | C | 0.573 | -0.007183 | 0.001296 | 3.08e-08 | 0.4509 | 18 | 911943 |
| 8 | 8684953 | G | A | 0.5211 | 0.007165 | 0.001293 | 3.09e-08 | 0.3888 | 18 | 911943 |
| 8 | 8660538 | C | A | 0.518 | 0.007136 | 0.001289 | 3.14e-08 | 0.4459 | 18 | 911943 |
| 8 | 8673736 | T | C | 0.5182 | 0.007136 | 0.001289 | 3.14e-08 | 0.4137 | 19 | 913246 |
| 8 | 11225168 | G | A | 0.5225 | -0.007119 | 0.001286 | 3.18e-08 | 0.3131 | 18 | 911943 |
| 8 | 8730488 | G | A | 0.5269 | 0.007178 | 0.001297 | 3.24e-08 | 0.5213 | 18 | 911943 |
| 8 | 11225910 | A | G | 0.5217 | -0.007125 | 0.001288 | 3.27e-08 | 0.3537 | 17 | 910655 |
| 8 | 8679176 | A | G | 0.5216 | 0.007118 | 0.00129 | 3.52e-08 | 0.3941 | 19 | 913240 |
| 8 | 8680477 | G | A | 0.5215 | 0.007109 | 0.00129 | 3.66e-08 | 0.3769 | 19 | 913236 |
| 8 | 11225480 | T | C | 0.5212 | -0.007088 | 0.001286 | 3.66e-08 | 0.3118 | 18 | 911943 |
| 8 | 8682878 | T | C | 0.5111 | 0.007108 | 0.00129 | 3.68e-08 | 0.5097 | 18 | 911943 |

| chromosome | position | other_allele | reference_allele | eaf | beta | se | p-value | i2 | n_studies | n_samples |
|---|---|---|---|---|---|---|---|---|---|---|
| 8 | 11213589 | T | G | 0.4306 | 0.007102 | 0.00129 | 3.74e-08 | 0.4307 | 17 | 910655 |
| 8 | 9569104 | T | C | 0.8201 | 0.009116 | 0.001657 | 3.84e-08 | 0 | 19 | 913239 |
| 8 | 11070360 | C | G | 0.5463 | -0.007068 | 0.001286 | 3.98e-08 | 0.5116 | 19 | 913241 |
| 8 | 11045161 | G | A | 0.5313 | -0.007055 | 0.001285 | 4.08e-08 | 0.4718 | 19 | 913257 |
| 8 | 11228672 | G | A | 0.478 | 0.00705 | 0.001285 | 4.16e-08 | 0.3448 | 18 | 911943 |
| 8 | 11054097 | C | A | 0.549 | -0.007068 | 0.001288 | 4.17e-08 | 0.416 | 19 | 913245 |
| 8 | 11046209 | T | C | 0.5994 | -0.007142 | 0.001303 | 4.37e-08 | 0.4603 | 18 | 911943 |
| 8 | 10986837 | T | G | 0.5729 | -0.007109 | 0.001298 | 4.41e-08 | 0.4971 | 17 | 910655 |
| 8 | 11041642 | A | T | 0.53 | -0.007042 | 0.001286 | 4.46e-08 | 0.4661 | 18 | 911943 |
| 8 | 11252425 | C | A | 0.506 | 0.00745 | 0.001361 | 4.46e-08 | 0.39 | 18 | 911943 |
| 8 | 8644274 | G | C | 0.5126 | 0.007074 | 0.001292 | 4.53e-08 | 0.4487 | 18 | 911943 |
| 8 | 11425809 | T | G | 0.5366 | -0.007038 | 0.001288 | 4.79e-08 | 0.5089 | 19 | 913234 |
| 8 | 8685190 | A | G | 0.5208 | 0.00706 | 0.001292 | 4.81e-08 | 0.386 | 18 | 911943 |
| 8 | 8654527 | T | C | 0.5158 | 0.007035 | 0.001289 | 4.91e-08 | 0.4002 | 18 | 911943 |
| 8 | 11039816 | T | G | 0.5275 | -0.007028 | 0.001288 | 4.94e-08 | 0.4789 | 17 | 910655 |
| 12 | 62989110 | A | C | 0.4966 | -0.00726 | 0.001281 | 1.5e-08 | 0.5437 | 19 | 913254 |
| 12 | 62995984 | C | T | 0.6721 | -0.008214 | 0.00145 | 1.52e-08 | 0.4616 | 14 | 885924 |
| 12 | 62986620 | G | A | 0.6689 | -0.008176 | 0.001445 | 1.57e-08 | 0.4594 | 15 | 887238 |
| 12 | 62990403 | A | G | 0.4964 | -0.007246 | 0.001281 | 1.6e-08 | 0.5448 | 19 | 913254 |
| 12 | 62990415 | T | C | 0.4964 | -0.007244 | 0.001281 | 1.61e-08 | 0.545 | 19 | 913254 |
| 12 | 62995340 | G | A | 0.6723 | -0.007497 | 0.001339 | 2.23e-08 | 0.3941 | 19 | 913254 |
| 12 | 62945970 | C | G | 0.6723 | -0.007478 | 0.001339 | 2.38e-08 | 0.4074 | 19 | 913256 |
| 12 | 62977163 | G | A | 0.6723 | -0.007489 | 0.001341 | 2.41e-08 | 0.3837 | 19 | 913247 |
| 12 | 62995269 | A | G | 0.6722 | -0.007479 | 0.001339 | 2.41e-08 | 0.3821 | 19 | 913255 |
| 12 | 62968280 | T | C | 0.6722 | -0.007464 | 0.001338 | 2.5e-08 | 0.3935 | 19 | 913255 |
| 12 | 62948736 | G | A | 0.6724 | -0.007468 | 0.00134 | 2.58e-08 | 0.4162 | 19 | 913256 |
| 12 | 62985871 | C | A | 0.6722 | -0.007461 | 0.001339 | 2.59e-08 | 0.3979 | 19 | 913254 |
| 12 | 62950556 | G | C | 0.6723 | -0.007452 | 0.001339 | 2.66e-08 | 0.4071 | 19 | 913256 |
| 12 | 62942258 | C | T | 0.6725 | -0.007438 | 0.001339 | 2.83e-08 | 0.4196 | 19 | 913256 |
| 12 | 62960880 | A | G | 0.669 | -0.007405 | 0.001335 | 2.98e-08 | 0.4136 | 19 | 913255 |
| 12 | 62961299 | C | T | 0.6724 | -0.007425 | 0.00134 | 3.05e-08 | 0.4013 | 19 | 913256 |
| 12 | 62957906 | G | C | 0.6689 | -0.007389 | 0.001336 | 3.25e-08 | 0.4206 | 19 | 913255 |
| 12 | 62996061 | G | A | 0.6723 | -0.007427 | 0.001343 | 3.29e-08 | 0.4224 | 18 | 911966 |
| 12 | 62993793 | G | A | 0.6692 | -0.007428 | 0.001344 | 3.31e-08 | 0.4273 | 18 | 717861 |
| 12 | 62952696 | C | T | 0.6722 | -0.007403 | 0.001339 | 3.33e-08 | 0.4048 | 19 | 913257 |
| 12 | 62949110 | A | C | 0.669 | -0.007376 | 0.001335 | 3.38e-08 | 0.4214 | 19 | 913256 |
| 12 | 62961801 | G | A | 0.669 | -0.007377 | 0.001336 | 3.42e-08 | 0.4179 | 19 | 913256 |
| 12 | 62975307 | C | T | 0.6722 | -0.007408 | 0.001342 | 3.48e-08 | 0.4223 | 18 | 911967 |
| 12 | 62937532 | C | T | 0.6662 | -0.007354 | 0.001333 | 3.52e-08 | 0.5161 | 18 | 911943 |
| 12 | 62990871 | C | A | 0.6722 | -0.007389 | 0.00134 | 3.56e-08 | 0.3996 | 19 | 913255 |
| 12 | 62964552 | T | C | 0.6611 | -0.007429 | 0.001347 | 3.57e-08 | 0.442 | 17 | 438286 |
| 12 | 62970482 | A | G | 0.669 | -0.007376 | 0.001338 | 3.63e-08 | 0.442 | 18 | 911967 |
| 12 | 62945245 | G | A | 0.6723 | -0.007386 | 0.001341 | 3.73e-08 | 0.4283 | 18 | 911969 |
| 12 | 62952910 | C | T | 0.6723 | -0.007379 | 0.001342 | 3.9e-08 | 0.4357 | 18 | 911968 |
| 12 | 62980580 | T | A | 0.6689 | -0.007328 | 0.001336 | 4.22e-08 | 0.4088 | 19 | 913255 |
| 12 | 62988288 | G | A | 0.6723 | -0.007354 | 0.001342 | 4.38e-08 | 0.431 | 18 | 911967 |
| 12 | 62870574 | C | G | 0.6679 | -0.007274 | 0.001331 | 4.74e-08 | 0.4334 | 19 | 913254 |
| 12 | 62941837 | A | G | 0.6714 | -0.007297 | 0.001337 | 4.93e-08 | 0.4417 | 19 | 913249 |
| 12 | 62941426 | T | C | 0.6712 | -0.007294 | 0.001337 | 4.97e-08 | 0.4393 | 19 | 913250 |
| 13 | 83312341 | C | T | 0.5983 | -0.007206 | 0.001303 | 3.23e-08 | 0 | 18 | 911943 |
| 13 | 83311141 | G | A | 0.5985 | -0.007179 | 0.001301 | 3.52e-08 | 0 | 18 | 911943 |
| 13 | 83312717 | G | C | 0.5979 | -0.007177 | 0.001303 | 3.74e-08 | 0 | 18 | 911943 |
| 13 | 83304898 | T | G | 0.5985 | -0.007095 | 0.001299 | 4.87e-08 | 0 | 18 | 911943 |
| 16 | 13040514 | C | T | 0.6805 | -0.008169 | 0.001459 | 2.19e-08 | 0 | 17 | 910655 |
| 16 | 13041027 | G | T | 0.6806 | -0.008021 | 0.001456 | 3.66e-08 | 0 | 18 | 911943 |
| 16 | 13042097 | G | A | 0.6807 | -0.00802 | 0.001456 | 3.66e-08 | 0 | 18 | 911943 |
| 16 | 13039154 | C | T | 0.6795 | -0.008713 | 0.001583 | 3.79e-08 | 0 | 15 | 887212 |
| 16 | 13040889 | C | G | 0.6806 | -0.008004 | 0.001455 | 3.86e-08 | 0 | 18 | 911943 |
| 16 | 13043452 | A | G | 0.6805 | -0.007971 | 0.001455 | 4.41e-08 | 0 | 18 | 911943 |
| 16 | 13041924 | A | G | 0.6806 | -0.007968 | 0.001455 | 4.44e-08 | 0 | 18 | 911943 |

Abbreviations: eaf = effect allele frequency; se = standard error

#### Supplementary Table S4

lead SNPs based on COJO analysis ordered by chromosome

| Chr | SNP | bp | refA | freq | b | se | p | n | freq_geno | bJ | bJ_se | pJ | LD_r |
|---|---|---|---|---|---|---|---|---|---|---|---|---|---|
| 2 | rs35098301 | 98580012 | T | 0.4898 | 0.008074 | 0.001427 | 1.53e-08 | 400971 | 0.6061 | 0.008074 | 0.001427 | 1.53e-08 | 0 |
| 8 | rs35009431 | 11232788 | G | 0.5716 | -0.007669 | 0.001292 | 2.92e-09 | 499184 | 0.5518 | -0.007669 | 0.001292 | 2.93e-09 | 0 |
| 12 | rs10877884 | 62989110 | A | 0.4966 | -0.00726 | 0.001281 | 1.45e-08 | 497403 | 0.5274 | -0.00726 | 0.001281 | 1.45e-08 | 0 |
| 13 | rs2147632 | 83311141 | G | 0.5985 | -0.007179 | 0.001301 | 3.43e-08 | 501657 | 0.5788 | -0.007179 | 0.001301 | 3.43e-08 | 0 |
| 16 | rs12922637 | 13040514 | C | 0.6805 | -0.008169 | 0.001459 | 2.16e-08 | 440877 | 0.7445 | -0.008169 | 0.001459 | 2.16e-08 | 0 |

Abbreviations: Chr = chromosome; SNP = single nucleotide polymorphism; bp = base position; refA = reference allele; freq = frequency effect allele; b = beta; se = standard error; p = p-value; n = number; freq_geno = frequency reference dataset; bJ = effect size joint analysis; bJ_se = standard error joint analysis; pJ = p-value joint analysis; LD_r = LD correlation between SNPs

#### Supplementary Table S5

Leave-one-out meta-analysis (LL_cytosnp) ordered from most significant to less significant per chromosome

| chromosome | position | beta | beta_LL_cytoSNP_out | p-value | p-value_LL_cytoSNP_out | i2 | i2_LL_cytoSNP_out |
|---|---|---|---|---|---|---|---|
| 2 | 98580012 | 0.008074 | 0.008075 | 1.57e-08 | 1.6e-08 | 0.3447 | 0.3884 |
| 2 | 98427382 | 0.008 | 0.007995 | 2.07e-08 | 2.16e-08 | 0.413 | 0.4547 |
| 8 | 11417582 | 0.01009 | 0.01009 | 9.09e-11 | 9.09e-11 | 0.4954 | 0.4954 |
| 8 | 11235150 | -0.00851 | -0.008543 | 1.15e-09 | 9.99e-10 | 0.3717 | 0.3878 |
| 8 | 11226456 | -0.007853 | -0.007882 | 1.29e-09 | 1.13e-09 | 0.4095 | 0.4226 |
| 8 | 10147398 | -0.007824 | -0.007852 | 1.47e-09 | 1.3e-09 | 0.2429 | 0.2691 |
| 8 | 11226071 | -0.007704 | -0.007733 | 2.43e-09 | 2.13e-09 | 0.4171 | 0.433 |
| 8 | 11232788 | -0.007669 | -0.007699 | 2.99e-09 | 2.62e-09 | 0.4309 | 0.4431 |
| 8 | 11231249 | 0.007613 | 0.007639 | 3.38e-09 | 3.02e-09 | 0.402 | 0.4222 |
| 8 | 11242632 | -0.007632 | -0.007653 | 3.57e-09 | 3.26e-09 | 0.441 | 0.4637 |
| 8 | 11233318 | -0.007603 | -0.007638 | 3.74e-09 | 3.2e-09 | 0.4 | 0.4032 |
| 8 | 11236681 | -0.007595 | -0.007623 | 3.92e-09 | 3.46e-09 | 0.3885 | 0.4018 |
| 8 | 11236685 | -0.007594 | -0.007622 | 3.93e-09 | 3.47e-09 | 0.3889 | 0.4022 |
| 8 | 11236419 | -0.007605 | -0.007633 | 4.02e-09 | 3.55e-09 | 0.4042 | 0.4177 |
| 8 | 11245303 | 0.007598 | 0.007624 | 4.04e-09 | 3.61e-09 | 0.3975 | 0.4152 |
| 8 | 11236413 | -0.007601 | -0.007629 | 4.1e-09 | 3.63e-09 | 0.4063 | 0.4197 |
| 8 | 11236572 | -0.00758 | -0.00761 | 4.12e-09 | 3.61e-09 | 0.3963 | 0.4069 |
| 8 | 11230574 | -0.007598 | -0.007628 | 4.15e-09 | 3.64e-09 | 0.415 | 0.4266 |
| 8 | 11237477 | -0.007577 | -0.007605 | 4.26e-09 | 3.76e-09 | 0.3769 | 0.3896 |
| 8 | 11237480 | -0.007577 | -0.007606 | 4.26e-09 | 3.75e-09 | 0.3781 | 0.3898 |
| 8 | 11233419 | -0.007571 | -0.007606 | 4.35e-09 | 3.72e-09 | 0.4006 | 0.4027 |
| 8 | 11231886 | -0.007569 | -0.007594 | 4.41e-09 | 3.96e-09 | 0.4239 | 0.4441 |
| 8 | 11236392 | -0.007572 | -0.007601 | 4.7e-09 | 4.13e-09 | 0.402 | 0.4136 |
| 8 | 11227406 | -0.007569 | -0.007591 | 4.75e-09 | 4.33e-09 | 0.4118 | 0.4346 |
| 8 | 11239762 | -0.007547 | -0.007574 | 4.78e-09 | 4.25e-09 | 0.3914 | 0.4076 |
| 8 | 11249010 | 0.007569 | 0.007602 | 4.78e-09 | 4.13e-09 | 0.4165 | 0.422 |
| 8 | 11232860 | -0.007565 | -0.00759 | 4.79e-09 | 4.3e-09 | 0.4451 | 0.4642 |
| 8 | 11080675 | -0.007576 | -0.007582 | 4.9e-09 | 4.8e-09 | 0.4587 | 0.4897 |
| 8 | 11227885 | -0.007563 | -0.007593 | 4.9e-09 | 4.3e-09 | 0.4246 | 0.4368 |
| 8 | 11228100 | -0.007556 | -0.007579 | 4.98e-09 | 4.51e-09 | 0.4132 | 0.4342 |
| 8 | 11234500 | -0.007536 | -0.007564 | 5.04e-09 | 4.47e-09 | 0.41 | 0.425 |
| 8 | 11234520 | -0.007548 | -0.007578 | 5.04e-09 | 4.43e-09 | 0.426 | 0.4386 |
| 8 | 11234885 | -0.007541 | -0.007571 | 5.12e-09 | 4.49e-09 | 0.4243 | 0.4359 |
| 8 | 11233582 | -0.007535 | -0.007569 | 5.16e-09 | 4.44e-09 | 0.3987 | 0.4029 |
| 8 | 11233659 | -0.007534 | -0.007568 | 5.18e-09 | 4.45e-09 | 0.3984 | 0.4025 |
| 8 | 11234298 | -0.007542 | -0.007572 | 5.18e-09 | 4.54e-09 | 0.4195 | 0.4312 |
| 8 | 11062882 | -0.007525 | -0.007545 | 5.19e-09 | 4.75e-09 | 0.4822 | 0.5015 |
| 8 | 11234844 | -0.007537 | -0.007568 | 5.31e-09 | 4.65e-09 | 0.4234 | 0.4344 |
| 8 | 11233958 | -0.007539 | -0.007569 | 5.37e-09 | 4.7e-09 | 0.4345 | 0.4456 |
| 8 | 11236975 | -0.007528 | -0.007556 | 5.4e-09 | 4.76e-09 | 0.4179 | 0.4313 |
| 8 | 11232343 | -0.007538 | -0.007561 | 5.42e-09 | 4.91e-09 | 0.4423 | 0.4628 |
| 8 | 11234780 | -0.007531 | -0.007561 | 5.47e-09 | 4.8e-09 | 0.4236 | 0.4352 |
| 8 | 11230206 | -0.007537 | -0.007565 | 5.51e-09 | 4.86e-09 | 0.4123 | 0.4261 |
| 8 | 11233917 | -0.007534 | -0.007564 | 5.51e-09 | 4.81e-09 | 0.435 | 0.4459 |
| 8 | 11234367 | -0.007527 | -0.007557 | 5.57e-09 | 4.88e-09 | 0.4166 | 0.4281 |
| 8 | 11235360 | -0.007524 | -0.007553 | 5.59e-09 | 4.92e-09 | 0.4291 | 0.4422 |
| 8 | 11234626 | -0.00753 | -0.007558 | 5.61e-09 | 4.96e-09 | 0.4236 | 0.4381 |
| 8 | 11235497 | -0.007514 | -0.007544 | 5.7e-09 | 5e-09 | 0.3965 | 0.4069 |
| 8 | 11234613 | -0.007523 | -0.007551 | 5.78e-09 | 5.12e-09 | 0.4237 | 0.4384 |
| 8 | 10988275 | -0.008199 | -0.008181 | 5.79e-09 | 6.32e-09 | 0.5068 | 0.5406 |
| 8 | 11230259 | -0.007527 | -0.007555 | 5.79e-09 | 5.12e-09 | 0.4141 | 0.4283 |
| 8 | 11236850 | -0.007503 | -0.007532 | 5.81e-09 | 5.12e-09 | 0.3909 | 0.4021 |
| 8 | 11236964 | -0.007504 | -0.007532 | 5.81e-09 | 5.12e-09 | 0.3878 | 0.3997 |
| 8 | 11235735 | -0.007512 | -0.007541 | 5.85e-09 | 5.15e-09 | 0.4263 | 0.4392 |
| 8 | 11229319 | -0.007531 | -0.007555 | 5.9e-09 | 5.34e-09 | 0.4259 | 0.4466 |
| 8 | 11080665 | -0.00753 | -0.007536 | 6.04e-09 | 5.93e-09 | 0.4613 | 0.4922 |
| 8 | 11235136 | -0.00751 | -0.007539 | 6.04e-09 | 5,33E+07 | 0.4269 | 0.4406 |
| 8 | 11249261 | 0.008415 | 0.008472 | 6.14e-09 | 4.89e-09 | 0.2665 | 0.2209 |
| 8 | 11243126 | -0.007524 | -0.007555 | 6.27e-09 | 5.48e-09 | 0.4757 | 0.4876 |
| 8 | 11236809 | -0.007484 | -0.007513 | 6.35e-09 | 5.59e-09 | 0.3937 | 0.405 |

| chromosome | position | beta | beta_LL_cytoSNP_out | p-value | p-value_LL_cytoSNP_out | i2 | i2_LL_cytoSNP_out |
|---|---|---|---|---|---|---|---|
| 8 | 11238029 | -0.007501 | -0.007536 | 6.4e-09 | 5.49e-09 | 0.4699 | 0.4755 |
| 8 | 11239640 | -0.00751 | -0.007538 | 6.44e-09 | 5.7e-09 | 0.4631 | 0.4781 |
| 8 | 11231354 | 0.007478 | 0.007504 | 6.46e-09 | 5.79e-09 | 0.4068 | 0.4267 |
| 8 | 11235605 | -0.007492 | -0.007522 | 6.55e-09 | 5.74e-09 | 0.3969 | 0.407 |
| 8 | 11235393 | -0.007485 | -0.007514 | 6.65e-09 | 5.85e-09 | 0.4288 | 0.4415 |
| 8 | 11235579 | -0.007475 | -0.007507 | 7.06e-09 | 6.14e-09 | 0.3914 | 0.3983 |
| 8 | 11046394 | -0.008162 | -0.00818 | 7.69e-09 | 7.2e-09 | 0.4063 | 0.4427 |
| 8 | 11239510 | -0.00746 | -0.007489 | 7.93e-09 | 7.02e-09 | 0.4666 | 0.4813 |
| 8 | 11239565 | -0.007459 | -0.007487 | 7.99e-09 | 7.07e-09 | 0.4658 | 0.4806 |
| 8 | 10903475 | -0.007448 | -0.00743 | 8.09e-09 | 8.82e-09 | 0.5474 | 0.57 |
| 8 | 11239352 | -0.00745 | -0.007478 | 8.18e-09 | 7.23e-09 | 0.4677 | 0.4822 |
| 8 | 11229638 | -0.007456 | -0.007478 | 8.2e-09 | 7.48e-09 | 0.4441 | 0.4673 |
| 8 | 8700851 | 0.007429 | 0.007432 | 8.41e-09 | 8.31e-09 | 0.51 | 0.5385 |
| 8 | 11239137 | -0.007434 | -0.007463 | 8.81e-09 | 7.78e-09 | 0.4672 | 0.4811 |
| 8 | 11238597 | -0.007428 | -0.007457 | 8.92e-09 | 7.85e-09 | 0.4628 | 0.476 |
| 8 | 11061792 | -0.007402 | -0.007424 | 8.98e-09 | 8.2e-09 | 0.4862 | 0.5047 |
| 8 | 11060311 | -0.007402 | -0.007423 | 9.05e-09 | 8.24e-09 | 0.477 | 0.4949 |
| 8 | 11071057 | -0.007484 | -0.007493 | 9.1e-09 | 8.84e-09 | 0.5376 | 0.5655 |
| 8 | 11237773 | -0.007413 | -0.007441 | 9.23e-09 | 8.17e-09 | 0.4664 | 0.4812 |
| 8 | 11235910 | -0.007423 | -0.007457 | 9.37e-09 | 8.1e-09 | 0.4585 | 0.4662 |
| 8 | 8646246 | 0.007416 | 0.007409 | 9.37e-09 | 9.68e-09 | 0.4478 | 0.4809 |
| 8 | 11239078 | -0.007419 | -0.007448 | 9.43e-09 | 8.33e-09 | 0.4687 | 0.4826 |
| 8 | 11237756 | -0.007376 | -0.007404 | 9.75e-09 | 8.63e-09 | 0.4688 | 0.483 |
| 8 | 11239017 | -0.007409 | -0.007438 | 9.87e-09 | 8.7e-09 | 0.4648 | 0.4782 |
| 8 | 11239054 | -0.007403 | -0.007432 | 1.03e-08 | 9.08e-09 | 0.4671 | 0.4808 |
| 8 | 11079367 | -0.007379 | -0.007394 | 1.08e-08 | 1.01e-08 | 0.4981 | 0.5224 |
| 8 | 11078949 | -0.007319 | -0.007331 | 1.42e-08 | 1.36e-08 | 0.5077 | 0.5338 |
| 8 | 11247814 | 0.007548 | 0.007582 | 1.46e-08 | 1.27e-08 | 0.3576 | 0.367 |
| 8 | 8633548 | -0.007318 | -0.007309 | 1.51e-08 | 1.58e-08 | 0.4658 | 0.4949 |
| 8 | 8664622 | 0.007302 | 0.007302 | 1.61e-08 | 1.62e-08 | 0.3344 | 0.376 |
| 8 | 11076635 | -0.007286 | -0.007298 | 1.63e-08 | 1.55e-08 | 0.5173 | 0.544 |
| 8 | 8661534 | 0.007262 | 0.007256 | 1.65e-08 | 1.71e-08 | 0.4618 | 0.4904 |
| 8 | 8652889 | 0.007269 | 0.007263 | 1.67e-08 | 1.72e-08 | 0.4619 | 0.4908 |
| 8 | 11078781 | -0.007282 | -0.007295 | 1.68e-08 | 1.58e-08 | 0.5045 | 0.5294 |
| 8 | 8661681 | 0.007243 | 0.007233 | 1.92e-08 | 2.02e-08 | 0.4413 | 0.471 |
| 8 | 8685854 | 0.007265 | 0.007263 | 1.92e-08 | 1.94e-08 | 0.4535 | 0.4876 |
| 8 | 8641145 | 0.007248 | 0.007235 | 2.01e-08 | 2.15e-08 | 0.439 | 0.4675 |
| 8 | 11219781 | 0.007242 | 0.007252 | 2.03e-08 | 1.95e-08 | 0.4003 | 0.4331 |
| 8 | 8637429 | -0.007226 | -0.007214 | 2.15e-08 | 2.28e-08 | 0.4501 | 0.4775 |
| 8 | 8721473 | 0.007253 | 0.007263 | 2.18e-08 | 2.1e-08 | 0.5224 | 0.5483 |
| 8 | 10987553 | -0.007267 | -0.007253 | 2.2e-08 | 2.37e-08 | 0.4775 | 0.5068 |
| 8 | 8770512 | 0.007282 | 0.007223 | 2.27e-08 | 2.98e-08 | 0.3497 | 0.3055 |
| 8 | 11242025 | -0.007269 | -0.007301 | 2.28e-08 | 1.99e-08 | 0.4195 | 0.4284 |
| 8 | 8678530 | 0.007207 | 0.007208 | 2.37e-08 | 2.37e-08 | 0.3986 | 0.4319 |
| 8 | 11069960 | -0.007195 | -0.007216 | 2.4e-08 | 2.2e-08 | 0.5208 | 0.5407 |
| 8 | 11251175 | 0.007353 | 0.007399 | 2.43e-08 | 2e-08 | 0.4007 | 0.3848 |
| 8 | 11215617 | 0.007177 | 0.007185 | 2.55e-08 | 2.48e-08 | 0.3867 | 0.4212 |
| 8 | 10938260 | -0.007176 | -0.007188 | 2.63e-08 | 2.51e-08 | 0.5288 | 0.5556 |
| 8 | 10987651 | -0.007225 | -0.007209 | 2.65e-08 | 2.86e-08 | 0.4855 | 0.5138 |
| 8 | 11055597 | -0.007165 | -0.007185 | 2.67e-08 | 2.46e-08 | 0.4161 | 0.4396 |
| 8 | 8685646 | 0.007186 | 0.00718 | 2.69e-08 | 2.78e-08 | 0.4889 | 0.5179 |
| 8 | 8673601 | 0.007168 | 0.007169 | 2.76e-08 | 2.76e-08 | 0.4025 | 0.4376 |
| 8 | 11214972 | 0.007146 | 0.007149 | 2.8e-08 | 2.8e-08 | 0.3389 | 0.3777 |
| 8 | 8639740 | -0.007166 | -0.007154 | 2.85e-08 | 3.02e-08 | 0.4663 | 0.4929 |
| 8 | 11219334 | 0.007168 | 0.007179 | 2.87e-08 | 2.75e-08 | 0.4405 | 0.4727 |
| 8 | 8704330 | 0.007156 | 0.007152 | 2.87e-08 | 2.95e-08 | 0.5078 | 0.5362 |
| 8 | 11214455 | 0.00714 | 0.007145 | 2.89e-08 | 2.84e-08 | 0.3137 | 0.3532 |
| 8 | 8682192 | 0.007164 | 0.007161 | 2.91e-08 | 2.97e-08 | 0.3828 | 0.4167 |
| 8 | 8679614 | 0.00716 | 0.007161 | 2.92e-08 | 2.93e-08 | 0.3824 | 0.4167 |
| 8 | 8675325 | 0.007152 | 0.007149 | 3e-08 | 3.06e-08 | 0.4612 | 0.4926 |
| 8 | 10987199 | -0.007183 | -0.007167 | 3.08e-08 | 3.35e-08 | 0.4509 | 0.4783 |
| 8 | 8684953 | 0.007165 | 0.007168 | 3.09e-08 | 3.07e-08 | 0.3888 | 0.4245 |
| 8 | 8660538 | 0.007136 | 0.007129 | 3.14e-08 | 3.25e-08 | 0.4459 | 0.4771 |

| chromosome | position | beta | beta_LL_cytoSNP_out | p-value | p-value_LL_cytoSNP_out | i2 | i2_LL_cytoSNP_out |
|---|---|---|---|---|---|---|---|
| 8 | 8673736 | 0.007136 | 0.007137 | 3.14e-08 | 3.14e-08 | 0.4137 | 0.4462 |
| 8 | 11225168 | -0.007119 | -0.007145 | 3.18e-08 | 2.87e-08 | 0.3131 | 0.338 |
| 8 | 8730488 | 0.007178 | 0.007185 | 3.24e-08 | 3.16e-08 | 0.5213 | 0.5483 |
| 8 | 11225910 | -0.007125 | -0.007151 | 3.27e-08 | 2.94e-08 | 0.3537 | 0.379 |
| 8 | 8679176 | 0.007118 | 0.007119 | 3.52e-08 | 3.52e-08 | 0.3941 | 0.4278 |
| 8 | 11225480 | -0.007088 | -0.007115 | 3.66e-08 | 3.28e-08 | 0.3118 | 0.3353 |
| 8 | 8680477 | 0.007109 | 0.007109 | 3.66e-08 | 3.68e-08 | 0.3769 | 0.4115 |
| 8 | 8682878 | 0.007108 | 0.007103 | 3.68e-08 | 3.79e-08 | 0.5097 | 0.5378 |
| 8 | 11213589 | 0.007102 | 0.007111 | 3.74e-08 | 3.62e-08 | 0.4307 | 0.4645 |
| 8 | 9569104 | 0.009116 | 0.009108 | 3.84e-08 | 4e-08 | 0 | 0 |
| 8 | 11070360 | -0.007068 | -0.007087 | 3.98e-08 | 3.69e-08 | 0.5116 | 0.5321 |
| 8 | 11045161 | -0.007055 | -0.00708 | 4.08e-08 | 3.67e-08 | 0.4718 | 0.4849 |
| 8 | 11228672 | 0.00705 | 0.007086 | 4.16e-08 | 3.59e-08 | 0.3448 | 0.3542 |
| 8 | 11054097 | -0.007068 | -0.007088 | 4.17e-08 | 3.83e-08 | 0.416 | 0.4369 |
| 8 | 11046209 | -0.007142 | -0.007158 | 4.37e-08 | 4.1e-08 | 0.4603 | 0.4875 |
| 8 | 10986837 | -0.007109 | -0.00709 | 4.41e-08 | 4.82e-08 | 0.4971 | 0.5232 |
| 8 | 11041642 | -0.007042 | -0.00707 | 4.46e-08 | 3.97e-08 | 0.4661 | 0.4771 |
| 8 | 11252425 | 0.00745 | 0.007451 | 4.46e-08 | 4.48e-08 | 0.39 | 0.4259 |
| 8 | 8644274 | 0.007074 | 0.007062 | 4.53e-08 | 4.8e-08 | 0.4487 | 0.4764 |
| 8 | 11425809 | -0.007038 | -0.007063 | 4.79e-08 | 4.3e-08 | 0.5089 | 0.523 |
| 8 | 8685190 | 0.00706 | 0.007061 | 4.81e-08 | 4.82e-08 | 0.386 | 0.4221 |
| 8 | 8654527 | 0.007035 | 0.007021 | 4.91e-08 | 5.24e-08 | 0.4002 | 0.4282 |
| 8 | 11039816 | -0.007028 | -0.007057 | 4.94e-08 | 4.37e-08 | 0.4789 | 0.4879 |
| 12 | 62989110 | -0.00726 | -0.007303 | 1.5e-08 | 1.26e-08 | 0.5437 | 0.5569 |
| 12 | 62995984 | -0.008214 | -0.008259 | 1.52e-08 | 1.29e-08 | 0.4616 | 0.4863 |
| 12 | 62986620 | -0.008176 | -0.008222 | 1.57e-08 | 1.33e-08 | 0.4594 | 0.4815 |
| 12 | 62990403 | -0.007246 | -0.007289 | 1.6e-08 | 1.34e-08 | 0.5448 | 0.5578 |
| 12 | 62990415 | -0.007244 | -0.007287 | 1.61e-08 | 1.35e-08 | 0.545 | 0.5581 |
| 12 | 62995340 | -0.007497 | -0.007535 | 2.23e-08 | 1.93e-08 | 0.3941 | 0.4129 |
| 12 | 62945970 | -0.007478 | -0.007514 | 2.38e-08 | 2.08e-08 | 0.4074 | 0.4281 |
| 12 | 62977163 | -0.007489 | -0.007525 | 2.41e-08 | 2.1e-08 | 0.3837 | 0.4041 |
| 12 | 62995269 | -0.007479 | -0.007515 | 2.41e-08 | 2.09e-08 | 0.3821 | 0.4019 |
| 12 | 62968280 | -0.007464 | -0.0075 | 2.5e-08 | 2.18e-08 | 0.3935 | 0.4143 |
| 12 | 62948736 | -0.007468 | -0.007503 | 2.58e-08 | 2.26e-08 | 0.4162 | 0.437 |
| 12 | 62985871 | -0.007461 | -0.007499 | 2.59e-08 | 2.24e-08 | 0.3979 | 0.4168 |
| 12 | 62950556 | -0.007452 | -0.007487 | 2.66e-08 | 2.33e-08 | 0.4071 | 0.4277 |
| 12 | 62942258 | -0.007438 | -0.007472 | 2.83e-08 | 2.5e-08 | 0.4196 | 0.4411 |
| 12 | 62960880 | -0.007405 | -0.007442 | 2.98e-08 | 2.59e-08 | 0.4136 | 0.4331 |
| 12 | 62961299 | -0.007425 | -0.00746 | 3.05e-08 | 2.67e-08 | 0.4013 | 0.4224 |
| 12 | 62957906 | -0.007389 | -0.007426 | 3.25e-08 | 2.83e-08 | 0.4206 | 0.44 |
| 12 | 62996061 | -0.007427 | -0.007465 | 3.29e-08 | 2.85e-08 | 0.4224 | 0.4422 |
| 12 | 62993793 | -0.007428 | -0.007466 | 3.31e-08 | 2.87e-08 | 0.4273 | 0.447 |
| 12 | 62952696 | -0.007403 | -0.007437 | 3.33e-08 | 2.93e-08 | 0.4048 | 0.4265 |
| 12 | 62949110 | -0.007376 | -0.007411 | 3.38e-08 | 2.95e-08 | 0.4214 | 0.4418 |
| 12 | 62961801 | -0.007377 | -0.007414 | 3.42e-08 | 2.98e-08 | 0.4179 | 0.4376 |
| 12 | 62975307 | -0.007408 | -0.007445 | 3.48e-08 | 3.02e-08 | 0.4223 | 0.4428 |
| 12 | 62937532 | -0.007354 | -0.007397 | 3.52e-08 | 2.98e-08 | 0.5161 | 0.532 |
| 12 | 62990871 | -0.007389 | -0.007426 | 3.56e-08 | 3.09e-08 | 0.3996 | 0.4188 |
| 12 | 62964552 | -0.007429 | -0.007464 | 3.57e-08 | 3.14e-08 | 0.442 | 0.4655 |
| 12 | 62970482 | -0.007376 | -0.007414 | 3.63e-08 | 3.14e-08 | 0.442 | 0.4614 |
| 12 | 62945245 | -0.007386 | -0.00742 | 3.73e-08 | 3.28e-08 | 0.4283 | 0.4506 |
| 12 | 62952910 | -0.007379 | -0.007415 | 3.9e-08 | 3.41e-08 | 0.4357 | 0.4571 |
| 12 | 62980580 | -0.007328 | -0.007365 | 4.22e-08 | 3.66e-08 | 0.4088 | 0.4278 |
| 12 | 62988288 | -0.007354 | -0.007392 | 4.38e-08 | 3.8e-08 | 0.431 | 0.4509 |
| 12 | 62870574 | -0.007274 | -0.007313 | 4.74e-08 | 4.07e-08 | 0.4334 | 0.4505 |
| 12 | 62941837 | -0.007297 | -0.007331 | 4.93e-08 | 4.33e-08 | 0.4417 | 0.4622 |
| 12 | 62941426 | -0.007294 | -0.007329 | 4.97e-08 | 4.37e-08 | 0.4393 | 0.4599 |
| 13 | 83312341 | -0.007206 | -0.007155 | 3.23e-08 | 4.11e-08 | 0 | 0 |
| 13 | 83311141 | -0.007179 | -0.007128 | 3.52e-08 | 4.48e-08 | 0 | 0 |
| 13 | 83312717 | -0.007177 | -0.007129 | 3.74e-08 | 4.69e-08 | 0 | 0 |
| 13 | 83304898 | -0.007095 | -0.007045 | 4.87e-08 | 6.14e-08 | 0 | 0 |
| 16 | 13040514 | -0.008169 | -0.008077 | 2.19e-08 | 3.21e-08 | 0 | 0 |
| 16 | 13041027 | -0.008021 | -0.007929 | 3.66e-08 | 5.32e-08 | 0 | 0 |

| chromosome | position | beta | beta_LL_cytoSNP_out | p-value | p-value_LL_cytoSNP_out | i2 | i2_LL_cytoSNP_out |
|---|---|---|---|---|---|---|---|
| 16 | 13042097 | -0.00802 | -0.007927 | 3.66e-08 | 5.36e-08 | 0 | 0 |
| 16 | 13039154 | -0.008713 | -0.008609 | 3.79e-08 | 5.59e-08 | 0 | 0 |
| 16 | 13040889 | -0.008004 | -0.007912 | 3.86e-08 | 5.6e-08 | 0 | 0 |
| 16 | 13043452 | -0.007971 | -0.00788 | 4.41e-08 | 6.36e-08 | 0 | 0 |
| 16 | 13041924 | -0.007968 | -0.007877 | 4.44e-08 | 6.42e-08 | 0 | 0 |

#### Supplementary Table S6

Genetic correlation across cohorts (N > 5000 individuals)

| trait_A | trait_B | gencov | gencov_se | mean_z1z2 | cov_intercept | cov_int_se | rg | rg_se | z_score | p_value | p_fdr |
|---|---|---|---|---|---|---|---|---|---|---|---|
| FinnGen | UKB | 0.0157 | 0.002 | 0.0812 | -0.0017 | 0.0061 | 0.7299 | 0.0851 | 8.578 | 9.62e-18 | 9.62e-17 |
| EBB | UKB | 0.0173 | 0.0031 | 0.0624 | 0.0017 | 0.0061 | 0.6928 | 0.1451 | 4.774 | 1.81e-06 | 6.03e-06 |
| LLgsa | UKB | 0.045 | 0.0067 | 0.0588 | 0.005 | 0.0058 | 0.6911 | 0.144 | 4.798 | 1.61e-06 | 6.03e-06 |
| LLaffy | UKB | 0.05 | 0.007 | 0.066 | 0.0049 | 0.0054 | 0.8764 | 0.1969 | 4.452 | 8.52e-06 | 2.13e-05 |
| EBB | FinnGen | 0.0066 | 0.0016 | 0.0441 | 0.003 | 0.0059 | 0.8138 | 0.1945 | 4.184 | 2.86e-05 | 5.72e-05 |
| EBB | LLaffy | 0.0137 | 0.0051 | 0.019 | -0.0009 | 0.005 | 0.64 | 0.2868 | 2.231 | 0.0257 | 0.03671 |
| FinnGen | LLgsa | 0.0091 | 0.0038 | 0.0177 | -0.0019 | 0.0053 | 0.432 | 0.1918 | 2.253 | 0.0243 | 0.03671 |
| LLaffy | LLgsa | 0.0326 | 0.0144 | 0.0328 | 0.0169 | 0.0057 | 0.6022 | 0.2873 | 2.096 | 0.036 | 0.045 |
| EBB | LLgsa | 0.006 | 0.006 | 0.014 | 0.0055 | 0.0055 | 0.2489 | 0.2478 | 1.004 | 0.3152 | 0.3222 |
| FinnGen | LLaffy | 0.0036 | 0.0035 | 0.0182 | 0.0102 | 0.0051 | 0.1967 | 0.1987 | 0.9899 | 0.3222 | 0.3222 |

#### Supplementary Table S7

Comparison of tinnitus phenotypes (meta-analysis vs self-report vs icd coded)

*part 1 of 3 — first column repeated in every part*

| rs_number | reference_allele_meta | other_allele_meta | eaf_meta | beta_meta | se_meta | z_meta | p-value_meta | _-log10_p-value_meta | q_statistic_meta | q_p-value_meta | i2_meta | n_studies_meta | n_samples_meta |
|---|---|---|---|---|---|---|---|---|---|---|---|---|---|
| 11:92526398 | G | A | 0.9999 | -0.7084 | 0.1263 | -5.609 | 2.09e-08 | 7.68 | 0 | 1 | NA | 1 | 146929 |
| 11:92529002 | A | T | 0.9999 | -0.7082 | 0.1263 | -5.607 | 2.11e-08 | 7.676 | 0 | 1 | NA | 1 | 146929 |
| 12:62870574 | C | G | 0.6679 | -0.007274 | 0.001331 | -5.465 | 4.74e-08 | 7.324 | 31.77 | 0.02341 | 0.4334 | 19 | 913254 |
| 12:62937532 | C | T | 0.6662 | -0.007354 | 0.001333 | -5.518 | 3.52e-08 | 7.454 | 35.13 | 0.005982 | 0.5161 | 18 | 911943 |
| 12:62941426 | T | C | 0.6712 | -0.007294 | 0.001337 | -5.456 | 4.97e-08 | 7.303 | 32.1 | 0.02138 | 0.4393 | 19 | 913250 |
| 12:62941837 | A | G | 0.6714 | -0.007297 | 0.001337 | -5.458 | 4.93e-08 | 7.307 | 32.24 | 0.0206 | 0.4417 | 19 | 913249 |
| 12:62942258 | C | T | 0.6725 | -0.007438 | 0.001339 | -5.556 | 2.83e-08 | 7.548 | 31.01 | 0.02869 | 0.4196 | 19 | 913256 |
| 12:62945245 | G | A | 0.6723 | -0.007386 | 0.001341 | -5.507 | 3.73e-08 | 7.428 | 29.73 | 0.02833 | 0.4283 | 18 | 911969 |
| 12:62945970 | C | G | 0.6723 | -0.007478 | 0.001339 | -5.586 | 2.38e-08 | 7.623 | 30.37 | 0.03397 | 0.4074 | 19 | 913256 |
| 12:62948736 | G | A | 0.6724 | -0.007468 | 0.00134 | -5.572 | 2.58e-08 | 7.589 | 30.83 | 0.0301 | 0.4162 | 19 | 913256 |
| 12:62949110 | A | C | 0.669 | -0.007376 | 0.001335 | -5.525 | 3.38e-08 | 7.471 | 31.11 | 0.02797 | 0.4214 | 19 | 913256 |
| 12:62950556 | G | C | 0.6723 | -0.007452 | 0.001339 | -5.566 | 2.66e-08 | 7.574 | 30.36 | 0.03412 | 0.4071 | 19 | 913256 |
| 12:62952696 | C | T | 0.6722 | -0.007403 | 0.001339 | -5.527 | 3.33e-08 | 7.478 | 30.24 | 0.03515 | 0.4048 | 19 | 913257 |
| 12:62952910 | C | T | 0.6723 | -0.007379 | 0.001342 | -5.499 | 3.9e-08 | 7.409 | 30.12 | 0.02547 | 0.4357 | 18 | 911968 |
| 12:62957906 | G | C | 0.6689 | -0.007389 | 0.001336 | -5.531 | 3.25e-08 | 7.488 | 31.06 | 0.0283 | 0.4206 | 19 | 913255 |
| 12:62960880 | A | G | 0.669 | -0.007405 | 0.001335 | -5.547 | 2.98e-08 | 7.526 | 30.7 | 0.03119 | 0.4136 | 19 | 913255 |
| 12:62961299 | C | T | 0.6724 | -0.007425 | 0.00134 | -5.543 | 3.05e-08 | 7.516 | 30.07 | 0.03681 | 0.4013 | 19 | 913256 |
| 12:62961801 | G | A | 0.669 | -0.007377 | 0.001336 | -5.523 | 3.42e-08 | 7.466 | 30.92 | 0.02937 | 0.4179 | 19 | 913256 |
| 12:62964552 | T | C | 0.6611 | -0.007429 | 0.001347 | -5.515 | 3.57e-08 | 7.447 | 28.67 | 0.02625 | 0.442 | 17 | 438286 |
| 12:62968280 | T | C | 0.6722 | -0.007464 | 0.001338 | -5.577 | 2.5e-08 | 7.602 | 29.68 | 0.04068 | 0.3935 | 19 | 913255 |
| 12:62970482 | A | G | 0.669 | -0.007376 | 0.001338 | -5.512 | 3.63e-08 | 7.44 | 30.46 | 0.02319 | 0.442 | 18 | 911967 |
| 12:62975307 | C | T | 0.6722 | -0.007408 | 0.001342 | -5.52 | 3.48e-08 | 7.459 | 29.43 | 0.03079 | 0.4223 | 18 | 911967 |
| 12:62977163 | G | A | 0.6723 | -0.007489 | 0.001341 | -5.584 | 2.41e-08 | 7.618 | 29.21 | 0.0459 | 0.3837 | 19 | 913247 |
| 12:62980580 | T | A | 0.6689 | -0.007328 | 0.001336 | -5.486 | 4.22e-08 | 7.375 | 30.45 | 0.03333 | 0.4088 | 19 | 913255 |
| 12:62985871 | C | A | 0.6722 | -0.007461 | 0.001339 | -5.571 | 2.59e-08 | 7.586 | 29.89 | 0.03849 | 0.3979 | 19 | 913254 |
| 12:62986620 | G | A | 0.6689 | -0.008176 | 0.001445 | -5.658 | 1.57e-08 | 7.805 | 25.9 | 0.02669 | 0.4594 | 15 | 887238 |
| 12:62988288 | G | A | 0.6723 | -0.007354 | 0.001342 | -5.479 | 4.38e-08 | 7.358 | 29.87 | 0.02726 | 0.431 | 18 | 911967 |
| 12:62989110 | A | C | 0.4966 | -0.00726 | 0.001281 | -5.666 | 1.5e-08 | 7.823 | 39.45 | 0.002477 | 0.5437 | 19 | 913254 |
| 12:62990403 | A | G | 0.4964 | -0.007246 | 0.001281 | -5.655 | 1.6e-08 | 7.796 | 39.54 | 0.00241 | 0.5448 | 19 | 913254 |
| 12:62990415 | T | C | 0.4964 | -0.007244 | 0.001281 | -5.653 | 1.61e-08 | 7.792 | 39.56 | 0.002394 | 0.545 | 19 | 913254 |
| 12:62990871 | C | A | 0.6722 | -0.007389 | 0.00134 | -5.516 | 3.56e-08 | 7.449 | 29.98 | 0.03762 | 0.3996 | 19 | 913255 |
| 12:62993793 | G | A | 0.6692 | -0.007428 | 0.001344 | -5.528 | 3.31e-08 | 7.48 | 29.69 | 0.02871 | 0.4273 | 18 | 717861 |
| 12:62995269 | A | G | 0.6722 | -0.007479 | 0.001339 | -5.584 | 2.41e-08 | 7.618 | 29.13 | 0.0468 | 0.3821 | 19 | 913255 |
| 12:62995340 | G | A | 0.6723 | -0.007497 | 0.001339 | -5.597 | 2.23e-08 | 7.651 | 29.71 | 0.04039 | 0.3941 | 19 | 913254 |
| 12:62995984 | C | T | 0.6721 | -0.008214 | 0.00145 | -5.664 | 1.52e-08 | 7.819 | 24.15 | 0.02982 | 0.4616 | 14 | 885924 |
| 12:62996061 | G | A | 0.6723 | -0.007427 | 0.001343 | -5.529 | 3.29e-08 | 7.483 | 29.43 | 0.03074 | 0.4224 | 18 | 911966 |
| 13:83304898 | T | G | 0.5985 | -0.007095 | 0.001299 | -5.46 | 4.87e-08 | 7.313 | 14.52 | 0.63 | 0 | 18 | 911943 |
| 13:83311141 | G | A | 0.5985 | -0.007179 | 0.001301 | -5.517 | 3.52e-08 | 7.453 | 14.15 | 0.6563 | 0 | 18 | 911943 |
| 13:83312341 | C | T | 0.5983 | -0.007206 | 0.001303 | -5.532 | 3.23e-08 | 7.49 | 13.98 | 0.6682 | 0 | 18 | 911943 |
| 13:83312717 | G | C | 0.5979 | -0.007177 | 0.001303 | -5.507 | 3.74e-08 | 7.427 | 13.55 | 0.6984 | 0 | 18 | 911943 |
| 16:13039154 | C | T | 0.6795 | -0.008713 | 0.001583 | -5.505 | 3.79e-08 | 7.422 | 11.92 | 0.6125 | 0 | 15 | 887212 |
| 16:13040514 | C | T | 0.6805 | -0.008169 | 0.001459 | -5.601 | 2.19e-08 | 7.66 | 11.86 | 0.7537 | 0 | 17 | 910655 |
| 16:13040889 | C | G | 0.6806 | -0.008004 | 0.001455 | -5.501 | 3.86e-08 | 7.414 | 14.56 | 0.6271 | 0 | 18 | 911943 |

| rs_number | reference_allele_meta | other_allele_meta | eaf_meta | beta_meta | se_meta | z_meta | p-value_meta | _-log10_p-value_meta | q_statistic_meta | q_p-value_meta | i2_meta | n_studies_meta | n_samples_meta |
|---|---|---|---|---|---|---|---|---|---|---|---|---|---|
| 16:13041027 | G | T | 0.6806 | -0.008021 | 0.001456 | -5.511 | 3.66e-08 | 7.437 | 14.56 | 0.6275 | 0 | 18 | 911943 |
| 16:13041924 | A | G | 0.6806 | -0.007968 | 0.001455 | -5.477 | 4.44e-08 | 7.353 | 14.65 | 0.6207 | 0 | 18 | 911943 |
| 16:13042097 | G | A | 0.6807 | -0.00802 | 0.001456 | -5.51 | 3.66e-08 | 7.436 | 14.76 | 0.6131 | 0 | 18 | 911943 |
| 16:13043452 | A | G | 0.6805 | -0.007971 | 0.001455 | -5.478 | 4.41e-08 | 7.356 | 14.98 | 0.597 | 0 | 18 | 911943 |
| 1:110500725 | C | T | 0.9999 | -0.5838 | 0.1016 | -5.746 | 9.37e-09 | 8.028 | 0 | 1 | NA | 1 | 146929 |
| 1:110501839 | A | G | 0.000156 | 0.4661 | 0.08529 | 5.465 | 4.73e-08 | 7.325 | 0.000136 | 0.9907 | 0 | 2 | 171401 |
| 2:98427382 | T | C | 0.4907 | 0.008 | 0.001426 | 5.61 | 2.07e-08 | 7.684 | 23.85 | 0.04776 | 0.413 | 15 | 887234 |
| 2:98580012 | T | C | 0.4898 | 0.008074 | 0.001427 | 5.658 | 1.57e-08 | 7.805 | 22.89 | 0.08647 | 0.3447 | 16 | 888525 |
| 8:10147398 | C | T | 0.4776 | -0.007824 | 0.001293 | -6.053 | 1.47e-09 | 8.834 | 22.45 | 0.1679 | 0.2429 | 18 | 911943 |
| 8:10903475 | T | A | 0.5141 | -0.007448 | 0.001291 | -5.771 | 8.09e-09 | 8.092 | 35.35 | 0.003556 | 0.5474 | 17 | 910655 |
| 8:10938260 | G | A | 0.5198 | -0.007176 | 0.001289 | -5.569 | 2.63e-08 | 7.58 | 33.96 | 0.005507 | 0.5288 | 17 | 910655 |
| 8:10986837 | T | G | 0.5729 | -0.007109 | 0.001298 | -5.478 | 4.41e-08 | 7.356 | 31.81 | 0.01058 | 0.4971 | 17 | 910655 |
| 8:10987199 | T | C | 0.573 | -0.007183 | 0.001296 | -5.541 | 3.08e-08 | 7.511 | 30.96 | 0.02021 | 0.4509 | 18 | 911943 |
| 8:10987553 | T | C | 0.5731 | -0.007267 | 0.001298 | -5.6 | 2.2e-08 | 7.657 | 30.62 | 0.01504 | 0.4775 | 17 | 910655 |
| 8:10987651 | T | G | 0.573 | -0.007225 | 0.001298 | -5.567 | 2.65e-08 | 7.577 | 31.1 | 0.01307 | 0.4855 | 17 | 910655 |
| 8:10988275 | A | G | 0.5736 | -0.008199 | 0.001407 | -5.827 | 5.79e-09 | 8.237 | 26.36 | 0.01522 | 0.5068 | 14 | 885924 |
| 8:11039816 | T | G | 0.5275 | -0.007028 | 0.001288 | -5.458 | 4.94e-08 | 7.307 | 30.7 | 0.01468 | 0.4789 | 17 | 910655 |
| 8:11041642 | A | T | 0.53 | -0.007042 | 0.001286 | -5.476 | 4.46e-08 | 7.351 | 31.84 | 0.01574 | 0.4661 | 18 | 911943 |
| 8:11045161 | G | A | 0.5313 | -0.007055 | 0.001285 | -5.491 | 4.08e-08 | 7.389 | 34.08 | 0.01232 | 0.4718 | 19 | 913257 |
| 8:11046209 | T | C | 0.5994 | -0.007142 | 0.001303 | -5.48 | 4.37e-08 | 7.36 | 31.5 | 0.01735 | 0.4603 | 18 | 911943 |
| 8:11046394 | C | G | 0.6006 | -0.008162 | 0.001412 | -5.78 | 7.69e-09 | 8.114 | 23.58 | 0.05146 | 0.4063 | 15 | 887212 |
| 8:11054097 | C | A | 0.549 | -0.007068 | 0.001288 | -5.488 | 4.17e-08 | 7.38 | 30.82 | 0.0302 | 0.416 | 19 | 913245 |
| 8:11055597 | C | A | 0.5451 | -0.007165 | 0.001287 | -5.566 | 2.67e-08 | 7.574 | 29.11 | 0.03351 | 0.4161 | 18 | 911943 |
| 8:11060311 | A | C | 0.5375 | -0.007402 | 0.001287 | -5.752 | 9.05e-09 | 8.044 | 34.42 | 0.01119 | 0.477 | 19 | 913243 |
| 8:11061792 | C | T | 0.5377 | -0.007402 | 0.001287 | -5.753 | 8.98e-09 | 8.047 | 35.03 | 0.009361 | 0.4862 | 19 | 913246 |
| 8:11062882 | T | C | 0.5367 | -0.007525 | 0.001287 | -5.845 | 5.19e-09 | 8.285 | 34.76 | 0.01013 | 0.4822 | 19 | 913242 |
| 8:11069960 | C | T | 0.5505 | -0.007195 | 0.001288 | -5.585 | 2.4e-08 | 7.62 | 35.47 | 0.005387 | 0.5208 | 18 | 911969 |
| 8:11070360 | C | G | 0.5463 | -0.007068 | 0.001286 | -5.496 | 3.98e-08 | 7.4 | 36.86 | 0.005468 | 0.5116 | 19 | 913241 |
| 8:11071057 | A | G | 0.5908 | -0.007484 | 0.001301 | -5.751 | 9.1e-09 | 8.041 | 34.6 | 0.004501 | 0.5376 | 17 | 910655 |
| 8:11076635 | T | G | 0.5317 | -0.007286 | 0.001289 | -5.651 | 1.63e-08 | 7.787 | 33.15 | 0.007062 | 0.5173 | 17 | 910655 |
| 8:11078781 | A | C | 0.5317 | -0.007282 | 0.00129 | -5.647 | 1.68e-08 | 7.776 | 34.31 | 0.007651 | 0.5045 | 18 | 911943 |
| 8:11078949 | G | A | 0.5332 | -0.007319 | 0.00129 | -5.675 | 1.42e-08 | 7.848 | 34.53 | 0.007158 | 0.5077 | 18 | 911943 |
| 8:11079367 | G | A | 0.5331 | -0.007379 | 0.00129 | -5.722 | 1.08e-08 | 7.967 | 33.87 | 0.00873 | 0.4981 | 18 | 911943 |
| 8:11080665 | T | A | 0.5402 | -0.00753 | 0.001294 | -5.82 | 6.04e-09 | 8.219 | 31.56 | 0.01707 | 0.4613 | 18 | 911943 |
| 8:11080675 | C | T | 0.5403 | -0.007576 | 0.001294 | -5.855 | 4.9e-09 | 8.31 | 31.41 | 0.01781 | 0.4587 | 18 | 911943 |
| 8:11213589 | T | G | 0.4306 | 0.007102 | 0.00129 | 5.507 | 3.74e-08 | 7.427 | 28.11 | 0.0307 | 0.4307 | 17 | 910655 |
| 8:11214455 | A | G | 0.47 | 0.00714 | 0.001286 | 5.552 | 2.89e-08 | 7.539 | 24.77 | 0.09996 | 0.3137 | 18 | 911943 |
| 8:11214972 | A | G | 0.4697 | 0.007146 | 0.001286 | 5.558 | 2.8e-08 | 7.552 | 25.72 | 0.07979 | 0.3389 | 18 | 911943 |
| 8:11215617 | C | G | 0.4304 | 0.007177 | 0.001288 | 5.574 | 2.55e-08 | 7.594 | 27.72 | 0.04835 | 0.3867 | 18 | 911943 |
| 8:11219334 | G | A | 0.4312 | 0.007168 | 0.001291 | 5.554 | 2.87e-08 | 7.543 | 28.6 | 0.0268 | 0.4405 | 17 | 910655 |
| 8:11219781 | C | T | 0.4314 | 0.007242 | 0.00129 | 5.614 | 2.03e-08 | 7.693 | 28.35 | 0.04105 | 0.4003 | 18 | 911943 |
| 8:11225168 | G | A | 0.5225 | -0.007119 | 0.001286 | -5.535 | 3.18e-08 | 7.497 | 24.75 | 0.1005 | 0.3131 | 18 | 911943 |
| 8:11225480 | T | C | 0.5212 | -0.007088 | 0.001286 | -5.511 | 3.66e-08 | 7.437 | 24.7 | 0.1016 | 0.3118 | 18 | 911943 |
| 8:11225910 | A | G | 0.5217 | -0.007125 | 0.001288 | -5.531 | 3.27e-08 | 7.486 | 24.76 | 0.07423 | 0.3537 | 17 | 910655 |
| 8:11226071 | A | G | 0.5664 | -0.007704 | 0.00129 | -5.971 | 2.43e-09 | 8.615 | 29.17 | 0.03303 | 0.4171 | 18 | 911943 |
| 8:11226456 | C | A | 0.5726 | -0.007853 | 0.001293 | -6.073 | 1.29e-09 | 8.888 | 28.79 | 0.03654 | 0.4095 | 18 | 911943 |
| 8:11227406 | G | A | 0.5709 | -0.007569 | 0.001292 | -5.86 | 4.75e-09 | 8.323 | 28.9 | 0.03546 | 0.4118 | 18 | 911943 |

| rs_number | reference_allele_meta | other_allele_meta | eaf_meta | beta_meta | se_meta | z_meta | p-value_meta | _-log10_p-value_meta | q_statistic_meta | q_p-value_meta | i2_meta | n_studies_meta | n_samples_meta |
|---|---|---|---|---|---|---|---|---|---|---|---|---|---|
| 8:11227885 | C | T | 0.5714 | -0.007563 | 0.001292 | -5.855 | 4.9e-09 | 8.31 | 29.54 | 0.02983 | 0.4246 | 18 | 911943 |
| 8:11228100 | G | A | 0.5709 | -0.007556 | 0.001291 | -5.852 | 4.98e-09 | 8.303 | 28.97 | 0.03478 | 0.4132 | 18 | 911943 |
| 8:11228672 | G | A | 0.478 | 0.00705 | 0.001285 | 5.488 | 4.16e-08 | 7.381 | 25.95 | 0.07544 | 0.3448 | 18 | 911943 |
| 8:11229319 | C | G | 0.5714 | -0.007531 | 0.001293 | -5.824 | 5.9e-09 | 8.229 | 29.61 | 0.02927 | 0.4259 | 18 | 911943 |
| 8:11229638 | C | T | 0.5705 | -0.007456 | 0.001293 | -5.769 | 8.2e-09 | 8.086 | 28.78 | 0.02547 | 0.4441 | 17 | 910655 |
| 8:11230206 | G | T | 0.5712 | -0.007537 | 0.001292 | -5.835 | 5.51e-09 | 8.259 | 28.93 | 0.03519 | 0.4123 | 18 | 911943 |
| 8:11230259 | A | T | 0.5711 | -0.007527 | 0.001292 | -5.827 | 5.79e-09 | 8.237 | 29.02 | 0.03436 | 0.4141 | 18 | 911943 |
| 8:11230574 | G | A | 0.5712 | -0.007598 | 0.001292 | -5.882 | 4.15e-09 | 8.382 | 29.06 | 0.03396 | 0.415 | 18 | 911943 |
| 8:11231249 | T | C | 0.4387 | 0.007613 | 0.001287 | 5.917 | 3.38e-09 | 8.472 | 28.43 | 0.04017 | 0.402 | 18 | 911943 |
| 8:11231354 | G | T | 0.4393 | 0.007478 | 0.001287 | 5.809 | 6.46e-09 | 8.19 | 28.66 | 0.03783 | 0.4068 | 18 | 911943 |
| 8:11231886 | A | G | 0.5657 | -0.007569 | 0.001289 | -5.872 | 4.41e-09 | 8.355 | 29.51 | 0.0301 | 0.4239 | 18 | 911943 |
| 8:11232343 | C | G | 0.5706 | -0.007538 | 0.001291 | -5.838 | 5.42e-09 | 8.266 | 30.48 | 0.02305 | 0.4423 | 18 | 911943 |
| 8:11232788 | G | A | 0.5716 | -0.007669 | 0.001292 | -5.937 | 2.99e-09 | 8.524 | 29.87 | 0.0273 | 0.4309 | 18 | 911943 |
| 8:11232860 | G | A | 0.5714 | -0.007565 | 0.001291 | -5.859 | 4.79e-09 | 8.32 | 30.64 | 0.02209 | 0.4451 | 18 | 911943 |
| 8:11233318 | A | C | 0.5659 | -0.007603 | 0.001289 | -5.9 | 3.74e-09 | 8.427 | 28.33 | 0.04119 | 0.4 | 18 | 911943 |
| 8:11233419 | T | G | 0.5659 | -0.007571 | 0.001289 | -5.875 | 4.35e-09 | 8.362 | 28.36 | 0.04087 | 0.4006 | 18 | 911943 |
| 8:11233582 | A | G | 0.5659 | -0.007535 | 0.001289 | -5.846 | 5.16e-09 | 8.287 | 28.27 | 0.04184 | 0.3987 | 18 | 911943 |
| 8:11233659 | A | G | 0.5659 | -0.007534 | 0.001289 | -5.846 | 5.18e-09 | 8.286 | 28.26 | 0.04199 | 0.3984 | 18 | 911943 |
| 8:11233917 | C | T | 0.5707 | -0.007534 | 0.001291 | -5.836 | 5.51e-09 | 8.259 | 30.09 | 0.02573 | 0.435 | 18 | 911943 |
| 8:11233958 | A | G | 0.5707 | -0.007539 | 0.001291 | -5.84 | 5.37e-09 | 8.27 | 30.06 | 0.02589 | 0.4345 | 18 | 911943 |
| 8:11234298 | C | T | 0.5706 | -0.007542 | 0.00129 | -5.846 | 5.18e-09 | 8.286 | 29.29 | 0.03198 | 0.4195 | 18 | 911943 |
| 8:11234367 | T | G | 0.57 | -0.007527 | 0.00129 | -5.834 | 5.57e-09 | 8.254 | 29.14 | 0.03325 | 0.4166 | 18 | 911943 |
| 8:11234500 | A | T | 0.5656 | -0.007536 | 0.001288 | -5.85 | 5.04e-09 | 8.297 | 28.81 | 0.03628 | 0.41 | 18 | 911943 |
| 8:11234520 | A | G | 0.5706 | -0.007548 | 0.00129 | -5.85 | 5.04e-09 | 8.297 | 29.62 | 0.02925 | 0.426 | 18 | 911943 |
| 8:11234613 | T | A | 0.5707 | -0.007523 | 0.001291 | -5.827 | 5.78e-09 | 8.238 | 29.5 | 0.03021 | 0.4237 | 18 | 911943 |
| 8:11234626 | T | C | 0.5707 | -0.00753 | 0.001291 | -5.832 | 5.61e-09 | 8.251 | 29.49 | 0.03025 | 0.4236 | 18 | 911943 |
| 8:11234780 | C | G | 0.5706 | -0.007531 | 0.00129 | -5.837 | 5.47e-09 | 8.262 | 29.49 | 0.03024 | 0.4236 | 18 | 911943 |
| 8:11234844 | G | A | 0.5706 | -0.007537 | 0.00129 | -5.842 | 5.31e-09 | 8.275 | 29.48 | 0.03034 | 0.4234 | 18 | 911943 |
| 8:11234885 | C | G | 0.5705 | -0.007541 | 0.00129 | -5.848 | 5.12e-09 | 8.291 | 29.53 | 0.02995 | 0.4243 | 18 | 911943 |
| 8:11235136 | C | G | 0.5707 | -0.00751 | 0.00129 | -5.82 | 6.04e-09 | 8.219 | 29.66 | 0.02887 | 0.4269 | 18 | 911943 |
| 8:11235150 | C | A | 0.5716 | -0.00851 | 0.001397 | -6.092 | 1.15e-09 | 8.94 | 22.28 | 0.07298 | 0.3717 | 15 | 887212 |
| 8:11235360 | A | C | 0.5706 | -0.007524 | 0.00129 | -5.833 | 5.59e-09 | 8.253 | 29.78 | 0.028 | 0.4291 | 18 | 911943 |
| 8:11235393 | A | G | 0.5705 | -0.007485 | 0.00129 | -5.804 | 6.65e-09 | 8.177 | 29.76 | 0.02812 | 0.4288 | 18 | 911943 |
| 8:11235497 | G | A | 0.5705 | -0.007514 | 0.001289 | -5.83 | 5.7e-09 | 8.244 | 29.83 | 0.03918 | 0.3965 | 19 | 913238 |
| 8:11235579 | T | G | 0.5701 | -0.007475 | 0.00129 | -5.794 | 7.06e-09 | 8.151 | 29.57 | 0.04179 | 0.3914 | 19 | 913239 |
| 8:11235605 | T | G | 0.5706 | -0.007492 | 0.00129 | -5.807 | 6.55e-09 | 8.184 | 29.85 | 0.03896 | 0.3969 | 19 | 913240 |
| 8:11235735 | T | C | 0.5705 | -0.007512 | 0.00129 | -5.825 | 5.85e-09 | 8.233 | 29.63 | 0.02914 | 0.4263 | 18 | 911943 |
| 8:11235910 | G | A | 0.5744 | -0.007423 | 0.001292 | -5.746 | 9.37e-09 | 8.028 | 31.39 | 0.01788 | 0.4585 | 18 | 911943 |
| 8:11236392 | T | C | 0.5711 | -0.007572 | 0.001292 | -5.862 | 4.7e-09 | 8.328 | 30.1 | 0.03648 | 0.402 | 19 | 913246 |
| 8:11236413 | C | T | 0.5711 | -0.007601 | 0.001292 | -5.885 | 4.1e-09 | 8.387 | 30.32 | 0.03448 | 0.4063 | 19 | 913246 |
| 8:11236419 | T | C | 0.5711 | -0.007605 | 0.001292 | -5.888 | 4.02e-09 | 8.396 | 30.21 | 0.03543 | 0.4042 | 19 | 913246 |
| 8:11236572 | T | A | 0.5704 | -0.00758 | 0.001288 | -5.884 | 4.12e-09 | 8.385 | 29.82 | 0.03927 | 0.3963 | 19 | 913247 |
| 8:11236681 | T | G | 0.5705 | -0.007595 | 0.001289 | -5.892 | 3.92e-09 | 8.407 | 29.43 | 0.04331 | 0.3885 | 19 | 913247 |
| 8:11236685 | C | T | 0.5705 | -0.007594 | 0.001289 | -5.892 | 3.93e-09 | 8.406 | 29.46 | 0.04308 | 0.3889 | 19 | 913247 |
| 8:11236809 | T | C | 0.5703 | -0.007484 | 0.001288 | -5.812 | 6.35e-09 | 8.197 | 29.69 | 0.0406 | 0.3937 | 19 | 913246 |
| 8:11236850 | C | T | 0.5705 | -0.007503 | 0.001288 | -5.827 | 5.81e-09 | 8.236 | 29.55 | 0.04202 | 0.3909 | 19 | 913247 |
| 8:11236964 | C | T | 0.5707 | -0.007504 | 0.001288 | -5.827 | 5.81e-09 | 8.236 | 29.4 | 0.04366 | 0.3878 | 19 | 913243 |

| rs_number | reference_allele_meta | other_allele_meta | eaf_meta | beta_meta | se_meta | z_meta | p-value_meta | _-log10_p-value_meta | q_statistic_meta | q_p-value_meta | i2_meta | n_studies_meta | n_samples_meta |
|---|---|---|---|---|---|---|---|---|---|---|---|---|---|
| 8:11236975 | A | G | 0.5705 | -0.007528 | 0.001289 | -5.839 | 5.4e-09 | 8.268 | 29.21 | 0.03267 | 0.4179 | 18 | 911969 |
| 8:11237477 | A | G | 0.5708 | -0.007577 | 0.001289 | -5.878 | 4.26e-09 | 8.371 | 28.89 | 0.04976 | 0.3769 | 19 | 913247 |
| 8:11237480 | A | G | 0.5708 | -0.007577 | 0.001289 | -5.878 | 4.26e-09 | 8.371 | 28.94 | 0.04907 | 0.3781 | 19 | 913247 |
| 8:11237756 | C | A | 0.5744 | -0.007376 | 0.001285 | -5.739 | 9.75e-09 | 8.011 | 32 | 0.01504 | 0.4688 | 18 | 911943 |
| 8:11237773 | T | C | 0.5746 | -0.007413 | 0.00129 | -5.749 | 9.23e-09 | 8.035 | 31.86 | 0.01568 | 0.4664 | 18 | 911943 |
| 8:11238029 | C | T | 0.5754 | -0.007501 | 0.001291 | -5.81 | 6.4e-09 | 8.194 | 32.07 | 0.01476 | 0.4699 | 18 | 911943 |
| 8:11238597 | C | T | 0.5751 | -0.007428 | 0.001291 | -5.754 | 8.92e-09 | 8.05 | 31.65 | 0.01664 | 0.4628 | 18 | 911943 |
| 8:11239017 | T | A | 0.5751 | -0.007409 | 0.001291 | -5.737 | 9.87e-09 | 8.006 | 31.76 | 0.01611 | 0.4648 | 18 | 911943 |
| 8:11239054 | C | T | 0.5752 | -0.007403 | 0.001292 | -5.73 | 1.03e-08 | 7.988 | 31.9 | 0.01549 | 0.4671 | 18 | 911943 |
| 8:11239078 | T | G | 0.5751 | -0.007419 | 0.001291 | -5.745 | 9.43e-09 | 8.025 | 32 | 0.01506 | 0.4687 | 18 | 911943 |
| 8:11239137 | G | A | 0.5749 | -0.007434 | 0.001291 | -5.757 | 8.81e-09 | 8.055 | 31.9 | 0.01546 | 0.4672 | 18 | 911943 |
| 8:11239352 | G | A | 0.575 | -0.00745 | 0.001291 | -5.769 | 8.18e-09 | 8.087 | 31.94 | 0.01532 | 0.4677 | 18 | 911943 |
| 8:11239510 | G | A | 0.575 | -0.00746 | 0.001292 | -5.774 | 7.93e-09 | 8.101 | 31.87 | 0.01563 | 0.4666 | 18 | 911943 |
| 8:11239565 | C | T | 0.575 | -0.007459 | 0.001292 | -5.773 | 7.99e-09 | 8.097 | 31.83 | 0.01582 | 0.4658 | 18 | 911943 |
| 8:11239640 | C | T | 0.5752 | -0.00751 | 0.001293 | -5.809 | 6.44e-09 | 8.191 | 31.66 | 0.01656 | 0.4631 | 18 | 911943 |
| 8:11239762 | A | T | 0.5659 | -0.007547 | 0.001288 | -5.859 | 4.78e-09 | 8.321 | 27.93 | 0.04571 | 0.3914 | 18 | 911943 |
| 8:11242025 | T | G | 0.5379 | -0.007269 | 0.001299 | -5.594 | 2.28e-08 | 7.643 | 27.56 | 0.03566 | 0.4195 | 17 | 438262 |
| 8:11242632 | G | A | 0.5709 | -0.007632 | 0.001292 | -5.907 | 3.57e-09 | 8.447 | 30.41 | 0.02352 | 0.441 | 18 | 911943 |
| 8:11243126 | C | G | 0.5755 | -0.007524 | 0.001294 | -5.814 | 6.27e-09 | 8.203 | 32.43 | 0.01331 | 0.4757 | 18 | 911943 |
| 8:11245303 | C | G | 0.4339 | 0.007598 | 0.001291 | 5.887 | 4.04e-09 | 8.393 | 28.22 | 0.04247 | 0.3975 | 18 | 911943 |
| 8:11247814 | G | A | 0.3784 | 0.007548 | 0.001331 | 5.671 | 1.46e-08 | 7.836 | 26.46 | 0.06645 | 0.3576 | 18 | 911943 |
| 8:11249010 | G | T | 0.4338 | 0.007569 | 0.001292 | 5.859 | 4.78e-09 | 8.32 | 29.13 | 0.03333 | 0.4165 | 18 | 911943 |
| 8:11249261 | T | C | 0.3757 | 0.008415 | 0.001447 | 5.817 | 6.14e-09 | 8.212 | 19.09 | 0.1617 | 0.2665 | 15 | 887212 |
| 8:11251175 | A | G | 0.4415 | 0.007353 | 0.001317 | 5.583 | 2.43e-08 | 7.615 | 25.03 | 0.04957 | 0.4007 | 16 | 715261 |
| 8:11252425 | C | A | 0.506 | 0.00745 | 0.001361 | 5.476 | 4.46e-08 | 7.351 | 27.87 | 0.0465 | 0.39 | 18 | 911943 |
| 8:11425809 | T | G | 0.5366 | -0.007038 | 0.001288 | -5.463 | 4.79e-08 | 7.32 | 36.65 | 0.005817 | 0.5089 | 19 | 913234 |
| 8:8633548 | T | G | 0.4989 | -0.007318 | 0.001292 | -5.665 | 1.51e-08 | 7.821 | 31.82 | 0.01583 | 0.4658 | 18 | 911943 |
| 8:8637429 | G | A | 0.4998 | -0.007226 | 0.00129 | -5.604 | 2.15e-08 | 7.667 | 30.92 | 0.02045 | 0.4501 | 18 | 911943 |
| 8:8639740 | A | G | 0.4998 | -0.007166 | 0.00129 | -5.555 | 2.85e-08 | 7.546 | 31.85 | 0.01569 | 0.4663 | 18 | 911943 |
| 8:8641145 | C | T | 0.5003 | 0.007248 | 0.001291 | 5.615 | 2.01e-08 | 7.696 | 28.52 | 0.02738 | 0.439 | 17 | 910655 |
| 8:8644274 | G | C | 0.5126 | 0.007074 | 0.001292 | 5.473 | 4.53e-08 | 7.344 | 30.84 | 0.0209 | 0.4487 | 18 | 911943 |
| 8:8646246 | T | C | 0.5115 | 0.007416 | 0.001291 | 5.746 | 9.37e-09 | 8.028 | 28.97 | 0.02412 | 0.4478 | 17 | 910655 |
| 8:8652889 | G | A | 0.5085 | 0.007269 | 0.001287 | 5.648 | 1.67e-08 | 7.778 | 33.45 | 0.01471 | 0.4619 | 19 | 913236 |
| 8:8654527 | T | C | 0.5158 | 0.007035 | 0.001289 | 5.459 | 4.91e-08 | 7.309 | 28.34 | 0.04107 | 0.4002 | 18 | 911943 |
| 8:8660538 | C | A | 0.518 | 0.007136 | 0.001289 | 5.538 | 3.14e-08 | 7.503 | 30.68 | 0.02184 | 0.4459 | 18 | 911943 |
| 8:8661534 | T | C | 0.5086 | 0.007262 | 0.001286 | 5.649 | 1.65e-08 | 7.782 | 33.44 | 0.01474 | 0.4618 | 19 | 913244 |
| 8:8661681 | C | G | 0.5125 | 0.007243 | 0.001288 | 5.623 | 1.92e-08 | 7.716 | 30.43 | 0.02344 | 0.4413 | 18 | 911943 |
| 8:8664622 | G | A | 0.5241 | 0.007302 | 0.001292 | 5.654 | 1.61e-08 | 7.794 | 24.04 | 0.0887 | 0.3344 | 17 | 910655 |
| 8:8673601 | A | C | 0.5173 | 0.007168 | 0.001289 | 5.56 | 2.76e-08 | 7.559 | 28.45 | 0.03992 | 0.4025 | 18 | 911969 |
| 8:8673736 | T | C | 0.5182 | 0.007136 | 0.001289 | 5.538 | 3.14e-08 | 7.504 | 30.7 | 0.03117 | 0.4137 | 19 | 913246 |
| 8:8675325 | A | T | 0.5129 | 0.007152 | 0.00129 | 5.546 | 3e-08 | 7.523 | 31.55 | 0.0171 | 0.4612 | 18 | 911943 |
| 8:8678530 | G | A | 0.5217 | 0.007207 | 0.00129 | 5.586 | 2.37e-08 | 7.624 | 29.93 | 0.03814 | 0.3986 | 19 | 913239 |
| 8:8679176 | A | G | 0.5216 | 0.007118 | 0.00129 | 5.518 | 3.52e-08 | 7.454 | 29.71 | 0.04036 | 0.3941 | 19 | 913240 |
| 8:8679614 | T | C | 0.5215 | 0.00716 | 0.00129 | 5.55 | 2.92e-08 | 7.534 | 29.15 | 0.04663 | 0.3824 | 19 | 913237 |
| 8:8680477 | G | A | 0.5215 | 0.007109 | 0.00129 | 5.511 | 3.66e-08 | 7.437 | 28.89 | 0.04978 | 0.3769 | 19 | 913236 |
| 8:8682192 | A | T | 0.5216 | 0.007164 | 0.001291 | 5.551 | 2.91e-08 | 7.536 | 29.17 | 0.04639 | 0.3828 | 19 | 913236 |
| 8:8682878 | T | C | 0.5111 | 0.007108 | 0.00129 | 5.51 | 3.68e-08 | 7.434 | 34.67 | 0.006862 | 0.5097 | 18 | 911943 |

| rs_number | reference_allele_meta | other_allele_meta | eaf_meta | beta_meta | se_meta | z_meta | p-value_meta | _-log10_p-value_meta | q_statistic_meta | q_p-value_meta | i2_meta | n_studies_meta | n_samples_meta |
|---|---|---|---|---|---|---|---|---|---|---|---|---|---|
| 8:8684953 | G | A | 0.5211 | 0.007165 | 0.001293 | 5.541 | 3.09e-08 | 7.511 | 27.81 | 0.04717 | 0.3888 | 18 | 911943 |
| 8:8685190 | A | G | 0.5208 | 0.00706 | 0.001292 | 5.462 | 4.81e-08 | 7.318 | 27.69 | 0.04874 | 0.386 | 18 | 911943 |
| 8:8685646 | T | C | 0.5102 | 0.007186 | 0.001291 | 5.565 | 2.69e-08 | 7.57 | 33.26 | 0.01045 | 0.4889 | 18 | 911943 |
| 8:8685854 | A | G | 0.5105 | 0.007265 | 0.001292 | 5.624 | 1.92e-08 | 7.718 | 29.28 | 0.02213 | 0.4535 | 17 | 910655 |
| 8:8700851 | C | T | 0.5142 | 0.007429 | 0.001289 | 5.765 | 8.41e-09 | 8.075 | 34.69 | 0.006824 | 0.51 | 18 | 911943 |
| 8:8704330 | G | C | 0.519 | 0.007156 | 0.001289 | 5.553 | 2.87e-08 | 7.542 | 34.54 | 0.007154 | 0.5078 | 18 | 911943 |
| 8:8721473 | G | A | 0.5303 | 0.007253 | 0.001295 | 5.601 | 2.18e-08 | 7.661 | 35.6 | 0.005192 | 0.5224 | 18 | 911943 |
| 8:8730488 | G | A | 0.5269 | 0.007178 | 0.001297 | 5.532 | 3.24e-08 | 7.489 | 35.51 | 0.005326 | 0.5213 | 18 | 911943 |
| 8:8770512 | C | T | 0.5757 | 0.007282 | 0.001302 | 5.594 | 2.27e-08 | 7.644 | 26.14 | 0.07195 | 0.3497 | 18 | 911943 |
| 8:9569104 | T | C | 0.8201 | 0.009116 | 0.001657 | 5.502 | 3.84e-08 | 7.415 | 12.42 | 0.8247 | 0 | 19 | 913239 |
| 11:71819437 | T | C | 0.9998 | -0.4691 | 0.0858 | -5.467 | 4.67e-08 | 7.33 | 0 | 1 | NA | 1 | 146929 |
| 11:90880166 | T | C | 0.9999 | -0.7792 | 0.1359 | -5.734 | 1.01e-08 | 7.996 | 0 | 1 | NA | 1 | 146929 |
| 13:36123276 | T | A | 0.9999 | -0.5076 | 0.09064 | -5.6 | 2.19e-08 | 7.659 | 0 | 1 | NA | 1 | 146929 |
| 15:53323213 | G | A | 0.9996 | -0.229 | 0.04036 | -5.672 | 1.45e-08 | 7.84 | 8.046 | 0.004561 | 0.8757 | 2 | 148709 |
| 16:79002432 | A | G | 0.9998 | -0.429 | 0.07089 | -6.052 | 1.48e-09 | 8.831 | 0 | 1 | 0 | 1 | 146929 |
| 18:38476471 | T | C | 0.9998 | -0.324 | 0.05653 | -5.731 | 1.02e-08 | 7.991 | 0 | 1 | NA | 1 | 146929 |
| 3:74581060 | G | A | 0.9998 | -0.3863 | 0.06473 | -5.968 | 2.47e-09 | 8.607 | 0 | 1 | NA | 1 | 146929 |
| 6:38953162 | C | A | 0.9997 | -0.2589 | 0.04682 | -5.53 | 3.29e-08 | 7.483 | 0 | 1 | NA | 1 | 146929 |
| 8:11417582 | T | C | 0.625 | 0.01009 | 0.001555 | 6.486 | 9.09e-11 | 10.04 | 9.909 | 0.07785 | 0.4954 | 6 | 821622 |
| 9:126164618 | C | T | 0.9999 | -0.494 | 0.08773 | -5.631 | 1.84e-08 | 7.736 | 0 | 1 | NA | 1 | 146929 |
| 5:121781695 | T | A | 0.01 | 0.5238 | 0.08928 | 5.867 | 4.56e-09 | 8.341 | 0 | 1 | NA | 1 | 1196 |
| 1:106531864 | A | G | 0.001422 | 0.3694 | 0.06708 | 5.507 | 3.74e-08 | 7.427 | 0 | 1 | NA | 1 | 8343 |

*part 2 of 3 — first column repeated in every part*

| rs_number | reference_allele_icd | other_allele_icd | eaf_icd | beta_icd | se_icd | z_icd | p-value_icd | _-log10_p-value_icd | q_statistic_icd | q_p-value_icd | i2_icd | n_studies_icd | n_samples_icd | reference_allele_selfrep |
|---|---|---|---|---|---|---|---|---|---|---|---|---|---|---|
| 11:92526398 | NA | NA | NA | NA | NA | NA | NA | NA | NA | NA | NA | NA | NA | G |
| 11:92529002 | NA | NA | NA | NA | NA | NA | NA | NA | NA | NA | NA | NA | NA | A |
| 12:62870574 | C | G | 0.6805 | -0.008288 | 0.01136 | -0.7295 | 0.4657 | 0.3319 | 0.4164 | 0.5188 | 0 | 2 | 669075 | C |
| 12:62937532 | C | T | 0.6796 | -0.004462 | 0.01137 | -0.3925 | 0.6947 | 0.1582 | 0.9486 | 0.3301 | 0 | 2 | 669075 | C |
| 12:62941426 | T | C | 0.6821 | -0.006026 | 0.01138 | -0.5296 | 0.5964 | 0.2245 | 0.4359 | 0.5091 | 0 | 2 | 669075 | T |
| 12:62941837 | A | G | 0.6824 | -0.00537 | 0.01138 | -0.4719 | 0.637 | 0.1959 | 0.5083 | 0.4759 | 0 | 2 | 669075 | A |
| 12:62942258 | C | T | 0.6831 | -0.005687 | 0.01139 | -0.4994 | 0.6175 | 0.2094 | 0.5186 | 0.4714 | 0 | 2 | 669075 | C |
| 12:62945245 | G | A | 0.683 | -0.005642 | 0.01139 | -0.4953 | 0.6204 | 0.2073 | 0.5464 | 0.4598 | 0 | 2 | 669075 | G |
| 12:62945970 | C | G | 0.6829 | -0.005874 | 0.01139 | -0.5157 | 0.6061 | 0.2175 | 0.5725 | 0.4493 | 0 | 2 | 669075 | C |
| 12:62948736 | G | A | 0.683 | -0.005796 | 0.01139 | -0.5088 | 0.6109 | 0.214 | 0.5674 | 0.4513 | 0 | 2 | 669075 | G |
| 12:62949110 | A | C | 0.6797 | -0.006291 | 0.01137 | -0.5536 | 0.5799 | 0.2367 | 0.2985 | 0.5849 | 0 | 2 | 669075 | A |
| 12:62950556 | G | C | 0.6829 | -0.005846 | 0.01139 | -0.5131 | 0.6079 | 0.2162 | 0.5786 | 0.4469 | 0 | 2 | 669075 | G |
| 12:62952696 | C | T | 0.6829 | -0.005711 | 0.01139 | -0.5014 | 0.6161 | 0.2103 | 0.5584 | 0.4549 | 0 | 2 | 669075 | C |
| 12:62952910 | C | T | 0.6829 | -0.00572 | 0.01139 | -0.502 | 0.6157 | 0.2106 | 0.56 | 0.4543 | 0 | 2 | 669075 | C |
| 12:62957906 | G | C | 0.6796 | -0.00627 | 0.01137 | -0.5517 | 0.5812 | 0.2357 | 0.296 | 0.5864 | 0 | 2 | 669075 | G |
| 12:62960880 | A | G | 0.6796 | -0.00628 | 0.01137 | -0.5526 | 0.5806 | 0.2361 | 0.2951 | 0.587 | 0 | 2 | 669075 | A |
| 12:62961299 | C | T | 0.6829 | -0.005632 | 0.0114 | -0.4941 | 0.6212 | 0.2067 | 0.5818 | 0.4456 | 0 | 2 | 669075 | C |
| 12:62961801 | G | A | 0.6796 | -0.00628 | 0.01137 | -0.5526 | 0.5806 | 0.2361 | 0.2951 | 0.587 | 0 | 2 | 669075 | G |
| 12:62964552 | T | C | 0.6835 | -0.01443 | 0.01691 | -0.8533 | 0.3935 | 0.4051 | 0 | 1 | NA | 1 | 195394 | T |
| 12:62968280 | T | C | 0.6829 | -0.005881 | 0.01139 | -0.5161 | 0.6058 | 0.2177 | 0.567 | 0.4514 | 0 | 2 | 669075 | T |
| 12:62970482 | A | G | 0.6796 | -0.006488 | 0.01137 | -0.5709 | 0.5681 | 0.2456 | 0.3174 | 0.5732 | 0 | 2 | 669075 | A |
| 12:62975307 | C | T | 0.6828 | -0.006049 | 0.01139 | -0.531 | 0.5954 | 0.2252 | 0.6075 | 0.4357 | 0 | 2 | 669075 | C |
| 12:62977163 | G | A | 0.6828 | -0.006062 | 0.01139 | -0.5322 | 0.5946 | 0.2258 | 0.6109 | 0.4345 | 0 | 2 | 669075 | G |
| 12:62980580 | T | A | 0.6796 | -0.006524 | 0.01137 | -0.5741 | 0.5659 | 0.2472 | 0.3296 | 0.5659 | 0 | 2 | 669075 | T |
| 12:62985871 | C | A | 0.6828 | -0.005978 | 0.01139 | -0.5246 | 0.5999 | 0.2219 | 0.598 | 0.4393 | 0 | 2 | 669075 | C |
| 12:62986620 | G | A | 0.6796 | -0.006507 | 0.01137 | -0.5726 | 0.5669 | 0.2465 | 0.3184 | 0.5726 | 0 | 2 | 669075 | G |
| 12:62988288 | G | A | 0.6828 | -0.005955 | 0.01139 | -0.5227 | 0.6012 | 0.221 | 0.6008 | 0.4383 | 0 | 2 | 669075 | G |
| 12:62989110 | A | C | 0.4879 | -0.01234 | 0.01061 | -1.163 | 0.2449 | 0.6111 | 1.388 | 0.2387 | 0.2796 | 2 | 669075 | A |
| 12:62990403 | A | G | 0.4876 | -0.01223 | 0.01062 | -1.152 | 0.2493 | 0.6034 | 1.363 | 0.243 | 0.2665 | 2 | 669075 | A |
| 12:62990415 | T | C | 0.4876 | -0.01223 | 0.01062 | -1.152 | 0.2493 | 0.6034 | 1.363 | 0.243 | 0.2665 | 2 | 669075 | T |
| 12:62990871 | C | A | 0.6828 | -0.005776 | 0.01139 | -0.5069 | 0.6122 | 0.2131 | 0.6232 | 0.4299 | 0 | 2 | 669075 | C |
| 12:62993793 | G | A | 0.6826 | 0.001747 | 0.01542 | 0.1133 | 0.9098 | 0.04106 | 0 | 1 | NA | 1 | 473681 | G |
| 12:62995269 | A | G | 0.6829 | -0.006013 | 0.01139 | -0.5278 | 0.5977 | 0.2235 | 0.5417 | 0.4617 | 0 | 2 | 669075 | A |
| 12:62995340 | G | A | 0.6828 | -0.006004 | 0.01139 | -0.5269 | 0.5983 | 0.2231 | 0.5429 | 0.4612 | 0 | 2 | 669075 | G |
| 12:62995984 | C | T | 0.6827 | -0.005924 | 0.01139 | -0.5199 | 0.6032 | 0.2196 | 0.5523 | 0.4574 | 0 | 2 | 669075 | C |
| 12:62996061 | G | A | 0.6828 | -0.006007 | 0.01139 | -0.5272 | 0.5981 | 0.2233 | 0.5424 | 0.4614 | 0 | 2 | 669075 | G |
| 13:83304898 | T | G | 0.6067 | 0.004271 | 0.0109 | 0.3917 | 0.6953 | 0.1578 | 0.1013 | 0.7502 | 0 | 2 | 669075 | T |
| 13:83311141 | G | A | 0.6068 | 0.004223 | 0.0109 | 0.3876 | 0.6983 | 0.1559 | 0.1174 | 0.7319 | 0 | 2 | 669075 | G |
| 13:83312341 | C | T | 0.6068 | 0.004047 | 0.01089 | 0.3715 | 0.7102 | 0.1486 | 0.1033 | 0.7479 | 0 | 2 | 669075 | C |
| 13:83312717 | G | C | 0.6063 | 0.004344 | 0.01089 | 0.399 | 0.6899 | 0.1612 | 0.1368 | 0.7115 | 0 | 2 | 669075 | G |
| 16:13039154 | C | T | 0.6607 | -0.002665 | 0.01143 | -0.2332 | 0.8156 | 0.08853 | 0.004627 | 0.9458 | 0 | 2 | 669075 | C |
| 16:13040514 | C | T | 0.6607 | -0.002607 | 0.01142 | -0.2283 | 0.8194 | 0.08649 | 0.00468 | 0.9455 | 0 | 2 | 669075 | C |
| 16:13040889 | C | G | 0.6607 | -0.002728 | 0.01142 | -0.2388 | 0.8113 | 0.09084 | 0.003637 | 0.9519 | 0 | 2 | 669075 | C |
| 16:13041027 | G | T | 0.6607 | -0.002641 | 0.01142 | -0.2312 | 0.8171 | 0.0877 | 0.004343 | 0.9475 | 0 | 2 | 669075 | G |
| 16:13041924 | A | G | 0.6607 | -0.00264 | 0.01143 | -0.231 | 0.8173 | 0.08762 | 0.004487 | 0.9466 | 0 | 2 | 669075 | A |
| 16:13042097 | G | A | 0.6608 | -0.002644 | 0.01143 | -0.2314 | 0.817 | 0.08778 | 0.004574 | 0.9461 | 0 | 2 | 669075 | G |

| rs_number | reference_allele_icd | other_allele_icd | eaf_icd | beta_icd | se_icd | z_icd | p-value_icd | _-log10_p-value_icd | q_statistic_icd | q_p-value_icd | i2_icd | n_studies_icd | n_samples_icd | reference_allele_selfrep |
|---|---|---|---|---|---|---|---|---|---|---|---|---|---|---|
| 16:13043452 | A | G | 0.6606 | -0.002281 | 0.01142 | -0.1997 | 0.8417 | 0.07483 | 0.00225 | 0.9622 | 0 | 2 | 669075 | A |
| 1:110500725 | NA | NA | NA | NA | NA | NA | NA | NA | NA | NA | NA | NA | NA | C |
| 1:110501839 | NA | NA | NA | NA | NA | NA | NA | NA | NA | NA | NA | NA | NA | A |
| 2:98427382 | T | C | 0.4582 | 0.04364 | 0.01068 | 4.087 | 4.42e-05 | 4.355 | 1.409 | 0.2352 | 0.2904 | 2 | 669075 | T |
| 2:98580012 | T | C | 0.4572 | 0.0426 | 0.01068 | 3.989 | 6.69e-05 | 4.174 | 1.437 | 0.2306 | 0.3042 | 2 | 669075 | T |
| 8:10147398 | C | T | 0.4849 | -0.03102 | 0.01063 | -2.918 | 0.003534 | 2.452 | 1.172 | 0.279 | 0.1466 | 2 | 669075 | C |
| 8:10903475 | T | A | 0.5244 | -0.04255 | 0.01074 | -3.96 | 7.57e-05 | 4.121 | 0.01497 | 0.9026 | 0 | 2 | 669075 | T |
| 8:10938260 | G | A | 0.5314 | -0.04062 | 0.01075 | -3.78 | 0.000158 | 3.801 | 0.2275 | 0.6334 | 0 | 2 | 669075 | G |
| 8:10986837 | T | G | 0.5729 | -0.0445 | 0.01077 | -4.131 | 3.65e-05 | 4.437 | 0.001051 | 0.9741 | 0 | 2 | 669075 | T |
| 8:10987199 | T | C | 0.5729 | -0.04446 | 0.01077 | -4.128 | 3.71e-05 | 4.431 | 0.001081 | 0.9738 | 0 | 2 | 669075 | T |
| 8:10987553 | T | C | 0.573 | -0.04439 | 0.01077 | -4.121 | 3.81e-05 | 4.419 | 0.001936 | 0.9649 | 0 | 2 | 669075 | T |
| 8:10987651 | T | G | 0.573 | -0.04438 | 0.01077 | -4.12 | 3.83e-05 | 4.416 | 0.001977 | 0.9645 | 0 | 2 | 669075 | T |
| 8:10988275 | A | G | 0.573 | -0.04426 | 0.01077 | -4.109 | 4.02e-05 | 4.395 | 0.001407 | 0.9701 | 0 | 2 | 669075 | A |
| 8:11039816 | T | G | 0.5398 | -0.03286 | 0.01078 | -3.048 | 0.002316 | 2.635 | 0.02528 | 0.8737 | 0 | 2 | 669075 | T |
| 8:11041642 | A | T | 0.5428 | -0.03312 | 0.01079 | -3.071 | 0.002142 | 2.669 | 0.001076 | 0.9738 | 0 | 2 | 669075 | A |
| 8:11045161 | G | A | 0.5444 | -0.03449 | 0.0108 | -3.195 | 0.001404 | 2.853 | 0.02074 | 0.8855 | 0 | 2 | 669075 | G |
| 8:11046209 | T | C | 0.6028 | -0.03935 | 0.01088 | -3.617 | 0.000301 | 3.522 | 0.2826 | 0.595 | 0 | 2 | 669075 | T |
| 8:11046394 | C | G | 0.6033 | -0.03909 | 0.01089 | -3.591 | 0.000331 | 3.48 | 0.2785 | 0.5977 | 0 | 2 | 669075 | C |
| 8:11054097 | C | A | 0.5596 | -0.03798 | 0.01081 | -3.513 | 0.000446 | 3.351 | 0.02289 | 0.8797 | 0 | 2 | 669075 | C |
| 8:11055597 | C | A | 0.5551 | -0.03713 | 0.01079 | -3.44 | 0.000585 | 3.233 | 0.04556 | 0.831 | 0 | 2 | 669075 | C |
| 8:11060311 | A | C | 0.5499 | -0.03754 | 0.01079 | -3.479 | 0.000507 | 3.295 | 0.01443 | 0.9044 | 0 | 2 | 669075 | A |
| 8:11061792 | C | T | 0.5501 | -0.03736 | 0.01079 | -3.463 | 0.000538 | 3.269 | 0.02091 | 0.885 | 0 | 2 | 669075 | C |
| 8:11062882 | T | C | 0.5492 | -0.03796 | 0.01079 | -3.516 | 0.00044 | 3.356 | 0.01684 | 0.8968 | 0 | 2 | 669075 | T |
| 8:11069960 | C | T | 0.5606 | -0.0383 | 0.01082 | -3.54 | 0.000402 | 3.396 | 3.1e-05 | 0.9956 | 0 | 2 | 669075 | C |
| 8:11070360 | C | G | 0.558 | -0.03841 | 0.01081 | -3.552 | 0.000384 | 3.415 | 0.00074 | 0.9783 | 0 | 2 | 669075 | C |
| 8:11071057 | A | G | 0.5962 | -0.03804 | 0.01085 | -3.507 | 0.000457 | 3.34 | 0.1027 | 0.7486 | 0 | 2 | 669075 | A |
| 8:11076635 | T | G | 0.5456 | -0.03504 | 0.0108 | -3.245 | 0.00118 | 2.928 | 0.00922 | 0.9235 | 0 | 2 | 669075 | T |
| 8:11078781 | A | C | 0.5456 | -0.03558 | 0.0108 | -3.295 | 0.00099 | 3.004 | 0.006672 | 0.9349 | 0 | 2 | 669075 | A |
| 8:11078949 | G | A | 0.5467 | -0.03583 | 0.0108 | -3.318 | 0.000912 | 3.04 | 0.01841 | 0.8921 | 0 | 2 | 669075 | G |
| 8:11079367 | G | A | 0.5467 | -0.03564 | 0.0108 | -3.3 | 0.000973 | 3.012 | 0.006597 | 0.9353 | 0 | 2 | 669075 | G |
| 8:11080665 | T | A | 0.5525 | -0.03691 | 0.01081 | -3.413 | 0.000646 | 3.19 | 0.009109 | 0.924 | 0 | 2 | 669075 | T |
| 8:11080675 | C | T | 0.5525 | -0.03706 | 0.01082 | -3.427 | 0.000615 | 3.211 | 0.01234 | 0.9116 | 0 | 2 | 669075 | C |
| 8:11213589 | T | G | 0.4204 | 0.03333 | 0.01078 | 3.092 | 0.001997 | 2.7 | 0.5117 | 0.4744 | 0 | 2 | 669075 | T |
| 8:11214455 | A | G | 0.4712 | 0.03161 | 0.01064 | 2.971 | 0.002978 | 2.526 | 0.7036 | 0.4016 | 0 | 2 | 669075 | A |
| 8:11214972 | A | G | 0.4707 | 0.03168 | 0.01064 | 2.978 | 0.002912 | 2.536 | 0.6542 | 0.4186 | 0 | 2 | 669075 | A |
| 8:11215617 | C | G | 0.42 | 0.03225 | 0.01078 | 2.991 | 0.002795 | 2.554 | 0.5197 | 0.471 | 0 | 2 | 669075 | C |
| 8:11219334 | G | A | 0.421 | 0.03226 | 0.01078 | 2.993 | 0.00277 | 2.558 | 0.3333 | 0.5637 | 0 | 2 | 669075 | G |
| 8:11219781 | C | T | 0.4211 | 0.03218 | 0.0108 | 2.98 | 0.002893 | 2.539 | 0.3501 | 0.5541 | 0 | 2 | 669075 | C |
| 8:11225168 | G | A | 0.5331 | -0.02419 | 0.01072 | -2.256 | 0.02409 | 1.618 | 1.85 | 0.1737 | 0.4596 | 2 | 669075 | G |
| 8:11225480 | T | C | 0.5315 | -0.02355 | 0.01073 | -2.194 | 0.02823 | 1.549 | 1.746 | 0.1864 | 0.4273 | 2 | 669075 | T |
| 8:11225910 | A | G | 0.5321 | -0.02349 | 0.01073 | -2.189 | 0.02865 | 1.543 | 1.723 | 0.1894 | 0.4195 | 2 | 669075 | A |
| 8:11226071 | A | G | 0.575 | -0.03197 | 0.01077 | -2.968 | 0.003004 | 2.522 | 0.3774 | 0.539 | 0 | 2 | 669075 | A |
| 8:11226456 | C | A | 0.579 | -0.03333 | 0.01078 | -3.092 | 0.001994 | 2.7 | 0.4601 | 0.4976 | 0 | 2 | 669075 | C |
| 8:11227406 | G | A | 0.5777 | -0.03238 | 0.01077 | -3.006 | 0.002657 | 2.576 | 0.3253 | 0.5685 | 0 | 2 | 669075 | G |
| 8:11227885 | C | T | 0.5777 | -0.03234 | 0.01077 | -3.003 | 0.002686 | 2.571 | 0.3338 | 0.5635 | 0 | 2 | 669075 | C |
| 8:11228100 | G | A | 0.5777 | -0.03237 | 0.01077 | -3.005 | 0.002665 | 2.574 | 0.3263 | 0.5678 | 0 | 2 | 669075 | G |
| 8:11228672 | G | A | 0.467 | 0.0238 | 0.01072 | 2.22 | 0.02641 | 1.578 | 2.079 | 0.1494 | 0.5189 | 2 | 669075 | G |

| rs_number | reference_allele_icd | other_allele_icd | eaf_icd | beta_icd | se_icd | z_icd | p-value_icd | _-log10_p-value_icd | q_statistic_icd | q_p-value_icd | i2_icd | n_studies_icd | n_samples_icd | reference_allele_selfrep |
|---|---|---|---|---|---|---|---|---|---|---|---|---|---|---|
| 8:11229319 | C | G | 0.5769 | -0.03014 | 0.01079 | -2.792 | 0.00525 | 2.28 | 0.3099 | 0.5777 | 0 | 2 | 669075 | C |
| 8:11229638 | C | T | 0.5774 | -0.03211 | 0.0108 | -2.974 | 0.002954 | 2.53 | 0.3252 | 0.5685 | 0 | 2 | 669075 | C |
| 8:11230206 | G | T | 0.5775 | -0.03223 | 0.01079 | -2.986 | 0.002841 | 2.547 | 0.2972 | 0.5856 | 0 | 2 | 669075 | G |
| 8:11230259 | A | T | 0.5775 | -0.03222 | 0.01079 | -2.985 | 0.002845 | 2.546 | 0.2967 | 0.5859 | 0 | 2 | 669075 | A |
| 8:11230574 | G | A | 0.5775 | -0.0322 | 0.01079 | -2.983 | 0.002863 | 2.543 | 0.2957 | 0.5866 | 0 | 2 | 669075 | G |
| 8:11231249 | T | C | 0.4286 | 0.03158 | 0.01078 | 2.929 | 0.003416 | 2.466 | 0.3756 | 0.54 | 0 | 2 | 669075 | T |
| 8:11231354 | G | T | 0.4292 | 0.03081 | 0.01078 | 2.858 | 0.004276 | 2.369 | 0.345 | 0.557 | 0 | 2 | 669075 | G |
| 8:11231886 | A | G | 0.5749 | -0.0322 | 0.01077 | -2.989 | 0.002809 | 2.551 | 0.3823 | 0.5364 | 0 | 2 | 669075 | A |
| 8:11232343 | C | G | 0.5772 | -0.03262 | 0.0108 | -3.021 | 0.002531 | 2.597 | 0.2816 | 0.5956 | 0 | 2 | 669075 | C |
| 8:11232788 | G | A | 0.5781 | -0.03274 | 0.01078 | -3.038 | 0.002391 | 2.621 | 0.3623 | 0.5472 | 0 | 2 | 669075 | G |
| 8:11232860 | G | A | 0.5783 | -0.03278 | 0.01078 | -3.041 | 0.002366 | 2.626 | 0.3092 | 0.5782 | 0 | 2 | 669075 | G |
| 8:11233318 | A | C | 0.5727 | -0.03129 | 0.01078 | -2.902 | 0.003721 | 2.429 | 0.207 | 0.6491 | 0 | 2 | 669075 | A |
| 8:11233419 | T | G | 0.5726 | -0.03126 | 0.01078 | -2.899 | 0.003758 | 2.425 | 0.2091 | 0.6475 | 0 | 2 | 669075 | T |
| 8:11233582 | A | G | 0.5726 | -0.03127 | 0.01078 | -2.9 | 0.003745 | 2.426 | 0.2108 | 0.6461 | 0 | 2 | 669075 | A |
| 8:11233659 | A | G | 0.5726 | -0.03124 | 0.01078 | -2.897 | 0.003775 | 2.423 | 0.2087 | 0.6478 | 0 | 2 | 669075 | A |
| 8:11233917 | C | T | 0.5771 | -0.03245 | 0.0108 | -3.005 | 0.002662 | 2.575 | 0.2719 | 0.6021 | 0 | 2 | 669075 | C |
| 8:11233958 | A | G | 0.5771 | -0.03245 | 0.01079 | -3.006 | 0.002653 | 2.576 | 0.2716 | 0.6023 | 0 | 2 | 669075 | A |
| 8:11234298 | C | T | 0.5771 | -0.03243 | 0.01079 | -3.005 | 0.002669 | 2.574 | 0.2716 | 0.6023 | 0 | 2 | 669075 | C |
| 8:11234367 | T | G | 0.5763 | -0.03153 | 0.01079 | -2.922 | 0.003494 | 2.457 | 0.1951 | 0.6587 | 0 | 2 | 669075 | T |
| 8:11234500 | A | T | 0.5726 | -0.03111 | 0.01078 | -2.885 | 0.003927 | 2.406 | 0.2172 | 0.6411 | 0 | 2 | 669075 | A |
| 8:11234520 | A | G | 0.5771 | -0.03243 | 0.01079 | -3.004 | 0.002674 | 2.573 | 0.2711 | 0.6026 | 0 | 2 | 669075 | A |
| 8:11234613 | T | A | 0.5771 | -0.03243 | 0.01079 | -3.004 | 0.002674 | 2.573 | 0.2711 | 0.6026 | 0 | 2 | 669075 | T |
| 8:11234626 | T | C | 0.5771 | -0.03243 | 0.01079 | -3.004 | 0.002674 | 2.573 | 0.2711 | 0.6026 | 0 | 2 | 669075 | T |
| 8:11234780 | C | G | 0.5772 | -0.03246 | 0.01079 | -3.007 | 0.002651 | 2.577 | 0.2682 | 0.6045 | 0 | 2 | 669075 | C |
| 8:11234844 | G | A | 0.5771 | -0.03243 | 0.01079 | -3.004 | 0.002674 | 2.573 | 0.2711 | 0.6026 | 0 | 2 | 669075 | G |
| 8:11234885 | C | G | 0.5771 | -0.03239 | 0.01079 | -3.001 | 0.002704 | 2.568 | 0.2678 | 0.6048 | 0 | 2 | 669075 | C |
| 8:11235136 | C | G | 0.5771 | -0.03238 | 0.01079 | -3 | 0.002708 | 2.567 | 0.2673 | 0.6052 | 0 | 2 | 669075 | C |
| 8:11235150 | C | A | 0.5771 | -0.03242 | 0.01079 | -3.004 | 0.002678 | 2.572 | 0.2706 | 0.6029 | 0 | 2 | 669075 | C |
| 8:11235360 | A | C | 0.5772 | -0.03245 | 0.01079 | -3.006 | 0.002655 | 2.576 | 0.2678 | 0.6048 | 0 | 2 | 669075 | A |
| 8:11235393 | A | G | 0.5771 | -0.03242 | 0.01079 | -3.003 | 0.002682 | 2.572 | 0.2701 | 0.6032 | 0 | 2 | 669075 | A |
| 8:11235497 | G | A | 0.577 | -0.03239 | 0.01079 | -3.001 | 0.002704 | 2.568 | 0.2678 | 0.6048 | 0 | 2 | 669075 | G |
| 8:11235579 | T | G | 0.5764 | -0.03154 | 0.01079 | -2.923 | 0.003475 | 2.459 | 0.1919 | 0.6613 | 0 | 2 | 669075 | T |
| 8:11235605 | T | G | 0.5771 | -0.03242 | 0.01079 | -3.003 | 0.002682 | 2.572 | 0.2701 | 0.6032 | 0 | 2 | 669075 | T |
| 8:11235735 | T | C | 0.5771 | -0.03242 | 0.01079 | -3.003 | 0.002682 | 2.572 | 0.2701 | 0.6032 | 0 | 2 | 669075 | T |
| 8:11235910 | G | A | 0.5791 | -0.03227 | 0.01078 | -2.994 | 0.002767 | 2.558 | 0.3387 | 0.5606 | 0 | 2 | 669075 | G |
| 8:11236392 | T | C | 0.5771 | -0.03242 | 0.01079 | -3.003 | 0.002682 | 2.572 | 0.2701 | 0.6032 | 0 | 2 | 669075 | T |
| 8:11236413 | C | T | 0.5771 | -0.03226 | 0.01079 | -2.988 | 0.002814 | 2.551 | 0.2559 | 0.6129 | 0 | 2 | 669075 | C |
| 8:11236419 | T | C | 0.5771 | -0.03241 | 0.01079 | -3.003 | 0.002686 | 2.571 | 0.2697 | 0.6036 | 0 | 2 | 669075 | T |
| 8:11236572 | T | A | 0.5771 | -0.03241 | 0.01079 | -3.003 | 0.002686 | 2.571 | 0.2697 | 0.6036 | 0 | 2 | 669075 | T |
| 8:11236681 | T | G | 0.5771 | -0.03241 | 0.01079 | -3.003 | 0.002686 | 2.571 | 0.2697 | 0.6036 | 0 | 2 | 669075 | T |
| 8:11236685 | C | T | 0.577 | -0.03243 | 0.01079 | -3.004 | 0.002674 | 2.573 | 0.2711 | 0.6026 | 0 | 2 | 669075 | C |
| 8:11236809 | T | C | 0.5769 | -0.03243 | 0.01079 | -3.005 | 0.002669 | 2.574 | 0.2716 | 0.6023 | 0 | 2 | 669075 | T |
| 8:11236850 | C | T | 0.5771 | -0.03241 | 0.01079 | -3.003 | 0.002686 | 2.571 | 0.2697 | 0.6036 | 0 | 2 | 669075 | C |
| 8:11236964 | C | T | 0.5774 | -0.03262 | 0.0108 | -3.021 | 0.002527 | 2.597 | 0.2567 | 0.6124 | 0 | 2 | 669075 | C |
| 8:11236975 | A | G | 0.5771 | -0.03229 | 0.01079 | -2.991 | 0.00279 | 2.554 | 0.263 | 0.6081 | 0 | 2 | 669075 | A |
| 8:11237477 | A | G | 0.5771 | -0.03242 | 0.0108 | -3.003 | 0.002688 | 2.571 | 0.269 | 0.604 | 0 | 2 | 669075 | A |
| 8:11237480 | A | G | 0.5771 | -0.03242 | 0.0108 | -3.003 | 0.002688 | 2.571 | 0.269 | 0.604 | 0 | 2 | 669075 | A |

| rs_number | reference_allele_icd | other_allele_icd | eaf_icd | beta_icd | se_icd | z_icd | p-value_icd | _-log10_p-value_icd | q_statistic_icd | q_p-value_icd | i2_icd | n_studies_icd | n_samples_icd | reference_allele_selfrep |
|---|---|---|---|---|---|---|---|---|---|---|---|---|---|---|
| 8:11237756 | C | A | 0.5799 | -0.03301 | 0.01078 | -3.061 | 0.002213 | 2.655 | 0.4139 | 0.52 | 0 | 2 | 669075 | C |
| 8:11237773 | T | C | 0.5799 | -0.03301 | 0.01078 | -3.061 | 0.002213 | 2.655 | 0.4139 | 0.52 | 0 | 2 | 669075 | T |
| 8:11238029 | C | T | 0.5804 | -0.03222 | 0.01078 | -2.988 | 0.002822 | 2.549 | 0.517 | 0.4721 | 0 | 2 | 669075 | C |
| 8:11238597 | C | T | 0.5804 | -0.03236 | 0.01078 | -3.001 | 0.002705 | 2.568 | 0.527 | 0.4679 | 0 | 2 | 669075 | C |
| 8:11239017 | T | A | 0.5804 | -0.03244 | 0.01078 | -3.008 | 0.002642 | 2.578 | 0.5304 | 0.4664 | 0 | 2 | 669075 | T |
| 8:11239054 | C | T | 0.5804 | -0.03244 | 0.01078 | -3.008 | 0.002642 | 2.578 | 0.5304 | 0.4664 | 0 | 2 | 669075 | C |
| 8:11239078 | T | G | 0.5804 | -0.03244 | 0.01078 | -3.008 | 0.002639 | 2.579 | 0.5297 | 0.4667 | 0 | 2 | 669075 | T |
| 8:11239137 | G | A | 0.5802 | -0.03307 | 0.01079 | -3.064 | 0.002194 | 2.659 | 0.4421 | 0.5061 | 0 | 2 | 669075 | G |
| 8:11239352 | G | A | 0.5804 | -0.03273 | 0.01078 | -3.035 | 0.002415 | 2.617 | 0.5632 | 0.453 | 0 | 2 | 669075 | G |
| 8:11239510 | G | A | 0.5804 | -0.03268 | 0.01078 | -3.03 | 0.002452 | 2.611 | 0.5529 | 0.4571 | 0 | 2 | 669075 | G |
| 8:11239565 | C | T | 0.5804 | -0.03262 | 0.01078 | -3.025 | 0.0025 | 2.602 | 0.5446 | 0.4605 | 0 | 2 | 669075 | C |
| 8:11239640 | C | T | 0.5804 | -0.03269 | 0.01078 | -3.031 | 0.002448 | 2.611 | 0.5522 | 0.4574 | 0 | 2 | 669075 | C |
| 8:11239762 | A | T | 0.5732 | -0.03056 | 0.01079 | -2.833 | 0.004627 | 2.335 | 0.3093 | 0.5781 | 0 | 2 | 669075 | A |
| 8:11242025 | T | G | 0.5082 | -0.0263 | 0.01579 | -1.666 | 0.09579 | 1.019 | 0 | 1 | NA | 1 | 195394 | T |
| 8:11242632 | G | A | 0.5776 | -0.03217 | 0.01081 | -2.977 | 0.002919 | 2.535 | 0.3002 | 0.5838 | 0 | 2 | 669075 | G |
| 8:11243126 | C | G | 0.5805 | -0.03296 | 0.01079 | -3.054 | 0.002266 | 2.645 | 0.4626 | 0.4964 | 0 | 2 | 669075 | C |
| 8:11245303 | C | G | 0.4267 | 0.03091 | 0.01079 | 2.864 | 0.004202 | 2.377 | 0.2371 | 0.6263 | 0 | 2 | 669075 | C |
| 8:11247814 | G | A | 0.3808 | 0.02347 | 0.01099 | 2.136 | 0.03273 | 1.485 | 1.501 | 0.2205 | 0.3339 | 2 | 669075 | G |
| 8:11249010 | G | T | 0.4262 | 0.03167 | 0.01081 | 2.93 | 0.003403 | 2.468 | 0.9326 | 0.3342 | 0 | 2 | 669075 | G |
| 8:11249261 | T | C | 0.3793 | 0.02386 | 0.01099 | 2.17 | 0.03003 | 1.522 | 1.073 | 0.3003 | 0.06806 | 2 | 669075 | T |
| 8:11251175 | A | G | 0.4155 | 0.03337 | 0.01486 | 2.246 | 0.02475 | 1.606 | 0 | 1 | NA | 1 | 473681 | A |
| 8:11252425 | C | A | 0.4863 | 0.02763 | 0.01104 | 2.502 | 0.01237 | 1.907 | 0.979 | 0.3225 | 0 | 2 | 669075 | C |
| 8:11425809 | T | G | 0.5412 | -0.03078 | 0.01074 | -2.866 | 0.00417 | 2.38 | 0.004785 | 0.9448 | 0 | 2 | 669075 | T |
| 8:8633548 | T | G | 0.5062 | -0.0377 | 0.01068 | -3.53 | 0.000419 | 3.378 | 0.01358 | 0.9072 | 0 | 2 | 669075 | T |
| 8:8637429 | G | A | 0.5069 | -0.03843 | 0.01068 | -3.599 | 0.000322 | 3.492 | 0.02454 | 0.8755 | 0 | 2 | 669075 | G |
| 8:8639740 | A | G | 0.5069 | -0.03829 | 0.01068 | -3.586 | 0.000339 | 3.47 | 0.02116 | 0.8843 | 0 | 2 | 669075 | A |
| 8:8641145 | C | T | 0.4931 | 0.03789 | 0.01068 | 3.547 | 0.000392 | 3.407 | 0.04657 | 0.8291 | 0 | 2 | 669075 | C |
| 8:8644274 | G | C | 0.5087 | 0.03992 | 0.01068 | 3.737 | 0.000188 | 3.726 | 0.1587 | 0.6904 | 0 | 2 | 669075 | G |
| 8:8646246 | T | C | 0.5079 | 0.03983 | 0.01068 | 3.729 | 0.000194 | 3.713 | 0.2269 | 0.6339 | 0 | 2 | 669075 | T |
| 8:8652889 | G | A | 0.5043 | 0.04199 | 0.01067 | 3.936 | 8.38e-05 | 4.077 | 0.3357 | 0.5623 | 0 | 2 | 669075 | G |
| 8:8654527 | T | C | 0.5107 | 0.04027 | 0.01068 | 3.77 | 0.000165 | 3.784 | 0.2674 | 0.6051 | 0 | 2 | 669075 | T |
| 8:8660538 | C | A | 0.5124 | 0.04235 | 0.01069 | 3.96 | 7.56e-05 | 4.122 | 0.1527 | 0.6959 | 0 | 2 | 669075 | C |
| 8:8661534 | T | C | 0.5044 | 0.04141 | 0.01068 | 3.879 | 0.000106 | 3.975 | 0.3364 | 0.5619 | 0 | 2 | 669075 | T |
| 8:8661681 | C | G | 0.5061 | 0.04244 | 0.0107 | 3.968 | 7.31e-05 | 4.136 | 0.2139 | 0.6437 | 0 | 2 | 669075 | C |
| 8:8664622 | G | A | 0.519 | 0.0408 | 0.01069 | 3.818 | 0.000136 | 3.867 | 0.1603 | 0.6889 | 0 | 2 | 669075 | G |
| 8:8673601 | A | C | 0.5138 | 0.0417 | 0.01066 | 3.91 | 9.33e-05 | 4.03 | 0.1698 | 0.6803 | 0 | 2 | 669075 | A |
| 8:8673736 | T | C | 0.5143 | 0.04167 | 0.01067 | 3.905 | 9.52e-05 | 4.021 | 0.1582 | 0.6908 | 0 | 2 | 669075 | T |
| 8:8675325 | A | T | 0.5082 | 0.04279 | 0.01068 | 4.009 | 6.17e-05 | 4.21 | 0.136 | 0.7123 | 0 | 2 | 669075 | A |
| 8:8678530 | G | A | 0.5178 | 0.0414 | 0.01068 | 3.876 | 0.000107 | 3.97 | 0.3938 | 0.5303 | 0 | 2 | 669075 | G |
| 8:8679176 | A | G | 0.5178 | 0.04106 | 0.01068 | 3.844 | 0.000122 | 3.913 | 0.3321 | 0.5644 | 0 | 2 | 669075 | A |
| 8:8679614 | T | C | 0.5177 | 0.04107 | 0.01068 | 3.845 | 0.000122 | 3.915 | 0.3332 | 0.5638 | 0 | 2 | 669075 | T |
| 8:8680477 | G | A | 0.5177 | 0.04103 | 0.01068 | 3.841 | 0.000123 | 3.909 | 0.2836 | 0.5944 | 0 | 2 | 669075 | G |
| 8:8682192 | A | T | 0.5177 | 0.04144 | 0.01069 | 3.879 | 0.000106 | 3.975 | 0.21 | 0.6468 | 0 | 2 | 669075 | A |
| 8:8682878 | T | C | 0.5079 | 0.04239 | 0.01067 | 3.973 | 7.18e-05 | 4.144 | 0.4551 | 0.4999 | 0 | 2 | 669075 | T |
| 8:8684953 | G | A | 0.5183 | 0.04132 | 0.01069 | 3.865 | 0.000112 | 3.951 | 0.3493 | 0.5545 | 0 | 2 | 669075 | G |
| 8:8685190 | A | G | 0.5183 | 0.0414 | 0.01069 | 3.873 | 0.000109 | 3.964 | 0.3576 | 0.5498 | 0 | 2 | 669075 | A |
| 8:8685646 | T | C | 0.5082 | 0.04282 | 0.01068 | 4.01 | 6.14e-05 | 4.212 | 0.5863 | 0.4439 | 0 | 2 | 669075 | T |

| rs_number | reference_allele_icd | other_allele_icd | eaf_icd | beta_icd | se_icd | z_icd | p-value_icd | _-log10_p-value_icd | q_statistic_icd | q_p-value_icd | i2_icd | n_studies_icd | n_samples_icd | reference_allele_selfrep |
|---|---|---|---|---|---|---|---|---|---|---|---|---|---|---|
| 8:8685854 | A | G | 0.5083 | 0.04247 | 0.01068 | 3.977 | 7.06e-05 | 4.151 | 0.587 | 0.4436 | 0 | 2 | 669075 | A |
| 8:8700851 | C | T | 0.5121 | 0.04445 | 0.01069 | 4.16 | 3.22e-05 | 4.492 | 0.3207 | 0.5712 | 0 | 2 | 669075 | C |
| 8:8704330 | G | C | 0.5176 | 0.0415 | 0.01068 | 3.885 | 0.000103 | 3.986 | 0.3584 | 0.5494 | 0 | 2 | 669075 | G |
| 8:8721473 | G | A | 0.5241 | 0.04914 | 0.01071 | 4.589 | 4.53e-06 | 5.344 | 0.2029 | 0.6524 | 0 | 2 | 669075 | G |
| 8:8730488 | G | A | 0.523 | 0.0486 | 0.01071 | 4.538 | 5.77e-06 | 5.239 | 0.07257 | 0.7876 | 0 | 2 | 669075 | G |
| 8:8770512 | C | T | 0.58 | 0.02021 | 0.01087 | 1.858 | 0.06318 | 1.199 | 4.469 | 0.03452 | 0.7762 | 2 | 669075 | C |
| 8:9569104 | T | C | 0.823 | 0.03531 | 0.01417 | 2.492 | 0.01272 | 1.895 | 0.598 | 0.4393 | 0 | 2 | 669075 | T |
| 11:71819437 | NA | NA | NA | NA | NA | NA | NA | NA | NA | NA | NA | NA | NA | T |
| 11:90880166 | NA | NA | NA | NA | NA | NA | NA | NA | NA | NA | NA | NA | NA | T |
| 13:36123276 | NA | NA | NA | NA | NA | NA | NA | NA | NA | NA | NA | NA | NA | T |
| 15:53323213 | NA | NA | NA | NA | NA | NA | NA | NA | NA | NA | NA | NA | NA | G |
| 16:79002432 | NA | NA | NA | NA | NA | NA | NA | NA | NA | NA | NA | NA | NA | A |
| 18:38476471 | NA | NA | NA | NA | NA | NA | NA | NA | NA | NA | NA | NA | NA | T |
| 3:74581060 | NA | NA | NA | NA | NA | NA | NA | NA | NA | NA | NA | NA | NA | G |
| 6:38953162 | NA | NA | NA | NA | NA | NA | NA | NA | NA | NA | NA | NA | NA | C |
| 8:11417582 | C | T | 0.3783 | -0.03741 | 0.01109 | -3.372 | 0.000751 | 3.125 | 2.254 | 0.1333 | 0.5564 | 2 | 669075 | T |
| 9:126164618 | NA | NA | NA | NA | NA | NA | NA | NA | NA | NA | NA | NA | NA | C |
| 5:121781695 | NA | NA | NA | NA | NA | NA | NA | NA | NA | NA | NA | NA | NA | T |
| 1:106531864 | NA | NA | NA | NA | NA | NA | NA | NA | NA | NA | NA | NA | NA | A |

*part 3 of 3 — first column repeated in every part*

| rs_number | other_allele_selfrep | eaf_selfrep | beta_selfrep | se_selfrep | z_selfrep | p-value_selfrep | _-log10_p-value_selfrep | q_statistic_selfrep | q_p-value_selfrep | i2_selfrep | n_studies_selfrep | n_samples_selfrep |
|---|---|---|---|---|---|---|---|---|---|---|---|---|
| 11:92526398 | A | 0.9999 | -0.7084 | 0.1263 | -5.609 | 2.09e-08 | 7.68 | 0 | 1 | NA | 1 | 146929 |
| 11:92529002 | T | 0.9999 | -0.7082 | 0.1263 | -5.607 | 2.11e-08 | 7.676 | 0 | 1 | NA | 1 | 146929 |
| 12:62870574 | G | 0.6333 | -0.007299 | 0.001341 | -5.444 | 5.34e-08 | 7.273 | 30.51 | 0.01021 | 0.5084 | 16 | 237896 |
| 12:62937532 | T | 0.6294 | -0.007438 | 0.001343 | -5.539 | 3.12e-08 | 7.506 | 33.17 | 0.002723 | 0.5779 | 15 | 236585 |
| 12:62941426 | C | 0.6414 | -0.007347 | 0.001347 | -5.455 | 5.01e-08 | 7.3 | 31.03 | 0.00871 | 0.5166 | 16 | 237892 |
| 12:62941837 | G | 0.6414 | -0.007359 | 0.001347 | -5.463 | 4.78e-08 | 7.32 | 31.07 | 0.008589 | 0.5173 | 16 | 237891 |
| 12:62942258 | T | 0.6434 | -0.007497 | 0.001349 | -5.558 | 2.8e-08 | 7.553 | 29.88 | 0.01238 | 0.4979 | 16 | 237898 |
| 12:62945245 | A | 0.6431 | -0.007445 | 0.001351 | -5.51 | 3.68e-08 | 7.434 | 28.55 | 0.01201 | 0.5096 | 15 | 236611 |
| 12:62945970 | G | 0.6433 | -0.007537 | 0.001349 | -5.587 | 2.36e-08 | 7.626 | 29.13 | 0.01547 | 0.4851 | 16 | 237898 |
| 12:62948736 | A | 0.6434 | -0.007527 | 0.00135 | -5.574 | 2.55e-08 | 7.594 | 29.6 | 0.01343 | 0.4933 | 16 | 237898 |
| 12:62949110 | C | 0.6398 | -0.007427 | 0.001345 | -5.521 | 3.44e-08 | 7.463 | 30.15 | 0.01141 | 0.5024 | 16 | 237898 |
| 12:62950556 | C | 0.6432 | -0.00751 | 0.001349 | -5.568 | 2.64e-08 | 7.579 | 29.11 | 0.01558 | 0.4847 | 16 | 237898 |
| 12:62952696 | T | 0.643 | -0.007461 | 0.001349 | -5.529 | 3.3e-08 | 7.482 | 29.06 | 0.0158 | 0.4838 | 16 | 237899 |
| 12:62952910 | T | 0.6431 | -0.007439 | 0.001352 | -5.502 | 3.84e-08 | 7.415 | 28.89 | 0.01081 | 0.5154 | 15 | 236610 |
| 12:62957906 | C | 0.6397 | -0.007442 | 0.001346 | -5.529 | 3.29e-08 | 7.483 | 30.05 | 0.01174 | 0.5008 | 16 | 237897 |
| 12:62960880 | G | 0.6398 | -0.007458 | 0.001345 | -5.545 | 3.01e-08 | 7.521 | 29.68 | 0.01312 | 0.4947 | 16 | 237897 |
| 12:62961299 | T | 0.6435 | -0.007486 | 0.00135 | -5.546 | 2.99e-08 | 7.525 | 28.82 | 0.01695 | 0.4796 | 16 | 237898 |
| 12:62961801 | A | 0.64 | -0.007429 | 0.001346 | -5.52 | 3.47e-08 | 7.46 | 29.92 | 0.0122 | 0.4987 | 16 | 237898 |
| 12:62964552 | C | 0.6432 | -0.007419 | 0.001352 | -5.487 | 4.18e-08 | 7.379 | 27.89 | 0.0147 | 0.4981 | 15 | 236609 |
| 12:62968280 | C | 0.6432 | -0.007522 | 0.001348 | -5.579 | 2.48e-08 | 7.606 | 28.44 | 0.01899 | 0.4725 | 16 | 237897 |
| 12:62970482 | G | 0.6398 | -0.007427 | 0.001348 | -5.509 | 3.7e-08 | 7.432 | 29.38 | 0.009274 | 0.5235 | 15 | 236609 |
| 12:62975307 | T | 0.6432 | -0.007464 | 0.001352 | -5.52 | 3.47e-08 | 7.46 | 28.09 | 0.01383 | 0.5017 | 15 | 236609 |
| 12:62977163 | A | 0.6435 | -0.007546 | 0.001351 | -5.584 | 2.41e-08 | 7.618 | 27.9 | 0.02219 | 0.4624 | 16 | 237889 |
| 12:62980580 | A | 0.6397 | -0.007376 | 0.001346 | -5.481 | 4.33e-08 | 7.364 | 29.38 | 0.01438 | 0.4894 | 16 | 237897 |
| 12:62985871 | A | 0.6432 | -0.00752 | 0.001349 | -5.573 | 2.56e-08 | 7.592 | 28.54 | 0.01845 | 0.4743 | 16 | 237896 |
| 12:62986620 | A | 0.6361 | -0.00825 | 0.001458 | -5.66 | 1.55e-08 | 7.809 | 24.73 | 0.009981 | 0.5552 | 12 | 211880 |
| 12:62988288 | A | 0.6433 | -0.007412 | 0.001352 | -5.48 | 4.34e-08 | 7.362 | 28.52 | 0.01212 | 0.5091 | 15 | 236609 |
| 12:62989110 | C | 0.5209 | -0.007229 | 0.001292 | -5.597 | 2.24e-08 | 7.65 | 36.75 | 0.001376 | 0.5918 | 16 | 237896 |
| 12:62990403 | G | 0.5207 | -0.007216 | 0.001291 | -5.588 | 2.36e-08 | 7.627 | 36.86 | 0.001325 | 0.5931 | 16 | 237896 |
| 12:62990415 | C | 0.5207 | -0.007214 | 0.001291 | -5.586 | 2.38e-08 | 7.623 | 36.89 | 0.001315 | 0.5933 | 16 | 237896 |
| 12:62990871 | A | 0.6432 | -0.007449 | 0.00135 | -5.519 | 3.48e-08 | 7.458 | 28.6 | 0.01808 | 0.4756 | 16 | 237897 |
| 12:62993793 | A | 0.6434 | -0.007536 | 0.001349 | -5.585 | 2.4e-08 | 7.62 | 28.57 | 0.01824 | 0.475 | 16 | 237897 |
| 12:62995269 | G | 0.6431 | -0.007536 | 0.001349 | -5.585 | 2.4e-08 | 7.621 | 27.87 | 0.02242 | 0.4617 | 16 | 237897 |
| 12:62995340 | A | 0.6434 | -0.007556 | 0.00135 | -5.599 | 2.21e-08 | 7.656 | 28.39 | 0.01923 | 0.4717 | 16 | 237896 |
| 12:62995984 | T | 0.6395 | -0.008297 | 0.001463 | -5.672 | 1.45e-08 | 7.839 | 22.77 | 0.01165 | 0.5607 | 11 | 210566 |
| 12:62996061 | A | 0.6433 | -0.007486 | 0.001353 | -5.531 | 3.26e-08 | 7.487 | 28.13 | 0.01369 | 0.5023 | 15 | 236608 |
| 13:83304898 | G | 0.576 | -0.007209 | 0.00131 | -5.505 | 3.79e-08 | 7.422 | 11.95 | 0.6106 | 0 | 15 | 236585 |
| 13:83311141 | A | 0.5757 | -0.007292 | 0.001311 | -5.561 | 2.75e-08 | 7.561 | 11.47 | 0.6485 | 0 | 15 | 236585 |
| 13:83312341 | T | 0.5753 | -0.007318 | 0.001313 | -5.575 | 2.54e-08 | 7.595 | 11.39 | 0.655 | 0 | 15 | 236585 |
| 13:83312717 | C | 0.5751 | -0.007296 | 0.001313 | -5.555 | 2.85e-08 | 7.545 | 11.03 | 0.6839 | 0 | 15 | 236585 |
| 16:13039154 | T | 0.7373 | -0.008726 | 0.001599 | -5.456 | 4.98e-08 | 7.303 | 8.39 | 0.678 | 0 | 12 | 211854 |
| 16:13040514 | T | 0.7357 | -0.008167 | 0.001472 | -5.55 | 2.92e-08 | 7.534 | 7.99 | 0.8442 | 0 | 14 | 235297 |
| 16:13040889 | G | 0.7356 | -0.007998 | 0.001468 | -5.449 | 5.17e-08 | 7.286 | 10.75 | 0.7053 | 0 | 15 | 236585 |
| 16:13041027 | T | 0.7359 | -0.008016 | 0.001468 | -5.46 | 4.88e-08 | 7.311 | 10.73 | 0.7074 | 0 | 15 | 236585 |
| 16:13041924 | G | 0.7357 | -0.007963 | 0.001468 | -5.425 | 5.92e-08 | 7.228 | 10.86 | 0.697 | 0 | 15 | 236585 |
| 16:13042097 | A | 0.7359 | -0.008014 | 0.001468 | -5.458 | 4.93e-08 | 7.307 | 10.78 | 0.7036 | 0 | 15 | 236585 |

| rs_number | other_allele_selfrep | eaf_selfrep | beta_selfrep | se_selfrep | z_selfrep | p-value_selfrep | _-log10_p-value_selfrep | q_statistic_selfrep | q_p-value_selfrep | i2_selfrep | n_studies_selfrep | n_samples_selfrep |
|---|---|---|---|---|---|---|---|---|---|---|---|---|
| 16:13043452 | G | 0.7357 | -0.007972 | 0.001468 | -5.431 | 5.72e-08 | 7.242 | 11.22 | 0.6687 | 0 | 15 | 236585 |
| 1:110500725 | T | 0.9999 | -0.5838 | 0.1016 | -5.746 | 9.37e-09 | 8.028 | 0 | 1 | NA | 1 | 146929 |
| 1:110501839 | G | 0.000156 | 0.4661 | 0.08529 | 5.465 | 4.73e-08 | 7.325 | 0.000136 | 0.9907 | 0 | 2 | 171401 |
| 2:98427382 | C | 0.5917 | 0.007347 | 0.00144 | 5.103 | 3.42e-07 | 6.466 | 11.09 | 0.4361 | 0.00769 | 12 | 211876 |
| 2:98580012 | C | 0.5907 | 0.007446 | 0.001441 | 5.168 | 2.42e-07 | 6.617 | 10.81 | 0.5452 | 0 | 13 | 213167 |
| 8:10147398 | T | 0.4568 | -0.007504 | 0.001303 | -5.759 | 8.67e-09 | 8.062 | 15.9 | 0.3196 | 0.1195 | 15 | 236585 |
| 8:10903475 | A | 0.4852 | -0.006916 | 0.0013 | -5.319 | 1.07e-07 | 6.973 | 24.03 | 0.03083 | 0.4591 | 14 | 235297 |
| 8:10938260 | A | 0.4873 | -0.0067 | 0.001298 | -5.16 | 2.51e-07 | 6.6 | 23.71 | 0.03393 | 0.4516 | 14 | 235297 |
| 8:10986837 | G | 0.5723 | -0.006539 | 0.001308 | -5 | 5.83e-07 | 6.235 | 19.22 | 0.1164 | 0.3237 | 14 | 235297 |
| 8:10987199 | C | 0.5728 | -0.006618 | 0.001306 | -5.066 | 4.13e-07 | 6.384 | 18.51 | 0.1846 | 0.2435 | 15 | 236585 |
| 8:10987553 | C | 0.5731 | -0.006705 | 0.001308 | -5.128 | 2.99e-07 | 6.524 | 18.35 | 0.1448 | 0.2915 | 14 | 235297 |
| 8:10987651 | G | 0.5728 | -0.006661 | 0.001308 | -5.095 | 3.56e-07 | 6.448 | 18.77 | 0.1304 | 0.3074 | 14 | 235297 |
| 8:10988275 | G | 0.5751 | -0.007554 | 0.00142 | -5.321 | 1.05e-07 | 6.977 | 14.71 | 0.1431 | 0.3201 | 11 | 210566 |
| 8:11039816 | G | 0.4926 | -0.006684 | 0.001297 | -5.152 | 2.62e-07 | 6.581 | 23.46 | 0.03649 | 0.4458 | 14 | 235297 |
| 8:11041642 | T | 0.4938 | -0.006694 | 0.001295 | -5.167 | 2.43e-07 | 6.615 | 24.68 | 0.0379 | 0.4327 | 15 | 236585 |
| 8:11045161 | A | 0.4946 | -0.006686 | 0.001294 | -5.167 | 2.43e-07 | 6.615 | 26.44 | 0.03365 | 0.4327 | 16 | 237899 |
| 8:11046209 | C | 0.5893 | -0.006689 | 0.001313 | -5.094 | 3.58e-07 | 6.446 | 22.06 | 0.07744 | 0.3653 | 15 | 236585 |
| 8:11046394 | G | 0.5915 | -0.007651 | 0.001425 | -5.37 | 8.05e-08 | 7.094 | 14.84 | 0.1897 | 0.259 | 12 | 211854 |
| 8:11054097 | A | 0.519 | -0.006643 | 0.001297 | -5.12 | 3.11e-07 | 6.507 | 21.89 | 0.1109 | 0.3146 | 16 | 237887 |
| 8:11055597 | A | 0.5169 | -0.006752 | 0.001297 | -5.207 | 1.96e-07 | 6.708 | 20.7 | 0.1096 | 0.3236 | 15 | 236585 |
| 8:11060311 | C | 0.5028 | -0.006989 | 0.001296 | -5.391 | 7.15e-08 | 7.146 | 25.74 | 0.0408 | 0.4173 | 16 | 237885 |
| 8:11061792 | T | 0.5028 | -0.006992 | 0.001296 | -5.394 | 7.04e-08 | 7.153 | 26.49 | 0.03315 | 0.4338 | 16 | 237888 |
| 8:11062882 | C | 0.5016 | -0.007106 | 0.001297 | -5.48 | 4.36e-08 | 7.36 | 26.03 | 0.03767 | 0.4238 | 16 | 237884 |
| 8:11069960 | T | 0.5216 | -0.006769 | 0.001298 | -5.215 | 1.88e-07 | 6.727 | 26.46 | 0.02259 | 0.4709 | 15 | 236611 |
| 8:11070360 | G | 0.5135 | -0.006637 | 0.001296 | -5.123 | 3.07e-07 | 6.513 | 27.82 | 0.02275 | 0.4607 | 16 | 237883 |
| 8:11071057 | G | 0.5753 | -0.007046 | 0.001311 | -5.374 | 7.87e-08 | 7.104 | 26.38 | 0.01512 | 0.5072 | 14 | 235297 |
| 8:11076635 | G | 0.4923 | -0.006897 | 0.001299 | -5.31 | 1.12e-07 | 6.951 | 26.19 | 0.01604 | 0.5036 | 14 | 235297 |
| 8:11078781 | C | 0.4926 | -0.006886 | 0.001299 | -5.301 | 1.18e-07 | 6.929 | 27.03 | 0.01907 | 0.4821 | 15 | 236585 |
| 8:11078949 | A | 0.4953 | -0.006918 | 0.001299 | -5.325 | 1.03e-07 | 6.987 | 27.24 | 0.01793 | 0.486 | 15 | 236585 |
| 8:11079367 | A | 0.4949 | -0.006985 | 0.001299 | -5.377 | 7.74e-08 | 7.111 | 26.55 | 0.02199 | 0.4728 | 15 | 236585 |
| 8:11080665 | A | 0.5056 | -0.007109 | 0.001303 | -5.454 | 5.04e-08 | 7.298 | 24.01 | 0.04568 | 0.4169 | 15 | 236585 |
| 8:11080675 | T | 0.5059 | -0.007153 | 0.001303 | -5.488 | 4.16e-08 | 7.381 | 23.81 | 0.04836 | 0.4119 | 15 | 236585 |
| 8:11213589 | G | 0.459 | 0.00673 | 0.001299 | 5.18 | 2.27e-07 | 6.644 | 21.5 | 0.06367 | 0.3953 | 14 | 235297 |
| 8:11214455 | G | 0.4671 | 0.006783 | 0.001296 | 5.234 | 1.69e-07 | 6.771 | 18.67 | 0.178 | 0.2501 | 15 | 236585 |
| 8:11214972 | G | 0.4673 | 0.006785 | 0.001296 | 5.236 | 1.67e-07 | 6.777 | 19.66 | 0.1413 | 0.2879 | 15 | 236585 |
| 8:11215617 | G | 0.4596 | 0.006823 | 0.001297 | 5.26 | 1.48e-07 | 6.831 | 21.64 | 0.08622 | 0.3532 | 15 | 236585 |
| 8:11219334 | A | 0.4597 | 0.006814 | 0.0013 | 5.24 | 1.64e-07 | 6.786 | 22.62 | 0.04649 | 0.4252 | 14 | 235297 |
| 8:11219781 | T | 0.4602 | 0.006891 | 0.0013 | 5.302 | 1.17e-07 | 6.933 | 22.47 | 0.06946 | 0.3769 | 15 | 236585 |
| 8:11225168 | A | 0.4931 | -0.006896 | 0.001296 | -5.321 | 1.05e-07 | 6.977 | 19.75 | 0.1381 | 0.2912 | 15 | 236585 |
| 8:11225480 | C | 0.4928 | -0.006875 | 0.001296 | -5.305 | 1.15e-07 | 6.939 | 19.95 | 0.1318 | 0.2981 | 15 | 236585 |
| 8:11225910 | G | 0.493 | -0.006912 | 0.001298 | -5.325 | 1.03e-07 | 6.986 | 20.08 | 0.09323 | 0.3526 | 14 | 235297 |
| 8:11226071 | G | 0.5423 | -0.00738 | 0.0013 | -5.677 | 1.4e-08 | 7.852 | 22.7 | 0.06526 | 0.3834 | 15 | 236585 |
| 8:11226456 | A | 0.5547 | -0.00751 | 0.001303 | -5.765 | 8.41e-09 | 8.075 | 21.59 | 0.08737 | 0.3516 | 15 | 236585 |
| 8:11227406 | A | 0.5518 | -0.007229 | 0.001301 | -5.555 | 2.84e-08 | 7.546 | 22.6 | 0.06706 | 0.3806 | 15 | 236585 |
| 8:11227885 | T | 0.5534 | -0.007231 | 0.001301 | -5.557 | 2.82e-08 | 7.55 | 22.72 | 0.06504 | 0.3837 | 15 | 236585 |
| 8:11228100 | A | 0.5519 | -0.007217 | 0.001301 | -5.549 | 2.95e-08 | 7.53 | 22.58 | 0.06749 | 0.3799 | 15 | 236585 |
| 8:11228672 | A | 0.5084 | 0.006843 | 0.001294 | 5.286 | 1.28e-07 | 6.894 | 20.23 | 0.123 | 0.308 | 15 | 236585 |

| rs_number | other_allele_selfrep | eaf_selfrep | beta_selfrep | se_selfrep | z_selfrep | p-value_selfrep | _-log10_p-value_selfrep | q_statistic_selfrep | q_p-value_selfrep | i2_selfrep | n_studies_selfrep | n_samples_selfrep |
|---|---|---|---|---|---|---|---|---|---|---|---|---|
| 8:11229319 | G | 0.5558 | -0.007226 | 0.001303 | -5.546 | 2.99e-08 | 7.524 | 24.16 | 0.04382 | 0.4205 | 15 | 236585 |
| 8:11229638 | T | 0.5512 | -0.00712 | 0.001302 | -5.468 | 4.67e-08 | 7.331 | 22.56 | 0.04732 | 0.4237 | 14 | 235297 |
| 8:11230206 | T | 0.5532 | -0.007207 | 0.001301 | -5.539 | 3.12e-08 | 7.506 | 22.29 | 0.07292 | 0.3718 | 15 | 236585 |
| 8:11230259 | T | 0.5531 | -0.007197 | 0.001301 | -5.53 | 3.28e-08 | 7.484 | 22.39 | 0.07089 | 0.3748 | 15 | 236585 |
| 8:11230574 | A | 0.5535 | -0.007271 | 0.001301 | -5.587 | 2.36e-08 | 7.627 | 22.35 | 0.0717 | 0.3736 | 15 | 236585 |
| 8:11231249 | C | 0.4669 | 0.007293 | 0.001296 | 5.626 | 1.89e-08 | 7.722 | 22.32 | 0.07234 | 0.3727 | 15 | 236585 |
| 8:11231354 | T | 0.4675 | 0.007167 | 0.001297 | 5.525 | 3.36e-08 | 7.473 | 22.82 | 0.06326 | 0.3865 | 15 | 236585 |
| 8:11231886 | G | 0.5398 | -0.007237 | 0.001299 | -5.573 | 2.57e-08 | 7.591 | 23.1 | 0.05859 | 0.3941 | 15 | 236585 |
| 8:11232343 | G | 0.552 | -0.007198 | 0.001301 | -5.533 | 3.22e-08 | 7.492 | 24.04 | 0.04535 | 0.4176 | 15 | 236585 |
| 8:11232788 | A | 0.5534 | -0.007334 | 0.001302 | -5.635 | 1.8e-08 | 7.746 | 22.89 | 0.06212 | 0.3883 | 15 | 236585 |
| 8:11232860 | A | 0.5521 | -0.007223 | 0.001301 | -5.552 | 2.89e-08 | 7.539 | 24.01 | 0.04571 | 0.4169 | 15 | 236585 |
| 8:11233318 | C | 0.5468 | -0.007295 | 0.001298 | -5.619 | 1.97e-08 | 7.705 | 21.72 | 0.08454 | 0.3555 | 15 | 236585 |
| 8:11233419 | G | 0.5468 | -0.007264 | 0.001298 | -5.594 | 2.27e-08 | 7.644 | 21.7 | 0.08507 | 0.3547 | 15 | 236585 |
| 8:11233582 | G | 0.5469 | -0.007225 | 0.001298 | -5.565 | 2.69e-08 | 7.57 | 21.69 | 0.0853 | 0.3544 | 15 | 236585 |
| 8:11233659 | G | 0.5469 | -0.007225 | 0.001298 | -5.564 | 2.69e-08 | 7.57 | 21.68 | 0.08545 | 0.3542 | 15 | 236585 |
| 8:11233917 | T | 0.5527 | -0.007203 | 0.001301 | -5.538 | 3.13e-08 | 7.505 | 23.21 | 0.05686 | 0.3969 | 15 | 236585 |
| 8:11233958 | G | 0.5527 | -0.007208 | 0.001301 | -5.542 | 3.06e-08 | 7.514 | 23.2 | 0.05714 | 0.3965 | 15 | 236585 |
| 8:11234298 | T | 0.5524 | -0.007212 | 0.0013 | -5.548 | 2.96e-08 | 7.529 | 22.47 | 0.06938 | 0.3771 | 15 | 236585 |
| 8:11234367 | G | 0.5523 | -0.007209 | 0.0013 | -5.546 | 2.99e-08 | 7.524 | 22.78 | 0.06398 | 0.3854 | 15 | 236585 |
| 8:11234500 | T | 0.5458 | -0.007223 | 0.001298 | -5.565 | 2.68e-08 | 7.572 | 22.77 | 0.06408 | 0.3852 | 15 | 236585 |
| 8:11234520 | G | 0.5522 | -0.007217 | 0.0013 | -5.552 | 2.89e-08 | 7.539 | 22.85 | 0.06267 | 0.3874 | 15 | 236585 |
| 8:11234613 | A | 0.5528 | -0.00719 | 0.001301 | -5.528 | 3.32e-08 | 7.479 | 22.83 | 0.06309 | 0.3868 | 15 | 236585 |
| 8:11234626 | C | 0.5528 | -0.007197 | 0.001301 | -5.533 | 3.22e-08 | 7.492 | 22.82 | 0.06331 | 0.3864 | 15 | 236585 |
| 8:11234780 | G | 0.5521 | -0.0072 | 0.0013 | -5.539 | 3.12e-08 | 7.506 | 22.66 | 0.066 | 0.3822 | 15 | 236585 |
| 8:11234844 | A | 0.5522 | -0.007207 | 0.0013 | -5.544 | 3.02e-08 | 7.52 | 22.63 | 0.06649 | 0.3815 | 15 | 236585 |
| 8:11234885 | G | 0.5521 | -0.007212 | 0.001299 | -5.551 | 2.91e-08 | 7.536 | 22.73 | 0.06479 | 0.3841 | 15 | 236585 |
| 8:11235136 | G | 0.5525 | -0.007178 | 0.0013 | -5.522 | 3.44e-08 | 7.464 | 22.96 | 0.06097 | 0.3902 | 15 | 236585 |
| 8:11235150 | A | 0.5545 | -0.008136 | 0.001409 | -5.774 | 7.95e-09 | 8.1 | 15.99 | 0.1416 | 0.312 | 12 | 211854 |
| 8:11235360 | C | 0.552 | -0.007192 | 0.001299 | -5.535 | 3.19e-08 | 7.496 | 23.02 | 0.05992 | 0.3919 | 15 | 236585 |
| 8:11235393 | G | 0.552 | -0.007153 | 0.001299 | -5.506 | 3.76e-08 | 7.425 | 22.98 | 0.06063 | 0.3907 | 15 | 236585 |
| 8:11235497 | A | 0.5521 | -0.007184 | 0.001298 | -5.533 | 3.23e-08 | 7.491 | 23.02 | 0.08368 | 0.3484 | 16 | 237880 |
| 8:11235579 | G | 0.5524 | -0.007158 | 0.0013 | -5.507 | 3.73e-08 | 7.428 | 23.03 | 0.08355 | 0.3486 | 16 | 237881 |
| 8:11235605 | G | 0.5524 | -0.007161 | 0.0013 | -5.509 | 3.69e-08 | 7.432 | 23 | 0.08407 | 0.3479 | 16 | 237882 |
| 8:11235735 | C | 0.5519 | -0.007181 | 0.001299 | -5.527 | 3.33e-08 | 7.478 | 22.87 | 0.06241 | 0.3879 | 15 | 236585 |
| 8:11235910 | A | 0.561 | -0.007095 | 0.001302 | -5.451 | 5.12e-08 | 7.291 | 24.26 | 0.04263 | 0.4229 | 15 | 236585 |
| 8:11236392 | C | 0.5543 | -0.00724 | 0.001301 | -5.564 | 2.7e-08 | 7.568 | 23.36 | 0.07686 | 0.3578 | 16 | 237888 |
| 8:11236413 | T | 0.5541 | -0.007271 | 0.001301 | -5.587 | 2.36e-08 | 7.627 | 23.76 | 0.06936 | 0.3686 | 16 | 237888 |
| 8:11236419 | C | 0.5541 | -0.007273 | 0.001301 | -5.589 | 2.34e-08 | 7.631 | 23.58 | 0.07261 | 0.3638 | 16 | 237888 |
| 8:11236572 | A | 0.5516 | -0.007252 | 0.001298 | -5.587 | 2.37e-08 | 7.626 | 23.04 | 0.08338 | 0.3488 | 16 | 237889 |
| 8:11236681 | G | 0.5522 | -0.007264 | 0.001299 | -5.594 | 2.28e-08 | 7.643 | 22.8 | 0.08853 | 0.342 | 16 | 237889 |
| 8:11236685 | T | 0.5522 | -0.007263 | 0.001299 | -5.593 | 2.28e-08 | 7.641 | 22.81 | 0.08824 | 0.3424 | 16 | 237889 |
| 8:11236809 | C | 0.5518 | -0.007153 | 0.001297 | -5.514 | 3.59e-08 | 7.444 | 22.9 | 0.08638 | 0.3448 | 16 | 237888 |
| 8:11236850 | T | 0.552 | -0.007173 | 0.001297 | -5.529 | 3.3e-08 | 7.482 | 22.77 | 0.08904 | 0.3414 | 16 | 237889 |
| 8:11236964 | T | 0.5519 | -0.00717 | 0.001297 | -5.527 | 3.34e-08 | 7.476 | 22.58 | 0.09339 | 0.3358 | 16 | 237885 |
| 8:11236975 | G | 0.5519 | -0.007198 | 0.001299 | -5.542 | 3.06e-08 | 7.514 | 22.55 | 0.06807 | 0.379 | 15 | 236611 |
| 8:11237477 | G | 0.5529 | -0.007247 | 0.001299 | -5.58 | 2.46e-08 | 7.609 | 22.22 | 0.1021 | 0.3251 | 16 | 237889 |
| 8:11237480 | G | 0.5529 | -0.007247 | 0.001299 | -5.581 | 2.45e-08 | 7.61 | 22.23 | 0.1018 | 0.3254 | 16 | 237889 |

| rs_number | other_allele_selfrep | eaf_selfrep | beta_selfrep | se_selfrep | z_selfrep | p-value_selfrep | _-log10_p-value_selfrep | q_statistic_selfrep | q_p-value_selfrep | i2_selfrep | n_studies_selfrep | n_samples_selfrep |
|---|---|---|---|---|---|---|---|---|---|---|---|---|
| 8:11237756 | A | 0.5588 | -0.007035 | 0.001295 | -5.434 | 5.64e-08 | 7.249 | 24.82 | 0.03643 | 0.4358 | 15 | 236585 |
| 8:11237773 | C | 0.5597 | -0.00707 | 0.001299 | -5.442 | 5.39e-08 | 7.269 | 24.72 | 0.03742 | 0.4337 | 15 | 236585 |
| 8:11238029 | T | 0.5614 | -0.007177 | 0.001301 | -5.518 | 3.51e-08 | 7.455 | 24.67 | 0.03795 | 0.4325 | 15 | 236585 |
| 8:11238597 | T | 0.5598 | -0.007095 | 0.0013 | -5.456 | 4.98e-08 | 7.303 | 24.6 | 0.03877 | 0.4308 | 15 | 236585 |
| 8:11239017 | A | 0.5601 | -0.007075 | 0.001301 | -5.438 | 5.52e-08 | 7.258 | 24.68 | 0.03788 | 0.4327 | 15 | 236585 |
| 8:11239054 | T | 0.5602 | -0.007068 | 0.001302 | -5.43 | 5.76e-08 | 7.24 | 24.83 | 0.03625 | 0.4362 | 15 | 236585 |
| 8:11239078 | G | 0.56 | -0.007084 | 0.001301 | -5.445 | 5.3e-08 | 7.276 | 24.95 | 0.03509 | 0.4388 | 15 | 236585 |
| 8:11239137 | A | 0.5599 | -0.007091 | 0.001301 | -5.45 | 5.15e-08 | 7.288 | 24.68 | 0.03786 | 0.4327 | 15 | 236585 |
| 8:11239352 | A | 0.5599 | -0.007111 | 0.001301 | -5.466 | 4.72e-08 | 7.326 | 24.77 | 0.03688 | 0.4349 | 15 | 236585 |
| 8:11239510 | A | 0.56 | -0.007122 | 0.001302 | -5.471 | 4.57e-08 | 7.34 | 24.76 | 0.03702 | 0.4346 | 15 | 236585 |
| 8:11239565 | T | 0.56 | -0.007121 | 0.001302 | -5.471 | 4.59e-08 | 7.338 | 24.75 | 0.03712 | 0.4343 | 15 | 236585 |
| 8:11239640 | T | 0.5607 | -0.007171 | 0.001302 | -5.506 | 3.75e-08 | 7.425 | 24.59 | 0.03889 | 0.4306 | 15 | 236585 |
| 8:11239762 | T | 0.5455 | -0.007242 | 0.001298 | -5.58 | 2.46e-08 | 7.609 | 22.1 | 0.07668 | 0.3664 | 15 | 236585 |
| 8:11242025 | G | 0.5614 | -0.007172 | 0.001304 | -5.499 | 3.91e-08 | 7.408 | 24.79 | 0.03674 | 0.4352 | 15 | 236585 |
| 8:11242632 | A | 0.552 | -0.007297 | 0.001302 | -5.606 | 2.12e-08 | 7.674 | 24.31 | 0.042 | 0.4241 | 15 | 236585 |
| 8:11243126 | G | 0.5613 | -0.007184 | 0.001304 | -5.51 | 3.68e-08 | 7.435 | 25.14 | 0.0332 | 0.4432 | 15 | 236585 |
| 8:11245303 | G | 0.4541 | 0.007286 | 0.0013 | 5.603 | 2.15e-08 | 7.667 | 22.4 | 0.07074 | 0.375 | 15 | 236585 |
| 8:11247814 | A | 0.3721 | 0.007346 | 0.001341 | 5.476 | 4.44e-08 | 7.352 | 21.66 | 0.08598 | 0.3535 | 15 | 236585 |
| 8:11249010 | T | 0.4552 | 0.007253 | 0.001301 | 5.573 | 2.56e-08 | 7.591 | 21.72 | 0.08461 | 0.3554 | 15 | 236585 |
| 8:11249261 | C | 0.3646 | 0.008201 | 0.00146 | 5.618 | 1.97e-08 | 7.705 | 13.62 | 0.2548 | 0.1923 | 12 | 211854 |
| 8:11251175 | G | 0.493 | 0.007194 | 0.001323 | 5.439 | 5.49e-08 | 7.261 | 19.68 | 0.1036 | 0.3393 | 14 | 235297 |
| 8:11252425 | A | 0.561 | 0.00714 | 0.001371 | 5.206 | 1.97e-07 | 6.707 | 23.5 | 0.05262 | 0.4042 | 15 | 236585 |
| 8:11425809 | G | 0.5232 | -0.006717 | 0.001298 | -5.176 | 2.32e-07 | 6.635 | 30.69 | 0.009677 | 0.5112 | 16 | 237876 |
| 8:8633548 | G | 0.4783 | -0.006858 | 0.001302 | -5.269 | 1.4e-07 | 6.853 | 23.45 | 0.05335 | 0.4029 | 15 | 236585 |
| 8:8637429 | A | 0.4795 | -0.006752 | 0.001299 | -5.197 | 2.07e-07 | 6.684 | 21.92 | 0.08018 | 0.3614 | 15 | 236585 |
| 8:8639740 | G | 0.4797 | -0.006692 | 0.0013 | -5.149 | 2.67e-07 | 6.573 | 22.91 | 0.06182 | 0.3888 | 15 | 236585 |
| 8:8641145 | T | 0.5207 | 0.00678 | 0.001301 | 5.213 | 1.89e-07 | 6.723 | 19.76 | 0.1013 | 0.3422 | 14 | 235297 |
| 8:8644274 | C | 0.5239 | 0.006573 | 0.001302 | 5.048 | 4.56e-07 | 6.341 | 20.8 | 0.107 | 0.3268 | 15 | 236585 |
| 8:8646246 | C | 0.5218 | 0.006929 | 0.0013 | 5.329 | 1.01e-07 | 6.996 | 19.32 | 0.1135 | 0.3271 | 14 | 235297 |
| 8:8652889 | A | 0.5205 | 0.00675 | 0.001297 | 5.206 | 1.97e-07 | 6.705 | 22.3 | 0.1002 | 0.3273 | 16 | 237878 |
| 8:8654527 | C | 0.5306 | 0.00653 | 0.001298 | 5.029 | 5.03e-07 | 6.299 | 17.88 | 0.2122 | 0.2171 | 15 | 236585 |
| 8:8660538 | A | 0.534 | 0.00661 | 0.001298 | 5.091 | 3.62e-07 | 6.441 | 19.44 | 0.1489 | 0.2797 | 15 | 236585 |
| 8:8661534 | C | 0.5205 | 0.006753 | 0.001295 | 5.214 | 1.88e-07 | 6.725 | 22.64 | 0.09208 | 0.3375 | 16 | 237886 |
| 8:8661681 | G | 0.5308 | 0.006715 | 0.001298 | 5.174 | 2.34e-07 | 6.632 | 19.04 | 0.1635 | 0.2646 | 15 | 236585 |
| 8:8664622 | A | 0.5387 | 0.006806 | 0.001301 | 5.23 | 1.73e-07 | 6.762 | 13.9 | 0.3806 | 0.06508 | 14 | 235297 |
| 8:8673601 | C | 0.5275 | 0.006657 | 0.001299 | 5.125 | 3.03e-07 | 6.518 | 17.64 | 0.2235 | 0.2065 | 15 | 236611 |
| 8:8673736 | C | 0.5295 | 0.006626 | 0.001298 | 5.103 | 3.4e-07 | 6.468 | 19.91 | 0.1752 | 0.2468 | 16 | 237888 |
| 8:8675325 | T | 0.5265 | 0.006621 | 0.001299 | 5.095 | 3.55e-07 | 6.45 | 20.08 | 0.1275 | 0.3029 | 15 | 236585 |
| 8:8678530 | A | 0.5329 | 0.006702 | 0.0013 | 5.156 | 2.57e-07 | 6.589 | 19.13 | 0.2077 | 0.2161 | 16 | 237881 |
| 8:8679176 | G | 0.5325 | 0.006616 | 0.0013 | 5.091 | 3.64e-07 | 6.439 | 19.13 | 0.2078 | 0.2159 | 16 | 237882 |
| 8:8679614 | C | 0.5325 | 0.006659 | 0.0013 | 5.123 | 3.06e-07 | 6.514 | 18.58 | 0.2332 | 0.1929 | 16 | 237879 |
| 8:8680477 | A | 0.5326 | 0.006606 | 0.0013 | 5.083 | 3.79e-07 | 6.422 | 18.37 | 0.2438 | 0.1833 | 16 | 237878 |
| 8:8682192 | T | 0.5327 | 0.006653 | 0.0013 | 5.116 | 3.18e-07 | 6.498 | 18.49 | 0.2379 | 0.1887 | 16 | 237878 |
| 8:8682878 | C | 0.5205 | 0.006579 | 0.0013 | 5.061 | 4.24e-07 | 6.372 | 23.07 | 0.05919 | 0.3931 | 15 | 236585 |
| 8:8684953 | A | 0.5292 | 0.00666 | 0.001303 | 5.112 | 3.26e-07 | 6.487 | 17.09 | 0.2512 | 0.181 | 15 | 236585 |
| 8:8685190 | G | 0.5282 | 0.006551 | 0.001302 | 5.03 | 4.99e-07 | 6.302 | 16.86 | 0.2639 | 0.1694 | 15 | 236585 |
| 8:8685646 | C | 0.5162 | 0.006651 | 0.001301 | 5.112 | 3.26e-07 | 6.487 | 21.3 | 0.09431 | 0.3426 | 15 | 236585 |

| rs_number | other_allele_selfrep | eaf_selfrep | beta_selfrep | se_selfrep | z_selfrep | p-value_selfrep | _-log10_p-value_selfrep | q_statistic_selfrep | q_p-value_selfrep | i2_selfrep | n_studies_selfrep | n_samples_selfrep |
|---|---|---|---|---|---|---|---|---|---|---|---|---|
| 8:8685854 | G | 0.5171 | 0.00674 | 0.001302 | 5.178 | 2.29e-07 | 6.64 | 17.66 | 0.171 | 0.2637 | 14 | 235297 |
| 8:8700851 | T | 0.5204 | 0.006886 | 0.001298 | 5.303 | 1.16e-07 | 6.935 | 22.17 | 0.07518 | 0.3685 | 15 | 236585 |
| 8:8704330 | C | 0.5235 | 0.006644 | 0.001298 | 5.117 | 3.16e-07 | 6.501 | 23.65 | 0.05054 | 0.4079 | 15 | 236585 |
| 8:8721473 | A | 0.548 | 0.006641 | 0.001305 | 5.09 | 3.64e-07 | 6.439 | 19.7 | 0.14 | 0.2893 | 15 | 236585 |
| 8:8730488 | A | 0.5383 | 0.006568 | 0.001307 | 5.024 | 5.16e-07 | 6.287 | 20.17 | 0.1248 | 0.3059 | 15 | 236585 |
| 8:8770512 | T | 0.5648 | 0.007034 | 0.001312 | 5.363 | 8.37e-08 | 7.077 | 17.12 | 0.2496 | 0.1825 | 15 | 236585 |
| 8:9569104 | C | 0.8127 | 0.008745 | 0.001669 | 5.24 | 1.64e-07 | 6.784 | 8.336 | 0.9096 | 0 | 16 | 237881 |
| 11:71819437 | C | 0.9998 | -0.4691 | 0.0858 | -5.467 | 4.67e-08 | 7.33 | 0 | 1 | NA | 1 | 146929 |
| 11:90880166 | C | 0.9999 | -0.7792 | 0.1359 | -5.734 | 1.01e-08 | 7.996 | 0 | 1 | NA | 1 | 146929 |
| 13:36123276 | A | 0.9999 | -0.5076 | 0.09064 | -5.6 | 2.19e-08 | 7.659 | 0 | 1 | NA | 1 | 146929 |
| 15:53323213 | A | 0.9996 | -0.229 | 0.04036 | -5.672 | 1.45e-08 | 7.84 | 8.046 | 0.004561 | 0.8757 | 2 | 148709 |
| 16:79002432 | G | 0.9998 | -0.429 | 0.07089 | -6.052 | 1.48e-09 | 8.831 | 0 | 1 | 0 | 1 | 146929 |
| 18:38476471 | C | 0.9998 | -0.324 | 0.05653 | -5.731 | 1.02e-08 | 7.991 | 0 | 1 | NA | 1 | 146929 |
| 3:74581060 | A | 0.9998 | -0.3863 | 0.06473 | -5.968 | 2.47e-09 | 8.607 | 0 | 1 | NA | 1 | 146929 |
| 6:38953162 | A | 0.9997 | -0.2589 | 0.04682 | -5.53 | 3.29e-08 | 7.483 | 0 | 1 | NA | 1 | 146929 |
| 8:11417582 | C | 0.6396 | 0.009539 | 0.001571 | 6.074 | 1.29e-09 | 8.89 | 1.468 | 0.6897 | 0 | 4 | 152547 |
| 9:126164618 | T | 0.9999 | -0.494 | 0.08773 | -5.631 | 1.84e-08 | 7.736 | 0 | 1 | NA | 1 | 146929 |
| 5:121781695 | A | 0.01 | 0.5238 | 0.08928 | 5.867 | 4.56e-09 | 8.341 | 0 | 1 | NA | 1 | 1196 |
| 1:106531864 | G | 0.001422 | 0.3694 | 0.06708 | 5.507 | 3.74e-08 | 7.427 | 0 | 1 | NA | 1 | 8343 |

#### Supplementary Table S8

VEGAS2 gene mapping

| Chr | Gene | nSNPs | nSims | Start | Stop | Test | Pvalue | Best.SNP | SNP.pvalue |
|---|---|---|---|---|---|---|---|---|---|
| 2 | TMEM131 | 717 | 1000000 | 98372800 | 98612354 | 4425 | 2e-06 | rs35098301 | 1.57e-08 |
| 2 | TEX41 | 1074 | 1000000 | 145425533 | 145834291 | 3886 | 1e-06 | rs2381687 | 5.28e-06 |
| 3 | ETV5 | 159 | 1000000 | 185764105 | 185826901 | 791.5 | 2e-06 | rs12638263 | 9.53e-06 |
| 4 | THAP6 | 48 | 1000000 | 76439653 | 76455236 | 221.2 | 2e-06 | rs13103482 | 1.7e-05 |
| 6 | HCG14 | 4 | 1000000 | 28864306 | 28865097 | 32.63 | 2e-06 | rs3135316 | 0.000104 |
| 8 | C8orf12 | 322 | 1000000 | 11225910 | 11296166 | 2253 | 1e-06 | rs35009431 | 2.99e-09 |
| 8 | XKR6 | 1206 | 1000000 | 10753656 | 11058875 | 4940 | 1e-06 | rs4841507 | 7.69e-09 |
| 8 | MFHAS1 | 586 | 1000000 | 8641998 | 8751131 | 4007 | 1e-06 | rs11249893 | 8.41e-09 |
| 8 | TDH | 110 | 1000000 | 11197145 | 11225961 | 1122 | 1e-06 | rs2249804 | 2.55e-08 |
| 8 | SLC35G5 | 9 | 1000000 | 11188494 | 11189695 | 133.9 | 1e-06 | rs12681987 | 2.05e-07 |
| 12 | MON2 | 450 | 1000000 | 62860596 | 62991363 | 3278 | 2e-06 | rs10877884 | 1.5e-08 |
| 12 | C12orf61 | 9 | 1000000 | 62995530 | 62997214 | 101.9 | 1e-06 | rs10877885 | 1.52e-08 |
| 14 | PTPN21 | 297 | 1000000 | 88932121 | 89021123 | 2730 | 1e-06 | rs1152376 | 8.93e-08 |
| 14 | SPATA7 | 117 | 1000000 | 88851987 | 88904804 | 659.1 | 2e-06 | rs2747081 | 1.78e-07 |
| 15 | WDR76 | 107 | 1000000 | 44119111 | 44160617 | 828.1 | 2e-06 | rs4597285 | 1.84e-06 |
| 15 | RNU6-28P_15 | 518 | 1000000 | 43675287 | 43894832 | 3396 | 2e-06 | rs2444247 | 2.75e-06 |
| 16 | SHISA9 | 1670 | 1000000 | 12995476 | 13334273 | 5250 | 1e-06 | rs12922637 | 2.19e-08 |

Abbreviations: Chr = chromosome; nSNPs = number of SNPs; nSims = number of simulations

### Supplementary Table S9

Gene mapping FUMA ordered from most significant to less significant per chromosome

*part 1 of 2 — first column repeated in every part*

| uniqID | rsID | chr | pos | non_effect allele | effect_allele | MAF | gwasP | beta | se | r2 | IndSigSNP | GenomicLocus |
|---|---|---|---|---|---|---|---|---|---|---|---|---|
| 2:98580012:C:T | 2:98580012 | 2 | 98580012 | C | T | 0.3907 | 1.57e-08 | 0.008074 | 0.001427 | 1 | 2:98580012 | 1 |
| 2:98427382:C:T | 2:98427382 | 2 | 98427382 | C | T | 0.3926 | 2.07e-08 | 0.008 | 0.001426 | 0.9926 | 2:98580012 | 1 |
| 2:98354511:A:G | 2:98354511 | 2 | 98354511 | A | G | 0.3618 | 6.5e-08 | 0.007867 | 0.001454 | 0.8494 | 2:98580012 | 1 |
| 2:98357163:C:T | 2:98357163 | 2 | 98357163 | C | T | 0.3748 | 7.2e-08 | 0.007207 | 0.001337 | 0.9002 | 2:98580012 | 1 |
| 2:98520564:C:T | 2:98520564 | 2 | 98520564 | C | T | 0.3897 | 7.39e-08 | 0.007052 | 0.00131 | 0.9963 | 2:98580012 | 1 |
| 2:98370698:A:T | 2:98370698 | 2 | 98370698 | A | T | 0.3907 | 7.6e-08 | 0.007116 | 0.001323 | 0.9705 | 2:98580012 | 1 |
| 2:98493359:A:G | 2:98493359 | 2 | 98493359 | G | A | 0.3897 | 7.7e-08 | 0.007042 | 0.00131 | 0.9963 | 2:98580012 | 1 |
| 2:98519014:C:T | 2:98519014 | 2 | 98519014 | C | T | 0.3907 | 8.13e-08 | 0.007028 | 0.001309 | 1 | 2:98580012 | 1 |
| 2:98495661:G:T | 2:98495661 | 2 | 98495661 | G | T | 0.3907 | 8.68e-08 | 0.007012 | 0.001309 | 1 | 2:98580012 | 1 |
| 2:98440234:C:T | 2:98440234 | 2 | 98440234 | T | C | 0.3926 | 8.69e-08 | 0.007015 | 0.00131 | 0.9853 | 2:98580012 | 1 |
| 2:98491750:C:T | 2:98491750 | 2 | 98491750 | C | T | 0.3907 | 8.69e-08 | 0.007011 | 0.001309 | 1 | 2:98580012 | 1 |
| 2:98506910:C:T | 2:98506910 | 2 | 98506910 | C | T | 0.3907 | 8.7e-08 | 0.007012 | 0.001309 | 1 | 2:98580012 | 1 |
| 2:98521823:A:C | 2:98521823 | 2 | 98521823 | A | C | 0.3907 | 8.75e-08 | 0.00701 | 0.001309 | 1 | 2:98580012 | 1 |
| 2:98365164:C:G | 2:98365164 | 2 | 98365164 | C | G | 0.3926 | 8.77e-08 | 0.007025 | 0.001312 | 0.978 | 2:98580012 | 1 |
| 2:98502987:C:T | 2:98502987 | 2 | 98502987 | C | T | 0.3907 | 8.89e-08 | 0.007007 | 0.001309 | 1 | 2:98580012 | 1 |
| 2:98510166:G:T | 2:98510166 | 2 | 98510166 | G | T | 0.3907 | 9.47e-08 | 0.006992 | 0.001309 | 1 | 2:98580012 | 1 |
| 2:98623227:A:G | 2:98623227 | 2 | 98623227 | A | G | 0.3887 | 9.79e-08 | 0.007081 | 0.001327 | 0.9926 | 2:98580012 | 1 |
| 2:98623406:A:G | 2:98623406 | 2 | 98623406 | G | A | 0.3897 | 9.93e-08 | 0.007077 | 0.001327 | 0.9889 | 2:98580012 | 1 |
| 2:98410769:G:T | 2:98410769 | 2 | 98410769 | T | G | 0.3917 | 1.05e-07 | 0.006968 | 0.001309 | 0.989 | 2:98580012 | 1 |
| 2:98435982:C:T | 2:98435982 | 2 | 98435982 | T | C | 0.3926 | 1.06e-07 | 0.006964 | 0.001309 | 0.9926 | 2:98580012 | 1 |
| 2:98382886:A:G | 2:98382886 | 2 | 98382886 | A | G | 0.3986 | 1.08e-07 | 0.006991 | 0.001315 | 0.9565 | 2:98580012 | 1 |
| 2:98461821:C:T | 2:98461821 | 2 | 98461821 | T | C | 0.3907 | 1.08e-07 | 0.006959 | 0.001309 | 1 | 2:98580012 | 1 |
| 2:98567820:A:G | 2:98567820 | 2 | 98567820 | A | G | 0.3907 | 1.09e-07 | 0.006969 | 0.001311 | 0.9926 | 2:98580012 | 1 |
| 2:98409565:C:T | 2:98409565 | 2 | 98409565 | C | T | 0.3936 | 1.12e-07 | 0.006948 | 0.001309 | 0.989 | 2:98580012 | 1 |
| 2:98517626:A:G | 2:98517626 | 2 | 98517626 | A | G | 0.3897 | 1.12e-07 | 0.00697 | 0.001313 | 0.9963 | 2:98580012 | 1 |
| 2:98461953:C:T | 2:98461953 | 2 | 98461953 | T | C | 0.3917 | 1.13e-07 | 0.006949 | 0.001309 | 0.9963 | 2:98580012 | 1 |
| 2:98534531:C:T | 2:98534531 | 2 | 98534531 | T | C | 0.3897 | 1.14e-07 | 0.006956 | 0.001311 | 0.9963 | 2:98580012 | 1 |
| 2:98621060:A:T | 2:98621060 | 2 | 98621060 | T | A | 0.3907 | 1.14e-07 | -0.008091 | 0.001525 | 1 | 2:98580012 | 1 |
| 2:98570999:A:G | 2:98570999 | 2 | 98570999 | A | G | 0.3907 | 1.15e-07 | 0.006958 | 0.001311 | 1 | 2:98580012 | 1 |
| 2:98447004:C:T | 2:98447004 | 2 | 98447004 | C | T | 0.3917 | 1.18e-07 | 0.006939 | 0.001309 | 0.989 | 2:98580012 | 1 |
| 2:98449024:A:G | 2:98449024 | 2 | 98449024 | A | G | 0.3917 | 1.18e-07 | 0.006939 | 0.001309 | 0.989 | 2:98580012 | 1 |
| 2:98407350:C:T | 2:98407350 | 2 | 98407350 | T | C | 0.3926 | 1.19e-07 | 0.006934 | 0.001309 | 0.9926 | 2:98580012 | 1 |
| 2:98580724:G:T | 2:98580724 | 2 | 98580724 | T | G | 0.3907 | 1.19e-07 | 0.006953 | 0.001312 | 1 | 2:98580012 | 1 |
| 2:98588372:A:C | 2:98588372 | 2 | 98588372 | A | C | 0.3897 | 1.19e-07 | 0.006967 | 0.001315 | 0.9963 | 2:98580012 | 1 |
| 2:98565400:A:G | 2:98565400 | 2 | 98565400 | A | G | 0.3907 | 1.2e-07 | 0.006943 | 0.001311 | 1 | 2:98580012 | 1 |
| 2:98612260:C:T | 2:98612260 | 2 | 98612260 | C | T | 0.3887 | 1.21e-07 | 0.006988 | 0.001319 | 0.9926 | 2:98580012 | 1 |
| 2:98455510:C:G | 2:98455510 | 2 | 98455510 | C | G | 0.3926 | 1.22e-07 | 0.006929 | 0.001309 | 0.9926 | 2:98580012 | 1 |
| 2:98454572:C:G | 2:98454572 | 2 | 98454572 | C | G | 0.3926 | 1.23e-07 | 0.006928 | 0.001309 | 0.9926 | 2:98580012 | 1 |
| 2:98443658:C:T | 2:98443658 | 2 | 98443658 | T | C | 0.3926 | 1.25e-07 | 0.006924 | 0.001309 | 0.9926 | 2:98580012 | 1 |
| 2:98479581:A:C | 2:98479581 | 2 | 98479581 | A | C | 0.3887 | 1.25e-07 | 0.006942 | 0.001312 | 0.9926 | 2:98580012 | 1 |
| 2:98442202:A:G | 2:98442202 | 2 | 98442202 | A | G | 0.3926 | 1.26e-07 | 0.00693 | 0.00131 | 0.9853 | 2:98580012 | 1 |
| 2:98466647:C:T | 2:98466647 | 2 | 98466647 | T | C | 0.3907 | 1.28e-07 | 0.006918 | 0.001309 | 1 | 2:98580012 | 1 |
| 2:98454473:C:T | 2:98454473 | 2 | 98454473 | T | C | 0.3926 | 1.3e-07 | 0.006941 | 0.001314 | 0.9926 | 2:98580012 | 1 |

| uniqID | rsID | chr | pos | non_effect allele | effect_allele | MAF | gwasP | beta | se | r2 | IndSigSNP | GenomicLocus |
|---|---|---|---|---|---|---|---|---|---|---|---|---|
| 2:98417089:C:T | 2:98417089 | 2 | 98417089 | T | C | 0.3926 | 1.32e-07 | 0.006909 | 0.001309 | 0.9926 | 2:98580012 | 1 |
| 2:98421364:A:G | 2:98421364 | 2 | 98421364 | G | A | 0.3917 | 1.32e-07 | 0.00691 | 0.001309 | 0.989 | 2:98580012 | 1 |
| 2:98541001:C:T | 2:98541001 | 2 | 98541001 | C | T | 0.3907 | 1.32e-07 | 0.006918 | 0.00131 | 1 | 2:98580012 | 1 |
| 2:98539588:A:C | 2:98539588 | 2 | 98539588 | A | C | 0.3907 | 1.33e-07 | 0.006915 | 0.00131 | 1 | 2:98580012 | 1 |
| 2:98424802:A:G | 2:98424802 | 2 | 98424802 | G | A | 0.3926 | 1.34e-07 | 0.006906 | 0.001309 | 0.9926 | 2:98580012 | 1 |
| 2:98552299:C:G | 2:98552299 | 2 | 98552299 | G | C | 0.3897 | 1.36e-07 | 0.006915 | 0.001311 | 0.9963 | 2:98580012 | 1 |
| 2:98405695:C:T | 2:98405695 | 2 | 98405695 | T | C | 0.3917 | 1.37e-07 | 0.006902 | 0.001309 | 0.989 | 2:98580012 | 1 |
| 2:98409046:A:G | 2:98409046 | 2 | 98409046 | A | G | 0.3917 | 1.38e-07 | 0.00692 | 0.001313 | 0.989 | 2:98580012 | 1 |
| 2:98454472:A:G | 2:98454472 | 2 | 98454472 | A | G | 0.3926 | 1.38e-07 | 0.006926 | 0.001314 | 0.9854 | 2:98580012 | 1 |
| 2:98565144:A:G | 2:98565144 | 2 | 98565144 | A | G | 0.3907 | 1.38e-07 | 0.00691 | 0.001311 | 1 | 2:98580012 | 1 |
| 2:98587288:A:G | 2:98587288 | 2 | 98587288 | G | A | 0.3887 | 1.38e-07 | 0.006955 | 0.001319 | 0.9927 | 2:98580012 | 1 |
| 2:98420431:C:T | 2:98420431 | 2 | 98420431 | C | T | 0.3926 | 1.39e-07 | 0.006896 | 0.001309 | 0.9926 | 2:98580012 | 1 |
| 2:98472610:C:T | 2:98472610 | 2 | 98472610 | C | T | 0.3887 | 1.39e-07 | 0.00692 | 0.001313 | 0.9927 | 2:98580012 | 1 |
| 2:98419726:A:T | 2:98419726 | 2 | 98419726 | A | T | 0.3926 | 1.4e-07 | 0.006895 | 0.001309 | 0.9926 | 2:98580012 | 1 |
| 2:98420142:C:G | 2:98420142 | 2 | 98420142 | C | G | 0.3926 | 1.4e-07 | 0.006894 | 0.001309 | 0.9926 | 2:98580012 | 1 |
| 2:98528689:A:G | 2:98528689 | 2 | 98528689 | A | G | 0.3897 | 1.4e-07 | 0.006915 | 0.001312 | 0.9963 | 2:98580012 | 1 |
| 2:98552271:C:G | 2:98552271 | 2 | 98552271 | C | G | 0.3887 | 1.4e-07 | 0.00693 | 0.001315 | 0.9927 | 2:98580012 | 1 |
| 2:98405929:A:G | 2:98405929 | 2 | 98405929 | G | A | 0.3926 | 1.42e-07 | 0.006891 | 0.001309 | 0.9926 | 2:98580012 | 1 |
| 2:98606770:C:T | 2:98606770 | 2 | 98606770 | C | T | 0.3877 | 1.46e-07 | 0.00695 | 0.001321 | 0.989 | 2:98580012 | 1 |
| 2:98413781:C:T | 2:98413781 | 2 | 98413781 | T | C | 0.3926 | 1.47e-07 | 0.006899 | 0.001312 | 0.9926 | 2:98580012 | 1 |
| 2:98455152:C:T | 2:98455152 | 2 | 98455152 | T | C | 0.3936 | 1.47e-07 | 0.006895 | 0.001311 | 0.989 | 2:98580012 | 1 |
| 2:98377512:A:T | 2:98377512 | 2 | 98377512 | T | A | 0.3926 | 1.56e-07 | 0.006878 | 0.00131 | 0.9926 | 2:98580012 | 1 |
| 2:98379813:A:G | 2:98379813 | 2 | 98379813 | A | G | 0.3926 | 1.56e-07 | 0.006875 | 0.00131 | 0.9926 | 2:98580012 | 1 |
| 2:98379267:C:T | 2:98379267 | 2 | 98379267 | T | C | 0.3926 | 1.57e-07 | 0.006874 | 0.00131 | 0.9926 | 2:98580012 | 1 |
| 2:98397301:C:T | 2:98397301 | 2 | 98397301 | T | C | 0.3917 | 1.58e-07 | 0.006866 | 0.001309 | 0.989 | 2:98580012 | 1 |
| 2:98402753:A:T | 2:98402753 | 2 | 98402753 | A | T | 0.3926 | 1.58e-07 | 0.006866 | 0.001309 | 0.9926 | 2:98580012 | 1 |
| 2:98402772:C:G | 2:98402772 | 2 | 98402772 | C | G | 0.3907 | 1.59e-07 | 0.006887 | 0.001313 | 0.9853 | 2:98580012 | 1 |
| 2:98395653:A:C | 2:98395653 | 2 | 98395653 | C | A | 0.3926 | 1.65e-07 | 0.006855 | 0.001309 | 0.9926 | 2:98580012 | 1 |
| 2:98374567:C:T | 2:98374567 | 2 | 98374567 | T | C | 0.3907 | 1.7e-07 | 0.006859 | 0.001311 | 0.9853 | 2:98580012 | 1 |
| 2:98561153:A:C | 2:98561153 | 2 | 98561153 | A | C | 0.3907 | 1.71e-07 | 0.006872 | 0.001313 | 1 | 2:98580012 | 1 |
| 2:98386731:C:T | 2:98386731 | 2 | 98386731 | C | T | 0.3917 | 1.82e-07 | 0.006839 | 0.00131 | 0.989 | 2:98580012 | 1 |
| 2:98393894:C:T | 2:98393894 | 2 | 98393894 | T | C | 0.3917 | 1.84e-07 | 0.006851 | 0.001313 | 0.9817 | 2:98580012 | 1 |
| 2:98557575:G:T | 2:98557575 | 2 | 98557575 | G | T | 0.3897 | 1.85e-07 | -0.007775 | 0.00149 | 0.9963 | 2:98580012 | 1 |
| 2:98605982:A:G | 2:98605982 | 2 | 98605982 | A | G | 0.3897 | 1.88e-07 | -0.007884 | 0.001512 | 0.9963 | 2:98580012 | 1 |
| 2:98501884:C:T | 2:98501884 | 2 | 98501884 | C | T | 0.3897 | 1.95e-07 | -0.007752 | 0.001489 | 0.9963 | 2:98580012 | 1 |
| 2:98416848:A:C | 2:98416848 | 2 | 98416848 | A | C | 0.3907 | 2.06e-07 | 0.006835 | 0.001315 | 0.9853 | 2:98580012 | 1 |
| 2:98416850:G:T | 2:98416850 | 2 | 98416850 | G | T | 0.3907 | 2.06e-07 | 0.006834 | 0.001315 | 0.9853 | 2:98580012 | 1 |
| 2:98554946:A:C | 2:98554946 | 2 | 98554946 | C | A | 0.3897 | 2.11e-07 | -0.007741 | 0.001491 | 0.9963 | 2:98580012 | 1 |
| 2:98448395:G:T | 2:98448395 | 2 | 98448395 | T | G | 0.3936 | 3.14e-07 | 0.006727 | 0.001314 | 0.989 | 2:98580012 | 1 |
| 2:98602165:A:G | 2:98602165 | 2 | 98602165 | G | A | 0.3897 | 3.77e-07 | 0.006718 | 0.001321 | 0.9889 | 2:98580012 | 1 |
| 2:98342323:A:C | 2:98342323 | 2 | 98342323 | A | C | 0.3926 | 3.83e-07 | 0.006673 | 0.001313 | 0.9274 | 2:98580012 | 1 |
| 2:98488949:C:G | 2:98488949 | 2 | 98488949 | C | G | 0.3907 | 4.15e-07 | 0.006657 | 0.001314 | 1 | 2:98580012 | 1 |
| 2:98454931:A:G | 2:98454931 | 2 | 98454931 | A | G | 0.3917 | 5.52e-07 | 0.007512 | 0.001499 | 0.989 | 2:98580012 | 1 |
| 2:98443039:C:G | 2:98443039 | 2 | 98443039 | C | G | 0.3926 | 5.71e-07 | 0.006573 | 0.001313 | 0.9926 | 2:98580012 | 1 |
| 2:98336200:A:C | 2:98336200 | 2 | 98336200 | C | A | 0.4066 | 5.87e-07 | 0.006567 | 0.001314 | 0.8861 | 2:98580012 | 1 |
| 2:98405034:C:T | 2:98405034 | 2 | 98405034 | C | T | 0.3917 | 6.38e-07 | 0.006544 | 0.001313 | 0.989 | 2:98580012 | 1 |
| 2:98355990:A:G | 2:98355990 | 2 | 98355990 | A | G | 0.3628 | 1.38e-06 | 0.00644 | 0.001333 | 0.8454 | 2:98580012 | 1 |

| uniqID | rsID | chr | pos | non_effect allele | effect_allele | MAF | gwasP | beta | se | r2 | IndSigSNP | GenomicLocus |
|---|---|---|---|---|---|---|---|---|---|---|---|---|
| 2:98534412:C:T | 2:98534412 | 2 | 98534412 | T | C | 0.3956 | 1.8e-06 | -0.007266 | 0.001521 | 0.9818 | 2:98580012 | 1 |
| 2:98354139:G:T | 2:98354139 | 2 | 98354139 | T | G | 0.3618 | 1.93e-06 | 0.006353 | 0.001333 | 0.8423 | 2:98580012 | 1 |
| 2:98353847:C:T | 2:98353847 | 2 | 98353847 | T | C | 0.3628 | 2.19e-06 | 0.006332 | 0.001336 | 0.8454 | 2:98580012 | 1 |
| 2:98339513:C:T | 2:98339513 | 2 | 98339513 | T | C | 0.3608 | 4.2e-06 | 0.006157 | 0.001337 | 0.8111 | 2:98580012 | 1 |
| 2:98325330:G:T | 2:98325330 | 2 | 98325330 | T | G | 0.4245 | 4.78e-06 | -0.00668 | 0.001459 | 0.71 | 2:98580012 | 1 |
| 2:98326867:A:G | 2:98326867 | 2 | 98326867 | A | G | 0.4245 | 6.08e-06 | 0.005863 | 0.001295 | 0.71 | 2:98580012 | 1 |
| 2:98330052:A:C | 2:98330052 | 2 | 98330052 | A | C | 0.4235 | 9.06e-06 | 0.005754 | 0.001295 | 0.7131 | 2:98580012 | 1 |
| 2:98356846:A:G | 2:98356846 | 2 | 98356846 | A | G | 0.4871 | 9.82e-06 | 0.005687 | 0.001285 | 0.6105 | 2:98580012 | 1 |
| 2:98361679:C:G | 2:98361679 | 2 | 98361679 | G | C | 0.4851 | 1.58e-05 | 0.005552 | 0.001285 | 0.605 | 2:98580012 | 1 |
| 2:98360443:A:G | 2:98360443 | 2 | 98360443 | G | A | 0.4831 | 1.82e-05 | 0.005517 | 0.001286 | 0.602 | 2:98580012 | 1 |
| 2:98363313:A:T | 2:98363313 | 2 | 98363313 | A | T | 0.4841 | 3.39e-05 | 0.00533 | 0.001285 | 0.6041 | 2:98580012 | 1 |
| 2:98334679:C:T | 2:98334679 | 2 | 98334679 | T | C | 0.3638 | 4.54e-05 | 0.005509 | 0.00135 | 0.7993 | 2:98580012 | 1 |
| 2:98351986:C:T | 2:98351986 | 2 | 98351986 | C | T | 0.498 | 8.75e-05 | 0.005052 | 0.001287 | 0.6453 | 2:98580012 | 1 |
| 2:98343258:A:G | 2:98343258 | 2 | 98343258 | G | A | 0.493 | 8.91e-05 | 0.005048 | 0.001288 | 0.6269 | 2:98580012 | 1 |
| 2:98345086:G:T | 2:98345086 | 2 | 98345086 | G | T | 0.4891 | 0.000109 | 0.005415 | 0.001399 | 0.6206 | 2:98580012 | 1 |
| 2:98393231:C:T | 2:98393231 | 2 | 98393231 | T | C | 0.3111 | 0.000698 | 0.004691 | 0.001383 | 0.6975 | 2:98580012 | 1 |
| 2:98346461:A:G | 2:98346461 | 2 | 98346461 | A | G | 0.498 | 0.000895 | -0.004907 | 0.001477 | 0.6404 | 2:98580012 | 1 |
| 2:98571084:A:G | 2:98571084 | 2 | 98571084 | G | A | 0.3897 | 1,00E-07 | 0.006996 | 0.001313 | 0.9963 | 2:98580012 | 1 |
| 2:98351654:T:TG | rs34532595 | 2 | 98351654 | TG | T | 0.3608 | NA | NA | NA | 0.846 | 2:98580012 | 1 |
| 2:98368551:A:ACT | rs34388573 | 2 | 98368551 | ACT | A | 0.4831 | NA | NA | NA | 0.6008 | 2:98580012 | 1 |
| 2:98384528:A:ATTTTC | rs149697530 | 2 | 98384528 | ATTTTC | A | 0.3996 | NA | NA | NA | 0.9679 | 2:98580012 | 1 |
| 2:98390117:G:GT | rs538393585 | 2 | 98390117 | G | GT | 0.3579 | NA | NA | NA | 0.8414 | 2:98580012 | 1 |
| 2:98434931:G:GGGGGC | rs147071183 | 2 | 98434931 | GGGGGC | G | 0.3897 | NA | NA | NA | 0.9816 | 2:98580012 | 1 |
| 2:98443081:G:GA | rs35808422 | 2 | 98443081 | GA | G | 0.3917 | NA | NA | NA | 0.989 | 2:98580012 | 1 |
| 2:98468892:A:AC | rs58300457 | 2 | 98468892 | AC | A | 0.3907 | NA | NA | NA | 1 | 2:98580012 | 1 |
| 2:98506011:T:TATA | rs71386038 | 2 | 98506011 | TATA | T | 0.3887 | NA | NA | NA | 0.9927 | 2:98580012 | 1 |
| 2:98507972:C:CA | rs35677645 | 2 | 98507972 | CA | C | 0.3907 | NA | NA | NA | 1 | 2:98580012 | 1 |
| 2:98526685:A:AAAAT | rs148038816 | 2 | 98526685 | AAAAT | A | 0.3897 | NA | NA | NA | 0.9963 | 2:98580012 | 1 |
| 2:98547069:A:AT | rs35058741 | 2 | 98547069 | AT | A | 0.3907 | NA | NA | NA | 1 | 2:98580012 | 1 |
| 2:98548604:T:TA | rs34976723 | 2 | 98548604 | TA | T | 0.4453 | NA | NA | NA | 0.7157 | 2:98580012 | 1 |
| 2:98551456:G:GA | rs545443077 | 2 | 98551456 | G | GA | 0.3857 | NA | NA | NA | 0.9668 | 2:98580012 | 1 |
| 2:98565115:C:CT | rs137894541 | 2 | 98565115 | C | CT | 0.3877 | NA | NA | NA | 0.989 | 2:98580012 | 1 |
| 2:98604570:G:GACAC | rs140523936 | 2 | 98604570 | GACAC | G | 0.3887 | NA | NA | NA | 0.9926 | 2:98580012 | 1 |
| 2:98616519:A:ATT | rs370334808 | 2 | 98616519 | ATT | A | 0.4702 | NA | NA | NA | 0.7289 | 2:98580012 | 1 |
| 2:98621058:A:AG | rs147329160 | 2 | 98621058 | AG | A | 0.3907 | NA | NA | NA | 1 | 2:98580012 | 1 |
| 8:11417582:C:T | 8:11417582 | 8 | 11417582 | T | C | 0.339 | 9.09e-11 | -0.01009 | 0.001555 | 1 | 8:11417582 | 3 |
| 8:11235150:A:C | 8:11235150 | 8 | 11235150 | A | C | 0.4672 | 1.15e-09 | -0.00851 | 0.001397 | 1 | 8:11235150 | 3 |
| 8:11226456:A:C | 8:11226456 | 8 | 11226456 | A | C | 0.4602 | 1.29e-09 | -0.007853 | 0.001293 | 0.9579 | 8:11235150 | 3 |
| 8:10147398:C:T | 8:10147398 | 8 | 10147398 | T | C | 0.4344 | 1.47e-09 | -0.007824 | 0.001293 | 1 | 8:10147398 | 3 |
| 8:11226071:A:G | 8:11226071 | 8 | 11226071 | G | A | 0.4692 | 2.43e-09 | -0.007704 | 0.00129 | 0.9238 | 8:11235150 | 3 |
| 8:11232788:A:G | 8:11232788 | 8 | 11232788 | A | G | 0.4642 | 2.99e-09 | -0.007669 | 0.001292 | 0.9885 | 8:11235150 | 3 |
| 8:11231249:C:T | 8:11231249 | 8 | 11231249 | C | T | 0.4891 | 3.38e-09 | 0.007613 | 0.001287 | 0.8832 | 8:11235150 | 3 |
| 8:11242632:A:G | 8:11242632 | 8 | 11242632 | A | G | 0.4662 | 3.57e-09 | -0.007632 | 0.001292 | 0.9733 | 8:11235150 | 3 |
| 8:11233318:A:C | 8:11233318 | 8 | 11233318 | C | A | 0.4781 | 3.74e-09 | -0.007603 | 0.001289 | 0.9438 | 8:11235150 | 3 |
| 8:11236681:G:T | 8:11236681 | 8 | 11236681 | G | T | 0.4672 | 3.92e-09 | -0.007595 | 0.001289 | 1 | 8:11235150 | 3 |
| 8:11236685:C:T | 8:11236685 | 8 | 11236685 | T | C | 0.4672 | 3.93e-09 | -0.007594 | 0.001289 | 1 | 8:11235150 | 3 |
| 8:11236419:C:T | 8:11236419 | 8 | 11236419 | C | T | 0.4672 | 4.02e-09 | -0.007605 | 0.001292 | 1 | 8:11235150 | 3 |

| uniqID | rsID | chr | pos | non_effect allele | effect_allele | MAF | gwasP | beta | se | r2 | IndSigSNP | GenomicLocus |
|---|---|---|---|---|---|---|---|---|---|---|---|---|
| 8:11245303:C:G | 8:11245303 | 8 | 11245303 | G | C | 0.4781 | 4.04e-09 | 0.007598 | 0.001291 | 0.9137 | 8:11235150 | 3 |
| 8:11236413:C:T | 8:11236413 | 8 | 11236413 | T | C | 0.4672 | 4.1e-09 | -0.007601 | 0.001292 | 1 | 8:11235150 | 3 |
| 8:11236572:A:T | 8:11236572 | 8 | 11236572 | A | T | 0.4672 | 4.12e-09 | -0.00758 | 0.001288 | 1 | 8:11235150 | 3 |
| 8:11230574:A:G | 8:11230574 | 8 | 11230574 | A | G | 0.4632 | 4.15e-09 | -0.007598 | 0.001292 | 0.9771 | 8:11235150 | 3 |
| 8:11237477:A:G | 8:11237477 | 8 | 11237477 | G | A | 0.4672 | 4.26e-09 | -0.007577 | 0.001289 | 1 | 8:11235150 | 3 |
| 8:11237480:A:G | 8:11237480 | 8 | 11237480 | G | A | 0.4672 | 4.26e-09 | -0.007577 | 0.001289 | 1 | 8:11235150 | 3 |
| 8:11233419:G:T | 8:11233419 | 8 | 11233419 | G | T | 0.4781 | 4.35e-09 | -0.007571 | 0.001289 | 0.9438 | 8:11235150 | 3 |
| 8:11231886:A:G | 8:11231886 | 8 | 11231886 | G | A | 0.4722 | 4.41e-09 | -0.007569 | 0.001289 | 0.9506 | 8:11235150 | 3 |
| 8:11236392:C:T | 8:11236392 | 8 | 11236392 | C | T | 0.4672 | 4.7e-09 | -0.007572 | 0.001292 | 1 | 8:11235150 | 3 |
| 8:11227406:A:G | 8:11227406 | 8 | 11227406 | A | G | 0.4622 | 4.75e-09 | -0.007569 | 0.001292 | 0.9655 | 8:11235150 | 3 |
| 8:11239762:A:T | 8:11239762 | 8 | 11239762 | T | A | 0.4801 | 4.78e-09 | -0.007547 | 0.001288 | 0.9292 | 8:11235150 | 3 |
| 8:11249010:G:T | 8:11249010 | 8 | 11249010 | T | G | 0.4722 | 4.78e-09 | 0.007569 | 0.001292 | 0.8479 | 8:11235150 | 3 |
| 8:11232860:A:G | 8:11232860 | 8 | 11232860 | A | G | 0.4672 | 4.79e-09 | -0.007565 | 0.001291 | 0.9847 | 8:11235150 | 3 |
| 8:11080675:C:T | 8:11080675 | 8 | 11080675 | T | C | 0.492 | 4.9e-09 | -0.007576 | 0.001294 | 1 | 8:11080675 | 3 |
| 8:11227885:C:T | 8:11227885 | 8 | 11227885 | T | C | 0.4592 | 4.9e-09 | -0.007563 | 0.001292 | 0.9619 | 8:11235150 | 3 |
| 8:11228100:A:G | 8:11228100 | 8 | 11228100 | A | G | 0.4622 | 4.98e-09 | -0.007556 | 0.001291 | 0.9655 | 8:11235150 | 3 |
| 8:11234500:A:T | 8:11234500 | 8 | 11234500 | T | A | 0.4801 | 5.04e-09 | -0.007536 | 0.001288 | 0.9367 | 8:11235150 | 3 |
| 8:11234520:A:G | 8:11234520 | 8 | 11234520 | G | A | 0.4672 | 5.04e-09 | -0.007548 | 0.00129 | 1 | 8:11235150 | 3 |
| 8:11234885:C:G | 8:11234885 | 8 | 11234885 | G | C | 0.4672 | 5.12e-09 | -0.007541 | 0.00129 | 1 | 8:11235150 | 3 |
| 8:11233582:A:G | 8:11233582 | 8 | 11233582 | G | A | 0.4781 | 5.16e-09 | -0.007535 | 0.001289 | 0.9438 | 8:11235150 | 3 |
| 8:11233659:A:G | 8:11233659 | 8 | 11233659 | G | A | 0.4781 | 5.18e-09 | -0.007534 | 0.001289 | 0.9438 | 8:11235150 | 3 |
| 8:11234298:C:T | 8:11234298 | 8 | 11234298 | T | C | 0.4672 | 5.18e-09 | -0.007542 | 0.00129 | 1 | 8:11235150 | 3 |
| 8:11062882:C:T | 8:11062882 | 8 | 11062882 | C | T | 0.496 | 5.19e-09 | -0.007525 | 0.001287 | 0.8732 | 8:11080675 | 3 |
| 8:11234844:A:G | 8:11234844 | 8 | 11234844 | A | G | 0.4672 | 5.31e-09 | -0.007537 | 0.00129 | 1 | 8:11235150 | 3 |
| 8:11233958:A:G | 8:11233958 | 8 | 11233958 | G | A | 0.4642 | 5.37e-09 | -0.007539 | 0.001291 | 0.9885 | 8:11235150 | 3 |
| 8:11236975:A:G | 8:11236975 | 8 | 11236975 | G | A | 0.4672 | 5.4e-09 | -0.007528 | 0.001289 | 1 | 8:11235150 | 3 |
| 8:11232343:C:G | 8:11232343 | 8 | 11232343 | G | C | 0.4662 | 5.42e-09 | -0.007538 | 0.001291 | 0.9808 | 8:11235150 | 3 |
| 8:11234780:C:G | 8:11234780 | 8 | 11234780 | G | C | 0.4672 | 5.47e-09 | -0.007531 | 0.00129 | 1 | 8:11235150 | 3 |
| 8:11230206:G:T | 8:11230206 | 8 | 11230206 | T | G | 0.4622 | 5.51e-09 | -0.007537 | 0.001292 | 0.9809 | 8:11235150 | 3 |
| 8:11233917:C:T | 8:11233917 | 8 | 11233917 | T | C | 0.4642 | 5.51e-09 | -0.007534 | 0.001291 | 0.9885 | 8:11235150 | 3 |
| 8:11234367:G:T | 8:11234367 | 8 | 11234367 | G | T | 0.4672 | 5.57e-09 | -0.007527 | 0.00129 | 1 | 8:11235150 | 3 |
| 8:11235360:A:C | 8:11235360 | 8 | 11235360 | C | A | 0.4682 | 5.59e-09 | -0.007524 | 0.00129 | 0.9961 | 8:11235150 | 3 |
| 8:11234626:C:T | 8:11234626 | 8 | 11234626 | C | T | 0.4672 | 5.61e-09 | -0.00753 | 0.001291 | 1 | 8:11235150 | 3 |
| 8:11235497:A:G | 8:11235497 | 8 | 11235497 | A | G | 0.4672 | 5.7e-09 | -0.007514 | 0.001289 | 1 | 8:11235150 | 3 |
| 8:11234613:A:T | 8:11234613 | 8 | 11234613 | A | T | 0.4672 | 5.78e-09 | -0.007523 | 0.001291 | 1 | 8:11235150 | 3 |
| 8:10988275:A:G | 8:10988275 | 8 | 10988275 | G | A | 0.4264 | 5.79e-09 | -0.008199 | 0.001407 | 1 | 8:10988275 | 3 |
| 8:11230259:A:T | 8:11230259 | 8 | 11230259 | T | A | 0.4622 | 5.79e-09 | -0.007527 | 0.001292 | 0.9809 | 8:11235150 | 3 |
| 8:11236850:C:T | 8:11236850 | 8 | 11236850 | T | C | 0.4672 | 5.81e-09 | -0.007503 | 0.001288 | 1 | 8:11235150 | 3 |
| 8:11236964:C:T | 8:11236964 | 8 | 11236964 | T | C | 0.4672 | 5.81e-09 | -0.007504 | 0.001288 | 1 | 8:11235150 | 3 |
| 8:11235735:C:T | 8:11235735 | 8 | 11235735 | C | T | 0.4672 | 5.85e-09 | -0.007512 | 0.00129 | 1 | 8:11235150 | 3 |
| 8:11229319:C:G | 8:11229319 | 8 | 11229319 | G | C | 0.4592 | 5.9e-09 | -0.007531 | 0.001293 | 0.9698 | 8:11235150 | 3 |
| 8:11080665:A:T | 8:11080665 | 8 | 11080665 | A | T | 0.492 | 6.04e-09 | -0.00753 | 0.001294 | 1 | 8:11080675 | 3 |
| 8:11235136:C:G | 8:11235136 | 8 | 11235136 | G | C | 0.4672 | 6.04e-09 | -0.00751 | 0.00129 | 1 | 8:11235150 | 3 |
| 8:11249261:C:T | 8:11249261 | 8 | 11249261 | C | T | 0.3797 | 6.14e-09 | 0.008415 | 0.001447 | 1 | 8:11249261 | 3 |
| 8:11243126:C:G | 8:11243126 | 8 | 11243126 | G | C | 0.4563 | 6.27e-09 | -0.007524 | 0.001294 | 0.9522 | 8:11235150 | 3 |
| 8:11236809:C:T | 8:11236809 | 8 | 11236809 | C | T | 0.4672 | 6.35e-09 | -0.007484 | 0.001288 | 1 | 8:11235150 | 3 |
| 8:11238029:C:T | 8:11238029 | 8 | 11238029 | T | C | 0.4553 | 6.4e-09 | -0.007501 | 0.001291 | 0.9557 | 8:11235150 | 3 |

| uniqID | rsID | chr | pos | non_effect allele | effect_allele | MAF | gwasP | beta | se | r2 | IndSigSNP | GenomicLocus |
|---|---|---|---|---|---|---|---|---|---|---|---|---|
| 8:11239640:C:T | 8:11239640 | 8 | 11239640 | T | C | 0.4612 | 6.44e-09 | -0.00751 | 0.001293 | 0.955 | 8:11235150 | 3 |
| 8:11231354:G:T | 8:11231354 | 8 | 11231354 | T | G | 0.4881 | 6.46e-09 | 0.007478 | 0.001287 | 0.8937 | 8:11235150 | 3 |
| 8:11235605:G:T | 8:11235605 | 8 | 11235605 | G | T | 0.4672 | 6.55e-09 | -0.007492 | 0.00129 | 1 | 8:11235150 | 3 |
| 8:11235393:A:G | 8:11235393 | 8 | 11235393 | G | A | 0.4662 | 6.65e-09 | -0.007485 | 0.00129 | 0.9962 | 8:11235150 | 3 |
| 8:11235579:G:T | 8:11235579 | 8 | 11235579 | G | T | 0.4672 | 7.06e-09 | -0.007475 | 0.00129 | 1 | 8:11235150 | 3 |
| 8:11046394:C:G | 8:11046394 | 8 | 11046394 | G | C | 0.4076 | 7.69e-09 | -0.008162 | 0.001412 | 0.7896 | 8:10988275 | 3 |
| 8:11239510:A:G | 8:11239510 | 8 | 11239510 | A | G | 0.4612 | 7.93e-09 | -0.00746 | 0.001292 | 0.955 | 8:11235150 | 3 |
| 8:11239565:C:T | 8:11239565 | 8 | 11239565 | T | C | 0.4612 | 7.99e-09 | -0.007459 | 0.001292 | 0.955 | 8:11235150 | 3 |
| 8:10903475:A:T | 8:10903475 | 8 | 10903475 | A | T | 0.4881 | 8.09e-09 | -0.007448 | 0.001291 | 0.7052 | 8:11080675 | 3 |
| 8:11239352:A:G | 8:11239352 | 8 | 11239352 | A | G | 0.4602 | 8.18e-09 | -0.00745 | 0.001291 | 0.9589 | 8:11235150 | 3 |
| 8:11229638:C:T | 8:11229638 | 8 | 11229638 | T | C | 0.4652 | 8.2e-09 | -0.007456 | 0.001293 | 0.9692 | 8:11235150 | 3 |
| 8:8700851:C:T | 8:8700851 | 8 | 8700851 | T | C | 0.4453 | 8.41e-09 | 0.007429 | 0.001289 | 1 | 8:8700851 | 2 |
| 8:11239137:A:G | 8:11239137 | 8 | 11239137 | A | G | 0.4602 | 8.81e-09 | -0.007434 | 0.001291 | 0.9589 | 8:11235150 | 3 |
| 8:11238597:C:T | 8:11238597 | 8 | 11238597 | T | C | 0.4602 | 8.92e-09 | -0.007428 | 0.001291 | 0.9589 | 8:11235150 | 3 |
| 8:11061792:C:T | 8:11061792 | 8 | 11061792 | T | C | 0.497 | 8.98e-09 | -0.007402 | 0.001287 | 0.8694 | 8:11080675 | 3 |
| 8:11060311:A:C | 8:11060311 | 8 | 11060311 | C | A | 0.498 | 9.05e-09 | -0.007402 | 0.001287 | 0.8656 | 8:11080675 | 3 |
| 8:11071057:A:G | 8:11071057 | 8 | 11071057 | G | A | 0.4165 | 9.1e-09 | -0.007484 | 0.001301 | 0.7369 | 8:10988275 | 3 |
| 8:11237773:C:T | 8:11237773 | 8 | 11237773 | C | T | 0.4622 | 9.23e-09 | -0.007413 | 0.00129 | 0.9585 | 8:11235150 | 3 |
| 8:8646246:C:T | 8:8646246 | 8 | 8646246 | C | T | 0.4483 | 9.37e-09 | 0.007416 | 0.001291 | 0.8949 | 8:8700851 | 2 |
| 8:11235910:A:G | 8:11235910 | 8 | 11235910 | A | G | 0.4592 | 9.37e-09 | -0.007423 | 0.001292 | 0.9702 | 8:11235150 | 3 |
| 8:11239078:G:T | 8:11239078 | 8 | 11239078 | G | T | 0.4602 | 9.43e-09 | -0.007419 | 0.001291 | 0.9589 | 8:11235150 | 3 |
| 8:11237756:A:C | 8:11237756 | 8 | 11237756 | A | C | 0.4612 | 9.75e-09 | -0.007376 | 0.001285 | 0.9625 | 8:11235150 | 3 |
| 8:11239017:A:T | 8:11239017 | 8 | 11239017 | A | T | 0.4602 | 9.87e-09 | -0.007409 | 0.001291 | 0.9589 | 8:11235150 | 3 |
| 8:11239054:C:T | 8:11239054 | 8 | 11239054 | T | C | 0.4602 | 1.03e-08 | -0.007403 | 0.001292 | 0.9589 | 8:11235150 | 3 |
| 8:11079367:A:G | 8:11079367 | 8 | 11079367 | A | G | 0.4841 | 1.08e-08 | -0.007379 | 0.00129 | 0.9467 | 8:11080675 | 3 |
| 8:11078949:A:G | 8:11078949 | 8 | 11078949 | A | G | 0.4861 | 1.42e-08 | -0.007319 | 0.00129 | 0.9389 | 8:11080675 | 3 |
| 8:11247814:A:G | 8:11247814 | 8 | 11247814 | A | G | 0.3887 | 1.46e-08 | 0.007548 | 0.001331 | 0.9562 | 8:11249261 | 3 |
| 8:8633548:G:T | 8:8633548 | 8 | 8633548 | G | T | 0.4553 | 1.51e-08 | -0.007318 | 0.001292 | 0.8625 | 8:8700851 | 2 |
| 8:8664622:A:G | 8:8664622 | 8 | 8664622 | A | G | 0.4245 | 1.61e-08 | 0.007302 | 0.001292 | 0.8145 | 8:8700851 | 2 |
| 8:11076635:G:T | 8:11076635 | 8 | 11076635 | G | T | 0.4781 | 1.63e-08 | -0.007286 | 0.001289 | 0.925 | 8:11080675 | 3 |
| 8:8661534:C:T | 8:8661534 | 8 | 8661534 | C | T | 0.4473 | 1.65e-08 | 0.007262 | 0.001286 | 0.898 | 8:8700851 | 2 |
| 8:8652889:A:G | 8:8652889 | 8 | 8652889 | A | G | 0.4463 | 1.67e-08 | 0.007269 | 0.001287 | 0.8943 | 8:8700851 | 2 |
| 8:11078781:A:C | 8:11078781 | 8 | 11078781 | C | A | 0.4821 | 1.68e-08 | -0.007282 | 0.00129 | 0.9243 | 8:11080675 | 3 |
| 8:8661681:C:G | 8:8661681 | 8 | 8661681 | G | C | 0.4304 | 1.92e-08 | 0.007243 | 0.001288 | 0.8435 | 8:8700851 | 2 |
| 8:8685854:A:G | 8:8685854 | 8 | 8685854 | G | A | 0.4543 | 1.92e-08 | 0.007265 | 0.001292 | 0.8938 | 8:8700851 | 2 |
| 8:8641145:C:T | 8:8641145 | 8 | 8641145 | T | C | 0.4553 | 2.01e-08 | 0.007248 | 0.001291 | 0.8694 | 8:8700851 | 2 |
| 8:11219781:C:T | 8:11219781 | 8 | 11219781 | T | C | 0.4652 | 2.03e-08 | 0.007242 | 0.00129 | 0.8863 | 8:11235150 | 3 |
| 8:8637429:A:G | 8:8637429 | 8 | 8637429 | A | G | 0.4543 | 2.15e-08 | -0.007226 | 0.00129 | 0.8583 | 8:8700851 | 2 |
| 8:8721473:A:G | 8:8721473 | 8 | 8721473 | A | G | 0.4095 | 2.18e-08 | 0.007253 | 0.001295 | 0.8532 | 8:8700851 | 2 |
| 8:10987553:C:T | 8:10987553 | 8 | 10987553 | C | T | 0.4264 | 2.2e-08 | -0.007267 | 0.001298 | 1 | 8:10988275 | 3 |
| 8:8770512:C:T | 8:8770512 | 8 | 8770512 | T | C | 0.4036 | 2.27e-08 | 0.007282 | 0.001302 | 1 | 8:8770512 | 2 |
| 8:11242025:G:T | 8:11242025 | 8 | 11242025 | G | T | 0.4553 | 2.28e-08 | -0.007269 | 0.001299 | 0.9485 | 8:11235150 | 3 |
| 8:8678530:A:G | 8:8678530 | 8 | 8678530 | A | G | 0.4384 | 2.37e-08 | 0.007207 | 0.00129 | 0.8791 | 8:8700851 | 2 |
| 8:11069960:C:T | 8:11069960 | 8 | 11069960 | T | C | 0.4861 | 2.4e-08 | -0.007195 | 0.001288 | 0.8174 | 8:11080675 | 3 |
| 8:11251175:A:G | 8:11251175 | 8 | 11251175 | G | A | 0.4881 | 2.43e-08 | 0.007353 | 0.001317 | 0.728 | 8:11235150 | 3 |
| 8:11215617:C:G | 8:11215617 | 8 | 11215617 | G | C | 0.4662 | 2.55e-08 | 0.007177 | 0.001288 | 0.8828 | 8:11235150 | 3 |
| 8:10938260:A:G | 8:10938260 | 8 | 10938260 | A | G | 0.4831 | 2.63e-08 | -0.007176 | 0.001289 | 0.7552 | 8:11080675 | 3 |

| uniqID | rsID | chr | pos | non_effect allele | effect_allele | MAF | gwasP | beta | se | r2 | IndSigSNP | GenomicLocus |
|---|---|---|---|---|---|---|---|---|---|---|---|---|
| 8:10987651:G:T | 8:10987651 | 8 | 10987651 | G | T | 0.4264 | 2.65e-08 | -0.007225 | 0.001298 | 1 | 8:10988275 | 3 |
| 8:11055597:A:C | 8:11055597 | 8 | 11055597 | A | C | 0.4871 | 2.67e-08 | -0.007165 | 0.001287 | 0.8699 | 8:11080675 | 3 |
| 8:8685646:C:T | 8:8685646 | 8 | 8685646 | C | T | 0.4533 | 2.69e-08 | 0.007186 | 0.001291 | 0.8977 | 8:8700851 | 2 |
| 8:8673601:A:C | 8:8673601 | 8 | 8673601 | C | A | 0.4433 | 2.76e-08 | 0.007168 | 0.001289 | 0.8755 | 8:8700851 | 2 |
| 8:11214972:A:G | 8:11214972 | 8 | 11214972 | G | A | 0.4831 | 2.8e-08 | 0.007146 | 0.001286 | 0.8372 | 8:11235150 | 3 |
| 8:8639740:A:G | 8:8639740 | 8 | 8639740 | G | A | 0.4553 | 2.85e-08 | -0.007166 | 0.00129 | 0.8631 | 8:8700851 | 2 |
| 8:8704330:C:G | 8:8704330 | 8 | 8704330 | C | G | 0.4394 | 2.87e-08 | 0.007156 | 0.001289 | 0.9602 | 8:8700851 | 2 |
| 8:11219334:A:G | 8:11219334 | 8 | 11219334 | A | G | 0.4662 | 2.87e-08 | 0.007168 | 0.001291 | 0.8824 | 8:11235150 | 3 |
| 8:11214455:A:G | 8:11214455 | 8 | 11214455 | G | A | 0.4831 | 2.89e-08 | 0.00714 | 0.001286 | 0.8372 | 8:11235150 | 3 |
| 8:8682192:A:T | 8:8682192 | 8 | 8682192 | T | A | 0.4374 | 2.91e-08 | 0.007164 | 0.001291 | 0.8752 | 8:8700851 | 2 |
| 8:8679614:C:T | 8:8679614 | 8 | 8679614 | C | T | 0.4414 | 2.92e-08 | 0.00716 | 0.00129 | 0.8826 | 8:8700851 | 2 |
| 8:10987199:C:T | 8:10987199 | 8 | 10987199 | C | T | 0.4274 | 3.08e-08 | -0.007183 | 0.001296 | 0.996 | 8:10988275 | 3 |
| 8:8684953:A:G | 8:8684953 | 8 | 8684953 | A | G | 0.4443 | 3.09e-08 | 0.007165 | 0.001293 | 0.8702 | 8:8700851 | 2 |
| 8:8660538:A:C | 8:8660538 | 8 | 8660538 | A | C | 0.4245 | 3.14e-08 | 0.007136 | 0.001289 | 0.8152 | 8:8700851 | 2 |
| 8:8673736:C:T | 8:8673736 | 8 | 8673736 | C | T | 0.4404 | 3.14e-08 | 0.007136 | 0.001289 | 0.864 | 8:8700851 | 2 |
| 8:11225168:A:G | 8:11225168 | 8 | 11225168 | A | G | 0.4801 | 3.18e-08 | -0.007119 | 0.001286 | 0.7339 | 8:11235150 | 3 |
| 8:8730488:A:G | 8:8730488 | 8 | 8730488 | A | G | 0.4175 | 3.24e-08 | 0.007178 | 0.001297 | 0.8491 | 8:8700851 | 2 |
| 8:11225910:A:G | 8:11225910 | 8 | 11225910 | G | A | 0.4761 | 3.27e-08 | -0.007125 | 0.001288 | 0.7519 | 8:11235150 | 3 |
| 8:8679176:A:G | 8:8679176 | 8 | 8679176 | G | A | 0.4404 | 3.52e-08 | 0.007118 | 0.00129 | 0.8867 | 8:8700851 | 2 |
| 8:8680477:A:G | 8:8680477 | 8 | 8680477 | A | G | 0.4414 | 3.66e-08 | 0.007109 | 0.00129 | 0.8826 | 8:8700851 | 2 |
| 8:11225480:C:T | 8:11225480 | 8 | 11225480 | C | T | 0.4771 | 3.66e-08 | -0.007088 | 0.001286 | 0.7473 | 8:11235150 | 3 |
| 8:8682878:C:T | 8:8682878 | 8 | 8682878 | C | T | 0.4493 | 3.68e-08 | 0.007108 | 0.00129 | 0.9055 | 8:8700851 | 2 |
| 8:11213589:G:T | 8:11213589 | 8 | 11213589 | G | T | 0.4652 | 3.74e-08 | 0.007102 | 0.00129 | 0.8863 | 8:11235150 | 3 |
| 8:9569104:C:T | 8:9569104 | 8 | 9569104 | C | T | 0.169 | 3.84e-08 | 0.009116 | 0.001657 | 1 | 8:9569104 | 2 |
| 8:11070360:C:G | 8:11070360 | 8 | 11070360 | G | C | 0.497 | 3.98e-08 | -0.007068 | 0.001286 | 0.8613 | 8:11080675 | 3 |
| 8:11045161:A:G | 8:11045161 | 8 | 11045161 | A | G | 0.4881 | 4.08e-08 | -0.007055 | 0.001285 | 0.8499 | 8:11080675 | 3 |
| 8:11228672:A:G | 8:11228672 | 8 | 11228672 | A | G | 0.4771 | 4.16e-08 | 0.00705 | 0.001285 | 0.7684 | 8:11235150 | 3 |
| 8:11054097:A:C | 8:11054097 | 8 | 11054097 | A | C | 0.4831 | 4.17e-08 | -0.007068 | 0.001288 | 0.8487 | 8:11080675 | 3 |
| 8:11046209:C:T | 8:11046209 | 8 | 11046209 | C | T | 0.4066 | 4.37e-08 | -0.007142 | 0.001303 | 0.7945 | 8:10988275 | 3 |
| 8:10986837:G:T | 8:10986837 | 8 | 10986837 | G | T | 0.4274 | 4.41e-08 | -0.007109 | 0.001298 | 0.996 | 8:10988275 | 3 |
| 8:11041642:A:T | 8:11041642 | 8 | 11041642 | T | A | 0.4871 | 4.46e-08 | -0.007042 | 0.001286 | 0.86 | 8:11080675 | 3 |
| 8:11252425:A:C | 8:11252425 | 8 | 11252425 | A | C | 0.4423 | 4.46e-08 | 0.00745 | 0.001361 | 0.6595 | 8:11235150 | 3 |
| 8:8644274:C:G | 8:8644274 | 8 | 8644274 | C | G | 0.4483 | 4.53e-08 | 0.007074 | 0.001292 | 0.8794 | 8:8700851 | 2 |
| 8:11425809:G:T | 8:11425809 | 8 | 11425809 | G | T | 0.4861 | 4.79e-08 | -0.007038 | 0.001288 | 1 | 8:11425809 | 3 |
| 8:8685190:A:G | 8:8685190 | 8 | 8685190 | G | A | 0.4453 | 4.81e-08 | 0.00706 | 0.001292 | 0.8743 | 8:8700851 | 2 |
| 8:8654527:C:T | 8:8654527 | 8 | 8654527 | C | T | 0.4304 | 4.91e-08 | 0.007035 | 0.001289 | 0.8356 | 8:8700851 | 2 |
| 8:11039816:G:T | 8:11039816 | 8 | 11039816 | G | T | 0.4851 | 4.94e-08 | -0.007028 | 0.001288 | 0.8529 | 8:11080675 | 3 |
| 8:8643938:C:T | 8:8643938 | 8 | 8643938 | T | C | 0.4483 | 5.01e-08 | 0.007051 | 0.001293 | 0.8794 | 8:8700851 | 2 |
| 8:11228254:A:G | 8:11228254 | 8 | 11228254 | A | G | 0.4791 | 5.05e-08 | -0.007003 | 0.001284 | 0.773 | 8:11235150 | 3 |
| 8:8729761:C:G | 8:8729761 | 8 | 8729761 | C | G | 0.4235 | 5.09e-08 | 0.007064 | 0.001296 | 0.8393 | 8:8700851 | 2 |
| 8:11037187:C:T | 8:11037187 | 8 | 11037187 | C | T | 0.4861 | 5.2e-08 | -0.007005 | 0.001286 | 0.8562 | 8:11080675 | 3 |
| 8:11422289:C:G | 8:11422289 | 8 | 11422289 | C | G | 0.334 | 5.21e-08 | 0.007315 | 0.001343 | 0.9476 | 8:11417582 | 3 |
| 8:11086942:C:T | 8:11086942 | 8 | 11086942 | C | T | 0.495 | 5.29e-08 | -0.007097 | 0.001303 | 0.8902 | 8:11080675 | 3 |
| 8:11247298:A:G | 8:11247298 | 8 | 11247298 | A | G | 0.3887 | 5.38e-08 | 0.007251 | 0.001332 | 0.933 | 8:11249261 | 3 |
| 8:8649881:C:T | 8:8649881 | 8 | 8649881 | T | C | 0.4414 | 5.67e-08 | 0.007025 | 0.001293 | 0.8685 | 8:8700851 | 2 |
| 8:8702875:C:T | 8:8702875 | 8 | 8702875 | C | T | 0.4165 | 5.84e-08 | 0.007028 | 0.001295 | 0.8193 | 8:8700851 | 2 |
| 8:11033976:C:T | 8:11033976 | 8 | 11033976 | C | T | 0.4851 | 6.52e-08 | -0.006967 | 0.001288 | 0.8601 | 8:11080675 | 3 |

| uniqID | rsID | chr | pos | non_effect allele | effect_allele | MAF | gwasP | beta | se | r2 | IndSigSNP | GenomicLocus |
|---|---|---|---|---|---|---|---|---|---|---|---|---|
| 8:8672429:C:G | 8:8672429 | 8 | 8672429 | C | G | 0.4304 | 6.63e-08 | 0.00697 | 0.00129 | 0.8136 | 8:8700851 | 2 |
| 8:8670082:C:G | 8:8670082 | 8 | 8670082 | C | G | 0.4294 | 6.67e-08 | 0.006963 | 0.001289 | 0.817 | 8:8700851 | 2 |
| 8:11223022:A:C | 8:11223022 | 8 | 11223022 | A | C | 0.4851 | 6.75e-08 | 0.006942 | 0.001285 | 0.7911 | 8:11235150 | 3 |
| 8:10985432:C:T | 8:10985432 | 8 | 10985432 | T | C | 0.4771 | 6.77e-08 | -0.006943 | 0.001285 | 0.7835 | 8:11080675 | 3 |
| 8:8671962:C:T | 8:8671962 | 8 | 8671962 | T | C | 0.4284 | 6.86e-08 | 0.006958 | 0.001289 | 0.8211 | 8:8700851 | 2 |
| 8:10121635:A:G | 8:10121635 | 8 | 10121635 | G | A | 0.4414 | 6.97e-08 | 0.007013 | 0.0013 | 0.6089 | 8:10147398 | 3 |
| 8:11245064:C:T | 8:11245064 | 8 | 11245064 | T | C | 0.3897 | 7.04e-08 | -0.007185 | 0.001332 | 0.9057 | 8:11249261 | 3 |
| 8:11235614:A:G | 8:11235614 | 8 | 11235614 | A | G | 0.4841 | 7.33e-08 | -0.006929 | 0.001286 | 0.8086 | 8:11235150 | 3 |
| 8:11415812:C:T | 8:11415812 | 8 | 11415812 | T | C | 0.4911 | 7.39e-08 | -0.006968 | 0.001294 | 0.9588 | 8:11425809 | 3 |
| 8:11427637:A:T | 8:11427637 | 8 | 11427637 | T | A | 0.4901 | 7.66e-08 | -0.00694 | 0.00129 | 0.985 | 8:11425809 | 3 |
| 8:8658540:A:G | 8:8658540 | 8 | 8658540 | G | A | 0.4324 | 7.71e-08 | 0.00693 | 0.001289 | 0.8353 | 8:8700851 | 2 |
| 8:10909193:A:C | 8:10909193 | 8 | 10909193 | A | C | 0.4374 | 7.91e-08 | -0.006971 | 0.001297 | 0.6588 | 8:11080675 | 3 |
| 8:8592845:C:G | 8:8592845 | 8 | 8592845 | G | C | 0.4404 | 8.02e-08 | -0.006916 | 0.001288 | 0.709 | 8:8700851 | 2 |
| 8:11073402:A:G | 8:11073402 | 8 | 11073402 | A | G | 0.498 | 8.03e-08 | -0.006912 | 0.001287 | 0.8645 | 8:11080675 | 3 |
| 8:8769708:C:T | 8:8769708 | 8 | 8769708 | T | C | 0.3996 | 8.08e-08 | 0.007008 | 0.001305 | 0.975 | 8:8770512 | 2 |
| 8:8672579:A:G | 8:8672579 | 8 | 8672579 | G | A | 0.4294 | 8.14e-08 | 0.006921 | 0.001289 | 0.817 | 8:8700851 | 2 |
| 8:10982051:A:G | 8:10982051 | 8 | 10982051 | G | A | 0.4672 | 8.22e-08 | -0.006908 | 0.001287 | 0.8006 | 8:11080675 | 3 |
| 8:8670177:A:T | 8:8670177 | 8 | 8670177 | A | T | 0.4294 | 8.34e-08 | 0.006915 | 0.001289 | 0.817 | 8:8700851 | 2 |
| 8:11238587:A:G | 8:11238587 | 8 | 11238587 | G | A | 0.492 | 8.36e-08 | -0.006905 | 0.001287 | 0.7985 | 8:11235150 | 3 |
| 8:8702827:A:G | 8:8702827 | 8 | 8702827 | G | A | 0.4165 | 8.6e-08 | 0.006942 | 0.001296 | 0.8193 | 8:8700851 | 2 |
| 8:10932203:C:G | 8:10932203 | 8 | 10932203 | C | G | 0.492 | 8.82e-08 | -0.006897 | 0.001288 | 0.7241 | 8:11080675 | 3 |
| 8:11074036:A:G | 8:11074036 | 8 | 11074036 | A | G | 0.499 | 8.95e-08 | -0.00689 | 0.001288 | 0.8683 | 8:11080675 | 3 |
| 8:11420221:A:G | 8:11420221 | 8 | 11420221 | A | G | 0.33 | 9.01e-08 | 0.007202 | 0.001346 | 0.9642 | 8:11417582 | 3 |
| 8:8547811:A:G | 8:8547811 | 8 | 8547811 | G | A | 0.5 | 9.03e-08 | -0.006885 | 0.001287 | 0.674 | 8:8700851 | 2 |
| 8:11425105:G:T | 8:11425105 | 8 | 11425105 | T | G | 0.492 | 9.06e-08 | -0.00689 | 0.001288 | 0.9776 | 8:11425809 | 3 |
| 8:11227104:A:G | 8:11227104 | 8 | 11227104 | G | A | 0.4751 | 9.09e-08 | -0.00687 | 0.001285 | 0.7699 | 8:11235150 | 3 |
| 8:11420104:C:G | 8:11420104 | 8 | 11420104 | G | C | 0.332 | 9.13e-08 | 0.007198 | 0.001346 | 0.964 | 8:11417582 | 3 |
| 8:11223793:G:T | 8:11223793 | 8 | 11223793 | G | T | 0.4801 | 9.29e-08 | -0.006872 | 0.001286 | 0.7906 | 8:11235150 | 3 |
| 8:11244841:A:G | 8:11244841 | 8 | 11244841 | A | G | 0.3678 | 9.37e-08 | -0.007179 | 0.001344 | 0.8324 | 8:11249261 | 3 |
| 8:11224313:C:T | 8:11224313 | 8 | 11224313 | T | C | 0.4811 | 9.68e-08 | 0.006866 | 0.001287 | 0.7801 | 8:11235150 | 3 |
| 8:10910066:C:T | 8:10910066 | 8 | 10910066 | C | T | 0.4364 | 9.88e-08 | 0.006924 | 0.001298 | 0.6556 | 8:11080675 | 3 |
| 8:11421358:A:C | 8:11421358 | 8 | 11421358 | A | C | 0.332 | 9.89e-08 | 0.007175 | 0.001345 | 0.964 | 8:11417582 | 3 |
| 8:8578794:A:T | 8:8578794 | 8 | 8578794 | T | A | 0.4384 | 9.96e-08 | -0.006875 | 0.00129 | 0.7215 | 8:8700851 | 2 |
| 8:8670599:A:G | 8:8670599 | 8 | 8670599 | G | A | 0.4294 | 9.99e-08 | 0.006877 | 0.00129 | 0.817 | 8:8700851 | 2 |
| 8:10986859:C:T | 8:10986859 | 8 | 10986859 | T | C | 0.4771 | 1.01e-07 | -0.006851 | 0.001286 | 0.7835 | 8:11080675 | 3 |
| 8:11411005:C:G | 8:11411005 | 8 | 11411005 | C | G | 0.4553 | 1.01e-07 | -0.006904 | 0.001296 | 0.7482 | 8:11425809 | 3 |
| 8:11420295:C:T | 8:11420295 | 8 | 11420295 | C | T | 0.332 | 1.02e-07 | 0.00718 | 0.001348 | 0.964 | 8:11417582 | 3 |
| 8:11421016:A:G | 8:11421016 | 8 | 11421016 | A | G | 0.332 | 1.02e-07 | 0.007167 | 0.001345 | 0.964 | 8:11417582 | 3 |
| 8:8578811:C:T | 8:8578811 | 8 | 8578811 | C | T | 0.4394 | 1.04e-07 | -0.006864 | 0.00129 | 0.7186 | 8:8700851 | 2 |
| 8:8706332:A:C | 8:8706332 | 8 | 8706332 | C | A | 0.4145 | 1.04e-07 | 0.006928 | 0.001302 | 0.8242 | 8:8700851 | 2 |
| 8:11033525:C:T | 8:11033525 | 8 | 11033525 | T | C | 0.4871 | 1.04e-07 | -0.006866 | 0.00129 | 0.8532 | 8:11080675 | 3 |
| 8:11238316:G:T | 8:11238316 | 8 | 11238316 | G | T | 0.494 | 1.04e-07 | 0.007931 | 0.00149 | 0.7788 | 8:11235150 | 3 |
| 8:11238332:G:T | 8:11238332 | 8 | 11238332 | G | T | 0.4911 | 1.07e-07 | -0.006844 | 0.001287 | 0.79 | 8:11235150 | 3 |
| 8:8692477:C:T | 8:8692477 | 8 | 8692477 | C | T | 0.4205 | 1.08e-07 | 0.006913 | 0.0013 | 0.8382 | 8:8700851 | 2 |
| 8:8548117:A:T | 8:8548117 | 8 | 8548117 | A | T | 0.4702 | 1.1e-07 | -0.006841 | 0.001288 | 0.7618 | 8:8700851 | 2 |
| 8:8670736:A:C | 8:8670736 | 8 | 8670736 | A | C | 0.4264 | 1.1e-07 | 0.006858 | 0.001291 | 0.8062 | 8:8700851 | 2 |
| 8:11421793:C:T | 8:11421793 | 8 | 11421793 | T | C | 0.333 | 1.1e-07 | 0.00715 | 0.001345 | 0.9517 | 8:11417582 | 3 |

| uniqID | rsID | chr | pos | non_effect allele | effect_allele | MAF | gwasP | beta | se | r2 | IndSigSNP | GenomicLocus |
|---|---|---|---|---|---|---|---|---|---|---|---|---|
| 8:8640172:A:C | 8:8640172 | 8 | 8640172 | A | C | 0.4414 | 1.12e-07 | -0.006871 | 0.001294 | 0.8243 | 8:8700851 | 2 |
| 8:10926892:A:C | 8:10926892 | 8 | 10926892 | A | C | 0.498 | 1.13e-07 | -0.00684 | 0.001288 | 0.7111 | 8:11080675 | 3 |
| 8:10975682:C:G | 8:10975682 | 8 | 10975682 | G | C | 0.4722 | 1.16e-07 | -0.006814 | 0.001285 | 0.7977 | 8:11080675 | 3 |
| 8:11040216:C:T | 8:11040216 | 8 | 11040216 | C | T | 0.4105 | 1.16e-07 | -0.006892 | 0.0013 | 0.7853 | 8:10988275 | 3 |
| 8:11251705:A:G | 8:11251705 | 8 | 11251705 | A | G | 0.493 | 1.18e-07 | -0.00695 | 0.001311 | 0.7433 | 8:11235150 | 3 |
| 8:8578120:A:T | 8:8578120 | 8 | 8578120 | A | T | 0.4453 | 1.2e-07 | -0.006842 | 0.001292 | 0.7045 | 8:8700851 | 2 |
| 8:11422130:C:T | 8:11422130 | 8 | 11422130 | C | T | 0.4901 | 1.2e-07 | -0.006846 | 0.001292 | 0.9699 | 8:11425809 | 3 |
| 8:10122423:A:G | 8:10122423 | 8 | 10122423 | G | A | 0.4423 | 1.24e-07 | 0.006867 | 0.001298 | 0.6117 | 8:10147398 | 3 |
| 8:10936811:A:G | 8:10936811 | 8 | 10936811 | G | A | 0.4722 | 1.25e-07 | -0.006817 | 0.001289 | 0.7668 | 8:11080675 | 3 |
| 8:11416171:C:T | 8:11416171 | 8 | 11416171 | C | T | 0.3539 | 1.27e-07 | -0.007027 | 0.001329 | 0.842 | 8:11417582 | 3 |
| 8:10935368:A:C | 8:10935368 | 8 | 10935368 | C | A | 0.4712 | 1.28e-07 | -0.006821 | 0.001291 | 0.7701 | 8:11080675 | 3 |
| 8:10936891:C:T | 8:10936891 | 8 | 10936891 | C | T | 0.4722 | 1.3e-07 | -0.006808 | 0.001289 | 0.7668 | 8:11080675 | 3 |
| 8:11087475:A:G | 8:11087475 | 8 | 11087475 | A | G | 0.4911 | 1.3e-07 | -0.006911 | 0.001308 | 0.8349 | 8:11080675 | 3 |
| 8:11428395:C:T | 8:11428395 | 8 | 11428395 | T | C | 0.4911 | 1.3e-07 | -0.006843 | 0.001295 | 0.9813 | 8:11425809 | 3 |
| 8:11416428:C:T | 8:11416428 | 8 | 11416428 | T | C | 0.3529 | 1.33e-07 | -0.00702 | 0.00133 | 0.8375 | 8:11417582 | 3 |
| 8:11422045:A:G | 8:11422045 | 8 | 11422045 | A | G | 0.4901 | 1.33e-07 | -0.006825 | 0.001293 | 0.9699 | 8:11425809 | 3 |
| 8:11423083:A:C | 8:11423083 | 8 | 11423083 | C | A | 0.492 | 1.34e-07 | -0.006813 | 0.001291 | 0.9776 | 8:11425809 | 3 |
| 8:11416635:A:G | 8:11416635 | 8 | 11416635 | A | G | 0.3529 | 1.35e-07 | -0.007011 | 0.001329 | 0.8375 | 8:11417582 | 3 |
| 8:11421384:C:T | 8:11421384 | 8 | 11421384 | T | C | 0.333 | 1.35e-07 | 0.007098 | 0.001345 | 0.9601 | 8:11417582 | 3 |
| 8:11053922:A:G | 8:11053922 | 8 | 11053922 | A | G | 0.4841 | 1.36e-07 | -0.006812 | 0.001292 | 0.8518 | 8:11080675 | 3 |
| 8:11415794:A:T | 8:11415794 | 8 | 11415794 | A | T | 0.3539 | 1.36e-07 | -0.00701 | 0.001329 | 0.842 | 8:11417582 | 3 |
| 8:8543324:C:T | 8:8543324 | 8 | 8543324 | C | T | 0.496 | 1.37e-07 | -0.006794 | 0.001288 | 0.6662 | 8:8700851 | 2 |
| 8:11417493:C:T | 8:11417493 | 8 | 11417493 | C | T | 0.4513 | 1.37e-07 | 0.00681 | 0.001292 | 0.8183 | 8:11425809 | 3 |
| 8:11417150:G:T | 8:11417150 | 8 | 11417150 | T | G | 0.4771 | 1.39e-07 | -0.006817 | 0.001293 | 0.9226 | 8:11425809 | 3 |
| 8:8595104:A:T | 8:8595104 | 8 | 8595104 | T | A | 0.4414 | 1.43e-07 | -0.00678 | 0.001288 | 0.7187 | 8:8700851 | 2 |
| 8:11415597:C:T | 8:11415597 | 8 | 11415597 | T | C | 0.3539 | 1.44e-07 | -0.006997 | 0.001329 | 0.842 | 8:11417582 | 3 |
| 8:11422936:C:T | 8:11422936 | 8 | 11422936 | T | C | 0.4473 | 1.44e-07 | 0.006787 | 0.001289 | 0.8071 | 8:11425809 | 3 |
| 8:10930069:A:C | 8:10930069 | 8 | 10930069 | C | A | 0.4771 | 1.45e-07 | -0.006794 | 0.001291 | 0.751 | 8:11080675 | 3 |
| 8:10963288:C:G | 8:10963288 | 8 | 10963288 | C | G | 0.4831 | 1.45e-07 | -0.006776 | 0.001288 | 0.7766 | 8:11080675 | 3 |
| 8:11426790:C:G | 8:11426790 | 8 | 11426790 | C | G | 0.4891 | 1.45e-07 | -0.006786 | 0.001289 | 0.9812 | 8:11425809 | 3 |
| 8:8549020:A:C | 8:8549020 | 8 | 8549020 | C | A | 0.5 | 1.47e-07 | -0.006773 | 0.001287 | 0.674 | 8:8700851 | 2 |
| 8:10983534:A:G | 8:10983534 | 8 | 10983534 | G | A | 0.4563 | 1.49e-07 | -0.006766 | 0.001287 | 0.796 | 8:11080675 | 3 |
| 8:11415572:A:G | 8:11415572 | 8 | 11415572 | G | A | 0.3539 | 1.5e-07 | -0.006987 | 0.001329 | 0.842 | 8:11417582 | 3 |
| 8:11422442:A:C | 8:11422442 | 8 | 11422442 | A | C | 0.4901 | 1.51e-07 | -0.006787 | 0.001292 | 0.9699 | 8:11425809 | 3 |
| 8:10979561:C:G | 8:10979561 | 8 | 10979561 | G | C | 0.4632 | 1.55e-07 | -0.006755 | 0.001287 | 0.7817 | 8:11080675 | 3 |
| 8:11422861:C:G | 8:11422861 | 8 | 11422861 | G | C | 0.4473 | 1.55e-07 | 0.006765 | 0.001289 | 0.8132 | 8:11425809 | 3 |
| 8:10935082:C:T | 8:10935082 | 8 | 10935082 | C | T | 0.4712 | 1.56e-07 | -0.006768 | 0.001289 | 0.7701 | 8:11080675 | 3 |
| 8:8598388:A:T | 8:8598388 | 8 | 8598388 | T | A | 0.4463 | 1.63e-07 | -0.006755 | 0.001289 | 0.7427 | 8:8700851 | 2 |
| 8:10960572:C:T | 8:10960572 | 8 | 10960572 | T | C | 0.4841 | 1.68e-07 | -0.006739 | 0.001287 | 0.7795 | 8:11080675 | 3 |
| 8:10962929:C:G | 8:10962929 | 8 | 10962929 | C | G | 0.4801 | 1.74e-07 | -0.006734 | 0.001288 | 0.78 | 8:11080675 | 3 |
| 8:11228006:A:G | 8:11228006 | 8 | 11228006 | A | G | 0.4761 | 1.74e-07 | -0.006708 | 0.001283 | 0.7722 | 8:11235150 | 3 |
| 8:11187434:C:T | 8:11187434 | 8 | 11187434 | C | T | 0.4891 | 1.79e-07 | 0.006721 | 0.001287 | 0.8049 | 8:11235150 | 3 |
| 8:10962800:C:T | 8:10962800 | 8 | 10962800 | C | T | 0.4831 | 1.82e-07 | -0.006723 | 0.001288 | 0.7629 | 8:11080675 | 3 |
| 8:11229889:A:G | 8:11229889 | 8 | 11229889 | G | A | 0.4841 | 1.82e-07 | 0.006827 | 0.001308 | 0.6557 | 8:11235150 | 3 |
| 8:10932695:C:G | 8:10932695 | 8 | 10932695 | C | G | 0.4801 | 1.83e-07 | -0.006733 | 0.00129 | 0.7391 | 8:11080675 | 3 |
| 8:11419852:C:T | 8:11419852 | 8 | 11419852 | C | T | 0.332 | 1.84e-07 | 0.007031 | 0.001347 | 0.964 | 8:11417582 | 3 |
| 8:11186674:A:G | 8:11186674 | 8 | 11186674 | G | A | 0.4881 | 1.86e-07 | 0.006708 | 0.001286 | 0.8014 | 8:11235150 | 3 |

| uniqID | rsID | chr | pos | non_effect allele | effect_allele | MAF | gwasP | beta | se | r2 | IndSigSNP | GenomicLocus |
|---|---|---|---|---|---|---|---|---|---|---|---|---|
| 8:11040647:A:G | 8:11040647 | 8 | 11040647 | G | A | 0.4821 | 1.88e-07 | -0.006713 | 0.001287 | 0.8432 | 8:11080675 | 3 |
| 8:10976569:A:G | 8:10976569 | 8 | 10976569 | A | G | 0.492 | 1.89e-07 | 0.007776 | 0.001492 | 0.7486 | 8:11080675 | 3 |
| 8:11417144:C:T | 8:11417144 | 8 | 11417144 | C | T | 0.4771 | 1.89e-07 | -0.00675 | 0.001295 | 0.9226 | 8:11425809 | 3 |
| 8:11425081:G:T | 8:11425081 | 8 | 11425081 | T | G | 0.3221 | 1.91e-07 | 0.007043 | 0.001351 | 0.8631 | 8:11417582 | 3 |
| 8:10935898:A:G | 8:10935898 | 8 | 10935898 | G | A | 0.4712 | 1.93e-07 | -0.006718 | 0.001289 | 0.7701 | 8:11080675 | 3 |
| 8:11415184:C:G | 8:11415184 | 8 | 11415184 | G | C | 0.3549 | 1.95e-07 | -0.006937 | 0.001332 | 0.8383 | 8:11417582 | 3 |
| 8:11427133:G:T | 8:11427133 | 8 | 11427133 | T | G | 0.4901 | 1.97e-07 | -0.006736 | 0.001294 | 0.985 | 8:11425809 | 3 |
| 8:8768895:A:G | 8:8768895 | 8 | 8768895 | G | A | 0.3976 | 2.01e-07 | 0.006778 | 0.001303 | 0.9753 | 8:8770512 | 2 |
| 8:8578229:A:G | 8:8578229 | 8 | 8578229 | G | A | 0.4414 | 2.02e-07 | -0.006717 | 0.001291 | 0.7187 | 8:8700851 | 2 |
| 8:8578067:C:T | 8:8578067 | 8 | 8578067 | C | T | 0.4394 | 2.04e-07 | -0.006715 | 0.001291 | 0.7186 | 8:8700851 | 2 |
| 8:11189488:C:T | 8:11189488 | 8 | 11189488 | C | T | 0.4861 | 2.05e-07 | 0.007409 | 0.001425 | 0.8141 | 8:11235150 | 3 |
| 8:11423072:A:G | 8:11423072 | 8 | 11423072 | G | A | 0.4483 | 2.05e-07 | 0.006704 | 0.00129 | 0.8165 | 8:11425809 | 3 |
| 8:11238315:C:T | 8:11238315 | 8 | 11238315 | T | C | 0.4751 | 2.07e-07 | 0.007624 | 0.001467 | 0.7524 | 8:11235150 | 3 |
| 8:11417257:A:G | 8:11417257 | 8 | 11417257 | G | A | 0.4781 | 2.11e-07 | -0.006716 | 0.001293 | 0.926 | 8:11425809 | 3 |
| 8:11204165:C:T | 8:11204165 | 8 | 11204165 | T | C | 0.4871 | 2.13e-07 | 0.006677 | 0.001286 | 0.8099 | 8:11235150 | 3 |
| 8:11426400:G:T | 8:11426400 | 8 | 11426400 | G | T | 0.333 | 2.13e-07 | 0.006984 | 0.001345 | 0.9199 | 8:11417582 | 3 |
| 8:11188532:C:G | 8:11188532 | 8 | 11188532 | G | C | 0.4811 | 2.25e-07 | -0.007598 | 0.001467 | 0.8379 | 8:11235150 | 3 |
| 8:10933699:A:T | 8:10933699 | 8 | 10933699 | A | T | 0.4821 | 2.29e-07 | -0.006674 | 0.001289 | 0.7388 | 8:11080675 | 3 |
| 8:11219386:G:T | 8:11219386 | 8 | 11219386 | T | G | 0.4682 | 2.3e-07 | 0.006663 | 0.001287 | 0.6888 | 8:11235150 | 3 |
| 8:10176506:A:T | 8:10176506 | 8 | 10176506 | T | A | 0.3738 | 2.39e-07 | 0.006827 | 0.001321 | 0.7192 | 8:10147398 | 3 |
| 8:11208903:A:G | 8:11208903 | 8 | 11208903 | A | G | 0.4781 | 2.41e-07 | 0.006655 | 0.001288 | 0.8472 | 8:11235150 | 3 |
| 8:11422170:A:G | 8:11422170 | 8 | 11422170 | G | A | 0.332 | 2.44e-07 | -0.006958 | 0.001347 | 0.9478 | 8:11417582 | 3 |
| 8:11207367:A:C | 8:11207367 | 8 | 11207367 | A | C | 0.4881 | 2.45e-07 | 0.006635 | 0.001285 | 0.8205 | 8:11235150 | 3 |
| 8:11031472:C:T | 8:11031472 | 8 | 11031472 | C | T | 0.4612 | 2.48e-07 | -0.006714 | 0.001301 | 0.767 | 8:11080675 | 3 |
| 8:11205602:C:T | 8:11205602 | 8 | 11205602 | C | T | 0.4881 | 2.5e-07 | 0.006647 | 0.001288 | 0.8205 | 8:11235150 | 3 |
| 8:11204755:C:T | 8:11204755 | 8 | 11204755 | C | T | 0.4891 | 2.55e-07 | 0.006629 | 0.001285 | 0.817 | 8:11235150 | 3 |
| 8:10910343:C:G | 8:10910343 | 8 | 10910343 | C | G | 0.4344 | 2.59e-07 | -0.006751 | 0.00131 | 0.6492 | 8:11080675 | 3 |
| 8:11217441:A:G | 8:11217441 | 8 | 11217441 | G | A | 0.4722 | 2.63e-07 | 0.006613 | 0.001284 | 0.6847 | 8:11235150 | 3 |
| 8:11423781:C:G | 8:11423781 | 8 | 11423781 | G | C | 0.3708 | 2.64e-07 | -0.006814 | 0.001323 | 0.8188 | 8:11417582 | 3 |
| 8:11423537:A:G | 8:11423537 | 8 | 11423537 | A | G | 0.325 | 2.68e-07 | 0.00694 | 0.001348 | 0.9053 | 8:11417582 | 3 |
| 8:10979821:A:G | 8:10979821 | 8 | 10979821 | G | A | 0.4622 | 2.73e-07 | -0.007193 | 0.001398 | 0.7851 | 8:11080675 | 3 |
| 8:10935366:C:T | 8:10935366 | 8 | 10935366 | C | T | 0.4712 | 2.81e-07 | -0.006652 | 0.001294 | 0.7701 | 8:11080675 | 3 |
| 8:11252170:C:T | 8:11252170 | 8 | 11252170 | T | C | 0.4632 | 2.87e-07 | -0.00685 | 0.001334 | 0.6717 | 8:11235150 | 3 |
| 8:11418385:A:G | 8:11418385 | 8 | 11418385 | A | G | 0.3211 | 3.01e-07 | 0.006938 | 0.001353 | 0.9296 | 8:11417582 | 3 |
| 8:11425077:C:T | 8:11425077 | 8 | 11425077 | T | C | 0.3549 | 3.03e-07 | 0.006804 | 0.001328 | 0.7754 | 8:11417582 | 3 |
| 8:11032228:G:T | 8:11032228 | 8 | 11032228 | G | T | 0.4751 | 3.19e-07 | -0.006618 | 0.001294 | 0.8184 | 8:11080675 | 3 |
| 8:11216761:G:T | 8:11216761 | 8 | 11216761 | G | T | 0.4692 | 3.29e-07 | 0.006563 | 0.001284 | 0.6963 | 8:11235150 | 3 |
| 8:11186639:C:G | 8:11186639 | 8 | 11186639 | C | G | 0.4801 | 3.4e-07 | 0.006564 | 0.001286 | 0.8344 | 8:11235150 | 3 |
| 8:11221313:C:T | 8:11221313 | 8 | 11221313 | T | C | 0.4702 | 3.41e-07 | 0.006576 | 0.001289 | 0.7315 | 8:11235150 | 3 |
| 8:11186453:C:T | 8:11186453 | 8 | 11186453 | C | T | 0.4801 | 3.44e-07 | 0.006561 | 0.001286 | 0.8344 | 8:11235150 | 3 |
| 8:8687325:A:G | 8:8687325 | 8 | 8687325 | G | A | 0.4732 | 3.52e-07 | -0.006599 | 0.001295 | 0.6821 | 8:8700851 | 2 |
| 8:11211302:G:T | 8:11211302 | 8 | 11211302 | G | T | 0.4742 | 3.52e-07 | 0.006559 | 0.001287 | 0.8394 | 8:11235150 | 3 |
| 8:11423434:A:G | 8:11423434 | 8 | 11423434 | G | A | 0.3231 | 3.55e-07 | 0.006881 | 0.001351 | 0.8973 | 8:11417582 | 3 |
| 8:10970773:A:T | 8:10970773 | 8 | 10970773 | A | T | 0.4761 | 3.59e-07 | -0.006554 | 0.001287 | 0.7814 | 8:11080675 | 3 |
| 8:11199938:A:C | 8:11199938 | 8 | 11199938 | A | C | 0.4801 | 3.66e-07 | 0.00654 | 0.001285 | 0.8409 | 8:11235150 | 3 |
| 8:8581408:C:G | 8:8581408 | 8 | 8581408 | G | C | 0.4453 | 3.67e-07 | -0.006555 | 0.001288 | 0.7123 | 8:8700851 | 2 |
| 8:11213250:C:T | 8:11213250 | 8 | 11213250 | T | C | 0.4672 | 3.69e-07 | 0.006543 | 0.001286 | 0.6858 | 8:11235150 | 3 |

| uniqID | rsID | chr | pos | non_effect allele | effect_allele | MAF | gwasP | beta | se | r2 | IndSigSNP | GenomicLocus |
|---|---|---|---|---|---|---|---|---|---|---|---|---|
| 8:11218893:A:G | 8:11218893 | 8 | 11218893 | G | A | 0.4702 | 3.8e-07 | 0.006532 | 0.001285 | 0.6818 | 8:11235150 | 3 |
| 8:9489417:A:G | 8:9489417 | 8 | 9489417 | A | G | 0.165 | 3.82e-07 | 0.008442 | 0.001661 | 0.8721 | 8:9569104 | 2 |
| 8:8699757:A:T | 8:8699757 | 8 | 8699757 | A | T | 0.4195 | 3.83e-07 | 0.006605 | 0.0013 | 0.8649 | 8:8700851 | 2 |
| 8:11418773:C:T | 8:11418773 | 8 | 11418773 | T | C | 0.3231 | 3.87e-07 | 0.006869 | 0.001353 | 0.9212 | 8:11417582 | 3 |
| 8:10932868:A:G | 8:10932868 | 8 | 10932868 | G | A | 0.4781 | 3.88e-07 | -0.006548 | 0.001289 | 0.7461 | 8:11080675 | 3 |
| 8:8691622:A:T | 8:8691622 | 8 | 8691622 | A | T | 0.4175 | 3.98e-07 | 0.006598 | 0.0013 | 0.8277 | 8:8700851 | 2 |
| 8:8699761:C:T | 8:8699761 | 8 | 8699761 | T | C | 0.4195 | 4.06e-07 | 0.00659 | 0.0013 | 0.8649 | 8:8700851 | 2 |
| 8:8713038:C:T | 8:8713038 | 8 | 8713038 | T | C | 0.3569 | 4.06e-07 | 0.006726 | 0.001327 | 0.6724 | 8:8700851 | 2 |
| 8:11213363:C:T | 8:11213363 | 8 | 11213363 | T | C | 0.4662 | 4.12e-07 | 0.006513 | 0.001285 | 0.6904 | 8:11235150 | 3 |
| 8:11416885:C:T | 8:11416885 | 8 | 11416885 | T | C | 0.4781 | 4.3e-07 | -0.006565 | 0.001298 | 0.926 | 8:11425809 | 3 |
| 8:11237330:C:G | 8:11237330 | 8 | 11237330 | G | C | 0.4046 | 4.35e-07 | 0.007593 | 0.001501 | 0.8843 | 8:11249261 | 3 |
| 8:11213389:C:T | 8:11213389 | 8 | 11213389 | T | C | 0.4662 | 4.44e-07 | 0.006497 | 0.001286 | 0.6904 | 8:11235150 | 3 |
| 8:8589117:C:T | 8:8589117 | 8 | 8589117 | T | C | 0.4404 | 4.54e-07 | -0.007033 | 0.001393 | 0.7147 | 8:8700851 | 2 |
| 8:11434176:A:T | 8:11434176 | 8 | 11434176 | T | A | 0.4473 | 4.54e-07 | -0.00652 | 0.001291 | 0.8331 | 8:11425809 | 3 |
| 8:11466745:A:T | 8:11466745 | 8 | 11466745 | T | A | 0.4592 | 4.59e-07 | -0.006621 | 0.001312 | 0.6355 | 8:11425809 | 3 |
| 8:11435927:C:G | 8:11435927 | 8 | 11435927 | C | G | 0.4473 | 4.73e-07 | -0.006511 | 0.001292 | 0.8395 | 8:11425809 | 3 |
| 8:11434232:C:T | 8:11434232 | 8 | 11434232 | T | C | 0.4473 | 4.74e-07 | -0.00651 | 0.001292 | 0.8331 | 8:11425809 | 3 |
| 8:11435049:A:G | 8:11435049 | 8 | 11435049 | A | G | 0.4473 | 4.78e-07 | -0.006507 | 0.001291 | 0.8325 | 8:11425809 | 3 |
| 8:8696449:G:T | 8:8696449 | 8 | 8696449 | G | T | 0.4225 | 4.86e-07 | 0.006543 | 0.001299 | 0.8377 | 8:8700851 | 2 |
| 8:11250848:A:C | 8:11250848 | 8 | 11250848 | C | A | 0.337 | 4.88e-07 | 0.00692 | 0.001374 | 0.8223 | 8:11249261 | 3 |
| 8:8548801:A:G | 8:8548801 | 8 | 8548801 | A | G | 0.495 | 5.01e-07 | -0.006471 | 0.001287 | 0.6566 | 8:8700851 | 2 |
| 8:11032240:C:T | 8:11032240 | 8 | 11032240 | T | C | 0.4751 | 5.02e-07 | -0.006518 | 0.001296 | 0.8184 | 8:11080675 | 3 |
| 8:10976494:C:G | 8:10976494 | 8 | 10976494 | C | G | 0.4732 | 5.06e-07 | 0.007356 | 0.001463 | 0.8005 | 8:11080675 | 3 |
| 8:11435564:C:T | 8:11435564 | 8 | 11435564 | T | C | 0.4473 | 5.09e-07 | -0.006492 | 0.001292 | 0.8395 | 8:11425809 | 3 |
| 8:10939490:G:T | 8:10939490 | 8 | 10939490 | T | G | 0.4901 | 5.13e-07 | -0.006475 | 0.001289 | 0.6692 | 8:11080675 | 3 |
| 8:11073578:C:T | 8:11073578 | 8 | 11073578 | T | C | 0.4622 | 5.13e-07 | -0.006516 | 0.001297 | 0.85 | 8:11080675 | 3 |
| 8:8544808:C:T | 8:8544808 | 8 | 8544808 | C | T | 0.4891 | 5.18e-07 | -0.006469 | 0.001288 | 0.651 | 8:8700851 | 2 |
| 8:11430485:A:G | 8:11430485 | 8 | 11430485 | A | G | 0.2962 | 5.23e-07 | 0.006922 | 0.001378 | 0.7481 | 8:11417582 | 3 |
| 8:11433909:C:T | 8:11433909 | 8 | 11433909 | C | T | 0.4473 | 5.25e-07 | -0.006483 | 0.001291 | 0.8331 | 8:11425809 | 3 |
| 8:8544872:A:G | 8:8544872 | 8 | 8544872 | A | G | 0.4891 | 5.31e-07 | -0.006463 | 0.001288 | 0.651 | 8:8700851 | 2 |
| 8:11080014:C:T | 8:11080014 | 8 | 11080014 | T | C | 0.4553 | 5.47e-07 | -0.006535 | 0.001304 | 0.8503 | 8:11080675 | 3 |
| 8:11423142:A:G | 8:11423142 | 8 | 11423142 | G | A | 0.3579 | 5.51e-07 | 0.006659 | 0.001329 | 0.7872 | 8:11417582 | 3 |
| 8:8584344:A:G | 8:8584344 | 8 | 8584344 | A | G | 0.4453 | 5.59e-07 | -0.006442 | 0.001286 | 0.7123 | 8:8700851 | 2 |
| 8:11192593:A:G | 8:11192593 | 8 | 11192593 | A | G | 0.4791 | 5.7e-07 | 0.006436 | 0.001286 | 0.844 | 8:11235150 | 3 |
| 8:11432946:A:C | 8:11432946 | 8 | 11432946 | A | C | 0.4483 | 5.74e-07 | -0.006463 | 0.001292 | 0.8358 | 8:11425809 | 3 |
| 8:11427341:G:T | 8:11427341 | 8 | 11427341 | G | T | 0.327 | 5.87e-07 | 0.006715 | 0.001343 | 0.8891 | 8:11417582 | 3 |
| 8:11213092:A:G | 8:11213092 | 8 | 11213092 | G | A | 0.4662 | 5.88e-07 | 0.006428 | 0.001286 | 0.6904 | 8:11235150 | 3 |
| 8:9492453:C:T | 8:9492453 | 8 | 9492453 | C | T | 0.164 | 6.04e-07 | 0.008294 | 0.001661 | 0.8535 | 8:9569104 | 2 |
| 8:11248956:C:T | 8:11248956 | 8 | 11248956 | T | C | 0.338 | 6.14e-07 | 0.006768 | 0.001356 | 0.8397 | 8:11249261 | 3 |
| 8:8587571:G:T | 8:8587571 | 8 | 8587571 | G | T | 0.4761 | 6.23e-07 | -0.006395 | 0.001282 | 0.6111 | 8:8700851 | 2 |
| 8:9525325:C:T | 8:9525325 | 8 | 9525325 | T | C | 0.16 | 6.26e-07 | 0.008354 | 0.001675 | 0.9044 | 8:9569104 | 2 |
| 8:11196970:C:T | 8:11196970 | 8 | 11196970 | T | C | 0.4771 | 6.34e-07 | 0.006417 | 0.001287 | 0.8509 | 8:11235150 | 3 |
| 8:9492426:G:T | 8:9492426 | 8 | 9492426 | G | T | 0.164 | 6.36e-07 | 0.008278 | 0.001661 | 0.8535 | 8:9569104 | 2 |
| 8:8549432:A:G | 8:8549432 | 8 | 8549432 | A | G | 0.495 | 6.47e-07 | -0.00641 | 0.001287 | 0.6631 | 8:8700851 | 2 |
| 8:10148447:A:G | 8:10148447 | 8 | 10148447 | G | A | 0.3171 | 6.47e-07 | 0.006846 | 0.001375 | 0.6166 | 8:10147398 | 3 |
| 8:11213881:C:G | 8:11213881 | 8 | 11213881 | C | G | 0.4662 | 6.51e-07 | 0.006392 | 0.001284 | 0.6904 | 8:11235150 | 3 |
| 8:11435516:C:T | 8:11435516 | 8 | 11435516 | C | T | 0.4384 | 6.63e-07 | -0.006428 | 0.001292 | 0.8134 | 8:11425809 | 3 |

| uniqID | rsID | chr | pos | non_effect allele | effect_allele | MAF | gwasP | beta | se | r2 | IndSigSNP | GenomicLocus |
|---|---|---|---|---|---|---|---|---|---|---|---|---|
| 8:11438064:G:T | 8:11438064 | 8 | 11438064 | T | G | 0.2903 | 6.71e-07 | 0.00689 | 0.001385 | 0.7238 | 8:11417582 | 3 |
| 8:8583872:C:T | 8:8583872 | 8 | 8583872 | C | T | 0.4453 | 6.86e-07 | -0.006392 | 0.001286 | 0.7123 | 8:8700851 | 2 |
| 8:8596731:G:T | 8:8596731 | 8 | 8596731 | T | G | 0.4771 | 6.88e-07 | -0.006382 | 0.001285 | 0.6154 | 8:8700851 | 2 |
| 8:8595838:G:T | 8:8595838 | 8 | 8595838 | G | T | 0.4771 | 6.96e-07 | -0.006376 | 0.001284 | 0.6154 | 8:8700851 | 2 |
| 8:8547110:C:G | 8:8547110 | 8 | 8547110 | C | G | 0.4871 | 7.04e-07 | -0.006391 | 0.001288 | 0.6449 | 8:8700851 | 2 |
| 8:10939273:G:T | 8:10939273 | 8 | 10939273 | T | G | 0.4891 | 7.22e-07 | -0.00639 | 0.001289 | 0.6732 | 8:11080675 | 3 |
| 8:11467557:C:G | 8:11467557 | 8 | 11467557 | G | C | 0.4612 | 7.39e-07 | -0.006544 | 0.001321 | 0.6282 | 8:11425809 | 3 |
| 8:10975081:A:T | 8:10975081 | 8 | 10975081 | A | T | 0.4553 | 7.41e-07 | -0.006367 | 0.001285 | 0.6053 | 8:11080675 | 3 |
| 8:8687054:C:G | 8:8687054 | 8 | 8687054 | C | G | 0.4175 | 7.49e-07 | 0.006463 | 0.001305 | 0.8044 | 8:8700851 | 2 |
| 8:11199584:A:G | 8:11199584 | 8 | 11199584 | G | A | 0.4652 | 8.34e-07 | 0.006362 | 0.00129 | 0.8278 | 8:11235150 | 3 |
| 8:10150070:C:G | 8:10150070 | 8 | 10150070 | C | G | 0.3191 | 8.4e-07 | 0.006772 | 0.001374 | 0.6211 | 8:10147398 | 3 |
| 8:11012977:C:T | 8:11012977 | 8 | 11012977 | T | C | 0.4801 | 8.51e-07 | -0.006351 | 0.001289 | 0.859 | 8:11080675 | 3 |
| 8:9527863:A:T | 8:9527863 | 8 | 9527863 | T | A | 0.16 | 8.67e-07 | 0.008237 | 0.001673 | 0.9044 | 8:9569104 | 2 |
| 8:11433780:C:T | 8:11433780 | 8 | 11433780 | C | T | 0.4384 | 8.69e-07 | -0.006359 | 0.001292 | 0.8072 | 8:11425809 | 3 |
| 8:11015338:C:T | 8:11015338 | 8 | 11015338 | T | C | 0.4781 | 8.8e-07 | -0.006338 | 0.001288 | 0.8593 | 8:11080675 | 3 |
| 8:9527869:A:G | 8:9527869 | 8 | 9527869 | A | G | 0.16 | 8.91e-07 | 0.008229 | 0.001673 | 0.9044 | 8:9569104 | 2 |
| 8:11248500:C:T | 8:11248500 | 8 | 11248500 | T | C | 0.337 | 8.99e-07 | 0.006669 | 0.001357 | 0.8286 | 8:11249261 | 3 |
| 8:11434792:C:G | 8:11434792 | 8 | 11434792 | G | C | 0.4513 | 9.03e-07 | -0.006345 | 0.001291 | 0.8441 | 8:11425809 | 3 |
| 8:11212778:C:T | 8:11212778 | 8 | 11212778 | T | C | 0.4682 | 9.23e-07 | 0.00634 | 0.001291 | 0.6833 | 8:11235150 | 3 |
| 8:8723651:C:G | 8:8723651 | 8 | 8723651 | G | C | 0.3976 | 9.29e-07 | 0.006376 | 0.001299 | 0.7712 | 8:8700851 | 2 |
| 8:11212811:A:C | 8:11212811 | 8 | 11212811 | A | C | 0.4682 | 9.36e-07 | 0.006331 | 0.00129 | 0.6833 | 8:11235150 | 3 |
| 8:11432438:C:G | 8:11432438 | 8 | 11432438 | G | C | 0.3042 | 9.39e-07 | -0.006752 | 0.001376 | 0.7524 | 8:11417582 | 3 |
| 8:11185096:C:G | 8:11185096 | 8 | 11185096 | C | G | 0.4771 | 9.81e-07 | 0.006302 | 0.001286 | 0.8519 | 8:11235150 | 3 |
| 8:11432453:C:T | 8:11432453 | 8 | 11432453 | T | C | 0.2893 | 9.86e-07 | 0.006789 | 0.001386 | 0.7326 | 8:11417582 | 3 |
| 8:11212812:C:G | 8:11212812 | 8 | 11212812 | G | C | 0.4682 | 9.88e-07 | 0.006318 | 0.00129 | 0.6833 | 8:11235150 | 3 |
| 8:11440019:A:G | 8:11440019 | 8 | 11440019 | A | G | 0.4423 | 1.03e-06 | -0.006315 | 0.001292 | 0.8043 | 8:11425809 | 3 |
| 8:11029039:G:T | 8:11029039 | 8 | 11029039 | G | T | 0.4602 | 1.06e-06 | -0.006332 | 0.001297 | 0.8069 | 8:11080675 | 3 |
| 8:10149212:A:C | 8:10149212 | 8 | 10149212 | C | A | 0.3191 | 1.07e-06 | 0.006713 | 0.001375 | 0.6211 | 8:10147398 | 3 |
| 8:8524474:C:G | 8:8524474 | 8 | 8524474 | C | G | 0.497 | 1.11e-06 | -0.006404 | 0.001314 | 0.659 | 8:8700851 | 2 |
| 8:11182455:A:G | 8:11182455 | 8 | 11182455 | G | A | 0.4612 | 1.13e-06 | 0.006786 | 0.001393 | 0.6531 | 8:11235150 | 3 |
| 8:11198792:A:G | 8:11198792 | 8 | 11198792 | G | A | 0.4642 | 1.17e-06 | 0.006274 | 0.00129 | 0.8251 | 8:11235150 | 3 |
| 8:9527707:A:G | 8:9527707 | 8 | 9527707 | A | G | 0.166 | 1.18e-06 | 0.008085 | 0.001663 | 0.9041 | 8:9569104 | 2 |
| 8:11212875:C:G | 8:11212875 | 8 | 11212875 | C | G | 0.4662 | 1.18e-06 | 0.006242 | 0.001284 | 0.6904 | 8:11235150 | 3 |
| 8:11430990:C:T | 8:11430990 | 8 | 11430990 | T | C | 0.2913 | 1.19e-06 | 0.00673 | 0.001385 | 0.7236 | 8:11417582 | 3 |
| 8:11023997:C:G | 8:11023997 | 8 | 11023997 | C | G | 0.4751 | 1.25e-06 | -0.006779 | 0.001397 | 0.8268 | 8:11080675 | 3 |
| 8:9494732:G:T | 8:9494732 | 8 | 9494732 | T | G | 0.171 | 1.28e-06 | -0.007973 | 0.001645 | 0.8485 | 8:9569104 | 2 |
| 8:11212599:C:T | 8:11212599 | 8 | 11212599 | T | C | 0.4662 | 1.35e-06 | 0.006219 | 0.001286 | 0.6904 | 8:11235150 | 3 |
| 8:10975629:A:C | 8:10975629 | 8 | 10975629 | A | C | 0.4553 | 1.39e-06 | -0.006215 | 0.001287 | 0.6066 | 8:11080675 | 3 |
| 8:11177126:A:C | 8:11177126 | 8 | 11177126 | A | C | 0.4483 | 1.47e-06 | 0.006232 | 0.001293 | 0.6867 | 8:11080675 | 3 |
| 8:11461111:A:G | 8:11461111 | 8 | 11461111 | G | A | 0.4652 | 1.47e-06 | -0.006266 | 0.0013 | 0.6446 | 8:11425809 | 3 |
| 8:8724276:C:T | 8:8724276 | 8 | 8724276 | T | C | 0.3777 | 1.52e-06 | 0.00629 | 0.001307 | 0.7285 | 8:8700851 | 2 |
| 8:11014616:G:T | 8:11014616 | 8 | 11014616 | G | T | 0.4911 | 1.52e-06 | -0.006219 | 0.001292 | 0.8526 | 8:11080675 | 3 |
| 8:11240571:C:T | 8:11240571 | 8 | 11240571 | T | C | 0.33 | 1.57e-06 | -0.006579 | 0.001369 | 0.7054 | 8:11249261 | 3 |
| 8:11020313:G:T | 8:11020313 | 8 | 11020313 | T | G | 0.3658 | 1.59e-06 | -0.006382 | 0.001329 | 0.7063 | 8:10988275 | 3 |
| 8:11179458:G:T | 8:11179458 | 8 | 11179458 | T | G | 0.4304 | 1.59e-06 | 0.006241 | 0.001299 | 0.6298 | 8:11235150 | 3 |
| 8:8726804:G:T | 8:8726804 | 8 | 8726804 | T | G | 0.3986 | 1.64e-06 | 0.006243 | 0.001301 | 0.6581 | 8:8700851 | 2 |
| 8:11444516:A:G | 8:11444516 | 8 | 11444516 | A | G | 0.4354 | 1.64e-06 | 0.006207 | 0.001294 | 0.7713 | 8:11425809 | 3 |

| uniqID | rsID | chr | pos | non_effect allele | effect_allele | MAF | gwasP | beta | se | r2 | IndSigSNP | GenomicLocus |
|---|---|---|---|---|---|---|---|---|---|---|---|---|
| 8:11210824:G:T | 8:11210824 | 8 | 11210824 | T | G | 0.4781 | 1.66e-06 | -0.007106 | 0.001482 | 0.8544 | 8:11235150 | 3 |
| 8:11447093:A:G | 8:11447093 | 8 | 11447093 | A | G | 0.4433 | 1.7e-06 | -0.006203 | 0.001295 | 0.8001 | 8:11425809 | 3 |
| 8:11024663:A:C | 8:11024663 | 8 | 11024663 | A | C | 0.5 | 1.72e-06 | -0.00617 | 0.001289 | 0.8565 | 8:11080675 | 3 |
| 8:10987967:A:G | 8:10987967 | 8 | 10987967 | A | G | 0.3708 | 1.78e-06 | -0.00635 | 0.001328 | 0.7845 | 8:10988275 | 3 |
| 8:10989057:A:G | 8:10989057 | 8 | 10989057 | A | G | 0.3688 | 1.82e-06 | -0.006335 | 0.001326 | 0.7785 | 8:10988275 | 3 |
| 8:8724257:C:T | 8:8724257 | 8 | 8724257 | T | C | 0.3787 | 1.85e-06 | 0.006232 | 0.001306 | 0.7309 | 8:8700851 | 2 |
| 8:10974917:C:T | 8:10974917 | 8 | 10974917 | T | C | 0.4543 | 1.85e-06 | -0.006135 | 0.001285 | 0.6034 | 8:11080675 | 3 |
| 8:11410513:A:C | 8:11410513 | 8 | 11410513 | A | C | 0.2982 | 1.85e-06 | 0.006624 | 0.001388 | 0.746 | 8:11417582 | 3 |
| 8:8707197:C:G | 8:8707197 | 8 | 8707197 | G | C | 0.3867 | 1.86e-06 | 0.006245 | 0.001309 | 0.761 | 8:8700851 | 2 |
| 8:8547313:A:C | 8:8547313 | 8 | 8547313 | C | A | 0.4811 | 1.89e-06 | -0.006146 | 0.001289 | 0.6478 | 8:8700851 | 2 |
| 8:11400628:C:G | 8:11400628 | 8 | 11400628 | G | C | 0.4712 | 1.94e-06 | 0.006636 | 0.001393 | 0.6995 | 8:11425809 | 3 |
| 8:11444837:C:T | 8:11444837 | 8 | 11444837 | T | C | 0.4354 | 1.94e-06 | 0.006164 | 0.001294 | 0.7788 | 8:11425809 | 3 |
| 8:11210823:G:T | 8:11210823 | 8 | 11210823 | T | G | 0.4781 | 1.95e-06 | -0.007059 | 0.001482 | 0.8544 | 8:11235150 | 3 |
| 8:10989521:A:G | 8:10989521 | 8 | 10989521 | A | G | 0.3688 | 1.96e-06 | -0.006323 | 0.001328 | 0.7792 | 8:10988275 | 3 |
| 8:8724415:C:T | 8:8724415 | 8 | 8724415 | T | C | 0.3509 | 1.97e-06 | 0.006297 | 0.001323 | 0.6624 | 8:8700851 | 2 |
| 8:10964969:A:T | 8:10964969 | 8 | 10964969 | A | T | 0.4483 | 2.07e-06 | -0.006114 | 0.001287 | 0.6002 | 8:11080675 | 3 |
| 8:11109269:C:T | 8:11109269 | 8 | 11109269 | C | T | 0.4712 | 2.13e-06 | -0.006122 | 0.00129 | 0.7177 | 8:11080675 | 3 |
| 8:10989206:C:T | 8:10989206 | 8 | 10989206 | C | T | 0.3708 | 2.15e-06 | -0.006289 | 0.001326 | 0.7845 | 8:10988275 | 3 |
| 8:11178093:A:C | 8:11178093 | 8 | 11178093 | C | A | 0.4523 | 2.21e-06 | 0.006122 | 0.001292 | 0.6979 | 8:11080675 | 3 |
| 8:10996089:A:G | 8:10996089 | 8 | 10996089 | G | A | 0.3787 | 2.23e-06 | -0.006274 | 0.001325 | 0.7236 | 8:10988275 | 3 |
| 8:8702607:C:G | 8:8702607 | 8 | 8702607 | C | G | 0.4274 | 2.3e-06 | 0.006129 | 0.001296 | 0.7906 | 8:8700851 | 2 |
| 8:8603160:A:G | 8:8603160 | 8 | 8603160 | G | A | 0.4801 | 2.34e-06 | -0.006093 | 0.00129 | 0.6524 | 8:8700851 | 2 |
| 8:11019578:C:G | 8:11019578 | 8 | 11019578 | G | C | 0.4821 | 2.35e-06 | -0.006092 | 0.00129 | 0.8288 | 8:11080675 | 3 |
| 8:8602344:C:T | 8:8602344 | 8 | 8602344 | C | T | 0.4801 | 2.36e-06 | -0.006579 | 0.001393 | 0.6524 | 8:8700851 | 2 |
| 8:10990164:A:G | 8:10990164 | 8 | 10990164 | A | G | 0.3688 | 2.55e-06 | -0.006243 | 0.001326 | 0.7792 | 8:10988275 | 3 |
| 8:11030892:A:G | 8:11030892 | 8 | 11030892 | G | A | 0.3728 | 2.56e-06 | -0.006261 | 0.00133 | 0.7493 | 8:10988275 | 3 |
| 8:10964906:C:T | 8:10964906 | 8 | 10964906 | T | C | 0.4483 | 2.72e-06 | -0.006043 | 0.001287 | 0.6002 | 8:11080675 | 3 |
| 8:10992605:A:C | 8:10992605 | 8 | 10992605 | A | C | 0.3678 | 2.74e-06 | -0.006224 | 0.001326 | 0.7614 | 8:10988275 | 3 |
| 8:11450422:A:G | 8:11450422 | 8 | 11450422 | G | A | 0.4453 | 2.79e-06 | 0.006067 | 0.001294 | 0.7916 | 8:11425809 | 3 |
| 8:11013025:A:C | 8:11013025 | 8 | 11013025 | A | C | 0.3628 | 2.8e-06 | -0.006212 | 0.001325 | 0.72 | 8:10988275 | 3 |
| 8:10992883:C:T | 8:10992883 | 8 | 10992883 | T | C | 0.3688 | 2.87e-06 | -0.00621 | 0.001326 | 0.7648 | 8:10988275 | 3 |
| 8:11188540:A:G | 8:11188540 | 8 | 11188540 | G | A | 0.4553 | 2.99e-06 | 0.006518 | 0.001394 | 0.6432 | 8:11235150 | 3 |
| 8:11206363:C:G | 8:11206363 | 8 | 11206363 | G | C | 0.4602 | 3.01e-06 | 0.006502 | 0.001391 | 0.6457 | 8:11235150 | 3 |
| 8:9568369:A:G | 8:9568369 | 8 | 9568369 | G | A | 0.1849 | 3.11e-06 | 0.00738 | 0.001581 | 0.8689 | 8:9569104 | 2 |
| 8:8725126:G:T | 8:8725126 | 8 | 8725126 | T | G | 0.3787 | 3.34e-06 | 0.00609 | 0.001309 | 0.6306 | 8:8700851 | 2 |
| 8:8722675:C:T | 8:8722675 | 8 | 8722675 | T | C | 0.3946 | 3.44e-06 | 0.006534 | 0.001406 | 0.7762 | 8:8700851 | 2 |
| 8:10990672:C:T | 8:10990672 | 8 | 10990672 | T | C | 0.3678 | 3.51e-06 | -0.006156 | 0.001326 | 0.7694 | 8:10988275 | 3 |
| 8:11024275:C:T | 8:11024275 | 8 | 11024275 | C | T | 0.3757 | 3.69e-06 | -0.006136 | 0.001325 | 0.749 | 8:10988275 | 3 |
| 8:8725229:A:G | 8:8725229 | 8 | 8725229 | A | G | 0.3797 | 3.7e-06 | 0.006064 | 0.001309 | 0.6327 | 8:8700851 | 2 |
| 8:10975733:A:G | 8:10975733 | 8 | 10975733 | G | A | 0.4563 | 3.73e-06 | -0.005973 | 0.00129 | 0.622 | 8:11080675 | 3 |
| 8:10994743:C:G | 8:10994743 | 8 | 10994743 | C | G | 0.3688 | 3.73e-06 | -0.006137 | 0.001326 | 0.7648 | 8:10988275 | 3 |
| 8:11024326:A:C | 8:11024326 | 8 | 11024326 | C | A | 0.4742 | 3.76e-06 | -0.005976 | 0.001291 | 0.8302 | 8:11080675 | 3 |
| 8:10953874:A:G | 8:10953874 | 8 | 10953874 | A | G | 0.493 | 3.89e-06 | -0.005956 | 0.001289 | 0.7438 | 8:11080675 | 3 |
| 8:10969075:C:T | 8:10969075 | 8 | 10969075 | T | C | 0.4463 | 3.89e-06 | -0.005952 | 0.001288 | 0.6089 | 8:11080675 | 3 |
| 8:11016889:A:G | 8:11016889 | 8 | 11016889 | G | A | 0.3738 | 3.9e-06 | -0.00612 | 0.001325 | 0.7518 | 8:10988275 | 3 |
| 8:10983921:C:G | 8:10983921 | 8 | 10983921 | G | C | 0.3847 | 3.96e-06 | -0.006093 | 0.00132 | 0.7155 | 8:10988275 | 3 |
| 8:11176403:A:G | 8:11176403 | 8 | 11176403 | A | G | 0.4523 | 3.97e-06 | 0.005965 | 0.001292 | 0.6979 | 8:11080675 | 3 |

| uniqID | rsID | chr | pos | non_effect allele | effect_allele | MAF | gwasP | beta | se | r2 | IndSigSNP | GenomicLocus |
|---|---|---|---|---|---|---|---|---|---|---|---|---|
| 8:11072020:A:G | 8:11072020 | 8 | 11072020 | G | A | 0.3449 | 4.01e-06 | -0.006161 | 0.001335 | 0.6074 | 8:10988275 | 3 |
| 8:10953092:C:T | 8:10953092 | 8 | 10953092 | C | T | 0.499 | 4.04e-06 | -0.005946 | 0.001289 | 0.7361 | 8:11080675 | 3 |
| 8:11023655:C:T | 8:11023655 | 8 | 11023655 | T | C | 0.3777 | 4.26e-06 | -0.006093 | 0.001324 | 0.7401 | 8:10988275 | 3 |
| 8:11030935:C:T | 8:11030935 | 8 | 11030935 | C | T | 0.4622 | 4.33e-06 | -0.005962 | 0.001297 | 0.7045 | 8:11080675 | 3 |
| 8:11022106:C:G | 8:11022106 | 8 | 11022106 | C | G | 0.3777 | 4.4e-06 | -0.00608 | 0.001323 | 0.733 | 8:10988275 | 3 |
| 8:10841858:A:G | 8:10841858 | 8 | 10841858 | A | G | 0.495 | 4.49e-06 | -0.005928 | 0.001291 | 0.6712 | 8:11080675 | 3 |
| 8:10842659:G:T | 8:10842659 | 8 | 10842659 | G | T | 0.4781 | 4.54e-06 | -0.005937 | 0.001294 | 0.6806 | 8:11080675 | 3 |
| 8:11022185:C:T | 8:11022185 | 8 | 11022185 | C | T | 0.3777 | 4.61e-06 | -0.006067 | 0.001323 | 0.733 | 8:10988275 | 3 |
| 8:8719513:A:G | 8:8719513 | 8 | 8719513 | A | G | 0.3698 | 4.65e-06 | 0.006028 | 0.001315 | 0.7225 | 8:8700851 | 2 |
| 8:11336781:A:G | 8:11336781 | 8 | 11336781 | A | G | 0.4185 | 4.83e-06 | -0.006471 | 0.001414 | 0.6378 | 8:11425809 | 3 |
| 8:11434929:C:T | 8:11434929 | 8 | 11434929 | T | C | 0.2942 | 5.19e-06 | -0.006317 | 0.001385 | 0.7144 | 8:11417582 | 3 |
| 8:8718850:A:G | 8:8718850 | 8 | 8718850 | A | G | 0.3489 | 5.27e-06 | 0.006073 | 0.001333 | 0.6722 | 8:8700851 | 2 |
| 8:9496118:A:T | 8:9496118 | 8 | 9496118 | A | T | 0.173 | 5.45e-06 | -0.007439 | 0.001635 | 0.8368 | 8:9569104 | 2 |
| 8:10839803:C:T | 8:10839803 | 8 | 10839803 | T | C | 0.4891 | 5.48e-06 | -0.005895 | 0.001296 | 0.6636 | 8:11080675 | 3 |
| 8:11033737:A:G | 8:11033737 | 8 | 11033737 | A | G | 0.3499 | 5.62e-06 | -0.006072 | 0.001337 | 0.6316 | 8:10988275 | 3 |
| 8:10993904:C:T | 8:10993904 | 8 | 10993904 | T | C | 0.3877 | 5.72e-06 | -0.005979 | 0.001317 | 0.6905 | 8:10988275 | 3 |
| 8:10950757:C:G | 8:10950757 | 8 | 10950757 | G | C | 0.499 | 5.79e-06 | -0.005844 | 0.001288 | 0.735 | 8:11080675 | 3 |
| 8:10950866:C:T | 8:10950866 | 8 | 10950866 | C | T | 0.499 | 5.96e-06 | -0.005836 | 0.001288 | 0.7342 | 8:11080675 | 3 |
| 8:10957243:A:G | 8:10957243 | 8 | 10957243 | G | A | 0.4891 | 6.39e-06 | -0.005824 | 0.00129 | 0.7823 | 8:11080675 | 3 |
| 8:11392093:A:C | 8:11392093 | 8 | 11392093 | C | A | 0.4652 | 6.5e-06 | -0.005808 | 0.001287 | 0.7079 | 8:11425809 | 3 |
| 8:11309192:C:T | 8:11309192 | 8 | 11309192 | C | T | 0.4433 | 6.59e-06 | -0.005835 | 0.001294 | 0.6216 | 8:11425809 | 3 |
| 8:11038244:A:T | 8:11038244 | 8 | 11038244 | T | A | 0.4404 | 6.62e-06 | -0.005826 | 0.001292 | 0.7113 | 8:11080675 | 3 |
| 8:11043236:C:T | 8:11043236 | 8 | 11043236 | T | C | 0.4394 | 6.85e-06 | -0.005819 | 0.001293 | 0.7102 | 8:11080675 | 3 |
| 8:10955225:C:G | 8:10955225 | 8 | 10955225 | G | C | 0.4901 | 6.96e-06 | -0.005799 | 0.001289 | 0.7501 | 8:11080675 | 3 |
| 8:11034859:A:G | 8:11034859 | 8 | 11034859 | G | A | 0.3499 | 7.07e-06 | -0.005992 | 0.001333 | 0.6316 | 8:10988275 | 3 |
| 8:11037034:A:G | 8:11037034 | 8 | 11037034 | A | G | 0.4543 | 7.22e-06 | -0.005788 | 0.001289 | 0.6969 | 8:11080675 | 3 |
| 8:10958824:C:T | 8:10958824 | 8 | 10958824 | C | T | 0.4881 | 7.38e-06 | -0.005781 | 0.001289 | 0.7854 | 8:11080675 | 3 |
| 8:11065003:A:C | 8:11065003 | 8 | 11065003 | A | C | 0.4284 | 7.5e-06 | -0.005829 | 0.0013 | 0.7054 | 8:11080675 | 3 |
| 8:10944809:G:T | 8:10944809 | 8 | 10944809 | T | G | 0.499 | 7.55e-06 | -0.00577 | 0.001288 | 0.7514 | 8:11080675 | 3 |
| 8:9394053:G:T | 8:9394053 | 8 | 9394053 | G | T | 0.161 | 7.78e-06 | -0.007565 | 0.001691 | 0.6724 | 8:9569104 | 2 |
| 8:10985140:A:G | 8:10985140 | 8 | 10985140 | G | A | 0.3767 | 7.79e-06 | -0.00592 | 0.001323 | 0.8101 | 8:10988275 | 3 |
| 8:10819854:C:T | 8:10819854 | 8 | 10819854 | T | C | 0.4831 | 8.02e-06 | -0.005751 | 0.001287 | 0.6914 | 8:11080675 | 3 |
| 8:10992252:C:G | 8:10992252 | 8 | 10992252 | G | C | 0.3509 | 8.02e-06 | -0.006052 | 0.001355 | 0.7144 | 8:10988275 | 3 |
| 8:11041661:C:T | 8:11041661 | 8 | 11041661 | C | T | 0.3499 | 8.09e-06 | -0.005951 | 0.001333 | 0.6238 | 8:10988275 | 3 |
| 8:11036799:C:G | 8:11036799 | 8 | 11036799 | C | G | 0.3489 | 8.17e-06 | -0.005946 | 0.001332 | 0.6217 | 8:10988275 | 3 |
| 8:9574830:G:T | 8:9574830 | 8 | 9574830 | G | T | 0.1044 | 8.21e-06 | 0.008945 | 0.002004 | 0.6079 | 8:9569104 | 2 |
| 8:10816772:A:G | 8:10816772 | 8 | 10816772 | G | A | 0.4771 | 8.29e-06 | -0.005751 | 0.001289 | 0.6833 | 8:11080675 | 3 |
| 8:11044689:A:G | 8:11044689 | 8 | 11044689 | G | A | 0.3489 | 8.46e-06 | -0.005939 | 0.001333 | 0.6217 | 8:10988275 | 3 |
| 8:11043926:C:T | 8:11043926 | 8 | 11043926 | T | C | 0.4394 | 8.55e-06 | -0.005758 | 0.001293 | 0.7102 | 8:11080675 | 3 |
| 8:9584598:C:G | 8:9584598 | 8 | 9584598 | G | C | 0.1054 | 8.61e-06 | 0.008939 | 0.002008 | 0.6127 | 8:9569104 | 2 |
| 8:9575445:A:G | 8:9575445 | 8 | 9575445 | G | A | 0.1044 | 8.65e-06 | 0.008917 | 0.002003 | 0.6079 | 8:9569104 | 2 |
| 8:10828909:C:T | 8:10828909 | 8 | 10828909 | C | T | 0.4702 | 8.81e-06 | -0.005752 | 0.001293 | 0.7045 | 8:11080675 | 3 |
| 8:11036843:C:T | 8:11036843 | 8 | 11036843 | C | T | 0.3489 | 8.84e-06 | -0.005923 | 0.001332 | 0.6217 | 8:10988275 | 3 |
| 8:9569109:C:T | 8:9569109 | 8 | 9569109 | C | T | 0.1083 | 8.97e-06 | 0.00886 | 0.001994 | 0.6147 | 8:9569104 | 2 |
| 8:11029029:G:T | 8:11029029 | 8 | 11029029 | G | T | 0.492 | 8.99e-06 | -0.005745 | 0.001293 | 0.7123 | 8:11080675 | 3 |
| 8:11043138:C:T | 8:11043138 | 8 | 11043138 | C | T | 0.3489 | 9.03e-06 | -0.005921 | 0.001333 | 0.6217 | 8:10988275 | 3 |
| 8:11038885:A:T | 8:11038885 | 8 | 11038885 | A | T | 0.3499 | 9.5e-06 | -0.005901 | 0.001332 | 0.6185 | 8:10988275 | 3 |

| uniqID | rsID | chr | pos | non_effect allele | effect_allele | MAF | gwasP | beta | se | r2 | IndSigSNP | GenomicLocus |
|---|---|---|---|---|---|---|---|---|---|---|---|---|
| 8:11036919:A:G | 8:11036919 | 8 | 11036919 | A | G | 0.3489 | 9.57e-06 | -0.0059 | 0.001332 | 0.6217 | 8:10988275 | 3 |
| 8:9583872:C:T | 8:9583872 | 8 | 9583872 | T | C | 0.1044 | 9.71e-06 | 0.008893 | 0.002009 | 0.6079 | 8:9569104 | 2 |
| 8:11074812:C:T | 8:11074812 | 8 | 11074812 | T | C | 0.3439 | 1.01e-05 | -0.005909 | 0.001338 | 0.6054 | 8:10988275 | 3 |
| 8:11396856:A:C | 8:11396856 | 8 | 11396856 | C | A | 0.4612 | 1.03e-05 | 0.005674 | 0.001286 | 0.6982 | 8:11425809 | 3 |
| 8:11188586:C:T | 8:11188586 | 8 | 11188586 | C | T | 0.4543 | 1.04e-05 | -0.006441 | 0.00146 | 0.6404 | 8:11235150 | 3 |
| 8:10817197:C:T | 8:10817197 | 8 | 10817197 | T | C | 0.4791 | 1.05e-05 | -0.005685 | 0.001289 | 0.6831 | 8:11080675 | 3 |
| 8:9586062:A:T | 8:9586062 | 8 | 9586062 | A | T | 0.1054 | 1.06e-05 | 0.008869 | 0.002012 | 0.6127 | 8:9569104 | 2 |
| 8:9593309:A:C | 8:9593309 | 8 | 9593309 | A | C | 0.1054 | 1.06e-05 | 0.008907 | 0.002021 | 0.6127 | 8:9569104 | 2 |
| 8:11034028:A:G | 8:11034028 | 8 | 11034028 | G | A | 0.4523 | 1.06e-05 | -0.00569 | 0.001291 | 0.6992 | 8:11080675 | 3 |
| 8:10813904:G:T | 8:10813904 | 8 | 10813904 | T | G | 0.4831 | 1.08e-05 | -0.00568 | 0.00129 | 0.6664 | 8:11080675 | 3 |
| 8:10815754:C:G | 8:10815754 | 8 | 10815754 | G | C | 0.4821 | 1.08e-05 | -0.005678 | 0.00129 | 0.679 | 8:11080675 | 3 |
| 8:10831868:G:T | 8:10831868 | 8 | 10831868 | G | T | 0.4732 | 1.12e-05 | -0.005688 | 0.001294 | 0.6888 | 8:11080675 | 3 |
| 8:11212081:A:G | 8:11212081 | 8 | 11212081 | G | A | 0.4612 | 1.13e-05 | 0.005654 | 0.001287 | 0.6701 | 8:11235150 | 3 |
| 8:10810451:A:G | 8:10810451 | 8 | 10810451 | G | A | 0.4712 | 1.14e-05 | -0.005678 | 0.001293 | 0.6892 | 8:11080675 | 3 |
| 8:11039159:C:G | 8:11039159 | 8 | 11039159 | C | G | 0.3489 | 1.17e-05 | -0.00584 | 0.001332 | 0.6217 | 8:10988275 | 3 |
| 8:11382367:A:G | 8:11382367 | 8 | 11382367 | A | G | 0.4165 | 1.19e-05 | 0.00572 | 0.001305 | 0.656 | 8:11425809 | 3 |
| 8:11446637:A:G | 8:11446637 | 8 | 11446637 | A | G | 0.4016 | 1.24e-05 | -0.006552 | 0.001498 | 0.6574 | 8:11425809 | 3 |
| 8:10813197:C:T | 8:10813197 | 8 | 10813197 | C | T | 0.4801 | 1.28e-05 | -0.005638 | 0.001291 | 0.6828 | 8:11080675 | 3 |
| 8:11187675:C:T | 8:11187675 | 8 | 11187675 | C | T | 0.4553 | 1.3e-05 | -0.006404 | 0.001468 | 0.6432 | 8:11235150 | 3 |
| 8:11021682:A:G | 8:11021682 | 8 | 11021682 | G | A | 0.3777 | 1.33e-05 | -0.005787 | 0.001328 | 0.733 | 8:10988275 | 3 |
| 8:11207672:C:G | 8:11207672 | 8 | 11207672 | G | C | 0.4583 | 1.33e-05 | 0.005619 | 0.001289 | 0.6485 | 8:11235150 | 3 |
| 8:10818657:A:G | 8:10818657 | 8 | 10818657 | G | A | 0.4771 | 1.34e-05 | -0.005612 | 0.001288 | 0.6845 | 8:11080675 | 3 |
| 8:11207431:C:T | 8:11207431 | 8 | 11207431 | T | C | 0.4592 | 1.34e-05 | 0.005597 | 0.001285 | 0.6503 | 8:11235150 | 3 |
| 8:11207508:C:T | 8:11207508 | 8 | 11207508 | C | T | 0.4592 | 1.34e-05 | 0.005598 | 0.001285 | 0.6503 | 8:11235150 | 3 |
| 8:10962099:A:T | 8:10962099 | 8 | 10962099 | T | A | 0.4702 | 1.35e-05 | -0.005623 | 0.001291 | 0.6856 | 8:11080675 | 3 |
| 8:11207326:A:C | 8:11207326 | 8 | 11207326 | A | C | 0.4592 | 1.35e-05 | 0.005596 | 0.001285 | 0.6503 | 8:11235150 | 3 |
| 8:10818607:A:G | 8:10818607 | 8 | 10818607 | G | A | 0.4851 | 1.36e-05 | -0.005608 | 0.001288 | 0.685 | 8:11080675 | 3 |
| 8:11206543:A:G | 8:11206543 | 8 | 11206543 | G | A | 0.4602 | 1.36e-05 | 0.005597 | 0.001286 | 0.6468 | 8:11235150 | 3 |
| 8:11205654:A:T | 8:11205654 | 8 | 11205654 | A | T | 0.4592 | 1.37e-05 | 0.005607 | 0.001289 | 0.6503 | 8:11235150 | 3 |
| 8:11206220:C:G | 8:11206220 | 8 | 11206220 | G | C | 0.4592 | 1.38e-05 | 0.005593 | 0.001286 | 0.6503 | 8:11235150 | 3 |
| 8:11206627:C:G | 8:11206627 | 8 | 11206627 | G | C | 0.4592 | 1.39e-05 | 0.005591 | 0.001286 | 0.6503 | 8:11235150 | 3 |
| 8:11037903:G:T | 8:11037903 | 8 | 11037903 | G | T | 0.3489 | 1.4e-05 | -0.005779 | 0.00133 | 0.6217 | 8:10988275 | 3 |
| 8:10837568:C:T | 8:10837568 | 8 | 10837568 | C | T | 0.4801 | 1.42e-05 | -0.005633 | 0.001297 | 0.6804 | 8:11080675 | 3 |
| 8:11205665:C:G | 8:11205665 | 8 | 11205665 | C | G | 0.4592 | 1.42e-05 | 0.005597 | 0.001289 | 0.6503 | 8:11235150 | 3 |
| 8:11206262:C:T | 8:11206262 | 8 | 11206262 | C | T | 0.4592 | 1.42e-05 | 0.005586 | 0.001286 | 0.6503 | 8:11235150 | 3 |
| 8:11185671:G:T | 8:11185671 | 8 | 11185671 | T | G | 0.4563 | 1.45e-05 | 0.005635 | 0.001299 | 0.6397 | 8:11235150 | 3 |
| 8:10982410:C:G | 8:10982410 | 8 | 10982410 | G | C | 0.3917 | 1.47e-05 | -0.005725 | 0.00132 | 0.7429 | 8:10988275 | 3 |
| 8:10837019:A:G | 8:10837019 | 8 | 10837019 | A | G | 0.4871 | 1.48e-05 | -0.005618 | 0.001296 | 0.6686 | 8:11080675 | 3 |
| 8:10988138:A:C | 8:10988138 | 8 | 10988138 | C | A | 0.3748 | 1.48e-05 | -0.005764 | 0.00133 | 0.8047 | 8:10988275 | 3 |
| 8:10961433:C:T | 8:10961433 | 8 | 10961433 | T | C | 0.4662 | 1.49e-05 | -0.005593 | 0.001291 | 0.6746 | 8:11080675 | 3 |
| 8:10837569:C:G | 8:10837569 | 8 | 10837569 | C | G | 0.4801 | 1.53e-05 | -0.00561 | 0.001297 | 0.6804 | 8:11080675 | 3 |
| 8:10945439:A:G | 8:10945439 | 8 | 10945439 | G | A | 0.493 | 1.53e-05 | -0.005573 | 0.001288 | 0.7365 | 8:11080675 | 3 |
| 8:11204184:A:G | 8:11204184 | 8 | 11204184 | A | G | 0.4592 | 1.54e-05 | 0.005561 | 0.001285 | 0.6503 | 8:11235150 | 3 |
| 8:11109303:C:G | 8:11109303 | 8 | 11109303 | G | C | 0.4612 | 1.56e-05 | -0.005598 | 0.001295 | 0.6974 | 8:11080675 | 3 |
| 8:11209499:C:T | 8:11209499 | 8 | 11209499 | C | T | 0.4612 | 1.58e-05 | 0.005555 | 0.001286 | 0.6562 | 8:11235150 | 3 |
| 8:11056388:C:T | 8:11056388 | 8 | 11056388 | C | T | 0.3439 | 1.59e-05 | -0.005767 | 0.001336 | 0.6015 | 8:10988275 | 3 |
| 8:11205593:C:T | 8:11205593 | 8 | 11205593 | C | T | 0.4592 | 1.59e-05 | 0.005566 | 0.001289 | 0.6503 | 8:11235150 | 3 |

| uniqID | rsID | chr | pos | non_effect allele | effect_allele | MAF | gwasP | beta | se | r2 | IndSigSNP | GenomicLocus |
|---|---|---|---|---|---|---|---|---|---|---|---|---|
| 8:11188752:A:G | 8:11188752 | 8 | 11188752 | A | G | 0.4553 | 1.6e-05 | 0.005555 | 0.001287 | 0.6432 | 8:11235150 | 3 |
| 8:11204532:A:G | 8:11204532 | 8 | 11204532 | G | A | 0.4592 | 1.66e-05 | 0.005537 | 0.001285 | 0.6503 | 8:11235150 | 3 |
| 8:10837414:A:C | 8:10837414 | 8 | 10837414 | C | A | 0.4861 | 1.74e-05 | -0.00557 | 0.001296 | 0.6887 | 8:11080675 | 3 |
| 8:11447119:C:T | 8:11447119 | 8 | 11447119 | T | C | 0.4016 | 1.76e-05 | -0.005611 | 0.001306 | 0.6574 | 8:11425809 | 3 |
| 8:10836024:C:T | 8:10836024 | 8 | 10836024 | C | T | 0.4861 | 1.78e-05 | -0.005567 | 0.001296 | 0.6662 | 8:11080675 | 3 |
| 8:10968926:A:G | 8:10968926 | 8 | 10968926 | A | G | 0.4443 | 1.79e-05 | -0.005543 | 0.001291 | 0.6115 | 8:11080675 | 3 |
| 8:10835917:C:T | 8:10835917 | 8 | 10835917 | T | C | 0.4861 | 1.8e-05 | -0.005559 | 0.001295 | 0.6662 | 8:11080675 | 3 |
| 8:11202960:A:G | 8:11202960 | 8 | 11202960 | A | G | 0.4573 | 1.84e-05 | 0.005506 | 0.001285 | 0.6532 | 8:11235150 | 3 |
| 8:11203107:A:T | 8:11203107 | 8 | 11203107 | T | A | 0.4583 | 1.92e-05 | 0.005498 | 0.001285 | 0.6485 | 8:11235150 | 3 |
| 8:10836069:C:T | 8:10836069 | 8 | 10836069 | T | C | 0.4861 | 1.95e-05 | -0.005535 | 0.001295 | 0.6662 | 8:11080675 | 3 |
| 8:11202154:A:G | 8:11202154 | 8 | 11202154 | A | G | 0.4573 | 2.01e-05 | 0.005482 | 0.001285 | 0.6532 | 8:11235150 | 3 |
| 8:11210983:A:G | 8:11210983 | 8 | 11210983 | G | A | 0.4592 | 2.05e-05 | 0.005482 | 0.001286 | 0.6579 | 8:11235150 | 3 |
| 8:10811829:C:T | 8:10811829 | 8 | 10811829 | C | T | 0.4712 | 2.06e-05 | -0.005503 | 0.001291 | 0.7006 | 8:11080675 | 3 |
| 8:10952500:A:T | 8:10952500 | 8 | 10952500 | T | A | 0.4771 | 2.08e-05 | -0.005494 | 0.00129 | 0.6735 | 8:11080675 | 3 |
| 8:11211068:C:T | 8:11211068 | 8 | 11211068 | C | T | 0.4592 | 2.1e-05 | 0.005475 | 0.001286 | 0.6579 | 8:11235150 | 3 |
| 8:11446868:C:T | 8:11446868 | 8 | 11446868 | C | T | 0.4016 | 2.12e-05 | -0.005558 | 0.001306 | 0.6574 | 8:11425809 | 3 |
| 8:10836508:A:G | 8:10836508 | 8 | 10836508 | A | G | 0.496 | 2.14e-05 | -0.005507 | 0.001295 | 0.6688 | 8:11080675 | 3 |
| 8:11056175:A:G | 8:11056175 | 8 | 11056175 | G | A | 0.3449 | 2.15e-05 | -0.005675 | 0.001335 | 0.6035 | 8:10988275 | 3 |
| 8:11187078:G:T | 8:11187078 | 8 | 11187078 | T | G | 0.4563 | 2.17e-05 | 0.00546 | 0.001285 | 0.6397 | 8:11235150 | 3 |
| 8:11182148:C:G | 8:11182148 | 8 | 11182148 | C | G | 0.4652 | 2.18e-05 | 0.005462 | 0.001286 | 0.6626 | 8:11235150 | 3 |
| 8:10981003:A:C | 8:10981003 | 8 | 10981003 | C | A | 0.4722 | 2.23e-05 | -0.005488 | 0.001294 | 0.7074 | 8:11080675 | 3 |
| 8:8665147:A:G | 8:8665147 | 8 | 8665147 | A | G | 0.4573 | 2.24e-05 | 0.005924 | 0.001396 | 0.6468 | 8:8700851 | 2 |
| 8:10836436:C:T | 8:10836436 | 8 | 10836436 | C | T | 0.4861 | 2.28e-05 | -0.005492 | 0.001296 | 0.6662 | 8:11080675 | 3 |
| 8:11446955:A:G | 8:11446955 | 8 | 11446955 | A | G | 0.4016 | 2.29e-05 | -0.005534 | 0.001306 | 0.6574 | 8:11425809 | 3 |
| 8:11450133:A:G | 8:11450133 | 8 | 11450133 | A | G | 0.3966 | 2.48e-05 | 0.00551 | 0.001306 | 0.6393 | 8:11425809 | 3 |
| 8:10836463:C:G | 8:10836463 | 8 | 10836463 | G | C | 0.4861 | 2.49e-05 | -0.005467 | 0.001296 | 0.6662 | 8:11080675 | 3 |
| 8:11191537:C:T | 8:11191537 | 8 | 11191537 | C | T | 0.4592 | 2.53e-05 | 0.00542 | 0.001286 | 0.6525 | 8:11235150 | 3 |
| 8:11184390:A:C | 8:11184390 | 8 | 11184390 | C | A | 0.4543 | 2.57e-05 | 0.005415 | 0.001286 | 0.6415 | 8:11235150 | 3 |
| 8:11446800:C:T | 8:11446800 | 8 | 11446800 | T | C | 0.4016 | 2.59e-05 | -0.005499 | 0.001306 | 0.6512 | 8:11425809 | 3 |
| 8:8725319:A:G | 8:8725319 | 8 | 8725319 | A | G | 0.3857 | 2.62e-05 | 0.00549 | 0.001305 | 0.6243 | 8:8700851 | 2 |
| 8:11184937:A:G | 8:11184937 | 8 | 11184937 | A | G | 0.4553 | 2.64e-05 | 0.005404 | 0.001285 | 0.6443 | 8:11235150 | 3 |
| 8:11182704:A:G | 8:11182704 | 8 | 11182704 | A | G | 0.4563 | 2.66e-05 | 0.005406 | 0.001286 | 0.6409 | 8:11235150 | 3 |
| 8:11010974:G:T | 8:11010974 | 8 | 11010974 | G | T | 0.4463 | 2.68e-05 | -0.005445 | 0.001296 | 0.6079 | 8:11080675 | 3 |
| 8:11184396:A:G | 8:11184396 | 8 | 11184396 | A | G | 0.4543 | 2.68e-05 | 0.005403 | 0.001286 | 0.6415 | 8:11235150 | 3 |
| 8:11187651:A:G | 8:11187651 | 8 | 11187651 | A | G | 0.4553 | 2.69e-05 | 0.005427 | 0.001292 | 0.6432 | 8:11235150 | 3 |
| 8:10837420:A:G | 8:10837420 | 8 | 10837420 | G | A | 0.4851 | 2.71e-05 | -0.005442 | 0.001296 | 0.6637 | 8:11080675 | 3 |
| 8:8667444:C:T | 8:8667444 | 8 | 8667444 | T | C | 0.4513 | 2.72e-05 | 0.005432 | 0.001294 | 0.6536 | 8:8700851 | 2 |
| 8:11446680:C:T | 8:11446680 | 8 | 11446680 | C | T | 0.4016 | 2.8e-05 | -0.005481 | 0.001307 | 0.6574 | 8:11425809 | 3 |
| 8:10835480:C:T | 8:10835480 | 8 | 10835480 | T | C | 0.4871 | 2.82e-05 | -0.005433 | 0.001297 | 0.6624 | 8:11080675 | 3 |
| 8:10968550:C:G | 8:10968550 | 8 | 10968550 | C | G | 0.4463 | 2.95e-05 | -0.006163 | 0.001474 | 0.6164 | 8:11080675 | 3 |
| 8:11197301:C:G | 8:11197301 | 8 | 11197301 | C | G | 0.4602 | 2.97e-05 | 0.005366 | 0.001284 | 0.6479 | 8:11235150 | 3 |
| 8:11193736:G:T | 8:11193736 | 8 | 11193736 | G | T | 0.4602 | 2.98e-05 | 0.005364 | 0.001284 | 0.6479 | 8:11235150 | 3 |
| 8:8672952:A:C | 8:8672952 | 8 | 8672952 | C | A | 0.4553 | 2.99e-05 | 0.005399 | 0.001293 | 0.6462 | 8:8700851 | 2 |
| 8:11190647:C:G | 8:11190647 | 8 | 11190647 | C | G | 0.4592 | 2.99e-05 | 0.005363 | 0.001284 | 0.6525 | 8:11235150 | 3 |
| 8:11192551:C:G | 8:11192551 | 8 | 11192551 | C | G | 0.4583 | 3.02e-05 | 0.005363 | 0.001285 | 0.6496 | 8:11235150 | 3 |
| 8:8673320:C:T | 8:8673320 | 8 | 8673320 | C | T | 0.4553 | 3.08e-05 | 0.005389 | 0.001292 | 0.6462 | 8:8700851 | 2 |
| 8:8824858:G:T | 8:8824858 | 8 | 8824858 | G | T | 0.4602 | 3.11e-05 | 0.005376 | 0.00129 | 0.6032 | 8:8700851 | 2 |

| uniqID | rsID | chr | pos | non_effect allele | effect_allele | MAF | gwasP | beta | se | r2 | IndSigSNP | GenomicLocus |
|---|---|---|---|---|---|---|---|---|---|---|---|---|
| 8:11197323:G:T | 8:11197323 | 8 | 11197323 | G | T | 0.4602 | 3.12e-05 | 0.005351 | 0.001284 | 0.6479 | 8:11235150 | 3 |
| 8:10836359:C:T | 8:10836359 | 8 | 10836359 | T | C | 0.495 | 3.17e-05 | -0.005395 | 0.001296 | 0.6663 | 8:11080675 | 3 |
| 8:11447679:C:G | 8:11447679 | 8 | 11447679 | C | G | 0.4036 | 3.17e-05 | -0.00544 | 0.001307 | 0.6546 | 8:11425809 | 3 |
| 8:8706209:A:C | 8:8706209 | 8 | 8706209 | C | A | 0.3728 | 3.22e-05 | 0.00552 | 0.001327 | 0.6893 | 8:8700851 | 2 |
| 8:11210828:C:T | 8:11210828 | 8 | 11210828 | C | T | 0.4592 | 3.24e-05 | -0.006162 | 0.001482 | 0.6579 | 8:11235150 | 3 |
| 8:11183505:A:G | 8:11183505 | 8 | 11183505 | A | G | 0.4553 | 3.26e-05 | 0.005351 | 0.001287 | 0.638 | 8:11235150 | 3 |
| 8:11450587:G:T | 8:11450587 | 8 | 11450587 | G | T | 0.3976 | 3.29e-05 | 0.005428 | 0.001306 | 0.641 | 8:11425809 | 3 |
| 8:11355602:C:G | 8:11355602 | 8 | 11355602 | G | C | 0.4632 | 3.34e-05 | -0.005364 | 0.001292 | 0.6064 | 8:11425809 | 3 |
| 8:11194911:A:G | 8:11194911 | 8 | 11194911 | G | A | 0.4602 | 3.36e-05 | 0.005329 | 0.001284 | 0.6479 | 8:11235150 | 3 |
| 8:11401116:A:G | 8:11401116 | 8 | 11401116 | A | G | 0.4801 | 3.37e-05 | -0.005338 | 0.001286 | 0.7035 | 8:11425809 | 3 |
| 8:11196295:C:G | 8:11196295 | 8 | 11196295 | G | C | 0.4702 | 3.43e-05 | 0.00533 | 0.001286 | 0.6224 | 8:11235150 | 3 |
| 8:11439225:A:G | 8:11439225 | 8 | 11439225 | A | G | 0.2793 | 3.56e-05 | 0.005831 | 0.00141 | 0.6106 | 8:11417582 | 3 |
| 8:11358156:C:T | 8:11358156 | 8 | 11358156 | C | T | 0.4692 | 3.61e-05 | -0.005349 | 0.001294 | 0.7135 | 8:11425809 | 3 |
| 8:11186215:A:C | 8:11186215 | 8 | 11186215 | C | A | 0.4602 | 3.62e-05 | 0.005311 | 0.001285 | 0.649 | 8:11235150 | 3 |
| 8:11446421:A:G | 8:11446421 | 8 | 11446421 | A | G | 0.4036 | 3.65e-05 | -0.005401 | 0.001307 | 0.6619 | 8:11425809 | 3 |
| 8:8668917:A:C | 8:8668917 | 8 | 8668917 | C | A | 0.4513 | 3.71e-05 | 0.005335 | 0.001293 | 0.6549 | 8:8700851 | 2 |
| 8:11201504:G:T | 8:11201504 | 8 | 11201504 | G | T | 0.4732 | 3.71e-05 | 0.005313 | 0.001287 | 0.6342 | 8:11235150 | 3 |
| 8:11400680:A:G | 8:11400680 | 8 | 11400680 | A | G | 0.4791 | 3.78e-05 | -0.0053 | 0.001285 | 0.7062 | 8:11425809 | 3 |
| 8:8675176:A:G | 8:8675176 | 8 | 8675176 | G | A | 0.4543 | 3.86e-05 | 0.005323 | 0.001292 | 0.6449 | 8:8700851 | 2 |
| 8:11400944:G:T | 8:11400944 | 8 | 11400944 | T | G | 0.4791 | 3.87e-05 | -0.005302 | 0.001288 | 0.7062 | 8:11425809 | 3 |
| 8:10837190:A:G | 8:10837190 | 8 | 10837190 | G | A | 0.495 | 3.92e-05 | -0.005329 | 0.001295 | 0.6663 | 8:11080675 | 3 |
| 8:11398865:A:G | 8:11398865 | 8 | 11398865 | G | A | 0.4791 | 3.94e-05 | -0.005281 | 0.001284 | 0.7062 | 8:11425809 | 3 |
| 8:11362275:A:C | 8:11362275 | 8 | 11362275 | A | C | 0.4672 | 3.98e-05 | -0.005321 | 0.001294 | 0.7128 | 8:11425809 | 3 |
| 8:8668486:A:G | 8:8668486 | 8 | 8668486 | G | A | 0.4513 | 4.07e-05 | 0.005308 | 0.001293 | 0.6549 | 8:8700851 | 2 |
| 8:11399484:A:T | 8:11399484 | 8 | 11399484 | T | A | 0.4791 | 4.13e-05 | -0.005267 | 0.001284 | 0.7062 | 8:11425809 | 3 |
| 8:11362277:G:T | 8:11362277 | 8 | 11362277 | T | G | 0.4672 | 4.16e-05 | -0.005308 | 0.001294 | 0.7128 | 8:11425809 | 3 |
| 8:8664940:A:G | 8:8664940 | 8 | 8664940 | A | G | 0.4573 | 4.18e-05 | 0.00529 | 0.00129 | 0.6468 | 8:8700851 | 2 |
| 8:11193530:C:T | 8:11193530 | 8 | 11193530 | T | C | 0.4592 | 4.35e-05 | 0.005252 | 0.001284 | 0.6525 | 8:11235150 | 3 |
| 8:10943276:C:T | 8:10943276 | 8 | 10943276 | T | C | 0.4722 | 4.38e-05 | -0.005265 | 0.001287 | 0.6738 | 8:11080675 | 3 |
| 8:11393764:A:G | 8:11393764 | 8 | 11393764 | G | A | 0.4801 | 4.43e-05 | -0.005257 | 0.001287 | 0.6951 | 8:11425809 | 3 |
| 8:11187770:C:G | 8:11187770 | 8 | 11187770 | G | C | 0.4573 | 4.47e-05 | 0.005282 | 0.001293 | 0.6415 | 8:11235150 | 3 |
| 8:11194457:A:G | 8:11194457 | 8 | 11194457 | A | G | 0.4592 | 4.68e-05 | 0.005248 | 0.001288 | 0.6525 | 8:11235150 | 3 |
| 8:11361261:C:G | 8:11361261 | 8 | 11361261 | G | C | 0.4563 | 4.75e-05 | -0.005248 | 0.001289 | 0.6958 | 8:11425809 | 3 |
| 8:11111462:A:T | 8:11111462 | 8 | 11111462 | T | A | 0.4284 | 4.95e-05 | -0.005259 | 0.001295 | 0.6215 | 8:11080675 | 3 |
| 8:11174484:C:G | 8:11174484 | 8 | 11174484 | C | G | 0.4016 | 5.2e-05 | 0.005358 | 0.001323 | 0.6625 | 8:11235150 | 3 |
| 8:11338146:A:G | 8:11338146 | 8 | 11338146 | G | A | 0.4175 | 5.23e-05 | -0.005278 | 0.001304 | 0.6359 | 8:11425809 | 3 |
| 8:11397073:A:C | 8:11397073 | 8 | 11397073 | C | A | 0.4811 | 5.52e-05 | -0.005176 | 0.001283 | 0.6988 | 8:11425809 | 3 |
| 8:8666916:C:T | 8:8666916 | 8 | 8666916 | T | C | 0.4543 | 5.56e-05 | 0.00521 | 0.001292 | 0.6463 | 8:8700851 | 2 |
| 8:11397086:A:T | 8:11397086 | 8 | 11397086 | A | T | 0.4811 | 5.79e-05 | -0.005162 | 0.001283 | 0.6988 | 8:11425809 | 3 |
| 8:11395079:A:G | 8:11395079 | 8 | 11395079 | A | G | 0.4831 | 6.06e-05 | -0.005153 | 0.001284 | 0.6977 | 8:11425809 | 3 |
| 8:11448659:C:G | 8:11448659 | 8 | 11448659 | C | G | 0.3847 | 6.46e-05 | -0.005279 | 0.001321 | 0.6111 | 8:11425809 | 3 |
| 8:11396874:A:G | 8:11396874 | 8 | 11396874 | G | A | 0.4811 | 7.14e-05 | -0.005095 | 0.001282 | 0.6988 | 8:11425809 | 3 |
| 8:10758213:A:G | 8:10758213 | 8 | 10758213 | G | A | 0.4692 | 7.16e-05 | -0.005127 | 0.00129 | 0.6029 | 8:11080675 | 3 |
| 8:8663215:C:T | 8:8663215 | 8 | 8663215 | T | C | 0.4583 | 8.66e-05 | 0.005086 | 0.001295 | 0.6432 | 8:8700851 | 2 |
| 8:11113089:A:G | 8:11113089 | 8 | 11113089 | A | G | 0.4245 | 9.37e-05 | -0.005078 | 0.001299 | 0.6129 | 8:11080675 | 3 |
| 8:11402347:A:G | 8:11402347 | 8 | 11402347 | A | G | 0.4712 | 9.45e-05 | 0.005033 | 0.001288 | 0.6943 | 8:11425809 | 3 |
| 8:11397457:C:G | 8:11397457 | 8 | 11397457 | G | C | 0.4732 | 9.84e-05 | 0.005004 | 0.001284 | 0.6919 | 8:11425809 | 3 |

| uniqID | rsID | chr | pos | non_effect allele | effect_allele | MAF | gwasP | beta | se | r2 | IndSigSNP | GenomicLocus |
|---|---|---|---|---|---|---|---|---|---|---|---|---|
| 8:11398953:C:T | 8:11398953 | 8 | 11398953 | C | T | 0.4851 | 0.000125 | -0.004939 | 0.001287 | 0.6862 | 8:11425809 | 3 |
| 8:9579318:C:T | 8:9579318 | 8 | 9579318 | C | T | 0.1034 | 0.000199 | -0.008522 | 0.00229 | 0.6031 | 8:9569104 | 2 |
| 8:11384556:C:T | 8:11384556 | 8 | 11384556 | T | C | 0.4404 | 0.000201 | 0.004812 | 0.001294 | 0.6251 | 8:11425809 | 3 |
| 8:8709971:C:T | 8:8709971 | 8 | 8709971 | T | C | 0.3569 | 0.000227 | 0.004943 | 0.00134 | 0.6343 | 8:8700851 | 2 |
| 8:8708974:C:G | 8:8708974 | 8 | 8708974 | G | C | 0.3549 | 0.000256 | 0.004875 | 0.001333 | 0.6383 | 8:8700851 | 2 |
| 8:8709756:C:G | 8:8709756 | 8 | 8709756 | C | G | 0.3569 | 0.00042 | 0.004733 | 0.001341 | 0.6355 | 8:8700851 | 2 |
| 8:11237591:A:C | 8:11237591 | 8 | 11237591 | C | A | 0.4592 | 0.000568 | -0.009538 | 0.002766 | 0.9702 | 8:11235150 | 3 |
| 8:10909936:C:T | 8:10909936 | 8 | 10909936 | C | T | 0.4364 | 1,00E-07 | 0.006917 | 0.001298 | 0.6556 | 8:11080675 | 3 |
| 8:11033517:C:G | 8:11033517 | 8 | 11033517 | C | G | 0.4871 | 1,00E-07 | -0.006875 | 0.00129 | 0.8532 | 8:11080675 | 3 |
| 8:11204503:C:G | 8:11204503 | 8 | 11204503 | C | G | 0.4592 | 2,00E-05 | 0.005486 | 0.001285 | 0.6503 | 8:11235150 | 3 |
| 8:11217284:C:T | 8:11217284 | 8 | 11217284 | T | C | 0.4662 | 2,00E-07 | -0.007657 | 0.001472 | 0.8969 | 8:11235150 | 3 |
| 8:11419861:G:T | 8:11419861 | 8 | 11419861 | G | T | 0.332 | 2,00E-07 | 0.007011 | 0.001347 | 0.964 | 8:11417582 | 3 |
| 8:8675325:A:T | 8:8675325 | 8 | 8675325 | T | A | 0.4463 | 3,00E-08 | 0.007152 | 0.00129 | 0.8868 | 8:8700851 | 2 |
| 8:8690787:C:T | 8:8690787 | 8 | 8690787 | T | C | 0.4225 | 5,00E-07 | 0.006535 | 0.001299 | 0.8371 | 8:8700851 | 2 |
| 8:10950396:C:G | 8:10950396 | 8 | 10950396 | G | C | 0.499 | 6,00E-06 | -0.005833 | 0.001288 | 0.735 | 8:11080675 | 3 |
| 8:9572099:A:G | 8:9572099 | 8 | 9572099 | A | G | 0.1044 | 9,00E-06 | 0.008905 | 0.002004 | 0.6079 | 8:9569104 | 2 |
| 8:8545624:T:TTTG | rs141541089 | 8 | 8545624 | TTTG | T | 0.4891 | NA | NA | NA | 0.6446 | 8:8700851 | 2 |
| 8:8546283:A:AAATT | rs3081202 | 8 | 8546283 | A | AAATT | 0.4851 | NA | NA | NA | 0.6479 | 8:8700851 | 2 |
| 8:8582155:A:AAAAAGAAAAG | rs144010740 | 8 | 8582155 | AAAAAGAAAAG | A | 0.4463 | NA | NA | NA | 0.7085 | 8:8700851 | 2 |
| 8:8582155:AAAAAG:AAAAAGAAAAG | rs555499155 | 8 | 8582155 | AAAAAGAAAAG | AAAAAG | 0.03579 | NA | NA | NA | 0.7085 | 8:8700851 | 2 |
| 8:8644213:G:GT | 8:8644213:G:GT | 8 | 8644213 | G | GT | 0.4056 | NA | NA | NA | 0.7681 | 8:8700851 | 2 |
| 8:8644213:G:GTT | 8:8644213:G:GTT | 8 | 8644213 | G | GTT | 0.2932 | NA | NA | NA | 0.7681 | 8:8700851 | 2 |
| 8:8644595:A:AT | rs371642880 | 8 | 8644595 | A | AT | 0.4761 | NA | NA | NA | 0.713 | 8:8700851 | 2 |
| 8:8654057:A:G | rs2409091 | 8 | 8654057 | G | A | 0.4463 | NA | NA | NA | 0.8943 | 8:8700851 | 2 |
| 8:8661026:C:CA | rs34602481 | 8 | 8661026 | CA | C | 0.4533 | NA | NA | NA | 0.8587 | 8:8700851 | 2 |
| 8:8661114:C:CA | rs34140883 | 8 | 8661114 | C | CA | 0.4463 | NA | NA | NA | 0.894 | 8:8700851 | 2 |
| 8:8665802:C:T | rs9644776 | 8 | 8665802 | T | C | 0.4443 | NA | NA | NA | 0.8698 | 8:8700851 | 2 |
| 8:8669681:C:CGTAA | rs34087173 | 8 | 8669681 | CGTAA | C | 0.4304 | NA | NA | NA | 0.8129 | 8:8700851 | 2 |
| 8:8669681:C:T | rs555149467 | 8 | 8669681 | C | T | 0.000994 | NA | NA | NA | 0.8129 | 8:8700851 | 2 |
| 8:8670322:C:CT | rs60073172 | 8 | 8670322 | C | CT | 0.4523 | NA | NA | NA | 0.6562 | 8:8700851 | 2 |
| 8:8676626:G:GAATC | rs5889248 | 8 | 8676626 | GAATC | G | 0.4394 | NA | NA | NA | 0.8675 | 8:8700851 | 2 |
| 8:8679141:C:CTT | rs34362508 | 8 | 8679141 | CTT | C | 0.4404 | NA | NA | NA | 0.8867 | 8:8700851 | 2 |
| 8:8698977:C:CA | rs11435426 | 8 | 8698977 | C | CA | 0.3926 | NA | NA | NA | 0.6914 | 8:8700851 | 2 |
| 8:8703781:G:GGA | rs201075941 | 8 | 8703781 | GGA | G | 0.4056 | NA | NA | NA | 0.7977 | 8:8700851 | 2 |
| 8:8706130:A:AT | rs35147353 | 8 | 8706130 | AT | A | 0.3688 | NA | NA | NA | 0.6796 | 8:8700851 | 2 |
| 8:9579144:C:CT | rs76984329 | 8 | 9579144 | C | CT | 0.1034 | NA | NA | NA | 0.6031 | 8:9569104 | 2 |
| 8:9579377:C:CA | rs67480155 | 8 | 9579377 | CA | C | 0.1044 | NA | NA | NA | 0.6079 | 8:9569104 | 2 |
| 8:10146490:C:CT | rs61082051 | 8 | 10146490 | C | CT | 0.4304 | NA | NA | NA | 0.967 | 8:10147398 | 3 |
| 8:10812333:G:GT | rs34180049 | 8 | 10812333 | G | GT | 0.4911 | NA | NA | NA | 0.6324 | 8:11080675 | 3 |
| 8:10821056:C:CT | rs35470964 | 8 | 10821056 | C | CT | 0.4751 | NA | NA | NA | 0.6285 | 8:11080675 | 3 |
| 8:10927234:C:CAGTA | rs33960758 | 8 | 10927234 | C | CAGTA | 0.498 | NA | NA | NA | 0.7111 | 8:11080675 | 3 |
| 8:10945767:C:CA | rs529577981 | 8 | 10945767 | CA | C | 0.4871 | NA | NA | NA | 0.7576 | 8:11080675 | 3 |
| 8:10948422:C:CA | rs34478855 | 8 | 10948422 | C | CA | 0.499 | NA | NA | NA | 0.7352 | 8:11080675 | 3 |
| 8:10948968:A:G | rs28722721 | 8 | 10948968 | G | A | 0.4861 | NA | NA | NA | 0.6352 | 8:11080675 | 3 |
| 8:10951175:G:GT | rs34623605 | 8 | 10951175 | G | GT | 0.496 | NA | NA | NA | 0.7327 | 8:11080675 | 3 |
| 8:10955383:G:GTT | rs546003990 | 8 | 10955383 | GTT | G | 0.03479 | NA | NA | NA | 0.6515 | 8:11080675 | 3 |
| 8:10955383:GT:GTT | rs559667973 | 8 | 10955383 | GTT | GT | 0.4592 | NA | NA | NA | 0.6515 | 8:11080675 | 3 |

| uniqID | rsID | chr | pos | non_effect allele | effect_allele | MAF | gwasP | beta | se | r2 | IndSigSNP | GenomicLocus |
|---|---|---|---|---|---|---|---|---|---|---|---|---|
| 8:10973149:C:CAA | 8:10973149:C:CAA | 8 | 10973149 | C | CAA | 0.003976 | NA | NA | NA | 0.7777 | 8:11080675 | 3 |
| 8:10973149:C:CA | 8:10973149:C:CA | 8 | 10973149 | CA | C | 0.4662 | NA | NA | NA | 0.7777 | 8:11080675 | 3 |
| 8:10976571:G:T | rs6995805 | 8 | 10976571 | T | G | 0.492 | NA | NA | NA | 0.7486 | 8:11080675 | 3 |
| 8:10978065:G:T | rs7005680 | 8 | 10978065 | T | G | 0.4742 | NA | NA | NA | 0.7957 | 8:11080675 | 3 |
| 8:10983579:G:T | rs60383089 | 8 | 10983579 | G | T | 0.4324 | NA | NA | NA | 0.6899 | 8:11080675 | 3 |
| 8:10990371:G:GA | rs33917510 | 8 | 10990371 | G | GA | 0.4463 | NA | NA | NA | 0.6531 | 8:11080675 | 3 |
| 8:10992544:A:AT | rs35184172 | 8 | 10992544 | AT | A | 0.4781 | NA | NA | NA | 0.6652 | 8:11080675 | 3 |
| 8:10993995:C:T | rs34309263 | 8 | 10993995 | C | T | 0.3688 | NA | NA | NA | 0.7648 | 8:10988275 | 3 |
| 8:11027491:T:TAA | rs113674355 | 8 | 11027491 | T | TAA | 0.4901 | NA | NA | NA | 0.7051 | 8:11080675 | 3 |
| 8:11035071:G:GAGGTCATAATGGAAT | rs113410502 | 8 | 11035071 | GAGGTCATAATGGAAT | G | 0.3509 | NA | NA | NA | 0.6336 | 8:10988275 | 3 |
| 8:11036052:T:TAA | rs35646464 | 8 | 11036052 | TAA | T | 0.3976 | NA | NA | NA | 0.6681 | 8:10988275 | 3 |
| 8:11041897:T:TACACACACACACAC | rs141816060 | 8 | 11041897 | TACACACACACACAC | T | 0.3519 | NA | NA | NA | 0.616 | 8:10988275 | 3 |
| 8:11042974:C:G | 8:11042974:C:G | 8 | 11042974 | G | C | 0.1014 | NA | NA | NA | 0.7771 | 8:10988275 | 3 |
| 8:11042974:G:T | 8:11042974:G:T | 8 | 11042974 | G | T | 0.4125 | NA | NA | NA | 0.7771 | 8:10988275 | 3 |
| 8:11060217:T:TA | rs36080897 | 8 | 11060217 | TA | T | 0.5 | NA | NA | NA | 0.858 | 8:11080675 | 3 |
| 8:11074365:C:CTA | rs36095784 | 8 | 11074365 | CTA | C | 0.4841 | NA | NA | NA | 0.9319 | 8:11080675 | 3 |
| 8:11098992:A:G | rs12334496 | 8 | 11098992 | G | A | 0.4821 | NA | NA | NA | 0.62 | 8:11080675 | 3 |
| 8:11098996:C:T | rs12334549 | 8 | 11098996 | C | T | 0.4801 | NA | NA | NA | 0.6128 | 8:11080675 | 3 |
| 8:11183765:C:CTTA | rs140914421 | 8 | 11183765 | CTTA | C | 0.4553 | NA | NA | NA | 0.638 | 8:11235150 | 3 |
| 8:11184478:C:CAA | rs34820607 | 8 | 11184478 | CAA | C | 0.4632 | NA | NA | NA | 0.6695 | 8:11235150 | 3 |
| 8:11185673:A:AT | rs33915676 | 8 | 11185673 | A | AT | 0.4901 | NA | NA | NA | 0.8021 | 8:11235150 | 3 |
| 8:11189535:C:T | rs12681991 | 8 | 11189535 | T | C | 0.4602 | NA | NA | NA | 0.6479 | 8:11235150 | 3 |
| 8:11197598:C:T | rs6601581 | 8 | 11197598 | T | C | 0.4602 | NA | NA | NA | 0.6479 | 8:11235150 | 3 |
| 8:11198579:C:CTTTT | rs143472078 | 8 | 11198579 | CTTTT | C | 0.4602 | NA | NA | NA | 0.6554 | 8:11235150 | 3 |
| 8:11200454:A:AAT | rs34785978 | 8 | 11200454 | AAT | A | 0.4573 | NA | NA | NA | 0.6415 | 8:11235150 | 3 |
| 8:11201605:A:ATT | rs370296579 | 8 | 11201605 | ATT | A | 0.4901 | NA | NA | NA | 0.7403 | 8:11235150 | 3 |
| 8:11205817:T:TA | rs35291885 | 8 | 11205817 | TA | T | 0.4632 | NA | NA | NA | 0.632 | 8:11235150 | 3 |
| 8:11212650:A:AT | rs3021507 | 8 | 11212650 | A | AT | 0.4662 | NA | NA | NA | 0.6904 | 8:11235150 | 3 |
| 8:11215868:A:AT | rs34369083 | 8 | 11215868 | AT | A | 0.4751 | NA | NA | NA | 0.6261 | 8:11235150 | 3 |
| 8:11220846:C:CAA | rs140232751 | 8 | 11220846 | C | CAA | 0.4364 | NA | NA | NA | 0.6323 | 8:11235150 | 3 |
| 8:11237587:C:CAAAA | rs55683451 | 8 | 11237587 | CAAAA | C | 0.4592 | NA | NA | NA | 0.9702 | 8:11235150 | 3 |
| 8:11239297:G:GATAG | 8:11239297:G:GATAG | 8 | 11239297 | G | GATAG | 0.001988 | NA | NA | NA | 0.9629 | 8:11235150 | 3 |
| 8:11239297:G:GATAT | 8:11239297:G:GATAT | 8 | 11239297 | G | GATAT | 0.4573 | NA | NA | NA | 0.9629 | 8:11235150 | 3 |
| 8:11239942:C:CT | rs35106439 | 8 | 11239942 | C | CT | 0.3628 | NA | NA | NA | 0.6493 | 8:11235150 | 3 |
| 8:11241935:G:GT | rs34637859 | 8 | 11241935 | GT | G | 0.4602 | NA | NA | NA | 0.9515 | 8:11235150 | 3 |
| 8:11242039:T:TA | rs35786152 | 8 | 11242039 | T | TA | 0.4583 | NA | NA | NA | 0.9591 | 8:11235150 | 3 |
| 8:11245562:A:T | rs73534344 | 8 | 11245562 | A | T | 0.3887 | NA | NA | NA | 0.933 | 8:11249261 | 3 |
| 8:11361850:G:GAGGA | rs200525228 | 8 | 11361850 | G | GAGGA | 0.4751 | NA | NA | NA | 0.7138 | 8:11425809 | 3 |
| 8:11417016:A:ACT | rs35482693 | 8 | 11417016 | A | ACT | 0.4791 | NA | NA | NA | 0.9296 | 8:11425809 | 3 |
| 8:11419335:A:AT | rs375515340 | 8 | 11419335 | A | AT | 0.4652 | NA | NA | NA | 0.7555 | 8:11425809 | 3 |
| 8:11422491:A:C | rs7823100 | 8 | 11422491 | A | C | 0.4901 | NA | NA | NA | 0.9699 | 8:11425809 | 3 |
| 8:11422492:C:G | rs7823101 | 8 | 11422492 | G | C | 0.4901 | NA | NA | NA | 0.9699 | 8:11425809 | 3 |
| 8:11422494:A:AT | rs34765125 | 8 | 11422494 | AT | A | 0.4463 | NA | NA | NA | 0.8106 | 8:11425809 | 3 |
| 8:11422521:G:GA | rs34883095 | 8 | 11422521 | GA | G | 0.3718 | NA | NA | NA | 0.8083 | 8:11417582 | 3 |
| 8:11431558:T:TAC | rs35580513 | 8 | 11431558 | T | TAC | 0.4523 | NA | NA | NA | 0.854 | 8:11425809 | 3 |
| 8:11431943:T:TAA | rs560993129 | 8 | 11431943 | T | TAA | 0.3141 | NA | NA | NA | 0.6755 | 8:11417582 | 3 |
| 8:11432085:A:ACG | 8:11432085:A:ACG | 8 | 11432085 | ACG | A | 0.325 | NA | NA | NA | 0.6674 | 8:11417582 | 3 |

| uniqID | rsID | chr | pos | non_effect allele | effect_allele | MAF | gwasP | beta | se | r2 | IndSigSNP | GenomicLocus |
|---|---|---|---|---|---|---|---|---|---|---|---|---|
| 8:11432085:A:ACACG | 8:11432085:A:ACACG | 8 | 11432085 | A | ACACG | 0.000994 | NA | NA | NA | 0.6674 | 8:11417582 | 3 |
| 8:11434415:C:CTCGGTTT | rs111613945 | 8 | 11434415 | CTCGGTTT | C | 0.4423 | NA | NA | NA | 0.79 | 8:11425809 | 3 |
| 8:11435291:G:GTGGC | rs140471203 | 8 | 11435291 | GTGGC | G | 0.4543 | NA | NA | NA | 0.817 | 8:11425809 | 3 |
| 8:11446652:A:G | rs13255662 | 8 | 11446652 | G | A | 0.3946 | NA | NA | NA | 0.6373 | 8:11425809 | 3 |
| 8:11460909:T:TA | rs34394602 | 8 | 11460909 | T | TA | 0.3926 | NA | NA | NA | 0.6175 | 8:11425809 | 3 |
| 12:62989110:A:C | 12:62989110 | 12 | 62989110 | C | A | 0.4761 | 1.5e-08 | -0.00726 | 0.001281 | 1 | 12:62989110 | 4 |
| 12:62995984:C:T | 12:62995984 | 12 | 62995984 | T | C | 0.3598 | 1.52e-08 | -0.008214 | 0.00145 | 0.9667 | 12:62995340 | 4 |
| 12:62986620:A:G | 12:62986620 | 12 | 62986620 | A | G | 0.3588 | 1.57e-08 | -0.008176 | 0.001445 | 0.9693 | 12:62995340 | 4 |
| 12:62990403:A:G | 12:62990403 | 12 | 62990403 | G | A | 0.4761 | 1.6e-08 | -0.007246 | 0.001281 | 1 | 12:62989110 | 4 |
| 12:62990415:C:T | 12:62990415 | 12 | 62990415 | C | T | 0.4761 | 1.61e-08 | -0.007244 | 0.001281 | 1 | 12:62989110 | 4 |
| 12:62995340:A:G | 12:62995340 | 12 | 62995340 | A | G | 0.3519 | 2.23e-08 | -0.007497 | 0.001339 | 1 | 12:62995340 | 4 |
| 12:62945970:C:G | 12:62945970 | 12 | 62945970 | G | C | 0.3549 | 2.38e-08 | -0.007478 | 0.001339 | 0.9868 | 12:62995340 | 4 |
| 12:62977163:A:G | 12:62977163 | 12 | 62977163 | A | G | 0.3549 | 2.41e-08 | -0.007489 | 0.001341 | 0.9868 | 12:62995340 | 4 |
| 12:62995269:A:G | 12:62995269 | 12 | 62995269 | G | A | 0.3539 | 2.41e-08 | -0.007479 | 0.001339 | 0.9911 | 12:62995340 | 4 |
| 12:62968280:C:T | 12:62968280 | 12 | 62968280 | C | T | 0.3539 | 2.5e-08 | -0.007464 | 0.001338 | 0.9911 | 12:62995340 | 4 |
| 12:62948736:A:G | 12:62948736 | 12 | 62948736 | A | G | 0.3519 | 2.58e-08 | -0.007468 | 0.00134 | 1 | 12:62995340 | 4 |
| 12:62985871:A:C | 12:62985871 | 12 | 62985871 | A | C | 0.3539 | 2.59e-08 | -0.007461 | 0.001339 | 0.9911 | 12:62995340 | 4 |
| 12:62950556:C:G | 12:62950556 | 12 | 62950556 | C | G | 0.3539 | 2.66e-08 | -0.007452 | 0.001339 | 0.9911 | 12:62995340 | 4 |
| 12:62942258:C:T | 12:62942258 | 12 | 62942258 | T | C | 0.3519 | 2.83e-08 | -0.007438 | 0.001339 | 1 | 12:62995340 | 4 |
| 12:62960880:A:G | 12:62960880 | 12 | 62960880 | G | A | 0.3579 | 2.98e-08 | -0.007405 | 0.001335 | 0.9737 | 12:62995340 | 4 |
| 12:62961299:C:T | 12:62961299 | 12 | 62961299 | T | C | 0.3519 | 3.05e-08 | -0.007425 | 0.00134 | 1 | 12:62995340 | 4 |
| 12:62957906:C:G | 12:62957906 | 12 | 62957906 | C | G | 0.3579 | 3.25e-08 | -0.007389 | 0.001336 | 0.9737 | 12:62995340 | 4 |
| 12:62996061:A:G | 12:62996061 | 12 | 62996061 | A | G | 0.3519 | 3.29e-08 | -0.007427 | 0.001343 | 1 | 12:62995340 | 4 |
| 12:62993793:A:G | 12:62993793 | 12 | 62993793 | A | G | 0.3519 | 3.31e-08 | -0.007428 | 0.001344 | 1 | 12:62995340 | 4 |
| 12:62952696:C:T | 12:62952696 | 12 | 62952696 | T | C | 0.3539 | 3.33e-08 | -0.007403 | 0.001339 | 0.9911 | 12:62995340 | 4 |
| 12:62949110:A:C | 12:62949110 | 12 | 62949110 | C | A | 0.3569 | 3.38e-08 | -0.007376 | 0.001335 | 0.9779 | 12:62995340 | 4 |
| 12:62961801:A:G | 12:62961801 | 12 | 62961801 | A | G | 0.3559 | 3.42e-08 | -0.007377 | 0.001336 | 0.9823 | 12:62995340 | 4 |
| 12:62975307:C:T | 12:62975307 | 12 | 62975307 | T | C | 0.3539 | 3.48e-08 | -0.007408 | 0.001342 | 0.9911 | 12:62995340 | 4 |
| 12:62937532:C:T | 12:62937532 | 12 | 62937532 | T | C | 0.3618 | 3.52e-08 | -0.007354 | 0.001333 | 0.93 | 12:62995340 | 4 |
| 12:62990871:A:C | 12:62990871 | 12 | 62990871 | A | C | 0.3539 | 3.56e-08 | -0.007389 | 0.00134 | 0.9911 | 12:62995340 | 4 |
| 12:62964552:C:T | 12:62964552 | 12 | 62964552 | C | T | 0.3539 | 3.57e-08 | -0.007429 | 0.001347 | 0.9911 | 12:62995340 | 4 |
| 12:62970482:A:G | 12:62970482 | 12 | 62970482 | G | A | 0.3579 | 3.63e-08 | -0.007376 | 0.001338 | 0.9737 | 12:62995340 | 4 |
| 12:62945245:A:G | 12:62945245 | 12 | 62945245 | A | G | 0.3539 | 3.73e-08 | -0.007386 | 0.001341 | 0.9911 | 12:62995340 | 4 |
| 12:62952910:C:T | 12:62952910 | 12 | 62952910 | T | C | 0.3539 | 3.9e-08 | -0.007379 | 0.001342 | 0.9911 | 12:62995340 | 4 |
| 12:62980580:A:T | 12:62980580 | 12 | 62980580 | A | T | 0.3579 | 4.22e-08 | -0.007328 | 0.001336 | 0.9737 | 12:62995340 | 4 |
| 12:62988288:A:G | 12:62988288 | 12 | 62988288 | A | G | 0.3519 | 4.38e-08 | -0.007354 | 0.001342 | 1 | 12:62995340 | 4 |
| 12:62870574:C:G | 12:62870574 | 12 | 62870574 | G | C | 0.3598 | 4.74e-08 | -0.007274 | 0.001331 | 0.9652 | 12:62995340 | 4 |
| 12:62941837:A:G | 12:62941837 | 12 | 62941837 | G | A | 0.3549 | 4.93e-08 | -0.007297 | 0.001337 | 0.9867 | 12:62995340 | 4 |
| 12:62941426:C:T | 12:62941426 | 12 | 62941426 | C | T | 0.3549 | 4.97e-08 | -0.007294 | 0.001337 | 0.9867 | 12:62995340 | 4 |
| 12:62921257:C:T | 12:62921257 | 12 | 62921257 | T | C | 0.3549 | 5.06e-08 | -0.007284 | 0.001336 | 0.9868 | 12:62995340 | 4 |
| 12:62941444:A:G | 12:62941444 | 12 | 62941444 | G | A | 0.3549 | 5.07e-08 | -0.00729 | 0.001337 | 0.9867 | 12:62995340 | 4 |
| 12:62898111:A:G | 12:62898111 | 12 | 62898111 | A | G | 0.3598 | 5.09e-08 | -0.007258 | 0.001331 | 0.9652 | 12:62995340 | 4 |
| 12:62917704:A:G | 12:62917704 | 12 | 62917704 | G | A | 0.3549 | 5.09e-08 | -0.007282 | 0.001336 | 0.9868 | 12:62995340 | 4 |
| 12:62865152:C:T | 12:62865152 | 12 | 62865152 | T | C | 0.3549 | 5.11e-08 | -0.007293 | 0.001338 | 0.9868 | 12:62995340 | 4 |
| 12:62852271:A:G | 12:62852271 | 12 | 62852271 | A | G | 0.3608 | 5.21e-08 | -0.007248 | 0.00133 | 0.9522 | 12:62995340 | 4 |
| 12:62847085:C:G | 12:62847085 | 12 | 62847085 | G | C | 0.3559 | 5.24e-08 | -0.007287 | 0.001338 | 0.9736 | 12:62995340 | 4 |
| 12:62920860:C:G | 12:62920860 | 12 | 62920860 | G | C | 0.3549 | 5.24e-08 | -0.007275 | 0.001336 | 0.9868 | 12:62995340 | 4 |

| uniqID | rsID | chr | pos | non_effect allele | effect_allele | MAF | gwasP | beta | se | r2 | IndSigSNP | GenomicLocus |
|---|---|---|---|---|---|---|---|---|---|---|---|---|
| 12:62939055:C:T | 12:62939055 | 12 | 62939055 | C | T | 0.3549 | 5.24e-08 | -0.007282 | 0.001337 | 0.9867 | 12:62995340 | 4 |
| 12:62922143:A:G | 12:62922143 | 12 | 62922143 | A | G | 0.3549 | 5.29e-08 | -0.007273 | 0.001336 | 0.9868 | 12:62995340 | 4 |
| 12:62941929:A:T | 12:62941929 | 12 | 62941929 | T | A | 0.3529 | 5.54e-08 | -0.007269 | 0.001337 | 0.9955 | 12:62995340 | 4 |
| 12:62886001:A:T | 12:62886001 | 12 | 62886001 | A | T | 0.3608 | 5.67e-08 | -0.007231 | 0.001331 | 0.9609 | 12:62995340 | 4 |
| 12:62902420:C:T | 12:62902420 | 12 | 62902420 | T | C | 0.3598 | 5.74e-08 | -0.007224 | 0.00133 | 0.9652 | 12:62995340 | 4 |
| 12:62904252:C:T | 12:62904252 | 12 | 62904252 | T | C | 0.3598 | 5.75e-08 | -0.007223 | 0.00133 | 0.9652 | 12:62995340 | 4 |
| 12:62868455:C:T | 12:62868455 | 12 | 62868455 | T | C | 0.3549 | 5.77e-08 | -0.007264 | 0.001338 | 0.9868 | 12:62995340 | 4 |
| 12:62884972:A:G | 12:62884972 | 12 | 62884972 | A | G | 0.3598 | 5.86e-08 | -0.007222 | 0.001331 | 0.9652 | 12:62995340 | 4 |
| 12:62932816:A:G | 12:62932816 | 12 | 62932816 | G | A | 0.3549 | 5.86e-08 | -0.007249 | 0.001336 | 0.9868 | 12:62995340 | 4 |
| 12:62888219:A:C | 12:62888219 | 12 | 62888219 | C | A | 0.3608 | 5.91e-08 | -0.00722 | 0.001331 | 0.9609 | 12:62995340 | 4 |
| 12:62930798:C:T | 12:62930798 | 12 | 62930798 | T | C | 0.3549 | 5.93e-08 | -0.007246 | 0.001336 | 0.9868 | 12:62995340 | 4 |
| 12:62865291:A:G | 12:62865291 | 12 | 62865291 | A | G | 0.3549 | 5.94e-08 | -0.007255 | 0.001337 | 0.9868 | 12:62995340 | 4 |
| 12:62880276:C:G | 12:62880276 | 12 | 62880276 | G | C | 0.3598 | 6.07e-08 | -0.007214 | 0.001331 | 0.9652 | 12:62995340 | 4 |
| 12:62858342:A:G | 12:62858342 | 12 | 62858342 | A | G | 0.3539 | 6.11e-08 | -0.00725 | 0.001338 | 0.9822 | 12:62995340 | 4 |
| 12:62868500:A:C | 12:62868500 | 12 | 62868500 | C | A | 0.3549 | 6.19e-08 | -0.007249 | 0.001338 | 0.9868 | 12:62995340 | 4 |
| 12:62898490:C:T | 12:62898490 | 12 | 62898490 | T | C | 0.3598 | 6.25e-08 | -0.007207 | 0.001331 | 0.9652 | 12:62995340 | 4 |
| 12:62903639:A:G | 12:62903639 | 12 | 62903639 | G | A | 0.3598 | 6.25e-08 | -0.007203 | 0.00133 | 0.9652 | 12:62995340 | 4 |
| 12:62897930:A:G | 12:62897930 | 12 | 62897930 | A | G | 0.3598 | 6.28e-08 | -0.007206 | 0.001331 | 0.9652 | 12:62995340 | 4 |
| 12:62903793:A:G | 12:62903793 | 12 | 62903793 | G | A | 0.3598 | 6.33e-08 | -0.0072 | 0.00133 | 0.9652 | 12:62995340 | 4 |
| 12:62858561:C:T | 12:62858561 | 12 | 62858561 | T | C | 0.3549 | 6.44e-08 | -0.007237 | 0.001338 | 0.9868 | 12:62995340 | 4 |
| 12:62875839:C:T | 12:62875839 | 12 | 62875839 | T | C | 0.3598 | 6.53e-08 | -0.007197 | 0.001331 | 0.9652 | 12:62995340 | 4 |
| 12:62894058:A:G | 12:62894058 | 12 | 62894058 | G | A | 0.3598 | 6.59e-08 | -0.007194 | 0.001331 | 0.9652 | 12:62995340 | 4 |
| 12:62926398:A:G | 12:62926398 | 12 | 62926398 | A | G | 0.3549 | 6.74e-08 | -0.007232 | 0.001339 | 0.9868 | 12:62995340 | 4 |
| 12:62862739:G:T | 12:62862739 | 12 | 62862739 | G | T | 0.3608 | 6.77e-08 | -0.007213 | 0.001335 | 0.9609 | 12:62995340 | 4 |
| 12:62890491:A:C | 12:62890491 | 12 | 62890491 | A | C | 0.3598 | 6.78e-08 | -0.007187 | 0.001331 | 0.9652 | 12:62995340 | 4 |
| 12:62848152:A:G | 12:62848152 | 12 | 62848152 | A | G | 0.3559 | 7.19e-08 | -0.007229 | 0.001341 | 0.9736 | 12:62995340 | 4 |
| 12:62855388:C:T | 12:62855388 | 12 | 62855388 | T | C | 0.3588 | 7.46e-08 | -0.007177 | 0.001333 | 0.9606 | 12:62995340 | 4 |
| 12:62852916:A:G | 12:62852916 | 12 | 62852916 | A | G | 0.3588 | 7.48e-08 | -0.007171 | 0.001332 | 0.9606 | 12:62995340 | 4 |
| 12:62910714:A:T | 12:62910714 | 12 | 62910714 | T | A | 0.3598 | 7.5e-08 | -0.00716 | 0.00133 | 0.9652 | 12:62995340 | 4 |
| 12:62935705:C:T | 12:62935705 | 12 | 62935705 | T | C | 0.3549 | 7.53e-08 | -0.007206 | 0.001339 | 0.9868 | 12:62995340 | 4 |
| 12:62889409:A:G | 12:62889409 | 12 | 62889409 | A | G | 0.3598 | 7.62e-08 | -0.007189 | 0.001336 | 0.9652 | 12:62995340 | 4 |
| 12:62906488:A:G | 12:62906488 | 12 | 62906488 | A | G | 0.3598 | 7.62e-08 | -0.007157 | 0.00133 | 0.9652 | 12:62995340 | 4 |
| 12:62889402:A:G | 12:62889402 | 12 | 62889402 | G | A | 0.3598 | 8.05e-08 | -0.007179 | 0.001337 | 0.9652 | 12:62995340 | 4 |
| 12:62928633:A:G | 12:62928633 | 12 | 62928633 | A | G | 0.3549 | 8.34e-08 | -0.007167 | 0.001336 | 0.9868 | 12:62995340 | 4 |
| 12:62886649:C:T | 12:62886649 | 12 | 62886649 | T | C | 0.3598 | 9.77e-08 | -0.00712 | 0.001335 | 0.9652 | 12:62995340 | 4 |
| 12:62849418:A:G | 12:62849418 | 12 | 62849418 | A | G | 0.3579 | 1.03e-07 | -0.007113 | 0.001336 | 0.9649 | 12:62995340 | 4 |
| 12:62868499:C:T | 12:62868499 | 12 | 62868499 | C | T | 0.3549 | 1.11e-07 | 0.008098 | 0.001524 | 0.9868 | 12:62995340 | 4 |
| 12:62886372:C:G | 12:62886372 | 12 | 62886372 | C | G | 0.3598 | 1.29e-07 | 0.007999 | 0.001514 | 0.9652 | 12:62995340 | 4 |
| 12:62999154:C:G | 12:62999154 | 12 | 62999154 | G | C | 0.3936 | 2.57e-07 | -0.006765 | 0.001312 | 0.7675 | 12:62995340 | 4 |
| 12:62997180:C:T | 12:62997180 | 12 | 62997180 | C | T | 0.4016 | 7.64e-07 | -0.006461 | 0.001306 | 0.7981 | 12:62995340 | 4 |
| 12:62937348:G:T | 12:62937348 | 12 | 62937348 | G | T | 0.3887 | 1.62e-06 | -0.006345 | 0.001322 | 0.6864 | 12:62995340 | 4 |
| 12:62837767:A:G | 12:62837767 | 12 | 62837767 | A | G | 0.3002 | 6.11e-05 | -0.005533 | 0.001379 | 0.6756 | 12:62995340 | 4 |
| 12:62831343:C:T | 12:62831343 | 12 | 62831343 | C | T | 0.3022 | 8.01e-05 | -0.005448 | 0.00138 | 0.6739 | 12:62995340 | 4 |
| 12:62830952:C:T | 12:62830952 | 12 | 62830952 | T | C | 0.3022 | 9.57e-05 | -0.005377 | 0.001378 | 0.6739 | 12:62995340 | 4 |
| 12:62977162:A:G | 12:62977162 | 12 | 62977162 | A | G | 0.3549 | 1,00E-07 | 0.00813 | 0.001525 | 0.9868 | 12:62995340 | 4 |
| 12:62930621:A:G | 12:62930621 | 12 | 62930621 | A | G | 0.3549 | 6,00E-08 | -0.007243 | 0.001336 | 0.9868 | 12:62995340 | 4 |
| 12:62851080:A:AC | rs36039527 | 12 | 62851080 | AC | A | 0.3608 | NA | NA | NA | 0.9522 | 12:62995340 | 4 |

| uniqID | rsID | chr | pos | non_effect allele | effect_allele | MAF | gwasP | beta | se | r2 | IndSigSNP | GenomicLocus |
|---|---|---|---|---|---|---|---|---|---|---|---|---|
| 12:62859241:A:AC | rs35579550 | 12 | 62859241 | AC | A | 0.3549 | NA | NA | NA | 0.9779 | 12:62995340 | 4 |
| 12:62861935:T:TA | rs35738119 | 12 | 62861935 | TA | T | 0.4165 | NA | NA | NA | 0.7579 | 12:62995340 | 4 |
| 12:62884714:A:ATTT | rs34591945 | 12 | 62884714 | ATTT | A | 0.3598 | NA | NA | NA | 0.9652 | 12:62995340 | 4 |
| 12:62891828:C:CT | rs34160326 | 12 | 62891828 | CT | C | 0.3787 | NA | NA | NA | 0.8617 | 12:62995340 | 4 |
| 12:62898463:C:CT | rs75092735 | 12 | 62898463 | C | CT | 0.3638 | NA | NA | NA | 0.9485 | 12:62995340 | 4 |
| 12:62903655:G:GT | rs574068633 | 12 | 62903655 | G | GT | 0.3698 | NA | NA | NA | 0.915 | 12:62995340 | 4 |
| 12:62905380:C:CA | rs35153665 | 12 | 62905380 | CA | C | 0.3598 | NA | NA | NA | 0.9652 | 12:62995340 | 4 |
| 12:62905772:A:AC | rs35759179 | 12 | 62905772 | A | AC | 0.3598 | NA | NA | NA | 0.9652 | 12:62995340 | 4 |
| 12:62914776:T:TA | rs71086606 | 12 | 62914776 | TA | T | 0.3519 | NA | NA | NA | 1 | 12:62995340 | 4 |
| 12:62928006:T:TA | rs545076659 | 12 | 62928006 | T | TA | 0.3529 | NA | NA | NA | 0.9598 | 12:62995340 | 4 |
| 12:62931846:A:ATTTGT | rs147032464 | 12 | 62931846 | ATTTGT | A | 0.3549 | NA | NA | NA | 0.9868 | 12:62995340 | 4 |
| 12:62935154:C:CA | rs531553553 | 12 | 62935154 | CA | C | 0.3897 | NA | NA | NA | 0.7746 | 12:62995340 | 4 |
| 12:62945158:G:GGAAACAACCTGACGAT | rs113510588 | 12 | 62945158 | GGAAACAACCTGACGAT | G | 0.3539 | NA | NA | NA | 0.9911 | 12:62995340 | 4 |
| 12:62968740:C:CA | rs11301095 | 12 | 62968740 | CA | C | 0.3539 | NA | NA | NA | 0.9911 | 12:62995340 | 4 |
| 12:62972210:G:GT | rs555030366 | 12 | 62972210 | G | GT | 0.3668 | NA | NA | NA | 0.6668 | 12:62995340 | 4 |
| 12:62977175:A:AG | rs34394452 | 12 | 62977175 | AG | A | 0.3549 | NA | NA | NA | 0.9868 | 12:62995340 | 4 |
| 12:62982152:C:CT | rs566612223 | 12 | 62982152 | C | CT | 0.3141 | NA | NA | NA | 0.7635 | 12:62995340 | 4 |
| 12:62992896:G:GA | rs34135257 | 12 | 62992896 | GA | G | 0.3539 | NA | NA | NA | 0.9911 | 12:62995340 | 4 |
| 12:63001068:A:AT | rs35168986 | 12 | 63001068 | AT | A | 0.3956 | NA | NA | NA | 0.7692 | 12:62995340 | 4 |
| 13:83312341:C:T | 13:83312341 | 13 | 83312341 | T | C | 0.4215 | 3.23e-08 | -0.007206 | 0.001303 | 1 | 13:83312341 | 5 |
| 13:83311141:A:G | 13:83311141 | 13 | 83311141 | A | G | 0.4205 | 3.52e-08 | -0.007179 | 0.001301 | 0.9962 | 13:83312341 | 5 |
| 13:83312717:C:G | 13:83312717 | 13 | 83312717 | C | G | 0.4225 | 3.74e-08 | -0.007177 | 0.001303 | 0.9737 | 13:83312341 | 5 |
| 13:83304898:G:T | 13:83304898 | 13 | 83304898 | G | T | 0.4205 | 4.87e-08 | -0.007095 | 0.001299 | 0.9962 | 13:83312341 | 5 |
| 13:83306586:A:G | 13:83306586 | 13 | 83306586 | A | G | 0.4205 | 5.07e-08 | -0.007087 | 0.0013 | 0.9962 | 13:83312341 | 5 |
| 13:83300359:A:G | 13:83300359 | 13 | 83300359 | G | A | 0.3728 | 9.13e-08 | -0.007131 | 0.001334 | 0.8286 | 13:83312341 | 5 |
| 13:83304274:C:T | 13:83304274 | 13 | 83304274 | C | T | 0.3757 | 1.01e-07 | -0.007094 | 0.001331 | 0.8374 | 13:83312341 | 5 |
| 13:83302594:C:T | 13:83302594 | 13 | 83302594 | T | C | 0.3728 | 1.08e-07 | -0.007089 | 0.001333 | 0.8286 | 13:83312341 | 5 |
| 13:83302795:A:T | 13:83302795 | 13 | 83302795 | T | A | 0.3757 | 1.12e-07 | -0.007075 | 0.001333 | 0.8374 | 13:83312341 | 5 |
| 13:83300269:C:T | 13:83300269 | 13 | 83300269 | T | C | 0.3757 | 1.94e-07 | -0.006959 | 0.001336 | 0.8374 | 13:83312341 | 5 |
| 13:83299269:C:T | 13:83299269 | 13 | 83299269 | C | T | 0.4274 | 3.93e-07 | -0.006623 | 0.001305 | 0.9625 | 13:83312341 | 5 |
| 13:83298475:A:G | 13:83298475 | 13 | 83298475 | A | G | 0.4235 | 7.82e-07 | -0.006459 | 0.001307 | 0.9473 | 13:83312341 | 5 |
| 13:83300084:A:G | rs9575156 | 13 | 83300084 | G | A | 0.3767 | NA | NA | NA | 0.8119 | 13:83312341 | 5 |
| 13:83300138:A:AGAGAGC | rs563056680 | 13 | 83300138 | AGAGAGC | A | 0.3936 | NA | NA | NA | 0.6099 | 13:83312341 | 5 |
| 16:13040514:C:T | 16:13040514 | 16 | 13040514 | T | C | 0.2932 | 2.19e-08 | -0.008169 | 0.001459 | 1 | 16:13040514 | 6 |
| 16:13041027:G:T | 16:13041027 | 16 | 13041027 | T | G | 0.2942 | 3.66e-08 | -0.008021 | 0.001456 | 0.995 | 16:13040514 | 6 |
| 16:13042097:A:G | 16:13042097 | 16 | 13042097 | A | G | 0.2942 | 3.66e-08 | -0.00802 | 0.001456 | 0.995 | 16:13040514 | 6 |
| 16:13039154:C:T | 16:13039154 | 16 | 13039154 | T | C | 0.2942 | 3.79e-08 | -0.008713 | 0.001583 | 0.995 | 16:13040514 | 6 |
| 16:13040889:C:G | 16:13040889 | 16 | 13040889 | G | C | 0.2942 | 3.86e-08 | -0.008004 | 0.001455 | 0.995 | 16:13040514 | 6 |
| 16:13043452:A:G | 16:13043452 | 16 | 13043452 | G | A | 0.2942 | 4.41e-08 | -0.007971 | 0.001455 | 0.995 | 16:13040514 | 6 |
| 16:13041924:A:G | 16:13041924 | 16 | 13041924 | G | A | 0.2942 | 4.44e-08 | -0.007968 | 0.001455 | 0.995 | 16:13040514 | 6 |
| 16:13039642:A:T | 16:13039642 | 16 | 13039642 | A | T | 0.2932 | 5.41e-08 | -0.007918 | 0.001455 | 1 | 16:13040514 | 6 |
| 16:13044033:C:T | 16:13044033 | 16 | 13044033 | C | T | 0.2942 | 5.82e-08 | -0.007898 | 0.001455 | 0.995 | 16:13040514 | 6 |
| 16:13030222:A:G | 16:13030222 | 16 | 13030222 | G | A | 0.3658 | 7.24e-08 | 0.00728 | 0.001351 | 0.7167 | 16:13040514 | 6 |
| 16:13032863:A:T | 16:13032863 | 16 | 13032863 | A | T | 0.3658 | 8.31e-08 | 0.007242 | 0.00135 | 0.7167 | 16:13040514 | 6 |
| 16:13032351:C:G | 16:13032351 | 16 | 13032351 | G | C | 0.3658 | 8.65e-08 | 0.007241 | 0.001352 | 0.7167 | 16:13040514 | 6 |
| 16:13038723:G:T | 16:13038723 | 16 | 13038723 | G | T | 0.2942 | 8.66e-08 | -0.007784 | 0.001453 | 0.995 | 16:13040514 | 6 |
| 16:13023207:C:T | 16:13023207 | 16 | 13023207 | T | C | 0.3668 | 1.05e-07 | 0.007209 | 0.001355 | 0.7152 | 16:13040514 | 6 |

| uniqID | rsID | chr | pos | non_effect allele | effect_allele | MAF | gwasP | beta | se | r2 | IndSigSNP | GenomicLocus |
|---|---|---|---|---|---|---|---|---|---|---|---|---|
| 16:13023388:A:G | 16:13023388 | 16 | 13023388 | G | A | 0.3658 | 1.07e-07 | 0.007205 | 0.001355 | 0.7105 | 16:13040514 | 6 |
| 16:13024150:A:G | 16:13024150 | 16 | 13024150 | G | A | 0.3658 | 1.12e-07 | 0.007203 | 0.001356 | 0.7105 | 16:13040514 | 6 |
| 16:13030875:A:G | 16:13030875 | 16 | 13030875 | G | A | 0.3658 | 1.18e-07 | 0.007142 | 0.001348 | 0.7167 | 16:13040514 | 6 |
| 16:13025315:G:T | 16:13025315 | 16 | 13025315 | G | T | 0.3658 | 1.37e-07 | 0.007146 | 0.001355 | 0.7105 | 16:13040514 | 6 |
| 16:13032547:A:G | 16:13032547 | 16 | 13032547 | G | A | 0.3658 | 1.51e-07 | 0.00708 | 0.001347 | 0.7167 | 16:13040514 | 6 |
| 16:13041940:A:C | 16:13041940 | 16 | 13041940 | A | C | 0.2684 | 1.87e-07 | -0.007811 | 0.001498 | 0.8733 | 16:13040514 | 6 |
| 16:13039646:C:T | 16:13039646 | 16 | 13039646 | T | C | 0.2704 | 1.94e-07 | -0.007794 | 0.001496 | 0.8824 | 16:13040514 | 6 |
| 16:13031195:A:C | 16:13031195 | 16 | 13031195 | C | A | 0.3658 | 1.98e-07 | 0.007048 | 0.001354 | 0.7167 | 16:13040514 | 6 |
| 16:13066833:C:T | 16:13066833 | 16 | 13066833 | T | C | 0.2793 | 1.98e-07 | -0.007585 | 0.001457 | 0.9126 | 16:13040514 | 6 |
| 16:13041921:C:T | 16:13041921 | 16 | 13041921 | C | T | 0.2714 | 2.03e-07 | -0.007785 | 0.001497 | 0.8772 | 16:13040514 | 6 |
| 16:13036811:C:T | 16:13036811 | 16 | 13036811 | T | C | 0.3608 | 2.3e-07 | -0.007613 | 0.001471 | 0.7313 | 16:13040514 | 6 |
| 16:13026502:A:G | 16:13026502 | 16 | 13026502 | A | G | 0.2724 | 2.35e-07 | 0.007756 | 0.001499 | 0.8714 | 16:13040514 | 6 |
| 16:13038196:A:G | 16:13038196 | 16 | 13038196 | G | A | 0.3628 | 2.36e-07 | -0.006961 | 0.001346 | 0.7264 | 16:13040514 | 6 |
| 16:13023394:A:G | 16:13023394 | 16 | 13023394 | G | A | 0.2704 | 2.45e-07 | 0.007774 | 0.001505 | 0.8528 | 16:13040514 | 6 |
| 16:13021889:C:T | 16:13021889 | 16 | 13021889 | C | T | 0.2753 | 2.51e-07 | 0.0078 | 0.001511 | 0.8668 | 16:13040514 | 6 |
| 16:13097206:C:G | 16:13097206 | 16 | 13097206 | G | C | 0.2793 | 2.57e-07 | -0.007516 | 0.001458 | 0.8833 | 16:13040514 | 6 |
| 16:13038054:G:T | 16:13038054 | 16 | 13038054 | T | G | 0.3608 | 2.68e-07 | -0.006939 | 0.001348 | 0.7313 | 16:13040514 | 6 |
| 16:13053951:C:T | 16:13053951 | 16 | 13053951 | T | C | 0.2803 | 2.76e-07 | -0.007646 | 0.001487 | 0.9277 | 16:13040514 | 6 |
| 16:13044827:C:T | 16:13044827 | 16 | 13044827 | C | T | 0.2833 | 3.03e-07 | -0.007597 | 0.001482 | 0.9415 | 16:13040514 | 6 |
| 16:13088905:C:T | 16:13088905 | 16 | 13088905 | C | T | 0.2932 | 3.51e-07 | -0.007378 | 0.001447 | 0.9403 | 16:13040514 | 6 |
| 16:13047734:C:T | 16:13047734 | 16 | 13047734 | C | T | 0.2833 | 3.58e-07 | -0.007546 | 0.001481 | 0.9415 | 16:13040514 | 6 |
| 16:13053882:A:G | 16:13053882 | 16 | 13053882 | A | G | 0.2803 | 3.6e-07 | -0.007557 | 0.001484 | 0.9277 | 16:13040514 | 6 |
| 16:13047598:C:G | 16:13047598 | 16 | 13047598 | C | G | 0.2833 | 3.63e-07 | -0.007542 | 0.001482 | 0.9415 | 16:13040514 | 6 |
| 16:13092220:C:T | 16:13092220 | 16 | 13092220 | T | C | 0.2932 | 3.83e-07 | -0.007327 | 0.001442 | 0.9403 | 16:13040514 | 6 |
| 16:13047022:C:G | 16:13047022 | 16 | 13047022 | G | C | 0.2823 | 3.86e-07 | -0.007566 | 0.00149 | 0.9365 | 16:13040514 | 6 |
| 16:13037305:C:T | 16:13037305 | 16 | 13037305 | C | T | 0.3608 | 3.88e-07 | -0.006847 | 0.001348 | 0.7313 | 16:13040514 | 6 |
| 16:13098440:C:G | 16:13098440 | 16 | 13098440 | C | G | 0.328 | 4.03e-07 | -0.006964 | 0.001373 | 0.7068 | 16:13040514 | 6 |
| 16:13048841:C:T | 16:13048841 | 16 | 13048841 | T | C | 0.2833 | 4.18e-07 | -0.007507 | 0.001482 | 0.9415 | 16:13040514 | 6 |
| 16:13049749:A:C | 16:13049749 | 16 | 13049749 | C | A | 0.2843 | 4.18e-07 | -0.007506 | 0.001482 | 0.9368 | 16:13040514 | 6 |
| 16:13099177:G:T | 16:13099177 | 16 | 13099177 | T | G | 0.3419 | 4.24e-07 | -0.006932 | 0.001369 | 0.767 | 16:13040514 | 6 |
| 16:13048395:C:T | 16:13048395 | 16 | 13048395 | C | T | 0.2833 | 4.42e-07 | -0.00749 | 0.001482 | 0.9415 | 16:13040514 | 6 |
| 16:13050340:A:C | 16:13050340 | 16 | 13050340 | A | C | 0.2833 | 4.45e-07 | -0.007488 | 0.001482 | 0.9415 | 16:13040514 | 6 |
| 16:13098508:A:G | 16:13098508 | 16 | 13098508 | A | G | 0.328 | 4.5e-07 | -0.006936 | 0.001373 | 0.7068 | 16:13040514 | 6 |
| 16:13061109:A:G | 16:13061109 | 16 | 13061109 | G | A | 0.2664 | 4.68e-07 | 0.008623 | 0.00171 | 0.8743 | 16:13040514 | 6 |
| 16:13054883:C:G | 16:13054883 | 16 | 13054883 | C | G | 0.2664 | 4.78e-07 | -0.007561 | 0.001501 | 0.8743 | 16:13040514 | 6 |
| 16:13097125:A:G | 16:13097125 | 16 | 13097125 | G | A | 0.2932 | 4.81e-07 | -0.00728 | 0.001445 | 0.9403 | 16:13040514 | 6 |
| 16:13052050:A:G | 16:13052050 | 16 | 13052050 | A | G | 0.2664 | 4.87e-07 | -0.007535 | 0.001497 | 0.8743 | 16:13040514 | 6 |
| 16:13098934:A:G | 16:13098934 | 16 | 13098934 | A | G | 0.328 | 4.91e-07 | -0.006913 | 0.001373 | 0.7068 | 16:13040514 | 6 |
| 16:13049557:C:T | 16:13049557 | 16 | 13049557 | T | C | 0.2803 | 5.71e-07 | -0.007419 | 0.001483 | 0.9277 | 16:13040514 | 6 |
| 16:13052714:C:G | 16:13052714 | 16 | 13052714 | G | C | 0.2664 | 5.76e-07 | -0.007487 | 0.001497 | 0.8743 | 16:13040514 | 6 |
| 16:13053072:A:G | 16:13053072 | 16 | 13053072 | G | A | 0.2664 | 5.82e-07 | -0.007478 | 0.001495 | 0.8743 | 16:13040514 | 6 |
| 16:13053187:C:T | 16:13053187 | 16 | 13053187 | C | T | 0.2803 | 5.91e-07 | -0.007411 | 0.001483 | 0.9277 | 16:13040514 | 6 |
| 16:13049906:A:T | 16:13049906 | 16 | 13049906 | T | A | 0.2803 | 6.22e-07 | -0.007398 | 0.001483 | 0.9277 | 16:13040514 | 6 |
| 16:13053270:A:G | 16:13053270 | 16 | 13053270 | G | A | 0.2803 | 6.34e-07 | -0.007378 | 0.00148 | 0.9277 | 16:13040514 | 6 |
| 16:13051686:A:C | 16:13051686 | 16 | 13051686 | A | C | 0.2803 | 6.66e-07 | -0.007378 | 0.001483 | 0.9277 | 16:13040514 | 6 |
| 16:13053924:C:G | 16:13053924 | 16 | 13053924 | G | C | 0.2803 | 7.03e-07 | -0.007366 | 0.001484 | 0.9277 | 16:13040514 | 6 |
| 16:13053576:A:T | 16:13053576 | 16 | 13053576 | T | A | 0.2813 | 7.04e-07 | -0.007348 | 0.00148 | 0.9226 | 16:13040514 | 6 |

| uniqID | rsID | chr | pos | non_effect allele | effect_allele | MAF | gwasP | beta | se | r2 | IndSigSNP | GenomicLocus |
|---|---|---|---|---|---|---|---|---|---|---|---|---|
| 16:13091332:C:T | 16:13091332 | 16 | 13091332 | T | C | 0.2664 | 7.04e-07 | -0.007426 | 0.001496 | 0.836 | 16:13040514 | 6 |
| 16:13054265:A:G | 16:13054265 | 16 | 13054265 | A | G | 0.2803 | 7.36e-07 | -0.007335 | 0.00148 | 0.9277 | 16:13040514 | 6 |
| 16:13059539:C:T | 16:13059539 | 16 | 13059539 | C | T | 0.2803 | 7.45e-07 | -0.007329 | 0.00148 | 0.9277 | 16:13040514 | 6 |
| 16:13054645:C:T | 16:13054645 | 16 | 13054645 | C | T | 0.2803 | 7.46e-07 | -0.00733 | 0.00148 | 0.9277 | 16:13040514 | 6 |
| 16:13055220:C:T | 16:13055220 | 16 | 13055220 | T | C | 0.2803 | 7.58e-07 | -0.007326 | 0.00148 | 0.9277 | 16:13040514 | 6 |
| 16:13054037:C:G | 16:13054037 | 16 | 13054037 | G | C | 0.2803 | 7.67e-07 | -0.007322 | 0.00148 | 0.9277 | 16:13040514 | 6 |
| 16:13054897:A:C | 16:13054897 | 16 | 13054897 | C | A | 0.2803 | 7.69e-07 | -0.007325 | 0.001481 | 0.9277 | 16:13040514 | 6 |
| 16:13057200:A:G | 16:13057200 | 16 | 13057200 | A | G | 0.2803 | 8.02e-07 | -0.007313 | 0.001481 | 0.918 | 16:13040514 | 6 |
| 16:13052846:A:G | 16:13052846 | 16 | 13052846 | G | A | 0.2813 | 8.16e-07 | -0.007315 | 0.001482 | 0.9323 | 16:13040514 | 6 |
| 16:13055766:C:T | 16:13055766 | 16 | 13055766 | T | C | 0.2803 | 8.17e-07 | -0.007303 | 0.00148 | 0.9277 | 16:13040514 | 6 |
| 16:13077901:A:G | 16:13077901 | 16 | 13077901 | A | G | 0.3419 | 8.17e-07 | -0.006759 | 0.00137 | 0.7754 | 16:13040514 | 6 |
| 16:13062232:A:G | 16:13062232 | 16 | 13062232 | A | G | 0.2803 | 8.93e-07 | -0.007277 | 0.00148 | 0.9277 | 16:13040514 | 6 |
| 16:13053140:C:G | 16:13053140 | 16 | 13053140 | G | C | 0.2803 | 9.15e-07 | -0.007299 | 0.001486 | 0.9277 | 16:13040514 | 6 |
| 16:13053136:G:T | 16:13053136 | 16 | 13053136 | T | G | 0.2803 | 9.22e-07 | -0.007297 | 0.001486 | 0.9277 | 16:13040514 | 6 |
| 16:13077351:C:T | 16:13077351 | 16 | 13077351 | C | T | 0.2674 | 9.35e-07 | -0.007298 | 0.001487 | 0.8401 | 16:13040514 | 6 |
| 16:13075745:A:G | 16:13075745 | 16 | 13075745 | G | A | 0.2674 | 9.78e-07 | -0.00726 | 0.001482 | 0.8401 | 16:13040514 | 6 |
| 16:13097084:G:T | 16:13097084 | 16 | 13097084 | T | G | 0.2664 | 1.08e-06 | -0.007252 | 0.001486 | 0.836 | 16:13040514 | 6 |
| 16:13079214:A:G | 16:13079214 | 16 | 13079214 | G | A | 0.3419 | 1.09e-06 | -0.006646 | 0.001362 | 0.7754 | 16:13040514 | 6 |
| 16:13079463:C:T | 16:13079463 | 16 | 13079463 | C | T | 0.3419 | 1.14e-06 | -0.006635 | 0.001362 | 0.7754 | 16:13040514 | 6 |
| 16:13089854:G:T | 16:13089854 | 16 | 13089854 | G | T | 0.2664 | 1.14e-06 | -0.007213 | 0.001481 | 0.836 | 16:13040514 | 6 |
| 16:13086810:C:T | 16:13086810 | 16 | 13086810 | T | C | 0.2664 | 1.17e-06 | -0.007218 | 0.001484 | 0.836 | 16:13040514 | 6 |
| 16:13099953:G:T | 16:13099953 | 16 | 13099953 | G | T | 0.2664 | 1.18e-06 | -0.007233 | 0.001487 | 0.836 | 16:13040514 | 6 |
| 16:13100021:C:T | 16:13100021 | 16 | 13100021 | C | T | 0.2664 | 1.24e-06 | -0.007219 | 0.001487 | 0.836 | 16:13040514 | 6 |
| 16:13096300:C:T | 16:13096300 | 16 | 13096300 | T | C | 0.2803 | 1.25e-06 | -0.007156 | 0.001475 | 0.8888 | 16:13040514 | 6 |
| 16:13099919:A:G | 16:13099919 | 16 | 13099919 | A | G | 0.2664 | 1.25e-06 | -0.007216 | 0.001487 | 0.836 | 16:13040514 | 6 |
| 16:13089059:A:G | 16:13089059 | 16 | 13089059 | A | G | 0.2664 | 1.29e-06 | -0.007187 | 0.001483 | 0.836 | 16:13040514 | 6 |
| 16:13070238:A:G | 16:13070238 | 16 | 13070238 | G | A | 0.2674 | 1.31e-06 | -0.007193 | 0.001485 | 0.8401 | 16:13040514 | 6 |
| 16:13099114:C:G | 16:13099114 | 16 | 13099114 | C | G | 0.2664 | 1.32e-06 | -0.007199 | 0.001487 | 0.836 | 16:13040514 | 6 |
| 16:13070671:C:T | 16:13070671 | 16 | 13070671 | T | C | 0.2674 | 1.34e-06 | -0.007183 | 0.001485 | 0.8401 | 16:13040514 | 6 |
| 16:13079082:A:C | 16:13079082 | 16 | 13079082 | C | A | 0.2674 | 1.34e-06 | -0.007232 | 0.001495 | 0.8401 | 16:13040514 | 6 |
| 16:13070809:C:G | 16:13070809 | 16 | 13070809 | G | C | 0.2674 | 1.35e-06 | -0.007181 | 0.001485 | 0.8401 | 16:13040514 | 6 |
| 16:13083055:G:T | 16:13083055 | 16 | 13083055 | G | T | 0.2674 | 1.37e-06 | -0.00717 | 0.001484 | 0.8401 | 16:13040514 | 6 |
| 16:13082445:A:G | 16:13082445 | 16 | 13082445 | A | G | 0.2674 | 1.39e-06 | -0.007167 | 0.001484 | 0.8401 | 16:13040514 | 6 |
| 16:13081923:C:G | 16:13081923 | 16 | 13081923 | G | C | 0.2674 | 1.45e-06 | -0.007147 | 0.001482 | 0.8401 | 16:13040514 | 6 |
| 16:13093774:G:T | 16:13093774 | 16 | 13093774 | T | G | 0.2803 | 1.59e-06 | -0.007073 | 0.001473 | 0.8888 | 16:13040514 | 6 |
| 16:13080515:C:T | 16:13080515 | 16 | 13080515 | T | C | 0.2674 | 1.8e-06 | -0.007092 | 0.001484 | 0.8401 | 16:13040514 | 6 |
| 16:13079389:G:T | 16:13079389 | 16 | 13079389 | T | G | 0.3161 | 2.09e-06 | -0.006608 | 0.001392 | 0.6814 | 16:13040514 | 6 |
| 16:13093778:C:T | 16:13093778 | 16 | 13093778 | T | C | 0.2803 | 2.1e-06 | -0.006984 | 0.001471 | 0.8888 | 16:13040514 | 6 |
| 16:13092663:C:T | 16:13092663 | 16 | 13092663 | C | T | 0.2803 | 2.13e-06 | -0.006969 | 0.001469 | 0.8888 | 16:13040514 | 6 |
| 16:13078044:A:T | 16:13078044 | 16 | 13078044 | A | T | 0.2674 | 2.14e-06 | -0.007044 | 0.001485 | 0.8401 | 16:13040514 | 6 |
| 16:13093858:C:T | 16:13093858 | 16 | 13093858 | T | C | 0.2803 | 2.2e-06 | -0.006971 | 0.001471 | 0.8888 | 16:13040514 | 6 |
| 16:13092100:A:G | 16:13092100 | 16 | 13092100 | G | A | 0.2803 | 2.24e-06 | -0.006954 | 0.001469 | 0.8888 | 16:13040514 | 6 |
| 16:13112335:C:T | 16:13112335 | 16 | 13112335 | T | C | 0.2932 | 2.42e-06 | -0.006661 | 0.001412 | 0.6643 | 16:13040514 | 6 |
| 16:13098762:A:C | 16:13098762 | 16 | 13098762 | A | C | 0.3151 | 2.5e-06 | -0.006587 | 0.001398 | 0.6772 | 16:13040514 | 6 |
| 16:13094769:G:T | 16:13094769 | 16 | 13094769 | G | T | 0.2803 | 2.53e-06 | -0.006931 | 0.001472 | 0.8888 | 16:13040514 | 6 |
| 16:13095296:A:T | 16:13095296 | 16 | 13095296 | T | A | 0.2803 | 2.54e-06 | -0.006961 | 0.001479 | 0.8888 | 16:13040514 | 6 |
| 16:13095739:A:C | 16:13095739 | 16 | 13095739 | C | A | 0.2803 | 2.68e-06 | -0.006907 | 0.001471 | 0.8888 | 16:13040514 | 6 |

| uniqID | rsID | chr | pos | non_effect allele | effect_allele | MAF | gwasP | beta | se | r2 | IndSigSNP | GenomicLocus |
|---|---|---|---|---|---|---|---|---|---|---|---|---|
| 16:13095171:C:T | 16:13095171 | 16 | 13095171 | T | C | 0.2803 | 2.81e-06 | -0.006906 | 0.001473 | 0.8888 | 16:13040514 | 6 |
| 16:13077915:A:G | 16:13077915 | 16 | 13077915 | A | G | 0.2674 | 2.83e-06 | -0.006974 | 0.001488 | 0.8401 | 16:13040514 | 6 |
| 16:13095142:C:G | 16:13095142 | 16 | 13095142 | G | C | 0.2803 | 2.83e-06 | -0.006904 | 0.001473 | 0.8888 | 16:13040514 | 6 |
| 16:13105091:A:C | 16:13105091 | 16 | 13105091 | A | C | 0.2932 | 2.99e-06 | -0.006607 | 0.001413 | 0.6643 | 16:13040514 | 6 |
| 16:13109594:G:T | 16:13109594 | 16 | 13109594 | T | G | 0.2932 | 3.08e-06 | -0.006593 | 0.001412 | 0.6643 | 16:13040514 | 6 |
| 16:13094897:C:G | 16:13094897 | 16 | 13094897 | C | G | 0.2803 | 3.16e-06 | -0.006868 | 0.001473 | 0.8888 | 16:13040514 | 6 |
| 16:13105863:C:T | 16:13105863 | 16 | 13105863 | T | C | 0.2922 | 3.25e-06 | -0.006583 | 0.001413 | 0.6603 | 16:13040514 | 6 |
| 16:13104924:C:T | 16:13104924 | 16 | 13104924 | T | C | 0.2922 | 3.68e-06 | -0.006547 | 0.001413 | 0.6685 | 16:13040514 | 6 |
| 16:13118299:A:T | 16:13118299 | 16 | 13118299 | A | T | 0.2942 | 3.7e-06 | -0.00656 | 0.001417 | 0.6601 | 16:13040514 | 6 |
| 16:13105998:C:T | 16:13105998 | 16 | 13105998 | C | T | 0.2932 | 3.75e-06 | -0.006564 | 0.001418 | 0.6643 | 16:13040514 | 6 |
| 16:13080252:A:G | 16:13080252 | 16 | 13080252 | A | G | 0.3161 | 3.84e-06 | -0.006421 | 0.001389 | 0.6814 | 16:13040514 | 6 |
| 16:13115755:A:G | 16:13115755 | 16 | 13115755 | G | A | 0.2942 | 4.05e-06 | -0.006505 | 0.00141 | 0.6601 | 16:13040514 | 6 |
| 16:13078807:C:T | 16:13078807 | 16 | 13078807 | T | C | 0.3161 | 4.07e-06 | -0.006399 | 0.001388 | 0.6814 | 16:13040514 | 6 |
| 16:13115702:C:G | 16:13115702 | 16 | 13115702 | C | G | 0.2942 | 4.39e-06 | -0.006481 | 0.00141 | 0.6601 | 16:13040514 | 6 |
| 16:13079698:A:T | 16:13079698 | 16 | 13079698 | T | A | 0.3161 | 4.47e-06 | -0.006375 | 0.001388 | 0.6814 | 16:13040514 | 6 |
| 16:13101555:C:T | 16:13101555 | 16 | 13101555 | C | T | 0.3161 | 9.22e-06 | -0.006237 | 0.001405 | 0.8527 | 16:13040514 | 6 |
| 16:13101618:A:G | 16:13101618 | 16 | 13101618 | A | G | 0.2883 | 3.65e-05 | -0.00594 | 0.001438 | 0.7401 | 16:13040514 | 6 |
| 16:13102906:A:T | 16:13102906 | 16 | 13102906 | T | A | 0.2803 | 4.72e-05 | -0.005879 | 0.001444 | 0.7112 | 16:13040514 | 6 |
| 16:13022033:A:AT | rs11361092 | 16 | 13022033 | A | AT | 0.2753 | NA | NA | NA | 0.8561 | 16:13040514 | 6 |
| 16:13029711:G:GA | rs11428944 | 16 | 13029711 | GA | G | 0.3131 | NA | NA | NA | 0.6838 | 16:13040514 | 6 |
| 16:13035206:T:TA | rs34236666 | 16 | 13035206 | TA | T | 0.3648 | NA | NA | NA | 0.7183 | 16:13040514 | 6 |
| 16:13045117:C:CA | rs113668400 | 16 | 13045117 | CA | C | 0.2833 | NA | NA | NA | 0.9415 | 16:13040514 | 6 |
| 16:13048916:G:GT | rs527441820 | 16 | 13048916 | G | GT | 0.2783 | NA | NA | NA | 0.897 | 16:13040514 | 6 |
| 16:13049490:T:TA | rs571415029 | 16 | 13049490 | T | TA | 0.2654 | NA | NA | NA | 0.7099 | 16:13040514 | 6 |
| 16:13053457:A:AC | rs11430580 | 16 | 13053457 | A | AC | 0.2803 | NA | NA | NA | 0.9277 | 16:13040514 | 6 |
| 16:13055621:T:TCC | rs35005963 | 16 | 13055621 | T | TCC | 0.2664 | NA | NA | NA | 0.8743 | 16:13040514 | 6 |
| 16:13058241:G:GT | rs566468808 | 16 | 13058241 | G | GT | 0.3012 | NA | NA | NA | 0.769 | 16:13040514 | 6 |
| 16:13079063:C:CTT | rs3075030 | 16 | 13079063 | C | CTT | 0.3419 | NA | NA | NA | 0.7754 | 16:13040514 | 6 |
| 16:13083068:G:GT | rs71147777 | 16 | 13083068 | G | GT | 0.2674 | NA | NA | NA | 0.8401 | 16:13040514 | 6 |
| 16:13097746:G:T | rs60560408 | 16 | 13097746 | G | T | 0.3131 | NA | NA | NA | 0.6032 | 16:13040514 | 6 |
| 16:13097749:G:T | rs8050866 | 16 | 13097749 | G | T | 0.2326 | NA | NA | NA | 0.6975 | 16:13040514 | 6 |
| 16:13102532:C:T | rs4781381 | 16 | 13102532 | C | T | 0.2793 | NA | NA | NA | 0.7059 | 16:13040514 | 6 |
| 16:13116871:T:TTTTA | rs144610339 | 16 | 13116871 | TTTTA | T | 0.2942 | NA | NA | NA | 0.6601 | 16:13040514 | 6 |

*part 2 of 2 — first column repeated in every part*

| uniqID | nearestGene | dist | func | CADD | RDB | minChrState | commonChrState | posMapFilt | eqtlMapFilt | ciMapFilt |
|---|---|---|---|---|---|---|---|---|---|---|
| 2:98580012:C:T | TMEM131 | 0 | intronic | 3.615 | 7 | 5 | 5 | 1 | 1 | 0 |
| 2:98427382:C:T | TMEM131 | 0 | intronic | 10.38 | 5 | 4 | 4 | 1 | 1 | 0 |
| 2:98354511:A:G | ZAP70 | 0 | exonic | 10.58 | 5 | 4 | 15 | 1 | 1 | 0 |
| 2:98357163:C:T | ZAP70 | 837 | downstream | 0.591 | 4 | 4 | 15 | 1 | 1 | 0 |
| 2:98520564:C:T | TMEM131 | 0 | intronic | 7.871 | 6 | 4 | 5 | 1 | 1 | 0 |
| 2:98370698:A:T | TMEM131 | 2100 | intergenic | 1.091 | 6 | 4 | 5 | 1 | 1 | 0 |
| 2:98493359:A:G | TMEM131 | 0 | intronic | 2.663 | 6 | 5 | 5 | 1 | 1 | 0 |
| 2:98519014:C:T | TMEM131 | 0 | intronic | 3.591 | 6 | 4 | 5 | 1 | 1 | 0 |
| 2:98495661:G:T | TMEM131 | 0 | intronic | 6.015 | 6 | 4 | 5 | 1 | 1 | 0 |
| 2:98440234:C:T | TMEM131 | 0 | intronic | 2.827 | 5 | 4 | 5 | 1 | 1 | 0 |
| 2:98491750:C:T | TMEM131 | 0 | intronic | 0.197 | 6 | 5 | 5 | 1 | 1 | 0 |
| 2:98506910:C:T | TMEM131 | 0 | intronic | 0.597 | 4 | 4 | 5 | 1 | 1 | 0 |
| 2:98521823:A:C | TMEM131 | 0 | intronic | 11.46 | 5 | 4 | 5 | 1 | 1 | 0 |
| 2:98365164:C:G | TMEM131 | 7634 | intergenic | 0.757 | 4 | 2 | 5 | 1 | 1 | 0 |
| 2:98502987:C:T | TMEM131 | 0 | intronic | 4.338 | 7 | 4 | 8 | 1 | 1 | 0 |
| 2:98510166:G:T | TMEM131 | 0 | intronic | 1.728 | 7 | 5 | 5 | 1 | 1 | 0 |
| 2:98623227:A:G | TMEM131 | 10838 | intergenic | 0.256 | 7 | 5 | 15 | 0 | 1 | 0 |
| 2:98623406:A:G | TMEM131 | 11017 | intergenic | 0.347 | 6 | 5 | 15 | 0 | 1 | 0 |
| 2:98410769:G:T | TMEM131 | 0 | intronic | 2.088 | 6 | 4 | 5 | 1 | 1 | 0 |
| 2:98435982:C:T | TMEM131 | 0 | intronic | 7.27 | 5 | 4 | 5 | 1 | 1 | 0 |
| 2:98382886:A:G | TMEM131 | 0 | intronic | 5.539 | 5 | 2 | 4 | 1 | 1 | 0 |
| 2:98461821:C:T | TMEM131 | 0 | intronic | 0.604 | 5 | 4 | 5 | 1 | 1 | 0 |
| 2:98567820:A:G | TMEM131 | 0 | intronic | 4.684 | 3a | 5 | 7 | 1 | 1 | 0 |
| 2:98409565:C:T | TMEM131 | 0 | intronic | 0.091 | 6 | 4 | 4 | 1 | 1 | 0 |
| 2:98517626:A:G | TMEM131 | 0 | intronic | 1.939 | 5 | 5 | 5 | 1 | 1 | 0 |
| 2:98461953:C:T | TMEM131 | 0 | intronic | 2.06 | 7 | 4 | 5 | 1 | 1 | 0 |
| 2:98534531:C:T | TMEM131 | 0 | intronic | 8.039 | 6 | 4 | 5 | 1 | 1 | 0 |
| 2:98621060:A:T | TMEM131 | 8671 | intergenic | 0.848 | 7 | 5 | 15 | 1 | 1 | 0 |
| 2:98570999:A:G | TMEM131 | 0 | intronic | 3.673 | 5 | 2 | 15 | 1 | 1 | 0 |
| 2:98447004:C:T | TMEM131 | 0 | intronic | 4.929 | NA | 4 | 5 | 1 | 1 | 0 |
| 2:98449024:A:G | TMEM131 | 0 | intronic | 1.746 | 6 | 4 | 5 | 1 | 1 | 0 |
| 2:98407350:C:T | TMEM131 | 0 | intronic | 0.663 | 7 | 4 | 5 | 1 | 1 | 0 |
| 2:98580724:G:T | TMEM131 | 0 | intronic | 0.478 | 7 | 5 | 5 | 1 | 1 | 0 |
| 2:98588372:A:C | TMEM131 | 0 | intronic | 0.983 | 7 | 5 | 15 | 1 | 1 | 0 |
| 2:98565400:A:G | TMEM131 | 0 | intronic | 3.026 | 7 | 2 | 15 | 1 | 1 | 0 |
| 2:98612260:C:T | TMEM131 | 0 | UTR5 | 14.06 | NA | 1 | 1 | 1 | 1 | 0 |
| 2:98455510:C:G | TMEM131 | 0 | intronic | 1.062 | 7 | 4 | 5 | 1 | 1 | 0 |
| 2:98454572:C:G | TMEM131 | 0 | intronic | 1.995 | 7 | 4 | 5 | 1 | 1 | 0 |
| 2:98443658:C:T | TMEM131 | 0 | intronic | 5.605 | 5 | 4 | 5 | 1 | 1 | 0 |
| 2:98479581:A:C | TMEM131 | 0 | intronic | 3.39 | 6 | 4 | 5 | 1 | 1 | 0 |
| 2:98442202:A:G | TMEM131 | 0 | intronic | 3.931 | 7 | 4 | 5 | 1 | 1 | 0 |
| 2:98466647:C:T | TMEM131 | 0 | intronic | 2.087 | 4 | 4 | 5 | 1 | 1 | 0 |
| 2:98454473:C:T | TMEM131 | 0 | intronic | 2.151 | 5 | 4 | 5 | 1 | 1 | 0 |
| 2:98417089:C:T | TMEM131 | 0 | intronic | 1.456 | 6 | 4 | 5 | 1 | 1 | 0 |
| 2:98421364:A:G | TMEM131 | 0 | intronic | 4.476 | 6 | 4 | 4 | 1 | 1 | 0 |
| 2:98541001:C:T | TMEM131 | 0 | intronic | 5.185 | 7 | 4 | 4 | 1 | 1 | 0 |

| uniqID | nearestGene | dist | func | CADD | RDB | minChrState | commonChrState | posMapFilt | eqtlMapFilt | ciMapFilt |
|---|---|---|---|---|---|---|---|---|---|---|
| 2:98539588:A:C | TMEM131 | 0 | intronic | 1.541 | 7 | 4 | 5 | 1 | 1 | 0 |
| 2:98424802:A:G | TMEM131 | 0 | intronic | 3.82 | 4 | 4 | 5 | 1 | 1 | 0 |
| 2:98552299:C:G | TMEM131 | 0 | intronic | 0.448 | 2b | 1 | 7 | 1 | 1 | 0 |
| 2:98405695:C:T | TMEM131 | 0 | intronic | 0.348 | 4 | 4 | 4 | 1 | 1 | 0 |
| 2:98409046:A:G | TMEM131 | 0 | exonic | 18.87 | 5 | 4 | 4 | 1 | 1 | 0 |
| 2:98454472:A:G | TMEM131 | 0 | intronic | 2.226 | 5 | 4 | 5 | 1 | 1 | 0 |
| 2:98565144:A:G | TMEM131 | 0 | intronic | 2.208 | 5 | 4 | 15 | 1 | 1 | 0 |
| 2:98587288:A:G | TMEM131 | 0 | intronic | 8.473 | 5 | 5 | 15 | 1 | 1 | 0 |
| 2:98420431:C:T | TMEM131 | 0 | intronic | 8.313 | 7 | 4 | 5 | 1 | 1 | 0 |
| 2:98472610:C:T | TMEM131 | 0 | intronic | 6.196 | 6 | 4 | 5 | 1 | 1 | 0 |
| 2:98419726:A:T | TMEM131 | 0 | intronic | 5.647 | 7 | 4 | 5 | 1 | 1 | 0 |
| 2:98420142:C:G | TMEM131 | 0 | intronic | 5.067 | 7 | 4 | 4 | 1 | 1 | 0 |
| 2:98528689:A:G | TMEM131 | 0 | intronic | 1.128 | 2c | 1 | 5 | 1 | 1 | 0 |
| 2:98552271:C:G | TMEM131 | 0 | intronic | 11.3 | 4 | 1 | 7 | 1 | 1 | 0 |
| 2:98405929:A:G | TMEM131 | 0 | intronic | 5.081 | 3a | 4 | 4 | 1 | 1 | 0 |
| 2:98606770:C:T | TMEM131 | 0 | intronic | 1.129 | NA | 2 | 15 | 1 | 1 | 0 |
| 2:98413781:C:T | TMEM131 | 0 | intronic | 0.711 | 7 | 4 | 5 | 1 | 1 | 0 |
| 2:98455152:C:T | TMEM131 | 0 | intronic | 2.056 | 7 | 4 | 5 | 1 | 1 | 0 |
| 2:98377512:A:T | TMEM131 | 0 | intronic | 1.357 | 5 | 3 | 4 | 1 | 1 | 0 |
| 2:98379813:A:G | TMEM131 | 0 | intronic | 5.987 | 5 | 2 | 4 | 1 | 1 | 0 |
| 2:98379267:C:T | TMEM131 | 0 | intronic | 3.889 | 7 | 4 | 4 | 1 | 1 | 0 |
| 2:98397301:C:T | TMEM131 | 0 | intronic | 6.049 | 7 | 4 | 5 | 1 | 1 | 0 |
| 2:98402753:A:T | TMEM131 | 0 | intronic | 3.156 | 6 | 4 | 5 | 1 | 1 | 0 |
| 2:98402772:C:G | TMEM131 | 0 | intronic | 1.59 | 6 | 4 | 5 | 1 | 1 | 0 |
| 2:98395653:A:C | TMEM131 | 0 | intronic | 1.114 | 3a | 4 | 5 | 1 | 1 | 0 |
| 2:98374567:C:T | TMEM131 | 0 | intronic | 1.088 | 7 | 4 | 4 | 1 | 1 | 0 |
| 2:98561153:A:C | TMEM131 | 0 | intronic | 7.356 | 7 | 5 | 15 | 1 | 1 | 0 |
| 2:98386731:C:T | TMEM131 | 0 | intronic | 0.083 | 5 | 2 | 5 | 1 | 1 | 0 |
| 2:98393894:C:T | TMEM131 | 0 | intronic | 0.124 | 7 | 4 | 5 | 1 | 1 | 0 |
| 2:98557575:G:T | TMEM131 | 0 | intronic | 1.301 | 6 | 5 | 15 | 1 | 1 | 0 |
| 2:98605982:A:G | TMEM131 | 0 | intronic | 5.225 | 5 | 2 | 15 | 1 | 1 | 0 |
| 2:98501884:C:T | TMEM131 | 0 | intronic | 0.77 | 6 | 4 | 5 | 1 | 1 | 0 |
| 2:98416848:A:C | TMEM131 | 0 | intronic | 3.475 | NA | 4 | 5 | 1 | 1 | 0 |
| 2:98416850:G:T | TMEM131 | 0 | intronic | 6.209 | NA | 4 | 5 | 1 | 1 | 0 |
| 2:98554946:A:C | TMEM131 | 0 | intronic | 1.708 | 7 | 5 | 15 | 1 | 1 | 0 |
| 2:98448395:G:T | TMEM131 | 0 | intronic | 9.048 | 7 | 4 | 5 | 1 | 1 | 0 |
| 2:98602165:A:G | TMEM131 | 0 | intronic | 0.322 | 6 | 5 | 5 | 1 | 1 | 0 |
| 2:98342323:A:C | ZAP70 | 0 | intronic | 1.587 | 5 | 4 | 15 | 1 | 1 | 0 |
| 2:98488949:C:G | TMEM131 | 0 | intronic | 0.16 | 6 | 4 | 5 | 1 | 1 | 0 |
| 2:98454931:A:G | TMEM131 | 0 | intronic | 0.645 | 7 | 4 | 5 | 1 | 1 | 0 |
| 2:98443039:C:G | TMEM131 | 0 | intronic | 1.206 | 4 | 4 | 5 | 1 | 1 | 0 |
| 2:98336200:A:C | ZAP70 | 0 | intronic | 1.961 | 5 | 2 | 15 | 1 | 1 | 0 |
| 2:98405034:C:T | TMEM131 | 0 | intronic | 0.387 | 7 | 4 | 4 | 1 | 1 | 0 |
| 2:98355990:A:G | ZAP70 | 0 | UTR3 | 0.484 | 5 | 4 | 15 | 1 | 1 | 0 |
| 2:98534412:C:T | TMEM131 | 0 | intronic | 0.101 | 3a | 4 | 5 | 1 | 1 | 0 |
| 2:98354139:G:T | ZAP70 | 0 | intronic | 2.278 | 5 | 4 | 15 | 1 | 1 | 0 |
| 2:98353847:C:T | ZAP70 | 0 | intronic | 0.322 | 5 | 4 | 15 | 1 | 1 | 0 |

| uniqID | nearestGene | dist | func | CADD | RDB | minChrState | commonChrState | posMapFilt | eqtlMapFilt | ciMapFilt |
|---|---|---|---|---|---|---|---|---|---|---|
| 2:98339513:C:T | ZAP70 | 0 | intronic | 10.26 | 5 | 4 | 15 | 1 | 1 | 0 |
| 2:98325330:G:T | ZAP70 | 4692 | intergenic | 2.612 | 3a | 5 | 15 | 1 | 1 | 0 |
| 2:98326867:A:G | ZAP70 | 3155 | intergenic | 10.46 | 7 | 5 | 15 | 1 | 1 | 0 |
| 2:98330052:A:C | ZAP70 | 0 | UTR5 | 3.694 | NA | 1 | 15 | 1 | 1 | 0 |
| 2:98356846:A:G | ZAP70 | 520 | downstream | 3.061 | 5 | 4 | 15 | 1 | 1 | 0 |
| 2:98361679:C:G | ZAP70 | 5353 | intergenic | 0.189 | 7 | 4 | 5 | 1 | 1 | 0 |
| 2:98360443:A:G | ZAP70 | 4117 | intergenic | 2.059 | 2b | 4 | 5 | 1 | 1 | 0 |
| 2:98363313:A:T | ZAP70 | 6987 | intergenic | 0.132 | 5 | 4 | 5 | 1 | 1 | 0 |
| 2:98334679:C:T | ZAP70 | 0 | intronic | 1.276 | 5 | 1 | 15 | 1 | 1 | 0 |
| 2:98351986:C:T | ZAP70 | 0 | intronic | 2.517 | 5 | 1 | 15 | 1 | 1 | 0 |
| 2:98343258:A:G | ZAP70 | 0 | intronic | 0.578 | 5 | 4 | 15 | 1 | 1 | 0 |
| 2:98345086:G:T | ZAP70 | 0 | intronic | 4.315 | 5 | 2 | 15 | 1 | 1 | 0 |
| 2:98393231:C:T | TMEM131 | 0 | intronic | 0.26 | 5 | 4 | 5 | 1 | 1 | 0 |
| 2:98346461:A:G | ZAP70 | 0 | intronic | 0.589 | 7 | 4 | 9 | 1 | 1 | 0 |
| 2:98571084:A:G | TMEM131 | 0 | intronic | 1.749 | 5 | 5 | 15 | 1 | 1 | 0 |
| 2:98351654:T:TG | ZAP70 | 0 | intronic | 6.648 | NA | 1 | 15 | 1 | 1 | 0 |
| 2:98368551:A:ACT | TMEM131 | 4247 | intergenic | 1.011 | NA | 4 | 5 | 1 | 1 | 0 |
| 2:98384528:A:ATTTTC | TMEM131 | 0 | intronic | 1.9 | NA | 4 | 5 | 1 | 1 | 0 |
| 2:98390117:G:GT | TMEM131 | 0 | intronic | 1.21 | NA | 4 | 4 | 1 | 1 | 0 |
| 2:98434931:G:GGGGGC | TMEM131 | 0 | intronic | 0.409 | NA | 4 | 5 | 1 | 1 | 0 |
| 2:98443081:G:GA | TMEM131 | 0 | intronic | 0.909 | NA | 4 | 5 | 1 | 1 | 0 |
| 2:98468892:A:AC | TMEM131 | 0 | intronic | 1.118 | NA | 4 | 5 | 1 | 1 | 0 |
| 2:98506011:T:TATA | TMEM131 | 0 | intronic | 7.701 | NA | 4 | 5 | 1 | 1 | 0 |
| 2:98507972:C:CA | TMEM131 | 0 | intronic | 7.161 | NA | 5 | 5 | 1 | 1 | 0 |
| 2:98526685:A:AAAAT | TMEM131 | 0 | intronic | 1.979 | NA | 2 | 5 | 1 | 1 | 0 |
| 2:98547069:A:AT | TMEM131 | 0 | intronic | 0.185 | NA | 2 | 5 | 1 | 1 | 0 |
| 2:98548604:T:TA | TMEM131 | 0 | intronic | 6.515 | NA | 5 | 5 | 1 | 1 | 0 |
| 2:98551456:G:GA | TMEM131 | 0 | intronic | 0.585 | NA | 5 | 5 | 1 | 1 | 0 |
| 2:98565115:C:CT | TMEM131 | 0 | intronic | 1.425 | NA | 4 | 15 | 1 | 1 | 0 |
| 2:98604570:G:GACAC | TMEM131 | 0 | intronic | 7.818 | NA | 5 | 5 | 1 | 1 | 0 |
| 2:98616519:A:ATT | TMEM131 | 4130 | intergenic | 3.339 | NA | 5 | 15 | 1 | 1 | 0 |
| 2:98621058:A:AG | TMEM131 | 8669 | intergenic | 0.525 | NA | 5 | 15 | 1 | 1 | 0 |
| 8:11417582:C:T | BLK | 0 | intronic | 6.773 | 5 | 4 | 14 | 1 | 1 | 0 |
| 8:11235150:A:C | C8orf12 | 0 | intronic | 0.082 | 6 | 5 | 14 | 1 | 1 | 0 |
| 8:11226456:A:C | C8orf12 | 0 | intronic | 0.53 | 6 | 5 | 14 | 1 | 1 | 0 |
| 8:10147398:C:T | MSRA | 0 | intronic | 1.289 | 7 | 5 | 15 | 1 | 1 | 0 |
| 8:11226071:A:G | C8orf12 | 0 | intronic | 2.691 | 5 | 5 | 14 | 1 | 1 | 0 |
| 8:11232788:A:G | C8orf12 | 0 | intronic | 5.899 | 7 | 5 | 14 | 1 | 1 | 0 |
| 8:11231249:C:T | C8orf12 | 0 | intronic | 0.646 | NA | 5 | 14 | 1 | 1 | 0 |
| 8:11242632:A:G | C8orf12 | 0 | intronic | 1.013 | 7 | 13 | 14 | 1 | 1 | 0 |
| 8:11233318:A:C | C8orf12 | 0 | intronic | 6.373 | 6 | 5 | 14 | 1 | 1 | 0 |
| 8:11236681:G:T | C8orf12 | 0 | intronic | 1.926 | 7 | 5 | 14 | 1 | 1 | 0 |
| 8:11236685:C:T | C8orf12 | 0 | intronic | 5.579 | 7 | 5 | 14 | 1 | 1 | 0 |
| 8:11236419:C:T | C8orf12 | 0 | intronic | 0.261 | 7 | 5 | 14 | 1 | 1 | 0 |
| 8:11245303:C:G | C8orf12 | 0 | intronic | 4.095 | 5 | 5 | 14 | 1 | 1 | 0 |
| 8:11236413:C:T | C8orf12 | 0 | intronic | 2.361 | 7 | 5 | 14 | 1 | 1 | 0 |
| 8:11236572:A:T | C8orf12 | 0 | intronic | 10.93 | 6 | 5 | 14 | 1 | 1 | 0 |

| uniqID | nearestGene | dist | func | CADD | RDB | minChrState | commonChrState | posMapFilt | eqtlMapFilt | ciMapFilt |
|---|---|---|---|---|---|---|---|---|---|---|
| 8:11230574:A:G | C8orf12 | 0 | intronic | 0.19 | 6 | 5 | 14 | 1 | 1 | 0 |
| 8:11237477:A:G | C8orf12 | 0 | intronic | 3.855 | NA | 5 | 14 | 1 | 1 | 0 |
| 8:11237480:A:G | C8orf12 | 0 | intronic | 0.548 | NA | 5 | 14 | 1 | 1 | 0 |
| 8:11233419:G:T | C8orf12 | 0 | intronic | 1.64 | 7 | 5 | 14 | 1 | 1 | 0 |
| 8:11231886:A:G | C8orf12 | 0 | intronic | 8.262 | 6 | 5 | 14 | 1 | 1 | 0 |
| 8:11236392:C:T | C8orf12 | 0 | intronic | 4.437 | NA | 5 | 14 | 1 | 1 | 0 |
| 8:11227406:A:G | C8orf12 | 0 | intronic | 3.042 | NA | 5 | 14 | 1 | 1 | 0 |
| 8:11239762:A:T | C8orf12 | 0 | intronic | 7.515 | 1f | 13 | 14 | 1 | 1 | 0 |
| 8:11249010:G:T | C8orf12 | 0 | intronic | 1.583 | NA | 5 | 14 | 1 | 1 | 0 |
| 8:11232860:A:G | C8orf12 | 0 | intronic | 1.562 | 6 | 5 | 14 | 1 | 1 | 0 |
| 8:11080675:C:T | AF131215.8 | 19494 | intergenic | 0.164 | 5 | 13 | 14 | 0 | 1 | 0 |
| 8:11227885:C:T | C8orf12 | 0 | intronic | 0.248 | 5 | 5 | 14 | 1 | 1 | 0 |
| 8:11228100:A:G | C8orf12 | 0 | intronic | 1.674 | 7 | 5 | 14 | 1 | 1 | 0 |
| 8:11234500:A:T | C8orf12 | 0 | intronic | 3.108 | 1f | 5 | 14 | 1 | 1 | 0 |
| 8:11234520:A:G | C8orf12 | 0 | intronic | 1.936 | NA | 5 | 14 | 1 | 1 | 0 |
| 8:11234885:C:G | C8orf12 | 0 | intronic | 2.009 | 6 | 5 | 14 | 1 | 1 | 0 |
| 8:11233582:A:G | C8orf12 | 0 | intronic | 3.772 | 7 | 5 | 14 | 1 | 1 | 0 |
| 8:11233659:A:G | C8orf12 | 0 | intronic | 4.043 | 6 | 5 | 14 | 1 | 1 | 0 |
| 8:11234298:C:T | C8orf12 | 0 | intronic | 0.087 | 5 | 5 | 14 | 1 | 1 | 0 |
| 8:11062882:C:T | AF131215.8 | 1701 | intergenic | 4.088 | 1f | 5 | 14 | 1 | 1 | 0 |
| 8:11234844:A:G | C8orf12 | 0 | intronic | 0.451 | 6 | 5 | 14 | 1 | 1 | 0 |
| 8:11233958:A:G | C8orf12 | 0 | intronic | 2.323 | 7 | 5 | 14 | 1 | 1 | 0 |
| 8:11236975:A:G | C8orf12:RN7SL293P | 00:00:00 | ncRNA_exonic | 4.398 | NA | 5 | 14 | 1 | 1 | 0 |
| 8:11232343:C:G | C8orf12 | 0 | intronic | 14.38 | 7 | 5 | 14 | 1 | 1 | 0 |
| 8:11234780:C:G | C8orf12 | 0 | intronic | 0.478 | 6 | 5 | 14 | 1 | 1 | 0 |
| 8:11230206:G:T | C8orf12 | 0 | intronic | 2.811 | 7 | 5 | 14 | 1 | 1 | 0 |
| 8:11233917:C:T | C8orf12 | 0 | intronic | 0.215 | 6 | 5 | 14 | 1 | 1 | 0 |
| 8:11234367:G:T | C8orf12 | 0 | intronic | 0.956 | NA | 5 | 14 | 1 | 1 | 0 |
| 8:11235360:A:C | C8orf12 | 0 | intronic | 1.486 | NA | 5 | 14 | 1 | 1 | 0 |
| 8:11234626:C:T | C8orf12 | 0 | intronic | 2.092 | 6 | 5 | 14 | 1 | 1 | 0 |
| 8:11235497:A:G | C8orf12 | 0 | intronic | 0.33 | NA | 5 | 14 | 1 | 1 | 0 |
| 8:11234613:A:T | C8orf12 | 0 | intronic | 5.325 | 7 | 5 | 14 | 1 | 1 | 0 |
| 8:10988275:A:G | XKR6 | 0 | intronic | 0.395 | 5 | 5 | 15 | 1 | 1 | 0 |
| 8:11230259:A:T | C8orf12 | 0 | intronic | 2.428 | 6 | 5 | 14 | 1 | 1 | 0 |
| 8:11236850:C:T | C8orf12 | 0 | intronic | 0.043 | 7 | 5 | 14 | 1 | 1 | 0 |
| 8:11236964:C:T | C8orf12:RN7SL293P | 00:00:00 | ncRNA_exonic | 3.381 | 7 | 5 | 14 | 1 | 1 | 0 |
| 8:11235735:C:T | C8orf12 | 0 | intronic | 0.148 | NA | 5 | 14 | 1 | 1 | 0 |
| 8:11229319:C:G | C8orf12 | 0 | intronic | 0.337 | 5 | 5 | 14 | 1 | 1 | 0 |
| 8:11080665:A:T | AF131215.8 | 19484 | intergenic | 1.242 | 5 | 13 | 14 | 0 | 1 | 0 |
| 8:11235136:C:G | C8orf12 | 0 | intronic | 1.53 | 7 | 5 | 14 | 1 | 1 | 0 |
| 8:11249261:C:T | C8orf12 | 0 | intronic | 1.008 | 5 | 5 | 14 | 1 | 1 | 0 |
| 8:11243126:C:G | C8orf12 | 0 | intronic | 0.907 | 6 | 13 | 14 | 1 | 1 | 0 |
| 8:11236809:C:T | C8orf12 | 0 | intronic | 5.18 | 6 | 5 | 14 | 1 | 1 | 0 |
| 8:11238029:C:T | C8orf12 | 0 | intronic | 4.941 | 5 | 9 | 14 | 1 | 1 | 0 |
| 8:11239640:C:T | C8orf12 | 0 | intronic | 1.125 | 3a | 13 | 14 | 1 | 1 | 0 |
| 8:11231354:G:T | C8orf12 | 0 | intronic | 4.995 | 7 | 5 | 14 | 1 | 1 | 0 |
| 8:11235605:G:T | C8orf12 | 0 | intronic | 1.698 | NA | 5 | 14 | 1 | 1 | 0 |

| uniqID | nearestGene | dist | func | CADD | RDB | minChrState | commonChrState | posMapFilt | eqtlMapFilt | ciMapFilt |
|---|---|---|---|---|---|---|---|---|---|---|
| 8:11235393:A:G | C8orf12 | 0 | intronic | 2.024 | NA | 5 | 14 | 1 | 1 | 0 |
| 8:11235579:G:T | C8orf12 | 0 | intronic | 4.501 | 7 | 5 | 14 | 1 | 1 | 0 |
| 8:11046394:C:G | XKR6 | 0 | intronic | 3.587 | 6 | 5 | 15 | 1 | 1 | 0 |
| 8:11239510:A:G | C8orf12 | 0 | intronic | 0.798 | NA | 13 | 14 | 1 | 1 | 0 |
| 8:11239565:C:T | C8orf12 | 0 | intronic | 2.887 | NA | 13 | 14 | 1 | 1 | 0 |
| 8:10903475:A:T | XKR6 | 0 | intronic | 4.33 | 7 | 5 | 15 | 1 | 1 | 0 |
| 8:11239352:A:G | C8orf12 | 0 | intronic | 4.714 | NA | 13 | 14 | 1 | 1 | 0 |
| 8:11229638:C:T | C8orf12 | 0 | intronic | 2.917 | NA | 5 | 14 | 1 | 1 | 0 |
| 8:8700851:C:T | MFHAS1 | 0 | intronic | 1.177 | 5 | 5 | 5 | 1 | 1 | 0 |
| 8:11239137:A:G | C8orf12 | 0 | intronic | 1.824 | NA | 13 | 14 | 1 | 1 | 0 |
| 8:11238597:C:T | C8orf12 | 0 | intronic | 2.289 | NA | 9 | 14 | 1 | 1 | 0 |
| 8:11061792:C:T | AF131215.8 | 611 | downstream | 6.566 | 6 | 5 | 14 | 1 | 1 | 0 |
| 8:11060311:A:C | AF131215.8 | 162 | upstream | 6.184 | 5 | 5 | 14 | 1 | 1 | 0 |
| 8:11071057:A:G | AF131215.8 | 9876 | intergenic | 0.822 | 7 | 9 | 14 | 0 | 1 | 0 |
| 8:11237773:C:T | C8orf12 | 0 | intronic | 1.65 | 5 | 9 | 14 | 1 | 1 | 0 |
| 8:8646246:C:T | MFHAS1 | 0 | intronic | 0.006 | 1d | 4 | 5 | 1 | 1 | 0 |
| 8:11235910:A:G | C8orf12 | 0 | intronic | 0.645 | NA | 5 | 14 | 1 | 1 | 0 |
| 8:11239078:G:T | C8orf12 | 0 | intronic | 0.718 | 5 | 13 | 14 | 1 | 1 | 0 |
| 8:11237756:A:C | C8orf12 | 0 | intronic | 1.401 | NA | 9 | 14 | 1 | 1 | 0 |
| 8:11239017:A:T | C8orf12 | 0 | intronic | 0.286 | 5 | 13 | 14 | 1 | 1 | 0 |
| 8:11239054:C:T | C8orf12 | 0 | intronic | 0.009 | 5 | 13 | 14 | 1 | 1 | 0 |
| 8:11079367:A:G | AF131215.8 | 18186 | intergenic | 7.258 | 7 | 12 | 14 | 0 | 1 | 0 |
| 8:11078949:A:G | AF131215.8 | 17768 | intergenic | 2.448 | 6 | 13 | 14 | 0 | 1 | 0 |
| 8:11247814:A:G | C8orf12 | 0 | intronic | 0.572 | 5 | 5 | 14 | 1 | 1 | 0 |
| 8:8633548:G:T | RP11-211C9.1 | 0 | ncRNA_intronic | 5.665 | NA | 5 | 15 | 1 | 1 | 0 |
| 8:8664622:A:G | MFHAS1 | 0 | intronic | 0.544 | 1f | 5 | 5 | 1 | 1 | 0 |
| 8:11076635:G:T | AF131215.8 | 15454 | intergenic | 1.543 | 6 | 13 | 14 | 0 | 1 | 0 |
| 8:8661534:C:T | MFHAS1 | 0 | intronic | 5.035 | 1f | 4 | 5 | 1 | 1 | 0 |
| 8:8652889:A:G | MFHAS1 | 0 | intronic | 2.542 | 6 | 4 | 5 | 1 | 1 | 0 |
| 8:11078781:A:C | AF131215.8 | 17600 | intergenic | 1.136 | 6 | 13 | 14 | 0 | 1 | 0 |
| 8:8661681:C:G | MFHAS1 | 0 | intronic | 0.053 | 7 | 4 | 5 | 1 | 1 | 0 |
| 8:8685854:A:G | MFHAS1 | 0 | intronic | 1.488 | NA | 2 | 15 | 1 | 1 | 0 |
| 8:8641145:C:T | MFHAS1 | 0 | UTR3 | 16.03 | 1f | 2 | 5 | 1 | 1 | 0 |
| 8:11219781:C:T | TDH | 0 | ncRNA_intronic | 4.25 | 6 | 2 | 14 | 1 | 1 | 0 |
| 8:8637429:A:G | RP11-211C9.1 | 0 | ncRNA_intronic | 1.071 | 2b | 2 | 7 | 1 | 1 | 0 |
| 8:8721473:A:G | MFHAS1 | 0 | intronic | 4.862 | 6 | 4 | 5 | 1 | 1 | 0 |
| 8:10987553:C:T | XKR6:AF131215.5 | 00:00:00 | UTR5 | 9.295 | 1b | 5 | 15 | 1 | 1 | 0 |
| 8:8770512:C:T | RNU6-682P | 17083 | intergenic | 0.105 | NA | 5 | 15 | 0 | 1 | 0 |
| 8:11242025:G:T | C8orf12 | 0 | intronic | 2.468 | 6 | 13 | 14 | 1 | 1 | 0 |
| 8:8678530:A:G | MFHAS1 | 0 | intronic | 5.451 | NA | 2 | 5 | 1 | 1 | 0 |
| 8:11069960:C:T | AF131215.8 | 8779 | intergenic | 5.453 | 6 | 9 | 14 | 0 | 1 | 0 |
| 8:11251175:A:G | C8orf12 | 0 | intronic | 0.986 | 6 | 7 | 14 | 1 | 1 | 0 |
| 8:11215617:C:G | TDH | 0 | ncRNA_intronic | 0.513 | 6 | 4 | 14 | 0 | 1 | 0 |
| 8:10938260:A:G | XKR6 | 0 | intronic | 4.101 | 1f | 5 | 15 | 1 | 1 | 0 |
| 8:10987651:G:T | XKR6:AF131215.5 | 00:00:00 | UTR5 | 9.078 | 5 | 5 | 15 | 1 | 1 | 0 |
| 8:11055597:A:C | XKR6 | 0 | intronic | 0.225 | 7 | 1 | 15 | 1 | 1 | 0 |
| 8:8685646:C:T | MFHAS1 | 0 | intronic | 1.281 | 1f | 2 | 5 | 1 | 1 | 0 |

| uniqID | nearestGene | dist | func | CADD | RDB | minChrState | commonChrState | posMapFilt | eqtlMapFilt | ciMapFilt |
|---|---|---|---|---|---|---|---|---|---|---|
| 8:8673601:A:C | MFHAS1 | 0 | intronic | 3.727 | 5 | 4 | 5 | 1 | 1 | 0 |
| 8:11214972:A:G | TDH | 0 | ncRNA_intronic | 0.262 | 1f | 4 | 14 | 0 | 1 | 0 |
| 8:8639740:A:G | RP11-211C9.1 | 0 | ncRNA_exonic | 0.189 | 4 | 1 | 5 | 1 | 1 | 0 |
| 8:8704330:C:G | MFHAS1 | 0 | intronic | 2.414 | 5 | 2 | 7 | 1 | 1 | 0 |
| 8:11219334:A:G | TDH | 0 | ncRNA_intronic | 0.398 | 2b | 2 | 14 | 1 | 1 | 0 |
| 8:11214455:A:G | TDH | 0 | ncRNA_intronic | 3.7 | 1f | 4 | 14 | 0 | 1 | 0 |
| 8:8682192:A:T | MFHAS1 | 0 | intronic | 5.001 | 4 | 4 | 5 | 1 | 1 | 0 |
| 8:8679614:C:T | MFHAS1 | 0 | intronic | 0.034 | 4 | 2 | 5 | 1 | 1 | 0 |
| 8:10987199:C:T | XKR6:AF131215.5 | 00:00:00 | UTR5 | 9.773 | NA | 5 | 15 | 1 | 1 | 0 |
| 8:8684953:A:G | MFHAS1 | 0 | intronic | 1.584 | NA | 5 | 5 | 1 | 1 | 0 |
| 8:8660538:A:C | MFHAS1 | 0 | intronic | 0.886 | 7 | 4 | 5 | 1 | 1 | 0 |
| 8:8673736:C:T | MFHAS1 | 0 | intronic | 2.268 | 6 | 4 | 5 | 1 | 1 | 0 |
| 8:11225168:A:G | TDH | 0 | ncRNA_intronic | 0.418 | NA | 4 | 14 | 1 | 1 | 0 |
| 8:8730488:A:G | MFHAS1 | 0 | intronic | 1.773 | NA | 2 | 7 | 1 | 1 | 0 |
| 8:11225910:A:G | TDH:C8orf12 | 00:00:00 | ncRNA_exonic | 0.293 | 1f | 5 | 14 | 1 | 1 | 0 |
| 8:8679176:A:G | MFHAS1 | 0 | intronic | 10.14 | 6 | 2 | 5 | 1 | 1 | 0 |
| 8:8680477:A:G | MFHAS1 | 0 | intronic | 3.841 | 5 | 4 | 5 | 1 | 1 | 0 |
| 8:11225480:C:T | TDH | 0 | ncRNA_intronic | 0.398 | 1f | 5 | 14 | 1 | 1 | 0 |
| 8:8682878:C:T | MFHAS1 | 0 | intronic | 3.001 | 7 | 4 | 5 | 1 | 1 | 0 |
| 8:11213589:G:T | TDH | 0 | ncRNA_intronic | 0.079 | 7 | 5 | 14 | 0 | 1 | 0 |
| 8:9569104:C:T | TNKS | 0 | intronic | 2.099 | 6 | 4 | 5 | 1 | 1 | 0 |
| 8:11070360:C:G | AF131215.8 | 9179 | intergenic | 0.57 | 7 | 9 | 13 | 0 | 1 | 0 |
| 8:11045161:A:G | XKR6 | 0 | intronic | 1.653 | 6 | 2 | 15 | 1 | 1 | 0 |
| 8:11228672:A:G | C8orf12 | 0 | intronic | 2.822 | 6 | 5 | 14 | 1 | 1 | 0 |
| 8:11054097:A:C | XKR6 | 0 | intronic | 1.367 | 5 | 2 | 15 | 1 | 1 | 0 |
| 8:11046209:C:T | XKR6 | 0 | intronic | 6.142 | 5 | 5 | 15 | 1 | 1 | 0 |
| 8:10986837:G:T | XKR6:AF131215.5 | 00:00:00 | UTR5 | 4.475 | 1f | 5 | 15 | 1 | 1 | 0 |
| 8:11041642:A:T | XKR6 | 0 | intronic | 10.77 | 7 | 5 | 15 | 1 | 1 | 0 |
| 8:11252425:A:C | C8orf12 | 0 | intronic | 0.718 | 7 | 5 | 14 | 1 | 1 | 0 |
| 8:8644274:C:G | MFHAS1 | 0 | intronic | 0.288 | 5 | 5 | 5 | 1 | 1 | 0 |
| 8:11425809:G:T | BLK | 3695 | intergenic | 3.475 | 6 | 5 | 14 | 1 | 1 | 0 |
| 8:8685190:A:G | MFHAS1 | 0 | intronic | 5.143 | NA | 5 | 5 | 1 | 1 | 0 |
| 8:8654527:C:T | MFHAS1 | 0 | intronic | 2.606 | 3a | 2 | 5 | 1 | 1 | 0 |
| 8:11039816:G:T | XKR6 | 0 | intronic | 1.235 | 6 | 4 | 15 | 1 | 1 | 0 |
| 8:8643938:C:T | MFHAS1 | 0 | intronic | 1.258 | 5 | 4 | 5 | 1 | 1 | 0 |
| 8:11228254:A:G | C8orf12 | 0 | intronic | 0.561 | NA | 5 | 14 | 1 | 1 | 0 |
| 8:8729761:C:G | MFHAS1 | 0 | intronic | 15.49 | 1b | 2 | 7 | 1 | 1 | 0 |
| 8:11037187:C:T | XKR6 | 0 | intronic | 1.406 | 7 | 5 | 15 | 1 | 1 | 0 |
| 8:11422289:C:G | BLK | 175 | downstream | 1.012 | 2b | 1 | 10 | 1 | 1 | 0 |
| 8:11086942:C:T | LINC00529 | 18192 | intergenic | 1.004 | 1f | 5 | 14 | 0 | 1 | 0 |
| 8:11247298:A:G | C8orf12 | 0 | intronic | 0.401 | 5 | 5 | 14 | 1 | 1 | 0 |
| 8:8649881:C:T | MFHAS1 | 0 | intronic | 3.127 | 7 | 4 | 5 | 1 | 1 | 0 |
| 8:8702875:C:T | MFHAS1 | 0 | intronic | 0.501 | 2b | 2 | 7 | 1 | 1 | 0 |
| 8:11033976:C:T | XKR6 | 0 | intronic | 8.343 | 6 | 5 | 15 | 1 | 1 | 0 |
| 8:8672429:C:G | MFHAS1 | 0 | intronic | 1.464 | 7 | 4 | 5 | 1 | 1 | 0 |
| 8:8670082:C:G | MFHAS1 | 0 | intronic | 7.687 | 1f | 4 | 5 | 1 | 1 | 0 |
| 8:11223022:A:C | TDH | 0 | ncRNA_intronic | 3.295 | 6 | 4 | 14 | 1 | 1 | 0 |

| uniqID | nearestGene | dist | func | CADD | RDB | minChrState | commonChrState | posMapFilt | eqtlMapFilt | ciMapFilt |
|---|---|---|---|---|---|---|---|---|---|---|
| 8:10985432:C:T | XKR6:AF131215.5 | 00:00:00 | intronic | 0.937 | 6 | 5 | 15 | 1 | 1 | 0 |
| 8:8671962:C:T | MFHAS1 | 0 | intronic | 2.474 | 1f | 4 | 5 | 1 | 1 | 0 |
| 8:10121635:A:G | MSRA | 0 | intronic | 0.878 | 5 | 5 | 15 | 1 | 1 | 0 |
| 8:11245064:C:T | C8orf12 | 0 | intronic | 2.829 | 5 | 7 | 14 | 1 | 1 | 0 |
| 8:11235614:A:G | C8orf12 | 0 | intronic | 2.104 | NA | 5 | 14 | 1 | 1 | 0 |
| 8:11415812:C:T | BLK | 0 | intronic | 3.952 | 1f | 2 | 14 | 1 | 1 | 0 |
| 8:11427637:A:T | BLK | 5523 | intergenic | 4.827 | 1f | 5 | 14 | 1 | 1 | 0 |
| 8:8658540:A:G | MFHAS1 | 0 | intronic | 4.468 | 7 | 5 | 5 | 1 | 1 | 0 |
| 8:10909193:A:C | XKR6 | 0 | intronic | 0.513 | 5 | 5 | 14 | 1 | 1 | 0 |
| 8:8592845:C:G | RP11-211C9.1 | 0 | ncRNA_intronic | 3.765 | 6 | 5 | 14 | 0 | 1 | 0 |
| 8:11073402:A:G | AF131215.8 | 12221 | intergenic | 1.258 | 7 | 9 | 13 | 0 | 1 | 0 |
| 8:8769708:C:T | RNU6-682P | 16279 | intergenic | 1.818 | NA | 5 | 15 | 0 | 1 | 0 |
| 8:8672579:A:G | MFHAS1 | 0 | intronic | 3.217 | 7 | 4 | 5 | 1 | 1 | 0 |
| 8:10982051:A:G | XKR6:AF131215.3 | 00:00:00 | ncRNA_exonic | 4.462 | 4 | 1 | 15 | 1 | 1 | 0 |
| 8:8670177:A:T | MFHAS1 | 0 | intronic | 3.257 | 5 | 4 | 5 | 1 | 1 | 0 |
| 8:11238587:A:G | C8orf12 | 0 | intronic | 4.59 | NA | 9 | 14 | 1 | 1 | 0 |
| 8:8702827:A:G | MFHAS1 | 0 | intronic | 2.907 | 1b | 2 | 7 | 1 | 1 | 0 |
| 8:10932203:C:G | XKR6 | 0 | intronic | 1.779 | 6 | 5 | 15 | 1 | 1 | 0 |
| 8:11074036:A:G | AF131215.8 | 12855 | intergenic | 1.458 | 7 | 9 | 14 | 0 | 1 | 0 |
| 8:11420221:A:G | BLK | 0 | intronic | 0.375 | 4 | 4 | 13 | 1 | 1 | 0 |
| 8:8547811:A:G | CLDN23 | 11636 | intergenic | 0.532 | 5 | 5 | 15 | 0 | 1 | 0 |
| 8:11425105:G:T | BLK | 2991 | intergenic | 0.373 | 5 | 2 | 14 | 1 | 1 | 0 |
| 8:11227104:A:G | C8orf12 | 0 | intronic | 1.426 | 5 | 5 | 14 | 1 | 1 | 0 |
| 8:11420104:C:G | BLK | 0 | intronic | 0.642 | 4 | 4 | 13 | 1 | 1 | 0 |
| 8:11223793:G:T | TDH | 0 | ncRNA_intronic | 2.87 | NA | 4 | 14 | 1 | 1 | 0 |
| 8:11244841:A:G | C8orf12 | 0 | intronic | 0.042 | 5 | 7 | 14 | 1 | 1 | 0 |
| 8:11224313:C:T | TDH | 0 | ncRNA_intronic | 0.17 | NA | 4 | 14 | 1 | 1 | 0 |
| 8:10910066:C:T | XKR6 | 0 | intronic | 5.728 | 5 | 5 | 14 | 1 | 1 | 0 |
| 8:11421358:A:C | BLK | 0 | intronic | 2.157 | 2b | 1 | 13 | 1 | 1 | 0 |
| 8:8578794:A:T | RP11-211C9.1 | 2408 | intergenic | 2.053 | 3a | 7 | 14 | 0 | 1 | 0 |
| 8:8670599:A:G | MFHAS1 | 0 | intronic | 4.763 | 7 | 4 | 5 | 1 | 1 | 0 |
| 8:10986859:C:T | XKR6:AF131215.5 | 00:00:00 | UTR5 | 0.648 | NA | 5 | 15 | 1 | 1 | 0 |
| 8:11411005:C:G | BLK:RP11-148O21.3 | 00:00:00 | ncRNA_exonic | 0.662 | 2b | 5 | 14 | 1 | 1 | 0 |
| 8:11420295:C:T | BLK | 0 | intronic | 3.616 | 4 | 4 | 13 | 1 | 1 | 0 |
| 8:11421016:A:G | BLK | 0 | intronic | 2.018 | 4 | 5 | 13 | 1 | 1 | 0 |
| 8:8578811:C:T | RP11-211C9.1 | 2391 | intergenic | 6.568 | NA | 7 | 14 | 0 | 1 | 0 |
| 8:8706332:A:C | MFHAS1 | 0 | intronic | 3.613 | 5 | 1 | 5 | 1 | 1 | 0 |
| 8:11033525:C:T | XKR6 | 0 | intronic | 6.262 | 5 | 5 | 15 | 1 | 1 | 0 |
| 8:11238316:G:T | C8orf12 | 0 | intronic | 2.607 | 5 | 9 | 14 | 1 | 1 | 0 |
| 8:11238332:G:T | C8orf12 | 0 | intronic | 5.788 | NA | 9 | 14 | 1 | 1 | 0 |
| 8:8692477:C:T | MFHAS1 | 0 | intronic | 4.006 | 6 | 5 | 15 | 1 | 1 | 0 |
| 8:8548117:A:T | CLDN23 | 11330 | intergenic | 0.38 | 6 | 5 | 15 | 0 | 1 | 0 |
| 8:8670736:A:C | MFHAS1 | 0 | intronic | 2.53 | 7 | 5 | 5 | 1 | 1 | 0 |
| 8:11421793:C:T | BLK | 0 | UTR3 | 10.12 | NA | 1 | 11 | 1 | 1 | 0 |
| 8:8640172:A:C | RP11-211C9.1 | 182 | upstream | 0.231 | NA | 1 | 5 | 1 | 1 | 0 |
| 8:10926892:A:C | XKR6 | 0 | intronic | 6.456 | 5 | 5 | 14 | 1 | 1 | 0 |
| 8:10975682:C:G | XKR6 | 0 | intronic | 0.667 | 7 | 5 | 15 | 1 | 1 | 0 |

| uniqID | nearestGene | dist | func | CADD | RDB | minChrState | commonChrState | posMapFilt | eqtlMapFilt | ciMapFilt |
|---|---|---|---|---|---|---|---|---|---|---|
| 8:11040216:C:T | XKR6 | 0 | intronic | 0.027 | 6 | 4 | 15 | 1 | 1 | 0 |
| 8:11251705:A:G | C8orf12 | 0 | intronic | 0.901 | NA | 7 | 14 | 1 | 1 | 0 |
| 8:8578120:A:T | RP11-211C9.1 | 3082 | intergenic | 1.801 | 5 | 9 | 14 | 0 | 1 | 0 |
| 8:11422130:C:T | BLK | 16 | downstream | 10.52 | 4 | 1 | 11 | 1 | 1 | 0 |
| 8:10122423:A:G | MSRA | 0 | intronic | 0.238 | 7 | 5 | 15 | 1 | 1 | 0 |
| 8:10936811:A:G | XKR6 | 0 | intronic | 7.974 | 5 | 5 | 15 | 1 | 1 | 0 |
| 8:11416171:C:T | BLK:RP11-148O21.2 | 00:00:00 | ncRNA_exonic | 0.797 | 1d | 2 | 13 | 1 | 1 | 0 |
| 8:10935368:A:C | XKR6 | 0 | intronic | 1.477 | 5 | 5 | 15 | 1 | 1 | 0 |
| 8:10936891:C:T | XKR6 | 0 | intronic | 13.99 | 6 | 5 | 15 | 1 | 1 | 0 |
| 8:11087475:A:G | LINC00529 | 17659 | intergenic | 1.164 | NA | 6 | 13 | 0 | 1 | 0 |
| 8:11428395:C:T | LINC00208 | 5426 | intergenic | 0.711 | 5 | 5 | 14 | 1 | 1 | 0 |
| 8:11416428:C:T | BLK:RP11-148O21.2 | 00:00:00 | ncRNA_exonic | 2.832 | 4 | 2 | 14 | 1 | 1 | 0 |
| 8:11422045:A:G | BLK | 0 | UTR3 | 10.74 | 2c | 1 | 11 | 1 | 1 | 0 |
| 8:11423083:A:C | BLK | 969 | downstream | 4.608 | 4 | 5 | 13 | 1 | 1 | 0 |
| 8:11416635:A:G | BLK:RP11-148O21.2 | 00:00:00 | ncRNA_intronic | 3.356 | 2b | 2 | 14 | 1 | 1 | 0 |
| 8:11421384:C:T | BLK | 0 | intronic | 6.665 | 2b | 1 | 13 | 1 | 1 | 0 |
| 8:11053922:A:G | XKR6 | 0 | intronic | 0.938 | 6 | 2 | 15 | 1 | 1 | 0 |
| 8:11415794:A:T | BLK | 0 | intronic | 12.6 | 1f | 2 | 14 | 1 | 1 | 0 |
| 8:8543324:C:T | CLDN23 | 16123 | intergenic | 3.038 | 5 | 2 | 15 | 0 | 1 | 0 |
| 8:11417493:C:T | BLK:RP11-148O21.2 | 00:00:00 | ncRNA_exonic | 4.147 | 7 | 4 | 14 | 1 | 1 | 0 |
| 8:11417150:G:T | BLK:RP11-148O21.2 | 00:00:00 | ncRNA_intronic | 3.414 | 2b | 2 | 14 | 1 | 1 | 0 |
| 8:8595104:A:T | RP11-211C9.1 | 0 | ncRNA_intronic | 3.341 | 6 | 5 | 14 | 0 | 1 | 0 |
| 8:11415597:C:T | BLK | 0 | intronic | 1.31 | 2a | 2 | 14 | 1 | 1 | 0 |
| 8:11422936:C:T | BLK | 822 | downstream | 6.434 | 3a | 2 | 13 | 1 | 1 | 0 |
| 8:10930069:A:C | XKR6 | 0 | intronic | 1.146 | 5 | 5 | 15 | 1 | 1 | 0 |
| 8:10963288:C:G | XKR6:AF131215.9 | 00:00:00 | ncRNA_exonic | 1.422 | 5 | 4 | 15 | 1 | 1 | 0 |
| 8:11426790:C:G | BLK | 4676 | intergenic | 1.063 | 1f | 5 | 14 | 1 | 1 | 0 |
| 8:8549020:A:C | CLDN23 | 10427 | intergenic | 0.637 | 6 | 5 | 15 | 0 | 1 | 0 |
| 8:10983534:A:G | XKR6:AF131215.3 | 00:00:00 | ncRNA_exonic | 0.551 | 5 | 5 | 15 | 1 | 1 | 0 |
| 8:11415572:A:G | BLK | 0 | intronic | 7 | 3a | 2 | 14 | 1 | 1 | 0 |
| 8:11422442:A:C | BLK | 328 | downstream | 8.058 | 4 | 1 | 11 | 1 | 1 | 0 |
| 8:10979561:C:G | XKR6 | 0 | intronic | 0.327 | 6 | 5 | 15 | 1 | 1 | 0 |
| 8:11422861:C:G | BLK | 747 | downstream | 1.233 | 1b | 2 | 13 | 1 | 1 | 0 |
| 8:10935082:C:T | XKR6 | 0 | intronic | 0.716 | 5 | 5 | 15 | 1 | 1 | 0 |
| 8:8598388:A:T | RP11-211C9.1 | 0 | ncRNA_intronic | 4.009 | 5 | 5 | 14 | 0 | 1 | 0 |
| 8:10960572:C:T | XKR6 | 0 | intronic | 0.232 | 6 | 5 | 15 | 1 | 1 | 0 |
| 8:10962929:C:G | XKR6:AF131215.9 | 00:00:00 | ncRNA_exonic | 2.251 | 2b | 4 | 15 | 1 | 1 | 0 |
| 8:11228006:A:G | C8orf12 | 0 | intronic | 2.856 | 5 | 5 | 14 | 1 | 1 | 0 |
| 8:11187434:C:T | SLC35G5 | 962 | upstream | 1.617 | 6 | 4 | 15 | 1 | 1 | 0 |
| 8:10962800:C:T | XKR6:AF131215.9 | 00:00:00 | ncRNA_exonic | 1.253 | NA | 5 | 15 | 1 | 1 | 0 |
| 8:11229889:A:G | C8orf12 | 0 | intronic | 0.115 | 6 | 5 | 14 | 1 | 1 | 0 |
| 8:10932695:C:G | XKR6 | 0 | intronic | 0.134 | 5 | 5 | 15 | 1 | 1 | 0 |
| 8:11419852:C:T | BLK | 0 | intronic | 0.213 | 2b | 4 | 13 | 1 | 1 | 0 |
| 8:11186674:A:G | MTMR9 | 1027 | intergenic | 1.684 | 5 | 5 | 15 | 1 | 1 | 0 |
| 8:11040647:A:G | XKR6 | 0 | intronic | 1.146 | 6 | 4 | 15 | 1 | 1 | 0 |
| 8:10976569:A:G | XKR6 | 0 | intronic | 4.532 | 7 | 5 | 15 | 1 | 1 | 0 |
| 8:11417144:C:T | BLK:RP11-148O21.2 | 00:00:00 | ncRNA_intronic | 6.135 | 2b | 2 | 14 | 1 | 1 | 0 |

| uniqID | nearestGene | dist | func | CADD | RDB | minChrState | commonChrState | posMapFilt | eqtlMapFilt | ciMapFilt |
|---|---|---|---|---|---|---|---|---|---|---|
| 8:11425081:G:T | BLK | 2967 | intergenic | 4.213 | 5 | 2 | 14 | 1 | 1 | 0 |
| 8:10935898:A:G | XKR6 | 0 | intronic | 0.29 | 5 | 5 | 15 | 1 | 1 | 0 |
| 8:11415184:C:G | BLK:RP11-148O21.4 | 00:00:00 | ncRNA_intronic | 1.476 | 3a | 2 | 14 | 1 | 1 | 0 |
| 8:11427133:G:T | BLK | 5019 | intergenic | 0.308 | 3a | 5 | 14 | 1 | 1 | 0 |
| 8:8768895:A:G | RNU6-682P | 15466 | intergenic | 1.316 | NA | 5 | 15 | 0 | 1 | 0 |
| 8:8578229:A:G | RP11-211C9.1 | 2973 | intergenic | 0.116 | 5 | 7 | 14 | 0 | 1 | 0 |
| 8:8578067:C:T | RP11-211C9.1 | 3135 | intergenic | 2.144 | 5 | 9 | 14 | 0 | 1 | 0 |
| 8:11189488:C:T | SLC35G5 | 0 | exonic | 6.484 | 1f | 5 | 9 | 1 | 1 | 0 |
| 8:11423072:A:G | BLK | 958 | downstream | 7.57 | 4 | 5 | 13 | 1 | 1 | 0 |
| 8:11238315:C:T | C8orf12 | 0 | intronic | 4.801 | 5 | 9 | 14 | 1 | 1 | 0 |
| 8:11417257:A:G | BLK:RP11-148O21.2 | 00:00:00 | ncRNA_intronic | 1.129 | 4 | 4 | 14 | 1 | 1 | 0 |
| 8:11204165:C:T | TDH:AF131216.5 | 00:00:00 | ncRNA_intronic | 1.208 | NA | 1 | 13 | 0 | 1 | 0 |
| 8:11426400:G:T | BLK | 4286 | intergenic | 3.736 | 3a | 5 | 14 | 1 | 1 | 0 |
| 8:11188532:C:G | SLC35G5 | 0 | UTR5 | 4.766 | 5 | 5 | 15 | 1 | 1 | 0 |
| 8:10933699:A:T | XKR6 | 0 | intronic | 4.451 | 7 | 5 | 15 | 1 | 1 | 0 |
| 8:11219386:G:T | TDH | 0 | ncRNA_intronic | 1.856 | 4 | 2 | 14 | 1 | 1 | 0 |
| 8:10176506:A:T | MSRA | 0 | intronic | 0.045 | 5 | 5 | 15 | 1 | 1 | 0 |
| 8:11208903:A:G | TDH | 0 | ncRNA_intronic | 0.953 | NA | 2 | 14 | 0 | 1 | 0 |
| 8:11422170:A:G | BLK | 56 | downstream | 8.243 | 4 | 1 | 11 | 1 | 1 | 0 |
| 8:11207367:A:C | TDH | 0 | ncRNA_intronic | 0.318 | 5 | 2 | 13 | 0 | 1 | 0 |
| 8:11031472:C:T | XKR6 | 0 | intronic | 0.606 | 7 | 5 | 15 | 1 | 1 | 0 |
| 8:11205602:C:T | TDH | 0 | ncRNA_intronic | 8.019 | 2a | 1 | 11 | 0 | 1 | 0 |
| 8:11204755:C:T | TDH:AF131216.5 | 00:00:00 | ncRNA_intronic | 8.921 | 4 | 1 | 11 | 0 | 1 | 0 |
| 8:10910343:C:G | XKR6 | 0 | intronic | 0.481 | 2b | 5 | 14 | 1 | 1 | 0 |
| 8:11217441:A:G | TDH | 0 | ncRNA_intronic | 3.963 | NA | 4 | 14 | 1 | 1 | 0 |
| 8:11423781:C:G | BLK | 1667 | intergenic | 2.101 | 4 | 5 | 13 | 1 | 1 | 0 |
| 8:11423537:A:G | BLK | 1423 | intergenic | 1.13 | 2b | 5 | 13 | 1 | 1 | 0 |
| 8:10979821:A:G | XKR6 | 0 | intronic | 0.52 | 6 | 5 | 15 | 1 | 1 | 0 |
| 8:10935366:C:T | XKR6 | 0 | intronic | 3.97 | 5 | 5 | 15 | 1 | 1 | 0 |
| 8:11252170:C:T | C8orf12 | 0 | intronic | 0.099 | 1f | 5 | 14 | 1 | 1 | 0 |
| 8:11418385:A:G | BLK | 0 | intronic | 3.446 | 4 | 4 | 13 | 1 | 1 | 0 |
| 8:11425077:C:T | BLK | 2963 | intergenic | 0.451 | 5 | 2 | 14 | 1 | 1 | 0 |
| 8:11032228:G:T | XKR6 | 0 | intronic | 7.268 | 6 | 5 | 15 | 1 | 1 | 0 |
| 8:11216761:G:T | TDH | 0 | ncRNA_exonic | 13.95 | 6 | 4 | 14 | 1 | 1 | 0 |
| 8:11186639:C:G | MTMR9 | 992 | downstream | 0.869 | 5 | 5 | 15 | 1 | 1 | 0 |
| 8:11221313:C:T | TDH | 0 | ncRNA_intronic | 0.445 | 7 | 4 | 14 | 1 | 1 | 0 |
| 8:11186453:C:T | MTMR9 | 806 | downstream | 0.35 | 5 | 5 | 15 | 1 | 1 | 0 |
| 8:8687325:A:G | MFHAS1 | 0 | intronic | 0.63 | 6 | 5 | 15 | 1 | 1 | 0 |
| 8:11211302:G:T | TDH | 0 | ncRNA_intronic | 2.824 | NA | 5 | 14 | 0 | 1 | 0 |
| 8:11423434:A:G | BLK | 1320 | intergenic | 7.415 | 3a | 5 | 13 | 1 | 1 | 0 |
| 8:10970773:A:T | XKR6 | 0 | intronic | 2.962 | 7 | 5 | 15 | 1 | 1 | 0 |
| 8:11199938:A:C | TDH | 0 | ncRNA_intronic | 1.801 | 5 | 5 | 14 | 0 | 1 | 0 |
| 8:8581408:C:G | RP11-211C9.1 | 0 | ncRNA_intronic | 2.184 | 5 | 7 | 13 | 0 | 1 | 0 |
| 8:11213250:C:T | TDH | 0 | ncRNA_intronic | 0.197 | 4 | 5 | 14 | 0 | 1 | 0 |
| 8:11218893:A:G | TDH | 0 | ncRNA_splicing | 14.74 | 5 | 2 | 14 | 1 | 1 | 0 |
| 8:9489417:A:G | TNKS | 0 | intronic | 1.908 | 6 | 5 | 15 | 1 | 1 | 0 |
| 8:8699757:A:T | MFHAS1 | 0 | intronic | 0.057 | 7 | 1 | 5 | 1 | 1 | 0 |

| uniqID | nearestGene | dist | func | CADD | RDB | minChrState | commonChrState | posMapFilt | eqtlMapFilt | ciMapFilt |
|---|---|---|---|---|---|---|---|---|---|---|
| 8:11418773:C:T | BLK | 0 | intronic | 5.889 | NA | 4 | 13 | 1 | 1 | 0 |
| 8:10932868:A:G | XKR6 | 0 | intronic | 0.127 | 1f | 5 | 15 | 1 | 1 | 0 |
| 8:8691622:A:T | MFHAS1 | 0 | intronic | 2.747 | 7 | 5 | 15 | 1 | 1 | 0 |
| 8:8699761:C:T | MFHAS1 | 0 | intronic | 0.311 | 7 | 1 | 5 | 1 | 1 | 0 |
| 8:8713038:C:T | MFHAS1 | 0 | intronic | 0.806 | 6 | 4 | 5 | 1 | 1 | 0 |
| 8:11213363:C:T | TDH | 0 | ncRNA_intronic | 4.861 | 4 | 5 | 14 | 0 | 1 | 0 |
| 8:11416885:C:T | BLK:RP11-148O21.2 | 00:00:00 | ncRNA_intronic | 1.527 | 4 | 2 | 14 | 1 | 1 | 0 |
| 8:11237330:C:G | C8orf12 | 0 | intronic | 4.354 | 6 | 5 | 14 | 1 | 1 | 0 |
| 8:11213389:C:T | TDH | 0 | ncRNA_intronic | 13.3 | 4 | 5 | 14 | 0 | 1 | 0 |
| 8:8589117:C:T | RP11-211C9.1 | 0 | ncRNA_intronic | 5.356 | NA | 13 | 14 | 0 | 1 | 0 |
| 8:11434176:A:T | LINC00208 | 0 | ncRNA_exonic | 2.916 | 5 | 5 | 14 | 0 | 1 | 0 |
| 8:11466745:A:T | LINC00208 | 27893 | intergenic | 2.164 | 6 | 5 | 14 | 0 | 1 | 0 |
| 8:11435927:C:G | LINC00208 | 0 | ncRNA_exonic | 3.254 | 5 | 5 | 14 | 0 | 1 | 0 |
| 8:11434232:C:T | LINC00208 | 0 | ncRNA_exonic | 0.306 | 5 | 5 | 14 | 0 | 1 | 0 |
| 8:11435049:A:G | LINC00208 | 0 | ncRNA_intronic | 0.072 | 5 | 5 | 14 | 0 | 1 | 0 |
| 8:8696449:G:T | MFHAS1 | 0 | intronic | 0.068 | 5 | 5 | 15 | 1 | 1 | 0 |
| 8:11250848:A:C | C8orf12 | 0 | intronic | 1.532 | 4 | 7 | 14 | 1 | 1 | 0 |
| 8:8548801:A:G | CLDN23 | 10646 | intergenic | 0.129 | 6 | 5 | 15 | 0 | 1 | 0 |
| 8:11032240:C:T | XKR6 | 0 | intronic | 1.397 | 6 | 5 | 15 | 1 | 1 | 0 |
| 8:10976494:C:G | XKR6 | 0 | intronic | 0.565 | 6 | 5 | 15 | 1 | 1 | 0 |
| 8:11435564:C:T | LINC00208 | 0 | ncRNA_intronic | 2.009 | 5 | 5 | 14 | 0 | 1 | 0 |
| 8:10939490:G:T | XKR6 | 0 | intronic | 1.783 | 5 | 2 | 15 | 1 | 1 | 0 |
| 8:11073578:C:T | AF131215.8 | 12397 | intergenic | 0.008 | 6 | 9 | 13 | 0 | 1 | 0 |
| 8:8544808:C:T | CLDN23 | 14639 | intergenic | 2.513 | NA | 1 | 15 | 0 | 1 | 0 |
| 8:11430485:A:G | LINC00208 | 3336 | intergenic | 0.543 | 4 | 2 | 14 | 1 | 1 | 0 |
| 8:11433909:C:T | LINC00208 | 0 | ncRNA_exonic | 3.539 | 5 | 5 | 14 | 0 | 1 | 0 |
| 8:8544872:A:G | CLDN23 | 14575 | intergenic | 5.216 | 6 | 1 | 15 | 0 | 1 | 0 |
| 8:11080014:C:T | AF131215.8 | 18833 | intergenic | 0.214 | 6 | 13 | 14 | 0 | 1 | 0 |
| 8:11423142:A:G | BLK | 1028 | intergenic | 4.855 | 4 | 5 | 13 | 1 | 1 | 0 |
| 8:8584344:A:G | RP11-211C9.1 | 0 | ncRNA_intronic | 2.425 | NA | 13 | 14 | 0 | 1 | 0 |
| 8:11192593:A:G | SLC35G5 | 2875 | intergenic | 1.855 | 6 | 5 | 15 | 1 | 1 | 0 |
| 8:11432946:A:C | LINC00208 | 875 | upstream | 2.867 | 7 | 5 | 14 | 0 | 1 | 0 |
| 8:11427341:G:T | BLK | 5227 | intergenic | 0.701 | 1f | 5 | 14 | 1 | 1 | 0 |
| 8:11213092:A:G | TDH | 0 | ncRNA_intronic | 0.89 | 4 | 5 | 14 | 0 | 1 | 0 |
| 8:9492453:C:T | TNKS | 0 | intronic | 0.86 | 7 | 5 | 15 | 1 | 1 | 0 |
| 8:11248956:C:T | C8orf12 | 0 | intronic | 3.157 | 5 | 2 | 14 | 1 | 1 | 0 |
| 8:8587571:G:T | RP11-211C9.1 | 0 | ncRNA_intronic | 0.377 | NA | 13 | 14 | 0 | 1 | 0 |
| 8:9525325:C:T | TNKS | 0 | intronic | 2.067 | 7 | 5 | 15 | 1 | 1 | 0 |
| 8:11196970:C:T | TDH | 175 | upstream | 2.749 | 7 | 5 | 15 | 1 | 1 | 0 |
| 8:9492426:G:T | TNKS | 0 | intronic | 6.813 | 7 | 5 | 15 | 1 | 1 | 0 |
| 8:8549432:A:G | CLDN23 | 10015 | intergenic | 1.549 | 7 | 2 | 15 | 0 | 1 | 0 |
| 8:10148447:A:G | MSRA | 0 | intronic | 6.046 | 5 | 5 | 15 | 1 | 1 | 0 |
| 8:11213881:C:G | TDH | 0 | ncRNA_intronic | 0.823 | 6 | 4 | 14 | 0 | 1 | 0 |
| 8:11435516:C:T | LINC00208 | 0 | ncRNA_intronic | 1.67 | 5 | 5 | 14 | 0 | 1 | 0 |
| 8:11438064:G:T | LINC00208 | 0 | ncRNA_exonic | 0.484 | 5 | 5 | 14 | 0 | 1 | 0 |
| 8:8583872:C:T | RP11-211C9.1 | 0 | ncRNA_intronic | 2.972 | 6 | 13 | 14 | 0 | 1 | 0 |
| 8:8596731:G:T | RP11-211C9.1 | 0 | ncRNA_intronic | 1.585 | 6 | 5 | 14 | 0 | 1 | 0 |

| uniqID | nearestGene | dist | func | CADD | RDB | minChrState | commonChrState | posMapFilt | eqtlMapFilt | ciMapFilt |
|---|---|---|---|---|---|---|---|---|---|---|
| 8:8595838:G:T | RP11-211C9.1 | 0 | ncRNA_intronic | 3.232 | 5 | 5 | 14 | 0 | 1 | 0 |
| 8:8547110:C:G | CLDN23 | 12337 | intergenic | 1.581 | 1f | 2 | 15 | 0 | 1 | 0 |
| 8:10939273:G:T | XKR6 | 0 | intronic | 5.647 | 1f | 2 | 15 | 1 | 1 | 0 |
| 8:11467557:C:G | LINC00208 | 28705 | intergenic | 8.517 | 6 | 5 | 14 | 0 | 1 | 0 |
| 8:10975081:A:T | XKR6 | 0 | intronic | 3.799 | 7 | 2 | 15 | 1 | 1 | 0 |
| 8:8687054:C:G | MFHAS1 | 0 | intronic | 0.078 | 7 | 5 | 15 | 1 | 1 | 0 |
| 8:11199584:A:G | TDH | 0 | ncRNA_intronic | 1.598 | 5 | 5 | 14 | 1 | 1 | 0 |
| 8:10150070:C:G | MSRA | 0 | intronic | 1.293 | 5 | 5 | 15 | 1 | 1 | 0 |
| 8:11012977:C:T | XKR6 | 0 | intronic | 0.68 | 7 | 5 | 15 | 1 | 1 | 0 |
| 8:9527863:A:T | TNKS | 0 | intronic | 5.511 | 6 | 5 | 15 | 1 | 1 | 0 |
| 8:11433780:C:T | LINC00208 | 41 | upstream | 0.025 | 5 | 5 | 14 | 0 | 1 | 0 |
| 8:11015338:C:T | XKR6 | 0 | intronic | 5.978 | 6 | 5 | 15 | 1 | 1 | 0 |
| 8:9527869:A:G | TNKS | 0 | intronic | 2.726 | 6 | 5 | 15 | 1 | 1 | 0 |
| 8:11248500:C:T | C8orf12 | 0 | intronic | 1.825 | 5 | 5 | 14 | 1 | 1 | 0 |
| 8:11434792:C:G | LINC00208 | 0 | ncRNA_exonic | 1.963 | 5 | 5 | 14 | 0 | 1 | 0 |
| 8:11212778:C:T | TDH | 0 | ncRNA_intronic | 5.929 | 5 | 5 | 14 | 0 | 1 | 0 |
| 8:8723651:C:G | MFHAS1 | 0 | intronic | 0.712 | 1f | 4 | 5 | 1 | 1 | 0 |
| 8:11212811:A:C | TDH | 0 | ncRNA_intronic | 1.117 | NA | 5 | 14 | 0 | 1 | 0 |
| 8:11432438:C:G | LINC00208 | 1383 | intergenic | 4.309 | 5 | 5 | 14 | 0 | 1 | 0 |
| 8:11185096:C:G | MTMR9 | 0 | UTR3 | 14.92 | 7 | 4 | 5 | 1 | 1 | 0 |
| 8:11432453:C:T | LINC00208 | 1368 | intergenic | 3.286 | 5 | 5 | 14 | 0 | 1 | 0 |
| 8:11212812:C:G | TDH | 0 | ncRNA_intronic | 1.498 | 5 | 5 | 14 | 0 | 1 | 0 |
| 8:11440019:A:G | LINC00208 | 1167 | intergenic | 2.025 | 7 | 5 | 14 | 0 | 1 | 0 |
| 8:11029039:G:T | XKR6 | 0 | intronic | 0.176 | 5 | 4 | 15 | 1 | 1 | 0 |
| 8:10149212:A:C | MSRA | 0 | intronic | 12.79 | 5 | 5 | 15 | 1 | 1 | 0 |
| 8:8524474:C:G | AC087269.2 | 33415 | intergenic | 0.307 | 6 | 5 | 15 | 0 | 1 | 0 |
| 8:11182455:A:G | MTMR9:AF131216.6 | 00:00:00 | ncRNA_intronic | 0.238 | 7 | 4 | 5 | 1 | 1 | 0 |
| 8:11198792:A:G | TDH | 0 | ncRNA_intronic | 2.13 | 7 | 5 | 14 | 1 | 1 | 0 |
| 8:9527707:A:G | TNKS | 0 | intronic | 0.807 | 5 | 5 | 15 | 1 | 1 | 0 |
| 8:11212875:C:G | TDH | 0 | ncRNA_intronic | 0.27 | 5 | 5 | 14 | 0 | 1 | 0 |
| 8:11430990:C:T | LINC00208 | 2831 | intergenic | 0.042 | 1d | 2 | 14 | 1 | 1 | 0 |
| 8:11023997:C:G | XKR6 | 0 | intronic | 0.096 | 7 | 5 | 15 | 1 | 1 | 0 |
| 8:9494732:G:T | TNKS | 0 | intronic | 1.74 | 5 | 1 | 15 | 1 | 1 | 0 |
| 8:11212599:C:T | TDH | 0 | ncRNA_exonic | 1.786 | NA | 5 | 14 | 0 | 1 | 0 |
| 8:10975629:A:C | XKR6 | 0 | intronic | 0.511 | 7 | 5 | 15 | 1 | 1 | 0 |
| 8:11177126:A:C | MTMR9:AF131216.6 | 00:00:00 | ncRNA_exonic | 0.057 | 7 | 4 | 4 | 1 | 1 | 0 |
| 8:11461111:A:G | LINC00208 | 22259 | intergenic | 0.311 | 6 | 5 | 14 | 0 | 1 | 0 |
| 8:8724276:C:T | MFHAS1 | 0 | intronic | 4.798 | NA | 4 | 5 | 1 | 1 | 0 |
| 8:11014616:G:T | XKR6 | 0 | intronic | 0.896 | 5 | 5 | 15 | 1 | 1 | 0 |
| 8:11240571:C:T | C8orf12 | 0 | intronic | 1.086 | 7 | 13 | 14 | 1 | 1 | 0 |
| 8:11020313:G:T | XKR6 | 0 | intronic | 2.826 | 5 | 2 | 15 | 1 | 1 | 0 |
| 8:11179458:G:T | MTMR9:AF131216.6 | 00:00:00 | ncRNA_intronic | 2.907 | NA | 4 | 4 | 1 | 1 | 0 |
| 8:8726804:G:T | MFHAS1 | 0 | intronic | 6.512 | 1d | 1 | 5 | 1 | 1 | 0 |
| 8:11444516:A:G | LINC00208 | 5664 | intergenic | 0.019 | 5 | 5 | 14 | 0 | 1 | 0 |
| 8:11210824:G:T | TDH | 0 | ncRNA_intronic | 2.595 | 7 | 5 | 14 | 0 | 1 | 0 |
| 8:11447093:A:G | LINC00208 | 8241 | intergenic | 0.836 | 1b | 1 | 14 | 0 | 1 | 0 |
| 8:11024663:A:C | XKR6 | 0 | intronic | 3.013 | 7 | 5 | 15 | 1 | 1 | 0 |

| uniqID | nearestGene | dist | func | CADD | RDB | minChrState | commonChrState | posMapFilt | eqtlMapFilt | ciMapFilt |
|---|---|---|---|---|---|---|---|---|---|---|
| 8:10987967:A:G | XKR6 | 0 | intronic | 0.223 | 6 | 5 | 15 | 1 | 1 | 0 |
| 8:10989057:A:G | XKR6 | 0 | intronic | 0.796 | 6 | 5 | 15 | 1 | 1 | 0 |
| 8:8724257:C:T | MFHAS1 | 0 | intronic | 0.004 | NA | 4 | 5 | 1 | 1 | 0 |
| 8:10974917:C:T | XKR6 | 0 | intronic | 0.214 | 6 | 2 | 15 | 1 | 1 | 0 |
| 8:11410513:A:C | BLK | 0 | intronic | 2.599 | 3a | 4 | 14 | 1 | 1 | 0 |
| 8:8707197:C:G | MFHAS1 | 0 | intronic | 11.1 | 1f | 2 | 5 | 1 | 1 | 0 |
| 8:8547313:A:C | CLDN23 | 12134 | intergenic | 0.412 | 5 | 5 | 15 | 0 | 1 | 0 |
| 8:11400628:C:G | BLK | 0 | intronic | 1.923 | 5 | 5 | 15 | 1 | 1 | 0 |
| 8:11444837:C:T | LINC00208 | 5985 | intergenic | 0.37 | 5 | 5 | 14 | 0 | 1 | 0 |
| 8:11210823:G:T | TDH | 0 | ncRNA_intronic | 3.326 | NA | 5 | 14 | 0 | 1 | 0 |
| 8:10989521:A:G | XKR6 | 0 | intronic | 1.299 | 5 | 5 | 15 | 1 | 1 | 0 |
| 8:8724415:C:T | MFHAS1 | 0 | intronic | 0.266 | 5 | 4 | 5 | 1 | 1 | 0 |
| 8:10964969:A:T | XKR6 | 0 | intronic | 3.513 | 7 | 4 | 15 | 1 | 1 | 0 |
| 8:11109269:C:T | LINC00529 | 0 | ncRNA_intronic | 5.097 | 6 | 2 | 14 | 0 | 1 | 0 |
| 8:10989206:C:T | XKR6 | 0 | intronic | 0.034 | 7 | 5 | 15 | 1 | 1 | 0 |
| 8:11178093:A:C | MTMR9:AF131216.6 | 00:00:00 | ncRNA_exonic | 2.319 | 7 | 4 | 4 | 1 | 1 | 0 |
| 8:10996089:A:G | XKR6:AF131215.4 | 00:00:00 | ncRNA_exonic | 1.477 | 7 | 5 | 15 | 1 | 1 | 0 |
| 8:8702607:C:G | MFHAS1 | 0 | intronic | 0.87 | 1f | 2 | 5 | 1 | 1 | 0 |
| 8:8603160:A:G | RP11-211C9.1 | 0 | ncRNA_intronic | 7.021 | 5 | 5 | 15 | 0 | 1 | 0 |
| 8:11019578:C:G | XKR6 | 0 | intronic | 12.47 | 5 | 1 | 15 | 1 | 1 | 0 |
| 8:8602344:C:T | RP11-211C9.1 | 0 | ncRNA_intronic | 6.657 | 2b | 5 | 15 | 0 | 1 | 0 |
| 8:10990164:A:G | XKR6 | 0 | intronic | 0.827 | 7 | 5 | 15 | 1 | 1 | 0 |
| 8:11030892:A:G | XKR6 | 0 | intronic | 1.376 | 6 | 5 | 15 | 1 | 1 | 0 |
| 8:10964906:C:T | XKR6 | 0 | intronic | 1.332 | 7 | 4 | 15 | 1 | 1 | 0 |
| 8:10992605:A:C | XKR6 | 0 | intronic | 0.079 | 7 | 5 | 15 | 1 | 1 | 0 |
| 8:11450422:A:G | LINC00208 | 11570 | intergenic | 1.775 | NA | 5 | 14 | 0 | 1 | 0 |
| 8:11013025:A:C | XKR6 | 0 | intronic | 3.158 | 7 | 5 | 15 | 1 | 1 | 0 |
| 8:10992883:C:T | XKR6 | 0 | intronic | 2.877 | 7 | 5 | 15 | 1 | 1 | 0 |
| 8:11188540:A:G | SLC35G5 | 0 | UTR5 | 7.904 | 5 | 5 | 15 | 1 | 1 | 0 |
| 8:11206363:C:G | TDH | 0 | ncRNA_intronic | 2.289 | 2b | 1 | 13 | 0 | 1 | 0 |
| 8:9568369:A:G | TNKS | 0 | intronic | 2.31 | 6 | 4 | 5 | 1 | 0 | 0 |
| 8:8725126:G:T | MFHAS1 | 0 | intronic | 0.21 | 3b | 4 | 5 | 1 | 1 | 0 |
| 8:8722675:C:T | MFHAS1 | 0 | intronic | 0.77 | NA | 4 | 5 | 1 | 1 | 0 |
| 8:10990672:C:T | XKR6 | 0 | intronic | 10.12 | 4 | 5 | 15 | 1 | 1 | 0 |
| 8:11024275:C:T | XKR6 | 0 | intronic | 3.118 | 7 | 5 | 15 | 1 | 1 | 0 |
| 8:8725229:A:G | MFHAS1 | 0 | intronic | 0.663 | 5 | 4 | 5 | 1 | 1 | 0 |
| 8:10975733:A:G | XKR6 | 0 | intronic | 0.367 | 7 | 5 | 15 | 1 | 1 | 0 |
| 8:10994743:C:G | XKR6:AF131215.4 | 00:00:00 | ncRNA_exonic | 3.883 | 4 | 5 | 15 | 1 | 1 | 0 |
| 8:11024326:A:C | XKR6 | 0 | intronic | 2.053 | 6 | 5 | 15 | 1 | 1 | 0 |
| 8:10953874:A:G | XKR6 | 0 | intronic | 3.045 | 5 | 5 | 15 | 1 | 1 | 0 |
| 8:10969075:C:T | XKR6 | 0 | intronic | 0.076 | 6 | 4 | 15 | 1 | 1 | 0 |
| 8:11016889:A:G | XKR6 | 0 | intronic | 9.357 | 3a | 2 | 15 | 1 | 1 | 0 |
| 8:10983921:C:G | XKR6 | 0 | intronic | 0.089 | 6 | 5 | 15 | 1 | 1 | 0 |
| 8:11176403:A:G | MTMR9:AF131216.6 | 00:00:00 | ncRNA_exonic | 2.582 | 6 | 4 | 5 | 1 | 1 | 0 |
| 8:11072020:A:G | AF131215.8 | 10839 | intergenic | 1.899 | 6 | 7 | 13 | 0 | 1 | 0 |
| 8:10953092:C:T | XKR6 | 0 | intronic | 0.934 | 1f | 5 | 15 | 1 | 1 | 0 |
| 8:11023655:C:T | XKR6 | 0 | intronic | 0.225 | 7 | 5 | 15 | 1 | 1 | 0 |

| uniqID | nearestGene | dist | func | CADD | RDB | minChrState | commonChrState | posMapFilt | eqtlMapFilt | ciMapFilt |
|---|---|---|---|---|---|---|---|---|---|---|
| 8:11030935:C:T | XKR6 | 0 | intronic | 1.584 | 6 | 5 | 15 | 1 | 1 | 0 |
| 8:11022106:C:G | XKR6 | 0 | intronic | 4.349 | 6 | 2 | 15 | 1 | 1 | 0 |
| 8:10841858:A:G | XKR6 | 0 | intronic | 3.723 | 7 | 14 | 14 | 1 | 1 | 0 |
| 8:10842659:G:T | XKR6 | 0 | intronic | 0.558 | 7 | 14 | 14 | 1 | 1 | 0 |
| 8:11022185:C:T | XKR6 | 0 | intronic | 0.972 | 6 | 2 | 15 | 1 | 1 | 0 |
| 8:8719513:A:G | MFHAS1 | 0 | intronic | 0.455 | 4 | 2 | 5 | 1 | 1 | 0 |
| 8:11336781:A:G | FAM167A | 4556 | intergenic | 0.231 | 3a | 5 | 15 | 1 | 1 | 0 |
| 8:11434929:C:T | LINC00208 | 0 | ncRNA_intronic | 0.875 | 5 | 5 | 14 | 0 | 1 | 0 |
| 8:8718850:A:G | MFHAS1 | 0 | intronic | 0.563 | 6 | 4 | 5 | 1 | 1 | 0 |
| 8:9496118:A:T | TNKS | 0 | intronic | 8.138 | 6 | 5 | 15 | 1 | 1 | 0 |
| 8:10839803:C:T | XKR6 | 0 | intronic | 1.947 | 7 | 9 | 14 | 1 | 1 | 0 |
| 8:11033737:A:G | XKR6 | 0 | intronic | 1.049 | 6 | 5 | 15 | 1 | 1 | 0 |
| 8:10993904:C:T | XKR6 | 0 | intronic | 1.881 | 7 | 5 | 15 | 1 | 1 | 0 |
| 8:10950757:C:G | XKR6 | 0 | intronic | 0.015 | 7 | 5 | 15 | 1 | 1 | 0 |
| 8:10950866:C:T | XKR6 | 0 | intronic | 3.266 | 5 | 5 | 15 | 1 | 1 | 0 |
| 8:10957243:A:G | XKR6 | 0 | intronic | 0.551 | 6 | 5 | 15 | 1 | 1 | 0 |
| 8:11392093:A:C | BLK | 0 | intronic | 3.651 | NA | 2 | 14 | 1 | 1 | 0 |
| 8:11309192:C:T | FAM167A | 0 | intronic | 10.02 | 3a | 5 | 15 | 1 | 1 | 0 |
| 8:11038244:A:T | XKR6 | 0 | intronic | 1.995 | 7 | 5 | 15 | 1 | 1 | 0 |
| 8:11043236:C:T | XKR6 | 0 | intronic | 0.426 | 5 | 5 | 15 | 1 | 1 | 0 |
| 8:10955225:C:G | XKR6 | 0 | intronic | 0.174 | 6 | 5 | 15 | 1 | 1 | 0 |
| 8:11034859:A:G | XKR6 | 0 | intronic | 2.613 | 1d | 5 | 15 | 1 | 1 | 0 |
| 8:11037034:A:G | XKR6 | 0 | intronic | 0.081 | 1f | 5 | 15 | 1 | 1 | 0 |
| 8:10958824:C:T | XKR6 | 0 | intronic | 5.397 | 6 | 5 | 15 | 1 | 1 | 0 |
| 8:11065003:A:C | AF131215.8 | 3822 | intergenic | 0.106 | 7 | 5 | 13 | 1 | 1 | 0 |
| 8:10944809:G:T | XKR6 | 0 | intronic | 1.283 | 6 | 5 | 15 | 1 | 1 | 0 |
| 8:9394053:G:T | RP11-375N15.2 | 18602 | intergenic | 1.673 | 7 | 5 | 15 | 0 | 1 | 0 |
| 8:10985140:A:G | XKR6:AF131215.5 | 00:00:00 | intronic | 1.52 | 5 | 5 | 15 | 1 | 1 | 0 |
| 8:10819854:C:T | XKR6 | 0 | intronic | 2.275 | 5 | 5 | 15 | 1 | 1 | 0 |
| 8:10992252:C:G | XKR6 | 0 | intronic | 0.791 | 7 | 5 | 15 | 1 | 1 | 0 |
| 8:11041661:C:T | XKR6 | 0 | intronic | 11.96 | 7 | 5 | 15 | 1 | 1 | 0 |
| 8:11036799:C:G | XKR6 | 0 | intronic | 1.041 | 7 | 5 | 15 | 1 | 1 | 0 |
| 8:9574830:G:T | TNKS | 0 | intronic | 6.472 | 7 | 4 | 5 | 1 | 0 | 0 |
| 8:10816772:A:G | XKR6 | 0 | intronic | 3.992 | 5 | 5 | 15 | 1 | 1 | 0 |
| 8:11044689:A:G | XKR6 | 0 | intronic | 7.705 | 5 | 5 | 15 | 1 | 1 | 0 |
| 8:11043926:C:T | XKR6 | 0 | intronic | 2.035 | 5 | 5 | 15 | 1 | 1 | 0 |
| 8:9584598:C:G | TNKS | 0 | intronic | 0.539 | 6 | 4 | 5 | 1 | 0 | 0 |
| 8:9575445:A:G | TNKS | 0 | intronic | 1.93 | 6 | 4 | 5 | 1 | 0 | 0 |
| 8:10828909:C:T | XKR6 | 0 | intronic | 2.26 | 7 | 5 | 15 | 1 | 1 | 0 |
| 8:11036843:C:T | XKR6 | 0 | intronic | 5.53 | 6 | 5 | 15 | 1 | 1 | 0 |
| 8:9569109:C:T | TNKS | 0 | intronic | 2.722 | 6 | 4 | 5 | 1 | 0 | 0 |
| 8:11029029:G:T | XKR6 | 0 | intronic | 1.178 | 5 | 4 | 15 | 1 | 1 | 0 |
| 8:11043138:C:T | XKR6 | 0 | intronic | 3.122 | 5 | 5 | 15 | 1 | 1 | 0 |
| 8:11038885:A:T | XKR6 | 0 | intronic | 1.307 | 7 | 5 | 15 | 1 | 1 | 0 |
| 8:11036919:A:G | XKR6 | 0 | intronic | 11.72 | 5 | 5 | 15 | 1 | 1 | 0 |
| 8:9583872:C:T | TNKS | 0 | intronic | 9.427 | 6 | 4 | 5 | 1 | 0 | 0 |
| 8:11074812:C:T | AF131215.8 | 13631 | intergenic | 1.156 | 7 | 13 | 14 | 0 | 1 | 0 |

| uniqID | nearestGene | dist | func | CADD | RDB | minChrState | commonChrState | posMapFilt | eqtlMapFilt | ciMapFilt |
|---|---|---|---|---|---|---|---|---|---|---|
| 8:11396856:A:C | BLK | 0 | intronic | 1.911 | 1d | 2 | 15 | 1 | 1 | 0 |
| 8:11188586:C:T | SLC35G5 | 0 | UTR5 | 8.567 | 5 | 5 | 15 | 1 | 1 | 0 |
| 8:10817197:C:T | XKR6 | 0 | intronic | 1.116 | 5 | 5 | 15 | 1 | 1 | 0 |
| 8:9586062:A:T | TNKS | 0 | intronic | 2.722 | 6 | 1 | 5 | 1 | 0 | 0 |
| 8:9593309:A:C | TNKS | 0 | intronic | 2.031 | 5 | 4 | 4 | 1 | 0 | 0 |
| 8:11034028:A:G | XKR6 | 0 | intronic | 7.031 | 6 | 5 | 15 | 1 | 1 | 0 |
| 8:10813904:G:T | XKR6 | 0 | intronic | 3.109 | 5 | 5 | 15 | 1 | 1 | 0 |
| 8:10815754:C:G | XKR6 | 0 | intronic | 14.04 | 5 | 5 | 15 | 1 | 1 | 0 |
| 8:10831868:G:T | XKR6 | 0 | intronic | 0.088 | 2b | 5 | 14 | 1 | 1 | 0 |
| 8:11212081:A:G | TDH | 0 | ncRNA_intronic | 2.264 | 5 | 5 | 14 | 0 | 1 | 0 |
| 8:10810451:A:G | XKR6 | 0 | intronic | 3.584 | 7 | 5 | 15 | 1 | 1 | 0 |
| 8:11039159:C:G | XKR6 | 0 | intronic | 3.977 | 7 | 5 | 15 | 1 | 1 | 0 |
| 8:11382367:A:G | BLK | 0 | intronic | 0.042 | NA | 1 | 14 | 1 | 1 | 0 |
| 8:11446637:A:G | LINC00208 | 7785 | intergenic | 0.282 | 5 | 5 | 14 | 0 | 1 | 0 |
| 8:10813197:C:T | XKR6 | 0 | intronic | 2.464 | 6 | 5 | 15 | 1 | 1 | 0 |
| 8:11187675:C:T | SLC35G5 | 721 | upstream | 0.354 | 5 | 5 | 15 | 1 | 1 | 0 |
| 8:11021682:A:G | XKR6 | 0 | intronic | 8.265 | 4 | 2 | 15 | 1 | 1 | 0 |
| 8:11207672:C:G | TDH | 0 | ncRNA_intronic | 0.255 | 4 | 2 | 13 | 0 | 1 | 0 |
| 8:10818657:A:G | XKR6 | 0 | intronic | 3.434 | 1f | 5 | 15 | 1 | 1 | 0 |
| 8:11207431:C:T | TDH | 0 | ncRNA_intronic | 1.138 | 5 | 2 | 13 | 0 | 1 | 0 |
| 8:11207508:C:T | TDH | 0 | ncRNA_intronic | 6.239 | 5 | 2 | 13 | 0 | 1 | 0 |
| 8:10962099:A:T | XKR6 | 0 | intronic | 5.806 | 6 | 5 | 15 | 1 | 1 | 0 |
| 8:11207326:A:C | TDH | 0 | ncRNA_intronic | 1.448 | 5 | 2 | 13 | 0 | 1 | 0 |
| 8:10818607:A:G | XKR6 | 0 | intronic | 4.565 | 5 | 5 | 15 | 1 | 1 | 0 |
| 8:11206543:A:G | TDH | 0 | ncRNA_intronic | 0.484 | 2b | 1 | 13 | 0 | 1 | 0 |
| 8:11205654:A:T | TDH | 0 | ncRNA_intronic | 9.436 | 2b | 1 | 11 | 0 | 1 | 0 |
| 8:11206220:C:G | TDH | 0 | ncRNA_intronic | 4.87 | 3a | 1 | 13 | 0 | 1 | 0 |
| 8:11206627:C:G | TDH | 0 | ncRNA_intronic | 6.061 | 2b | 1 | 13 | 0 | 1 | 0 |
| 8:11037903:G:T | XKR6 | 0 | intronic | 1.457 | 1f | 5 | 15 | 1 | 1 | 0 |
| 8:10837568:C:T | XKR6 | 0 | intronic | 1.276 | 7 | 9 | 14 | 1 | 1 | 0 |
| 8:11205665:C:G | TDH | 0 | ncRNA_intronic | 4.565 | 2b | 1 | 11 | 0 | 1 | 0 |
| 8:11206262:C:T | TDH | 0 | ncRNA_intronic | 7.808 | 4 | 1 | 13 | 0 | 1 | 0 |
| 8:11185671:G:T | MTMR9 | 24 | downstream | 7.622 | 5 | 5 | 15 | 1 | 1 | 0 |
| 8:10982410:C:G | XKR6:AF131215.3 | 00:00:00 | ncRNA_intronic | 0.649 | 5 | 1 | 15 | 1 | 1 | 0 |
| 8:10837019:A:G | XKR6 | 0 | intronic | 2.3 | 7 | 7 | 14 | 1 | 1 | 0 |
| 8:10988138:A:C | XKR6 | 0 | intronic | 5.227 | 6 | 5 | 15 | 1 | 1 | 0 |
| 8:10961433:C:T | XKR6 | 0 | intronic | 1.648 | 7 | 5 | 15 | 1 | 1 | 0 |
| 8:10837569:C:G | XKR6 | 0 | intronic | 0.182 | 7 | 9 | 14 | 1 | 1 | 0 |
| 8:10945439:A:G | XKR6 | 0 | intronic | 6.536 | 5 | 5 | 15 | 1 | 1 | 0 |
| 8:11204184:A:G | TDH:AF131216.5 | 00:00:00 | ncRNA_intronic | 4.89 | 5 | 1 | 13 | 0 | 1 | 0 |
| 8:11109303:C:G | LINC00529 | 0 | ncRNA_intronic | 0.469 | 6 | 2 | 14 | 0 | 1 | 0 |
| 8:11209499:C:T | TDH | 0 | ncRNA_intronic | 9.636 | NA | 5 | 14 | 0 | 1 | 0 |
| 8:11056388:C:T | XKR6 | 0 | intronic | 3.66 | 5 | 1 | 15 | 1 | 1 | 0 |
| 8:11205593:C:T | TDH | 0 | ncRNA_intronic | 9.341 | 2b | 1 | 10 | 0 | 1 | 0 |
| 8:11188752:A:G | SLC35G5 | 0 | exonic | 22 | 5 | 5 | 15 | 1 | 1 | 0 |
| 8:11204532:A:G | TDH:AF131216.5 | 00:00:00 | ncRNA_intronic | 5.608 | 4 | 1 | 13 | 0 | 1 | 0 |
| 8:10837414:A:C | XKR6 | 0 | intronic | 1.612 | 5 | 9 | 14 | 1 | 1 | 0 |

| uniqID | nearestGene | dist | func | CADD | RDB | minChrState | commonChrState | posMapFilt | eqtlMapFilt | ciMapFilt |
|---|---|---|---|---|---|---|---|---|---|---|
| 8:11447119:C:T | LINC00208 | 8267 | intergenic | 3.088 | 4 | 1 | 14 | 0 | 1 | 0 |
| 8:10836024:C:T | XKR6 | 0 | intronic | 7.428 | 6 | 5 | 14 | 1 | 1 | 0 |
| 8:10968926:A:G | XKR6 | 0 | intronic | 2.187 | 6 | 4 | 15 | 1 | 1 | 0 |
| 8:10835917:C:T | XKR6 | 0 | intronic | 5.889 | 6 | 5 | 14 | 1 | 1 | 0 |
| 8:11202960:A:G | TDH | 0 | ncRNA_intronic | 1.895 | 4 | 1 | 13 | 0 | 1 | 0 |
| 8:11203107:A:T | TDH | 0 | ncRNA_intronic | 2.258 | 4 | 1 | 13 | 0 | 1 | 0 |
| 8:10836069:C:T | XKR6 | 0 | intronic | 1.915 | 7 | 5 | 14 | 1 | 1 | 0 |
| 8:11202154:A:G | TDH | 0 | ncRNA_intronic | 3.985 | 4 | 5 | 13 | 0 | 1 | 0 |
| 8:11210983:A:G | TDH | 0 | ncRNA_intronic | 2.152 | 6 | 5 | 14 | 0 | 1 | 0 |
| 8:10811829:C:T | XKR6 | 0 | intronic | 0.783 | 5 | 5 | 15 | 1 | 1 | 0 |
| 8:10952500:A:T | XKR6 | 0 | intronic | 0.494 | 7 | 5 | 15 | 1 | 1 | 0 |
| 8:11211068:C:T | TDH | 0 | ncRNA_intronic | 8.026 | 6 | 5 | 14 | 0 | 1 | 0 |
| 8:11446868:C:T | LINC00208 | 8016 | intergenic | 7.794 | 2b | 2 | 14 | 0 | 1 | 0 |
| 8:10836508:A:G | XKR6 | 0 | intronic | 4.494 | 5 | 7 | 14 | 1 | 1 | 0 |
| 8:11056175:A:G | XKR6 | 0 | intronic | 5.555 | 1f | 1 | 15 | 1 | 1 | 0 |
| 8:11187078:G:T | SLC35G5 | 1318 | intergenic | 0.495 | 3a | 4 | 15 | 1 | 1 | 0 |
| 8:11182148:C:G | MTMR9:AF131216.6 | 00:00:00 | ncRNA_intronic | 3.447 | 1f | 4 | 5 | 1 | 1 | 0 |
| 8:10981003:A:C | XKR6:AF131215.3 | 00:00:00 | ncRNA_exonic | 0.53 | 5 | 2 | 15 | 1 | 1 | 0 |
| 8:8665147:A:G | MFHAS1 | 0 | intronic | 1.321 | 1f | 5 | 5 | 1 | 1 | 0 |
| 8:10836436:C:T | XKR6 | 0 | intronic | 2.966 | 5 | 7 | 14 | 1 | 1 | 0 |
| 8:11446955:A:G | LINC00208 | 8103 | intergenic | 1.776 | 4 | 2 | 14 | 0 | 1 | 0 |
| 8:11450133:A:G | LINC00208 | 11281 | intergenic | 0.998 | 5 | 5 | 14 | 0 | 1 | 0 |
| 8:10836463:C:G | XKR6 | 0 | intronic | 3.938 | 5 | 7 | 14 | 1 | 1 | 0 |
| 8:11191537:C:T | SLC35G5 | 1819 | intergenic | 3.346 | 7 | 5 | 9 | 1 | 1 | 0 |
| 8:11184390:A:C | MTMR9 | 0 | UTR3 | 0.244 | 6 | 4 | 5 | 1 | 1 | 0 |
| 8:11446800:C:T | LINC00208 | 7948 | intergenic | 2.108 | 5 | 5 | 14 | 0 | 1 | 0 |
| 8:8725319:A:G | MFHAS1 | 0 | intronic | 0.824 | 6 | 4 | 5 | 1 | 1 | 0 |
| 8:11184937:A:G | MTMR9 | 0 | UTR3 | 7.477 | 5 | 4 | 5 | 1 | 1 | 0 |
| 8:11182704:A:G | MTMR9:AF131216.6 | 00:00:00 | ncRNA_intronic | 0.579 | 5 | 4 | 5 | 1 | 1 | 0 |
| 8:11010974:G:T | XKR6 | 0 | intronic | 7.035 | 7 | 5 | 15 | 1 | 1 | 0 |
| 8:11184396:A:G | MTMR9 | 0 | UTR3 | 0.277 | 6 | 4 | 5 | 1 | 1 | 0 |
| 8:11187651:A:G | SLC35G5 | 745 | upstream | 0.354 | 5 | 5 | 15 | 1 | 1 | 0 |
| 8:10837420:A:G | XKR6 | 0 | intronic | 5.108 | 5 | 9 | 14 | 1 | 1 | 0 |
| 8:8667444:C:T | MFHAS1 | 0 | intronic | 3.913 | 7 | 5 | 5 | 1 | 1 | 0 |
| 8:11446680:C:T | LINC00208 | 7828 | intergenic | 3.541 | 5 | 5 | 14 | 0 | 1 | 0 |
| 8:10835480:C:T | XKR6 | 0 | intronic | 1.023 | 7 | 5 | 14 | 1 | 1 | 0 |
| 8:10968550:C:G | XKR6 | 0 | intronic | 0.6 | NA | 5 | 15 | 1 | 1 | 0 |
| 8:11197301:C:G | TDH | 0 | ncRNA_intronic | 5.741 | 7 | 5 | 15 | 1 | 1 | 0 |
| 8:11193736:G:T | TDH | 3409 | intergenic | 4.232 | 5 | 7 | 15 | 1 | 1 | 0 |
| 8:8672952:A:C | MFHAS1 | 0 | intronic | 3.414 | 7 | 4 | 5 | 1 | 1 | 0 |
| 8:11190647:C:G | SLC35G5 | 929 | downstream | 1.131 | 4 | 5 | 9 | 1 | 1 | 0 |
| 8:11192551:C:G | SLC35G5 | 2833 | intergenic | 1.302 | 7 | 5 | 15 | 1 | 1 | 0 |
| 8:8673320:C:T | MFHAS1 | 0 | intronic | 2.843 | 6 | 4 | 5 | 1 | 1 | 0 |
| 8:8824858:G:T | RP11-62H7.2 | 0 | ncRNA_intronic | 2.74 | 7 | 5 | 15 | 0 | 1 | 0 |
| 8:11197323:G:T | TDH | 0 | ncRNA_intronic | 3.534 | 7 | 5 | 15 | 1 | 1 | 0 |
| 8:10836359:C:T | XKR6 | 0 | intronic | 0.384 | 5 | 5 | 14 | 1 | 1 | 0 |
| 8:11447679:C:G | LINC00208 | 8827 | intergenic | 1.295 | 4 | 2 | 14 | 0 | 1 | 0 |

| uniqID | nearestGene | dist | func | CADD | RDB | minChrState | commonChrState | posMapFilt | eqtlMapFilt | ciMapFilt |
|---|---|---|---|---|---|---|---|---|---|---|
| 8:8706209:A:C | MFHAS1 | 0 | intronic | 5.242 | 5 | 1 | 5 | 1 | 1 | 0 |
| 8:11210828:C:T | TDH | 0 | ncRNA_intronic | 2.072 | 7 | 5 | 14 | 0 | 1 | 0 |
| 8:11183505:A:G | MTMR9 | 0 | UTR3 | 0.564 | NA | 4 | 5 | 1 | 1 | 0 |
| 8:11450587:G:T | LINC00208 | 11735 | intergenic | 0.463 | 5 | 5 | 14 | 0 | 1 | 0 |
| 8:11355602:C:G | BLK | 0 | intronic | 0.254 | 1f | 2 | 15 | 1 | 1 | 0 |
| 8:11194911:A:G | TDH | 2234 | intergenic | 2.87 | 7 | 2 | 15 | 1 | 1 | 0 |
| 8:11401116:A:G | BLK | 0 | intronic | 9.069 | 5 | 5 | 15 | 1 | 1 | 0 |
| 8:11196295:C:G | TDH | 850 | upstream | 0.718 | 6 | 5 | 15 | 1 | 1 | 0 |
| 8:11439225:A:G | LINC00208 | 373 | downstream | 0.142 | 5 | 5 | 14 | 0 | 1 | 0 |
| 8:11358156:C:T | BLK | 0 | intronic | 3.056 | 7 | 5 | 15 | 1 | 1 | 0 |
| 8:11186215:A:C | MTMR9 | 568 | downstream | 1.755 | 5 | 5 | 15 | 1 | 1 | 0 |
| 8:11446421:A:G | LINC00208 | 7569 | intergenic | 2.107 | 5 | 5 | 14 | 0 | 1 | 0 |
| 8:8668917:A:C | MFHAS1 | 0 | intronic | 1.227 | 5 | 4 | 5 | 1 | 1 | 0 |
| 8:11201504:G:T | TDH | 0 | ncRNA_intronic | 1.86 | 7 | 5 | 14 | 0 | 1 | 0 |
| 8:11400680:A:G | BLK | 0 | intronic | 2.637 | NA | 5 | 15 | 1 | 1 | 0 |
| 8:8675176:A:G | MFHAS1 | 0 | intronic | 0.987 | 5 | 1 | 5 | 1 | 1 | 0 |
| 8:11400944:G:T | BLK | 0 | intronic | 8.189 | NA | 5 | 15 | 1 | 1 | 0 |
| 8:10837190:A:G | XKR6 | 0 | intronic | 8.094 | 5 | 7 | 14 | 1 | 1 | 0 |
| 8:11398865:A:G | BLK | 0 | intronic | 0.855 | 6 | 5 | 15 | 1 | 1 | 0 |
| 8:11362275:A:C | BLK | 0 | intronic | 0.265 | 5 | 5 | 15 | 1 | 1 | 0 |
| 8:8668486:A:G | MFHAS1 | 0 | intronic | 4.816 | 1f | 2 | 5 | 1 | 1 | 0 |
| 8:11399484:A:T | BLK | 0 | intronic | 0.514 | 7 | 5 | 15 | 1 | 1 | 0 |
| 8:11362277:G:T | BLK | 0 | intronic | 1.093 | 5 | 5 | 15 | 1 | 1 | 0 |
| 8:8664940:A:G | MFHAS1 | 0 | intronic | 6.776 | 1f | 5 | 5 | 1 | 1 | 0 |
| 8:11193530:C:T | TDH | 3615 | intergenic | 1.777 | 7 | 5 | 15 | 1 | 1 | 0 |
| 8:10943276:C:T | XKR6 | 0 | intronic | 0.551 | 1f | 5 | 15 | 1 | 1 | 0 |
| 8:11393764:A:G | BLK | 0 | intronic | 1.294 | NA | 5 | 14 | 1 | 1 | 0 |
| 8:11187770:C:G | SLC35G5 | 626 | upstream | 0.005 | 5 | 5 | 15 | 1 | 1 | 0 |
| 8:11194457:A:G | TDH | 2688 | intergenic | 0.36 | 4 | 2 | 15 | 1 | 1 | 0 |
| 8:11361261:C:G | BLK | 0 | intronic | 0.43 | 6 | 3 | 15 | 1 | 1 | 0 |
| 8:11111462:A:T | LINC00529 | 0 | ncRNA_intronic | 1.428 | 6 | 5 | 14 | 0 | 1 | 0 |
| 8:11174484:C:G | MTMR9:AF131216.6 | 00:00:00 | ncRNA_exonic | 0.323 | 6 | 4 | 4 | 1 | 1 | 0 |
| 8:11338146:A:G | FAM167A | 5921 | intergenic | 2.425 | 4 | 2 | 15 | 1 | 1 | 0 |
| 8:11397073:A:C | BLK | 0 | intronic | 6.644 | NA | 2 | 15 | 1 | 1 | 0 |
| 8:8666916:C:T | MFHAS1 | 0 | intronic | 1.862 | 4 | 2 | 5 | 1 | 1 | 0 |
| 8:11397086:A:T | BLK | 0 | intronic | 6.509 | 5 | 2 | 15 | 1 | 1 | 0 |
| 8:11395079:A:G | BLK | 0 | intronic | 0.367 | NA | 2 | 15 | 1 | 1 | 0 |
| 8:11448659:C:G | LINC00208 | 9807 | intergenic | 4.266 | 5 | 5 | 14 | 0 | 1 | 0 |
| 8:11396874:A:G | BLK | 0 | intronic | 0.553 | 4 | 2 | 15 | 1 | 1 | 0 |
| 8:10758213:A:G | XKR6 | 0 | intronic | 0.556 | 7 | 5 | 15 | 1 | 1 | 0 |
| 8:8663215:C:T | MFHAS1 | 0 | intronic | 1.424 | 5 | 5 | 5 | 1 | 1 | 0 |
| 8:11113089:A:G | LINC00529 | 0 | ncRNA_intronic | 0.595 | 7 | 5 | 14 | 0 | 1 | 0 |
| 8:11402347:A:G | BLK | 0 | intronic | 0.031 | 6 | 5 | 15 | 1 | 1 | 0 |
| 8:11397457:C:G | BLK | 0 | intronic | 0.433 | 5 | 2 | 15 | 1 | 1 | 0 |
| 8:11398953:C:T | BLK | 0 | intronic | 3.613 | NA | 5 | 15 | 1 | 1 | 0 |
| 8:9579318:C:T | TNKS | 0 | intronic | 3.74 | 7 | 4 | 5 | 1 | 0 | 0 |
| 8:11384556:C:T | BLK | 0 | intronic | 2.083 | NA | 5 | 14 | 1 | 1 | 0 |

| uniqID | nearestGene | dist | func | CADD | RDB | minChrState | commonChrState | posMapFilt | eqtlMapFilt | ciMapFilt |
|---|---|---|---|---|---|---|---|---|---|---|
| 8:8709971:C:T | MFHAS1 | 0 | intronic | 1.462 | 7 | 5 | 5 | 1 | 1 | 0 |
| 8:8708974:C:G | MFHAS1 | 0 | intronic | 1.073 | 5 | 1 | 7 | 1 | 1 | 0 |
| 8:8709756:C:G | MFHAS1 | 0 | intronic | 1.515 | 5 | 5 | 5 | 1 | 1 | 0 |
| 8:11237591:A:C | C8orf12 | 0 | intronic | 3.028 | NA | 5 | 14 | 1 | 1 | 0 |
| 8:10909936:C:T | XKR6 | 0 | intronic | 0.715 | 5 | 5 | 14 | 1 | 1 | 0 |
| 8:11033517:C:G | XKR6 | 0 | intronic | 1.862 | 5 | 5 | 15 | 1 | 1 | 0 |
| 8:11204503:C:G | TDH:AF131216.5 | 00:00:00 | ncRNA_intronic | 8.084 | 4 | 1 | 13 | 0 | 1 | 0 |
| 8:11217284:C:T | TDH | 0 | ncRNA_intronic | 1.709 | 7 | 4 | 14 | 1 | 1 | 0 |
| 8:11419861:G:T | BLK | 0 | intronic | 1.014 | 2b | 4 | 13 | 1 | 1 | 0 |
| 8:8675325:A:T | MFHAS1 | 0 | intronic | 6.869 | 5 | 5 | 5 | 1 | 1 | 0 |
| 8:8690787:C:T | MFHAS1 | 0 | intronic | 2.52 | 7 | 5 | 15 | 1 | 1 | 0 |
| 8:10950396:C:G | XKR6 | 0 | intronic | 2.2 | 4 | 1 | 15 | 1 | 1 | 0 |
| 8:9572099:A:G | TNKS | 0 | intronic | 5.956 | 5 | 4 | 5 | 1 | 0 | 0 |
| 8:8545624:T:TTTG | CLDN23 | 13823 | intergenic | 1.643 | NA | 5 | 15 | 0 | 1 | 0 |
| 8:8546283:A:AAATT | CLDN23 | 13164 | intergenic | 3.898 | NA | 5 | 15 | 0 | 1 | 0 |
| 8:8582155:A:AAAAAGAAAAG | RP11-211C9.1 | 0 | ncRNA_intronic | 0.435 | NA | 13 | 13 | 0 | 1 | 0 |
| 8:8582155:AAAAAG:AAAAAGAAAAG | RP11-211C9.1 | 0 | NA | NA | NA | 13 | 13 | 0 | 0 | 0 |
| 8:8644213:G:GT | MFHAS1 | 0 | intronic | 0.986 | NA | 5 | 5 | 1 | 1 | 0 |
| 8:8644213:G:GTT | MFHAS1 | 0 | NA | 0.961 | NA | 5 | 5 | 1 | 1 | 0 |
| 8:8644595:A:AT | MFHAS1 | 0 | intronic | 0.622 | NA | 5 | 5 | 1 | 1 | 0 |
| 8:8654057:A:G | MFHAS1 | 0 | intronic | 2.046 | 7 | 4 | 5 | 1 | 1 | 0 |
| 8:8661026:C:CA | MFHAS1 | 0 | intronic | 1.165 | NA | 5 | 5 | 1 | 1 | 0 |
| 8:8661114:C:CA | MFHAS1 | 0 | intronic | 1.142 | NA | 5 | 5 | 1 | 1 | 0 |
| 8:8665802:C:T | MFHAS1 | 0 | intronic | 3.787 | 3a | 5 | 5 | 1 | 1 | 0 |
| 8:8669681:C:CGTAA | MFHAS1 | 0 | intronic | 3.15 | NA | 2 | 5 | 1 | 1 | 0 |
| 8:8669681:C:T | MFHAS1 | 0 | NA | 0.033 | NA | 2 | 5 | 1 | 0 | 0 |
| 8:8670322:C:CT | MFHAS1 | 0 | intronic | 0.841 | NA | 4 | 5 | 1 | 1 | 0 |
| 8:8676626:G:GAATC | MFHAS1 | 0 | intronic | 2.03 | NA | 5 | 5 | 1 | 1 | 0 |
| 8:8679141:C:CTT | MFHAS1 | 0 | intronic | 1.974 | NA | 2 | 5 | 1 | 1 | 0 |
| 8:8698977:C:CA | MFHAS1 | 0 | intronic | 1.834 | NA | 5 | 15 | 1 | 1 | 0 |
| 8:8703781:G:GGA | MFHAS1 | 0 | intronic | 0.094 | NA | 5 | 5 | 1 | 1 | 0 |
| 8:8706130:A:AT | MFHAS1 | 0 | intronic | 1.522 | NA | 1 | 5 | 1 | 1 | 0 |
| 8:9579144:C:CT | TNKS | 0 | intronic | 0.957 | NA | 4 | 5 | 1 | 0 | 0 |
| 8:9579377:C:CA | TNKS | 0 | intronic | 2.513 | NA | 4 | 5 | 1 | 0 | 0 |
| 8:10146490:C:CT | MSRA | 0 | intronic | 1.365 | NA | 5 | 15 | 1 | 1 | 0 |
| 8:10812333:G:GT | XKR6 | 0 | intronic | 0.761 | NA | 5 | 15 | 1 | 1 | 0 |
| 8:10821056:C:CT | XKR6 | 0 | intronic | 1.424 | NA | 5 | 15 | 1 | 1 | 0 |
| 8:10927234:C:CAGTA | XKR6 | 0 | intronic | 0.504 | NA | 5 | 14 | 1 | 1 | 0 |
| 8:10945767:C:CA | XKR6 | 0 | intronic | 0.627 | NA | 5 | 15 | 1 | 1 | 0 |
| 8:10948422:C:CA | XKR6 | 0 | intronic | 0.578 | NA | 7 | 15 | 1 | 1 | 0 |
| 8:10948968:A:G | XKR6 | 0 | intronic | 0.514 | 6 | 7 | 15 | 1 | 1 | 0 |
| 8:10951175:G:GT | XKR6 | 0 | intronic | 0.28 | NA | 5 | 15 | 1 | 1 | 0 |
| 8:10955383:G:GTT | XKR6 | 0 | intronic | 0.948 | NA | 5 | 15 | 1 | 0 | 0 |
| 8:10955383:GT:GTT | XKR6 | 0 | NA | NA | NA | 5 | 15 | 1 | 1 | 0 |
| 8:10973149:C:CAA | XKR6 | 0 | intronic | 3.379 | NA | 5 | 15 | 1 | 0 | 0 |
| 8:10973149:C:CA | XKR6 | 0 | NA | 3.444 | NA | 5 | 15 | 1 | 1 | 0 |
| 8:10976571:G:T | XKR6 | 0 | intronic | 0.657 | 7 | 5 | 15 | 1 | 1 | 0 |

| uniqID | nearestGene | dist | func | CADD | RDB | minChrState | commonChrState | posMapFilt | eqtlMapFilt | ciMapFilt |
|---|---|---|---|---|---|---|---|---|---|---|
| 8:10978065:G:T | XKR6 | 0 | intronic | 1.49 | 6 | 5 | 15 | 1 | 1 | 0 |
| 8:10983579:G:T | XKR6 | 0 | intronic | 0.982 | 6 | 5 | 15 | 1 | 1 | 0 |
| 8:10990371:G:GA | XKR6 | 0 | intronic | 1.821 | NA | 5 | 15 | 1 | 1 | 0 |
| 8:10992544:A:AT | XKR6 | 0 | intronic | 0.275 | NA | 5 | 15 | 1 | 1 | 0 |
| 8:10993995:C:T | XKR6 | 0 | intronic | 1.127 | 6 | 5 | 15 | 1 | 1 | 0 |
| 8:11027491:T:TAA | XKR6 | 0 | intronic | 0.751 | NA | 5 | 15 | 1 | 1 | 0 |
| 8:11035071:G:GAGGTCATAATGGAAT | XKR6 | 0 | intronic | 6.345 | NA | 5 | 15 | 1 | 1 | 0 |
| 8:11036052:T:TAA | XKR6 | 0 | intronic | 0.389 | NA | 5 | 15 | 1 | 1 | 0 |
| 8:11041897:T:TACACACACACACAC | XKR6 | 0 | intronic | 0.421 | NA | 5 | 15 | 1 | 1 | 0 |
| 8:11042974:C:G | XKR6 | 0 | intronic | 0.038 | 7 | 5 | 15 | 1 | 0 | 0 |
| 8:11042974:G:T | XKR6 | 0 | NA | 0.036 | NA | 5 | 15 | 1 | 1 | 0 |
| 8:11060217:T:TA | AF131215.8 | 256 | upstream | 2.243 | NA | 5 | 14 | 1 | 1 | 0 |
| 8:11074365:C:CTA | AF131215.8 | 13184 | intergenic | 0.606 | NA | 13 | 14 | 0 | 1 | 0 |
| 8:11098992:A:G | LINC00529 | 6142 | intergenic | 2.279 | 4 | 9 | 14 | 0 | 1 | 0 |
| 8:11098996:C:T | LINC00529 | 6138 | intergenic | 1.865 | 4 | 9 | 14 | 0 | 1 | 0 |
| 8:11183765:C:CTTA | MTMR9 | 0 | UTR3 | 8.068 | NA | 4 | 5 | 1 | 1 | 0 |
| 8:11184478:C:CAA | MTMR9 | 0 | UTR3 | 2.867 | NA | 4 | 5 | 1 | 1 | 0 |
| 8:11185673:A:AT | MTMR9 | 26 | downstream | 7.945 | NA | 5 | 15 | 1 | 1 | 0 |
| 8:11189535:C:T | SLC35G5 | 0 | exonic | 4.662 | 5 | 5 | 9 | 1 | 1 | 0 |
| 8:11197598:C:T | TDH | 0 | ncRNA_intronic | 3.494 | 7 | 5 | 15 | 1 | 1 | 0 |
| 8:11198579:C:CTTTT | TDH | 0 | ncRNA_intronic | 0.47 | NA | 5 | 14 | 1 | 1 | 0 |
| 8:11200454:A:AAT | TDH | 0 | ncRNA_intronic | 6.214 | NA | 5 | 14 | 0 | 1 | 0 |
| 8:11201605:A:ATT | TDH | 0 | ncRNA_intronic | 0.02 | NA | 5 | 14 | 0 | 1 | 0 |
| 8:11205817:T:TA | TDH | 0 | ncRNA_intronic | 8.083 | NA | 1 | 11 | 0 | 1 | 0 |
| 8:11212650:A:AT | TDH | 0 | ncRNA_exonic | 0.321 | NA | 5 | 14 | 0 | 1 | 0 |
| 8:11215868:A:AT | TDH | 0 | ncRNA_intronic | 0.479 | NA | 4 | 14 | 0 | 1 | 0 |
| 8:11220846:C:CAA | TDH | 0 | ncRNA_intronic | 1.568 | NA | 4 | 14 | 1 | 1 | 0 |
| 8:11237587:C:CAAAA | C8orf12 | 0 | intronic | 0.904 | NA | 5 | 14 | 1 | 1 | 0 |
| 8:11239297:G:GATAG | C8orf12 | 0 | intronic | 4.732 | NA | 13 | 14 | 1 | 0 | 0 |
| 8:11239297:G:GATAT | C8orf12 | 0 | NA | 4.732 | NA | 13 | 14 | 1 | 1 | 0 |
| 8:11239942:C:CT | C8orf12 | 0 | intronic | 0.696 | NA | 13 | 14 | 1 | 1 | 0 |
| 8:11241935:G:GT | C8orf12 | 0 | intronic | 10.26 | NA | 13 | 14 | 1 | 1 | 0 |
| 8:11242039:T:TA | C8orf12 | 0 | intronic | 1.535 | NA | 13 | 14 | 1 | 1 | 0 |
| 8:11245562:A:T | C8orf12 | 0 | intronic | 0.919 | 5 | 5 | 14 | 1 | 1 | 0 |
| 8:11361850:G:GAGGA | BLK | 0 | intronic | 0.638 | NA | 5 | 15 | 1 | 1 | 0 |
| 8:11417016:A:ACT | BLK:RP11-148O21.2 | 00:00:00 | ncRNA_intronic | 2.756 | NA | 2 | 14 | 1 | 1 | 0 |
| 8:11419335:A:AT | BLK | 0 | intronic | 0.088 | NA | 4 | 14 | 1 | 1 | 0 |
| 8:11422491:A:C | BLK | 377 | downstream | 2.275 | 3a | 1 | 11 | 1 | 1 | 0 |
| 8:11422492:C:G | BLK | 378 | downstream | 0.574 | 3a | 1 | 11 | 1 | 1 | 0 |
| 8:11422494:A:AT | BLK | 380 | downstream | 0.572 | NA | 1 | 11 | 1 | 1 | 0 |
| 8:11422521:G:GA | BLK | 407 | downstream | 4.036 | NA | 1 | 11 | 1 | 1 | 0 |
| 8:11431558:T:TAC | LINC00208 | 2263 | intergenic | 0.204 | NA | 5 | 14 | 1 | 1 | 0 |
| 8:11431943:T:TAA | LINC00208 | 1878 | intergenic | 0.02 | NA | 5 | 14 | 1 | 1 | 0 |
| 8:11432085:A:ACG | LINC00208 | 1736 | intergenic | 1.509 | NA | 5 | 14 | 1 | 1 | 0 |
| 8:11432085:A:ACACG | LINC00208 | 1736 | NA | 1.587 | NA | 5 | 14 | 1 | 0 | 0 |
| 8:11434415:C:CTCGGTTT | LINC00208 | 0 | ncRNA_exonic | 4.114 | NA | 5 | 14 | 0 | 1 | 0 |
| 8:11435291:G:GTGGC | LINC00208 | 0 | ncRNA_intronic | 0.725 | NA | 5 | 14 | 0 | 1 | 0 |

| uniqID | nearestGene | dist | func | CADD | RDB | minChrState | commonChrState | posMapFilt | eqtlMapFilt | ciMapFilt |
|---|---|---|---|---|---|---|---|---|---|---|
| 8:11446652:A:G | LINC00208 | 7800 | intergenic | 1.483 | 5 | 5 | 14 | 0 | 1 | 0 |
| 8:11460909:T:TA | LINC00208 | 22057 | intergenic | 0.944 | NA | 5 | 14 | 0 | 1 | 0 |
| 12:62989110:A:C | MON2 | 0 | UTR3 | 2.64 | 7 | 4 | 5 | 1 | 0 | 0 |
| 12:62995984:C:T | C12orf61 | 0 | UTR3 | 4.366 | 4 | 1 | 2 | 1 | 1 | 0 |
| 12:62986620:A:G | MON2 | 0 | UTR3 | 5.166 | 7 | 4 | 5 | 1 | 1 | 0 |
| 12:62990403:A:G | MON2 | 0 | UTR3 | 1.187 | 6 | 5 | 5 | 1 | 0 | 0 |
| 12:62990415:C:T | MON2 | 0 | UTR3 | 5.356 | 6 | 5 | 5 | 1 | 0 | 0 |
| 12:62995340:A:G | C12orf61 | 190 | downstream | 10.81 | 3a | 1 | 5 | 1 | 1 | 0 |
| 12:62945970:C:G | MON2 | 0 | intronic | 0.022 | 7 | 4 | 5 | 1 | 1 | 0 |
| 12:62977163:A:G | MON2 | 0 | intronic | 2.991 | 7 | 4 | 5 | 1 | 1 | 0 |
| 12:62995269:A:G | C12orf61 | 261 | downstream | 3.248 | 3a | 1 | 5 | 1 | 1 | 0 |
| 12:62968280:C:T | MON2 | 0 | intronic | 3.801 | 7 | 4 | 5 | 1 | 1 | 0 |
| 12:62948736:A:G | MON2 | 0 | intronic | 1.539 | 7 | 4 | 4 | 1 | 1 | 0 |
| 12:62985871:A:C | MON2 | 0 | intronic | 1.573 | 6 | 4 | 5 | 1 | 1 | 0 |
| 12:62950556:C:G | MON2 | 0 | intronic | 0.964 | 5 | 4 | 4 | 1 | 1 | 0 |
| 12:62942258:C:T | MON2 | 0 | intronic | 3.639 | 7 | 4 | 5 | 1 | 1 | 0 |
| 12:62960880:A:G | MON2 | 0 | intronic | 0.317 | 7 | 4 | 4 | 1 | 1 | 0 |
| 12:62961299:C:T | MON2 | 0 | intronic | 0.418 | 7 | 4 | 4 | 1 | 1 | 0 |
| 12:62957906:C:G | MON2 | 0 | intronic | 1.425 | 7 | 4 | 5 | 1 | 1 | 0 |
| 12:62996061:A:G | C12orf61 | 0 | UTR3 | 7.271 | 4 | 1 | 1 | 1 | 1 | 0 |
| 12:62993793:A:G | C12orf61 | 1737 | intergenic | 2.935 | 4 | 5 | 5 | 1 | 1 | 0 |
| 12:62952696:C:T | MON2 | 0 | intronic | 0.665 | 7 | 4 | 5 | 1 | 1 | 0 |
| 12:62949110:A:C | MON2 | 0 | intronic | 3.79 | 6 | 4 | 4 | 1 | 1 | 0 |
| 12:62961801:A:G | MON2 | 0 | intronic | 14.42 | 7 | 4 | 5 | 1 | 1 | 0 |
| 12:62975307:C:T | MON2 | 0 | intronic | 8.418 | 7 | 4 | 5 | 1 | 1 | 0 |
| 12:62937532:C:T | MON2 | 0 | intronic | 0.549 | 7 | 4 | 4 | 1 | 0 | 0 |
| 12:62990871:A:C | MON2 | 0 | UTR3 | 0.274 | 6 | 5 | 5 | 1 | 1 | 0 |
| 12:62964552:C:T | MON2 | 0 | intronic | 0.725 | 7 | 4 | 5 | 1 | 1 | 0 |
| 12:62970482:A:G | MON2 | 0 | intronic | 8.697 | 3b | 4 | 5 | 1 | 1 | 0 |
| 12:62945245:A:G | MON2 | 0 | intronic | 2.468 | 6 | 4 | 5 | 1 | 1 | 0 |
| 12:62952910:C:T | MON2 | 0 | intronic | 0.387 | 7 | 4 | 5 | 1 | 1 | 0 |
| 12:62980580:A:T | MON2 | 0 | intronic | 1.964 | 7 | 4 | 5 | 1 | 1 | 0 |
| 12:62988288:A:G | MON2 | 0 | UTR3 | 3.65 | 6 | 4 | 5 | 1 | 1 | 0 |
| 12:62870574:C:G | MON2 | 0 | intronic | 12.04 | 6 | 4 | 5 | 1 | 1 | 0 |
| 12:62941837:A:G | MON2 | 0 | intronic | 0.389 | NA | 4 | 4 | 1 | 1 | 0 |
| 12:62941426:C:T | MON2 | 0 | intronic | 4.232 | 6 | 4 | 4 | 1 | 1 | 0 |
| 12:62921257:C:T | MON2 | 0 | intronic | 0.604 | 7 | 4 | 5 | 1 | 1 | 0 |
| 12:62941444:A:G | MON2 | 0 | intronic | 1.013 | 7 | 4 | 4 | 1 | 1 | 0 |
| 12:62898111:A:G | MON2 | 0 | intronic | 0.439 | 6 | 4 | 5 | 1 | 1 | 0 |
| 12:62917704:A:G | MON2 | 0 | intronic | 7.458 | 5 | 4 | 5 | 1 | 1 | 0 |
| 12:62865152:C:T | MON2 | 0 | intronic | 0.057 | 7 | 4 | 5 | 1 | 1 | 0 |
| 12:62852271:A:G | MON2 | 8325 | intergenic | 1.554 | 7 | 14 | 15 | 1 | 1 | 0 |
| 12:62847085:C:G | MON2 | 13511 | intergenic | 0.205 | 5 | 14 | 15 | 0 | 1 | 0 |
| 12:62920860:C:G | MON2 | 0 | intronic | 0.331 | 6 | 4 | 5 | 1 | 1 | 0 |
| 12:62939055:C:T | MON2 | 0 | intronic | 2.072 | 7 | 4 | 5 | 1 | 1 | 0 |
| 12:62922143:A:G | MON2 | 0 | intronic | 1.086 | 7 | 4 | 5 | 1 | 1 | 0 |
| 12:62941929:A:T | MON2 | 0 | intronic | 1.022 | 7 | 4 | 4 | 1 | 1 | 0 |

| uniqID | nearestGene | dist | func | CADD | RDB | minChrState | commonChrState | posMapFilt | eqtlMapFilt | ciMapFilt |
|---|---|---|---|---|---|---|---|---|---|---|
| 12:62886001:A:T | MON2 | 0 | intronic | 6.743 | 7 | 5 | 5 | 1 | 1 | 0 |
| 12:62902420:C:T | MON2 | 0 | intronic | 5.76 | 6 | 4 | 4 | 1 | 1 | 0 |
| 12:62904252:C:T | MON2 | 0 | intronic | 0.16 | 6 | 4 | 5 | 1 | 1 | 0 |
| 12:62868455:C:T | MON2 | 0 | intronic | 6.552 | 6 | 4 | 5 | 1 | 1 | 0 |
| 12:62884972:A:G | MON2 | 0 | intronic | 2.49 | 7 | 4 | 5 | 1 | 1 | 0 |
| 12:62932816:A:G | MON2 | 0 | intronic | 0.594 | 6 | 4 | 5 | 1 | 1 | 0 |
| 12:62888219:A:C | MON2 | 0 | intronic | 8.628 | 7 | 4 | 5 | 1 | 1 | 0 |
| 12:62930798:C:T | MON2 | 0 | intronic | 1.385 | 5 | 4 | 5 | 1 | 1 | 0 |
| 12:62865291:A:G | MON2 | 0 | intronic | 17.38 | 6 | 4 | 5 | 1 | 1 | 0 |
| 12:62880276:C:G | MON2 | 0 | intronic | 4.482 | 7 | 4 | 5 | 1 | 1 | 0 |
| 12:62858342:A:G | MON2 | 2254 | intergenic | 0.806 | 6 | 5 | 15 | 1 | 1 | 0 |
| 12:62868500:A:C | MON2 | 0 | intronic | 1.141 | 6 | 4 | 5 | 1 | 1 | 0 |
| 12:62898490:C:T | MON2 | 0 | intronic | 13.47 | 5 | 4 | 5 | 1 | 1 | 0 |
| 12:62903639:A:G | MON2 | 0 | intronic | 2.129 | 5 | 4 | 5 | 1 | 1 | 0 |
| 12:62897930:A:G | MON2 | 0 | intronic | 0.788 | 6 | 4 | 5 | 1 | 1 | 0 |
| 12:62903793:A:G | MON2 | 0 | intronic | 5.794 | 6 | 4 | 5 | 1 | 1 | 0 |
| 12:62858561:C:T | MON2 | 2035 | intergenic | 0.25 | 6 | 5 | 15 | 1 | 1 | 0 |
| 12:62875839:C:T | MON2 | 0 | intronic | 0.415 | 4 | 1 | 5 | 1 | 1 | 0 |
| 12:62894058:A:G | MON2 | 0 | intronic | 4.943 | 5 | 4 | 5 | 1 | 1 | 0 |
| 12:62926398:A:G | MON2 | 0 | exonic | 4.094 | 5 | 4 | 5 | 1 | 1 | 0 |
| 12:62862739:G:T | MON2 | 0 | intronic | 2.853 | 5 | 1 | 7 | 1 | 1 | 0 |
| 12:62890491:A:C | MON2 | 0 | intronic | 0.608 | 6 | 4 | 5 | 1 | 1 | 0 |
| 12:62848152:A:G | MON2 | 12444 | intergenic | 5.294 | 6 | 9 | 15 | 0 | 1 | 0 |
| 12:62855388:C:T | MON2 | 5208 | intergenic | 0.035 | NA | 5 | 15 | 1 | 1 | 0 |
| 12:62852916:A:G | MON2 | 7680 | intergenic | 1.166 | 4 | 9 | 15 | 1 | 1 | 0 |
| 12:62910714:A:T | MON2 | 0 | intronic | 3.425 | 6 | 4 | 5 | 1 | 1 | 0 |
| 12:62935705:C:T | MON2 | 0 | intronic | 1.512 | 7 | 4 | 5 | 1 | 1 | 0 |
| 12:62889409:A:G | MON2 | 0 | intronic | 2.917 | 7 | 4 | 5 | 1 | 1 | 0 |
| 12:62906488:A:G | MON2 | 0 | intronic | 0.206 | 6 | 4 | 5 | 1 | 1 | 0 |
| 12:62889402:A:G | MON2 | 0 | intronic | 0.547 | 7 | 4 | 5 | 1 | 1 | 0 |
| 12:62928633:A:G | MON2 | 0 | exonic | 22.2 | 7 | 4 | 5 | 1 | 1 | 0 |
| 12:62886649:C:T | MON2 | 0 | intronic | 0.64 | 6 | 5 | 5 | 1 | 1 | 0 |
| 12:62849418:A:G | MON2 | 11178 | intergenic | 5.096 | 6 | 9 | 15 | 0 | 1 | 0 |
| 12:62868499:C:T | MON2 | 0 | intronic | 0.688 | 6 | 4 | 5 | 1 | 1 | 0 |
| 12:62886372:C:G | MON2 | 0 | intronic | 1.624 | 7 | 5 | 5 | 1 | 1 | 0 |
| 12:62999154:C:G | RP11-631N16.2 | 0 | ncRNA_intronic | 6.226 | 6 | 1 | 1 | 1 | 1 | 0 |
| 12:62997180:C:T | C12orf61:RP11-631N16.2 | 00:00:00 | ncRNA_intronic | 8.091 | 4 | 1 | 1 | 1 | 1 | 0 |
| 12:62937348:G:T | MON2 | 0 | intronic | 1.592 | 5 | 4 | 5 | 1 | 1 | 0 |
| 12:62837767:A:G | MON2 | 22829 | intergenic | 8.124 | 7 | 9 | 15 | 0 | 1 | 0 |
| 12:62831343:C:T | USP15 | 20131 | intergenic | 3.851 | 6 | 5 | 15 | 0 | 1 | 0 |
| 12:62830952:C:T | USP15 | 19740 | intergenic | 0.366 | NA | 5 | 15 | 0 | 1 | 0 |
| 12:62977162:A:G | MON2 | 0 | intronic | 4.379 | 7 | 4 | 5 | 1 | 1 | 0 |
| 12:62930621:A:G | MON2 | 0 | intronic | 1.7 | 7 | 4 | 5 | 1 | 1 | 0 |
| 12:62851080:A:AC | MON2 | 9516 | intergenic | 0.161 | NA | 14 | 15 | 1 | 1 | 0 |
| 12:62859241:A:AC | MON2 | 1355 | intergenic | 0.209 | NA | 1 | 15 | 1 | 1 | 0 |
| 12:62861935:T:TA | MON2 | 0 | intronic | 4.258 | NA | 1 | 1 | 1 | 1 | 0 |
| 12:62884714:A:ATTT | MON2 | 0 | intronic | 5.955 | NA | 4 | 5 | 1 | 1 | 0 |

| uniqID | nearestGene | dist | func | CADD | RDB | minChrState | commonChrState | posMapFilt | eqtlMapFilt | ciMapFilt |
|---|---|---|---|---|---|---|---|---|---|---|
| 12:62891828:C:CT | MON2 | 0 | intronic | 2.052 | NA | 4 | 5 | 1 | 1 | 0 |
| 12:62898463:C:CT | MON2 | 0 | intronic | 4.501 | NA | 4 | 5 | 1 | 1 | 0 |
| 12:62903655:G:GT | MON2 | 0 | intronic | 0.527 | NA | 4 | 5 | 1 | 1 | 0 |
| 12:62905380:C:CA | MON2 | 0 | intronic | 11.44 | NA | 4 | 5 | 1 | 1 | 0 |
| 12:62905772:A:AC | MON2 | 0 | intronic | 0.954 | NA | 4 | 5 | 1 | 1 | 0 |
| 12:62914776:T:TA | MON2 | 0 | intronic | 0.035 | NA | 4 | 5 | 1 | 1 | 0 |
| 12:62928006:T:TA | MON2 | 0 | intronic | 9.725 | NA | 4 | 5 | 1 | 1 | 0 |
| 12:62931846:A:ATTTGT | MON2 | 0 | intronic | 5.01 | NA | 4 | 4 | 1 | 1 | 0 |
| 12:62935154:C:CA | MON2 | 0 | intronic | 0.829 | NA | 4 | 5 | 1 | 1 | 0 |
| 12:62945158:G:GGAAACAACCTGACGAT | MON2 | 0 | intronic | 0.824 | NA | 4 | 5 | 1 | 1 | 0 |
| 12:62968740:C:CA | MON2 | 0 | intronic | 1.134 | NA | 4 | 5 | 1 | 1 | 0 |
| 12:62972210:G:GT | MON2 | 0 | intronic | 6.131 | NA | 4 | 5 | 1 | 1 | 0 |
| 12:62977175:A:AG | MON2 | 0 | intronic | 0.663 | NA | 4 | 5 | 1 | 1 | 0 |
| 12:62982152:C:CT | MON2 | 0 | intronic | 0.434 | NA | 4 | 5 | 1 | 1 | 0 |
| 12:62992896:G:GA | MON2 | 1532 | intergenic | 0.466 | NA | 5 | 5 | 1 | 1 | 0 |
| 12:63001068:A:AT | RP11-631N16.2 | 0 | ncRNA_intronic | 0.137 | NA | 1 | 15 | 1 | 1 | 0 |
| 13:83312341:C:T | GYG1P2 | 84924 | intergenic | 1.149 | 6 | 9 | 15 | 0 | 0 | 0 |
| 13:83311141:A:G | GYG1P2 | 86124 | intergenic | 1.455 | 7 | 9 | 15 | 0 | 0 | 0 |
| 13:83312717:C:G | GYG1P2 | 84548 | intergenic | 0.67 | 7 | 9 | 15 | 0 | 0 | 0 |
| 13:83304898:G:T | GYG1P2 | 92367 | intergenic | 0.832 | 7 | 5 | 15 | 0 | 0 | 0 |
| 13:83306586:A:G | GYG1P2 | 90679 | intergenic | 0.529 | 7 | 5 | 15 | 0 | 0 | 0 |
| 13:83300359:A:G | GYG1P2 | 96906 | intergenic | 0.331 | 7 | 5 | 15 | 0 | 0 | 0 |
| 13:83304274:C:T | GYG1P2 | 92991 | intergenic | 6.255 | 7 | 5 | 15 | 0 | 0 | 0 |
| 13:83302594:C:T | GYG1P2 | 94671 | intergenic | 0.263 | 7 | 5 | 15 | 0 | 0 | 0 |
| 13:83302795:A:T | GYG1P2 | 94470 | intergenic | 2.481 | 5 | 5 | 15 | 0 | 0 | 0 |
| 13:83300269:C:T | GYG1P2 | 96996 | intergenic | 0.096 | 7 | 5 | 15 | 0 | 0 | 0 |
| 13:83299269:C:T | GYG1P2 | 97996 | intergenic | 0.528 | 6 | 5 | 15 | 0 | 0 | 0 |
| 13:83298475:A:G | GYG1P2 | 98790 | intergenic | 2.716 | 6 | 5 | 15 | 0 | 0 | 0 |
| 13:83300084:A:G | GYG1P2 | 97181 | intergenic | 0.305 | 6 | 5 | 15 | 0 | 0 | 0 |
| 13:83300138:A:AGAGAGC | GYG1P2 | 97127 | intergenic | 0.646 | NA | 5 | 15 | 0 | 0 | 0 |
| 16:13040514:C:T | SHISA9 | 0 | intronic | 2.022 | 5 | 5 | 15 | 1 | 0 | 0 |
| 16:13041027:G:T | SHISA9 | 0 | intronic | 2.795 | 7 | 5 | 15 | 1 | 0 | 0 |
| 16:13042097:A:G | SHISA9 | 0 | intronic | 13.23 | 7 | 5 | 15 | 1 | 0 | 0 |
| 16:13039154:C:T | SHISA9 | 0 | intronic | 0.774 | 7 | 5 | 15 | 1 | 0 | 0 |
| 16:13040889:C:G | SHISA9 | 0 | intronic | 0.618 | 6 | 5 | 15 | 1 | 0 | 0 |
| 16:13043452:A:G | SHISA9 | 0 | intronic | 5.192 | 6 | 5 | 15 | 1 | 0 | 0 |
| 16:13041924:A:G | SHISA9 | 0 | intronic | 6.099 | 5 | 5 | 15 | 1 | 0 | 0 |
| 16:13039642:A:T | SHISA9 | 0 | intronic | 0.153 | 6 | 5 | 15 | 1 | 0 | 0 |
| 16:13044033:C:T | SHISA9 | 0 | intronic | 2.077 | 7 | 5 | 15 | 1 | 0 | 0 |
| 16:13030222:A:G | SHISA9 | 0 | intronic | 0.303 | NA | 5 | 15 | 1 | 0 | 0 |
| 16:13032863:A:T | SHISA9 | 0 | intronic | 6.471 | 5 | 5 | 15 | 1 | 0 | 0 |
| 16:13032351:C:G | SHISA9 | 0 | intronic | 1.596 | 7 | 5 | 15 | 1 | 0 | 0 |
| 16:13038723:G:T | SHISA9 | 0 | intronic | 4.269 | 7 | 5 | 15 | 1 | 0 | 0 |
| 16:13023207:C:T | SHISA9 | 0 | intronic | 1.744 | 7 | 5 | 15 | 1 | 0 | 0 |
| 16:13023388:A:G | SHISA9 | 0 | intronic | 0.702 | NA | 5 | 15 | 1 | 0 | 0 |
| 16:13024150:A:G | SHISA9 | 0 | intronic | 2.228 | NA | 5 | 15 | 1 | 0 | 0 |
| 16:13030875:A:G | SHISA9 | 0 | intronic | 7.135 | NA | 5 | 15 | 1 | 0 | 0 |

| uniqID | nearestGene | dist | func | CADD | RDB | minChrState | commonChrState | posMapFilt | eqtlMapFilt | ciMapFilt |
|---|---|---|---|---|---|---|---|---|---|---|
| 16:13025315:G:T | SHISA9 | 0 | intronic | 0.331 | 7 | 5 | 15 | 1 | 0 | 0 |
| 16:13032547:A:G | SHISA9 | 0 | intronic | 0.354 | NA | 5 | 15 | 1 | 0 | 0 |
| 16:13041940:A:C | SHISA9 | 0 | intronic | 0.074 | 5 | 5 | 15 | 1 | 0 | 0 |
| 16:13039646:C:T | SHISA9 | 0 | intronic | 0.227 | 6 | 5 | 15 | 1 | 0 | 0 |
| 16:13031195:A:C | SHISA9 | 0 | intronic | 1.656 | NA | 5 | 15 | 1 | 0 | 0 |
| 16:13066833:C:T | SHISA9 | 0 | intronic | 0.149 | 7 | 5 | 15 | 1 | 0 | 0 |
| 16:13041921:C:T | SHISA9 | 0 | intronic | 2.982 | 5 | 5 | 15 | 1 | 0 | 0 |
| 16:13036811:C:T | SHISA9 | 0 | intronic | 6.27 | 5 | 5 | 15 | 1 | 0 | 0 |
| 16:13026502:A:G | SHISA9 | 0 | intronic | 3.705 | NA | 5 | 15 | 1 | 0 | 0 |
| 16:13038196:A:G | SHISA9 | 0 | intronic | 2.321 | 7 | 5 | 15 | 1 | 0 | 0 |
| 16:13023394:A:G | SHISA9 | 0 | intronic | 0.612 | 6 | 5 | 15 | 1 | 0 | 0 |
| 16:13021889:C:T | SHISA9 | 0 | intronic | 5.19 | NA | 5 | 15 | 1 | 0 | 0 |
| 16:13097206:C:G | SHISA9 | 0 | intronic | 10.25 | 7 | 5 | 15 | 1 | 0 | 0 |
| 16:13038054:G:T | SHISA9 | 0 | intronic | 0.838 | 7 | 5 | 15 | 1 | 0 | 0 |
| 16:13053951:C:T | SHISA9 | 0 | intronic | 13.54 | 6 | 5 | 15 | 1 | 0 | 0 |
| 16:13044827:C:T | SHISA9 | 0 | intronic | 0.396 | 6 | 5 | 15 | 1 | 0 | 0 |
| 16:13088905:C:T | SHISA9 | 0 | intronic | 3.136 | 6 | 5 | 15 | 1 | 0 | 0 |
| 16:13047734:C:T | SHISA9 | 0 | intronic | 0.025 | 7 | 5 | 15 | 1 | 0 | 0 |
| 16:13053882:A:G | SHISA9 | 0 | intronic | 6.437 | 7 | 5 | 15 | 1 | 0 | 0 |
| 16:13047598:C:G | SHISA9 | 0 | intronic | 5.302 | 7 | 5 | 15 | 1 | 0 | 0 |
| 16:13092220:C:T | SHISA9 | 0 | intronic | 1.601 | 5 | 7 | 15 | 1 | 0 | 0 |
| 16:13047022:C:G | SHISA9 | 0 | intronic | 1.271 | 7 | 5 | 15 | 1 | 0 | 0 |
| 16:13037305:C:T | SHISA9 | 0 | intronic | 6.084 | 7 | 5 | 15 | 1 | 0 | 0 |
| 16:13098440:C:G | SHISA9 | 0 | intronic | 1.645 | 7 | 5 | 15 | 1 | 0 | 0 |
| 16:13048841:C:T | SHISA9 | 0 | intronic | 2.929 | 7 | 5 | 15 | 1 | 0 | 0 |
| 16:13049749:A:C | SHISA9 | 0 | intronic | 2.541 | 6 | 5 | 15 | 1 | 0 | 0 |
| 16:13099177:G:T | SHISA9 | 0 | intronic | 1.897 | 7 | 5 | 15 | 1 | 0 | 0 |
| 16:13048395:C:T | SHISA9 | 0 | intronic | 6.099 | 6 | 5 | 15 | 1 | 0 | 0 |
| 16:13050340:A:C | SHISA9 | 0 | intronic | 8.215 | 6 | 5 | 15 | 1 | 0 | 0 |
| 16:13098508:A:G | SHISA9 | 0 | intronic | 0.372 | 7 | 5 | 15 | 1 | 0 | 0 |
| 16:13061109:A:G | SHISA9 | 0 | intronic | 2.338 | 7 | 5 | 15 | 1 | 0 | 0 |
| 16:13054883:C:G | SHISA9 | 0 | intronic | 1.632 | 6 | 5 | 15 | 1 | 0 | 0 |
| 16:13097125:A:G | SHISA9 | 0 | intronic | 4.784 | 7 | 5 | 15 | 1 | 0 | 0 |
| 16:13052050:A:G | SHISA9 | 0 | intronic | 0.245 | 6 | 5 | 15 | 1 | 0 | 0 |
| 16:13098934:A:G | SHISA9 | 0 | intronic | 6.741 | 6 | 5 | 15 | 1 | 0 | 0 |
| 16:13049557:C:T | SHISA9 | 0 | intronic | 2.982 | 7 | 5 | 15 | 1 | 0 | 0 |
| 16:13052714:C:G | SHISA9 | 0 | intronic | 0.427 | 6 | 5 | 15 | 1 | 0 | 0 |
| 16:13053072:A:G | SHISA9 | 0 | intronic | 11.53 | 5 | 5 | 15 | 1 | 0 | 0 |
| 16:13053187:C:T | SHISA9 | 0 | intronic | 8.439 | 6 | 5 | 15 | 1 | 0 | 0 |
| 16:13049906:A:T | SHISA9 | 0 | intronic | 0.557 | 6 | 5 | 15 | 1 | 0 | 0 |
| 16:13053270:A:G | SHISA9 | 0 | intronic | 0.116 | 7 | 5 | 15 | 1 | 0 | 0 |
| 16:13051686:A:C | SHISA9 | 0 | intronic | 10.84 | 5 | 5 | 15 | 1 | 0 | 0 |
| 16:13053924:C:G | SHISA9 | 0 | intronic | 0.189 | 7 | 5 | 15 | 1 | 0 | 0 |
| 16:13053576:A:T | SHISA9 | 0 | intronic | 4.662 | 7 | 5 | 15 | 1 | 0 | 0 |
| 16:13091332:C:T | SHISA9 | 0 | intronic | 0.023 | 6 | 5 | 15 | 1 | 0 | 0 |
| 16:13054265:A:G | SHISA9 | 0 | intronic | 1.912 | 5 | 5 | 15 | 1 | 0 | 0 |
| 16:13059539:C:T | SHISA9 | 0 | intronic | 1.061 | 5 | 5 | 15 | 1 | 0 | 0 |

| uniqID | nearestGene | dist | func | CADD | RDB | minChrState | commonChrState | posMapFilt | eqtlMapFilt | ciMapFilt |
|---|---|---|---|---|---|---|---|---|---|---|
| 16:13054645:C:T | SHISA9 | 0 | intronic | 5.104 | 7 | 5 | 15 | 1 | 0 | 0 |
| 16:13055220:C:T | SHISA9 | 0 | intronic | 1.037 | 7 | 5 | 15 | 1 | 0 | 0 |
| 16:13054037:C:G | SHISA9 | 0 | intronic | 6.68 | 6 | 5 | 15 | 1 | 0 | 0 |
| 16:13054897:A:C | SHISA9 | 0 | intronic | 3.295 | 7 | 5 | 15 | 1 | 0 | 0 |
| 16:13057200:A:G | SHISA9 | 0 | intronic | 0.058 | 7 | 5 | 15 | 1 | 0 | 0 |
| 16:13052846:A:G | SHISA9 | 0 | intronic | 5.403 | 7 | 5 | 15 | 1 | 0 | 0 |
| 16:13055766:C:T | SHISA9 | 0 | intronic | 11.99 | 7 | 5 | 15 | 1 | 0 | 0 |
| 16:13077901:A:G | SHISA9 | 0 | intronic | 5.813 | 7 | 5 | 15 | 1 | 0 | 0 |
| 16:13062232:A:G | SHISA9 | 0 | intronic | 0.804 | 7 | 5 | 15 | 1 | 0 | 0 |
| 16:13053140:C:G | SHISA9 | 0 | intronic | 7.368 | 5 | 5 | 15 | 1 | 0 | 0 |
| 16:13053136:G:T | SHISA9 | 0 | intronic | 0.977 | 5 | 5 | 15 | 1 | 0 | 0 |
| 16:13077351:C:T | SHISA9 | 0 | intronic | 6.158 | 7 | 5 | 15 | 1 | 0 | 0 |
| 16:13075745:A:G | SHISA9 | 0 | intronic | 5.049 | 7 | 5 | 15 | 1 | 0 | 0 |
| 16:13097084:G:T | SHISA9 | 0 | intronic | 2.854 | 7 | 5 | 15 | 1 | 0 | 0 |
| 16:13079214:A:G | SHISA9 | 0 | intronic | 1.107 | NA | 5 | 15 | 1 | 0 | 0 |
| 16:13079463:C:T | SHISA9 | 0 | intronic | 4.718 | NA | 5 | 15 | 1 | 0 | 0 |
| 16:13089854:G:T | SHISA9 | 0 | intronic | 4.927 | 5 | 5 | 15 | 1 | 0 | 0 |
| 16:13086810:C:T | SHISA9 | 0 | intronic | 3.294 | 7 | 5 | 15 | 1 | 0 | 0 |
| 16:13099953:G:T | SHISA9 | 0 | intronic | 4.047 | 5 | 5 | 15 | 1 | 0 | 0 |
| 16:13100021:C:T | SHISA9 | 0 | intronic | 9.243 | 5 | 5 | 15 | 1 | 0 | 0 |
| 16:13096300:C:T | SHISA9 | 0 | intronic | 0.266 | 5 | 5 | 15 | 1 | 0 | 0 |
| 16:13099919:A:G | SHISA9 | 0 | intronic | 1.291 | 3a | 5 | 15 | 1 | 0 | 0 |
| 16:13089059:A:G | SHISA9 | 0 | intronic | 0.275 | 7 | 5 | 15 | 1 | 0 | 0 |
| 16:13070238:A:G | SHISA9 | 0 | intronic | 0.72 | 7 | 5 | 15 | 1 | 0 | 0 |
| 16:13099114:C:G | SHISA9 | 0 | intronic | 7.072 | 7 | 5 | 15 | 1 | 0 | 0 |
| 16:13070671:C:T | SHISA9 | 0 | intronic | 5.193 | 5 | 5 | 15 | 1 | 0 | 0 |
| 16:13079082:A:C | SHISA9 | 0 | intronic | 3.279 | NA | 5 | 15 | 1 | 0 | 0 |
| 16:13070809:C:G | SHISA9 | 0 | intronic | 0.578 | 5 | 5 | 15 | 1 | 0 | 0 |
| 16:13083055:G:T | SHISA9 | 0 | intronic | 7.401 | 6 | 5 | 15 | 1 | 0 | 0 |
| 16:13082445:A:G | SHISA9 | 0 | intronic | 2.422 | 6 | 5 | 15 | 1 | 0 | 0 |
| 16:13081923:C:G | SHISA9 | 0 | intronic | 2.457 | 7 | 5 | 15 | 1 | 0 | 0 |
| 16:13093774:G:T | SHISA9 | 0 | intronic | 2.987 | 3a | 7 | 15 | 1 | 0 | 0 |
| 16:13080515:C:T | SHISA9 | 0 | intronic | 3.566 | 7 | 5 | 15 | 1 | 0 | 0 |
| 16:13079389:G:T | SHISA9 | 0 | intronic | 3.309 | 7 | 5 | 15 | 1 | 0 | 0 |
| 16:13093778:C:T | SHISA9 | 0 | intronic | 2.641 | 3a | 7 | 15 | 1 | 0 | 0 |
| 16:13092663:C:T | SHISA9 | 0 | intronic | 4.331 | 5 | 7 | 15 | 1 | 0 | 0 |
| 16:13078044:A:T | SHISA9 | 0 | intronic | 11.07 | 5 | 5 | 15 | 1 | 0 | 0 |
| 16:13093858:C:T | SHISA9 | 0 | intronic | 5.85 | 5 | 7 | 15 | 1 | 0 | 0 |
| 16:13092100:A:G | SHISA9 | 0 | intronic | 4.241 | 6 | 7 | 15 | 1 | 0 | 0 |
| 16:13112335:C:T | SHISA9 | 0 | intronic | 1.172 | 5 | 5 | 15 | 1 | 0 | 0 |
| 16:13098762:A:C | SHISA9 | 0 | intronic | 2.113 | 7 | 5 | 15 | 1 | 0 | 0 |
| 16:13094769:G:T | SHISA9 | 0 | intronic | 7.55 | 5 | 7 | 15 | 1 | 0 | 0 |
| 16:13095296:A:T | SHISA9 | 0 | intronic | 4.542 | 5 | 5 | 15 | 1 | 0 | 0 |
| 16:13095739:A:C | SHISA9 | 0 | intronic | 9.648 | 6 | 5 | 15 | 1 | 0 | 0 |
| 16:13095171:C:T | SHISA9 | 0 | intronic | 7.956 | 7 | 7 | 15 | 1 | 0 | 0 |
| 16:13077915:A:G | SHISA9 | 0 | intronic | 6.365 | 6 | 5 | 15 | 1 | 0 | 0 |
| 16:13095142:C:G | SHISA9 | 0 | intronic | 9.344 | 7 | 7 | 15 | 1 | 0 | 0 |

| uniqID | nearestGene | dist | func | CADD | RDB | minChrState | commonChrState | posMapFilt | eqtlMapFilt | ciMapFilt |
|---|---|---|---|---|---|---|---|---|---|---|
| 16:13105091:A:C | SHISA9 | 0 | intronic | 0.902 | 6 | 5 | 15 | 1 | 0 | 0 |
| 16:13109594:G:T | SHISA9 | 0 | intronic | 9.678 | 5 | 5 | 15 | 1 | 0 | 0 |
| 16:13094897:C:G | SHISA9 | 0 | intronic | 3.361 | 6 | 7 | 15 | 1 | 0 | 0 |
| 16:13105863:C:T | SHISA9 | 0 | intronic | 1.341 | 6 | 5 | 15 | 1 | 0 | 0 |
| 16:13104924:C:T | SHISA9 | 0 | intronic | 0.494 | 6 | 5 | 15 | 1 | 0 | 0 |
| 16:13118299:A:T | SHISA9 | 0 | intronic | 12.68 | 5 | 7 | 15 | 1 | 0 | 0 |
| 16:13105998:C:T | SHISA9 | 0 | intronic | 2.094 | 6 | 5 | 15 | 1 | 0 | 0 |
| 16:13080252:A:G | SHISA9 | 0 | intronic | 0.757 | 6 | 5 | 15 | 1 | 0 | 0 |
| 16:13115755:A:G | SHISA9 | 0 | intronic | 3.883 | 7 | 2 | 15 | 1 | 0 | 0 |
| 16:13078807:C:T | SHISA9 | 0 | intronic | 0.588 | NA | 5 | 15 | 1 | 0 | 0 |
| 16:13115702:C:G | SHISA9 | 0 | intronic | 1.687 | 7 | 2 | 15 | 1 | 0 | 0 |
| 16:13079698:A:T | SHISA9 | 0 | intronic | 0.363 | 7 | 5 | 15 | 1 | 0 | 0 |
| 16:13101555:C:T | SHISA9 | 0 | intronic | 0.07 | NA | 5 | 15 | 1 | 0 | 0 |
| 16:13101618:A:G | SHISA9 | 0 | intronic | 4.209 | 6 | 5 | 15 | 1 | 0 | 0 |
| 16:13102906:A:T | SHISA9 | 0 | intronic | 1.494 | 6 | 5 | 15 | 1 | 0 | 0 |
| 16:13022033:A:AT | SHISA9 | 0 | intronic | 0.394 | NA | 5 | 15 | 1 | 0 | 0 |
| 16:13029711:G:GA | SHISA9 | 0 | intronic | 1.02 | NA | 5 | 15 | 1 | 0 | 0 |
| 16:13035206:T:TA | SHISA9 | 0 | intronic | 0.103 | NA | 5 | 15 | 1 | 0 | 0 |
| 16:13045117:C:CA | SHISA9 | 0 | intronic | 0.646 | NA | 5 | 15 | 1 | 0 | 0 |
| 16:13048916:G:GT | SHISA9 | 0 | intronic | 0.18 | NA | 5 | 15 | 1 | 0 | 0 |
| 16:13049490:T:TA | SHISA9 | 0 | intronic | 3.55 | NA | 5 | 15 | 1 | 0 | 0 |
| 16:13053457:A:AC | SHISA9 | 0 | intronic | 0.367 | NA | 5 | 15 | 1 | 0 | 0 |
| 16:13055621:T:TCC | SHISA9 | 0 | intronic | 2.733 | NA | 5 | 15 | 1 | 0 | 0 |
| 16:13058241:G:GT | SHISA9 | 0 | intronic | 1.705 | NA | 5 | 15 | 1 | 0 | 0 |
| 16:13079063:C:CTT | SHISA9 | 0 | intronic | 2.194 | NA | 5 | 15 | 1 | 0 | 0 |
| 16:13083068:G:GT | SHISA9 | 0 | intronic | 0.631 | NA | 5 | 15 | 1 | 0 | 0 |
| 16:13097746:G:T | SHISA9 | 0 | intronic | 8.377 | 6 | 5 | 15 | 1 | 0 | 0 |
| 16:13097749:G:T | SHISA9 | 0 | intronic | 6.476 | 6 | 5 | 15 | 1 | 0 | 0 |
| 16:13102532:C:T | SHISA9 | 0 | intronic | 2.573 | 6 | 5 | 15 | 1 | 0 | 0 |
| 16:13116871:T:TTTTA | SHISA9 | 0 | intronic | 0.442 | NA | 5 | 15 | 1 | 0 | 0 |

The fine line separates genome-wide significant from non-significant loci. The thick line separates chromosomes. Abbreviations: chr = chromosome; pos = position; MAF = minor allele frequency; gwasP = p-value GWAS; se = standard error; r2 = correlation coefficient; IndSigSNP = rsid of independent significant SNP; dist = distance to nearest gene; func = function; CADD = Combined Annotation Dependent Depletion score; RDB = RegulomeDB score; minChrState = minimum 15-core chromatin state across 127 tissue/cell type; commonChrState = most common 15-core chromatin state across 127 tissue/cell types; Filt = filter

#### Supplementary Table S10

MAGMA gene mapping

| GENE | CHR | START | STOP | NSNPS | NPARAM | N | ZSTAT | P | SYMBOL | P_bonferroni |
|---|---|---|---|---|---|---|---|---|---|---|
| ENSG00000142599 | 1 | 8412457 | 8877702 | 1077 | 50 | 864581 | 46.28 | 0.0185 | RERE | 0.03513 |
| ENSG00000075568 | 2 | 98372799 | 98612388 | 726 | 37 | 867258 | 48.76 | 0.00543 | TMEM131 | 0.01031 |
| ENSG00000175806 | 8 | 9911778 | 10286401 | 2323 | 113 | 875864 | 66.65 | 1.32e-07 | MSRA | 2.51e-07 |
| ENSG00000171044 | 8 | 10753555 | 11058875 | 1537 | 60 | 869954 | 61.47 | 3.94e-06 | XKR6 | 7.49e-06 |
| ENSG00000147324 | 8 | 8640864 | 8751155 | 729 | 52 | 877684 | 60.59 | 6.85e-06 | MFHAS1 | 1.3e-05 |
| ENSG00000184608 | 8 | 11225911 | 11296167 | 484 | 55 | 850654 | 59.69 | 1.19e-05 | C8orf12 | 2.26e-05 |
| ENSG00000215346 | 8 | 10983980 | 10987745 | 16 | 5 | 874968 | 53.66 | 0.000402 | AF131215.5 | 0.0007629 |
| ENSG00000177710 | 8 | 11188397 | 11189717 | 12 | 3 | 799029 | 51.89 | 0.000106 | SLC35G5 | 0.002006 |
| ENSG00000136573 | 8 | 11351510 | 11422113 | 543 | 49 | 852722 | 49.87 | 0.00307 | BLK | 0.005829 |
| ENSG00000104626 | 8 | 8859657 | 8974256 | 734 | 60 | 850658 | 4.556 | 0.0261 | ERI1 | 0.04952 |
| ENSG00000168959 | 11 | 88237744 | 88799113 | 2445 | 66 | 873387 | 46.48 | 0.0168 | GRM5 | 0.03192 |
| ENSG00000061987 | 12 | 62860597 | 62991363 | 455 | 43 | 865430 | 55.47 | 0.000145 | MON2 | 0.0002757 |
| ENSG00000221949 | 12 | 62995531 | 62997214 | 9 | 4 | 900877 | 53.82 | 3.68e-05 | C12orf61 | 0.0006984 |
| ENSG00000070778 | 14 | 88932122 | 89021077 | 304 | 23 | 870589 | 51.01 | 0.00169 | PTPN21 | 0.003212 |
| ENSG00000042317 | 14 | 88851268 | 88936694 | 202 | 47 | 859970 | 48.01 | 7.88e-07 | SPATA7 | 0.01497 |
| ENSG00000237515 | 16 | 12995477 | 13334272 | 1900 | 140 | 872998 | 53.09 | 0.00055 | SHISA9 | 0.001045 |
| ENSG00000108465 | 17 | 46045176 | 46059140 | 52 | 12 | 896719 | 46.49 | 0.0167 | CDK5RAP3 | 0.03173 |

Abbreviations: CHR = chromosome; NSNPS = number of SNPs; NPARAM = number of relevant parameters used in the model; P_bonferroni = Bonferroni corrected P-value

#### Supplementary Table S11

Single-nucleus enrichment analysis for tinnitus ordered from most significant to less significant.

| Trait | Annotation | Prop._SNPs | Prop._h2 | Prop._h2_std_error | Enrichment | Enrichment_std_error | Enrichment_p | Coefficient | Coefficient_std_error | Coefficient_z-score | P | P.fdr |
|---|---|---|---|---|---|---|---|---|---|---|---|---|
| tinnitus2025 | Amygdala_excitatory | 0.1529 | 0.2269 | 0.01863 | 1.484 | 0.1219 | 7.86e-05 | 8.73e-10 | 3.06e-10 | 2.849 | 0.002191 | 0.06736 |
| tinnitus2025 | MGE_interneuron | 0.1334 | 0.2064 | 0.01902 | 1.548 | 0.1426 | 8.61e-05 | 8.51e-10 | 3.43e-10 | 2.483 | 0.006519 | 0.06736 |
| tinnitus2025 | Midbrain_derived_inhibitory | 0.1301 | 0.2014 | 0.01636 | 1.548 | 0.1257 | 8.63e-05 | 8.34e-10 | 3.23e-10 | 2.582 | 0.004905 | 0.06736 |
| tinnitus2025 | Eccentric_medium_spiny_neuron | 0.1399 | 0.2052 | 0.01816 | 1.466 | 0.1298 | 0.0002132 | 6.43e-10 | 3.07e-10 | 2.095 | 0.01809 | 0.1122 |
| tinnitus2025 | Hippocampal_dentate_gyrus | 0.1335 | 0.1894 | 0.0162 | 1.419 | 0.1213 | 0.0005105 | 6.07e-10 | 2.89e-10 | 2.103 | 0.01772 | 0.1122 |
| tinnitus2025 | Miscellaneous | 0.1457 | 0.2014 | 0.01704 | 1.382 | 0.1169 | 0.001437 | 5.84e-10 | 2.95e-10 | 1.983 | 0.0237 | 0.1224 |
| tinnitus2025 | LAMP5_LHX6_and_Chandelier | 0.1337 | 0.1961 | 0.01821 | 1.467 | 0.1363 | 0.0005798 | 6.23e-10 | 3.35e-10 | 1.861 | 0.0314 | 0.139 |
| tinnitus2025 | Astrocyte | 0.1196 | 0.1832 | 0.01593 | 1.532 | 0.1331 | 9.16e-05 | 5.34e-10 | 3.15e-10 | 1.696 | 0.04499 | 0.155 |
| tinnitus2025 | Medium_spiny_neuron | 0.1311 | 0.1822 | 0.01575 | 1.39 | 0.1201 | 0.002132 | 5.07e-10 | 2.97e-10 | 1.704 | 0.04423 | 0.155 |
| tinnitus2025 | Upper_layer_intratelencephalic | 0.1267 | 0.1753 | 0.0166 | 1.384 | 0.1311 | 0.003676 | 4.33e-10 | 3.22e-10 | 1.342 | 0.08976 | 0.2782 |
| tinnitus2025 | Hippocampal_CA4 | 0.1387 | 0.1838 | 0.01465 | 1.326 | 0.1056 | 0.002036 | 3.29e-10 | 2.64e-10 | 1.242 | 0.107 | 0.3016 |
| tinnitus2025 | Cerebellar_inhibitory | 0.1158 | 0.1605 | 0.01669 | 1.386 | 0.1441 | 0.009615 | 3.34e-10 | 3.45e-10 | 0.9699 | 0.166 | 0.3721 |
| tinnitus2025 | Deep_layer_corticothalamic_and_6b | 0.1417 | 0.1815 | 0.01645 | 1.281 | 0.1161 | 0.0137 | 2.76e-10 | 2.86e-10 | 0.9636 | 0.1676 | 0.3721 |
| tinnitus2025 | Hippocampal_CA1_3 | 0.1399 | 0.178 | 0.01651 | 1.272 | 0.118 | 0.02131 | 2.8e-10 | 2.91e-10 | 0.9619 | 0.168 | 0.3721 |
| tinnitus2025 | Mammillary_body | 0.1414 | 0.1826 | 0.01789 | 1.292 | 0.1265 | 0.02333 | 2.82e-10 | 3.18e-10 | 0.8872 | 0.1875 | 0.3875 |
| tinnitus2025 | Deep_layer_intratelencephalic | 0.1356 | 0.1769 | 0.01682 | 1.304 | 0.1241 | 0.01426 | 2.29e-10 | 3.09e-10 | 0.7417 | 0.2291 | 0.4178 |
| tinnitus2025 | Upper_rhombic_lip | 0.1047 | 0.1493 | 0.01562 | 1.425 | 0.1491 | 0.006188 | 2.83e-10 | 3.65e-10 | 0.7751 | 0.2191 | 0.4178 |
| tinnitus2025 | Deep_layer_near_projecting | 0.1317 | 0.1682 | 0.01559 | 1.277 | 0.1183 | 0.02104 | 1.97e-10 | 2.87e-10 | 0.6848 | 0.2467 | 0.4249 |
| tinnitus2025 | Oligodendrocyte_precursor | 0.1431 | 0.1723 | 0.01431 | 1.204 | 0.09998 | 0.0435 | 7.65e-11 | 2.62e-10 | 0.2923 | 0.385 | 0.6282 |
| tinnitus2025 | Lower_rhombic_lip | 0.1209 | 0.1495 | 0.01582 | 1.237 | 0.1309 | 0.07902 | 1.6e-11 | 3.13e-10 | 0.05104 | 0.4796 | 0.7435 |
| tinnitus2025 | Bergmann_glia | 0.1169 | 0.1535 | 0.0163 | 1.313 | 0.1394 | 0.02458 | -2.71e-11 | 3.37e-10 | -0.08041 | 0.532 | 0.7854 |
| tinnitus2025 | Thalamic_excitatory | 0.1334 | 0.1565 | 0.01786 | 1.173 | 0.1339 | 0.1899 | -5.4e-11 | 3.29e-10 | -0.1642 | 0.5652 | 0.7965 |
| tinnitus2025 | CGE_interneuron | 0.1231 | 0.1452 | 0.01663 | 1.18 | 0.1352 | 0.1726 | -1.23e-10 | 3.29e-10 | -0.3726 | 0.6453 | 0.8697 |
| tinnitus2025 | Choroid_plexus | 0.1219 | 0.1307 | 0.01515 | 1.072 | 0.1243 | 0.5626 | -4.93e-10 | 3.09e-10 | -1.594 | 0.9445 | 0.9996 |
| tinnitus2025 | Committed_oligodendrocyte_precursor | 0.1174 | 0.1224 | 0.01451 | 1.042 | 0.1235 | 0.7348 | -6.79e-10 | 3.17e-10 | -2.142 | 0.9839 | 0.9996 |
| tinnitus2025 | Ependymal | 0.1128 | 0.1215 | 0.01447 | 1.077 | 0.1283 | 0.5421 | -5.19e-10 | 3.13e-10 | -1.659 | 0.9514 | 0.9996 |
| tinnitus2025 | Fibroblast | 0.1201 | 0.1446 | 0.01538 | 1.205 | 0.1281 | 0.1128 | -2.78e-10 | 3.27e-10 | -0.8483 | 0.8019 | 0.9996 |
| tinnitus2025 | Microglia | 0.1035 | 0.1096 | 0.01505 | 1.058 | 0.1454 | 0.6898 | -5.16e-10 | 3.54e-10 | -1.458 | 0.9275 | 0.9996 |
| tinnitus2025 | Oligodendrocyte | 0.1106 | 0.09672 | 0.01437 | 0.8744 | 0.1299 | 0.3352 | -1.08e-09 | 3.23e-10 | -3.346 | 0.9996 | 0.9996 |
| tinnitus2025 | Splatter | 0.1212 | 0.1323 | 0.01422 | 1.091 | 0.1173 | 0.4375 | -3.29e-10 | 2.76e-10 | -1.192 | 0.8833 | 0.9996 |
| tinnitus2025 | Vascular | 0.1085 | 0.1049 | 0.0155 | 0.9671 | 0.1429 | 0.818 | -9.64e-10 | 3.71e-10 | -2.597 | 0.9953 | 0.9996 |

Line separates suggestive significant enrichments from non-enriched superclusters. Abbreviation: Prop = probability; h2 = heritability coefficient

#### Supplementary Table S12

Single nucleus enrichment analysis on hearing loss, major depressive disorder, insomnia, neuroticism, schizophrenia and bipolar disorder ordered from most significant to less significant per trait

| Trait | Supercluster | dataset_name | Supercluster_ID | disorder_type | P | P.fdr | BETA | SE | Enrichment | Coefficient_z-score | if.sig.fdr | P.adj |
|---|---|---|---|---|---|---|---|---|---|---|---|---|
| bip2021 | Deep-layer intratelencephalic | bip2021 | Deep_layer_intratelencephalic | psychiatric | 2.33e-06 | 7.23e-05 | 5.57e-09 | 1.22e-09 | 1.618 | 4.579 | yes | 0.0001084 |
| bip2021 | Eccentric medium spiny neuron | bip2021 | Eccentric_medium_spiny_neuron | psychiatric | 0.0001257 | 0.001323 | 3.96e-09 | 1.08e-09 | 1.495 | 3.661 | yes | 0.001558 |
| bip2021 | Amygdala excitatory | bip2021 | Amygdala_excitatory | psychiatric | 0.0001506 | 0.001323 | 4.41e-09 | 1.22e-09 | 1.477 | 3.614 | yes | 0.001648 |
| bip2021 | Upper-layer intratelencephalic | bip2021 | Upper_layer_intratelencephalic | psychiatric | 0.1919 | 0.3305 | 1.07e-09 | 1.23e-09 | 1.345 | 0.871 | yes | 0.001728 |
| bip2021 | Hippocampal dentate gyrus | bip2021 | Hippocampal_dentate_gyrus | psychiatric | 0.000277 | 0.001718 | 4.02e-09 | 1.16e-09 | 1.475 | 3.453 | yes | 0.00224 |
| bip2021 | Hippocampal CA1-3 | bip2021 | Hippocampal_CA1_3 | psychiatric | 0.0005018 | 0.002592 | 3.87e-09 | 1.18e-09 | 1.452 | 3.29 | yes | 0.003733 |
| bip2021 | CGE interneuron | bip2021 | CGE_interneuron | psychiatric | 0.0006325 | 0.002801 | 3.81e-09 | 1.18e-09 | 1.498 | 3.224 | yes | 0.004202 |
| bip2021 | Hippocampal CA4 | bip2021 | Hippocampal_CA4 | psychiatric | 0.0008164 | 0.003163 | 3.42e-09 | 1.09e-09 | 1.438 | 3.15 | yes | 0.005236 |
| bip2021 | MGE interneuron | bip2021 | MGE_interneuron | psychiatric | 0.001215 | 0.004183 | 3.39e-09 | 1.12e-09 | 1.455 | 3.032 | yes | 0.007287 |
| bip2021 | Cerebellar inhibitory | bip2021 | Cerebellar_inhibitory | psychiatric | 0.004476 | 0.01388 | 2.81e-09 | 1.07e-09 | 1.441 | 2.614 | yes | 0.01856 |
| bip2021 | Miscellaneous | bip2021 | Miscellaneous | psychiatric | 0.006948 | 0.01958 | 3.05e-09 | 1.24e-09 | 1.385 | 2.46 | yes | 0.02563 |
| bip2021 | LAMP5-LHX6 and Chandelier | bip2021 | LAMP5_LHX6_and_Chandelier | psychiatric | 0.01394 | 0.036 | 2.22e-09 | 1.01e-09 | 1.369 | 2.199 | yes | 0.04469 |
| bip2021 | Deep-layer near-projecting | bip2021 | Deep_layer_near_projecting | psychiatric | 0.01632 | 0.03893 | 2.33e-09 | 1.09e-09 | 1.356 | 2.136 | yes | 0.05054 |
| bip2021 | Medium spiny neuron | bip2021 | Medium_spiny_neuron | psychiatric | 0.01765 | 0.03909 | 2.3e-09 | 1.09e-09 | 1.35 | 2.105 | yes | 0.05296 |
| bip2021 | Deep-layer corticothalamic and 6b | bip2021 | Deep_layer_corticothalamic_and_6b | psychiatric | 0.02113 | 0.04367 | 2.18e-09 | 1.08e-09 | 1.326 | 2.031 | yes | 0.06047 |
| bip2021 | Thalamic excitatory | bip2021 | Thalamic_excitatory | psychiatric | 0.07125 | 0.138 | 1.67e-09 | 1.14e-09 | 1.294 | 1.467 | no | 0.1642 |
| bip2021 | Midbrain-derived inhibitory | bip2021 | Midbrain_derived_inhibitory | psychiatric | 0.1296 | 0.2363 | 1.27e-09 | 1.12e-09 | 1.28 | 1.128 | no | 0.2708 |
| bip2021 | Upper rhombic lip | bip2021 | Upper_rhombic_lip | psychiatric | 0.0001707 | 0.001323 | 4.46e-09 | 1.24e-09 | 1.541 | 3.582 | no | 0.3465 |
| bip2021 | Mammillary body | bip2021 | Mammillary_body | psychiatric | 0.6806 | 0.9999 | -4.51e-10 | 9.62e-10 | 1.119 | -0.4694 | no | 0.9705 |
| bip2021 | Astrocyte | bip2021 | Astrocyte | psychiatric | 0.9874 | 0.9999 | -2.44e-09 | 1.09e-09 | 1.095 | -2.238 | no | 1 |
| bip2021 | Bergmann glia | bip2021 | Bergmann_glia | psychiatric | 0.7876 | 0.9999 | -9.02e-10 | 1.13e-09 | 1.21 | -0.798 | no | 1 |
| bip2021 | Choroid plexus | bip2021 | Choroid_plexus | psychiatric | 0.9917 | 0.9999 | -2.48e-09 | 1.03e-09 | 1.036 | -2.396 | no | 1 |
| bip2021 | Committed oligodendrocyte precursor | bip2021 | Committed_oligodendrocyte_precursor | psychiatric | 0.9417 | 0.9999 | -1.86e-09 | 1.18e-09 | 1.137 | -1.569 | no | 1 |
| bip2021 | Ependymal | bip2021 | Ependymal | psychiatric | 0.9996 | 0.9999 | -3.44e-09 | 1.03e-09 | 0.9709 | -3.338 | no | 1 |
| bip2021 | Fibroblast | bip2021 | Fibroblast | psychiatric | 0.9995 | 0.9999 | -3.58e-09 | 1.08e-09 | 0.9945 | -3.305 | no | 1 |
| bip2021 | Lower rhombic lip | bip2021 | Lower_rhombic_lip | psychiatric | 0.9376 | 0.9999 | -1.57e-09 | 1.02e-09 | 1.067 | -1.535 | no | 1 |
| bip2021 | Microglia | bip2021 | Microglia | psychiatric | 0.9429 | 0.9999 | -2.03e-09 | 1.29e-09 | 1.087 | -1.58 | no | 1 |
| bip2021 | Oligodendrocyte | bip2021 | Oligodendrocyte | psychiatric | 0.8963 | 0.9999 | -1.19e-09 | 9.41e-10 | 1.065 | -1.261 | no | 1 |
| bip2021 | Oligodendrocyte precursor | bip2021 | Oligodendrocyte_precursor | psychiatric | 0.9844 | 0.9999 | -2.36e-09 | 1.1e-09 | 1.074 | -2.154 | no | 1 |
| bip2021 | Splatter | bip2021 | Splatter | psychiatric | 0.9792 | 0.9999 | -2.05e-09 | 1.01e-09 | 1.021 | -2.037 | no | 1 |
| bip2021 | Vascular | bip2021 | Vascular | psychiatric | 0.9999 | 0.9999 | -4.54e-09 | 1.21e-09 | 0.9335 | -3.742 | no | 1 |
| hearing_loss | Amygdala excitatory | hearing_loss | Amygdala_excitatory | neurological | 0.01333 | 0.2533 | 1.38e-09 | 6.23e-10 | 1.363 | 2.216 | yes | 0.0435 |
| hearing_loss | Hippocampal CA1-3 | hearing_loss | Hippocampal_CA1_3 | neurological | 0.01891 | 0.2533 | 1.23e-09 | 5.94e-10 | 1.359 | 2.077 | yes | 0.05582 |
| hearing_loss | Deep-layer intratelencephalic | hearing_loss | Deep_layer_intratelencephalic | neurological | 0.0256 | 0.2533 | 1.36e-09 | 6.97e-10 | 1.447 | 1.95 | yes | 0.07106 |
| hearing_loss | Deep-layer near-projecting | hearing_loss | Deep_layer_near_projecting | neurological | 0.03268 | 0.2533 | 1.09e-09 | 5.92e-10 | 1.372 | 1.843 | yes | 0.08809 |
| hearing_loss | LAMP5-LHX6 and Chandelier | hearing_loss | LAMP5_LHX6_and_Chandelier | neurological | 0.04785 | 0.254 | 8.76e-10 | 5.26e-10 | 1.36 | 1.666 | no | 0.1219 |
| hearing_loss | Upper-layer intratelencephalic | hearing_loss | Upper_layer_intratelencephalic | neurological | 0.3971 | 0.6305 | 1.53e-10 | 5.85e-10 | 1.345 | 0.2608 | no | 0.1236 |
| hearing_loss | Medium spiny neuron | hearing_loss | Medium_spiny_neuron | neurological | 0.07414 | 0.3283 | 7.54e-10 | 5.21e-10 | 1.311 | 1.446 | no | 0.1661 |
| hearing_loss | Deep-layer corticothalamic and 6b | hearing_loss | Deep_layer_corticothalamic_and_6b | neurological | 0.117 | 0.4019 | 6.43e-10 | 5.41e-10 | 1.261 | 1.19 | no | 0.2502 |
| hearing_loss | Fibroblast | hearing_loss | Fibroblast | neurological | 0.1463 | 0.4019 | 7.97e-10 | 7.57e-10 | 1.548 | 1.052 | no | 0.2886 |
| hearing_loss | Mammillary body | hearing_loss | Mammillary_body | neurological | 0.1452 | 0.4019 | 5.77e-10 | 5.46e-10 | 1.24 | 1.057 | no | 0.2886 |

| Trait | Supercluster | dataset_name | Supercluster_ID | disorder_type | P | P.fdr | BETA | SE | Enrichment | Coefficient_z-score | if.sig.fdr | P.adj |
|---|---|---|---|---|---|---|---|---|---|---|---|---|
| hearing_loss | CGE interneuron | hearing_loss | CGE_interneuron | neurological | 0.1519 | 0.4019 | 5.3e-10 | 5.15e-10 | 1.317 | 1.028 | no | 0.2944 |
| hearing_loss | MGE interneuron | hearing_loss | MGE_interneuron | neurological | 0.1556 | 0.4019 | 5.87e-10 | 5.79e-10 | 1.305 | 1.013 | no | 0.2983 |
| hearing_loss | Midbrain-derived inhibitory | hearing_loss | Midbrain_derived_inhibitory | neurological | 0.1765 | 0.4141 | 5.19e-10 | 5.59e-10 | 1.279 | 0.9287 | no | 0.3317 |
| hearing_loss | Splatter | hearing_loss | Splatter | neurological | 0.191 | 0.4141 | 4.89e-10 | 5.6e-10 | 1.282 | 0.8743 | no | 0.3465 |
| hearing_loss | Eccentric medium spiny neuron | hearing_loss | Eccentric_medium_spiny_neuron | neurological | 0.2004 | 0.4141 | 4.18e-10 | 4.97e-10 | 1.249 | 0.8403 | no | 0.3561 |
| hearing_loss | Miscellaneous | hearing_loss | Miscellaneous | neurological | 0.2446 | 0.4739 | 3.56e-10 | 5.15e-10 | 1.199 | 0.6916 | no | 0.4174 |
| hearing_loss | Hippocampal dentate gyrus | hearing_loss | Hippocampal_dentate_gyrus | neurological | 0.342 | 0.6236 | 2.55e-10 | 6.25e-10 | 1.196 | 0.4071 | no | 0.5579 |
| hearing_loss | Upper rhombic lip | hearing_loss | Upper_rhombic_lip | neurological | 0.04916 | 0.254 | 8.84e-10 | 5.35e-10 | 1.367 | 1.653 | no | 0.6368 |
| hearing_loss | Bergmann glia | hearing_loss | Bergmann_glia | neurological | 0.4159 | 0.6305 | 1.42e-10 | 6.67e-10 | 1.383 | 0.2125 | no | 0.6609 |
| hearing_loss | Lower rhombic lip | hearing_loss | Lower_rhombic_lip | neurological | 0.4193 | 0.6305 | 9.68e-11 | 4.75e-10 | 1.217 | 0.2037 | no | 0.6609 |
| hearing_loss | Committed oligodendrocyte precursor | hearing_loss | Committed_oligodendrocyte_precursor | neurological | 0.4291 | 0.6305 | 1.03e-10 | 5.77e-10 | 1.392 | 0.1787 | no | 0.6691 |
| hearing_loss | Ependymal | hearing_loss | Ependymal | neurological | 0.4474 | 0.6305 | 7.33e-11 | 5.55e-10 | 1.31 | 0.1322 | no | 0.6878 |
| hearing_loss | Hippocampal CA4 | hearing_loss | Hippocampal_CA4 | neurological | 0.5265 | 0.7097 | -3.11e-11 | 4.67e-10 | 1.15 | -0.06658 | no | 0.7962 |
| hearing_loss | Thalamic excitatory | hearing_loss | Thalamic_excitatory | neurological | 0.6651 | 0.8592 | -2.18e-10 | 5.1e-10 | 1.101 | -0.4266 | no | 0.9665 |
| hearing_loss | Oligodendrocyte precursor | hearing_loss | Oligodendrocyte_precursor | neurological | 0.8226 | 0.8998 | -5.63e-10 | 6.08e-10 | 1.224 | -0.9252 | no | 0.9819 |
| hearing_loss | Astrocyte | hearing_loss | Astrocyte | neurological | 0.8418 | 0.8998 | -5.53e-10 | 5.52e-10 | 1.25 | -1.002 | no | 1 |
| hearing_loss | Cerebellar inhibitory | hearing_loss | Cerebellar_inhibitory | neurological | 0.9025 | 0.9025 | -6.63e-10 | 5.11e-10 | 1.106 | -1.296 | no | 1 |
| hearing_loss | Choroid plexus | hearing_loss | Choroid_plexus | neurological | 0.8047 | 0.8998 | -5.17e-10 | 6.02e-10 | 1.203 | -0.8585 | no | 1 |
| hearing_loss | Microglia | hearing_loss | Microglia | neurological | 0.8754 | 0.9025 | -6.71e-10 | 5.82e-10 | 1.247 | -1.152 | no | 1 |
| hearing_loss | Oligodendrocyte | hearing_loss | Oligodendrocyte | neurological | 0.7074 | 0.8771 | -2.43e-10 | 4.45e-10 | 1.123 | -0.5457 | no | 1 |
| hearing_loss | Vascular | hearing_loss | Vascular | neurological | 0.7568 | 0.8998 | -5.13e-10 | 7.36e-10 | 1.353 | -0.696 | no | 1 |
| insomnia | Amygdala excitatory | insomnia | Amygdala_excitatory | brain traits | 0.001824 | 0.0423 | 2.15e-09 | 7.4e-10 | 1.33 | 2.907 | yes | 0.009423 |
| insomnia | CGE interneuron | insomnia | CGE_interneuron | brain traits | 0.002729 | 0.0423 | 2.45e-09 | 8.82e-10 | 1.425 | 2.779 | yes | 0.01209 |
| insomnia | Upper-layer intratelencephalic | insomnia | Upper_layer_intratelencephalic | brain traits | 0.2226 | 0.4888 | 6.88e-10 | 9.01e-10 | 1.225 | 0.7636 | yes | 0.02563 |
| insomnia | Deep-layer intratelencephalic | insomnia | Deep_layer_intratelencephalic | brain traits | 0.008405 | 0.06514 | 2.03e-09 | 8.49e-10 | 1.355 | 2.391 | yes | 0.03006 |
| insomnia | MGE interneuron | insomnia | MGE_interneuron | brain traits | 0.01246 | 0.07727 | 1.78e-09 | 7.92e-10 | 1.354 | 2.243 | yes | 0.04139 |
| insomnia | LAMP5-LHX6 and Chandelier | insomnia | LAMP5_LHX6_and_Chandelier | brain traits | 0.02196 | 0.1134 | 1.65e-09 | 8.19e-10 | 1.335 | 2.015 | yes | 0.06188 |
| insomnia | Deep-layer near-projecting | insomnia | Deep_layer_near_projecting | brain traits | 0.02781 | 0.1232 | 1.61e-09 | 8.39e-10 | 1.299 | 1.914 | yes | 0.07608 |
| insomnia | Midbrain-derived inhibitory | insomnia | Midbrain_derived_inhibitory | brain traits | 0.1205 | 0.467 | 9.73e-10 | 8.3e-10 | 1.25 | 1.172 | no | 0.2547 |
| insomnia | Deep-layer corticothalamic and 6b | insomnia | Deep_layer_corticothalamic_and_6b | brain traits | 0.1443 | 0.4888 | 8.6e-10 | 8.11e-10 | 1.189 | 1.061 | no | 0.2886 |
| insomnia | Hippocampal CA1-3 | insomnia | Hippocampal_CA1_3 | brain traits | 0.166 | 0.4888 | 7.26e-10 | 7.48e-10 | 1.161 | 0.9702 | no | 0.315 |
| insomnia | Thalamic excitatory | insomnia | Thalamic_excitatory | brain traits | 0.186 | 0.4888 | 7.74e-10 | 8.67e-10 | 1.192 | 0.8928 | no | 0.3459 |
| insomnia | Hippocampal CA4 | insomnia | Hippocampal_CA4 | brain traits | 0.1899 | 0.4888 | 6.7e-10 | 7.63e-10 | 1.194 | 0.8783 | no | 0.3465 |
| insomnia | Upper rhombic lip | insomnia | Upper_rhombic_lip | brain traits | 0.007029 | 0.06514 | 1.99e-09 | 8.12e-10 | 1.349 | 2.456 | no | 0.3905 |
| insomnia | Lower rhombic lip | insomnia | Lower_rhombic_lip | brain traits | 0.2308 | 0.4888 | 6.33e-10 | 8.6e-10 | 1.183 | 0.7363 | no | 0.4012 |
| insomnia | Cerebellar inhibitory | insomnia | Cerebellar_inhibitory | brain traits | 0.2392 | 0.4888 | 6.2e-10 | 8.74e-10 | 1.208 | 0.7089 | no | 0.412 |
| insomnia | Eccentric medium spiny neuron | insomnia | Eccentric_medium_spiny_neuron | brain traits | 0.2523 | 0.4888 | 5.65e-10 | 8.46e-10 | 1.183 | 0.6673 | no | 0.4266 |
| insomnia | Splatter | insomnia | Splatter | brain traits | 0.3292 | 0.6003 | 3.64e-10 | 8.24e-10 | 1.154 | 0.4422 | no | 0.5418 |
| insomnia | Medium spiny neuron | insomnia | Medium_spiny_neuron | brain traits | 0.3841 | 0.6615 | 2.25e-10 | 7.65e-10 | 1.108 | 0.2948 | no | 0.6212 |
| insomnia | Hippocampal dentate gyrus | insomnia | Hippocampal_dentate_gyrus | brain traits | 0.4317 | 0.7043 | 1.21e-10 | 7.02e-10 | 1.1 | 0.1722 | no | 0.6691 |
| insomnia | Miscellaneous | insomnia | Miscellaneous | brain traits | 0.5348 | 0.8289 | -6.56e-11 | 7.52e-10 | 1.067 | -0.08731 | no | 0.8022 |
| insomnia | Bergmann glia | insomnia | Bergmann_glia | brain traits | 0.6308 | 0.9121 | -2.81e-10 | 8.41e-10 | 1.15 | -0.3339 | no | 0.9311 |
| insomnia | Ependymal | insomnia | Ependymal | brain traits | 0.6473 | 0.9121 | -3.09e-10 | 8.18e-10 | 1.076 | -0.3781 | no | 0.9481 |
| insomnia | Committed oligodendrocyte precursor | insomnia | Committed_oligodendrocyte_precursor | brain traits | 0.6833 | 0.9209 | -3.96e-10 | 8.31e-10 | 1.097 | -0.4769 | no | 0.9705 |
| insomnia | Astrocyte | insomnia | Astrocyte | brain traits | 0.9576 | 0.9876 | -1.6e-09 | 9.27e-10 | 0.9786 | -1.723 | no | 1 |

| Trait | Supercluster | dataset_name | Supercluster_ID | disorder_type | P | P.fdr | BETA | SE | Enrichment | Coefficient_z-score | if.sig.fdr | P.adj |
|---|---|---|---|---|---|---|---|---|---|---|---|---|
| insomnia | Choroid plexus | insomnia | Choroid_plexus | brain traits | 0.9313 | 0.9876 | -1.41e-09 | 9.46e-10 | 0.9368 | -1.485 | no | 1 |
| insomnia | Fibroblast | insomnia | Fibroblast | brain traits | 0.9876 | 0.9876 | -1.83e-09 | 8.13e-10 | 0.9076 | -2.246 | no | 1 |
| insomnia | Mammillary body | insomnia | Mammillary_body | brain traits | 0.8375 | 0.9876 | -7.38e-10 | 7.5e-10 | 1.01 | -0.9843 | no | 1 |
| insomnia | Microglia | insomnia | Microglia | brain traits | 0.8272 | 0.9876 | -8.75e-10 | 9.27e-10 | 0.9652 | -0.9432 | no | 1 |
| insomnia | Oligodendrocyte | insomnia | Oligodendrocyte | brain traits | 0.8077 | 0.9876 | -6.8e-10 | 7.82e-10 | 1.022 | -0.8693 | no | 1 |
| insomnia | Oligodendrocyte precursor | insomnia | Oligodendrocyte_precursor | brain traits | 0.9769 | 0.9876 | -1.67e-09 | 8.38e-10 | 0.9403 | -1.994 | no | 1 |
| insomnia | Vascular | insomnia | Vascular | brain traits | 0.9811 | 0.9876 | -2e-09 | 9.62e-10 | 0.8749 | -2.078 | no | 1 |
| mdd2019 | MGE interneuron | mdd2019 | MGE_interneuron | psychiatric | 3.65e-06 | 0.0001131 | 4.71e-09 | 1.05e-09 | 1.518 | 4.485 | yes | 0.0001357 |
| mdd2019 | CGE interneuron | mdd2019 | CGE_interneuron | psychiatric | 1.42e-05 | 0.0002198 | 4.63e-09 | 1.11e-09 | 1.474 | 4.186 | yes | 0.0003768 |
| mdd2019 | Amygdala excitatory | mdd2019 | Amygdala_excitatory | psychiatric | 6.17e-05 | 0.0006379 | 3.34e-09 | 8.69e-10 | 1.357 | 3.839 | yes | 0.0009568 |
| mdd2019 | LAMP5-LHX6 and Chandelier | mdd2019 | LAMP5_LHX6_and_Chandelier | psychiatric | 0.0001765 | 0.001368 | 3.47e-09 | 9.7e-10 | 1.405 | 3.573 | yes | 0.001728 |
| mdd2019 | Thalamic excitatory | mdd2019 | Thalamic_excitatory | psychiatric | 0.0002463 | 0.001527 | 3.24e-09 | 9.31e-10 | 1.355 | 3.485 | yes | 0.002085 |
| mdd2019 | Hippocampal CA1-3 | mdd2019 | Hippocampal_CA1_3 | psychiatric | 0.001759 | 0.007789 | 2.95e-09 | 1.01e-09 | 1.311 | 2.918 | yes | 0.009347 |
| mdd2019 | Miscellaneous | mdd2019 | Miscellaneous | psychiatric | 0.001744 | 0.007789 | 2.65e-09 | 9.08e-10 | 1.268 | 2.921 | yes | 0.009347 |
| mdd2019 | Eccentric medium spiny neuron | mdd2019 | Eccentric_medium_spiny_neuron | psychiatric | 0.002399 | 0.009297 | 2.8e-09 | 9.94e-10 | 1.333 | 2.82 | yes | 0.01123 |
| mdd2019 | Cerebellar inhibitory | mdd2019 | Cerebellar_inhibitory | psychiatric | 0.00449 | 0.01546 | 2.67e-09 | 1.02e-09 | 1.28 | 2.613 | yes | 0.01856 |
| mdd2019 | Deep-layer intratelencephalic | mdd2019 | Deep_layer_intratelencephalic | psychiatric | 0.01009 | 0.03128 | 2.32e-09 | 9.99e-10 | 1.276 | 2.323 | yes | 0.03476 |
| mdd2019 | Upper-layer intratelencephalic | mdd2019 | Upper_layer_intratelencephalic | psychiatric | 0.6835 | 0.9445 | -4.29e-10 | 8.99e-10 | 0.9549 | -0.4775 | yes | 0.05054 |
| mdd2019 | Hippocampal CA4 | mdd2019 | Hippocampal_CA4 | psychiatric | 0.01928 | 0.0498 | 2.12e-09 | 1.02e-09 | 1.266 | 2.069 | yes | 0.05603 |
| mdd2019 | Deep-layer corticothalamic and 6b | mdd2019 | Deep_layer_corticothalamic_and_6b | psychiatric | 0.03626 | 0.08646 | 1.74e-09 | 9.71e-10 | 1.206 | 1.796 | yes | 0.09634 |
| mdd2019 | Oligodendrocyte precursor | mdd2019 | Oligodendrocyte_precursor | psychiatric | 0.9587 | 1 | -1.59e-09 | 9.14e-10 | 0.8466 | -1.736 | no | 0.111 |
| mdd2019 | Deep-layer near-projecting | mdd2019 | Deep_layer_near_projecting | psychiatric | 0.07181 | 0.132 | 1.4e-09 | 9.54e-10 | 1.179 | 1.462 | no | 0.1642 |
| mdd2019 | Hippocampal dentate gyrus | mdd2019 | Hippocampal_dentate_gyrus | psychiatric | 0.07188 | 0.132 | 1.26e-09 | 8.6e-10 | 1.161 | 1.462 | no | 0.1642 |
| mdd2019 | Midbrain-derived inhibitory | mdd2019 | Midbrain_derived_inhibitory | psychiatric | 0.0724 | 0.132 | 1.48e-09 | 1.01e-09 | 1.231 | 1.458 | no | 0.1642 |
| mdd2019 | Medium spiny neuron | mdd2019 | Medium_spiny_neuron | psychiatric | 0.08547 | 0.1472 | 1.25e-09 | 9.12e-10 | 1.134 | 1.369 | no | 0.1893 |
| mdd2019 | Splatter | mdd2019 | Splatter | psychiatric | 0.1068 | 0.1743 | 1.22e-09 | 9.78e-10 | 1.137 | 1.244 | no | 0.2337 |
| mdd2019 | Mammillary body | mdd2019 | Mammillary_body | psychiatric | 0.1467 | 0.2274 | 9.2e-10 | 8.76e-10 | 1.161 | 1.051 | no | 0.2886 |
| mdd2019 | Upper rhombic lip | mdd2019 | Upper_rhombic_lip | psychiatric | 0.01657 | 0.04671 | 2.15e-09 | 1.01e-09 | 1.235 | 2.13 | no | 0.9705 |
| mdd2019 | Bergmann glia | mdd2019 | Bergmann_glia | psychiatric | 0.6958 | 0.9445 | -4.39e-10 | 8.56e-10 | 1.017 | -0.5123 | no | 0.98 |
| mdd2019 | Lower rhombic lip | mdd2019 | Lower_rhombic_lip | psychiatric | 0.7007 | 0.9445 | -5.17e-10 | 9.81e-10 | 0.972 | -0.5266 | no | 0.98 |
| mdd2019 | Astrocyte | mdd2019 | Astrocyte | psychiatric | 0.8522 | 1 | -1.07e-09 | 1.02e-09 | 0.9263 | -1.046 | no | 1 |
| mdd2019 | Choroid plexus | mdd2019 | Choroid_plexus | psychiatric | 1 | 1 | -3.26e-09 | 7.45e-10 | 0.683 | -4.379 | no | 1 |
| mdd2019 | Committed oligodendrocyte precursor | mdd2019 | Committed_oligodendrocyte_precursor | psychiatric | 0.9961 | 1 | -2.22e-09 | 8.33e-10 | 0.7637 | -2.662 | no | 1 |
| mdd2019 | Ependymal | mdd2019 | Ependymal | psychiatric | 0.9997 | 1 | -2.96e-09 | 8.57e-10 | 0.7014 | -3.451 | no | 1 |
| mdd2019 | Fibroblast | mdd2019 | Fibroblast | psychiatric | 0.9993 | 1 | -2.57e-09 | 8.01e-10 | 0.7191 | -3.211 | no | 1 |
| mdd2019 | Microglia | mdd2019 | Microglia | psychiatric | 0.9942 | 1 | -2.01e-09 | 7.98e-10 | 0.6798 | -2.522 | no | 1 |
| mdd2019 | Oligodendrocyte | mdd2019 | Oligodendrocyte | psychiatric | 0.04239 | 0.09386 | 1.46e-09 | 8.46e-10 | 1.202 | 1.724 | no | 1 |
| mdd2019 | Vascular | mdd2019 | Vascular | psychiatric | 0.9957 | 1 | -2.38e-09 | 9.08e-10 | 0.6737 | -2.624 | no | 1 |
| neuroticism | Deep-layer intratelencephalic | neuroticism | Deep_layer_intratelencephalic | brain traits | 1.19e-06 | 3.69e-05 | 6.97e-09 | 1.48e-09 | 1.448 | 4.718 | yes | 0.0001084 |
| neuroticism | Miscellaneous | neuroticism | Miscellaneous | brain traits | 2.37e-05 | 0.0003675 | 5.01e-09 | 1.23e-09 | 1.309 | 4.068 | yes | 0.0005513 |
| neuroticism | Medium spiny neuron | neuroticism | Medium_spiny_neuron | brain traits | 3.76e-05 | 0.000389 | 5.29e-09 | 1.34e-09 | 1.327 | 3.959 | yes | 0.000727 |
| neuroticism | Upper-layer intratelencephalic | neuroticism | Upper_layer_intratelencephalic | brain traits | 0.4832 | 0.6809 | 5.91e-11 | 1.4e-09 | 1.068 | 0.04213 | yes | 0.001221 |
| neuroticism | Deep-layer corticothalamic and 6b | neuroticism | Deep_layer_corticothalamic_and_6b | brain traits | 0.0001437 | 0.0008911 | 5.26e-09 | 1.45e-09 | 1.338 | 3.626 | yes | 0.001648 |
| neuroticism | Eccentric medium spiny neuron | neuroticism | Eccentric_medium_spiny_neuron | brain traits | 0.0002296 | 0.001092 | 4.54e-09 | 1.3e-09 | 1.336 | 3.503 | yes | 0.002085 |
| neuroticism | LAMP5-LHX6 and Chandelier | neuroticism | LAMP5_LHX6_and_Chandelier | brain traits | 0.0002466 | 0.001092 | 4.71e-09 | 1.35e-09 | 1.357 | 3.484 | yes | 0.002085 |

| Trait | Supercluster | dataset_name | Supercluster_ID | disorder_type | P | P.fdr | BETA | SE | Enrichment | Coefficient_z-score | if.sig.fdr | P.adj |
|---|---|---|---|---|---|---|---|---|---|---|---|---|
| neuroticism | Amygdala excitatory | neuroticism | Amygdala_excitatory | brain traits | 0.0003374 | 0.001307 | 4.55e-09 | 1.34e-09 | 1.301 | 3.4 | yes | 0.002614 |
| neuroticism | Hippocampal CA1-3 | neuroticism | Hippocampal_CA1_3 | brain traits | 0.001079 | 0.003718 | 4.04e-09 | 1.32e-09 | 1.269 | 3.067 | yes | 0.006693 |
| neuroticism | Hippocampal dentate gyrus | neuroticism | Hippocampal_dentate_gyrus | brain traits | 0.001685 | 0.004897 | 3.57e-09 | 1.22e-09 | 1.256 | 2.932 | yes | 0.009347 |
| neuroticism | MGE interneuron | neuroticism | MGE_interneuron | brain traits | 0.001738 | 0.004897 | 4.49e-09 | 1.54e-09 | 1.347 | 2.922 | yes | 0.009347 |
| neuroticism | CGE interneuron | neuroticism | CGE_interneuron | brain traits | 0.002082 | 0.005374 | 4.2e-09 | 1.47e-09 | 1.302 | 2.865 | yes | 0.01047 |
| neuroticism | Mammillary body | neuroticism | Mammillary_body | brain traits | 0.002254 | 0.005374 | 3.48e-09 | 1.22e-09 | 1.269 | 2.84 | yes | 0.01103 |
| neuroticism | Thalamic excitatory | neuroticism | Thalamic_excitatory | brain traits | 0.002618 | 0.005796 | 3.93e-09 | 1.41e-09 | 1.293 | 2.792 | yes | 0.01187 |
| neuroticism | Deep-layer near-projecting | neuroticism | Deep_layer_near_projecting | brain traits | 0.005566 | 0.0115 | 3.73e-09 | 1.47e-09 | 1.274 | 2.538 | yes | 0.02203 |
| neuroticism | Hippocampal CA4 | neuroticism | Hippocampal_CA4 | brain traits | 0.01007 | 0.01951 | 3.12e-09 | 1.34e-09 | 1.256 | 2.324 | yes | 0.03476 |
| neuroticism | Cerebellar inhibitory | neuroticism | Cerebellar_inhibitory | brain traits | 0.07181 | 0.1245 | 2.02e-09 | 1.38e-09 | 1.189 | 1.462 | no | 0.1642 |
| neuroticism | Midbrain-derived inhibitory | neuroticism | Midbrain_derived_inhibitory | brain traits | 0.07228 | 0.1245 | 2.15e-09 | 1.48e-09 | 1.228 | 1.459 | no | 0.1642 |
| neuroticism | Oligodendrocyte precursor | neuroticism | Oligodendrocyte_precursor | brain traits | 0.9931 | 1 | -3.19e-09 | 1.3e-09 | 0.9059 | -2.461 | no | 0.2447 |
| neuroticism | Lower rhombic lip | neuroticism | Lower_rhombic_lip | brain traits | 0.1337 | 0.2073 | 1.53e-09 | 1.38e-09 | 1.145 | 1.109 | no | 0.2764 |
| neuroticism | Splatter | neuroticism | Splatter | brain traits | 0.201 | 0.2967 | 1.26e-09 | 1.5e-09 | 1.133 | 0.838 | no | 0.3561 |
| neuroticism | Upper rhombic lip | neuroticism | Upper_rhombic_lip | brain traits | 9.19e-05 | 0.0007122 | 4.87e-09 | 1.3e-09 | 1.328 | 3.74 | no | 0.7367 |
| neuroticism | Bergmann glia | neuroticism | Bergmann_glia | brain traits | 0.7135 | 0.9617 | -8.22e-10 | 1.46e-09 | 1.086 | -0.5636 | no | 0.983 |
| neuroticism | Astrocyte | neuroticism | Astrocyte | brain traits | 0.9454 | 1 | -2.12e-09 | 1.32e-09 | 0.9979 | -1.602 | no | 1 |
| neuroticism | Choroid plexus | neuroticism | Choroid_plexus | brain traits | 0.7596 | 0.9811 | -9.74e-10 | 1.38e-09 | 0.9934 | -0.705 | no | 1 |
| neuroticism | Committed oligodendrocyte precursor | neuroticism | Committed_oligodendrocyte_precursor | brain traits | 0.9742 | 1 | -2.34e-09 | 1.2e-09 | 0.9293 | -1.946 | no | 1 |
| neuroticism | Ependymal | neuroticism | Ependymal | brain traits | 0.9573 | 1 | -2.26e-09 | 1.31e-09 | 0.9264 | -1.72 | no | 1 |
| neuroticism | Fibroblast | neuroticism | Fibroblast | brain traits | 1 | 1 | -5.02e-09 | 1.28e-09 | 0.7825 | -3.92 | no | 1 |
| neuroticism | Microglia | neuroticism | Microglia | brain traits | 0.9914 | 1 | -2.97e-09 | 1.25e-09 | 0.7978 | -2.381 | no | 1 |
| neuroticism | Oligodendrocyte | neuroticism | Oligodendrocyte | brain traits | 0.1131 | 0.1846 | 1.68e-09 | 1.39e-09 | 1.179 | 1.21 | no | 1 |
| neuroticism | Vascular | neuroticism | Vascular | brain traits | 0.9994 | 1 | -4.45e-09 | 1.36e-09 | 0.7639 | -3.263 | no | 1 |
| scz2022 | CGE interneuron | scz2022_eur | CGE_interneuron | psychiatric | 2.28e-06 | 3.53e-05 | 2.14e-08 | 4.68e-09 | 1.485 | 4.584 | yes | 0.0001084 |
| scz2022 | Hippocampal CA1-3 | scz2022_eur | Hippocampal_CA1_3 | psychiatric | 8.12e-07 | 2.52e-05 | 2.1e-08 | 4.37e-09 | 1.435 | 4.795 | yes | 0.0001084 |
| scz2022 | Deep-layer intratelencephalic | scz2022_eur | Deep_layer_intratelencephalic | psychiatric | 1.27e-05 | 0.0001313 | 2.3e-08 | 5.46e-09 | 1.502 | 4.211 | yes | 0.0003768 |
| scz2022 | Amygdala excitatory | scz2022_eur | Amygdala_excitatory | psychiatric | 3.91e-05 | 0.0002877 | 1.87e-08 | 4.74e-09 | 1.395 | 3.95 | yes | 0.000727 |
| scz2022 | Eccentric medium spiny neuron | scz2022_eur | Eccentric_medium_spiny_neuron | psychiatric | 4.64e-05 | 0.0002877 | 1.8e-08 | 4.61e-09 | 1.428 | 3.909 | yes | 0.0007846 |
| scz2022 | Hippocampal CA4 | scz2022_eur | Hippocampal_CA4 | psychiatric | 6.75e-05 | 0.000349 | 1.6e-08 | 4.19e-09 | 1.393 | 3.817 | yes | 0.0009663 |
| scz2022 | Upper-layer intratelencephalic | scz2022_eur | Upper_layer_intratelencephalic | psychiatric | 0.2684 | 0.416 | 3.13e-09 | 5.06e-09 | 1.228 | 0.6176 | yes | 0.003865 |
| scz2022 | Deep-layer corticothalamic and 6b | scz2022_eur | Deep_layer_corticothalamic_and_6b | psychiatric | 0.000612 | 0.002372 | 1.48e-08 | 4.57e-09 | 1.349 | 3.233 | yes | 0.004202 |
| scz2022 | LAMP5-LHX6 and Chandelier | scz2022_eur | LAMP5_LHX6_and_Chandelier | psychiatric | 0.002414 | 0.008316 | 1.36e-08 | 4.83e-09 | 1.377 | 2.818 | yes | 0.01123 |
| scz2022 | Miscellaneous | scz2022_eur | Miscellaneous | psychiatric | 0.003092 | 0.009585 | 1.28e-08 | 4.69e-09 | 1.31 | 2.738 | yes | 0.01337 |
| scz2022 | Hippocampal dentate gyrus | scz2022_eur | Hippocampal_dentate_gyrus | psychiatric | 0.005303 | 0.01495 | 1.25e-08 | 4.9e-09 | 1.323 | 2.555 | yes | 0.02144 |
| scz2022 | Medium spiny neuron | scz2022_eur | Medium_spiny_neuron | psychiatric | 0.005986 | 0.01546 | 1.21e-08 | 4.83e-09 | 1.311 | 2.513 | yes | 0.0232 |
| scz2022 | MGE interneuron | scz2022_eur | MGE_interneuron | psychiatric | 0.006725 | 0.01604 | 1.21e-08 | 4.88e-09 | 1.356 | 2.472 | yes | 0.02553 |
| scz2022 | Thalamic excitatory | scz2022_eur | Thalamic_excitatory | psychiatric | 0.01065 | 0.02357 | 9.54e-09 | 4.14e-09 | 1.289 | 2.303 | yes | 0.036 |
| scz2022 | Cerebellar inhibitory | scz2022_eur | Cerebellar_inhibitory | psychiatric | 0.01423 | 0.0294 | 1.08e-08 | 4.95e-09 | 1.335 | 2.191 | yes | 0.04485 |
| scz2022 | Deep-layer near-projecting | scz2022_eur | Deep_layer_near_projecting | psychiatric | 0.04595 | 0.08903 | 7.64e-09 | 4.53e-09 | 1.261 | 1.685 | no | 0.1187 |
| scz2022 | Oligodendrocyte precursor | scz2022_eur | Oligodendrocyte_precursor | psychiatric | 0.5825 | 0.8208 | -1.14e-09 | 5.45e-09 | 1.182 | -0.2084 | no | 0.1367 |
| scz2022 | Midbrain-derived inhibitory | scz2022_eur | Midbrain_derived_inhibitory | psychiatric | 0.05669 | 0.09764 | 6.9e-09 | 4.36e-09 | 1.268 | 1.583 | no | 0.1388 |
| scz2022 | Mammillary body | scz2022_eur | Mammillary_body | psychiatric | 0.1474 | 0.2405 | 4.71e-09 | 4.5e-09 | 1.215 | 1.048 | no | 0.2886 |
| scz2022 | Upper rhombic lip | scz2022_eur | Upper_rhombic_lip | psychiatric | 0.0005403 | 0.002372 | 1.78e-08 | 5.45e-09 | 1.424 | 3.269 | no | 0.4498 |
| scz2022 | Lower rhombic lip | scz2022_eur | Lower_rhombic_lip | psychiatric | 0.3165 | 0.4672 | 2.07e-09 | 4.34e-09 | 1.178 | 0.4775 | no | 0.5256 |

| Trait | Supercluster | dataset_name | Supercluster_ID | disorder_type | P | P.fdr | BETA | SE | Enrichment | Coefficient_z-score | if.sig.fdr | P.adj |
|---|---|---|---|---|---|---|---|---|---|---|---|---|
| scz2022 | Oligodendrocyte | scz2022_eur | Oligodendrocyte | psychiatric | 0.05514 | 0.09764 | 6.96e-09 | 4.36e-09 | 1.253 | 1.597 | no | 0.8668 |
| scz2022 | Astrocyte | scz2022_eur | Astrocyte | psychiatric | 0.9525 | 1 | -8.93e-09 | 5.35e-09 | 1.093 | -1.67 | no | 1 |
| scz2022 | Bergmann glia | scz2022_eur | Bergmann_glia | psychiatric | 0.9098 | 1 | -7.47e-09 | 5.58e-09 | 1.125 | -1.339 | no | 1 |
| scz2022 | Choroid plexus | scz2022_eur | Choroid_plexus | psychiatric | 0.9857 | 1 | -1.06e-08 | 4.83e-09 | 1.014 | -2.19 | no | 1 |
| scz2022 | Committed oligodendrocyte precursor | scz2022_eur | Committed_oligodendrocyte_precursor | psychiatric | 0.8251 | 1 | -4.12e-09 | 4.41e-09 | 1.129 | -0.9351 | no | 1 |
| scz2022 | Ependymal | scz2022_eur | Ependymal | psychiatric | 0.9978 | 1 | -1.39e-08 | 4.86e-09 | 0.972 | -2.851 | no | 1 |
| scz2022 | Fibroblast | scz2022_eur | Fibroblast | psychiatric | 1 | 1 | -2.51e-08 | 5.1e-09 | 0.8131 | -4.924 | no | 1 |
| scz2022 | Microglia | scz2022_eur | Microglia | psychiatric | 0.9999 | 1 | -1.64e-08 | 4.28e-09 | 0.8919 | -3.836 | no | 1 |
| scz2022 | Splatter | scz2022_eur | Splatter | psychiatric | 0.8331 | 1 | -4.21e-09 | 4.36e-09 | 1.075 | -0.9667 | no | 1 |
| scz2022 | Vascular | scz2022_eur | Vascular | psychiatric | 0.9992 | 1 | -1.55e-08 | 4.91e-09 | 0.9299 | -3.146 | no | 1 |

Fine line separates suggestive significant enrichments from non-enriched superclusters. Thick line separates traits. Abbreviation: P = p value; P.fdr = FDR adjusted P value Yao et al.,; BETA = beta coefficient; SE = standard error; P.adj = adjusted p value for included studies in the current analysis

#### Supplementary Table S13

LD score regression genetic correlation analysis ordered from most significant to less significant

| Trait1 | Trait2 | PMID | Rg | Rg_se | Pval | Pval_FDR | is_sig |
|---|---|---|---|---|---|---|---|
| Tinnitus | Hearing loss | 35580588 | 0.5008 | 0.0296 | 2.64e-64 | 3.43e-63 | yes |
| Tinnitus | Major Depressive Disorder | 30718901 | 0.3249 | 0.0281 | 5.7e-31 | 3.7e-30 | yes |
| Tinnitus | Neuroticism | 29942085 | 0.2624 | 0.0239 | 4.93e-28 | 2.14e-27 | yes |
| Tinnitus | Anxiety Disorder | 31748690 | 0.3671 | 0.044 | 6.87e-17 | 2.23e-16 | yes |
| Tinnitus | Insomnia | 30804565 | 0.1908 | 0.0339 | 1.8e-08 | 4.68e-08 | yes |
| Tinnitus | Vertigo | 34620984 | 0.2196 | 0.0513 | 1.85e-05 | 4.01e-05 | yes |
| Tinnitus | Chronic Pain | 31194737 | 0.2962 | 0.0931 | 0.0015 | 0.002786 | yes |
| Tinnitus | Post-traumatic stress syndrome | 31594949 | 0.2359 | 0.0778 | 0.0024 | 0.0039 | yes |
| Tinnitus | Schizophrenia | 35396580 | 0.0676 | 0.0294 | 0.0215 | 0.03106 | yes |
| Tinnitus | Bipolar disorder | 34002096 | 0.0798 | 0.0367 | 0.0297 | 0.03861 | yes |
| Tinnitus | Alzheimer's disease | 35379992 | 0.0485 | 0.0527 | 0.3577 | 0.4227 | no |
| Tinnitus | Type 2 Diabetes | 35551307 | 0.0242 | 0.0329 | 0.4616 | 0.5001 | no |
| Tinnitus | Cerebellar volume | 35842455 | 0.0272 | 0.051 | 0.5942 | 0.5942 | no |

Line separates significant correlations and non-significant correlations. Abbreviations: PMID = pubmed ID; Rg = correlation coefficient; se = standard error; Pval = p-value; FDR = false discovery rate

#### Supplementary Table S14

Brain cell-type enrichment of tinnitus heritability in the meta-analysis and in the Million Veteran Program

**Stratified LD score regression coefficient z-scores and Benjamini-Hochberg FDR across 31 superclusters of the human brain atlas. One-sided test on the regression coefficient; FDR computed within each phenotype.**

*part 1 of 2 — first column repeated in every part*

| Supercluster | Meta-analysis (this study) coefficient z | Meta-analysis (this study) FDR | MVP self-report (HVTin) coefficient z | MVP self-report (HVTin) FDR |
|---|---|---|---|---|
| Upper layer intratelencephalic | 1.4 | 0.2501 | 3.1 | 0.0298 |
| MGE interneuron | 2.49 | 0.0659 | 2.56 | 0.0526 |
| Amygdala excitatory | 2.86 | 0.0658 | 2.54 | 0.0526 |
| Deep layer intratelencephalic | 0.77 | 0.4015 | 1.58 | 0.1982 |
| LAMP5 LHX6 and Chandelier | 1.83 | 0.1338 | 2.32 | 0.0526 |
| Deep layer corticothalamic and 6b | 0.97 | 0.367 | 1.26 | 0.2876 |
| Medium spiny neuron | 1.81 | 0.1338 | 2.33 | 0.0526 |
| Hippocampal CA1 3 | 0.92 | 0.367 | 0.48 | 0.5406 |
| Miscellaneous | 2.14 | 0.0839 | 0.8 | 0.4178 |
| Deep layer near projecting | 0.73 | 0.4015 | 1.18 | 0.2876 |
| Eccentric medium spiny neuron | 2.21 | 0.0839 | 2.44 | 0.0526 |
| Hippocampal dentate gyrus | 2.22 | 0.0839 | 1.27 | 0.2876 |
| Midbrain derived inhibitory | 2.59 | 0.0659 | 1.64 | 0.1949 |
| CGE interneuron | -0.38 | 0.8712 | 1.89 | 0.1293 |
| Cerebellar inhibitory | 1.03 | 0.367 | 1.17 | 0.2876 |
| Upper rhombic lip | 0.78 | 0.4015 | -0.21 | 0.8199 |
| Hippocampal CA4 | 1.15 | 0.3513 | -0.57 | 0.8295 |
| Mammillary body | 0.93 | 0.367 | -1.27 | 0.9495 |
| Thalamic excitatory | -0.14 | 0.7829 | -0.49 | 0.8295 |
| Oligodendrocyte precursor | 0.47 | 0.5229 | 0.4 | 0.5609 |
| Astrocyte | 1.76 | 0.1338 | -0.35 | 0.8295 |
| Vascular | -2.48 | 0.9996 | -0.57 | 0.8295 |
| Committed oligodendrocyte precursor | -1.94 | 0.9996 | 0.74 | 0.4178 |
| Bergmann glia | 0.05 | 0.7111 | 0.75 | 0.4178 |
| Lower rhombic lip | 0.12 | 0.7009 | -0.59 | 0.8295 |
| Choroid plexus | -1.43 | 0.9996 | 0.09 | 0.7187 |
| Splatter | -1.17 | 0.9996 | 0.95 | 0.3794 |
| Oligodendrocyte | -3.35 | 0.9996 | -0.17 | 0.8199 |
| Microglia | -1.5 | 0.9996 | -1.25 | 0.9495 |
| Fibroblast | -0.86 | 0.9996 | -1.4 | 0.9495 |
| Ependymal | -1.41 | 0.9996 | -2.28 | 0.9888 |

*part 2 of 2 — first column repeated in every part*

| Supercluster | MVP diagnostic code (phecode 389.4) coefficient z | MVP diagnostic code (phecode 389.4) FDR | MVP medication (HVTinMed) coefficient z | MVP medication (HVTinMed) FDR |
|---|---|---|---|---|
| Upper layer intratelencephalic | 3.36 | 0.0059 | 2.23 | 0.2019 |
| MGE interneuron | 3.41 | 0.0059 | 1 | 0.728 |
| Amygdala excitatory | 2.52 | 0.0303 | 0.13 | 0.9655 |
| Deep layer intratelencephalic | 2.53 | 0.0303 | 2.93 | 0.0524 |
| LAMP5 LHX6 and Chandelier | 2.4 | 0.0362 | 0.95 | 0.728 |
| Deep layer corticothalamic and 6b | 3.09 | 0.0103 | 1.08 | 0.728 |
| Medium spiny neuron | 1.35 | 0.1942 | 0.25 | 0.9655 |
| Hippocampal CA1 3 | 2.99 | 0.0108 | 0.76 | 0.7584 |
| Miscellaneous | 1.5 | 0.1592 | 0.69 | 0.7584 |
| Deep layer near projecting | 2.19 | 0.055 | 0.97 | 0.728 |
| Eccentric medium spiny neuron | 0.91 | 0.3495 | -0.56 | 0.9655 |
| Hippocampal dentate gyrus | 1.92 | 0.0858 | -0.67 | 0.9655 |
| Midbrain derived inhibitory | 1.52 | 0.1592 | -1.1 | 0.9722 |
| CGE interneuron | 1.75 | 0.1133 | -0.64 | 0.9655 |
| Cerebellar inhibitory | 0.06 | 0.7 | -0.08 | 0.9655 |
| Upper rhombic lip | 1.93 | 0.0858 | -0.34 | 0.9655 |
| Hippocampal CA4 | 0.95 | 0.3495 | -0.52 | 0.9655 |
| Mammillary body | 0.66 | 0.438 | 0.59 | 0.782 |
| Thalamic excitatory | -0.16 | 0.7667 | 1.15 | 0.728 |
| Oligodendrocyte precursor | -0.17 | 0.7667 | -1.19 | 0.9722 |
| Astrocyte | -1.18 | 0.9109 | -1.21 | 0.9722 |
| Vascular | 0.87 | 0.3495 | 0.89 | 0.728 |
| Committed oligodendrocyte precursor | 0.11 | 0.7 | -0.5 | 0.9655 |
| Bergmann glia | -0.39 | 0.7956 | -1.99 | 0.9768 |
| Lower rhombic lip | -0.77 | 0.8829 | -0.51 | 0.9655 |
| Choroid plexus | -0.31 | 0.7956 | -0.22 | 0.9655 |
| Splatter | -0.43 | 0.7956 | -1.8 | 0.9768 |
| Oligodendrocyte | 0.18 | 0.6961 | -0.13 | 0.9655 |
| Microglia | -0.98 | 0.8938 | -0.63 | 0.9655 |
| Fibroblast | -0.83 | 0.8829 | -1.34 | 0.9722 |
| Ependymal | -1.48 | 0.9302 | -1.31 | 0.9722 |

#### SNP-based heritability (observed scale), LDSC intercept and mean chi-square for each dataset

| Dataset | h2 | SE | h2 z | Intercept | Mean chi2 | Cases | Controls |
|---|---|---|---|---|---|---|---|
| Meta-analysis (this study) | 0.0123 | 0.001 | 12.8 | 1.013 | 1.257 | 75584 | 885501 |
| MVP self-report (HVTin) | 0.044 | 0.0023 | 18.9 | 1.131 | 1.412 | 114325 | 201343 |
| MVP diagnostic code (phecode 389.4) | 0.0286 | 0.0017 | 16.9 | 1.118 | 1.345 | 77601 | 310953 |
| MVP medication (HVTinMed) | 0.0032 | 0.0014 | 2.3 | 1.025 | 1.044 | | |

#### Supplementary Table S15

Cluster-level enrichment of tinnitus heritability across the 461 Siletti clusters, with cluster annotation

*part 1 of 3 — first column repeated in every part*

| Cluster (Yao naming) | Siletti cluster name | Siletti cluster ID | Supercluster | Supercluster abbreviation | Class auto-annotation | Neurotransmitter auto-annotation | Subtype auto-annotation |
|---|---|---|---|---|---|---|---|
| Amyg-Exc 161 | Amex_161 | 161 | Amygdala excitatory | Amyg-Exc | NEUR | VGLUT1 VGLUT2 | |
| Misc 164 | Misc_164 | 164 | Miscellaneous | Misc | NEUR | VGLUT1 VGLUT2 | |
| Amyg-Exc 156 | Amex_156 | 156 | Amygdala excitatory | Amyg-Exc | NEUR | VGLUT1 VGLUT2 | |
| HiDG 203 | DG_203 | 203 | Hippocampal dentate gyrus | HiDG | NEUR | VGLUT1 | DG-GRAN |
| MGE-IN 259 | MGE_259 | 259 | MGE interneuron | MGE-IN | NEUR | GABA | INT-PVALB |
| Amyg-Exc 155 | Amex_155 | 155 | Amygdala excitatory | Amyg-Exc | NEUR | VGLUT1 VGLUT2 | |
| Amyg-Exc 153 | Amex_153 | 153 | Amygdala excitatory | Amyg-Exc | NEUR | VGLUT1 VGLUT2 | |
| Amyg-Exc 159 | Amex_159 | 159 | Amygdala excitatory | Amyg-Exc | NEUR | VGLUT1 VGLUT2 | |
| MGE-IN 236 | MGE_236 | 236 | MGE interneuron | MGE-IN | NEUR | GABA | INT-SST |
| Misc 168 | Misc_168 | 168 | Miscellaneous | Misc | NEUR | VGLUT1 VGLUT2 | |
| MB-In 439 | Midi_439 | 439 | Midbrain-derived inhibitory | MB-In | NEUR | GABA | |
| MGE-IN 260 | MGE_260 | 260 | MGE interneuron | MGE-IN | NEUR | GABA | INT-PVALB |
| MB-In 440 | Midi_440 | 440 | Midbrain-derived inhibitory | MB-In | NEUR | GABA | |
| MB-In 438 | Midi_438 | 438 | Midbrain-derived inhibitory | MB-In | NEUR | GABA | |
| DL-IT 144 | DLIT_144 | 144 | Deep-layer intratelencephalic | DL-IT | NEUR | VGLUT1 | |
| LLC-IN 265 | LLC_265 | 265 | LAMP5-LHX6 and Chandelier | LLC-IN | NEUR | GABA | CHAND |
| Amyg-Exc 173 | Amex_173 | 173 | Amygdala excitatory | Amyg-Exc | NEUR | VGLUT1 VGLUT2 | |
| UL-IT 120 | ULIT_120 | 120 | Upper-layer intratelencephalic | UL-IT | NEUR | VGLUT1 VGLUT2 | |
| UL-IT 124 | ULIT_124 | 124 | Upper-layer intratelencephalic | UL-IT | NEUR | VGLUT1 VGLUT2 | |
| UL-IT 126 | ULIT_126 | 126 | Upper-layer intratelencephalic | UL-IT | NEUR | VGLUT1 VGLUT2 | |
| Misc 404 | Misc_404 | 404 | Miscellaneous | Misc | NEUR | VGLUT1 | |
| Misc 170 | Misc_170 | 170 | Miscellaneous | Misc | NEUR | VGLUT1 VGLUT2 | |
| MSN 216 | MSN_216 | 216 | Medium spiny neuron | MSN | NEUR | GABA | MSN-D2 |
| MGE-IN 242 | MGE_242 | 242 | MGE interneuron | MGE-IN | NEUR | | |
| Amyg-Exc 405 | Amex_405 | 405 | Amygdala excitatory | Amyg-Exc | NEUR | VGLUT1 VGLUT2 | |
| DL-IT 136 | DLIT_136 | 136 | Deep-layer intratelencephalic | DL-IT | NEUR | VGLUT1 VGLUT2 | |
| eMSN 234 | EMSN_234 | 234 | Eccentric medium spiny neuron | eMSN | NEUR | GABA | MSN-D1 |
| MGE-IN 261 | MGE_261 | 261 | MGE interneuron | MGE-IN | NEUR | GABA | INT-PVALB |
| MSN 214 | MSN_214 | 214 | Medium spiny neuron | MSN | NEUR | GABA | MSN-D1 |
| LLC-IN 266 | LLC_266 | 266 | LAMP5-LHX6 and Chandelier | LLC-IN | NEUR | GABA | CHAND |
| eMSN 228 | EMSN_228 | 228 | Eccentric medium spiny neuron | eMSN | NEUR | GABA | MSN-D1 |
| MGE-IN 253 | MGE_253 | 253 | MGE interneuron | MGE-IN | NEUR | GABA | INT-SST |
| MSN 208 | MSN_208 | 208 | Medium spiny neuron | MSN | NEUR | GABA | MSN-D2 |
| Misc 166 | Misc_166 | 166 | Miscellaneous | Misc | NEUR | VGLUT1 VGLUT2 | |
| CA1-3 3 | Mono_3 | 3 | Hippocampal CA1-3 | CA1-3 | MONO | | |
| Amyg-Exc 172 | Amex_172 | 172 | Amygdala excitatory | Amyg-Exc | NEUR | VGLUT1 VGLUT2 | |
| MB-In 436 | Midi_436 | 436 | Midbrain-derived inhibitory | MB-In | NEUR | GABA | |
| Amyg-Exc 157 | Amex_157 | 157 | Amygdala excitatory | Amyg-Exc | NEUR | VGLUT1 VGLUT2 | |
| UL-IT 127 | ULIT_127 | 127 | Upper-layer intratelencephalic | UL-IT | NEUR | VGLUT1 VGLUT2 | |
| Amyg-Exc 158 | Amex_158 | 158 | Amygdala excitatory | Amyg-Exc | NEUR | VGLUT1 VGLUT2 | |
| MB-In 443 | Midi_443 | 443 | Midbrain-derived inhibitory | MB-In | NEUR | GABA | |
| MSN 212 | MSN_212 | 212 | Medium spiny neuron | MSN | NEUR | GABA | MSN-D2 |
| MSN 211 | MSN_211 | 211 | Medium spiny neuron | MSN | NEUR | GABA | MSN-D1 |

| Cluster (Yao naming) | Siletti cluster name | Siletti cluster ID | Supercluster | Supercluster abbreviation | Class auto-annotation | Neurotransmitter auto-annotation | Subtype auto-annotation |
|---|---|---|---|---|---|---|---|
| eMSN 231 | EMSN_231 | 231 | Eccentric medium spiny neuron | eMSN | NEUR | GABA | MSN-D1 |
| CA4 196 | CA4_196 | 196 | Hippocampal CA4 | CA4 | NEUR | VGLUT1 | |
| Misc 165 | Misc_165 | 165 | Miscellaneous | Misc | NEUR | VGLUT1 VGLUT2 | |
| DL-IT 146 | DLIT_146 | 146 | Deep-layer intratelencephalic | DL-IT | NEUR | VGLUT1 VGLUT2 | |
| Amyg-Exc 175 | Amex_175 | 175 | Amygdala excitatory | Amyg-Exc | NEUR | VGLUT1 VGLUT2 | |
| MGE-IN 243 | MGE_243 | 243 | MGE interneuron | MGE-IN | NEUR | GABA | INT-SST |
| MB-In 437 | Midi_437 | 437 | Midbrain-derived inhibitory | MB-In | NEUR | GABA | |
| Amyg-Exc 154 | Amex_154 | 154 | Amygdala excitatory | Amyg-Exc | NEUR | VGLUT1 VGLUT2 | |
| CA1-3 3 | Mono_3 | 3 | Hippocampal CA1-3 | CA1-3 | MONO | | |
| Splatter 420 | Splat_420 | 420 | Splatter | Splatter | NEUR | VGLUT2 | |
| MGE-IN 249 | MGE_249 | 249 | MGE interneuron | MGE-IN | NEUR | GABA | INT-SST |
| DL-IT 143 | DLIT_143 | 143 | Deep-layer intratelencephalic | DL-IT | NEUR | VGLUT1 | |
| eMSN 226 | EMSN_226 | 226 | Eccentric medium spiny neuron | eMSN | NEUR | GABA | MSN-D1 MSN-ECC |
| MB-In 435 | Midi_435 | 435 | Midbrain-derived inhibitory | MB-In | NEUR | GABA | |
| MGE-IN 257 | MGE_257 | 257 | MGE interneuron | MGE-IN | NEUR | GABA | INT-PVALB |
| MB-In 441 | Midi_441 | 441 | Midbrain-derived inhibitory | MB-In | NEUR | GABA | |
| eMSN 225 | EMSN_225 | 225 | Eccentric medium spiny neuron | eMSN | NEUR | GABA | MSN-D1 MSN-ECC |
| MGE-IN 263 | MGE_263 | 263 | MGE interneuron | MGE-IN | NEUR | GABA | |
| UL-IT 122 | ULIT_122 | 122 | Upper-layer intratelencephalic | UL-IT | NEUR | VGLUT1 VGLUT2 | |
| MB-In 442 | Midi_442 | 442 | Midbrain-derived inhibitory | MB-In | NEUR | GABA | |
| MSN 221 | MSN_221 | 221 | Medium spiny neuron | MSN | NEUR | GABA | MSN-D2 |
| eMSN 222 | EMSN_222 | 222 | Eccentric medium spiny neuron | eMSN | NEUR | GABA | MSN-D1 MSN-D2 MSN-ECC |
| DL-CT/6b 102 | DLCT6b_102 | 102 | Deep-layer corticothalamic and 6b | DL-CT/6b | NEUR | VGLUT1 | |
| MGE-IN 241 | MGE_241 | 241 | MGE interneuron | MGE-IN | NEUR | GABA | INT-SST |
| CA4 197 | CA4_197 | 197 | Hippocampal CA4 | CA4 | NEUR | VGLUT1 | |
| Amyg-Exc 174 | Amex_174 | 174 | Amygdala excitatory | Amyg-Exc | NEUR | VGLUT1 VGLUT2 | |
| MGE-IN 258 | MGE_258 | 258 | MGE interneuron | MGE-IN | NEUR | GABA VGLUT3 | INT-SST |
| HiDG 200 | DG_200 | 200 | Hippocampal dentate gyrus | HiDG | NEUR | VGLUT1 | DG-GRAN |
| Amyg-Exc 407 | Amex_407 | 407 | Amygdala excitatory | Amyg-Exc | NEUR | VGLUT2 VGLUT3 | |
| Misc 178 | Misc_178 | 178 | Miscellaneous | Misc | NEUR | VGLUT1 VGLUT2 | |
| DL-IT 148 | DLIT_148 | 148 | Deep-layer intratelencephalic | DL-IT | NEUR | VGLUT1 | |
| CA1-3 3 | Mono_3 | 3 | Hippocampal CA1-3 | CA1-3 | MONO | | |
| HiDG 202 | DG_202 | 202 | Hippocampal dentate gyrus | HiDG | NEUR | VGLUT1 | DG-GRAN |
| LLC-IN 269 | LLC_269 | 269 | LAMP5-LHX6 and Chandelier | LLC-IN | NEUR | GABA | INT-LAMP5-LHX6 |
| U RhombLip 308 | URL_308 | 308 | Upper rhombic lip | U RhombLip | | VGLUT1 | UBC |
| DL-NP 92 | DLNP_92 | 92 | Deep-layer near-projecting | DL-NP | NEUR | VGLUT1 | |
| MB-In 444 | Midi_444 | 444 | Midbrain-derived inhibitory | MB-In | NEUR | GABA | |
| Splatter 422 | Splat_422 | 422 | Splatter | Splatter | NEUR | GABA | |
| Astr 59 | Astro_59 | 59 | Astrocyte | Astr | ASTRO | | |
| Splatter 386 | Splat_386 | 386 | Splatter | Splatter | NEUR | VGLUT2 | |
| eMSN 233 | EMSN_233 | 233 | Eccentric medium spiny neuron | eMSN | NEUR | GABA | MSN-D1 MSN-ECC |
| UL-IT 121 | ULIT_121 | 121 | Upper-layer intratelencephalic | UL-IT | NEUR | VGLUT1 VGLUT2 | |
| Splatter 428 | Splat_428 | 428 | Splatter | Splatter | NEUR | GABA | |
| UL-IT 135 | ULIT_135 | 135 | Upper-layer intratelencephalic | UL-IT | NEUR | VGLUT1 VGLUT2 | |
| Misc 176 | Misc_176 | 176 | Miscellaneous | Misc | NEUR | VGLUT1 VGLUT2 | |
| Amyg-Exc 162 | Amex_162 | 162 | Amygdala excitatory | Amyg-Exc | NEUR | VGLUT1 VGLUT2 | |
| HiDG 201 | DG_201 | 201 | Hippocampal dentate gyrus | HiDG | NEUR | VGLUT1 | DG-GRAN |

| Cluster (Yao naming) | Siletti cluster name | Siletti cluster ID | Supercluster | Supercluster abbreviation | Class auto-annotation | Neurotransmitter auto-annotation | Subtype auto-annotation |
|---|---|---|---|---|---|---|---|
| MSN 209 | MSN_209 | 209 | Medium spiny neuron | MSN | NEUR | GABA | MSN-D1 |
| MGE-IN 240 | MGE_240 | 240 | MGE interneuron | MGE-IN | NEUR | GABA | INT-SST |
| CGE-IN 292 | CGE_292 | 292 | CGE interneuron | CGE-IN | NEUR | GABA | INT-VIP |
| Amyg-Exc 171 | Amex_171 | 171 | Amygdala excitatory | Amyg-Exc | NEUR | VGLUT1 VGLUT2 | |
| Splatter 364 | Splat_364 | 364 | Splatter | Splatter | NEUR | VGLUT2 | |
| Misc 177 | Misc_177 | 177 | Miscellaneous | Misc | NEUR | VGLUT1 VGLUT2 | |
| Mam 327 | Mmb_327 | 327 | Mammillary body | Mam | NEUR | VGLUT2 | |
| MGE-IN 255 | MGE_255 | 255 | MGE interneuron | MGE-IN | NEUR | GABA | |
| Cer-In 302 | CBI_302 | 302 | Cerebellar inhibitory | Cer-In | NEUR | GLY | CB-GranLayerInt |
| Astr 60 | Astro_60 | 60 | Astrocyte | Astr | ASTRO | | |
| eMSN 229 | EMSN_229 | 229 | Eccentric medium spiny neuron | eMSN | NEUR | GABA | MSN-D1 |
| Misc 132 | Misc_132 | 132 | Miscellaneous | Misc | NEUR | VGLUT1 VGLUT2 | |
| DL-NP 95 | DLNP_95 | 95 | Deep-layer near-projecting | DL-NP | NEUR | VGLUT1 | |
| UL-IT 123 | ULIT_123 | 123 | Upper-layer intratelencephalic | UL-IT | NEUR | VGLUT1 | |
| CA1-3 3 | Mono_3 | 3 | Hippocampal CA1-3 | CA1-3 | MONO | | |
| MB-In 434 | Midi_434 | 434 | Midbrain-derived inhibitory | MB-In | NEUR | GABA | |
| MSN 215 | MSN_215 | 215 | Medium spiny neuron | MSN | NEUR | GABA | MSN-D1 |
| MGE-IN 262 | MGE_262 | 262 | MGE interneuron | MGE-IN | NEUR | GABA | INT-PVALB |
| MSN 217 | MSN_217 | 217 | Medium spiny neuron | MSN | NEUR | GABA | MSN-D2 |
| MSN 213 | MSN_213 | 213 | Medium spiny neuron | MSN | NEUR | GABA | MSN-D2 |
| CGE-IN 289 | CGE_289 | 289 | CGE interneuron | CGE-IN | NEUR | GABA | INT-VIP |
| DL-IT 140 | DLIT_140 | 140 | Deep-layer intratelencephalic | DL-IT | NEUR | VGLUT1 VGLUT2 | |
| Splatter 237 | Splat_237 | 237 | Splatter | Splatter | NEUR | GABA | INT-SST |
| MSN 207 | MSN_207 | 207 | Medium spiny neuron | MSN | NEUR | GABA | MSN-D2 |
| Mam 328 | Mmb_328 | 328 | Mammillary body | Mam | NEUR | HDC VGLUT2 | |
| DL-CT/6b 109 | DLCT6b_109 | 109 | Deep-layer corticothalamic and 6b | DL-CT/6b | NEUR | VGLUT1 | |
| MGE-IN 254 | MGE_254 | 254 | MGE interneuron | MGE-IN | NEUR | GABA VGLUT3 | INT-SST |
| MGE-IN 250 | MGE_250 | 250 | MGE interneuron | MGE-IN | NEUR | GABA | |
| Misc 167 | Misc_167 | 167 | Miscellaneous | Misc | NEUR | VGLUT1 | |
| Mam 333 | Mmb_333 | 333 | Mammillary body | Mam | NEUR | HDC VGLUT2 | |
| Splatter 429 | Splat_429 | 429 | Splatter | Splatter | NEUR | GABA | |
| MGE-IN 244 | MGE_244 | 244 | MGE interneuron | MGE-IN | NEUR | GABA | INT-SST INT-SST-CHODL |
| MGE-IN 239 | MGE_239 | 239 | MGE interneuron | MGE-IN | NEUR | GABA | INT-SST |
| Splatter 403 | Splat_403 | 403 | Splatter | Splatter | NEUR | GABA | |
| DL-NP 87 | DLNP_87 | 87 | Deep-layer near-projecting | DL-NP | NEUR | VGLUT1 VGLUT2 | |
| CA1-3 3 | Mono_3 | 3 | Hippocampal CA1-3 | CA1-3 | MONO | | |
| MGE-IN 248 | MGE_248 | 248 | MGE interneuron | MGE-IN | NEUR | GABA | INT-SST |
| CA1-3 3 | Mono_3 | 3 | Hippocampal CA1-3 | CA1-3 | MONO | | |
| UL-IT 134 | ULIT_134 | 134 | Upper-layer intratelencephalic | UL-IT | NEUR | VGLUT1 VGLUT2 | |
| Splatter 313 | Splat_313 | 313 | Splatter | Splatter | NEUR | VGLUT2 | |
| Splatter 425 | Splat_425 | 425 | Splatter | Splatter | NEUR | GABA | |
| DL-IT 147 | DLIT_147 | 147 | Deep-layer intratelencephalic | DL-IT | NEUR | VGLUT1 | |
| Splatter 410 | Splat_410 | 410 | Splatter | Splatter | NEUR | VGLUT2 | |
| CGE-IN 286 | CGE_286 | 286 | CGE interneuron | CGE-IN | NEUR | GABA | INT-LAMP5 |
| CA4 192 | CA4_192 | 192 | Hippocampal CA4 | CA4 | NEUR | VGLUT1 | |
| DL-NP 93 | DLNP_93 | 93 | Deep-layer near-projecting | DL-NP | NEUR | VGLUT1 | |
| CA1-3 3 | Mono_3 | 3 | Hippocampal CA1-3 | CA1-3 | MONO | | |

| Cluster (Yao naming) | Siletti cluster name | Siletti cluster ID | Supercluster | Supercluster abbreviation | Class auto-annotation | Neurotransmitter auto-annotation | Subtype auto-annotation |
|---|---|---|---|---|---|---|---|
| Astr 58 | Astro_58 | 58 | Astrocyte | Astr | ASTRO | | |
| CA1-3 3 | Mono_3 | 3 | Hippocampal CA1-3 | CA1-3 | MONO | | |
| CA4 198 | CA4_198 | 198 | Hippocampal CA4 | CA4 | NEUR | VGLUT1 VGLUT2 | |
| CA1-3 3 | Mono_3 | 3 | Hippocampal CA1-3 | CA1-3 | MONO | | |
| Thal-Exc 459 | Thex_459 | 459 | Thalamic excitatory | Thal-Exc | NEUR | VGLUT2 | |
| Splatter 387 | Splat_387 | 387 | Splatter | Splatter | NEUR | VGLUT2 | |
| Splatter 396 | Splat_396 | 396 | Splatter | Splatter | NEUR | VGLUT3 | |
| Astr 53 | Astro_53 | 53 | Astrocyte | Astr | ASTRO | | |
| MSN 430 | MSN_430 | 430 | Medium spiny neuron | MSN | NEUR | GABA | |
| DL-IT 137 | DLIT_137 | 137 | Deep-layer intratelencephalic | DL-IT | NEUR | VGLUT1 VGLUT2 | |
| Splatter 367 | Splat_367 | 367 | Splatter | Splatter | NEUR | VGLUT2 | |
| MGE-IN 245 | MGE_245 | 245 | MGE interneuron | MGE-IN | NEUR | GABA | INT-SST INT-SST-CHODL |
| LLC-IN 264 | LLC_264 | 264 | LAMP5-LHX6 and Chandelier | LLC-IN | | GABA | CHAND |
| LLC-IN 268 | LLC_268 | 268 | LAMP5-LHX6 and Chandelier | LLC-IN | NEUR | GABA | INT-LAMP5-LHX6 |
| Mam 329 | Mmb_329 | 329 | Mammillary body | Mam | NEUR | HDC VGLUT2 | |
| DL-IT 149 | DLIT_149 | 149 | Deep-layer intratelencephalic | DL-IT | NEUR | VGLUT1 | |
| DL-IT 151 | DLIT_151 | 151 | Deep-layer intratelencephalic | DL-IT | NEUR | VGLUT1 VGLUT2 VGLUT3 | |
| eMSN 223 | EMSN_223 | 223 | Eccentric medium spiny neuron | eMSN | NEUR | GABA | MSN-D1 MSN-ECC |
| DL-NP 91 | DLNP_91 | 91 | Deep-layer near-projecting | DL-NP | NEUR | VGLUT1 | |
| Splatter 393 | Splat_393 | 393 | Splatter | Splatter | NEUR | GABA | |
| Astr 54 | Astro_54 | 54 | Astrocyte | Astr | ASTRO | | |
| DL-NP 96 | DLNP_96 | 96 | Deep-layer near-projecting | DL-NP | NEUR | VGLUT1 VGLUT2 | |
| DL-IT 141 | DLIT_141 | 141 | Deep-layer intratelencephalic | DL-IT | NEUR | VGLUT1 VGLUT2 | |
| Amyg-Exc 406 | Amex_406 | 406 | Amygdala excitatory | Amyg-Exc | NEUR | VGLUT1 VGLUT2 | |
| MGE-IN 252 | MGE_252 | 252 | MGE interneuron | MGE-IN | NEUR | GABA VGLUT3 | INT-SST |
| CA1-3 3 | Mono_3 | 3 | Hippocampal CA1-3 | CA1-3 | MONO | | |
| eMSN 227 | EMSN_227 | 227 | Eccentric medium spiny neuron | eMSN | NEUR | GABA | MSN-D1 |
| Astr 64 | Astro_64 | 64 | Astrocyte | Astr | ASTRO | | |
| MSN 218 | MSN_218 | 218 | Medium spiny neuron | MSN | NEUR | | |
| Splatter 395 | Splat_395 | 395 | Splatter | Splatter | NEUR | DA VGLUT2 | |
| Mam 332 | Mmb_332 | 332 | Mammillary body | Mam | NEUR | VGLUT2 | |
| eMSN 224 | EMSN_224 | 224 | Eccentric medium spiny neuron | eMSN | NEUR | GABA | MSN-D1 MSN-D2 MSN-ECC |
| MSN 210 | MSN_210 | 210 | Medium spiny neuron | MSN | NEUR | GABA | MSN-D1 |
| Astr 56 | Astro_56 | 56 | Astrocyte | Astr | ASTRO | | |
| Misc 117 | L5ET_117 | 117 | Miscellaneous | Misc | NEUR | VGLUT1 | L5-ET VonEcon |
| DL-IT 145 | DLIT_145 | 145 | Deep-layer intratelencephalic | DL-IT | NEUR | VGLUT1 VGLUT2 | |
| Amyg-Exc 419 | Amex_419 | 419 | Amygdala excitatory | Amyg-Exc | NEUR | VGLUT1 VGLUT2 | |
| UL-IT 129 | ULIT_129 | 129 | Upper-layer intratelencephalic | UL-IT | NEUR | VGLUT1 VGLUT2 | |
| UL-IT 125 | ULIT_125 | 125 | Upper-layer intratelencephalic | UL-IT | NEUR | VGLUT1 VGLUT2 | |
| CGE-IN 296 | CGE_296 | 296 | CGE interneuron | CGE-IN | NEUR | GABA | INT-VIP |
| Astr 52 | Astro_52 | 52 | Astrocyte | Astr | ASTRO | | |
| Astr 63 | Astro_63 | 63 | Astrocyte | Astr | ASTRO | | |
| CA4 191 | CA4_191 | 191 | Hippocampal CA4 | CA4 | NEUR | VGLUT1 | |
| MGE-IN 247 | MGE_247 | 247 | MGE interneuron | MGE-IN | NEUR | GABA | |
| Mam 326 | Mmb_326 | 326 | Mammillary body | Mam | NEUR | VGLUT2 | |
| Splatter 392 | Splat_392 | 392 | Splatter | Splatter | NEUR | GABA | |
| Thal-Exc 448 | Thex_448 | 448 | Thalamic excitatory | Thal-Exc | NEUR | VGLUT2 | |

| Cluster (Yao naming) | Siletti cluster name | Siletti cluster ID | Supercluster | Supercluster abbreviation | Class auto-annotation | Neurotransmitter auto-annotation | Subtype auto-annotation |
|---|---|---|---|---|---|---|---|
| DL-CT/6b 103 | DLCT6b_103 | 103 | Deep-layer corticothalamic and 6b | DL-CT/6b | NEUR | VGLUT1 | |
| CA1-3 3 | Mono_3 | 3 | Hippocampal CA1-3 | CA1-3 | MONO | | |
| Splatter 340 | Splat_340 | 340 | Splatter | Splatter | NEUR | VGLUT2 | |
| Amyg-Exc 160 | Amex_160 | 160 | Amygdala excitatory | Amyg-Exc | NEUR | VGLUT1 VGLUT2 | |
| CGE-IN 295 | CGE_295 | 295 | CGE interneuron | CGE-IN | NEUR | GABA | INT-VIP |
| Splatter 400 | Splat_400 | 400 | Splatter | Splatter | NEUR | CHOL VGLUT3 | |
| Astr 62 | Astro_62 | 62 | Astrocyte | Astr | ASTRO | | |
| DL-IT 142 | DLIT_142 | 142 | Deep-layer intratelencephalic | DL-IT | NEUR | VGLUT1 | |
| MSN 427 | MSN_427 | 427 | Medium spiny neuron | MSN | NEUR | GABA | MSN-D1 |
| HiDG 199 | DG_199 | 199 | Hippocampal dentate gyrus | HiDG | NEUR | VGLUT1 | DG-GRAN |
| UL-IT 128 | ULIT_128 | 128 | Upper-layer intratelencephalic | UL-IT | NEUR | VGLUT1 VGLUT2 | |
| Splatter 235 | Splat_235 | 235 | Splatter | Splatter | NEUR | GABA | INT-SST-CHODL |
| Splatter 349 | Splat_349 | 349 | Splatter | Splatter | NEUR | GLY | |
| Astr 61 | Astro_61 | 61 | Astrocyte | Astr | ASTRO | | |
| DL-NP 85 | DLNP_85 | 85 | Deep-layer near-projecting | DL-NP | NEUR | VGLUT1 VGLUT2 VGLUT3 | |
| LLC-IN 270 | LLC_270 | 270 | LAMP5-LHX6 and Chandelier | LLC-IN | NEUR | GABA | INT-LAMP5-LHX6 |
| eMSN 426 | EMSN_426 | 426 | Eccentric medium spiny neuron | eMSN | NEUR | GABA | MSN-D1 |
| LLC-IN 271 | LLC_271 | 271 | LAMP5-LHX6 and Chandelier | LLC-IN | NEUR | GABA | INT-LAMP5-LHX6 |
| Mam 331 | Mmb_331 | 331 | Mammillary body | Mam | NEUR | VGLUT2 | |
| CA1-3 3 | Mono_3 | 3 | Hippocampal CA1-3 | CA1-3 | MONO | | |
| Splatter 421 | Splat_421 | 421 | Splatter | Splatter | NEUR | GABA | |
| Astr 57 | Astro_57 | 57 | Astrocyte | Astr | ASTRO | | |
| CGE-IN 290 | CGE_290 | 290 | CGE interneuron | CGE-IN | NEUR | GABA | INT-VIP |
| CGE-IN 276 | CGE_276 | 276 | CGE interneuron | CGE-IN | NEUR | GABA VGLUT3 | INT-VIP |
| Splatter 341 | Splat_341 | 341 | Splatter | Splatter | NEUR | VGLUT2 | |
| DL-NP 90 | DLNP_90 | 90 | Deep-layer near-projecting | DL-NP | NEUR | VGLUT1 | |
| eMSN 232 | EMSN_232 | 232 | Eccentric medium spiny neuron | eMSN | NEUR | GABA | MSN-D1 |
| Splatter 378 | Splat_378 | 378 | Splatter | Splatter | NEUR | HDC VGLUT2 | |
| LLC-IN 273 | LLC_273 | 273 | LAMP5-LHX6 and Chandelier | LLC-IN | NEUR | GABA | INT-LAMP5-LHX6 |
| CGE-IN 287 | CGE_287 | 287 | CGE interneuron | CGE-IN | NEUR | GABA | INT-LAMP5 |
| MGE-IN 251 | MGE_251 | 251 | MGE interneuron | MGE-IN | NEUR | GABA | |
| Cer-In 300 | CBI_300 | 300 | Cerebellar inhibitory | Cer-In | NEUR | GABA | CB-PurkLayerInt |
| Splatter 418 | Splat_418 | 418 | Splatter | Splatter | NEUR | HDC VGLUT2 | |
| Splatter 371 | Splat_371 | 371 | Splatter | Splatter | NEUR | VGLUT2 | |
| L RhombLip 322 | LRL_322 | 322 | Lower rhombic lip | L RhombLip | NEUR | VGLUT1 VGLUT2 | |
| HiDG 205 | DG_205 | 205 | Hippocampal dentate gyrus | HiDG | NEUR | VGLUT1 | DG-GRAN |
| Thal-Exc 450 | Thex_450 | 450 | Thalamic excitatory | Thal-Exc | | VGLUT2 | |
| Thal-Exc 449 | Thex_449 | 449 | Thalamic excitatory | Thal-Exc | | VGLUT2 | |
| LLC-IN 267 | LLC_267 | 267 | LAMP5-LHX6 and Chandelier | LLC-IN | NEUR | GABA VGLUT3 | INT-LAMP5-LHX6 |
| U RhombLip 309 | URL_309 | 309 | Upper rhombic lip | U RhombLip | NEUR | VGLUT1 VGLUT2 | |
| Splatter 363 | Splat_363 | 363 | Splatter | Splatter | NEUR | VGLUT2 | |
| DL-CT/6b 98 | DLCT6b_98 | 98 | Deep-layer corticothalamic and 6b | DL-CT/6b | NEUR | VGLUT1 | |
| DL-CT/6b 110 | DLCT6b_110 | 110 | Deep-layer corticothalamic and 6b | DL-CT/6b | NEUR | VGLUT1 | |
| Splatter 412 | Splat_412 | 412 | Splatter | Splatter | NEUR | GABA HDC VGLUT2 | |
| Mam 325 | Mmb_325 | 325 | Mammillary body | Mam | NEUR | VGLUT2 | |
| Misc 116 | Misc_116 | 116 | Miscellaneous | Misc | NEUR | VGLUT1 | |
| CA4 190 | CA4_190 | 190 | Hippocampal CA4 | CA4 | NEUR | VGLUT1 | |

| Cluster (Yao naming) | Siletti cluster name | Siletti cluster ID | Supercluster | Supercluster abbreviation | Class auto-annotation | Neurotransmitter auto-annotation | Subtype auto-annotation |
|---|---|---|---|---|---|---|---|
| DL-CT/6b 101 | DLCT6b_101 | 101 | Deep-layer corticothalamic and 6b | DL-CT/6b | NEUR | VGLUT1 | |
| DL-CT/6b 100 | DLCT6b_100 | 100 | Deep-layer corticothalamic and 6b | DL-CT/6b | NEUR | VGLUT1 | |
| DL-NP 88 | DLNP_88 | 88 | Deep-layer near-projecting | DL-NP | NEUR | VGLUT1 | |
| DL-NP 83 | DLNP_83 | 83 | Deep-layer near-projecting | DL-NP | NEUR | VGLUT1 | |
| U RhombLip 297 | URL_297 | 297 | Upper rhombic lip | U RhombLip | | VGLUT1 | |
| Cer-In 303 | CBI_303 | 303 | Cerebellar inhibitory | Cer-In | | GABA | CB-MolLayerInt1 |
| DL-NP 86 | DLNP_86 | 86 | Deep-layer near-projecting | DL-NP | NEUR | VGLUT1 VGLUT2 | |
| Splatter 402 | Splat_402 | 402 | Splatter | Splatter | NEUR | GABA | |
| Splatter 375 | Splat_375 | 375 | Splatter | Splatter | NEUR | VGLUT2 VGLUT3 | |
| Cer-In 299 | CBI_299 | 299 | Cerebellar inhibitory | Cer-In | NEUR | GLY | CB-PurkLayerInt |
| DL-CT/6b 107 | DLCT6b_107 | 107 | Deep-layer corticothalamic and 6b | DL-CT/6b | NEUR | VGLUT1 | |
| CA1-3 3 | Mono_3 | 3 | Hippocampal CA1-3 | CA1-3 | MONO | | |
| Splatter 343 | Splat_343 | 343 | Splatter | Splatter | NEUR | VGLUT2 | |
| Cer-In 304 | CBI_304 | 304 | Cerebellar inhibitory | Cer-In | | GABA | CB-MolLayerInt1 |
| MB-In 433 | Midi_433 | 433 | Midbrain-derived inhibitory | MB-In | NEUR | GABA | |
| MSN 219 | MSN_219 | 219 | Medium spiny neuron | MSN | NEUR | GABA | MSN-D1 |
| CGE-IN 288 | CGE_288 | 288 | CGE interneuron | CGE-IN | NEUR | GABA | INT-LAMP5 |
| UL-IT 130 | ULIT_130 | 130 | Upper-layer intratelencephalic | UL-IT | NEUR | VGLUT1 VGLUT2 | |
| DL-IT 139 | DLIT_139 | 139 | Deep-layer intratelencephalic | DL-IT | NEUR | VGLUT1 VGLUT2 | |
| CA1-3 3 | Mono_3 | 3 | Hippocampal CA1-3 | CA1-3 | MONO | | |
| CA1-3 3 | Mono_3 | 3 | Hippocampal CA1-3 | CA1-3 | MONO | | |
| DL-NP 89 | DLNP_89 | 89 | Deep-layer near-projecting | DL-NP | NEUR | VGLUT1 VGLUT2 | |
| CGE-IN 277 | CGE_277 | 277 | CGE interneuron | CGE-IN | NEUR | GABA VGLUT3 | |
| Splatter 391 | Splat_391 | 391 | Splatter | Splatter | NEUR | GABA | |
| OPC 33 | OPC_33 | 33 | Oligodendrocyte precursor | OPC | OPC | | |
| Splatter 346 | Splat_346 | 346 | Splatter | Splatter | NEUR | VGLUT2 | |
| MSN 206 | MSN_206 | 206 | Medium spiny neuron | MSN | NEUR | | |
| DL-CT/6b 112 | DLCT6b_112 | 112 | Deep-layer corticothalamic and 6b | DL-CT/6b | NEUR | VGLUT1 | |
| UL-IT 138 | ULIT_138 | 138 | Upper-layer intratelencephalic | UL-IT | NEUR | VGLUT1 VGLUT2 | |
| CGE-IN 293 | CGE_293 | 293 | CGE interneuron | CGE-IN | NEUR | GABA | INT-VIP |
| DL-CT/6b 108 | DLCT6b_108 | 108 | Deep-layer corticothalamic and 6b | DL-CT/6b | NEUR | VGLUT1 | |
| DL-CT/6b 104 | DLCT6b_104 | 104 | Deep-layer corticothalamic and 6b | DL-CT/6b | NEUR | VGLUT1 | |
| Splatter 423 | Splat_423 | 423 | Splatter | Splatter | NEUR | GABA | INT-LAMP5 |
| Splatter 366 | Splat_366 | 366 | Splatter | Splatter | NEUR | VGLUT2 | |
| Mam 330 | Mmb_330 | 330 | Mammillary body | Mam | NEUR | VGLUT2 | |
| Splatter 385 | Splat_385 | 385 | Splatter | Splatter | NEUR | VGLUT2 | |
| UL-IT 133 | ULIT_133 | 133 | Upper-layer intratelencephalic | UL-IT | NEUR | VGLUT1 VGLUT2 | |
| Misc 114 | L5ET_114 | 114 | Miscellaneous | Misc | NEUR | VGLUT1 | L5-ET |
| MGE-IN 256 | MGE_256 | 256 | MGE interneuron | MGE-IN | NEUR | GABA VGLUT3 | INT-PVALB |
| OPC 32 | OPC_32 | 32 | Oligodendrocyte precursor | OPC | OPC | | |
| DL-CT/6b 106 | DLCT6b_106 | 106 | Deep-layer corticothalamic and 6b | DL-CT/6b | NEUR | VGLUT1 | |
| Cer-In 306 | CBI_306 | 306 | Cerebellar inhibitory | Cer-In | | GABA | CB-MolLayerInt1 |
| LLC-IN 272 | LLC_272 | 272 | LAMP5-LHX6 and Chandelier | LLC-IN | NEUR | GABA | INT-LAMP5-LHX6 |
| Splatter 373 | Splat_373 | 373 | Splatter | Splatter | NEUR | VGLUT2 | |
| Splatter 334 | Splat_334 | 334 | Splatter | Splatter | NEUR | CHOL VGLUT1 VGLUT2 | |
| Cer-In 301 | CBI_301 | 301 | Cerebellar inhibitory | Cer-In | NEUR | GLY | CB-MolLayerInt1 |
| Splatter 424 | Splat_424 | 424 | Splatter | Splatter | NEUR | GABA | |

| Cluster (Yao naming) | Siletti cluster name | Siletti cluster ID | Supercluster | Supercluster abbreviation | Class auto-annotation | Neurotransmitter auto-annotation | Subtype auto-annotation |
|---|---|---|---|---|---|---|---|
| CA4 193 | CA4_193 | 193 | Hippocampal CA4 | CA4 | NEUR | VGLUT1 | |
| DL-IT 150 | DLIT_150 | 150 | Deep-layer intratelencephalic | DL-IT | NEUR | VGLUT1 VGLUT2 | |
| DL-CT/6b 97 | DLCT6b_97 | 97 | Deep-layer corticothalamic and 6b | DL-CT/6b | NEUR | VGLUT1 | |
| Splatter 368 | Splat_368 | 368 | Splatter | Splatter | NEUR | VGLUT2 | |
| CA4 195 | CA4_195 | 195 | Hippocampal CA4 | CA4 | NEUR | VGLUT1 | |
| Splatter 354 | Splat_354 | 354 | Splatter | Splatter | NEUR | GABA | |
| Splatter 389 | Splat_389 | 389 | Splatter | Splatter | NEUR | GABA | |
| MSN 220 | MSN_220 | 220 | Medium spiny neuron | MSN | NEUR | GABA | MSN-D1 |
| Splatter 348 | Splat_348 | 348 | Splatter | Splatter | NEUR | GLY | |
| Thal-Exc 457 | Thex_457 | 457 | Thalamic excitatory | Thal-Exc | NEUR | VGLUT2 | |
| Thal-Exc 372 | Thex_372 | 372 | Thalamic excitatory | Thal-Exc | NEUR | VGLUT2 | |
| eMSN 230 | EMSN_230 | 230 | Eccentric medium spiny neuron | eMSN | NEUR | GABA | MSN-D1 |
| DL-CT/6b 99 | DLCT6b_99 | 99 | Deep-layer corticothalamic and 6b | DL-CT/6b | NEUR | VGLUT1 | |
| Thal-Exc 452 | Thex_452 | 452 | Thalamic excitatory | Thal-Exc | | VGLUT2 | |
| Cer-In 307 | CBI_307 | 307 | Cerebellar inhibitory | Cer-In | NEUR | GABA | CB-MolLayerInt2 |
| Mam 323 | Mmb_323 | 323 | Mammillary body | Mam | NEUR | VGLUT2 | |
| Thal-Exc 454 | Thex_454 | 454 | Thalamic excitatory | Thal-Exc | | VGLUT2 | |
| Amyg-Exc 408 | Amex_408 | 408 | Amygdala excitatory | Amyg-Exc | NEUR | VGLUT1 VGLUT2 | |
| Splatter 417 | Splat_417 | 417 | Splatter | Splatter | NEUR | GABA HDC | |
| DL-CT/6b 105 | DLCT6b_105 | 105 | Deep-layer corticothalamic and 6b | DL-CT/6b | NEUR | VGLUT1 | |
| Splatter 376 | Splat_376 | 376 | Splatter | Splatter | NEUR | VGLUT2 | |
| CGE-IN 284 | CGE_284 | 284 | CGE interneuron | CGE-IN | NEUR | GABA | INT-VIP |
| Splatter 382 | Splat_382 | 382 | Splatter | Splatter | NEUR | VGLUT2 | |
| Astr 55 | Astro_55 | 55 | Astrocyte | Astr | ASTRO | | |
| OPC 34 | OPC_34 | 34 | Oligodendrocyte precursor | OPC | OPC | | |
| B 51 | Bgl_51 | 51 | Bergmann glia | B | ASTRO | | |
| CGE-IN 294 | CGE_294 | 294 | CGE interneuron | CGE-IN | NEUR | GABA | INT-VIP |
| Splatter 415 | Splat_415 | 415 | Splatter | Splatter | NEUR | GABA VGLUT2 | |
| Thal-Exc 456 | Thex_456 | 456 | Thalamic excitatory | Thal-Exc | NEUR | VGLUT2 | |
| Splatter 365 | Splat_365 | 365 | Splatter | Splatter | NEUR | GABA VGLUT2 | |
| Mam 324 | Mmb_324 | 324 | Mammillary body | Mam | NEUR | VGLUT2 | |
| Splatter 374 | Splat_374 | 374 | Splatter | Splatter | NEUR | VGLUT2 | |
| Splatter 383 | Splat_383 | 383 | Splatter | Splatter | NEUR | VGLUT2 VGLUT3 | |
| Thal-Exc 447 | Thex_447 | 447 | Thalamic excitatory | Thal-Exc | | VGLUT2 | |
| OPC 35 | OPC_35 | 35 | Oligodendrocyte precursor | OPC | OPC | | |
| Fibr 30 | Fbl_30 | 30 | Fibroblast | Fibr | CHRP | | ARACH |
| U RhombLip 311 | URL_311 | 311 | Upper rhombic lip | U RhombLip | NEUR | VGLUT1 | CB-GRAN |
| Misc 118 | L5ET_118 | 118 | Miscellaneous | Misc | NEUR | VGLUT1 | L5-ET |
| Thal-Exc 460 | Thex_460 | 460 | Thalamic excitatory | Thal-Exc | NEUR | VGLUT2 | |
| Splatter 355 | Splat_355 | 355 | Splatter | Splatter | NEUR | GABA | |
| Splatter 394 | Splat_394 | 394 | Splatter | Splatter | NEUR | VGLUT2 | |
| CGE-IN 282 | CGE_282 | 282 | CGE interneuron | CGE-IN | NEUR | GABA | |
| L RhombLip 319 | LRL_319 | 319 | Lower rhombic lip | L RhombLip | NEUR | VGLUT1 VGLUT2 | |
| MGE-IN 246 | MGE_246 | 246 | MGE interneuron | MGE-IN | NEUR | GABA | INT-SST |
| Splatter 345 | Splat_345 | 345 | Splatter | Splatter | NEUR | VGLUT2 | |
| Splatter 358 | Splat_358 | 358 | Splatter | Splatter | NEUR | GABA | |
| U RhombLip 310 | URL_310 | 310 | Upper rhombic lip | U RhombLip | NEUR | VGLUT1 | CB-GRAN |

| Cluster (Yao naming) | Siletti cluster name | Siletti cluster ID | Supercluster | Supercluster abbreviation | Class auto-annotation | Neurotransmitter auto-annotation | Subtype auto-annotation |
|---|---|---|---|---|---|---|---|
| Fibr 31 | Fbl_31 | 31 | Fibroblast | Fibr | CHRP | | ABC PIA |
| Misc 3 | Mono_3 | 3 | Miscellaneous | Misc | MONO | | |
| L RhombLip 321 | LRL_321 | 321 | Lower rhombic lip | L RhombLip | NEUR | VGLUT1 VGLUT2 | |
| L RhombLip 318 | LRL_318 | 318 | Lower rhombic lip | L RhombLip | NEUR | VGLUT1 VGLUT2 | |
| UL-IT 131 | ULIT_131 | 131 | Upper-layer intratelencephalic | UL-IT | NEUR | VGLUT1 VGLUT2 | |
| Splatter 388 | Splat_388 | 388 | Splatter | Splatter | NEUR | VGLUT2 | |
| Splatter 413 | Splat_413 | 413 | Splatter | Splatter | NEUR | VGLUT2 | |
| L RhombLip 315 | LRL_315 | 315 | Lower rhombic lip | L RhombLip | NEUR | VGLUT1 VGLUT2 | |
| Misc 113 | L5ET_113 | 113 | Miscellaneous | Misc | NEUR | VGLUT1 | L5-ET |
| L RhombLip 317 | LRL_317 | 317 | Lower rhombic lip | L RhombLip | NEUR | VGLUT1 VGLUT2 | |
| CGE-IN 279 | CGE_279 | 279 | CGE interneuron | CGE-IN | NEUR | GABA VGLUT3 | INT-VIP |
| Splatter 369 | Splat_369 | 369 | Splatter | Splatter | NEUR | VGLUT2 | |
| MiG 11 | Mgl_11 | 11 | Microglia | MiG | MGL | | |
| Cer-In 305 | CBI_305 | 305 | Cerebellar inhibitory | Cer-In | NEUR | GABA | CB-MolLayerInt1 CB-MolLayerInt2 |
| CGE-IN 278 | CGE_278 | 278 | CGE interneuron | CGE-IN | NEUR | GABA VGLUT3 | |
| DL-CT/6b 111 | DLCT6b_111 | 111 | Deep-layer corticothalamic and 6b | DL-CT/6b | NEUR | VGLUT1 VGLUT2 | |
| Splatter 432 | Splat_432 | 432 | Splatter | Splatter | NEUR | HDC | |
| Splatter 359 | Splat_359 | 359 | Splatter | Splatter | NEUR | GABA | |
| CGE-IN 283 | CGE_283 | 283 | CGE interneuron | CGE-IN | NEUR | GABA | |
| L RhombLip 316 | LRL_316 | 316 | Lower rhombic lip | L RhombLip | NEUR | VGLUT1 VGLUT2 | |
| DL-IT 152 | DLIT_152 | 152 | Deep-layer intratelencephalic | DL-IT | NEUR | VGLUT1 VGLUT2 VGLUT3 | |
| Thal-Exc 446 | Thex_446 | 446 | Thalamic excitatory | Thal-Exc | | VGLUT2 | |
| DL-NP 94 | DLNP_94 | 94 | Deep-layer near-projecting | DL-NP | NEUR | VGLUT1 | |
| LLC-IN 274 | LLC_274 | 274 | LAMP5-LHX6 and Chandelier | LLC-IN | NEUR | GABA | INT-LAMP5-LHX6 |
| Splatter 238 | Splat_238 | 238 | Splatter | Splatter | NEUR | GABA | |
| Thal-Exc 453 | Thex_453 | 453 | Thalamic excitatory | Thal-Exc | | VGLUT2 | |
| CGE-IN 285 | CGE_285 | 285 | CGE interneuron | CGE-IN | NEUR | GABA | INT-VIP |
| Splatter 338 | Splat_338 | 338 | Splatter | Splatter | NEUR | VGLUT2 | |
| L RhombLip 320 | LRL_320 | 320 | Lower rhombic lip | L RhombLip | NEUR | VGLUT1 VGLUT2 | |
| Splatter 360 | Splat_360 | 360 | Splatter | Splatter | NEUR | GABA | |
| Splatter 370 | Splat_370 | 370 | Splatter | Splatter | NEUR | VGLUT2 | |
| CGE-IN 281 | CGE_281 | 281 | CGE interneuron | CGE-IN | NEUR | GABA | INT-VIP |
| Splatter 431 | Splat_431 | 431 | Splatter | Splatter | NEUR | HDC VGLUT2 | |
| Thal-Exc 458 | Thex_458 | 458 | Thalamic excitatory | Thal-Exc | NEUR | VGLUT2 | |
| Thal-Exc 451 | Thex_451 | 451 | Thalamic excitatory | Thal-Exc | | VGLUT2 | |
| Thal-Exc 445 | Thex_445 | 445 | Thalamic excitatory | Thal-Exc | | VGLUT2 | |
| MiG 12 | Mgl_12 | 12 | Microglia | MiG | MAC | | |
| Splatter 399 | Splat_399 | 399 | Splatter | Splatter | NEUR | VGLUT2 | |
| COP 37 | COP_37 | 37 | Committed oligodendrocyte precursor | COP | OLIGO | | COP |
| Misc 0 | Bcell_0 | 0 | Miscellaneous | Misc | BCELL | | |
| Splatter 352 | Splat_352 | 352 | Splatter | Splatter | | GABA | |
| U RhombLip 312 | URL_312 | 312 | Upper rhombic lip | U RhombLip | NEUR | VGLUT1 | CB-GRAN |
| Splatter 377 | Splat_377 | 377 | Splatter | Splatter | NEUR | VGLUT2 | |
| Splatter 356 | Splat_356 | 356 | Splatter | Splatter | NEUR | GABA | |
| Splatter 357 | Splat_357 | 357 | Splatter | Splatter | NEUR | GABA | |
| DL-CT/6b 84 | DLCT6b_84 | 84 | Deep-layer corticothalamic and 6b | DL-CT/6b | NEUR | VGLUT1 VGLUT2 VGLUT3 | |
| LLC-IN 275 | LLC_275 | 275 | LAMP5-LHX6 and Chandelier | LLC-IN | NEUR | GABA | INT-LAMP5-LHX6 |

| Cluster (Yao naming) | Siletti cluster name | Siletti cluster ID | Supercluster | Supercluster abbreviation | Class auto-annotation | Neurotransmitter auto-annotation | Subtype auto-annotation |
|---|---|---|---|---|---|---|---|
| Fibr 24 | Fbl_24 | 24 | Fibroblast | Fibr | FIB | | PIA PVF |
| CGE-IN 280 | CGE_280 | 280 | CGE interneuron | CGE-IN | NEUR | GABA | INT-VIP |
| Splatter 384 | Splat_384 | 384 | Splatter | Splatter | NEUR | VGLUT2 | |
| CGE-IN 291 | CGE_291 | 291 | CGE interneuron | CGE-IN | NEUR | GABA | INT-VIP |
| HiDG 204 | DG_204 | 204 | Hippocampal dentate gyrus | HiDG | NEUR | VGLUT1 | DG-GRAN |
| Thal-Exc 455 | Thex_455 | 455 | Thalamic excitatory | Thal-Exc | NEUR | VGLUT2 | |
| Splatter 347 | Splat_347 | 347 | Splatter | Splatter | NEUR | GLY | |
| OPC 36 | OPC_36 | 36 | Oligodendrocyte precursor | OPC | OPC | | |
| Splatter 381 | Splat_381 | 381 | Splatter | Splatter | NEUR | GABA | |
| Splatter 411 | Splat_411 | 411 | Splatter | Splatter | NEUR | VGLUT2 | |
| Splatter 362 | Splat_362 | 362 | Splatter | Splatter | NEUR | GABA | |
| Splatter 409 | Splat_409 | 409 | Splatter | Splatter | NEUR | GABA | |
| Splatter 361 | Splat_361 | 361 | Splatter | Splatter | NEUR | GABA | |
| MiG 8 | Mgl_8 | 8 | Microglia | MiG | MGL | | |
| Splatter 350 | Splat_350 | 350 | Splatter | Splatter | NEUR | GLY | |
| Fibr 26 | Fbl_26 | 26 | Fibroblast | Fibr | FIB | | PIA |
| MiG 4 | Mgl_4 | 4 | Microglia | MiG | MGL | | |
| Splatter 414 | Splat_414 | 414 | Splatter | Splatter | NEUR | VGLUT2 | |
| Epend 74 | Epen_74 | 74 | Ependymal | Epend | CHRP EPEN | | |
| MiG 10 | Mgl_10 | 10 | Microglia | MiG | MGL | | |
| Misc 2 | Nkcell_2 | 2 | Miscellaneous | Misc | NK | | |
| Vasc 13 | VendPLVAP_13 | 13 | Vascular | Vasc | CHRP ENDO | | |
| Splatter 380 | Splat_380 | 380 | Splatter | Splatter | NEUR | GLY | |
| Cer-In 298 | CBI_298 | 298 | Cerebellar inhibitory | Cer-In | NEUR | GLY | CB-PurkLayerInt |
| Epend 68 | Epen_68 | 68 | Ependymal | Epend | CHRP EPEN | | |
| Splatter 335 | Splat_335 | 335 | Splatter | Splatter | NEUR | VGLUT2 | |
| Vasc 14 | VendAC_14 | 14 | Vascular | Vasc | ENDO | | ARTERIOLE CAP |
| Vasc 18 | VendA_18 | 18 | Vascular | Vasc | ENDO | | ARTERIOLE ARTERY |
| Splatter 342 | Splat_342 | 342 | Splatter | Splatter | NEUR | VGLUT2 | |
| Splatter 416 | Splat_416 | 416 | Splatter | Splatter | NEUR | GABA | |
| Misc 1 | Tcell_1 | 1 | Miscellaneous | Misc | TCELL | | |
| COP 38 | COP_38 | 38 | Committed oligodendrocyte precursor | COP | OLIGO | | COP |
| Epend 71 | Epen_71 | 71 | Ependymal | Epend | EPEN | | |
| CA4 194 | CA4_194 | 194 | Hippocampal CA4 | CA4 | NEUR | VGLUT1 | |
| MiG 7 | Mgl_7 | 7 | Microglia | MiG | MGL | | |
| Misc 401 | Misc_401 | 401 | Miscellaneous | Misc | NEUR | VGLUT2 | |
| Epend 67 | Epen_67 | 67 | Ependymal | Epend | CHRP EPEN | | |
| MiG 5 | Mgl_5 | 5 | Microglia | MiG | MGL | | |
| Splatter 339 | Splat_339 | 339 | Splatter | Splatter | NEUR | GLY VGLUT2 | |
| Misc 314 | Misc_314 | 314 | Miscellaneous | Misc | NEUR | GABA | CB-PURK |
| MiG 6 | Mgl_6 | 6 | Microglia | MiG | MGL | | |
| Splatter 344 | Splat_344 | 344 | Splatter | Splatter | CHRP NEUR | VGLUT2 | |
| COP 39 | COP_39 | 39 | Committed oligodendrocyte precursor | COP | OLIGO | | COP |
| Vasc 16 | VendVC_16 | 16 | Vascular | Vasc | ENDO | | CAP |
| Vasc 15 | VendC_15 | 15 | Vascular | Vasc | ENDO | | CAP |
| COP 42 | COP_42 | 42 | Committed oligodendrocyte precursor | COP | OLIGO | | |
| Splatter 397 | Splat_397 | 397 | Splatter | Splatter | NEUR | SER VGLUT3 | |

| Cluster (Yao naming) | Siletti cluster name | Siletti cluster ID | Supercluster | Supercluster abbreviation | Class auto-annotation | Neurotransmitter auto-annotation | Subtype auto-annotation |
|---|---|---|---|---|---|---|---|
| MiG 9 | Mgl_9 | 9 | Microglia | MiG | MGL | | |
| Splatter 353 | Splat_353 | 353 | Splatter | Splatter | NEUR | GABA | |
| Splatter 398 | Splat_398 | 398 | Splatter | Splatter | NEUR | VGLUT2 | |
| Olig 40 | Oligo_40 | 40 | Oligodendrocyte | Olig | OLIGO | | |
| Olig 46 | Oligo_46 | 46 | Oligodendrocyte | Olig | OLIGO | | |
| Splatter 379 | Splat_379 | 379 | Splatter | Splatter | NEUR | GLY | |
| Vasc 20 | Vsmc_20 | 20 | Vascular | Vasc | VSMC | | |
| CPlx 78 | Chrp_78 | 78 | Choroid plexus | CPlx | CHRP | | |
| Epend 65 | Epen_65 | 65 | Ependymal | Epend | EPEN | | |
| Epend 70 | Epen_70 | 70 | Ependymal | Epend | EPEN | | |
| CPlx 80 | Chrp_80 | 80 | Choroid plexus | CPlx | CHRP | | |
| Vasc 23 | Per_23 | 23 | Vascular | Vasc | | | |
| Epend 72 | Epen_72 | 72 | Ependymal | Epend | EPEN | | |
| CPlx 82 | Chrp_82 | 82 | Choroid plexus | CPlx | CHRP | | |
| Splatter 337 | Splat_337 | 337 | Splatter | Splatter | NEUR | GABA | |
| Splatter 351 | Splat_351 | 351 | Splatter | Splatter | NEUR | GABA | |
| Olig 48 | Oligo_48 | 48 | Oligodendrocyte | Olig | OLIGO | | |
| Epend 66 | Epen_66 | 66 | Ependymal | Epend | CHRP EPEN | | |
| Epend 73 | Epen_73 | 73 | Ependymal | Epend | EPEN | | |
| Olig 47 | Oligo_47 | 47 | Oligodendrocyte | Olig | OLIGO | | |
| CPlx 77 | Chrp_77 | 77 | Choroid plexus | CPlx | CHRP | | |
| COP 41 | COP_41 | 41 | Committed oligodendrocyte precursor | COP | OLIGO | | |
| Fibr 29 | Fbl_29 | 29 | Fibroblast | Fibr | | | |
| Fibr 28 | Fbl_28 | 28 | Fibroblast | Fibr | FIB | | PIA |
| CPlx 79 | Chrp_79 | 79 | Choroid plexus | CPlx | CHRP | | |
| COP 75 | COP_75 | 75 | Committed oligodendrocyte precursor | COP | CHRP EPEN NEUR | | |
| Splatter 390 | Splat_390 | 390 | Splatter | Splatter | NEUR | GABA | |
| Vasc 19 | Vsmc_19 | 19 | Vascular | Vasc | ENDO VSMC | | FIBMYO |
| Vasc 17 | VendV_17 | 17 | Vascular | Vasc | ENDO | | VENOUS |
| CPlx 81 | Chrp_81 | 81 | Choroid plexus | CPlx | CHRP | | |
| Fibr 27 | Fbl_27 | 27 | Fibroblast | Fibr | FIB | | PIA |
| Epend 69 | Epen_69 | 69 | Ependymal | Epend | EPEN | | |
| Olig 49 | Oligo_49 | 49 | Oligodendrocyte | Olig | OLIGO | | |
| Fibr 25 | Fbl_25 | 25 | Fibroblast | Fibr | FIB | | PIA |
| Olig 45 | Oligo_45 | 45 | Oligodendrocyte | Olig | OLIGO | | |
| COP 43 | COP_43 | 43 | Committed oligodendrocyte precursor | COP | OLIGO | | |
| Splatter 336 | Splat_336 | 336 | Splatter | Splatter | NEUR | VGLUT2 VGLUT3 | |
| CPlx 76 | Chrp_76 | 76 | Choroid plexus | CPlx | CHRP EPEN OLIGO | | |
| Vasc 21 | Per_21 | 21 | Vascular | Vasc | PER | | |
| Vasc 22 | Per_22 | 22 | Vascular | Vasc | PER | | |
| Olig 44 | Oligo_44 | 44 | Oligodendrocyte | Olig | OLIGO | | |
| Olig 50 | Oligo_50 | 50 | Oligodendrocyte | Olig | OLIGO | | |

| Cluster (Yao naming) | Top three regions | Top three dissections | Top Enriched Genes | Number of cells | Number of donors |
|---|---|---|---|---|---|
| Amyg-Exc 161 | Amygdala: 66.2%, Cerebral cortex: 31.6%, Hippocampus: 1.3% | Human CMN: 57.7%, Human Pir: 21.2%, Human A35-A36: 10.4% | MEPE, P2RX2, OTOF, MOXD1, CARM1P1, TG, S100Z, MS4A8, LINC02338, CNGB1 | 7442 | 3 |
| Misc 164 | Cerebral cortex: 66.8%, Amygdala: 18.2%, Hippocampus: 15.0% | Human LEC: 42.3%, Human Pir: 23.8%, Human Sub: 15.0% | TMEM45B, LINC02752, LINC02105, BARX2, AC005482.1, DSC3, COL6A2, LINC01482, LINC02653, HMCN2 | 659 | 3 |
| Amyg-Exc 156 | Amygdala: 78.4%, Hippocampus: 19.2%, Cerebral cortex: 2.3% | Human CMN: 38.0%, Human BL: 24.7%, Human CA1U-CA2U-CA3U: 16.5% | CAPN8, TACR3, TG, AC099565.1, CERS3-AS1, TRHR, AL392086.3, AL136985.3, AL136441.1, DSP | 5433 | 4 |
| HiDG 203 | Hippocampus: 98.3%, Midbrain: 1.4%, Cerebral cortex: 0.2% | Human DGU-CA4Upy: 42.8%, Human DGR-CA4: 13.9%, Human DGR-CA4Rpy: 13.6% | AC097528.1, LINC02248, AC097487.1, AC092110.1, AC079772.1, AC009878.2, COL6A3, AL445985.1, GRP, AL161629.1 | 8378 | 4 |
| MGE-IN 259 | Cerebral cortex: 89.3%, Amygdala: 3.4%, Hippocampus: 3.3% | Human MTG: 6.4%, Human M1C: 6.3%, Human A43: 4.0% | RGS5, AC022905.1, MYO5B, AL136114.1, TCIM, KCNS3, AC011586.2, MAFB, LHX6, CRHBP | 17841 | 4 |
| Amyg-Exc 155 | Amygdala: 99.6%, Cerebral cortex: 0.2%, Hippocampus: 0.2% | Human La: 77.0%, Human BL: 17.2%, Human CEN: 5.2% | AC005909.2, LINC02770, AL445207.1, LINC01903, AC099517.1, AC022655.1, DCSTAMP, AC068112.1, LINC01821, AC115100.1 | 11716 | 4 |
| Amyg-Exc 153 | Amygdala: 93.4%, Hippocampus: 5.3%, Cerebral cortex: 1.3% | Human BL: 77.7%, Human La: 9.7%, Human CA1U-CA2U-CA3U: 5.3% | LINC02770, DCSTAMP, AC005909.2, AC068112.1, LINC01903, AL445207.1, AC022655.1, TG, PPP1R1C, LINC02073 | 2227 | 3 |
| Amyg-Exc 159 | Amygdala: 80.8%, Cerebral cortex: 17.6%, Hippocampus: 1.3% | Human CMN: 32.2%, Human BM: 26.7%, Human CEN: 17.3% | POSTN, AC018742.1, LINC02653, AL583785.1, DCSTAMP, PWWP3B, NPFFR2, AL139340.1, KCNG1, AC068633.1 | 2148 | 4 |
| MGE-IN 236 | Hippocampus: 61.2%, Cerebral cortex: 21.0%, Amygdala: 16.5% | Human CA1C-CA3C: 17.3%, Human CA1U-CA2U-CA3U: 11.6%, Human CA1R-CA2R-CA3R: 8.6% | LHX6, MYO5B, CRHBP, PRTG, AC022905.1, NGF, TRG-AS1, RGS5, TAC1, RASGRF2-AS1 | 4303 | 4 |
| Misc 168 | Cerebral cortex: 92.4%, Hippocampus: 7.6%, Amygdala: 0.0% | Human TH-TL: 56.3%, Human MEC: 34.4%, Human Sub: 4.4% | GRB14, DUXAP9, SCHLAP1, MET, TWIST2, LINC01297, LINC01524, DUXAP10, FER1L6-AS2, DCN | 631 | 3 |
| MB-In 439 | Midbrain: 52.8%, Thalamus: 31.3%, Pons: 10.7% | Human SC: 11.2%, Human PAG: 10.3%, Human PTR: 9.4% | DMBX1, OTX2-AS1, OTX2, LINC01210, GATA3, AC008163.1, AL161757.4, LINC02055, SOX14, PRSS12 | 21004 | 4 |
| MGE-IN 260 | Cerebral cortex: 86.4%, Amygdala: 4.6%, Basal forebrain: 4.2% | Human MEC: 9.0%, Human FI: 6.2%, Human ACC: 5.8% | OSTN, IQGAP2, TCIM, AC022905.1, C1QL1, ACADL, WNT16, LHX6, CRHBP, MYO5B | 6212 | 4 |
| MB-In 440 | Thalamus: 87.1%, Midbrain: 10.2%, Pons: 1.9% | Human ETH: 15.2%, Human Pul: 11.7%, Human MD: 10.5% | AC008163.1, OTX2-AS1, DMBX1, AL161757.4, OTX2, AL137100.3, LINC01210, CASR, GATA3, SOX14 | 7366 | 3 |
| MB-In 438 | Midbrain: 44.9%, Thalamus: 38.0%, Hypothalamus: 8.8% | Human SN-RN: 11.6%, Human CM-Pf: 8.9%, Human PAG: 7.1% | AC008163.1, AL161757.4, LINC01210, OTX2, OTX2-AS1, DMBX1, CASR, AL137100.3, SOX14, GATA3 | 11928 | 3 |
| DL-IT 144 | Cerebral cortex: 96.2%, Basal forebrain: 1.4%, Amygdala: 1.2% | Human MTG: 13.0%, Human M1C: 10.4%, Human A43: 5.0% | AC021134.1, AL033539.2, TRABD2A, AC079052.1, AL138927.1, LINC01680, AL158835.1, AC073091.1, AL445207.1, Z68323.1 | 27079 | 4 |
| LLC-IN 265 | Cerebral cortex: 72.8%, Amygdala: 17.1%, Hippocampus: 5.1% | Human MEC: 6.1%, Human M1C: 4.8%, Human BL: 4.2% | AC046195.2, LINC00943, PCDH18, COL15A1, PTHLH, C1QL1, LINC01837, NOG, CA8, PLEKHH2 | 4251 | 4 |
| Amyg-Exc 173 | Hippocampus: 55.8%, Amygdala: 36.5%, Cerebral cortex: 7.7% | Human Sub: 55.8%, Human BM: 30.5%, Human CMN: 4.7% | AC116345.1, AC016766.1, MCOLN2, AC018742.1, S100Z, WDR63, LINC00578, LINC02296, ALKAL2, LINC02015 | 233 | 3 |
| UL-IT 120 | Cerebral cortex: 91.5%, Hypothalamus: 4.6%, Basal forebrain: 2.9% | Human FI: 14.3%, Human M1C: 13.2%, Human ACC: 12.6% | GLP2R, LINC02248, LINC02306, AC009878.2, LINC01622, CBLN2, SLCO2A1, TTC6, LINC01378, AL132633.1 | 21417 | 4 |
| UL-IT 124 | Cerebral cortex: 98.9%, Basal forebrain: 0.5%, Hippocampus: 0.2% | Human MTG: 12.5%, Human A44-A45: 10.1%, Human A13: 9.0% | LINC00507, LINC00508, AC097512.1, AC110614.1, LINC02263, LINC02306, LINC00326, AC092440.1, AC093607.1, AC110014.1 | 28224 | 4 |
| UL-IT 126 | Cerebral cortex: 90.8%, Basal forebrain: 3.3%, Hypothalamus: 2.9% | Human FI: 8.2%, Human ACC: 7.9%, Human A25: 7.1% | LINC01090, ADAMTS9-AS1, AC117453.1, AJ009632.2, ADGRF5, AC110014.1, LINC00390, LINC02301, AC060765.1, LINC01478 | 36970 | 4 |
| Misc 404 | Cerebral cortex: 94.4%, Amygdala: 2.5%, Basal forebrain: 1.2% | Human A14: 60.0%, Human M1C: 11.2%, Human A43: 3.2% | AC016687.2, AC021134.1, LINC02378, LINC02263, LINC01915, CXCL14, AL033539.2, AC004862.1, AC073578.2, LINC02306 | 1276 | 4 |
| Misc 170 | Hippocampus: 68.6%, Hypothalamus: 21.7%, Cerebral cortex: 6.4% | Human CA1: 57.7%, Human HTHtub: 21.5%, Human CA1U-CA2U-CA3U: 9.3% | LINC02752, POSTN, DCN, ITPRID1, LINC02309, LINC02316, AC124290.1, GRB14, IFNG-AS1, MAFA | 503 | 3 |
| MSN 216 | Basal forebrain: 40.9%, Amygdala: 26.3%, Hypothalamus: 18.3% | Human SI: 36.9%, Human BNST: 19.4%, Human HTHpo: 16.1% | DRD2, MYO3A, PENK, ADORA2A, LINC01833, GPR6, LINC02462, AL020994.2, P2RY1, SIX3 | 5720 | 3 |

| Cluster (Yao naming) | Top three regions | Top three dissections | Top Enriched Genes | Number of cells | Number of donors |
|---|---|---|---|---|---|
| MGE-IN 242 | Cerebral cortex: 90.8%, Hippocampus: 4.6%, Basal forebrain: 1.8% | Human M1C: 6.8%, Human MTG: 6.4%, Human A32: 5.4% | AC022905.1, AC113347.4, MCUB, ELFN1, DCN, FBN2, SLC27A6, TNFSF15, SST, LINC02397 | 15796 | 4 |
| Amyg-Exc 405 | Cerebral cortex: 41.9%, Hypothalamus: 27.8%, Amygdala: 27.3% | Human AON: 31.1%, Human HTHso: 26.6%, Human CoA: 12.2% | ROR2, TACR3, AC022523.1, AC020704.1, WDR86, AC025470.2, AL136119.1, NFIA-AS2, RXFP2, AC087516.2 | 6125 | 4 |
| DL-IT 136 | Amygdala: 54.6%, Cerebral cortex: 36.2%, Hypothalamus: 8.6% | Human CoA: 54.6%, Human AON: 28.3%, Human HTHso: 8.6% | AC099517.1, LINC02196, AC079380.1, LINC02465, AL138927.1, AC073578.2, AL450352.1, ARHGAP15, TNNT2, LINC02378 | 152 | 4 |
| eMSN 234 | Amygdala: 75.9%, Cerebral cortex: 14.6%, Thalamus: 5.4% | Human BM: 29.1%, Human CEN: 16.6%, Human CMN: 14.3% | NPFFR2, ZNF736P9Y, GABRQ, LMNTD1, EYA2, AC012078.2, AC087516.2, PCDH11Y, LINC00354, NMBR | 3211 | 3 |
| MGE-IN 261 | Cerebral cortex: 95.5%, Hippocampus: 1.4%, Basal forebrain: 1.4% | Human M1C: 13.2%, Human MTG: 6.5%, Human A29-A30: 4.8% | TCIM, RNF144B, LRRC38, ADAMTS17, PRDM1, KCNS3, MYO5B, RGS5, OSTN, LHX6 | 21457 | 4 |
| MSN 214 | Basal forebrain: 61.9%, Amygdala: 15.7%, Cerebral cortex: 13.3% | Human SI: 37.9%, Human NAC: 15.7%, Human BNST: 14.8% | DRD3, AC010478.1, PDYN, AL020994.2, GPR101, AL359220.1, AC092604.1, MYO3A, DRD1, C6orf141 | 14016 | 3 |
| LLC-IN 266 | Amygdala: 42.8%, Hippocampus: 27.1%, Cerebral cortex: 25.1% | Human CMN: 12.5%, Human BL: 8.8%, Human La: 8.4% | IL18R1, C1QL1, COL15A1, IBSP, LINC01837, CPED1, LINC00943, GRIN2C, LHX6, PTHLH | 3566 | 4 |
| eMSN 228 | Amygdala: 83.7%, Cerebral cortex: 12.2%, Thalamus: 1.9% | Human La: 25.3%, Human BM: 17.5%, Human CEN: 16.1% | NPFFR2, AC087516.2, AC083837.1, AC008127.1, AL365255.1, AC097459.1, AC092590.1, EYA2, DBX2, SCN5A | 3227 | 3 |
| MGE-IN 253 | Cerebral cortex: 81.1%, Amygdala: 8.4%, Basal forebrain: 6.1% | Human M1C: 8.3%, Human FI: 6.3%, Human MTG: 5.3% | LINC00943, SCARA5, AC092353.2, AL450345.2, SST, LHX6, PAWR, TH, TLL2, NMU | 5703 | 4 |
| MSN 208 | Basal forebrain: 89.6%, Amygdala: 5.6%, Cerebral cortex: 3.5% | Human CaB: 31.1%, Human NAC: 28.8%, Human GPe: 12.5% | PENK, GPR6, ADORA2A, DRD2, LINC02137, AC004551.1, SMOC2, AC107208.1, LPL, EYA1 | 13170 | 3 |
| Misc 166 | Cerebral cortex: 79.4%, Hippocampus: 16.1%, Amygdala: 3.1% | Human MEC: 71.3%, Human Sub: 14.9%, Human LEC: 7.8% | LINC02752, IFNG-AS1, SNHG31, LINC01297, BARX2, AC072062.1, AC104689.2, TMEM45B, AC116345.1, AL589693.1 | 944 | 3 |
| CA1-3 3 | Thalamus: 21.9%, Pons: 16.1%, Basal forebrain: 13.9% | Human PnEN: 8.9%, Human MoAN: 6.6%, Human PnRF: 3.8% | FCN1, CLEC12A, S100A4, TNFRSF1B, IFI30, LYZ, CYTIP, FGR, LILRB2, KYNU | 503 | 3 |
| Amyg-Exc 172 | Cerebral cortex: 90.6%, Hippocampus: 8.8%, Amygdala: 0.6% | Human MEC: 81.6%, Human Sub: 8.8%, Human LEC: 8.6% | TGFBI, AC116345.1, AL391117.1, S100Z, AOX1, CRHR2, LINC01886, FBXL21P, AC092162.2, PLEKHG4B | 8592 | 4 |
| MB-In 436 | Thalamus: 82.2%, Midbrain: 17.5%, Hypothalamus: 0.1% | Human LG: 28.3%, Human Pul: 26.3%, Human SC: 7.5% | HMCN1, LINC01210, OTX2, SOX14, DMBX1, AC246817.2, OTX2-AS1, AC245123.1, AL133493.1, AC007159.1 | 13890 | 3 |
| Amyg-Exc 157 | Amygdala: 65.8%, Cerebral cortex: 28.7%, Hippocampus: 5.5% | Human BL: 31.3%, Human BM: 23.4%, Human Pir: 14.2% | TG, CERS3-AS1, AC099565.1, CAPN8, AL136441.1, SLA, MOXD1, DSP, AL136985.3, SEMA3D | 14855 | 4 |
| UL-IT 127 | Cerebral cortex: 90.2%, Basal forebrain: 4.2%, Amygdala: 3.4% | Human A38: 10.6%, Human ACC: 9.3%, Human FI: 9.0% | ADGRF5, AC018697.1, CARM1P1, AC124290.1, AL162493.1, AJ009632.2, AC124290.2, AC092848.2, AC117453.1, AC115100.1 | 10430 | 4 |
| Amyg-Exc 158 | Amygdala: 68.1%, Cerebral cortex: 25.8%, Hippocampus: 6.0% | Human CMN: 38.7%, Human BM: 16.0%, Human CoA: 12.6% | TACR3, TG, AC002480.2, SEMA3D, ALKAL2, TRHR, NPFFR2, LINC01865, GPC4, WDR86 | 6409 | 3 |
| MB-In 443 | Thalamus: 98.3%, Midbrain: 1.6%, Pons: 0.1% | Human Pul: 26.6%, Human MD: 14.7%, Human LP: 13.7% | CASR, LINC01210, OTX2-AS1, OTX2, AL133493.1, AL161757.4, AL137100.3, NPPC, DMBX1, SOX14 | 14414 | 3 |
| MSN 212 | Basal forebrain: 90.4%, Cerebral cortex: 3.6%, Amygdala: 3.6% | Human CaB: 25.4%, Human Pu: 22.3%, Human NAC: 18.4% | PENK, AL139379.1, GPR6, DRD2, AL450332.1, ADORA2A, AC107208.1, GPR88, AC092979.1, LINC02137 | 6867 | 3 |
| MSN 211 | Basal forebrain: 85.7%, Amygdala: 6.3%, Cerebral cortex: 4.2% | Human CaB: 26.7%, Human NAC: 18.8%, Human Pu: 17.8% | AL445584.2, AL450332.1, LINC02137, TH, CA12, AC138646.1, PDYN, AC107208.1, AL139379.1, MME | 9093 | 3 |
| eMSN 231 | Amygdala: 60.2%, Basal forebrain: 16.4%, Cerebral cortex: 13.7% | Human CMN: 15.9%, Human CEN: 12.8%, Human SI: 11.6% | LMNTD1, NPFFR2, AC012078.2, ZNF736P9Y, GABRQ, EYA2, PCDH11Y, CAPSL, DRAXIN, ITGA6 | 2372 | 3 |
| CA4 196 | Hippocampus: 99.9%, Midbrain: 0.1%, Cerebral cortex: 0.0% | Human CA4C-DGC: 55.3%, Human DGR-CA4Rpy: 39.4%, Human DGR-CA4: 3.0% | MORC1, ABCA13, LINC02196, GUCA1C, ITGA4, LATS2, TSPAN18, MYPN, AC021594.2, AC127526.2 | 1360 | 3 |
| Misc 165 | Cerebral cortex: 92.1%, Hippocampus: 5.0%, Amygdala: 2.8% | Human MEC: 72.0%, Human LEC: 18.2%, Human Sub: 5.0% | LINC02752, BARX2, IFNG-AS1, DSC3, AC116345.1, SNHG31, AC104689.2, AC006974.2, TMEM45B, AC072062.1 | 2064 | 3 |
| DL-IT 146 | Amygdala: 65.4%, Cerebral cortex: 33.1%, Hypothalamus: 0.8% | Human Pir: 22.2%, Human CEN: 19.8%, Human BL: 17.2% | CCN4, STPG2-AS1, AC105450.1, LINC00683, ATP10A, SNTG2, UCMA, TMEM244, FGF10, LINC02008 | 5654 | 4 |

| Cluster (Yao naming) | Top three regions | Top three dissections | Top Enriched Genes | Number of cells | Number of donors |
|---|---|---|---|---|---|
| Amyg-Exc 175 | Cerebral cortex: 91.3%, Hippocampus: 7.3%, Amygdala: 1.2% | Human MEC: 82.6%, Human LEC: 8.2%, Human Sub: 6.4% | AL035446.2, LINC00383, AC009081.2, AL138694.1, AC116345.1, SMIM36, FBLN2, AC113137.1, AC009878.2, CARM1P1 | 8112 | 4 |
| MGE-IN 243 | Cerebral cortex: 96.3%, Hippocampus: 1.4%, Basal forebrain: 1.1% | Human MTG: 8.2%, Human A40: 5.9%, Human M1C: 4.5% | LINC01687, AC011586.2, AC113347.4, MSR1, HPGD, AC011586.1, CMTM8, SST, DCN, AOX1 | 16126 | 4 |
| MB-In 437 | Thalamus: 85.5%, Midbrain: 13.8%, Pons: 0.5% | Human Pul: 19.7%, Human ETH: 12.5%, Human MD: 11.9% | AC245123.1, OTX2-AS1, LINC01210, OTX2, AC245187.2, AC246817.2, DMBX1, AL133493.1, GATA3, CASR | 6147 | 3 |
| Amyg-Exc 154 | Amygdala: 87.5%, Cerebral cortex: 6.8%, Hippocampus: 5.7% | Human BL: 55.4%, Human La: 20.5%, Human BM: 8.2% | DCSTAMP, AL445207.1, PTGER3, AC005909.2, LINC01821, SMIM36, AC099517.1, AC018742.1, AC022655.1, AC004704.1 | 7970 | 4 |
| CA1-3 3 | Thalamus: 21.9%, Pons: 16.1%, Basal forebrain: 13.9% | Human PnEN: 8.9%, Human MoAN: 6.6%, Human PnRF: 3.8% | FCN1, CLEC12A, S100A4, TNFRSF1B, IFI30, LYZ, CYTIP, FGR, LILRB2, KYNU | 503 | 3 |
| Splatter 420 | Thalamus: 46.3%, Hypothalamus: 26.8%, Amygdala: 24.7% | Human ANC: 45.1%, Human BNST: 19.3%, Human HTHpo-HTHso: 17.0% | OTP, SIM1, ARHGAP36, GABRQ, IGSF1, FEZF1, FEZF1-AS1, AC012078.2, CALCR, AL591463.1 | 922 | 3 |
| MGE-IN 249 | Cerebral cortex: 61.8%, Hippocampus: 23.3%, Amygdala: 10.2% | Human MEC: 10.2%, Human LEC: 7.8%, Human CA1C-CA3C: 6.1% | SCARA5, SST, ELFN1, DCN, ITGA1, PAWR, FAM89A, TNFSF15, MCUB, CRHBP | 5821 | 4 |
| DL-IT 143 | Cerebral cortex: 89.0%, Basal forebrain: 4.2%, Amygdala: 4.2% | Human MEC: 14.2%, Human ACC: 10.5%, Human A38: 9.9% | AC021134.1, Z68323.1, AL033539.2, AC079052.1, AC021134.2, LINC01821, TRABD2A, LINC01606, ADTRP, LINC01320 | 13468 | 4 |
| eMSN 226 | Basal forebrain: 33.6%, Amygdala: 33.3%, Hypothalamus: 19.3% | Human SI: 25.6%, Human BNST: 15.8%, Human HTHpo: 14.4% | NPFFR2, OTOF, GDNF-AS1, PDYN, ZNF736P9Y, DRD1, LINC01550, FBXL21P, AC034226.1, GFRA1 | 5927 | 3 |
| MB-In 435 | Midbrain: 54.9%, Thalamus: 43.2%, Pons: 0.8% | Human SN-RN: 24.2%, Human CM-Pf: 13.1%, Human PAG: 7.4% | NPSR1, LINC01210, CASR, SOX14, OTX2, OTX2-AS1, AC007159.1, DMBX1, GATA3, AL161757.4 | 7340 | 3 |
| MGE-IN 257 | Cerebral cortex: 73.5%, Hippocampus: 19.7%, Amygdala: 3.5% | Human M1C: 9.5%, Human MTG: 5.2%, Human CA1U-CA2U-CA3U: 4.6% | MEPE, WNT16, MTND6P22, RGS5, HGF, RNF144B, AC006974.2, AC006364.1, LHX6, CPED1 | 12242 | 4 |
| MB-In 441 | Thalamus: 89.9%, Midbrain: 8.8%, Pons: 0.7% | Human Pul: 15.3%, Human MD: 13.6%, Human ETH: 13.0% | DMBX1, OTX2-AS1, AC008163.1, AL161757.4, OTX2, TSPAN12, AL137100.3, LINC01210, CASR, MYO3B | 22931 | 3 |
| eMSN 225 | Amygdala: 47.2%, Basal forebrain: 33.3%, Cerebral cortex: 10.2% | Human SI: 15.2%, Human BNST: 14.1%, Human La: 12.0% | AC008786.1, ZNF736P9Y, LMNTD1, CASZ1, OTOF, S100Z, AC068633.1, DNAH8, CALCRL, MUC19 | 2296 | 3 |
| MGE-IN 263 | Cerebral cortex: 98.5%, Basal forebrain: 0.6%, Hippocampus: 0.4% | Human M1C: 18.6%, Human A29-A30: 10.6%, Human V1C: 9.3% | LRRC38, OSTN, SULF1, TCIM, PVALB, ADAMTS17, RNF144B, AC118942.1, PRDM1, AL096865.1 | 6587 | 4 |
| UL-IT 122 | Cerebral cortex: 93.9%, Basal forebrain: 2.6%, Hippocampus: 1.7% | Human MTG: 13.0%, Human Ig: 10.2%, Human TF: 8.4% | LINC01500, CASP4LP, LINC01484, AC097512.1, PDGFD, ART3, MUC19, AC093607.1, AC021613.1, AL353604.1 | 29434 | 4 |
| MB-In 442 | Thalamus: 99.1%, Midbrain: 0.8%, Pons: 0.1% | Human Pul: 22.3%, Human MD: 18.2%, Human LP: 13.8% | AL137100.3, CASR, AL161757.4, OTX2-AS1, OTX2, DMBX1, TSPAN12, LINC01210, NPSR1, VASH2 | 7951 | 3 |
| MSN 221 | Basal forebrain: 78.8%, Amygdala: 9.0%, Cerebral cortex: 6.5% | Human SI: 19.8%, Human Pu: 16.4%, Human NAC: 15.6% | AC048352.1, GPR6, LINC01305, DRD2, AC005482.1, RDH10, SIX3, LOXHD1, AC023905.1, AL392086.1 | 2571 | 3 |
| eMSN 222 | Basal forebrain: 93.9%, Thalamus: 1.9%, Cerebral cortex: 1.8% | Human Pu: 38.7%, Human CaB: 28.3%, Human NAC: 12.1% | AL445584.2, CASZ1, AC022133.2, LINC01735, TAC1, LOXHD1, C6orf141, PENK, FBXL21P, SLC35D3 | 2826 | 3 |
| DL-CT/6b 102 | Amygdala: 66.6%, Cerebral cortex: 28.9%, Hippocampus: 4.2% | Human La: 18.0%, Human BL: 17.0%, Human LEC: 16.4% | AC092131.1, LINC01194, CCN2, VWA2, AC064875.1, SLC15A5, HCP5, SNHG31, AC023669.2, AP003100.2 | 1920 | 4 |
| MGE-IN 241 | Cerebral cortex: 90.2%, Basal forebrain: 3.5%, Hippocampus: 2.5% | Human MTG: 6.3%, Human M1C: 5.3%, Human A43: 5.1% | AC011586.2, MSR1, AC022905.1, AC113347.4, MCOLN2, AC011586.1, CMTM8, MYO5B, AL355672.1, SLC27A6 | 5583 | 4 |
| CA4 197 | Hippocampus: 99.7%, Midbrain: 0.3%, Cerebral cortex: 0.0% | Human DGR-CA4Rpy: 50.3%, Human CA3R: 19.4%, Human DGU-CA4Upy: 14.3% | MORC1, LINC02196, GUCA1C, ABCA13, LATS2, TSPAN18, HLX-AS1, WNK4, AC019127.1, MYPN | 2367 | 3 |
| Amyg-Exc 174 | Cerebral cortex: 84.3%, Amygdala: 10.2%, Hippocampus: 5.2% | Human LEC: 47.8%, Human Pir: 27.5%, Human Sub: 5.2% | AC116345.1, P2RX2, AL031073.2, MS4A8, TMEM108-AS1, AL136441.1, AL136985.3, LINC02248, CARM1P1, SEMA3D | 10364 | 4 |
| MGE-IN 258 | Cerebral cortex: 80.1%, Hippocampus: 6.7%, Amygdala: 6.3% | Human M1C: 8.8%, Human MEC: 6.6%, Human FI: 5.5% | WNT16, LINC01035, HGF, LHX6, TCIM, AC006364.1, AC046195.2, MYO5B, CRHBP, ARX | 12943 | 4 |
| HiDG 200 | Hippocampus: 99.4%, Thalamus: 0.5%, Midbrain: 0.1% | Human CA4C-DGC: 59.1%, Human DGR-CA4Rpy: 18.1%, Human DGU-CA4Upy: 11.3% | LINC01697, AC097487.1, AJ006995.1, CHRNA1, TARID, AC097528.1, AC079793.1, NTF3, MCOLN3, COL6A3 | 23293 | 4 |
| Amyg-Exc 407 | Amygdala: 72.6%, Cerebral cortex: 23.5%, Hippocampus: 2.7% | Human CEN: 31.1%, Human CMN: 27.2%, Human A35-A36: 18.1% | TFAP2C, PAPPA2, CARTPT, BARHL2, LINC01798, MEIS1-AS3, AC006065.4, LHX9, GALR1, POU2F3 | 2370 | 4 |

| Cluster (Yao naming) | Top three regions | Top three dissections | Top Enriched Genes | Number of cells | Number of donors |
|---|---|---|---|---|---|
| Misc 178 | Hippocampus: 43.8%, Midbrain: 29.4%, Cerebral cortex: 26.8% | Human Sub: 37.9%, Human SC: 29.4%, Human TH-TL: 21.6% | AC008662.1, AC091096.1, AP003066.1, AL391117.1, COBLL1, AC099517.1, AC097528.1, LINC02143, AC008415.1, AC004052.1 | 691 | 3 |
| DL-IT 148 | Cerebral cortex: 85.2%, Basal forebrain: 6.0%, Amygdala: 5.0% | Human MTG: 7.7%, Human ACC: 7.4%, Human FI: 6.6% | LINC00343, AC011754.1, THEMIS, TMEM233, AL356432.3, AC092131.1, AC079163.2, AC004917.1, AC015943.1, SH2D1B | 16196 | 4 |
| CA1-3 3 | Thalamus: 21.9%, Pons: 16.1%, Basal forebrain: 13.9% | Human PnEN: 8.9%, Human MoAN: 6.6%, Human PnRF: 3.8% | FCN1, CLEC12A, S100A4, TNFRSF1B, IFI30, LYZ, CYTIP, FGR, LILRB2, KYNU | 503 | 3 |
| HiDG 202 | Hippocampus: 100.0%, Cerebral cortex: 0.0%, Amygdala: 0.0% | Human DGR-CA4Rpy: 87.7%, Human CA1R-CA2R-CA3R: 12.3% | TTR, FOLR1, LINC01697, AC097528.1, AC097487.1, AL672167.1, NTF3, AL591518.1, TARID, AJ006995.1 | 398 | 1 |
| LLC-IN 269 | Cerebral cortex: 43.7%, Hippocampus: 25.7%, Amygdala: 22.8% | Human MEC: 8.5%, Human CMN: 6.9%, Human CA1U-CA2U-CA3U: 6.2% | CHRDL1, C1QL1, ANO1, TMEM255A, ACADL, LINC01344, ISM1, CHST9, NGF, GRPR | 3214 | 4 |
| U RhombLip 308 | Cerebellum: 84.5%, Pons: 10.2%, Medulla: 3.6% | Human CBV: 77.8%, Human PnAN: 6.8%, Human CBL: 5.8% | AC017037.5, AC084740.1, EOMES, LMX1A, AL359208.1, CSRP2, AC092957.1, AC108517.2, MPP3, ITGBL1 | 920 | 4 |
| DL-NP 92 | Cerebral cortex: 94.7%, Hippocampus: 2.5%, Basal forebrain: 2.2% | Human MTG: 12.1%, Human Ig: 8.6%, Human A1C: 6.9% | CD200R1L, ERG, IFNG-AS1, NPSR1-AS1, BARX2, LINC02042, AC009081.2, AC129807.1, CD200R1L-AS1, CCDC80 | 2119 | 4 |
| MB-In 444 | Thalamus: 83.2%, Midbrain: 14.0%, Pons: 1.9% | Human LG: 57.2%, Human Pul: 4.7%, Human MD: 4.2% | OTX2-AS1, DMBX1, OTX2, CASR, NPPC, AL137100.3, AL161757.4, AL133493.1, LINC01210, LINC02253 | 3991 | 4 |
| Splatter 422 | Hypothalamus: 44.2%, Amygdala: 23.2%, Thalamus: 20.2% | Human BNST: 22.3%, Human ANC: 20.1%, Human HTHpo: 19.8% | CALCR, LHX8, TMEM255A, DGKK, AL110292.1, ARHGAP36, GABRE, ONECUT3, STK26, LINC00540 | 2976 | 3 |
| Astr 59 | Pons: 36.2%, Cerebellum: 36.1%, Medulla: 10.9% | Human CbDN: 36.1%, Human PnEN: 13.6%, Human PnRF: 13.0% | LGR6, SDC4, ITPRID1, FGFR3, SLC14A1, SLC7A10, AC090809.1, GJB6, GFAP, PYGM | 4062 | 4 |
| Splatter 386 | Medulla: 98.2%, Pons: 1.2%, Midbrain: 0.6% | Human MoRF-MoEN: 55.1%, Human MoSR: 38.9%, Human MoAN: 4.2% | TACR3, LINC01876, AC074099.1, HOXB3, HSPD1P8, POU4F1, PGR-AS1, PGR, LINC02742, HOXA3 | 167 | 3 |
| eMSN 233 | Amygdala: 73.7%, Thalamus: 9.5%, Hypothalamus: 5.8% | Human La: 23.4%, Human CEN: 17.3%, Human BNST: 12.8% | ZNF736P9Y, NPFFR2, GABRQ, NMBR, LMNTD1, PRSS35, FRMD7, PCDH11Y, CAPSL, LINC00354 | 3128 | 3 |
| UL-IT 121 | Cerebral cortex: 91.7%, Amygdala: 3.9%, Basal forebrain: 2.7% | Human A38: 13.5%, Human TH-TL: 11.4%, Human MTG: 9.6% | MUC19, LINC01331, GLP2R, AL136985.3, TTC6, AC009878.2, LINC02248, AC011287.2, LINC01622, AL353604.1 | 36308 | 4 |
| Splatter 428 | Basal forebrain: 59.7%, Amygdala: 16.1%, Hypothalamus: 13.8% | Human SEP: 55.1%, Human BNST: 12.4%, Human ANC: 5.7% | FREM2, AC092958.2, SCGN, AC092957.1, ZIC2, LHX8, PRDM12, PRDM16, ZIC5, TTR | 3182 | 4 |
| UL-IT 135 | Cerebral cortex: 95.4%, Basal forebrain: 2.7%, Hippocampus: 1.0% | Human M1C: 10.7%, Human A1C: 7.5%, Human MTG: 6.2% | LINC01915, COL22A1, AL353811.1, LINC01164, IZUMO3, AL157769.1, LINC01781, ITGA11, TWIST2, AC008459.1 | 28248 | 4 |
| Misc 176 | Hippocampus: 98.8%, Cerebral cortex: 1.0%, Thalamus: 0.2% | Human CA1U-CA2U-CA3U: 90.1%, Human CA1-2R: 4.3%, Human CA1: 2.7% | AC008662.1, AC097528.1, LINC01524, TMC1, AL391097.3, AC091096.1, AL391097.1, COBLL1, ABCA12, LINC02143 | 414 | 3 |
| Amyg-Exc 162 | Amygdala: 84.9%, Hippocampus: 8.9%, Cerebral cortex: 6.2% | Human BL: 29.2%, Human CMN: 27.0%, Human BM: 25.5% | AC018742.1, POSTN, MEPE, MCOLN3, P2RX2, LINC01885, LINC01606, AC009411.2, DCDC2C, SYTL1 | 807 | 3 |
| HiDG 201 | Hippocampus: 99.9%, Midbrain: 0.1%, Thalamus: 0.0% | Human DGU-CA4Upy: 34.3%, Human DGR-CA4: 34.3%, Human DGR-CA4Rpy: 14.6% | AC097487.1, AC097528.1, COL6A3, NTF3, LINC01697, ANKRD20A19P, AL672167.1, LINC01324, AC092110.1, AL445985.1 | 18996 | 3 |
| MSN 209 | Basal forebrain: 96.3%, Thalamus: 2.9%, Amygdala: 0.6% | Human CaB: 77.9%, Human Pu: 8.2%, Human NAC: 5.1% | AL450332.1, AL139379.1, LINC02137, AC004551.1, AC015631.1, LINC02462, AL445584.2, LPL, PLEK, CA12 | 11410 | 3 |
| MGE-IN 240 | Cerebral cortex: 92.3%, Amygdala: 3.0%, Hippocampus: 2.3% | Human M1C: 10.5%, Human MTG: 7.6%, Human A40: 5.0% | AC011586.2, AC022905.1, MSR1, IL18R1, PRDM1, MYO5B, RGS5, CDKN2B-AS1, CMTM8, CDCA7 | 1327 | 4 |
| CGE-IN 292 | Cerebral cortex: 86.1%, Hippocampus: 7.4%, Amygdala: 3.1% | Human MTG: 7.0%, Human M1C: 6.1%, Human A29-A30: 4.3% | NPR3, VIP, PPP1R1C, AC020930.1, CHRNA2, AC013265.1, SP8, CRH, AL391832.4, AC062021.1 | 12439 | 4 |
| Amyg-Exc 171 | Hippocampus: 93.7%, Amygdala: 5.7%, Cerebral cortex: 0.3% | Human CA1: 86.7%, Human Sub: 6.9%, Human CMN: 5.7% | SERPINB13, TGFBI, AOX1, AC116345.1, ABCA12, LINC01886, AC082650.2, AOX3P, AC068305.2, CRHR2 | 332 | 2 |
| Splatter 364 | Midbrain: 99.6%, Thalamus: 0.2%, Medulla: 0.1% | Human IC: 97.5%, Human SC: 1.7%, Human PTR: 0.2% | TFAP2D, LHX9, LINC01956, HNF4G, TMEM176B, LINC01568, AL138720.1, IRX2, WDR72, ZFHX4-AS1 | 7200 | 4 |
| Misc 177 | Hippocampus: 68.2%, Midbrain: 20.5%, Cerebral cortex: 10.8% | Human Sub: 39.5%, Human CA1-2R: 23.8%, Human SC: 20.4% | AC008662.1, AC097528.1, LINC01339, COBLL1, AC018680.1, VIPR2, AC091096.1, LINC01524, LINC02143, AL109930.1 | 3941 | 3 |

| Cluster (Yao naming) | Top three regions | Top three dissections | Top Enriched Genes | Number of cells | Number of donors |
|---|---|---|---|---|---|
| Mam 327 | Hypothalamus: 99.9%, Midbrain: 0.1%, Cerebral cortex: 0.0% | Human MN: 66.7%, Human HTHma: 27.8%, Human HTHma-HTHtub: 5.4% | LINC00410, PITX2, AC244258.1, AL691420.1, FOXB1, TNFRSF11B, LHX1-DT, LHX1, DRD3, MYO5B | 1581 | 3 |
| MGE-IN 255 | Cerebral cortex: 67.2%, Amygdala: 13.0%, Hippocampus: 12.1% | Human M1C: 5.6%, Human CEN: 5.0%, Human MTG: 4.3% | AC046195.2, WNT16, GLP1R, HGF, AL138899.1, MYBPC1, TRDN, LINC01885, AC046195.1, AC006364.1 | 3341 | 4 |
| Cer-In 302 | Cerebellum: 60.1%, Medulla: 19.0%, Pons: 15.3% | Human CBV: 42.3%, Human CBL: 12.9%, Human IO: 7.7% | CPLX4, AC010476.2, SLC6A5, PAX2, AC008725.1, AC068672.2, TFAP2B, LINC01179, AL139351.3, LGI2 | 248 | 3 |
| Astr 60 | Thalamus: 37.1%, Midbrain: 31.3%, Pons: 16.8% | Human SN-RN: 10.3%, Human SN: 9.3%, Human Pul: 8.4% | TNC, SLC14A1, GJA1, SDC4, OBI1-AS1, HPSE2, RASL12, AC012405.1, EFEMP1, EDNRB | 25366 | 4 |
| eMSN 229 | Amygdala: 56.1%, Basal forebrain: 28.8%, Cerebral cortex: 11.2% | Human CEN: 21.7%, Human CMN: 17.8%, Human Cla: 12.9% | NPFFR2, AC087516.2, CACNG5, EYA2, PDYN, ISM2, ZNF736P9Y, LINC00354, FOXP2, GPR88 | 1857 | 4 |
| Misc 132 | Cerebral cortex: 98.4%, Hippocampus: 0.8%, Midbrain: 0.8% | Human A29-A30: 90.9%, Human Pro: 5.1%, Human M1C: 1.2% | CD53, DPP4, AC060765.1, LINC01721, AC112770.1, AL157769.1, VIPR2, PCSK1, AC016687.2, QRFPR | 492 | 4 |
| DL-NP 95 | Cerebral cortex: 89.0%, Basal forebrain: 7.0%, Hypothalamus: 3.3% | Human A25: 21.2%, Human ACC: 13.9%, Human AON: 12.5% | NPSR1-AS1, AC009081.2, ERG, CCDC80, HMGA2, MYOCD, PDLIM1, CD200R1L, FSHR, GNAT3 | 1307 | 4 |
| UL-IT 123 | Cerebral cortex: 99.2%, Hippocampus: 0.4%, Basal forebrain: 0.3% | Human A5-A7: 8.0%, Human V2: 7.9%, Human A40: 6.9% | LINC02263, AP001994.2, LINC02306, LINC00507, LINC00508, AC021613.1, ART3, AC110614.1, LINC01484, COL5A1 | 55595 | 4 |
| CA1-3 3 | Thalamus: 21.9%, Pons: 16.1%, Basal forebrain: 13.9% | Human PnEN: 8.9%, Human MoAN: 6.6%, Human PnRF: 3.8% | FCN1, CLEC12A, S100A4, TNFRSF1B, IFI30, LYZ, CYTIP, FGR, LILRB2, KYNU | 503 | 3 |
| MB-In 434 | Thalamus: 89.3%, Midbrain: 8.9%, Pons: 1.3% | Human VLN: 16.5%, Human LG: 15.2%, Human LP: 14.1% | NPSR1, CASR, LINC01210, OTX2, AL161757.4, SOX14, DMBX1, AL137100.3, NPPC, AL133493.1 | 8469 | 3 |
| MSN 215 | Basal forebrain: 85.0%, Amygdala: 9.2%, Cerebral cortex: 3.6% | Human NAC: 43.8%, Human GPe: 16.9%, Human BNST: 8.9% | LINC02137, AC090124.1, AC068875.1, DRD3, AC092604.1, AL049830.3, C6orf141, AL445584.2, AL020994.2, AC107208.1 | 16654 | 3 |
| MGE-IN 262 | Cerebral cortex: 94.5%, Basal forebrain: 2.1%, Hippocampus: 1.8% | Human M1C: 8.7%, Human MTG: 7.9%, Human A29-A30: 4.5% | RNF144B, AL136114.1, PRDM1, SULF1, LRRC38, TCIM, WNT16, MYO5B, KCNS3, AL096865.1 | 10119 | 4 |
| MSN 217 | Amygdala: 69.5%, Basal forebrain: 15.7%, Hypothalamus: 7.4% | Human CEN: 36.4%, Human BNST: 21.2%, Human SI: 12.3% | DCDC2C, PENK, AC015631.1, SIX3, P2RY1, LINC02462, LINC01305, CALCR, CACNG5, LINC01833 | 7260 | 4 |
| MSN 213 | Basal forebrain: 75.5%, Cerebral cortex: 12.2%, Amygdala: 9.4% | Human SI: 30.9%, Human NAC: 26.6%, Human GPe: 14.4% | PENK, AC008786.1, GPR6, DRD3, DRD2, P2RY1, SCIN, AC092604.1, ENTHD1, AL020994.2 | 13450 | 3 |
| CGE-IN 289 | Cerebral cortex: 82.1%, Hippocampus: 7.9%, Amygdala: 5.8% | Human M1C: 7.0%, Human MTG: 6.6%, Human MEC: 4.8% | TAC3, AC011990.1, KMO, AJ006995.1, SCTR, SHISA8, SLC22A3, AL450338.2, LINC02488, DCN | 14326 | 4 |
| DL-IT 140 | Cerebral cortex: 99.2%, Hippocampus: 0.5%, Basal forebrain: 0.2% | Human M1C: 11.6%, Human V1C: 8.4%, Human MTG: 7.5% | AC016687.2, LINC01680, AP003066.1, AL138927.1, AC016687.3, AL033539.2, AC023503.1, AC060765.2, AC073091.1, AL353709.1 | 12455 | 4 |
| Splatter 237 | Amygdala: 57.4%, Cerebral cortex: 20.2%, Basal forebrain: 8.4% | Human BL: 16.0%, Human CMN: 13.9%, Human CEN: 8.6% | FBN2, LHX6, PDGFD, SNTG2, SOX6, CDCA7, AKR1C8P, AC022905.1, IGFBP4, AP002989.1 | 2234 | 4 |
| MSN 207 | Basal forebrain: 92.9%, Thalamus: 4.1%, Amygdala: 1.9% | Human Pu: 54.1%, Human CaB: 28.2%, Human GPe: 5.8% | GPR6, AL139379.1, DRD2, AC073325.1, ADORA2A, LINC02137, PENK, GPR88, AL450332.1, LPL | 20569 | 3 |
| Mam 328 | Hypothalamus: 100.0%, Cerebral cortex: 0.0%, Amygdala: 0.0% | Human MN: 40.4%, Human HTHma: 29.5%, Human HTHtub: 26.0% | LINC00410, PITX2, AL691420.1, SIM1, LHX1-DT, FOXB1, AC087664.2, TNFRSF11B, LHX1, FSHR | 2346 | 3 |
| DL-CT/6b 109 | Cerebral cortex: 94.0%, Hippocampus: 3.3%, Basal forebrain: 1.8% | Human M1C: 16.5%, Human MTG: 9.8%, Human TH-TL: 5.7% | AC105021.1, AC073968.2, VWA2, IL4R, C5AR2, SEMA3E, SH2D1B, ABCC9, SGCG, ADAMTSL1 | 10646 | 4 |
| MGE-IN 254 | Cerebral cortex: 77.9%, Hippocampus: 13.0%, Amygdala: 4.5% | Human M1C: 8.3%, Human MTG: 6.2%, Human MEC: 4.6% | LINC01035, KLHL14, AL450345.2, ITGA1, PAWR, FBN2, AL590608.1, AC022133.2, SLC17A8, SST | 7539 | 4 |
| MGE-IN 250 | Cerebral cortex: 92.5%, Hippocampus: 2.4%, Basal forebrain: 2.2% | Human MEC: 7.1%, Human M1C: 7.0%, Human MTG: 5.6% | PROM1, NMU, ADGRG6, AC022905.1, KCNJ5, MCUB, PAWR, SST, SCARA5, AC113347.4 | 16604 | 4 |
| Misc 167 | Hippocampus: 76.8%, Cerebral cortex: 14.3%, Midbrain: 8.2% | Human Sub: 29.3%, Human CA1U-CA2U-CA3U: 25.4%, Human CA1-2R: 15.9% | LINC01524, ANO2, AL109930.1, FRMD6-AS2, AL391097.3, AL031674.1, TRPC6, RASSF9, AL391097.1, SEMA3D | 1973 | 3 |
| Mam 333 | Hypothalamus: 97.7%, Cerebral cortex: 1.0%, Thalamus: 0.8% | Human HTHma-HTHtub: 36.0%, Human HTHtub: 28.7%, Human MN: 27.0% | SIM1, AC244258.1, LHX1-DT, LTBP1, FOXB1, ZFHX4-AS1, CD36, LHX1, PMFBP1, PITX2 | 711 | 4 |
| Splatter 429 | Basal forebrain: 55.9%, Amygdala: 24.1%, Thalamus: 12.9% | Human SEP: 48.3%, Human BNST: 23.8%, Human ANC: 12.9% | PRDM16, FREM2, TTR, PRDM16-DT, AC092957.1, ZIC2, SCN5A, Z96074.1, C14orf39, ZIC5 | 9792 | 3 |
| MGE-IN 244 | Cerebral cortex: 77.4%, Amygdala: 13.7%, Hippocampus: 6.5% | Human MEC: 5.8%, Human MTG: 5.5%, Human LEC: 5.4% | NPY, SST, AC011586.2, LINC01687, ELFN1, MSR1, MCUB, ADAMTS12, CORT, TAC3 | 1172 | 4 |

| Cluster (Yao naming) | Top three regions | Top three dissections | Top Enriched Genes | Number of cells | Number of donors |
|---|---|---|---|---|---|
| MGE-IN 239 | Cerebral cortex: 95.4%, Hippocampus: 1.5%, Basal forebrain: 1.2% | Human M1C: 10.0%, Human MTG: 6.9%, Human A29-A30: 5.1% | AC022905.1, NMU, MAFB, SCARA5, PRDM1, TRBC2, MYO5B, PDCL2, BLM, PNOC | 6114 | 4 |
| Splatter 403 | Cerebral cortex: 45.6%, Amygdala: 33.5%, Hypothalamus: 9.8% | Human AON: 25.7%, Human CoA: 18.1%, Human Pir: 14.1% | FRMD7, TRDN, AC007422.1, KANK3, CACNG5, SP8, ROR2, LINC01812, LINC01411, SCGN | 2608 | 4 |
| DL-NP 87 | Cerebral cortex: 83.9%, Basal forebrain: 12.2%, Hypothalamus: 2.8% | Human ACC: 25.8%, Human A25: 14.5%, Human FI: 11.5% | IFNG-AS1, NPSR1-AS1, LINC02218, MROH2A, NPSR1, AC244502.1, SLC9C1, CDCP1, PIWIL1, ERG | 1203 | 4 |
| CA1-3 3 | Thalamus: 21.9%, Pons: 16.1%, Basal forebrain: 13.9% | Human PnEN: 8.9%, Human MoAN: 6.6%, Human PnRF: 3.8% | FCN1, CLEC12A, S100A4, TNFRSF1B, IFI30, LYZ, CYTIP, FGR, LILRB2, KYNU | 503 | 3 |
| MGE-IN 248 | Cerebral cortex: 78.8%, Hippocampus: 12.5%, Amygdala: 5.0% | Human V1C: 11.2%, Human M1C: 10.1%, Human MTG: 4.4% | HPGD, TAC1, FAM89A, AL450345.2, DCN, ALDH1A3, AC096751.2, CORT, BMP3, LHX6 | 2535 | 4 |
| CA1-3 3 | Thalamus: 21.9%, Pons: 16.1%, Basal forebrain: 13.9% | Human PnEN: 8.9%, Human MoAN: 6.6%, Human PnRF: 3.8% | FCN1, CLEC12A, S100A4, TNFRSF1B, IFI30, LYZ, CYTIP, FGR, LILRB2, KYNU | 503 | 3 |
| UL-IT 134 | Cerebral cortex: 99.3%, Hippocampus: 0.5%, Basal forebrain: 0.2% | Human M1C: 13.0%, Human MTG: 9.4%, Human A29-A30: 8.2% | AL157769.1, LINC02444, VIPR2, LINC01721, COBLL1, CCDC168, CCDC68, LINC02296, SLC38A11, AC246817.1 | 12095 | 4 |
| Splatter 313 | Hippocampus: 41.6%, Cerebral cortex: 36.5%, Amygdala: 7.6% | Human MEC: 14.5%, Human CA4C-DGC: 10.7%, Human DGR-CA4Rpy: 6.7% | AL592463.1, AGTR1, NDNF, TP73, AIM2, PRPH, LINC02770, AC005064.1, LINC01133, SCUBE1 | 449 | 3 |
| Splatter 425 | Thalamus: 35.4%, Hypothalamus: 29.5%, Amygdala: 29.1% | Human ANC: 34.9%, Human BNST: 23.4%, Human HTHpo-HTHso: 16.4% | SLC7A3, SHISAL2B, ARHGAP36, GABRQ, DGKK, NPY, CYP19A1, ECEL1, CALCR, CITED1 | 4253 | 4 |
| DL-IT 147 | Cerebral cortex: 65.1%, Amygdala: 19.4%, Basal forebrain: 11.8% | Human Cla: 9.4%, Human FI: 6.8%, Human Idg: 5.9% | CCN4, TMEM233, AP000331.1, THEMIS, AC079163.2, RXFP2, VIT, AC011754.1, SH2D1B, PXDN | 13265 | 4 |
| Splatter 410 | Hypothalamus: 82.0%, Thalamus: 11.6%, Amygdala: 5.2% | Human HTHpo-HTHso: 58.9%, Human HTHtub: 15.3%, Human ANC: 11.5% | AVP, OXT, HCRT, SIM1, OTP, TRH, CARTPT, TH, AC092675.1, PGF | 2511 | 3 |
| CGE-IN 286 | Cerebral cortex: 70.6%, Hippocampus: 13.8%, Amygdala: 10.2% | Human MEC: 5.8%, Human M1C: 5.5%, Human MTG: 4.3% | NDNF, CXCL14, AC005064.1, RELN, NMBR, LINC00298, MYH11, AC092819.3, NR2E1, SLC25A48 | 15597 | 4 |
| CA4 192 | Hippocampus: 99.6%, Cerebral cortex: 0.4%, Amygdala: 0.0% | Human CA4C-DGC: 79.6%, Human CA1C-CA3C: 9.2%, Human DGR-CA4Rpy: 3.8% | MORC1, HLX-AS1, AL450311.1, GHSR, C16orf74, SCGN, LINC02237, AC011754.1, LINC00210, GUCA1C | 1635 | 3 |
| DL-NP 93 | Cerebral cortex: 97.2%, Basal forebrain: 1.1%, Hypothalamus: 0.7% | Human M1C: 28.5%, Human A23: 15.2%, Human A29-A30: 9.3% | IFNG-AS1, NPSR1-AS1, ERG, CD200R1L, LINC02218, BARX2, CCDC80, IL26, LINC02042, ROBO3 | 2217 | 4 |
| CA1-3 3 | Thalamus: 21.9%, Pons: 16.1%, Basal forebrain: 13.9% | Human PnEN: 8.9%, Human MoAN: 6.6%, Human PnRF: 3.8% | FCN1, CLEC12A, S100A4, TNFRSF1B, IFI30, LYZ, CYTIP, FGR, LILRB2, KYNU | 503 | 3 |
| Astr 58 | Cerebral cortex: 57.1%, Hippocampus: 37.8%, Amygdala: 3.0% | Human CA4C-DGC: 9.8%, Human TH-TL: 5.9%, Human DGR-CA4Rpy: 5.6% | ID3, AQP4, GFAP, FGFR3, MLC1, F3, AGT, MT1G, AL137139.2, EMX2 | 8021 | 4 |
| CA1-3 3 | Thalamus: 21.9%, Pons: 16.1%, Basal forebrain: 13.9% | Human PnEN: 8.9%, Human MoAN: 6.6%, Human PnRF: 3.8% | FCN1, CLEC12A, S100A4, TNFRSF1B, IFI30, LYZ, CYTIP, FGR, LILRB2, KYNU | 503 | 3 |
| CA4 198 | Hippocampus: 89.9%, Midbrain: 10.1%, Cerebral cortex: 0.0% | Human CA4C-DGC: 45.1%, Human DGR-CA4Rpy: 21.8%, Human DGR-CA4: 15.3% | MORC1, IGF2BP2, KREMEN2, ARL6IP1P3, AC006487.1, AP001978.1, LINC02720, AC021594.2, ABCA13, AC019127.1 | 417 | 3 |
| CA1-3 3 | Thalamus: 21.9%, Pons: 16.1%, Basal forebrain: 13.9% | Human PnEN: 8.9%, Human MoAN: 6.6%, Human PnRF: 3.8% | FCN1, CLEC12A, S100A4, TNFRSF1B, IFI30, LYZ, CYTIP, FGR, LILRB2, KYNU | 503 | 3 |
| Thal-Exc 459 | Thalamus: 89.9%, Amygdala: 9.2%, Midbrain: 0.6% | Human MD: 26.0%, Human CM-Pf: 20.8%, Human ANC: 20.7% | SLC15A1, COL6A5, LHX9, FSHR, LINC01997, SLITRK6, AC069410.1, GBX2, RERGL, CYP27C1 | 4507 | 4 |
| Splatter 387 | Pons: 89.9%, Midbrain: 8.9%, Medulla: 0.7% | Human PB: 68.9%, Human DTg: 16.1%, Human PAG: 7.9% | PAX5, EBF2, LINC01798, AC090993.1, F13A1, DLK1, AC004080.1, TMEM255A, GLP1R, EBF3 | 1804 | 3 |
| Splatter 396 | Pons: 66.2%, Midbrain: 31.2%, Hypothalamus: 1.2% | Human DTg: 32.0%, Human PnEN: 26.2%, Human PAG: 17.4% | LMX1B, FEV, TPH2, SLC17A8, RLN3, GATA3, LINC01956, SLC6A4, GATA2, VWA5B1 | 1608 | 3 |
| Astr 53 | Hippocampus: 96.3%, Cerebral cortex: 1.5%, Basal forebrain: 1.3% | Human CA4C-DGC: 23.9%, Human DGR-CA4Rpy: 18.3%, Human CA1: 18.1% | LINC00499, GLI3, OAF, ETNPPL, PRDM16-DT, CD38, AC097518.2, FGFR3, OBI1-AS1, AL137139.2 | 4803 | 4 |
| MSN 430 | Amygdala: 87.9%, Hypothalamus: 6.5%, Thalamus: 3.4% | Human CEN: 41.5%, Human BM: 23.7%, Human BNST: 19.7% | FREM2, IQGAP2, P2RY1, PENK, TRHR, GABRG1, ANKRD34B, TMEM215, LINC01305, FREM2-AS1 | 2057 | 4 |
| DL-IT 137 | Cerebral cortex: 94.4%, Basal forebrain: 2.1%, Hippocampus: 1.9% | Human MTG: 7.5%, Human A13: 5.6%, Human M1C: 4.6% | LINC02196, AC099517.1, AC079380.1, AC073578.2, ADGRL4, PKD2L1, AC021134.1, AL450352.1, TRIM22, ARHGAP15 | 17737 | 4 |
| Splatter 367 | Midbrain: 96.9%, Thalamus: 2.6%, Pons: 0.3% | Human SC: 38.5%, Human IC: 29.3%, Human PAG: 15.2% | LHX9, TFAP2D, TAC1, SAMD11, POU4F1, FAM9B, BARHL1, AC092114.1, SLC17A6, LINC01568 | 4065 | 3 |
| MGE-IN 245 | Amygdala: 50.6%, Cerebral cortex: 31.0%, Hippocampus: 15.3% | Human CMN: 23.8%, Human BL: 9.2%, Human BM: 7.6% | NPY, LINC02397, ELFN1, SST, MCUB, AC022905.1, STK32B, TNFSF15, AC105411.1, AC113347.4 | 1566 | 4 |

| Cluster (Yao naming) | Top three regions | Top three dissections | Top Enriched Genes | Number of cells | Number of donors |
|---|---|---|---|---|---|
| LLC-IN 264 | Cerebral cortex: 84.5%, Amygdala: 5.7%, Basal forebrain: 4.7% | Human M1C: 6.5%, Human S1C: 5.6%, Human A29-A30: 4.8% | COL15A1, LINC00943, AC046195.2, NOG, LINC01837, C1QL1, CA8, PTHLH, CRH, ZNF385D-AS2 | 6669 | 4 |
| LLC-IN 268 | Hippocampus: 95.5%, Cerebral cortex: 2.4%, Amygdala: 1.1% | Human CA1U-CA2U-CA3U: 17.8%, Human CA3R: 17.6%, Human CA4C-DGC: 13.9% | CHRDL1, AL136456.1, EYA4, HAPLN1, LINC01344, CHST9, ANO1, TACR1, GRPR, NPPC | 3843 | 4 |
| Mam 329 | Hypothalamus: 99.7%, Midbrain: 0.3%, Thalamus: 0.1% | Human HTHma-HTHtub: 33.4%, Human HTHtub: 25.8%, Human MN: 23.5% | SIM1, AL691420.1, FOXB1, PITX2, AC244258.1, LHX1-DT, ADGRL4, DAPL1, ANXA3, AC093912.1 | 1487 | 3 |
| DL-IT 149 | Cerebral cortex: 98.6%, Basal forebrain: 0.6%, Hippocampus: 0.5% | Human M1C: 24.6%, Human MTG: 8.2%, Human S1C: 6.3% | THEMIS, LINC00343, AL356432.3, SH2D1B, AC011754.1, AC099517.1, ISLR, MIR3976HG, AC079380.1, AL117329.1 | 31428 | 4 |
| DL-IT 151 | Cerebral cortex: 44.8%, Amygdala: 33.6%, Basal forebrain: 21.4% | Human Cla: 20.0%, Human CEN: 18.2%, Human Pir: 10.5% | SMYD1, AC073225.1, AL136119.1, AL512452.1, ATP10A, NTNG2, CCN4, SMIM32, THEMIS, STPG2-AS1 | 18750 | 4 |
| eMSN 223 | Basal forebrain: 74.1%, Amygdala: 12.2%, Thalamus: 9.6% | Human CaB: 31.8%, Human SEP: 12.3%, Human NAC: 10.9% | FRMD7, LINC01735, FBXL21P, CASZ1, AL445584.2, SCN5A, LECT2, CALCRL, AC022133.2, C6orf141 | 2239 | 3 |
| DL-NP 91 | Cerebral cortex: 89.4%, Basal forebrain: 4.7%, Hippocampus: 3.3% | Human MTG: 16.0%, Human Idg: 7.9%, Human TF: 7.8% | CD200R1L, NPSR1-AS1, IFNG-AS1, LINC02042, CCDC80, ERG, BARX2, CD200R1L-AS1, TRIM22, PDLIM1 | 1523 | 4 |
| Splatter 393 | Thalamus: 76.5%, Hypothalamus: 10.6%, Midbrain: 6.0% | Human ANC: 35.5%, Human VLN: 13.9%, Human LG: 9.9% | AC006065.4, ISL1, LINC00943, LINC02058, SST, AC010478.1, SIX3, AC093298.2, COL8A1, INHBB | 2754 | 4 |
| Astr 54 | Cerebral cortex: 38.8%, Hippocampus: 33.1%, Amygdala: 22.1% | Human A35r: 18.3%, Human CA1: 15.4%, Human Sub: 12.0% | ETNPPL, LINC00499, GLI3, OBI1-AS1, FGFR3, PAPLN, AC097518.2, GJA1, AL160272.2, RANBP3L | 24168 | 4 |
| DL-NP 96 | Cerebral cortex: 85.1%, Amygdala: 7.0%, Basal forebrain: 5.7% | Human A38: 17.5%, Human MTG: 9.1%, Human Idg: 8.3% | HMGA2, AC129807.1, MYOCD, NPSR1-AS1, CD200R1L, ERG, COL8A1, TRIM22, FER1L6-AS2, CCDC80 | 685 | 4 |
| DL-IT 141 | Cerebral cortex: 95.4%, Basal forebrain: 2.0%, Hippocampus: 1.4% | Human MTG: 7.9%, Human Ig: 6.0%, Human TF: 5.7% | AP003066.1, AC080079.2, BARX2, LINC01680, AL033539.2, AL353709.1, AL138927.1, AC073091.1, AC073091.2, TRABD2A | 9992 | 4 |
| Amyg-Exc 406 | Amygdala: 50.5%, Cerebral cortex: 46.9%, Hypothalamus: 2.4% | Human Pir: 27.1%, Human A35-A36: 18.8%, Human CoA: 17.9% | RXFP2, AC112178.1, SCUBE1, FBXL21P, TFAP2D, IRAG2, AC109439.1, PLEKHG4B, CARTPT, NDST4 | 4858 | 4 |
| MGE-IN 252 | Cerebral cortex: 85.8%, Hippocampus: 8.0%, Basal forebrain: 3.6% | Human M1C: 11.7%, Human MTG: 6.1%, Human S1C: 4.3% | EDNRA, AL450345.2, NMU, SST, PAWR, TAC3, DCN, PNOC, CYP26A1, ITGA1 | 16079 | 4 |
| CA1-3 3 | Thalamus: 21.9%, Pons: 16.1%, Basal forebrain: 13.9% | Human PnEN: 8.9%, Human MoAN: 6.6%, Human PnRF: 3.8% | FCN1, CLEC12A, S100A4, TNFRSF1B, IFI30, LYZ, CYTIP, FGR, LILRB2, KYNU | 503 | 3 |
| eMSN 227 | Amygdala: 52.6%, Basal forebrain: 20.6%, Hypothalamus: 20.3% | Human CMN: 22.0%, Human SI: 19.2%, Human CEN: 13.6% | NPFFR2, FAM160A1, AC002072.1, NTS, FAM9B, NPNT, GDNF-AS1, RD3, WFS1, OTOF | 359 | 3 |
| Astr 64 | Hypothalamus: 93.0%, Amygdala: 1.6%, Thalamus: 1.6% | Human MN: 92.5%, Human CEN: 1.2%, Human CA3R: 0.6% | AC112253.1, AC098656.1, NAA11, LINC01094, CD44, AL627316.1, AC012405.1, AC073941.1, AC072022.1, AC093535.1 | 1181 | 3 |
| MSN 218 | Basal forebrain: 50.9%, Amygdala: 22.3%, Hypothalamus: 18.5% | Human SI: 38.1%, Human BNST: 19.5%, Human HTHpo: 16.2% | AC093515.1, AC079052.1, AC022433.1, ISL1, AC010478.1, DRD3, RXFP2, GDNF-AS1, AC006972.1, COL1A1 | 6712 | 3 |
| Splatter 395 | Midbrain: 92.6%, Hypothalamus: 6.5%, Thalamus: 0.7% | Human SN-RN: 58.2%, Human SN: 19.2%, Human PAG-DR: 6.2% | SLC6A3, LINC01956, TH, EN1, SLC18A2, LMX1B, SLC18A1, LMX1A, LINC00261, ASB4 | 998 | 3 |
| Mam 332 | Hypothalamus: 99.9%, Thalamus: 0.1%, Cerebral cortex: 0.0% | Human MN: 44.7%, Human HTHma: 37.0%, Human HTHtub: 9.6% | SIM1, FOXB1, PITX2, LHX1-DT, FRMPD2, LHX1, ANXA3, AC012501.2, AC083837.1, AL691420.1 | 1061 | 3 |
| eMSN 224 | Basal forebrain: 70.9%, Amygdala: 13.6%, Cerebral cortex: 8.2% | Human SI: 29.8%, Human NAC: 26.6%, Human BNST: 13.0% | SLC35D3, CASZ1, AL590867.1, AC022133.2, FBXL21P, AC022126.1, DRD1, LECT2, C6orf141, RXRG | 4297 | 3 |
| MSN 210 | Basal forebrain: 97.9%, Thalamus: 1.9%, Hippocampus: 0.2% | Human Pu: 80.1%, Human CaB: 12.7%, Human GPe: 4.5% | LINC02137, AL445584.2, AL450332.1, DDIT4L, PTPN7, LPL, AC092979.1, MME, PLEK, SLC25A24P1 | 10299 | 3 |
| Astr 56 | Hypothalamus: 30.0%, Thalamus: 21.5%, Amygdala: 14.3% | Human HTHso: 21.4%, Human VLN: 10.7%, Human HTHtub: 7.1% | TNC, GFAP, OBI1-AS1, FGFR3, GJA1, AC012405.1, EFEMP1, AQP4, SLC14A1, ETNPPL | 1672 | 4 |
| Misc 117 | Cerebral cortex: 82.3%, Basal forebrain: 7.9%, Hypothalamus: 7.7% | Human ACC: 23.6%, Human FI: 16.3%, Human AON: 14.0% | ABCB11, AC016074.2, ATP6V1C2, MIR548XHG, GABRQ, LINC00607, AP003062.2, LINC02717, FEZF2, AC068787.2 | 2655 | 3 |
| DL-IT 145 | Cerebral cortex: 65.1%, Hippocampus: 19.6%, Amygdala: 15.0% | Human MEC: 29.7%, Human LEC: 27.4%, Human Sub: 15.1% | AC079163.2, ONECUT1, AC016987.1, LINC02237, TWIST1, LINC01320, DCN, AGBL1, MYBPC1, CLEC19A | 4953 | 4 |
| Amyg-Exc 419 | Amygdala: 78.9%, Cerebral cortex: 14.0%, Thalamus: 5.1% | Human CMN: 29.0%, Human BM: 24.6%, Human CEN: 18.4% | AC025244.1, AC096759.1, SCN5A, FAM9B, GABRQ, LINC01920, VWA5B1, CYP19A1, CARM1P1, LINC02498 | 2170 | 3 |

| Cluster (Yao naming) | Top three regions | Top three dissections | Top Enriched Genes | Number of cells | Number of donors |
|---|---|---|---|---|---|
| UL-IT 129 | Cerebral cortex: 99.1%, Hippocampus: 0.4%, Basal forebrain: 0.4% | Human MTG: 14.4%, Human A44-A45: 8.5%, Human A46: 6.7% | LINC02296, LINC00326, AC110014.1, AC016598.2, AJ009632.2, LINC01478, AC016598.1, AC008696.2, LINC00508, LINC00507 | 46343 | 4 |
| UL-IT 125 | Cerebral cortex: 98.3%, Basal forebrain: 0.9%, Amygdala: 0.4% | Human MTG: 12.0%, Human V2: 10.3%, Human A5-A7: 9.0% | LINC02306, AC110614.1, LINC00507, LINC00326, AC097512.1, LINC00508, AC092440.1, AL132633.1, CYP1B1, AC093607.1 | 13397 | 4 |
| CGE-IN 296 | Cerebral cortex: 72.9%, Amygdala: 10.5%, Hippocampus: 9.0% | Human M1C: 8.1%, Human MEC: 6.2%, Human MTG: 4.7% | EREG, VIP, TAC3, IQGAP2, AREG, CXCL14, PTHLH, AC011990.1, SP8, NPR3 | 9524 | 4 |
| Astr 52 | Cerebral cortex: 90.3%, Hypothalamus: 3.9%, Hippocampus: 2.2% | Human MTG: 14.9%, Human STG: 7.9%, Human TH-TL: 6.2% | LINC01727, FGFR3, LINC00499, ZNF98, PRDM16-DT, F3, GJB6, PAPLN, GJA1, OAF | 25484 | 4 |
| Astr 63 | Medulla: 38.6%, Spinal cord: 20.0%, Pons: 15.4% | Human SpC: 20.0%, Human MoAN: 19.7%, Human IO: 9.6% | LINC01094, CD44, GFAP, AC012405.1, APLNR, AL627316.1, ID3, AL096709.1, AC073941.1, AEBP1 | 11325 | 4 |
| CA4 191 | Hippocampus: 98.1%, Cerebral cortex: 1.6%, Midbrain: 0.2% | Human CA4C-DGC: 41.8%, Human CA3R: 41.8%, Human CA1C-CA3C: 8.8% | AC021594.2, LINC02237, ITGA4, OR7E14P, LATS2, COL6A3, AC068985.1, AL450311.1, KCNG2, PTGFR | 486 | 4 |
| MGE-IN 247 | Hippocampus: 91.0%, Amygdala: 4.8%, Cerebral cortex: 3.4% | Human CA1C-CA3C: 20.3%, Human CA1U-CA2U-CA3U: 19.1%, Human CA1: 16.6% | AC022133.2, AC022126.1, AL450345.2, KCNJ5, TNFSF15, LINC02624, MYHAS, ZAP70, LINC02192, FAM89A | 1110 | 4 |
| Mam 326 | Hypothalamus: 100.0%, Cerebral cortex: 0.0%, Amygdala: 0.0% | Human MN: 64.1%, Human HTHma: 24.5%, Human HTHma-HTHtub: 11.0% | AC244258.1, SIM1, LINC00410, PITX2, FOXB1, AC087664.2, LHX1-DT, LHX1, AL691420.1, AC012501.2 | 1553 | 3 |
| Splatter 392 | Amygdala: 39.6%, Basal forebrain: 32.7%, Hypothalamus: 21.2% | Human BNST: 24.0%, Human SI: 17.5%, Human HTHpo: 11.4% | LHX8, GBX1, FGF10-AS1, FGF10, STAC, LHX6, DIAPH3, ELFN1, LINC00886, ASAH2 | 2659 | 4 |
| Thal-Exc 448 | Thalamus: 99.7%, Amygdala: 0.3%, Basal forebrain: 0.0% | Human VLN: 59.6%, Human CM-Pf: 18.2%, Human LP: 13.2% | CASQ2, AC093298.2, GDNF, CHRNA2, AL033530.1, FAM20A, TAFA4, MATN3, ST8SIA6, COL4A4 | 5500 | 3 |
| DL-CT/6b 103 | Cerebral cortex: 62.0%, Hippocampus: 25.8%, Amygdala: 12.1% | Human MEC: 45.4%, Human LEC: 14.7%, Human CA1U-CA2U-CA3U: 11.3% | CCN2, AC092131.1, VWA2, GUCA1C, SLC15A5, ITPRID1, C12orf42, F2R, LINC01735, LINC02218 | 1268 | 4 |
| CA1-3 3 | Thalamus: 21.9%, Pons: 16.1%, Basal forebrain: 13.9% | Human PnEN: 8.9%, Human MoAN: 6.6%, Human PnRF: 3.8% | FCN1, CLEC12A, S100A4, TNFRSF1B, IFI30, LYZ, CYTIP, FGR, LILRB2, KYNU | 503 | 3 |
| Splatter 340 | Pons: 88.3%, Midbrain: 9.5%, Thalamus: 1.6% | Human PB: 67.1%, Human DTg: 14.6%, Human PnEN: 5.1% | PTH2, ONECUT1, LHX9, MAB21L2, PRDM6, AC016044.1, DLK1, LINC01798, C1QL1, ONECUT3 | 547 | 3 |
| Amyg-Exc 160 | Amygdala: 90.3%, Cerebral cortex: 8.9%, Hippocampus: 0.8% | Human BL: 49.4%, Human BM: 21.7%, Human CMN: 13.3% | TFAP2D, DCSTAMP, PTGER3, LINC00334, TG, KLK7, NPFFR2, PLEKHG4B, FAM9B, MSC-AS1 | 7244 | 4 |
| CGE-IN 295 | Cerebral cortex: 67.0%, Amygdala: 17.8%, Hippocampus: 8.6% | Human M1C: 5.2%, Human MEC: 5.2%, Human BL: 4.2% | VIP, EREG, CRH, AC013265.1, TAC3, SLC6A2, AC062021.1, AL391832.4, SP8, PTHLH | 4922 | 4 |
| Splatter 400 | Basal forebrain: 54.0%, Amygdala: 22.9%, Hypothalamus: 14.7% | Human SI: 18.9%, Human BNST: 13.9%, Human HTHpo: 11.7% | SLC5A7, LHX8, CHAT, NTRK1, ECEL1, ISL1, AC091576.1, INSRR, OR8A1, CNBD1 | 769 | 3 |
| Astr 62 | Basal forebrain: 40.2%, Cerebellum: 19.3%, Midbrain: 16.2% | Human GPi: 30.1%, Human CbDN: 18.1%, Human SN: 10.1% | TNC, ANGPTL4, AC012405.1, GFAP, CPAMD8, AC073941.1, SLC14A1, GJA1, AL627316.1, CD44 | 24821 | 4 |
| DL-IT 142 | Cerebral cortex: 96.0%, Hippocampus: 1.3%, Basal forebrain: 1.2% | Human M1C: 26.9%, Human MTG: 11.3%, Human S1C: 4.8% | AC099517.1, Z68323.1, ADTRP, AC060765.2, AC021594.2, AL022724.3, AL359636.2, AL391117.1, AC008127.1, ALPL | 14326 | 4 |
| MSN 427 | Amygdala: 56.6%, Hypothalamus: 19.3%, Thalamus: 14.0% | Human BNST: 46.5%, Human ANC: 13.2%, Human HTHpo: 9.9% | AC022433.1, ISL1, AC010478.1, SCML4, GPR101, GABRG1, AC096759.1, TMEM215, AC025244.1, SCN5A | 3436 | 4 |
| HiDG 199 | Hippocampus: 96.8%, Thalamus: 3.2%, Cerebral cortex: 0.0% | Human CA4C-DGC: 50.3%, Human DGR-CA4Rpy: 22.4%, Human CA1R-CA2R-CA3R: 16.8% | AJ006995.1, LINC01697, AC112721.2, CR382285.1, POSTN, CHRNA1, AQP9, LINC01338, TARID, LINC02237 | 1365 | 3 |
| UL-IT 128 | Cerebral cortex: 99.9%, Hypothalamus: 0.0%, Basal forebrain: 0.0% | Human M1C: 73.1%, Human S1C: 17.9%, Human A32: 4.1% | AJ009632.2, MIR3976HG, LAMA2, TWIST1, VIPR2, AC087883.2, LINC02055, COL5A2, AL138694.1, SCTR | 10650 | 4 |
| Splatter 235 | Cerebral cortex: 31.8%, Amygdala: 28.5%, Basal forebrain: 25.2% | Human SI: 7.7%, Human CEN: 6.9%, Human Pu: 5.7% | NPY, CORT, SST, NOS1, AC005837.1, TLL2, CRHBP, TENT5A, TACR1, LHX6 | 3850 | 4 |
| Splatter 349 | Medulla: 84.7%, Spinal cord: 12.7%, Pons: 1.7% | Human MoRF-MoEN: 42.5%, Human MoAN: 28.5%, Human MoSR: 13.7% | SLC6A5, PAX2, HOXB3, HOXD3, PAX8, LINC00540, HOXB2, HOXA3, MAB21L2, GBX2 | 1557 | 3 |
| Astr 61 | Hippocampus: 47.7%, Cerebral cortex: 21.1%, Basal forebrain: 19.2% | Human CA1: 14.3%, Human Cla: 8.9%, Human CA4C-DGC: 6.7% | AC012405.1, TNC, GFAP, ID3, AC073941.1, AL627316.1, AQP4, CD38, CD44, ANGPTL4 | 13621 | 4 |
| DL-NP 85 | Cerebral cortex: 47.1%, Basal forebrain: 37.0%, Hypothalamus: 10.9% | Human ACC: 32.3%, Human SEP: 25.9%, Human SI: 9.7% | AC005909.2, FER1L6-AS2, PRELP, AC093912.1, NPSR1-AS1, VWA2, LPIN3, ABCC9, CDCP1, MROH2A | 514 | 4 |
| LLC-IN 270 | Cerebral cortex: 86.2%, Hippocampus: 6.7%, Thalamus: 3.3% | Human M1C: 12.3%, Human MTG: 5.2%, Human S1C: 4.5% | LINC01344, SFRP1, ANO1, EYA4, LINC00943, CA1, PDGFD, NOS1, CHRDL1, CDCA7 | 4751 | 4 |

| Cluster (Yao naming) | Top three regions | Top three dissections | Top Enriched Genes | Number of cells | Number of donors |
|---|---|---|---|---|---|
| eMSN 426 | Amygdala: 81.9%, Cerebral cortex: 10.0%, Thalamus: 3.2% | Human CEN: 36.3%, Human CMN: 31.4%, Human BM: 8.7% | LINC01497, NPFFR2, DGKK, PAX6, SCN5A, EYA2, AL161629.1, LINC01305, NPNT, AL592156.1 | 1600 | 4 |
| LLC-IN 271 | Cerebral cortex: 63.9%, Amygdala: 16.6%, Hippocampus: 9.9% | Human MEC: 11.6%, Human LEC: 5.7%, Human CEN: 4.6% | LINC01344, SFRP1, ANO1, CHRDL1, EYA4, HAPLN1, ADGRG6, AL136456.1, MYB, CASP4LP | 4045 | 4 |
| Mam 331 | Hypothalamus: 96.6%, Midbrain: 2.3%, Cerebral cortex: 1.1% | Human MN: 45.7%, Human HTHma: 22.9%, Human HTHma-HTHtub: 18.9% | PITX2, SIM1, PANCR, AC083837.1, FOXB1, AL691420.1, LHX1-DT, C14orf39, PKHD1L1, NPSR1 | 175 | 4 |
| CA1-3 3 | Thalamus: 21.9%, Pons: 16.1%, Basal forebrain: 13.9% | Human PnEN: 8.9%, Human MoAN: 6.6%, Human PnRF: 3.8% | FCN1, CLEC12A, S100A4, TNFRSF1B, IFI30, LYZ, CYTIP, FGR, LILRB2, KYNU | 503 | 3 |
| Splatter 421 | Hypothalamus: 64.1%, Amygdala: 19.7%, Thalamus: 14.8% | Human HTHpo: 29.3%, Human HTHso-HTHtub: 19.3%, Human BNST: 16.3% | ECEL1, FAM9B, TMEM114, ARHGAP36, AL035425.2, PGR, DGKK, ESR1, DIRAS3, PGR-AS1 | 1417 | 3 |
| Astr 57 | Hippocampus: 66.8%, Basal forebrain: 25.3%, Cerebral cortex: 4.0% | Human DGR-CA4Rpy: 26.6%, Human SEP: 21.1%, Human DGU-CA4Upy: 19.9% | CD38, OBI1-AS1, FGFR3, AL627316.1, AC012405.1, AL137139.2, TNC, LINC02649, GLI3, SLC14A1 | 6925 | 4 |
| CGE-IN 290 | Cerebral cortex: 83.0%, Hippocampus: 8.5%, Amygdala: 5.2% | Human MTG: 7.1%, Human M1C: 5.4%, Human A40: 3.9% | SHISA8, VIP, SCTR, FLT1, SLC22A3, KMO, PTHLH, GGT4P, CXCL12, LINC02488 | 20794 | 4 |
| CGE-IN 276 | Hippocampus: 73.7%, Amygdala: 13.0%, Cerebral cortex: 10.8% | Human CA3R: 12.6%, Human CA4C-DGC: 12.2%, Human CA1U-CA2U-CA3U: 11.2% | SHISAL2B, HTR3A, SLC17A8, LINC01997, GAS2L3, SCGN, AC112178.1, GPC3, PTGFR, ACADL | 2080 | 4 |
| Splatter 341 | Pons: 98.8%, Medulla: 1.2%, Cerebral cortex: 0.0% | Human DTg: 81.3%, Human PnEN: 11.1%, Human PnAN: 4.7% | EBF2, EBF3, AC079160.1, EBF1, DRGX, LINC01798, LINC01645, CNGA3, AC096719.1, LBX1-AS1 | 342 | 3 |
| DL-NP 90 | Cerebral cortex: 95.7%, Basal forebrain: 3.4%, Hypothalamus: 0.6% | Human M1C: 21.6%, Human FI: 10.5%, Human A25: 9.5% | IFNG-AS1, AC244502.1, NPSR1-AS1, CCDC80, CD200R1L, ERG, LINC02218, PDLIM1, BARX2, AC093912.1 | 1804 | 4 |
| eMSN 232 | Basal forebrain: 32.8%, Amygdala: 30.3%, Hypothalamus: 20.9% | Human SI: 25.7%, Human HTHpo: 17.0%, Human BNST: 16.4% | ZNF736P9Y, CAPSL, GABRQ, LMNTD1, EYA2, NPFFR2, OTOF, PCDH11Y, RD3, LINC01550 | 3682 | 3 |
| Splatter 378 | Amygdala: 29.4%, Basal forebrain: 25.2%, Hypothalamus: 22.3% | Human BNST: 27.8%, Human ANC: 18.1%, Human SEP: 16.8% | ONECUT3, C14orf39, ONECUT1, MIR217HG, ISL1, VWA5B1, AC022433.1, AC092957.1, COL15A1, AC010478.1 | 493 | 3 |
| LLC-IN 273 | Amygdala: 44.9%, Cerebral cortex: 33.4%, Hippocampus: 7.3% | Human BL: 11.5%, Human CEN: 9.9%, Human CMN: 9.0% | LINC01344, SFRP1, HAPLN1, CHRDL1, EYA4, ANO1, ADGRG6, TMEM255A, MYB, CHST9 | 3254 | 4 |
| CGE-IN 287 | Cerebral cortex: 53.6%, Amygdala: 17.5%, Hippocampus: 15.0% | Human M1C: 6.2%, Human MEC: 5.8%, Human BL: 4.1% | MYH11, LINC00298, HAPLN1, AC092819.3, AC137770.1, EYA4, KIT, TRPC3, AC132803.1, RELN | 18571 | 4 |
| MGE-IN 251 | Cerebral cortex: 94.0%, Hippocampus: 2.5%, Basal forebrain: 2.4% | Human MTG: 6.6%, Human A29-A30: 5.6%, Human M1C: 4.6% | EDNRA, NMU, MAFB, CYP26A1, PNOC, PDCL2, KCNJ5, SST, TRBC2, ADGRG6 | 16688 | 4 |
| Cer-In 300 | Cerebellum: 61.4%, Pons: 32.9%, Midbrain: 3.2% | Human CbDN: 58.9%, Human PnEN: 12.4%, Human PnRF: 8.1% | PAX2, SLC4A11, TFAP2B, NDNF, TFAP2A, IL15, SRD5A2, BHLHE22, ANGPT1, SLC22A3 | 1194 | 3 |
| Splatter 418 | Hypothalamus: 99.3%, Midbrain: 0.5%, Thalamus: 0.1% | Human HTHso-HTHtub: 52.0%, Human HTHtub: 32.9%, Human HTHma-HTHtub: 10.3% | FEZF1, FEZF1-AS1, PGR, LINC00540, DGKK, ZCCHC12, PRLR, PGR-AS1, SYTL4, ANKRD34B | 5390 | 3 |
| Splatter 371 | Midbrain: 99.7%, Cerebellum: 0.3%, Cerebral cortex: 0.0% | Human SC: 93.5%, Human IC: 6.0%, Human PAG-DR: 0.3% | CPA6, BNC2, NPSR1, EBF2, PGM5, OTOS, SHISAL2B, EBF1, LEF1, TAGLN2 | 398 | 3 |
| L RhombLip 322 | Pons: 83.9%, Medulla: 15.5%, Cerebral cortex: 0.4% | Human PnEN: 48.0%, Human DTg: 22.6%, Human MoRF-MoEN: 8.5% | LINC01982, AL359208.1, BMP5, AC007422.1, ADAMTS20, SLC5A7, EGFLAM, AL139351.3, INSRR, NTRK1 | 504 | 4 |
| HiDG 205 | Hippocampus: 96.5%, Midbrain: 3.4%, Thalamus: 0.0% | Human DGU-CA4Upy: 74.0%, Human CA1U-CA2U-CA3U: 17.4%, Human SN: 3.4% | AC097528.1, AC097487.1, LINC01697, AJ006995.1, AC079772.1, LINC02237, LINC02030, GRP, AC021088.1, LINC01324 | 5813 | 3 |
| Thal-Exc 450 | Thalamus: 99.9%, Basal forebrain: 0.0%, Hypothalamus: 0.0% | Human MD: 45.8%, Human ANC: 20.8%, Human ETH: 14.5% | CASQ2, RGS16, AC016766.1, SHOX2, GDNF, TGFBR2, FAM20A, VANGL1, IRAG1, LINC00868 | 3092 | 3 |
| Thal-Exc 449 | Thalamus: 98.7%, Amygdala: 1.0%, Midbrain: 0.3% | Human LP: 53.9%, Human ANC: 23.2%, Human Pul: 7.5% | CASQ2, RGS16, AC105213.1, MATN3, FAM20A, SHOX2, TAFA4, NTS, IRAG1, VANGL1 | 3539 | 3 |
| LLC-IN 267 | Hippocampus: 80.6%, Cerebral cortex: 13.8%, Amygdala: 1.9% | Human CA4C-DGC: 23.2%, Human CA1U-CA2U-CA3U: 10.4%, Human CA1C-CA3C: 9.5% | EXOC1L, CASP4LP, AC092819.3, ACADL, LINC01344, ANO1, CHST9, CHRDL1, ROR1, MYB | 1018 | 4 |
| U RhombLip 309 | Pons: 89.0%, Cerebellum: 8.6%, Medulla: 1.6% | Human PnEN: 37.8%, Human PnAN: 28.2%, Human PB: 12.3% | AC017037.5, AC108517.2, AC084740.1, LMX1A, ABCA13, AC092957.1, TMEM255B, CBLN1, CPLX4, AC080079.2 | 373 | 3 |
| Splatter 363 | Thalamus: 64.4%, Basal forebrain: 20.9%, Hypothalamus: 7.4% | Human ANC: 64.2%, Human SEP: 20.8%, Human BNST: 6.7% | EBF2, EBF3, AC009081.2, EOMES, CNGA3, AC092675.1, AC079160.1, GLP1R, ST8SIA6, CPA6 | 1482 | 3 |
| DL-CT/6b 98 | Cerebral cortex: 75.6%, Basal forebrain: 10.3%, Amygdala: 7.8% | Human MTG: 8.1%, Human FI: 8.1%, Human M1C: 5.9% | ERG, CCN2, AC079949.3, SLC15A5, SCUBE1, AC002331.1, CDH24, DPP4, TNIP3, OLFML2B | 4953 | 4 |

| Cluster (Yao naming) | Top three regions | Top three dissections | Top Enriched Genes | Number of cells | Number of donors |
|---|---|---|---|---|---|
| DL-CT/6b 110 | Cerebral cortex: 84.7%, Basal forebrain: 6.3%, Amygdala: 5.0% | Human FI: 9.6%, Human ACC: 9.2%, Human A38: 7.1% | AC105021.1, TRIM22, SULF1, C5AR2, AC073968.2, LGR6, CAV1, AC079380.1, S100Z, ABCC9 | 8324 | 4 |
| Splatter 412 | Hypothalamus: 97.4%, Midbrain: 2.2%, Thalamus: 0.3% | Human MN: 45.2%, Human HTHma-HTHtub: 26.3%, Human HTHtub: 14.1% | PMCH, AC022433.1, ISL1, TBX3, AC010478.1, C14orf39, AC026765.3, GAL, VWA5B1, RADX | 5062 | 3 |
| Mam 325 | Hypothalamus: 100.0%, Cerebral cortex: 0.0%, Amygdala: 0.0% | Human MN: 50.6%, Human HTHma-HTHtub: 43.4%, Human HTHma: 6.0% | LINC00410, AC244258.1, FSHR, PITX2, FOXB1, TNFRSF11B, AL691420.1, AC009975.1, LHX1-DT, LHX1 | 2155 | 3 |
| Misc 116 | Cerebral cortex: 93.4%, Amygdala: 3.3%, Basal forebrain: 2.6% | Human Idg: 22.6%, Human A38: 12.2%, Human M1C: 8.3% | ERG, MIR548XHG, ABCB11, CSN1S1, ATP6V1C2, AC016074.2, MIR1-1HG-AS1, FEZF2, POU3F1, AL158058.2 | 1341 | 3 |
| CA4 190 | Hippocampus: 100.0%, Cerebral cortex: 0.0%, Amygdala: 0.0% | Human CA3R: 68.8%, Human DGR-CA4Rpy: 17.3%, Human DGU-CA4Upy: 4.5% | AC019127.1, USH1C, LINC02720, AP006295.1, LATS2, AP001978.1, MLPH, HGF, HLX-AS1, WNK4 | 336 | 3 |
| DL-CT/6b 101 | Cerebral cortex: 65.6%, Hippocampus: 29.5%, Amygdala: 4.8% | Human MEC: 48.6%, Human Sub: 17.6%, Human LEC: 16.3% | AC092131.1, CCN2, AL596257.1, AC064875.1, KRT17, AC005482.1, SNHG31, AP003100.2, OLFML2B, AC022133.2 | 2972 | 4 |
| DL-CT/6b 100 | Cerebral cortex: 63.0%, Amygdala: 27.0%, Basal forebrain: 5.9% | Human AON: 9.6%, Human CMN: 9.2%, Human CEN: 7.7% | SLC15A5, VWA2, SULF1, AC010511.1, AC023503.1, LINC01194, LGR6, CPZ, ADTRP, CCN2 | 4900 | 4 |
| DL-NP 88 | Cerebral cortex: 92.2%, Basal forebrain: 4.8%, Amygdala: 1.9% | Human MTG: 15.3%, Human Idg: 10.9%, Human FI: 7.9% | IFNG-AS1, LINC02218, NPSR1-AS1, NPSR1, CD200R1L, AC244502.1, AC005909.2, IL26, CD36, CDCP1 | 1548 | 4 |
| DL-NP 83 | Cerebral cortex: 92.0%, Basal forebrain: 3.2%, Hypothalamus: 1.9% | Human M1C: 22.3%, Human S1C: 5.3%, Human MTG: 4.6% | AC022535.1, AL359313.1, AC044893.1, ERG, AL353072.2, MYH4, AC005909.2, NPSR1-AS1, ABCC9, LPO | 476 | 4 |
| U RhombLip 297 | Midbrain: 97.1%, Thalamus: 2.9%, Cerebral cortex: 0.0% | Human SC: 73.5%, Human SN: 20.6%, Human PAG: 2.9% | TPH1, RBP3, AC004852.2, GABRR3, AQP3, NEUROG1, ETV3L, AC063980.1, CACNA2D4, ASB15-AS1 | 34 | 2 |
| Cer-In 303 | Cerebellum: 96.8%, Pons: 1.5%, Medulla: 1.0% | Human CBV: 93.3%, Human CBL: 3.5%, Human PnEN: 0.7% | TFAP2B, AC010476.2, SCGN, AL162718.1, LIPG, XDH, LMCD1, TFAP2A, IL15, LRRC38 | 1439 | 3 |
| DL-NP 86 | Cerebral cortex: 97.0%, Basal forebrain: 1.8%, Hippocampus: 1.1% | Human M1C: 30.8%, Human S1C: 8.6%, Human Ig: 6.4% | IFNG-AS1, NPSR1-AS1, IL26, ROBO3, AC087863.2, LINC02444, SLC9C1, LINC02218, NPSR1, LINC01479 | 1508 | 4 |
| Splatter 402 | Midbrain: 58.2%, Hypothalamus: 41.6%, Amygdala: 0.1% | Human SN: 58.2%, Human HTHtub: 19.4%, Human HTHso-HTHtub: 19.2% | C14orf39, LMNTD1, AC004704.1, OTP, ISL1, P2RX2, LINC01920, FAM238B, AC010478.1, SLC10A4 | 1764 | 3 |
| Splatter 375 | Thalamus: 94.3%, Midbrain: 5.4%, Pons: 0.1% | Human ETH: 77.2%, Human MD-Re: 14.9%, Human PAG-DR: 3.3% | GPR151, POU4F1, MMRN1, DLK1, LHX9, LRRC55, LINC01497, EBF1, LINC01876, ONECUT1 | 2762 | 4 |
| Cer-In 299 | Cerebellum: 71.1%, Pons: 20.4%, Midbrain: 4.5% | Human CbDN: 64.6%, Human PnEN: 7.1%, Human PB: 6.6% | SLC6A5, PAX2, TFAP2B, IGF2BP2, TFAP2A, GBA3, INHBA, LIPG, ANGPT1, BHLHE22 | 560 | 4 |
| DL-CT/6b 107 | Cerebral cortex: 100.0%, Amygdala: 0.0%, Hippocampus: 0.0% | Human V1C: 68.8%, Human M1C: 9.8%, Human Pro: 9.2% | AC023503.1, LINC02699, ITGA11, VWA2, AL121904.2, SYT6, DPP4, CELSR1, AACSP1, AC016598.1 | 998 | 4 |
| CA1-3 3 | Thalamus: 21.9%, Pons: 16.1%, Basal forebrain: 13.9% | Human PnEN: 8.9%, Human MoAN: 6.6%, Human PnRF: 3.8% | FCN1, CLEC12A, S100A4, TNFRSF1B, IFI30, LYZ, CYTIP, FGR, LILRB2, KYNU | 503 | 3 |
| Splatter 343 | Medulla: 57.2%, Spinal cord: 34.1%, Pons: 5.9% | Human MoAN: 47.7%, Human SpC: 34.1%, Human MoSR: 9.2% | EBF2, DRGX, LMX1B, EBF1, MAFA, EBF3, HOXB7, HOXB3, AC024230.1, HOXD3 | 2797 | 4 |
| Cer-In 304 | Cerebellum: 95.5%, Pons: 2.4%, Medulla: 1.8% | Human CBV: 87.2%, Human CBL: 8.3%, Human PnAN: 1.4% | SCGN, TFAP2B, LMCD1, LIPG, AL162718.1, CLCNKA, LRRC38, TFAP2A, BHLHE22, AC010476.2 | 719 | 3 |
| MB-In 433 | Midbrain: 89.4%, Thalamus: 8.3%, Hypothalamus: 1.3% | Human SC: 51.7%, Human PTR: 13.8%, Human IC: 9.1% | OTX2, LINC01210, OTX2-AS1, AL137100.3, AL161757.4, CHODL, PRSS12, AL109761.1, GALR1, SOX14 | 1351 | 4 |
| MSN 219 | Amygdala: 33.2%, Basal forebrain: 32.8%, Hypothalamus: 18.6% | Human SI: 31.0%, Human BNST: 23.2%, Human HTHpo: 13.6% | ISL1, NPNT, AC010478.1, LINC02462, PROK2, AC015631.1, DRD3, AC022433.1, NMBR, LINC01951 | 8342 | 3 |
| CGE-IN 288 | Cerebral cortex: 70.6%, Amygdala: 13.7%, Hippocampus: 6.3% | Human M1C: 7.3%, Human MTG: 5.0%, Human MEC: 4.7% | HAPLN1, AC137770.1, LINC00298, AC132803.1, KIT, PKP2, EYA4, ROR2, NXPH2, GRPR | 24831 | 4 |
| UL-IT 130 | Cerebral cortex: 97.8%, Amygdala: 0.7%, Basal forebrain: 0.7% | Human A29-A30: 18.1%, Human MTG: 12.0%, Human A23: 7.7% | AC008568.1, AC025839.1, LINC02296, AJ009632.2, SLC38A11, AL117329.1, ENPEP, MIR3976HG, AC016598.2, MFGE8 | 25496 | 4 |
| DL-IT 139 | Cerebral cortex: 95.2%, Basal forebrain: 1.9%, Hippocampus: 1.5% | Human MTG: 12.0%, Human M1C: 7.8%, Human A43: 4.4% | AC016687.2, AC016687.3, AP003066.1, AL033539.2, AC099517.1, AC073578.2, Z68323.1, AC060765.2, AC021134.1, RCSD1 | 24657 | 4 |
| CA1-3 3 | Thalamus: 21.9%, Pons: 16.1%, Basal forebrain: 13.9% | Human PnEN: 8.9%, Human MoAN: 6.6%, Human PnRF: 3.8% | FCN1, CLEC12A, S100A4, TNFRSF1B, IFI30, LYZ, CYTIP, FGR, LILRB2, KYNU | 503 | 3 |

| Cluster (Yao naming) | Top three regions | Top three dissections | Top Enriched Genes | Number of cells | Number of donors |
|---|---|---|---|---|---|
| CA1-3 3 | Thalamus: 21.9%, Pons: 16.1%, Basal forebrain: 13.9% | Human PnEN: 8.9%, Human MoAN: 6.6%, Human PnRF: 3.8% | FCN1, CLEC12A, S100A4, TNFRSF1B, IFI30, LYZ, CYTIP, FGR, LILRB2, KYNU | 503 | 3 |
| DL-NP 89 | Cerebral cortex: 98.1%, Basal forebrain: 1.6%, Amygdala: 0.1% | Human M1C: 40.1%, Human FI: 8.0%, Human S1C: 7.7% | AC244502.1, NPSR1-AS1, IFNG-AS1, LINC02218, CD200R1L, CCDC80, LINC02042, CD36, AL161782.1, AC074139.1 | 1405 | 4 |
| CGE-IN 277 | Cerebral cortex: 77.4%, Amygdala: 13.3%, Hippocampus: 6.1% | Human M1C: 6.8%, Human LEC: 5.1%, Human FI: 4.8% | SCGN, SP8, LINC01497, DDR2, AC005064.1, LINC01162, ABCB5, TNFSF11, ANGPT1, RELN | 1650 | 4 |
| Splatter 391 | Hypothalamus: 92.3%, Thalamus: 3.3%, Midbrain: 2.8% | Human HTHpo-HTHso: 48.5%, Human HTHma-HTHtub: 13.7%, Human HTHma: 12.2% | SFRP1, AVP, PDGFD, AL158013.1, EGFL6, ABI3BP, GBX1, NFIA-AS2, AC008415.1, LINC01938 | 827 | 3 |
| OPC 33 | Pons: 23.8%, Midbrain: 22.9%, Thalamus: 19.2% | Human PnRF: 8.4%, Human CBV: 7.8%, Human SN-RN: 7.8% | BX284613.2, MROH9, FMO6P, PDGFRA, VCAN, FERMT1, AL512308.1, OLIG2, AC004852.2, COL9A1 | 48139 | 4 |
| Splatter 346 | Medulla: 60.2%, Spinal cord: 39.4%, Pons: 0.3% | Human SpC: 39.4%, Human MoAN: 34.3%, Human MoSR: 22.4% | LMX1B, HOXB3, HOXD3, AL161908.1, POU4F1, EBF3, HOXB7, HSPD1P8, NHLH1, TAC3 | 635 | 3 |
| MSN 206 | Hippocampus: 97.1%, Thalamus: 2.3%, Cerebral cortex: 0.6% | Human CA4C-DGC: 96.5%, Human Pul: 1.7%, Human LG: 0.6% | LINC02137, AC015631.1, GLI3, PLEK, AP003121.1, LINC02462, RGS9, MME, PTPN7, AL445584.2 | 172 | 2 |
| DL-CT/6b 112 | Cerebral cortex: 65.2%, Hippocampus: 33.8%, Amygdala: 0.6% | Human MEC: 57.0%, Human Sub: 17.6%, Human CA1U-CA2U-CA3U: 9.7% | AC092131.1, LINC01720, AC004917.1, RXFP2, CCN4, AL137191.1, CPZ, AC073968.2, AL596257.1, ADAMTSL1 | 6334 | 4 |
| UL-IT 138 | Cerebral cortex: 96.4%, Basal forebrain: 1.2%, Hippocampus: 1.2% | Human MTG: 14.3%, Human V2: 5.2%, Human A19: 4.5% | AL450352.1, AC073578.2, AC112770.1, AC016687.2, LINC01721, AP003066.1, SCHLAP1, COBLL1, AL355672.1, VIPR2 | 58263 | 4 |
| CGE-IN 293 | Cerebral cortex: 70.3%, Hippocampus: 15.1%, Amygdala: 8.5% | Human MEC: 5.0%, Human MTG: 5.0%, Human M1C: 4.3% | VIP, PTHLH, TAC3, AC020930.1, IQGAP2, HS3ST3A1, AC013265.1, AL391832.4, LAMA3, SLC22A3 | 20104 | 4 |
| DL-CT/6b 108 | Cerebral cortex: 89.4%, Hippocampus: 6.0%, Basal forebrain: 2.1% | Human M1C: 19.4%, Human A29-A30: 16.6%, Human A44-A45: 5.6% | LINC02699, AC105021.1, LINC02726, SH2D1B, IL4R, SEMA3E, LINC02718, AC046195.1, VWA2, TMC3 | 9206 | 4 |
| DL-CT/6b 104 | Cerebral cortex: 47.1%, Hippocampus: 34.6%, Amygdala: 18.1% | Human MEC: 31.3%, Human CA1U-CA2U-CA3U: 12.2%, Human Sub: 10.7% | AC092131.1, LINC02550, SLC15A5, CCN2, VWA2, AL663058.2, LINC00607, ADTRP, LINC02218, TRHR | 1973 | 4 |
| Splatter 423 | Amygdala: 50.6%, Thalamus: 19.8%, Hypothalamus: 15.4% | Human ANC: 19.2%, Human CEN: 17.4%, Human CMN: 13.8% | CYP19A1, SCN5A, DGKK, GABRQ, ARHGAP36, DIRAS3, NTS, FBXL21P, AL035425.2, VWA5B1 | 3114 | 3 |
| Splatter 366 | Midbrain: 57.4%, Pons: 19.9%, Thalamus: 15.2% | Human PAG: 17.9%, Human PTR: 12.3%, Human ETH: 9.4% | EBF2, EBF3, EBF1, KLHL1, SHOX2, AC007656.2, TFAP2D, RMST, SLC17A6, NCAPG | 11033 | 4 |
| Mam 330 | Hypothalamus: 100.0%, Cerebral cortex: 0.0%, Amygdala: 0.0% | Human MN: 44.6%, Human HTHma-HTHtub: 36.1%, Human HTHma: 19.2% | AC244258.1, AL691420.1, PITX2, FOXB1, SIM1, ANXA3, LHX1-DT, AC087664.2, AC093912.1, LHX1 | 1808 | 3 |
| Splatter 385 | Hypothalamus: 93.4%, Midbrain: 4.5%, Amygdala: 0.8% | Human HTHma-HTHtub: 46.9%, Human HTHtub: 19.6%, Human HTHma: 9.9% | LMX1A, LMX1B, LINC01667, CARTPT, IRX3, CRNDE, AR, TMEM255A, GABRE, IRX6 | 1800 | 3 |
| UL-IT 133 | Cerebral cortex: 100.0%, Basal forebrain: 0.0%, Hypothalamus: 0.0% | Human V1C: 62.9%, Human V2: 17.6%, Human Pro: 14.7% | LINC02388, AC112770.1, AC092448.1, AC126182.3, VIPR2, TRPC3, AC020637.1, IGFBP2, SLC38A11, LINC02296 | 20086 | 4 |
| Misc 114 | Cerebral cortex: 98.4%, Hippocampus: 1.2%, Basal forebrain: 0.3% | Human M1C: 23.2%, Human MTG: 16.1%, Human A23: 9.7% | CSN1S1, ERG, AP003062.2, ABCB11, MMP26, MIR1-1HG-AS1, LINC02433, AC016074.2, AL021396.1, ASGR2 | 915 | 4 |
| MGE-IN 256 | Cerebral cortex: 68.6%, Hippocampus: 21.8%, Basal forebrain: 5.0% | Human M1C: 8.3%, Human Sub: 6.3%, Human CA1U-CA2U-CA3U: 6.0% | WNT16, AC046195.2, MEPE, MYBPC1, AC006364.1, SRPK3, CPED1, AC106865.1, LHX6, AC022905.1 | 5526 | 4 |
| OPC 32 | Hypothalamus: 54.7%, Thalamus: 16.6%, Amygdala: 8.5% | Human MN: 54.6%, Human CM-Pf: 12.1%, Human CEN: 8.3% | MROH9, VCAN-AS1, FMO6P, FMO3, BX284613.2, AL512308.1, LHFPL3-AS1, Z94057.1, AC004852.2, LINC00402 | 929 | 4 |
| DL-CT/6b 106 | Cerebral cortex: 99.0%, Hippocampus: 0.6%, Basal forebrain: 0.4% | Human M1C: 20.6%, Human MTG: 8.8%, Human A23: 6.8% | AC023503.1, MDFIC, AC022133.2, AL138927.1, AC104689.2, DPP4, NPFFR2, KRT17, OLFML2B, CCN2 | 9905 | 4 |
| Cer-In 306 | Cerebellum: 91.6%, Pons: 3.4%, Medulla: 3.3% | Human CBV: 84.2%, Human CBL: 7.2%, Human PnEN: 1.5% | SCGN, TFAP2B, LIPG, LMCD1, LRRC38, BHLHE22, TFAP2A, LBX1-AS1, EN2, CHRNA3 | 6586 | 3 |
| LLC-IN 272 | Cerebral cortex: 67.5%, Amygdala: 16.1%, Basal forebrain: 7.8% | Human MEC: 6.2%, Human FI: 5.9%, Human LEC: 4.8% | LINC01344, SFRP1, ANO1, PDGFD, EYA4, FREM2, AP002989.1, CHRDL1, HAPLN1, ADGRG6 | 1960 | 4 |
| Splatter 373 | Midbrain: 61.4%, Thalamus: 36.6%, Amygdala: 0.7% | Human PTR: 32.8%, Human MG: 13.0%, Human SN-RN: 9.9% | PTH2, SHOX2, LHX9, FAM230I, SAMD11, VWA5B1, CALB2, NPB, LINC01920, AC074237.1 | 3543 | 3 |
| Splatter 334 | Thalamus: 91.7%, Midbrain: 8.2%, Pons: 0.1% | Human ETH: 62.8%, Human MD-Re: 27.9%, Human SC: 6.3% | GNG8, GPR151, LINC01307, CHRNB3, POU4F1, SLC5A7, SCUBE1, TRPM8, ACOXL, F13A1 | 1998 | 3 |
| Cer-In 301 | Pons: 92.6%, Medulla: 6.6%, Cerebral cortex: 0.7% | Human PnEN: 37.5%, Human PnAN: 26.5%, Human DTg: 16.9% | SLC6A5, TFAP2B, AC104574.2, PAX2, TFAP2A, BHLHE22, AC090796.1, XDH, AC018618.1, LBX1-AS1 | 136 | 4 |

| Cluster (Yao naming) | Top three regions | Top three dissections | Top Enriched Genes | Number of cells | Number of donors |
|---|---|---|---|---|---|
| Splatter 424 | Amygdala: 34.9%, Hypothalamus: 32.8%, Thalamus: 29.6% | Human ANC: 29.4%, Human BNST: 16.9%, Human HTHpo-HTHso: 15.5% | CYP19A1, CALCR, ARHGAP36, DGKK, GABRE, SCN5A, GABRQ, TMEM255A, LHX6, RADX | 2384 | 4 |
| CA4 193 | Hippocampus: 98.3%, Amygdala: 0.9%, Cerebral cortex: 0.5% | Human CA4C-DGC: 76.9%, Human CA1C-CA3C: 12.5%, Human DGR-CA4Rpy: 4.3% | SCGN, MORC1, LINC02237, AL450311.1, ITGA4, C16orf74, HLX-AS1, GUCA1C, TSPAN18, AC127526.2 | 1343 | 3 |
| DL-IT 150 | Cerebral cortex: 95.3%, Hippocampus: 2.2%, Basal forebrain: 1.3% | Human M1C: 11.9%, Human MTG: 7.3%, Human A23: 4.7% | SMYD1, OLFML2B, SMIM32, POSTN, THEMIS, AL136119.1, MCTP2, GAS2L3, AL356432.3, NTNG2 | 12307 | 4 |
| DL-CT/6b 97 | Cerebral cortex: 54.4%, Hippocampus: 42.5%, Amygdala: 2.1% | Human MEC: 52.6%, Human CA1: 22.6%, Human CA1U-CA2U-CA3U: 13.9% | LINC01194, GUCA1C, AC046195.2, AC023503.1, LINC02067, SLC15A5, ERG, AC092131.1, SCUBE1, IGFBP3 | 287 | 4 |
| Splatter 368 | Midbrain: 84.5%, Thalamus: 13.4%, Pons: 1.5% | Human PTR: 24.0%, Human PAG: 18.1%, Human IC: 15.7% | EBF3, TFAP2B, TFAP2D, LHX9, BNC2, EBF1, EBF2, ONECUT1, KLHL1, SCN7A | 10857 | 3 |
| CA4 195 | Hippocampus: 99.4%, Thalamus: 0.5%, Cerebral cortex: 0.1% | Human CA4C-DGC: 87.2%, Human CA3R: 4.8%, Human CA1C-CA3C: 4.7% | SCGN, MORC1, ITGA4, AC127526.2, ABCA13, AC127526.1, LINC02237, AL450311.1, AC021594.2, CASC6 | 2159 | 3 |
| Splatter 354 | Midbrain: 86.4%, Thalamus: 9.6%, Pons: 2.2% | Human PTR: 31.6%, Human SN-RN: 20.7%, Human SN: 19.1% | GATA3, LHX1, LHX1-DT, LHX5-AS1, GATA3-AS1, ASAH2, ST8SIA6, SAMD11, SIX3, GATA2 | 1441 | 3 |
| Splatter 389 | Pons: 67.3%, Midbrain: 29.6%, Thalamus: 2.4% | Human PnEN: 56.0%, Human PAG: 12.6%, Human DTg: 9.7% | NMB, GATA3, AL121974.1, IRX2, CALCR, GATA2, TACR3, TRPM8, VWA5B1, LMO1 | 759 | 3 |
| MSN 220 | Amygdala: 91.6%, Thalamus: 8.4%, Cerebral cortex: 0.0% | Human CEN: 87.5%, Human VA: 6.4%, Human La: 2.6% | FAM9B, ISL1, AC010478.1, PTGDR, NPNT, EBF1, DRD1, MYO3A, LPL, GDNF-AS1 | 391 | 3 |
| Splatter 348 | Medulla: 65.6%, Spinal cord: 26.9%, Pons: 6.0% | Human MoAN: 39.0%, Human SpC: 26.9%, Human MoSR: 19.3% | HOXB3, PAX2, GBX1, HOXD3, GPC3, PAX8, NPPC, HOXB7, SLC6A5, SALL3 | 1861 | 4 |
| Thal-Exc 457 | Thalamus: 98.3%, Amygdala: 1.4%, Cerebral cortex: 0.3% | Human ANC: 93.9%, Human ETH: 1.7%, Human BNST: 1.4% | LINC01036, SPTLC3, AC105213.1, SHOX2, LINC00922, NTS, TGFBI, AC082650.2, AC068989.1, LHX9 | 296 | 4 |
| Thal-Exc 372 | Midbrain: 53.2%, Thalamus: 44.2%, Cerebral cortex: 2.5% | Human PTR: 40.8%, Human MG: 22.6%, Human SN-RN: 10.0% | PTH2, FAM9B, CALCA, CALCB, GBX2, AC019068.1, AC012535.1, LINC01568, LHX9, CALB2 | 2593 | 4 |
| eMSN 230 | Amygdala: 41.5%, Basal forebrain: 26.8%, Cerebral cortex: 15.8% | Human SI: 21.0%, Human BNST: 15.9%, Human CMN: 12.9% | EYA2, NPFFR2, CAPSL, ZNF736P9Y, PRSS35, LINC00354, PCDH11Y, AL359649.1, ITGA6, ISM2 | 3123 | 3 |
| DL-CT/6b 99 | Cerebral cortex: 80.0%, Basal forebrain: 11.2%, Hippocampus: 6.5% | Human FI: 11.3%, Human ACC: 10.3%, Human Cla: 8.0% | CCN2, VWA2, SLC15A5, AC092131.1, DPP4, CDH24, AL596257.1, AC002331.1, KRT17, ADTRP | 4421 | 4 |
| Thal-Exc 452 | Thalamus: 98.9%, Midbrain: 1.1%, Amygdala: 0.0% | Human LG: 73.7%, Human VPL: 7.2%, Human MG: 6.6% | PVALB, ST8SIA6-AS1, PPP1R17, RGS16, SHOX2, TGFBR2, DEFB1, CASQ2, ITM2A, ST8SIA6 | 5044 | 3 |
| Cer-In 307 | Cerebellum: 70.3%, Medulla: 12.1%, Pons: 11.7% | Human CBV: 58.9%, Human CBL: 10.0%, Human PnEN: 5.7% | SCGN, TFAP2B, LINC00492, TFAP2A, TPTE2P4, CLCNKA, LIPG, LRRC38, LINC02552, LBX1-AS1 | 3020 | 3 |
| Mam 323 | Hypothalamus: 99.7%, Thalamus: 0.2%, Midbrain: 0.2% | Human MN: 49.2%, Human HTHma-HTHtub: 39.6%, Human HTHma: 7.5% | AC244258.1, FOXB1, AL691420.1, PITX2, LHX1-DT, LHX1, DRD3, ADGRD1, STAC, AL139231.1 | 654 | 3 |
| Thal-Exc 454 | Thalamus: 99.5%, Midbrain: 0.5%, Cerebral cortex: 0.0% | Human Pul: 76.9%, Human LP: 8.6%, Human ETH: 7.5% | RGS16, ST8SIA6-AS1, AP000331.1, AL162718.1, ST8SIA6, LEF1, VANGL1, LINC00868, CYP27C1, MATN3 | 11893 | 3 |
| Amyg-Exc 408 | Cerebral cortex: 82.2%, Amygdala: 13.3%, Hypothalamus: 4.4% | Human AON: 77.8%, Human CoA: 8.9%, Human Pir: 4.4% | EOMES, RXFP2, CDHR1, CARTPT, LINC01866, LINC01965, SHISA3, LGR6, AKAIN1, LINC01621 | 45 | 2 |
| Splatter 417 | Hypothalamus: 95.8%, Midbrain: 4.0%, Amygdala: 0.1% | Human HTHtub: 32.3%, Human HTHso-HTHtub: 31.4%, Human HTHma-HTHtub: 26.7% | GHRH, OTP, CALCR, ECEL1, DLK1, C14orf39, ISL1, GAL, GABRE, SIX6 | 3885 | 4 |
| DL-CT/6b 105 | Cerebral cortex: 80.5%, Basal forebrain: 8.4%, Hippocampus: 7.2% | Human ACC: 11.0%, Human FI: 10.2%, Human A25: 9.1% | AL138927.1, KRT17, CCN2, AC023503.1, MDFIC, AC104689.2, SNHG31, OLFML2B, AL358335.2, LINC01680 | 4269 | 4 |
| Splatter 376 | Hypothalamus: 51.0%, Midbrain: 37.9%, Thalamus: 6.6% | Human SN-RN: 18.1%, Human HTHma: 17.5%, Human HTHso-HTHtub: 14.7% | TRH, LMX1A, SIM1, OTP, MIR217HG, FEZF1-AS1, ONECUT1, DLK1, PITX2, ONECUT3 | 2573 | 3 |
| CGE-IN 284 | Cerebral cortex: 74.7%, Amygdala: 9.9%, Hippocampus: 9.3% | Human M1C: 5.9%, Human MEC: 5.8%, Human MTG: 4.9% | AC011990.1, PCDH18, CXCL14, LINC01931, KMO, PTGFR, LINC02397, AL136114.1, LINC02144, VIP | 9109 | 4 |
| Splatter 382 | Amygdala: 32.1%, Hypothalamus: 30.4%, Basal forebrain: 21.9% | Human BNST: 19.6%, Human SI: 14.2%, Human HTHpo: 13.5% | FST, LINC00348, CALCR, RSPO3, KCNJ5, DMRTA2, LHX5-AS1, PGM5, LHX5, FAM9B | 713 | 3 |
| Astr 55 | Basal forebrain: 82.4%, Thalamus: 7.0%, Hypothalamus: 5.0% | Human Pu: 19.1%, Human CaB: 17.0%, Human SI: 15.7% | ETNPPL, AC097518.2, PRDM16-DT, AL445259.1, CD38, OBI1-AS1, FGFR3, F3, APOC1, EFEMP1 | 3576 | 4 |
| OPC 34 | Hippocampus: 33.6%, Cerebral cortex: 30.9%, Amygdala: 16.5% | Human CA1: 11.7%, Human CEN: 8.5%, Human A35r: 6.4% | AC004852.2, AL512308.1, PDGFRA, BX284613.2, GPR17, VCAN, CSPG4, AC117464.1, OLIG2, FERMT1 | 31058 | 4 |

| Cluster (Yao naming) | Top three regions | Top three dissections | Top Enriched Genes | Number of cells | Number of donors |
|---|---|---|---|---|---|
| B 51 | Cerebellum: 95.8%, Pons: 1.9%, Medulla: 1.9% | Human CBV: 82.3%, Human CBL: 13.3%, Human MoAN: 1.3% | LINC01727, PAX3, AL627316.1, LCAT, AL355306.2, LINC01726, SMTN, AC098656.1, LGR6, AL449403.3 | 8041 | 3 |
| CGE-IN 294 | Cerebral cortex: 64.4%, Hippocampus: 14.7%, Amygdala: 10.8% | Human MEC: 5.3%, Human M1C: 5.2%, Human Cla: 4.7% | VIP, ABI3BP, PTHLH, IQGAP2, TAC3, CRH, DLX1, AC104129.1, NPR3, LAMA3 | 14134 | 4 |
| Splatter 415 | Hypothalamus: 78.7%, Midbrain: 7.6%, Thalamus: 3.7% | Human HTHpo-HTHso: 30.8%, Human HTHso-HTHtub: 26.6%, Human HTHtub: 7.5% | AVP, ECEL1, ARHGAP36, C14orf39, HMX3, ISL1, SIX6, DIRAS3, PMCH, ZCCHC12 | 25816 | 4 |
| Thal-Exc 456 | Thalamus: 99.8%, Hippocampus: 0.2%, Cerebral cortex: 0.0% | Human LG: 92.5%, Human Pul: 3.7%, Human MG: 2.0% | ST8SIA6, AC012535.1, AC093334.1, ST8SIA6-AS1, AP000331.1, AC006487.1, SHOX2, ANXA3, AC087482.1, VANGL1 | 509 | 3 |
| Splatter 365 | Pons: 43.0%, Midbrain: 30.2%, Medulla: 17.1% | Human DTg: 13.8%, Human PnEN: 12.8%, Human MoRF-MoEN: 11.6% | SLC6A5, GLRA1, ONECUT1, LHX5-AS1, MAB21L2, LHX1, LINC01933, ASAH2, KLHL1, IGDCC4 | 28081 | 4 |
| Mam 324 | Hypothalamus: 100.0%, Cerebral cortex: 0.0%, Amygdala: 0.0% | Human MN: 52.3%, Human HTHma-HTHtub: 34.2%, Human HTHma: 13.5% | LINC00410, AC244258.1, PITX2, FOXB1, TNFRSF11B, AL691420.1, LHX1-DT, LHX1, DRD3, MYO5B | 3071 | 3 |
| Splatter 374 | Pons: 77.9%, Midbrain: 19.6%, Medulla: 2.0% | Human PB: 67.0%, Human PTR: 16.2%, Human DTg: 8.6% | LMX1A, TFAP2B, LMX1B, CALCB, CALCA, EBF2, AC007614.1, ONECUT1, BNC2, CBLN1 | 1666 | 3 |
| Splatter 383 | Hypothalamus: 68.0%, Midbrain: 24.6%, Thalamus: 6.6% | Human HTHma: 35.5%, Human MN: 17.1%, Human HTHma-HTHtub: 15.2% | LMX1A, PITX2, EBF2, EBF3, AP005242.1, FOXA1, LMX1B, LINC01443, LINC01876, AC022034.3 | 4024 | 3 |
| Thal-Exc 447 | Thalamus: 100.0%, Cerebral cortex: 0.0%, Amygdala: 0.0% | Human LG: 98.6%, Human Pul: 0.6%, Human VPL: 0.2% | SHOX2, PVALB, CHRNA2, AL355612.1, DEFB1, AP001605.1, DCDC2C, PPP1R17, TGFBR2, LINC01695 | 2924 | 3 |
| OPC 35 | Cerebral cortex: 62.3%, Hippocampus: 21.4%, Basal forebrain: 6.5% | Human A35r: 17.0%, Human Sub: 9.1%, Human MTG: 8.0% | AC004852.2, PDGFRA, GPNMB, GPR17, AL512308.1, OLIG2, COL20A1, CSPG4, OLIG1, AC117464.1 | 23965 | 4 |
| Fibr 30 | Cerebral cortex: 35.3%, Basal forebrain: 29.7%, Medulla: 10.6% | Human SEP: 29.0%, Human A35r: 21.2%, Human MN: 6.4% | CFH, SLC6A20, COL1A2, KDR, VIM, SLC5A5, COL3A1, SLC6A13, SLC16A12, SLC6A12 | 283 | 3 |
| U RhombLip 311 | Cerebellum: 72.7%, Medulla: 10.3%, Pons: 8.2% | Human CBV: 33.7%, Human CBL: 23.0%, Human CbDN: 16.1% | CBLN3, AC073225.1, AC092957.1, CRTAM, SLC22A31, CDH15, PRR35, LINC01798, FAT2, AC044798.3 | 97038 | 4 |
| Misc 118 | Hippocampus: 60.6%, Cerebral cortex: 26.6%, Midbrain: 11.9% | Human Sub: 36.7%, Human TH-TL: 23.9%, Human SC: 11.9% | CSN1S1, ERG, AC005482.1, ATP6V1C2, SLC5A7, LINC02433, LINC00607, ABCB11, POU3F1, EHF | 109 | 3 |
| Thal-Exc 460 | Thalamus: 98.9%, Midbrain: 0.9%, Cerebral cortex: 0.1% | Human CM-Pf: 41.9%, Human CM: 30.5%, Human ETH: 13.0% | LINC01239, DMRTA1, MUC7, AC092445.1, AL159156.1, LINC01151, AC037486.1, ITIH5, SMILR, COL6A5 | 7103 | 4 |
| Splatter 355 | Midbrain: 87.7%, Pons: 9.3%, Thalamus: 2.4% | Human IC: 35.8%, Human SC: 26.8%, Human PTR: 14.3% | PAX5, GATA3, LINC01956, LHX5-AS1, LINC01497, PAX7, LINC01210, GATA3-AS1, GATA2, LHX1 | 4371 | 4 |
| Splatter 394 | Pons: 92.2%, Midbrain: 6.7%, Medulla: 0.5% | Human PB: 78.3%, Human DTg: 10.3%, Human PAG: 4.6% | NPS, LINC01798, IRX4, SLC5A7, LHX9, AC006296.3, AC074099.1, IRX2, LINC01876, TH | 1791 | 3 |
| CGE-IN 282 | Cerebral cortex: 82.6%, Hippocampus: 11.9%, Amygdala: 2.2% | Human A32: 5.2%, Human MTG: 5.1%, Human M1C: 4.8% | AC004862.1, AC091576.1, ADAM33, CXCL14, NDNF, AL139090.1, AL391832.4, AC073316.1, ARHGAP36, AC068633.1 | 10065 | 4 |
| L RhombLip 319 | Pons: 97.8%, Midbrain: 1.2%, Medulla: 1.0% | Human PN: 45.9%, Human PnEN: 25.5%, Human PnRF: 15.0% | LINC00355, IRX2, AL139351.3, AC136431.1, AC093765.2, S100A10, ADAMTS20, LINC02476, CDH23, BARHL1 | 8164 | 3 |
| MGE-IN 246 | Hippocampus: 72.3%, Cerebral cortex: 16.5%, Amygdala: 8.9% | Human CA4C-DGC: 21.5%, Human CA3R: 12.6%, Human CA1C-CA3C: 9.5% | AC113347.4, PROM1, ELFN1, SST, LINC02192, TNFSF15, TAC1, LINC01687, DEPDC1, NGF | 2100 | 4 |
| Splatter 345 | Medulla: 52.3%, Spinal cord: 35.4%, Midbrain: 6.8% | Human MoAN: 42.2%, Human SpC: 35.4%, Human MoSR: 9.3% | LMX1B, DRGX, SKOR2, HOXB7, HOXB3, EBF3, EBF2, EBF1, TLX3, HOXD3 | 2720 | 4 |
| Splatter 358 | Midbrain: 91.5%, Thalamus: 7.8%, Pons: 0.4% | Human SC: 30.4%, Human PTR: 26.0%, Human PAG: 14.0% | TFAP2B, PAX3, GATA3, LINC01497, PAX7, AC005208.1, TFAP2A, LHX5-AS1, GATA3-AS1, AC025946.1 | 3175 | 4 |
| U RhombLip 310 | Cerebellum: 52.4%, Pons: 26.2%, Medulla: 15.1% | Human CBV: 34.8%, Human PnAN: 19.7%, Human CbDN: 13.1% | CBLN3, AC092957.1, CRTAM, AC021698.1, LINC01036, AC073225.1, LINC01798, SLC22A31, AC044798.3, AC020659.1 | 33405 | 4 |
| Fibr 31 | Basal forebrain: 47.3%, Medulla: 20.3%, Hypothalamus: 12.4% | Human SEP: 47.2%, Human MN: 11.8%, Human MoAN: 8.8% | PTGDR, BNC2, BMP5, FOXC1, COL3A1, COL1A2, SLPI, SFRP2, SPTLC3, TBX15 | 659 | 3 |
| Misc 3 | Thalamus: 21.9%, Pons: 16.1%, Basal forebrain: 13.9% | Human PnEN: 8.9%, Human MoAN: 6.6%, Human PnRF: 3.8% | FCN1, CLEC12A, S100A4, TNFRSF1B, IFI30, LYZ, CYTIP, FGR, LILRB2, KYNU | 503 | 3 |
| L RhombLip 321 | Pons: 95.9%, Medulla: 3.3%, Midbrain: 0.4% | Human PnEN: 40.4%, Human PN: 30.9%, Human DTg: 13.1% | IRX2, AL139351.3, GADL1, AC136431.1, NLRP1, CDH23, LINC02476, LINC01798, INSRR, VEPH1 | 1119 | 4 |
| L RhombLip 318 | Pons: 98.8%, Medulla: 0.7%, Midbrain: 0.5% | Human PN: 54.9%, Human PnEN: 17.1%, Human DTg: 14.1% | AC093765.2, AL139351.3, ADAMTS20, LINC00836, LINC00210, BMP5, AC136431.1, BARHL1, LINC00355, IRX2 | 7999 | 3 |

| Cluster (Yao naming) | Top three regions | Top three dissections | Top Enriched Genes | Number of cells | Number of donors |
|---|---|---|---|---|---|
| UL-IT 131 | Cerebral cortex: 99.9%, Hypothalamus: 0.0%, Basal forebrain: 0.0% | Human M1C: 71.0%, Human S1C: 11.1%, Human A43: 3.7% | AC092353.2, LINC02306, MIR3976HG, AL132633.1, AC018742.1, AC087883.2, LAMA2, AL450313.1, DPP4, LINC01965 | 22050 | 4 |
| Splatter 388 | Pons: 90.9%, Hypothalamus: 4.1%, Amygdala: 3.6% | Human PnEN: 81.7%, Human PnAN: 4.6%, Human DTg: 4.1% | LINC01798, SCTR, LHX5-AS1, VSX2, APELA, VWA5B1, HPGD, LINC01828, LINC01799, ADCYAP1 | 197 | 3 |
| Splatter 413 | Hypothalamus: 92.1%, Midbrain: 5.8%, Pons: 1.4% | Human MN: 38.6%, Human HTHma-HTHtub: 21.6%, Human HTHma: 18.2% | SIM1, PITX2, EBF3, AL158013.1, LHX1-DT, NTS, LHX1, ADGRG4, PANCR, FOXB1 | 4634 | 3 |
| L RhombLip 315 | Pons: 95.9%, Medulla: 1.9%, Cerebral cortex: 1.5% | Human PnEN: 47.2%, Human DTg: 31.1%, Human PN: 9.4% | ADAMTS20, LINC00616, AL139351.3, BARHL1, IRX2, ADGRD2, EGFLAM, AC093765.2, C6orf141, LINC02381 | 267 | 4 |
| Misc 113 | Cerebral cortex: 100.0%, Amygdala: 0.0%, Hippocampus: 0.0% | Human M1C: 73.3%, Human S1C: 14.1%, Human A43: 8.9% | AC005100.1, LINC02424, AC105213.1, LINC00922, OR51E2, ERG, NAALADL2-AS2, AC128707.1, LINC02008, LINC00943 | 630 | 4 |
| L RhombLip 317 | Pons: 97.6%, Medulla: 2.1%, Midbrain: 0.2% | Human PnRF: 36.1%, Human PnEN: 23.6%, Human PN: 22.5% | BMP5, ADAMTS20, AL139351.3, AC093765.2, INSRR, LINC00836, CDH23, AC136431.1, IRX2, EGFLAM | 4858 | 3 |
| CGE-IN 279 | Cerebral cortex: 54.6%, Hippocampus: 26.0%, Amygdala: 14.6% | Human MEC: 5.4%, Human CMN: 5.0%, Human CA4C-DGC: 5.0% | LINC01162, AC006305.1, HTR3A, HGFAC, SP8, CXCL14, SLC17A8, CNR1, LINC01470, LINC01416 | 7444 | 4 |
| Splatter 369 | Midbrain: 96.5%, Thalamus: 2.3%, Pons: 0.6% | Human PTR: 42.2%, Human SC: 39.6%, Human IC: 6.3% | TFAP2B, FOXB1, LHX9, TFAP2D, SHOX2, NTS, EBF3, FAM9B, LEF1, AC074237.1 | 2975 | 4 |
| MiG 11 | Cerebral cortex: 37.4%, Hippocampus: 24.2%, Thalamus: 11.1% | Human DGR-CA4Rpy: 6.3%, Human MTG: 5.7%, Human DGU-CA4Upy: 5.7% | CSF1R, CX3CR1, APBB1IP, FYB1, AL392172.1, LINC00996, LNCAROD, SYK, DOCK8, C3 | 20036 | 4 |
| Cer-In 305 | Cerebellum: 88.0%, Pons: 4.7%, Midbrain: 3.7% | Human CBV: 79.1%, Human CBL: 7.9%, Human IC: 3.7% | SCGN, TFAP2B, LIPG, LRRC38, LBX1-AS1, BHLHE22, LINC02058, LHX1, LMCD1, EN2 | 191 | 3 |
| CGE-IN 278 | Cerebral cortex: 78.0%, Amygdala: 10.4%, Hippocampus: 8.1% | Human MEC: 5.9%, Human LEC: 4.8%, Human M1C: 4.7% | SP8, CXCL14, DDR2, AC006305.1, AC005064.1, RELN, ABCB5, GRPR, AC092819.3, NDNF | 7177 | 4 |
| DL-CT/6b 111 | Hippocampus: 92.5%, Amygdala: 6.3%, Cerebral cortex: 1.2% | Human CA1: 33.5%, Human CA1U-CA2U-CA3U: 27.2%, Human CA2U-CA3U: 9.3% | CLPSL1, VWA2, CPZ, SULF1, AL353072.2, AC092546.1, ADTRP, MC4R, GABRQ, AC091576.1 | 1546 | 4 |
| Splatter 432 | Hypothalamus: 79.8%, Midbrain: 19.6%, Thalamus: 0.6% | Human HTHma-HTHtub: 31.5%, Human SN: 19.6%, Human HTHtub: 19.5% | HDC, MIR3663HG, LINC02789, SLC18A2, TECRL, QRFPR, TBX3, PPP1R1C, GMNC, AC022433.1 | 2555 | 3 |
| Splatter 359 | Midbrain: 99.1%, Thalamus: 0.4%, Pons: 0.3% | Human SC: 70.1%, Human IC: 14.3%, Human PTR: 8.5% | TFAP2B, PAX7, GATA3, TFAP2A, ADAMTS20, LHX5-AS1, EMX2, LINC01210, GATA3-AS1, AC025946.1 | 6660 | 4 |
| CGE-IN 283 | Cerebral cortex: 81.0%, Hippocampus: 9.3%, Amygdala: 5.0% | Human MTG: 5.2%, Human A32: 4.5%, Human MEC: 4.1% | CXCL14, AC004862.1, BAGE2, PCDH18, AL139090.1, AL356585.4, AL391832.4, SGO1-AS1, AC073316.1, KMO | 10636 | 4 |
| L RhombLip 316 | Pons: 76.6%, Medulla: 22.0%, Midbrain: 0.6% | Human PnEN: 53.4%, Human MoAN: 10.7%, Human DTg: 8.7% | ADAMTS20, NTRK1, AL139351.3, INSRR, BARHL1, AC022239.1, AL359208.1, CDH23, IRX2, HOXD3 | 1939 | 4 |
| DL-IT 152 | Basal forebrain: 76.2%, Amygdala: 19.9%, Cerebral cortex: 3.8% | Human Cla: 70.7%, Human CEN: 17.6%, Human Pu: 5.6% | AC012593.2, ANXA1, SMIM32, LINC01876, AC074099.1, LINC02505, AC112481.2, PLA2G4A, SMYD1, AL136119.1 | 6048 | 4 |
| Thal-Exc 446 | Thalamus: 99.9%, Hippocampus: 0.0%, Midbrain: 0.0% | Human LG: 99.9%, Human PAG-DR: 0.0%, Human LP: 0.0% | DCDC2C, AL355612.1, LINC01491, AC092078.3, SHOX2, AC044810.3, DEFB1, LCP1, AC092078.2, PPP1R17 | 15148 | 3 |
| DL-NP 94 | Cerebral cortex: 94.1%, Hippocampus: 2.6%, Amygdala: 2.0% | Human MTG: 18.0%, Human TF: 6.6%, Human V2: 6.2% | IFNG-AS1, CD200R1L, LINC02218, NPSR1-AS1, LINC02042, CCDC80, IL26, BARX2, CD200R1L-AS1, ERG | 2547 | 4 |
| LLC-IN 274 | Cerebral cortex: 84.6%, Basal forebrain: 5.9%, Hippocampus: 3.1% | Human M1C: 11.7%, Human MTG: 7.6%, Human Cla: 4.8% | LINC01344, AC137770.1, SFRP1, AC132803.1, HAPLN1, EYA4, PDGFD, ANO1, AP002989.1, LINC00298 | 6373 | 4 |
| Splatter 238 | Basal forebrain: 57.1%, Amygdala: 27.7%, Cerebral cortex: 6.8% | Human CaB: 15.6%, Human NAC: 12.7%, Human SI: 10.8% | CRABP1, KLB, AP002989.1, HTR3A, PLSCR5, PDGFD, HAPLN1, EXOC1L, IL1RAPL2, AL356108.1 | 7651 | 4 |
| Thal-Exc 453 | Thalamus: 82.7%, Midbrain: 17.3%, Cerebral cortex: 0.0% | Human LG: 29.5%, Human MG: 24.7%, Human PTR: 14.4% | CARTPT, CASQ2, ST8SIA6-AS1, PLEKHG4B, RGS16, LEF1, PVALB, PPP1R17, CYP27C1, LINC00868 | 5556 | 3 |
| CGE-IN 285 | Cerebral cortex: 62.9%, Hippocampus: 22.3%, Amygdala: 9.5% | Human M1C: 7.7%, Human CA1C-CA3C: 5.2%, Human MEC: 4.2% | PTGFR, PENK, PCDH18, LINC01497, NR2E1, OSTM1-AS1, VIP, CXCL14, AC008798.3, HTR3A | 4688 | 4 |
| Splatter 338 | Medulla: 99.1%, Basal forebrain: 0.6%, Pons: 0.3% | Human MoAN: 53.4%, Human MoSR: 34.0%, Human MoRF-MoEN: 11.7% | LINC02058, AC004080.1, FSHR, HOXD3, ASAH2, EBF1, PPP1R1C, HOXB3, IGFBP7, LNCTAM34A | 350 | 3 |
| L RhombLip 320 | Pons: 90.4%, Medulla: 6.2%, Basal forebrain: 3.2% | Human PN: 27.7%, Human PnEN: 25.8%, Human PnRF: 21.5% | AL139351.3, AC093765.2, IRX2, LINC00355, LINC02476, AC136431.1, BMP5, TINAG, HOXC4, LINC00363 | 27800 | 3 |
| Splatter 360 | Midbrain: 99.1%, Pons: 0.8%, Thalamus: 0.1% | Human SC: 87.1%, Human IC: 10.6%, Human PTR: 0.6% | IGFL1, AP000146.1, LINC01692, OTX2, GATA3, IRX2, DMBX1, COL6A2, IRX3, FST | 1215 | 3 |

| Cluster (Yao naming) | Top three regions | Top three dissections | Top Enriched Genes | Number of cells | Number of donors |
|---|---|---|---|---|---|
| Splatter 370 | Midbrain: 98.8%, Pons: 0.5%, Thalamus: 0.5% | Human SC: 76.1%, Human IC: 11.3%, Human PTR: 7.9% | EBF2, TFAP2B, EBF3, EBF1, LHX9, POU4F1, HSPD1P8, LEF1, TFAP2D, LGR5 | 3622 | 4 |
| CGE-IN 281 | Cerebral cortex: 70.5%, Amygdala: 13.0%, Hippocampus: 10.6% | Human Pir: 5.7%, Human MEC: 4.6%, Human M1C: 4.5% | HTR3A, AC004862.1, AL139090.1, HGFAC, AL391832.4, CHRNA6, EDNRA, AC006305.1, CHRNB3, TNFAIP8L3 | 2234 | 4 |
| Splatter 431 | Hypothalamus: 99.7%, Midbrain: 0.2%, Cerebral cortex: 0.0% | Human MN: 57.0%, Human HTHma-HTHtub: 14.2%, Human HTHso-HTHtub: 13.8% | PGR, TBX3, PGR-AS1, AC004870.4, AC026765.3, CALCR, POMC, NR5A2, ESR1, C14orf39 | 2691 | 3 |
| Thal-Exc 458 | Thalamus: 93.6%, Midbrain: 6.4%, Amygdala: 0.0% | Human ETH: 22.6%, Human MD: 19.4%, Human ANC: 18.3% | SPTLC3, NTS, COL6A5, AC105213.1, AL033530.1, CCDC141, CYP27C1, LINC02775, LHX9, ADGRG6 | 7494 | 3 |
| Thal-Exc 451 | Thalamus: 99.9%, Midbrain: 0.1%, Hypothalamus: 0.0% | Human MD: 29.1%, Human ETH: 20.1%, Human ANC: 18.7% | CASQ2, AC016766.1, LINC00868, RGS16, C2orf83, TGFBR2, CYP27C1, AC106874.1, AOX2P, SHOX2 | 9740 | 3 |
| Thal-Exc 445 | Thalamus: 99.3%, Hippocampus: 0.7%, Cerebral cortex: 0.0% | Human LG: 98.5%, Human MG: 0.7%, Human DGR-CA4Rpy: 0.7% | LINC01491, AL355612.1, DCDC2C, AC092078.3, LCP1, SHOX2, CHRNA2, FZD10-AS1, VEGFC, DEFB1 | 134 | 1 |
| MiG 12 | Basal forebrain: 29.4%, Cerebral cortex: 14.0%, Cerebellum: 12.2% | Human GPi: 11.5%, Human CbDN: 9.7%, Human SEP: 9.2% | MRC1, CD163, STAB1, MS4A6A, MS4A4A, F13A1, SIGLEC1, MS4A4E, FCGR2C, LILRB5 | 3344 | 4 |
| Splatter 399 | Medulla: 99.3%, Pons: 0.5%, Cerebral cortex: 0.1% | Human MoAN: 42.7%, Human MoRF-MoEN: 36.3%, Human MoSR: 20.3% | PHOX2B-AS1, PHOX2B, TFAP2B, C5orf58, NPFF, LINC00682, HOXB3, LMX1B, DBH, RP1 | 1007 | 3 |
| COP 37 | Cerebral cortex: 35.6%, Hippocampus: 19.5%, Amygdala: 8.2% | Human A35r: 6.3%, Human CA1: 4.8%, Human MTG: 4.5% | GPR17, AL512308.1, TNS3, BCAS1, MIR503HG, AC008080.4, NKX2-2, GPNMB, INAVA, FIBIN | 796 | 3 |
| Misc 0 | Midbrain: 21.0%, Basal forebrain: 19.0%, Pons: 14.3% | Human GPi: 11.4%, Human SN: 11.4%, Human SN-RN: 4.8% | IGHM, MS4A1, FCRL1, AC244205.1, IGLL5, IGHA1, BLK, IGLC3, IGLC2, IKZF3 | 105 | 3 |
| Splatter 352 | Midbrain: 97.3%, Pons: 0.9%, Medulla: 0.8% | Human SC: 81.8%, Human IC: 10.4%, Human PAG-DR: 2.7% | LHX1, EBF1, EBF3, GATA3, TRAC, GATA3-AS1, PVALB, HAS2-AS1, HAS2, LHX1-DT | 862 | 4 |
| U RhombLip 312 | Medulla: 61.0%, Pons: 34.8%, Cerebellum: 3.6% | Human MoAN: 47.2%, Human PnAN: 33.2%, Human MoSR: 8.2% | TTR, CBLN3, AC073225.1, AC092957.1, MSR1, CRTAM, LINC01798, AL359208.1, AC044798.3, SLC22A31 | 5392 | 3 |
| Splatter 377 | Hypothalamus: 33.0%, Amygdala: 32.4%, Thalamus: 31.4% | Human BNST: 32.1%, Human ANC: 30.6%, Human HTHpo: 14.3% | MIR217HG, AGTR1, ARHGAP36, DLK1, ONECUT3, LHX5-AS1, C14orf39, TFPI, CALCR, ONECUT1 | 1964 | 3 |
| Splatter 356 | Midbrain: 67.5%, Pons: 20.1%, Thalamus: 9.5% | Human PTR: 30.3%, Human IC: 11.1%, Human SC: 8.1% | MECOM, COL8A1, GATA3, LHX5-AS1, AC005100.1, GATA3-AS1, LMO1, EBF3, GATA2, TFAP2A | 6063 | 4 |
| Splatter 357 | Midbrain: 97.4%, Pons: 1.6%, Thalamus: 0.9% | Human SC: 32.6%, Human PTR: 30.3%, Human PAG: 17.5% | TFAP2B, EMX2, LINC01497, PAX7, EMX2OS, FAM9B, GATA3, AC005208.1, LEF1, LHX5-AS1 | 2674 | 3 |
| DL-CT/6b 84 | Hippocampus: 98.5%, Amygdala: 0.8%, Cerebral cortex: 0.7% | Human CA1U-CA2U-CA3U: 20.1%, Human CA2U-CA3U: 19.3%, Human CA1-2R: 13.5% | PIWIL1, NPSR1-AS1, SLC9C1, IFNG-AS1, LINC02218, LINC01479, CPZ, ABCA12, LPO, C3orf52 | 4474 | 4 |
| LLC-IN 275 | Cerebral cortex: 66.0%, Amygdala: 16.5%, Basal forebrain: 9.9% | Human Cla: 7.5%, Human MEC: 6.9%, Human MTG: 5.9% | LINC01344, SFRP1, HAPLN1, PDGFD, EYA4, ADGRG6, ANO1, AC137770.1, AP002989.1, CRABP1 | 2174 | 4 |
| Fibr 24 | Basal forebrain: 38.0%, Medulla: 32.3%, Hippocampus: 25.1% | Human SEP: 38.0%, Human MoAN: 17.6%, Human CA1R-CA2R-CA3R: 16.8% | C7, CFH, IGF2, COL1A2, LINC02126, MGP, COL3A1, TBX18, FBLN1, TTR | 279 | 3 |
| CGE-IN 280 | Cerebral cortex: 73.7%, Amygdala: 13.3%, Hippocampus: 7.8% | Human MEC: 7.2%, Human Pir: 6.0%, Human LEC: 5.6% | AC006305.1, LINC01416, AL391832.4, LINC01905, SP8, DCN, CHRNB3, CXCL14, CHRNA7, CRH | 5106 | 4 |
| Splatter 384 | Hypothalamus: 43.2%, Midbrain: 39.3%, Pons: 9.4% | Human SN: 20.0%, Human SN-RN: 17.3%, Human HTHma-HTHtub: 17.0% | NTS, LMX1A, PITX2, LMX1B, CDH23, PANCR, GIPR, FOXA1, LINC02520, AL161908.1 | 3100 | 3 |
| CGE-IN 291 | Cerebral cortex: 75.1%, Hippocampus: 14.3%, Amygdala: 7.1% | Human MTG: 6.3%, Human M1C: 5.2%, Human MEC: 4.6% | Z83818.2, NPR3, KMO, AL391832.4, AL136114.1, EDNRA, SLC10A4, AC013265.1, PCDH18, TMEM100 | 12240 | 4 |
| HiDG 204 | Hippocampus: 98.3%, Thalamus: 1.7%, Cerebral cortex: 0.0% | Human DGR-CA4Rpy: 39.0%, Human CA1R-CA2R-CA3R: 32.3%, Human CA4C-DGC: 22.8% | TTR, LINC02030, LINC01697, LINC02237, AJ006995.1, CHRNA1, AC097487.1, AC097528.1, AC021088.1, CASC6 | 9290 | 3 |
| Thal-Exc 455 | Thalamus: 97.9%, Midbrain: 2.1%, Cerebral cortex: 0.0% | Human Pul: 40.5%, Human ETH: 38.2%, Human MD: 5.5% | AC037486.1, LINC01151, SMILR, MATN3, FSHR, COL6A5, CYP27C1, CLEC3A, ITIH5, AC012535.1 | 474 | 3 |
| Splatter 347 | Medulla: 68.6%, Spinal cord: 29.3%, Pons: 0.9% | Human MoAN: 33.4%, Human SpC: 29.3%, Human MoSR: 21.6% | PAX2, HOXB3, EBF3, NPPC, SLC6A5, HOXB7, AC024230.1, HOXB-AS3, PAX8, PDYN | 2434 | 3 |
| OPC 36 | Cerebral cortex: 33.7%, Thalamus: 12.5%, Midbrain: 11.5% | Human A35r: 20.8%, Human SpC: 8.0%, Human SN: 5.4% | AC004852.2, BX284613.2, PDGFRA, AL512308.1, VCAN, OLIG2, OLIG1, GPR17, CSPG4, FERMT1 | 1643 | 4 |
| Splatter 381 | Pons: 83.9%, Midbrain: 15.4%, Medulla: 0.5% | Human DTg: 34.1%, Human PB: 30.1%, Human PnEN: 16.2% | EBF2, OTP, PAX7, PAX3, SLC5A7, EBF1, PENK, LHX5-AS1, AP005242.1, AC124864.2 | 1544 | 3 |
| Splatter 411 | Hypothalamus: 99.2%, Amygdala: 0.2%, Midbrain: 0.2% | Human HTHma-HTHtub: 37.3%, Human HTHpo-HTHso: 36.7%, Human MN: 10.8% | PPP1R17, GABRE, ECEL1, LHX5-AS1, PRDM13, RERGL, RADX, KC877982.1, PIK3C2G, SLITRK6 | 1665 | 4 |

| Cluster (Yao naming) | Top three regions | Top three dissections | Top Enriched Genes | Number of cells | Number of donors |
|---|---|---|---|---|---|
| Splatter 362 | Pons: 39.4%, Cerebellum: 34.7%, Midbrain: 21.7% | Human CbDN: 34.7%, Human DTg: 24.0%, Human PTR: 8.7% | LHX5-AS1, OTP, LHX1, EBF3, S100A10, ZPLD1, ECEL1, LINC02520, AL359915.1, TRPC7 | 2288 | 4 |
| Splatter 409 | Basal forebrain: 66.5%, Amygdala: 14.2%, Thalamus: 9.9% | Human CaB: 17.8%, Human SI: 13.0%, Human NAC: 11.5% | GLP1R, CRABP1, CER1, TAC3, AL096799.1, TRH, LINC02203, NANOS1, NPR3, CHRNA3 | 6395 | 4 |
| Splatter 361 | Midbrain: 83.4%, Thalamus: 14.3%, Pons: 1.8% | Human PTR: 49.6%, Human PAG: 14.9%, Human ETH: 8.9% | LINC01497, GATA3, AC005208.1, KCNJ16, GATA2, AC074099.1, LINC01876, LHX5-AS1, SIX3, LHX1 | 1067 | 3 |
| MiG 8 | Hippocampus: 19.2%, Midbrain: 18.4%, Basal forebrain: 11.1% | Human SN: 8.2%, Human CBV: 4.6%, Human CA4C-DGC: 4.6% | CCDC26, LNCAROD, FYB1, CSF1R, APBB1IP, CX3CR1, SYK, AL392172.1, DOCK8, C3 | 7750 | 3 |
| Splatter 350 | Medulla: 98.2%, Spinal cord: 1.0%, Pons: 0.7% | Human MoRF-MoEN: 48.6%, Human MoAN: 35.1%, Human MoSR: 14.5% | SLC6A5, HOXB3, SKOR2, TFAP2A, LBX1-AS1, CRABP1, HOXA3, MAB21L2, HOXB2, HOXD3 | 3486 | 3 |
| Fibr 26 | Thalamus: 20.7%, Basal forebrain: 15.4%, Pons: 13.1% | Human SEP: 6.0%, Human CBV: 5.7%, Human SN: 4.6% | COL1A2, TBX18, FMO2, IGF2, ITIH5, C7, FBLN1, FLVCR2, SLC6A20, COL15A1 | 1340 | 4 |
| MiG 4 | Spinal cord: 31.8%, Pons: 26.0%, Medulla: 13.2% | Human SpC: 31.8%, Human MN: 11.5%, Human PnEN: 11.4% | SRGN, RGS1, GPR183, CD69, HLA-DRA, OLR1, TNFRSF1B, IFI30, CXCR4, CD74 | 745 | 3 |
| Splatter 414 | Hypothalamus: 90.6%, Midbrain: 8.5%, Thalamus: 0.8% | Human HTHma-HTHtub: 43.2%, Human MN: 14.9%, Human HTHma: 13.8% | SIM1, OTP, METTL11B, EBF3, LINC02505, AL110292.1, STK32B, LHX5-AS1, AL132857.1, AL158013.1 | 2915 | 3 |
| Epend 74 | Basal forebrain: 68.3%, Thalamus: 13.3%, Medulla: 7.3% | Human SEP: 67.8%, Human DGR-CA4Rpy: 6.0%, Human MD: 4.3% | DTHD1, ADGB, AC020718.1, AL672167.1, AC013470.2, CFAP157, AC093689.1, CROCC2, TTC29, RASSF9 | 795 | 3 |
| MiG 10 | Basal forebrain: 27.7%, Midbrain: 22.1%, Cerebral cortex: 10.3% | Human GPi: 17.4%, Human SN: 9.3%, Human GPe: 7.9% | LNCAROD, APBB1IP, CX3CR1, FYB1, CSF1R, CD74, DOCK8, C3, SYK, PTPRC | 16707 | 4 |
| Misc 2 | Thalamus: 17.6%, Cerebral cortex: 15.5%, Basal forebrain: 14.8% | Human GPe: 5.7%, Human GPi: 5.5%, Human CbDN: 5.1% | NKG7, GNLY, PRF1, KLRF1, PYHIN1, CCL4, RUNX3, CST7, GZMA, GZMB | 671 | 3 |
| Vasc 13 | Basal forebrain: 74.0%, Medulla: 23.3%, Hippocampus: 1.4% | Human SEP: 74.0%, Human MoSR: 11.0%, Human MoAN: 8.2% | FLT1, VWF, PLVAP, KDR, CFH, SERPINH1, CLEC1A, PKHD1L1, ANO2, TGM2 | 73 | 3 |
| Splatter 380 | Pons: 62.5%, Medulla: 34.7%, Spinal cord: 1.1% | Human PnEN: 23.4%, Human MoRF-MoEN: 17.0%, Human PB: 15.8% | SLC6A5, PAX2, EBF3, OTP, TFAP2A, HOXB3, LINC01297, LBX1-AS1, LHX5-AS1, EBF2 | 4748 | 4 |
| Cer-In 298 | Pons: 82.7%, Cerebellum: 11.0%, Medulla: 4.1% | Human PnEN: 27.0%, Human PB: 21.4%, Human PnAN: 18.2% | SLC6A5, PAX2, TRPM8, AL357507.1, LINC00410, TFAP2B, AC080079.2, TFAP2A, IGFBP7-AS1, CHAT | 318 | 4 |
| Epend 68 | Medulla: 29.8%, Pons: 23.1%, Thalamus: 13.7% | Human MoRF-MoEN: 17.9%, Human PnEN: 12.2%, Human SEP: 11.5% | FAM183A, C11orf88, RASSF9, DTHD1, ODF3B, FOXJ1, CFAP157, ADGB, AC013470.2, CAPSL | 459 | 3 |
| Splatter 335 | Medulla: 78.3%, Pons: 15.9%, Midbrain: 4.8% | Human IO: 70.8%, Human PnRF: 13.5%, Human MoSR: 3.2% | HSPD1P8, AC092445.1, LINC00331, POU4F1, TES, HOXD3, AC096773.1, PDLIM1, AC093772.1, LINC00581 | 993 | 3 |
| Vasc 14 | Hippocampus: 23.8%, Cerebral cortex: 23.3%, Thalamus: 10.3% | Human A35r: 14.0%, Human CA1: 8.7%, Human CBV: 5.5% | CLDN5, FLT1, ABCG2, MECOM, CD34, SMAD6, SOX18, EPAS1, SEMA3G, HLA-E | 748 | 4 |
| Vasc 18 | Cerebral cortex: 23.3%, Medulla: 16.4%, Thalamus: 14.6% | Human A35r: 10.4%, Human IO: 7.3%, Human MN: 6.9% | MECOM, CLDN5, MGP, FLT1, HLA-E, TM4SF1, SMAD6, EPAS1, KLF2, CXCL2 | 451 | 3 |
| Splatter 342 | Cerebellum: 97.0%, Pons: 1.8%, Midbrain: 0.8% | Human CbDN: 97.0%, Human PnAN: 1.0%, Human PB: 0.6% | ASAH2, VENTXP1, AF130359.1, LTBP2, LINC01036, AC090993.1, AC007614.1, LINC01524, DIAPH3, AC087855.2 | 868 | 3 |
| Splatter 416 | Hypothalamus: 65.4%, Thalamus: 17.9%, Amygdala: 8.1% | Human HTHso-HTHtub: 19.1%, Human ANC: 17.7%, Human HTHpo: 16.0% | GAL, HMX3, ECEL1, ISL1, ARHGAP36, DLK1, HMX2, DIRAS3, AC010478.1, C14orf39 | 3344 | 3 |
| Misc 1 | Midbrain: 15.0%, Basal forebrain: 14.0%, Pons: 13.2% | Human SpC: 8.2%, Human CbDN: 7.6%, Human SN: 7.4% | CD2, IL7R, PTPRC, SLFN12L, IL32, CCL5, GRAP2, RUNX3, CD69, CD3E | 2474 | 4 |
| COP 38 | Hippocampus: 29.9%, Cerebral cortex: 29.5%, Pons: 7.8% | Human CA1: 8.9%, Human A35r: 6.5%, Human PnEN: 5.6% | GPR17, AC008080.4, FRG2DP, AL512308.1, MDFI, LINC01447, TNS3, BCAS1, ADAM33, MIR503HG | 779 | 4 |
| Epend 71 | Basal forebrain: 41.0%, Hypothalamus: 26.7%, Thalamus: 14.5% | Human CaB: 20.4%, Human SEP: 17.0%, Human BNST: 12.2% | DTHD1, ADGB, AC013470.2, AL672167.1, CFAP157, AC093689.1, CROCC2, TTC29, RASSF9, LMNTD1 | 476 | 3 |
| CA4 194 | Hippocampus: 99.8%, Cerebral cortex: 0.2%, Amygdala: 0.0% | Human CA4C-DGC: 90.0%, Human DGR-CA4Rpy: 8.0%, Human CA3R: 1.5% | SCGN, MORC1, AL450311.1, LINC02237, ITGA4, CASC6, ZBTB20-AS2, AC127526.1, ZBTB20-AS1, LINC02192 | 551 | 3 |
| MiG 7 | Basal forebrain: 32.0%, Midbrain: 19.4%, Pons: 12.5% | Human GPi: 20.8%, Human SN: 10.3%, Human GPe: 9.1% | CD74, CX3CR1, APBB1IP, HLA-DRA, LNCAROD, C3, ITGAX, FYB1, DOCK8, PTPRC | 34829 | 4 |
| Misc 401 | Hippocampus: 76.2%, Amygdala: 19.4%, Cerebral cortex: 4.4% | Human CA1U-CA2U-CA3U: 73.9%, Human BL: 8.9%, Human CMN: 7.1% | FST, SOX11, AC087516.2, BHLHE22, AL138826.1, LINC01163, SOX4, AC012409.2, AC112236.1, GPC2 | 2038 | 3 |
| Epend 67 | Medulla: 52.7%, Spinal cord: 22.3%, Pons: 17.8% | Human SpC: 22.3%, Human MoRF-MoEN: 21.5%, Human MoAN: 18.7% | DTHD1, ADGB, RASSF9, CFAP157, AC013470.2, AC093689.1, AL672167.1, TTC29, LRRC77P, ODF3B | 651 | 3 |

| Cluster (Yao naming) | Top three regions | Top three dissections | Top Enriched Genes | Number of cells | Number of donors |
|---|---|---|---|---|---|
| MiG 5 | Hippocampus: 40.4%, Medulla: 29.6%, Basal forebrain: 20.1% | Human MoAN: 26.6%, Human DGR-CA4Rpy: 25.2%, Human SEP: 18.8% | HLA-DRA, TTR, CD74, HLA-DRB1, SRGN, RGS1, HLA-DPA1, MS4A6A, HLA-DQA1, HLA-DRB5 | 777 | 3 |
| Splatter 339 | Pons: 99.4%, Medulla: 0.4%, Midbrain: 0.2% | Human DTg: 39.1%, Human PnRF: 35.2%, Human PB: 16.6% | SLC6A5, GLRA1, LINC01956, LGI2, EN1, LINC01798, ADGRD2, ASAH2, HOXB-AS1, MAB21L2 | 489 | 3 |
| Misc 314 | Medulla: 35.7%, Pons: 33.3%, Cerebellum: 26.2% | Human CBV: 23.8%, Human PnAN: 21.4%, Human MoAN: 16.7% | PCP2, TRABD2B, ZNF385C, AL109659.1, ATP2A3, LINC02794, IL20RA, NCAM1-AS1, AC092448.1, GNG13 | 42 | 3 |
| MiG 6 | Hippocampus: 25.1%, Basal forebrain: 15.2%, Thalamus: 11.0% | Human CA1: 7.0%, Human GPi: 6.9%, Human CA4C-DGC: 6.5% | LNCAROD, ITGAX, CD74, LINC00278, FYB1, APBB1IP, CD84, PIK3R5, HLA-DRA, C1QC | 2485 | 3 |
| Splatter 344 | Medulla: 61.0%, Spinal cord: 21.9%, Pons: 14.9% | Human MoAN: 29.1%, Human SpC: 21.9%, Human MoRF-MoEN: 17.3% | LMX1B, EBF3, EBF1, PHOX2B-AS1, HOXB3, AC009411.2, EBF2, AC018742.1, PHOX2B, TLX3 | 3736 | 3 |
| COP 39 | Cerebral cortex: 28.1%, Hippocampus: 19.7%, Amygdala: 14.6% | Human CA1: 7.3%, Human A35r: 5.6%, Human Pir: 3.9% | AC008080.4, GPR17, AC020909.2, APCDD1, ACAN, DCT, AC022433.1, KANK2, FRG2DP, DOCK6 | 178 | 3 |
| Vasc 16 | Cerebral cortex: 25.5%, Hippocampus: 17.8%, Midbrain: 8.4% | Human A35r: 14.8%, Human CA1: 5.6%, Human SpC: 4.1% | CLDN5, MECOM, VWF, FLT1, ABCG2, FLI1, NOSTRIN, LINC02147, TGM2, CFH | 1046 | 4 |
| Vasc 15 | Cerebral cortex: 23.2%, Hippocampus: 15.7%, Thalamus: 15.3% | Human A35r: 8.6%, Human CBV: 6.6%, Human CA1: 4.8% | CLDN5, ABCG2, FLT1, MECOM, IFI27, TM4SF18, HLA-E, CAVIN2, FLI1, SLC38A5 | 2746 | 4 |
| COP 42 | Thalamus: 92.7%, Midbrain: 2.3%, Pons: 1.5% | Human VLN: 69.8%, Human ANC: 6.2%, Human LG: 5.0% | CNDP1, LINC01608, GREM1, OPALIN, FA2H, AC110285.1, MYRF, CD22, LINC01792, LINC00906 | 1170 | 3 |
| Splatter 397 | Pons: 80.1%, Medulla: 8.1%, Midbrain: 7.6% | Human PnRF: 26.5%, Human PnEN: 26.2%, Human DTg: 23.2% | TPH2, SLC6A4, FEV, SLC18A2, LMX1B, DDC, GATA3, NDNF, VWA5B1, AMIGO2 | 1316 | 3 |
| MiG 9 | Cerebral cortex: 27.2%, Midbrain: 16.6%, Hippocampus: 15.0% | Human SN-RN: 8.1%, Human GPi: 5.5%, Human SN: 5.4% | AIF1, HLA-DRA, CD74, C1QC, C1QB, CX3CR1, TYROBP, CSF1R, C3, APBB1IP | 5165 | 3 |
| Splatter 353 | Thalamus: 97.2%, Midbrain: 2.2%, Hypothalamus: 0.5% | Human LG: 93.9%, Human VLN: 1.4%, Human CM-Pf: 1.1% | GATA3, FABP4, HPGD, LHX5-AS1, NPY, GATA3-AS1, LINC00237, AC096751.2, AL138899.1, NDNF | 852 | 3 |
| Splatter 398 | Medulla: 86.6%, Pons: 12.8%, Midbrain: 0.2% | Human MoRF-MoEN: 40.5%, Human MoAN: 37.3%, Human MoSR: 8.9% | PHOX2B-AS1, PHOX2B, LINC00682, HOXB3, SLC5A7, SLC6A2, GAL, TH, PHOX2A, HOTAIRM1 | 2093 | 3 |
| Olig 40 | Hippocampus: 24.8%, Cerebral cortex: 23.4%, Amygdala: 10.0% | Human CA4C-DGC: 4.7%, Human DGR-CA4Rpy: 4.6%, Human CA1: 4.3% | OPALIN, CD9, AC020909.2, MAL, LINC00639, AC110285.1, PLLP, LINC01170, MAG, SLCO1A2 | 4913 | 4 |
| Olig 46 | Thalamus: 22.7%, Midbrain: 19.9%, Cerebral cortex: 13.7% | Human SN-RN: 9.7%, Human SN: 6.7%, Human CbDN: 5.9% | CNDP1, LINC01608, OPALIN, RNASE1, LINC01792, NKX6-2, MAG, PPP1R14A, CD9, TMEM98 | 60616 | 4 |
| Splatter 379 | Pons: 99.2%, Medulla: 0.8%, Cerebral cortex: 0.0% | Human PnEN: 45.0%, Human DTg: 31.7%, Human PnRF: 17.5% | OTP, SLC6A5, EBF2, GLI2, MAB21L2, EBF1, GPR101, AC022034.3, HEPACAM2, NPBWR1 | 120 | 3 |
| Vasc 20 | Thalamus: 21.8%, Cerebral cortex: 13.7%, Medulla: 12.4% | Human MN: 10.5%, Human IO: 7.0%, Human Pul: 4.0% | MYL9, CARMN, ADIRF, FHL5, ACTA2, VIM, TBX18, MUSTN1, TBX2, NOTCH3 | 845 | 4 |
| CPlx 78 | Basal forebrain: 92.5%, Medulla: 6.3%, Thalamus: 0.6% | Human SEP: 92.5%, Human MoAN: 4.9%, Human MoSR: 1.2% | AC004052.1, TRPV4, AC016152.1, LINC00880, CLIC6, NWD1, AP003500.1, LMX1A, ABCA4, SLC39A12 | 2653 | 3 |
| Epend 65 | Midbrain: 28.4%, Hippocampus: 12.5%, Pons: 12.5% | Human PnEN: 10.2%, Human PAG-DR: 9.1%, Human PAG: 9.1% | DTHD1, ADGB, AC093689.1, CFAP157, AC013470.2, LMNTD1, RASSF9, TTC29, AL672167.1, MAP3K19 | 88 | 3 |
| Epend 70 | Thalamus: 41.0%, Amygdala: 21.3%, Midbrain: 17.1% | Human MD: 22.3%, Human BNST: 21.0%, Human MD-Re: 7.1% | DTHD1, ADGB, AC020718.1, AL672167.1, AC013470.2, CFAP157, AC093689.1, CROCC2, RASSF9, TTC29 | 310 | 3 |
| CPlx 80 | Basal forebrain: 94.8%, Medulla: 4.5%, Hippocampus: 0.4% | Human SEP: 94.8%, Human MoAN: 3.0%, Human MoSR: 0.9% | AC016152.1, TRPV4, AC004052.1, CLIC6, SLC5A5, ABCA4, LINC00880, NWD1, AP003500.1, LMX1A | 1347 | 3 |
| Vasc 23 | Thalamus: 48.8%, Cerebral cortex: 25.0%, Basal forebrain: 10.7% | Human Pul: 31.0%, Human STG: 7.1%, Human LP: 6.0% | ITIH5, SLC6A12, SLC6A13, ATP1A2, DCN, PRELP, COLEC12, IFITM2, MUSTN1, TFPI | 84 | 3 |
| Epend 72 | Hippocampus: 53.2%, Amygdala: 20.2%, Basal forebrain: 7.9% | Human CA1: 38.5%, Human Sub: 11.1%, Human BNST: 7.2% | DTHD1, ADGB, AC020718.1, CFAP157, AL672167.1, AC093689.1, AC013470.2, LMNTD1, TTC29, CROCC2 | 1027 | 3 |
| CPlx 82 | Basal forebrain: 76.6%, Medulla: 22.2%, Pons: 1.2% | Human SEP: 76.6%, Human MoAN: 11.7%, Human MoSR: 10.5% | TRPV4, CLIC6, AC012078.2, LINC00113, LMX1A, SLC28A3, SLC5A5, FBP2, LINC00880, RBM47 | 171 | 3 |
| Splatter 337 | Basal forebrain: 81.9%, Midbrain: 17.6%, Hypothalamus: 0.5% | Human GPi: 53.9%, Human GPe: 26.4%, Human SN: 16.1% | ONECUT1, ASAH2, AC016044.1, LHX1-DT, KANK4, LINC00922, LINC02520, ZNF676, LHX1, MARCHF11-AS1 | 193 | 3 |
| Splatter 351 | Midbrain: 90.0%, Thalamus: 8.2%, Hypothalamus: 1.3% | Human SN-RN: 27.9%, Human PAG: 19.7%, Human PTR: 13.1% | LINC00261, FOXA2, TM4SF18, NDNF, PAX5, LINC01956, EMX2OS, ASAH2, EBF3, EMX2 | 559 | 3 |
| Olig 48 | Basal forebrain: 47.8%, Thalamus: 42.2%, Hypothalamus: 5.3% | Human GPe: 47.6%, Human CM-Pf: 36.8%, Human MN: 5.3% | AC061958.1, PDE4B-AS1, AL589787.1, LINC01608, MUSK, AC103831.1, AC026746.1, OPALIN, AC093330.1, SPATA22 | 2190 | 3 |

| Cluster (Yao naming) | Top three regions | Top three dissections | Top Enriched Genes | Number of cells | Number of donors |
|---|---|---|---|---|---|
| Epend 66 | Medulla: 83.1%, Pons: 16.0%, Midbrain: 0.4% | Human MoAN: 42.7%, Human MoSR: 22.2%, Human MoRF-MoEN: 18.2% | DTHD1, ADGB, RASSF9, AC020718.1, CFAP157, AC013470.2, ODF3B, FAM183A, C11orf88, AC093689.1 | 445 | 3 |
| Epend 73 | Hippocampus: 85.9%, Amygdala: 8.2%, Basal forebrain: 1.9% | Human CA1: 69.3%, Human BL: 5.4%, Human DGU-CA4Upy: 3.0% | DTHD1, ADGB, AC020718.1, CFAP157, AL672167.1, RASSF9, AC093689.1, AC019330.1, AC013470.2, CROCC2 | 907 | 3 |
| Olig 47 | Thalamus: 24.9%, Cerebral cortex: 20.9%, Hippocampus: 13.8% | Human GPe: 5.4%, Human STH: 4.8%, Human MTG: 4.2% | OPALIN, LINC01608, CD9, LINC01792, RNASE1, CNDP1, AC110285.1, PLLP, NKX6-2, MAG | 124837 | 4 |
| CPlx 77 | Basal forebrain: 49.7%, Medulla: 46.2%, Hippocampus: 3.0% | Human SEP: 49.7%, Human MoAN: 33.9%, Human MoSR: 11.8% | TTR, CLIC6, TRPV4, AC016152.1, AC004052.1, LINC00880, ABCA4, SLC39A12, AC104809.2, C1QTNF5 | 1659 | 3 |
| COP 41 | Amygdala: 24.7%, Hypothalamus: 23.3%, Basal forebrain: 20.2% | Human HTHso: 18.1%, Human La: 17.3%, Human GPe: 16.1% | OPALIN, LINC01608, CD22, CDH1, FA2H, LINC01099, LINC01792, MYRF, LINC00639, AC110285.1 | 1310 | 4 |
| Fibr 29 | Medulla: 41.3%, Spinal cord: 39.7%, Thalamus: 4.8% | Human SpC: 39.7%, Human IO: 25.4%, Human MoRF-MoEN: 7.9% | IGFBP6, VIM, BNC2, FLVCR2, IGF2, COL1A2, CFH, EBF2, CD177, KLF2 | 63 | 3 |
| Fibr 28 | Thalamus: 36.4%, Cerebral cortex: 12.1%, Pons: 11.0% | Human LG: 7.9%, Human Pul: 6.0%, Human LP: 4.6% | COL1A2, TBX18, C7, ITIH5, IGF2, FMO2, ISLR, LINC02147, FBLN1, DCN | 3767 | 4 |
| CPlx 79 | Basal forebrain: 94.0%, Medulla: 5.0%, Hippocampus: 0.5% | Human SEP: 94.0%, Human MoAN: 2.9%, Human MoSR: 1.6% | AC004052.1, TRPV4, AC016152.1, LINC00880, CLIC6, ABCA4, SLC5A5, NWD1, AP003500.1, LMX1A | 383 | 3 |
| COP 75 | Medulla: 38.6%, Basal forebrain: 32.9%, Thalamus: 15.7% | Human SEP: 32.9%, Human MoSR: 21.4%, Human Pul: 12.9% | TTR, FOLR1, SOD3, IGFBP7, FBLN1, C1QTNF5, FBP2, ECRG4, CLIC6, SLC39A12 | 70 | 3 |
| Splatter 390 | Thalamus: 81.4%, Amygdala: 5.6%, Pons: 5.3% | Human LG: 42.0%, Human ANC: 34.7%, Human BNST: 5.4% | ECEL1, SST, LINC00540, PAX6, DLK1, ARHGAP36, EGFL6, GPC3, LHX1, SIX3 | 3366 | 3 |
| Vasc 19 | Thalamus: 28.4%, Midbrain: 12.2%, Medulla: 11.8% | Human ANC: 7.4%, Human SN-RN: 5.7%, Human Pul: 5.7% | MYL9, CARMN, CCL19, IFITM2, FHL5, ADIRF, CLDN5, IFITM3, NOTCH3, CFH | 229 | 4 |
| Vasc 17 | Hypothalamus: 26.7%, Medulla: 21.8%, Pons: 15.8% | Human MN: 13.9%, Human IO: 9.9%, Human HTHso: 8.9% | IL1R1, VCAM1, CLDN5, VWF, CLEC2B, ICAM1, SERPINE1, ZFP36, PLA1A, MECOM | 101 | 3 |
| CPlx 81 | Basal forebrain: 73.7%, Medulla: 25.6%, Pons: 0.6% | Human SEP: 73.7%, Human MoAN: 20.9%, Human MoSR: 3.9% | AC004052.1, TRPV4, AC016152.1, SLC5A5, CLIC6, ABCA4, LINC00880, NWD1, LMX1A, RBM47 | 1375 | 3 |
| Fibr 27 | Thalamus: 34.3%, Cerebral cortex: 15.0%, Pons: 13.3% | Human CM-Pf: 8.7%, Human LG: 4.4%, Human VLN: 4.3% | TBX18, COL1A2, FMO2, ITIH5, LINC02147, COL15A1, FLVCR2, COLEC12, C7, LINC02058 | 2056 | 4 |
| Epend 69 | Hippocampus: 69.2%, Basal forebrain: 11.2%, Amygdala: 7.6% | Human CA1: 17.4%, Human CA1U-CA2U-CA3U: 16.7%, Human CA4C-DGC: 11.0% | DTHD1, CFAP157, RASSF9, ADGB, FAM183A, C11orf88, AC013470.2, AC020718.1, FOXJ1, AC093689.1 | 724 | 3 |
| Olig 49 | Pons: 30.3%, Midbrain: 20.2%, Medulla: 12.2% | Human PnRF: 10.0%, Human SN: 7.6%, Human CbDN: 7.3% | MBOAT1, AC110809.1, AC008945.2, S100A1, AC114485.1, AC007106.2, LINC01445, LINC01549, AC004690.2, S100B | 84282 | 4 |
| Fibr 25 | Thalamus: 22.3%, Cerebral cortex: 20.0%, Pons: 15.0% | Human MN: 8.3%, Human A35r: 8.2%, Human CM-Pf: 6.6% | COL1A2, TBX18, CFH, IGF2, COL3A1, ITIH5, FLVCR2, FMO2, LINC02147, BMP5 | 709 | 4 |
| Olig 45 | Cerebral cortex: 20.0%, Basal forebrain: 18.5%, Midbrain: 15.8% | Human GPe: 8.4%, Human SN: 7.4%, Human SN-RN: 4.8% | LINC01608, OPALIN, LINC01792, SLCO1A2, LINC02882, FOLH1, MAL, ASPA, AC108721.2, SH3TC2 | 109325 | 4 |
| COP 43 | Thalamus: 36.2%, Hypothalamus: 18.0%, Amygdala: 10.3% | Human VLN: 24.5%, Human HTHso: 11.5%, Human La: 6.0% | SLC5A11, FA2H, AL033523.1, CD22, AC012494.1, MOBP, MYRF, SH3TC2-DT, PRR5L, CNDP1 | 417 | 4 |
| Splatter 336 | Midbrain: 91.5%, Thalamus: 8.1%, Medulla: 0.3% | Human SN-RN: 59.7%, Human PAG: 18.3%, Human CM-Pf: 7.7% | NKX6-1, FAM83B, ASAH2, LINC00261, EBF3, SIM1, ONECUT1, FOXA2, AC024230.1, AC079148.3 | 1263 | 3 |
| CPlx 76 | Basal forebrain: 55.4%, Medulla: 31.7%, Thalamus: 8.9% | Human SEP: 55.4%, Human MoSR: 23.8%, Human Pul: 7.9% | TTR, AC004052.1, CLIC6, AC016152.1, LINC00880, TRPV4, LMX1A, NWD1, C1QTNF5, ABCA4 | 101 | 4 |
| Vasc 21 | Thalamus: 28.3%, Pons: 17.9%, Basal forebrain: 9.8% | Human LG: 8.0%, Human Pul: 7.0%, Human PnEN: 6.7% | IFITM2, CARMN, MUSTN1, NOTCH3, GGT5, IFITM3, SLC6A12, MYL9, BGN, ITIH5 | 1753 | 4 |
| Vasc 22 | Thalamus: 34.2%, Cerebral cortex: 15.9%, Basal forebrain: 10.2% | Human CBV: 6.7%, Human CM: 5.4%, Human LP: 5.3% | CARMN, NOTCH3, SLC6A12, ITIH5, GGT5, SLC12A7, IFITM2, LINC02147, CYSLTR2, BGN | 1856 | 4 |
| Olig 44 | Thalamus: 20.9%, Pons: 19.2%, Midbrain: 19.1% | Human SN: 10.9%, Human PnRF: 6.9%, Human CbDN: 5.8% | AC004690.2, SLC5A11, ITGA2, AL033523.1, LINC02882, MOG, FOLH1, CARNS1, LINC01949, LRP2 | 101039 | 4 |
| Olig 50 | Hypothalamus: 90.7%, Thalamus: 3.6%, Basal forebrain: 3.6% | Human MN: 90.7%, Human GPe: 3.6%, Human CM-Pf: 3.5% | PTCSC3, AC012494.2, AL591115.1, AC103831.1, AC093330.1, AL589787.1, AC061958.1, PDE4B-AS1, AC026746.1, AC108721.1 | 3044 | 3 |

*part 3 of 3 — first column repeated in every part*

| Cluster (Yao naming) | z (19 cohorts) | p (19 cohorts) | FDR (19 cohorts) | z (MVP diagnostic code) | p (MVP diagnostic code) | FDR (MVP diagnostic code) | z (MVP self-report) | p (MVP self-report) | FDR (MVP self-report) |
|---|---|---|---|---|---|---|---|---|---|
| Amyg-Exc 161 | 4.823 | 7.09e-07 | 0.0003267 | 2.555 | 0.005306 | 0.08434 | 1.984 | 0.02361 | 0.1512 |
| Misc 164 | 3.587 | 0.000167 | 0.03849 | 1.201 | 0.1148 | 0.3082 | 1.667 | 0.04773 | 0.2076 |
| Amyg-Exc 156 | 3.334 | 0.000428 | 0.06346 | 0.3427 | 0.3659 | 0.6448 | 1.417 | 0.07817 | 0.2592 |
| HiDG 203 | 3.23 | 0.0006183 | 0.06346 | 1.635 | 0.05107 | 0.1946 | 1.443 | 0.07453 | 0.2583 |
| MGE-IN 259 | 3.2 | 0.0006883 | 0.06346 | 2.014 | 0.02202 | 0.1253 | 2.634 | 0.004224 | 0.07788 |
| Amyg-Exc 155 | 3.132 | 0.0008689 | 0.06676 | 1.211 | 0.1129 | 0.308 | 1.842 | 0.03277 | 0.1737 |
| Amyg-Exc 153 | 3.033 | 0.001212 | 0.07982 | 2.208 | 0.01362 | 0.1072 | 2.784 | 0.002684 | 0.07788 |
| Amyg-Exc 159 | 2.965 | 0.001512 | 0.0871 | 1.496 | 0.06738 | 0.2269 | 2.026 | 0.02139 | 0.1497 |
| MGE-IN 236 | 2.927 | 0.00171 | 0.08757 | 2.064 | 0.01949 | 0.1185 | 2.682 | 0.003654 | 0.07788 |
| Misc 168 | 2.84 | 0.002253 | 0.1038 | 1.627 | 0.05184 | 0.1957 | 1.925 | 0.02714 | 0.1625 |
| MB-In 439 | 2.749 | 0.00299 | 0.1197 | 1.274 | 0.1013 | 0.2865 | 1.203 | 0.1144 | 0.3171 |
| MGE-IN 260 | 2.664 | 0.00386 | 0.1197 | 2.262 | 0.01185 | 0.1045 | 3.023 | 0.001252 | 0.07788 |
| MB-In 440 | 2.642 | 0.00412 | 0.1197 | 1.495 | 0.06743 | 0.2269 | 1.917 | 0.02764 | 0.1633 |
| MB-In 438 | 2.636 | 0.004201 | 0.1197 | 1.681 | 0.04638 | 0.1883 | 1.126 | 0.1301 | 0.3329 |
| DL-IT 144 | 2.633 | 0.004232 | 0.1197 | 1.902 | 0.02862 | 0.1481 | 1.868 | 0.03088 | 0.1695 |
| LLC-IN 265 | 2.606 | 0.004575 | 0.1197 | 3.095 | 0.0009831 | 0.04532 | 2.057 | 0.01984 | 0.1479 |
| Amyg-Exc 173 | 2.581 | 0.004928 | 0.1197 | 1.249 | 0.1057 | 0.2954 | 1.934 | 0.02655 | 0.161 |
| UL-IT 120 | 2.574 | 0.005032 | 0.1197 | 1.529 | 0.06315 | 0.2189 | 1.814 | 0.0348 | 0.1788 |
| UL-IT 124 | 2.564 | 0.00517 | 0.1197 | 2.318 | 0.01024 | 0.1045 | 3.411 | 0.0003233 | 0.07788 |
| UL-IT 126 | 2.563 | 0.005194 | 0.1197 | 2.69 | 0.003571 | 0.07667 | 2.301 | 0.01071 | 0.1122 |
| Misc 404 | 2.514 | 0.005976 | 0.1197 | 4.399 | 5.44e-06 | 0.001255 | 2.647 | 0.004055 | 0.07788 |
| Misc 170 | 2.51 | 0.006035 | 0.1197 | 1.659 | 0.04854 | 0.1929 | 2.015 | 0.02195 | 0.1497 |
| MSN 216 | 2.485 | 0.006472 | 0.1197 | 0.4674 | 0.3201 | 0.5764 | 1.687 | 0.04578 | 0.2039 |
| MGE-IN 242 | 2.457 | 0.007001 | 0.1197 | 1.402 | 0.08047 | 0.2524 | 2.072 | 0.01911 | 0.1468 |
| Amyg-Exc 405 | 2.452 | 0.007101 | 0.1197 | 2.174 | 0.01484 | 0.1113 | 2.126 | 0.01674 | 0.1378 |
| DL-IT 136 | 2.451 | 0.00713 | 0.1197 | 4.41 | 5.17e-06 | 0.001255 | 3.158 | 0.0007946 | 0.07788 |
| eMSN 234 | 2.447 | 0.007198 | 0.1197 | 1.177 | 0.1197 | 0.3135 | 1.582 | 0.0568 | 0.2182 |
| MGE-IN 261 | 2.432 | 0.007505 | 0.1197 | 2.242 | 0.01247 | 0.1045 | 2.168 | 0.01509 | 0.1364 |
| MSN 214 | 2.431 | 0.007533 | 0.1197 | 0.02334 | 0.4907 | 0.7642 | 1.143 | 0.1266 | 0.3316 |
| LLC-IN 266 | 2.395 | 0.008303 | 0.1225 | 2.761 | 0.002884 | 0.07132 | 2.012 | 0.02213 | 0.1497 |
| eMSN 228 | 2.39 | 0.008426 | 0.1225 | 2.205 | 0.01372 | 0.1072 | 1.903 | 0.0285 | 0.1663 |
| MGE-IN 253 | 2.387 | 0.008503 | 0.1225 | 1.716 | 0.0431 | 0.1857 | 1.68 | 0.04645 | 0.2039 |
| MSN 208 | 2.366 | 0.008998 | 0.1257 | 0.965 | 0.1673 | 0.3934 | 2.769 | 0.002813 | 0.07788 |
| Misc 166 | 2.347 | 0.009454 | 0.1263 | 1.075 | 0.1411 | 0.3594 | 1.971 | 0.02437 | 0.1539 |
| CA1-3 3 | 2.328 | 0.00995 | 0.1263 | 2.766 | 0.002834 | 0.07132 | 0.4351 | 0.3317 | 0.5793 |
| Amyg-Exc 172 | 2.32 | 0.01018 | 0.1263 | 1.284 | 0.09961 | 0.2835 | 2.649 | 0.004034 | 0.07788 |
| MB-In 436 | 2.313 | 0.01035 | 0.1263 | 1.197 | 0.1156 | 0.3082 | 1.121 | 0.1312 | 0.3329 |
| Amyg-Exc 157 | 2.311 | 0.01041 | 0.1263 | 1.68 | 0.0465 | 0.1883 | 0.9868 | 0.1619 | 0.3735 |
| UL-IT 127 | 2.286 | 0.01112 | 0.1315 | 2.654 | 0.003982 | 0.07667 | 2.484 | 0.006492 | 0.09522 |
| Amyg-Exc 158 | 2.264 | 0.01178 | 0.1358 | 2.89 | 0.001926 | 0.06343 | 2.639 | 0.004156 | 0.07788 |
| MB-In 443 | 2.234 | 0.01275 | 0.1434 | 1.349 | 0.08864 | 0.2636 | 1.598 | 0.05501 | 0.2182 |
| MSN 212 | 2.211 | 0.01351 | 0.1478 | 1.461 | 0.07199 | 0.2354 | 1.74 | 0.04097 | 0.187 |
| MSN 211 | 2.201 | 0.01387 | 0.1478 | 0.5705 | 0.2842 | 0.537 | 1.538 | 0.06202 | 0.2287 |
| eMSN 231 | 2.184 | 0.01449 | 0.1478 | 1.829 | 0.03366 | 0.1634 | 2.452 | 0.007106 | 0.09522 |
| CA4 196 | 2.182 | 0.01457 | 0.1478 | 1.724 | 0.04232 | 0.1857 | -0.8402 | 0.7996 | 0.9209 |
| Misc 165 | 2.177 | 0.01475 | 0.1478 | 0.8179 | 0.2067 | 0.4474 | 1.021 | 0.1537 | 0.3647 |

| Cluster (Yao naming) | z (19 cohorts) | p (19 cohorts) | FDR (19 cohorts) | z (MVP diagnostic code) | p (MVP diagnostic code) | FDR (MVP diagnostic code) | z (MVP self-report) | p (MVP self-report) | FDR (MVP self-report) |
|---|---|---|---|---|---|---|---|---|---|
| DL-IT 146 | 2.155 | 0.01557 | 0.1527 | 3.278 | 0.0005219 | 0.03557 | 2.124 | 0.01684 | 0.1378 |
| Amyg-Exc 175 | 2.142 | 0.01609 | 0.1546 | 1.874 | 0.03045 | 0.1542 | 1.887 | 0.02959 | 0.1679 |
| MGE-IN 243 | 2.123 | 0.01686 | 0.1569 | 2.095 | 0.01806 | 0.1185 | 2.448 | 0.007177 | 0.09522 |
| MB-In 437 | 2.12 | 0.01701 | 0.1569 | 0.958 | 0.169 | 0.3954 | 1.508 | 0.06583 | 0.239 |
| Amyg-Exc 154 | 2.111 | 0.01738 | 0.1571 | 0.8274 | 0.204 | 0.4474 | 1.027 | 0.1523 | 0.3645 |
| CA1-3 3 | 2.094 | 0.01812 | 0.1606 | 0.09796 | 0.461 | 0.7436 | -0.2754 | 0.6085 | 0.8084 |
| Splatter 420 | 2.075 | 0.01902 | 0.1632 | -0.1392 | 0.5554 | 0.8127 | 2.817 | 0.002427 | 0.07788 |
| MGE-IN 249 | 2.07 | 0.01923 | 0.1632 | 1.315 | 0.09428 | 0.2734 | 1.315 | 0.09422 | 0.2915 |
| DL-IT 143 | 2.061 | 0.01966 | 0.1632 | 2.271 | 0.01156 | 0.1045 | 1.17 | 0.121 | 0.3242 |
| eMSN 226 | 2.056 | 0.01989 | 0.1632 | 0.454 | 0.3249 | 0.5828 | 0.5572 | 0.2887 | 0.5264 |
| MB-In 435 | 2.05 | 0.02018 | 0.1632 | 2.085 | 0.01854 | 0.1185 | 1.569 | 0.05831 | 0.2221 |
| MGE-IN 257 | 2.042 | 0.02056 | 0.1634 | 1.72 | 0.04275 | 0.1857 | 2.056 | 0.01988 | 0.1479 |
| MB-In 441 | 1.995 | 0.02302 | 0.1794 | 0.8691 | 0.1924 | 0.4369 | 1.005 | 0.1575 | 0.3667 |
| eMSN 225 | 1.979 | 0.02391 | 0.1794 | 0.9549 | 0.1698 | 0.3954 | 1.693 | 0.04526 | 0.2039 |
| MGE-IN 263 | 1.965 | 0.02472 | 0.1794 | 2.019 | 0.02176 | 0.1253 | 1.809 | 0.03519 | 0.1788 |
| UL-IT 122 | 1.964 | 0.02476 | 0.1794 | 2.285 | 0.01115 | 0.1045 | 2.342 | 0.009584 | 0.1028 |
| MB-In 442 | 1.962 | 0.02486 | 0.1794 | 1.018 | 0.1543 | 0.3808 | 0.9651 | 0.1672 | 0.3817 |
| MSN 221 | 1.958 | 0.02512 | 0.1794 | -0.4108 | 0.6594 | 0.8785 | 2.008 | 0.02231 | 0.1497 |
| eMSN 222 | 1.955 | 0.0253 | 0.1794 | 0.5954 | 0.2758 | 0.5284 | 2.494 | 0.006307 | 0.09522 |
| DL-CT/6b 102 | 1.937 | 0.02637 | 0.1816 | 1.516 | 0.06476 | 0.2228 | 0.8322 | 0.2027 | 0.4214 |
| MGE-IN 241 | 1.937 | 0.02639 | 0.1816 | 1.466 | 0.07126 | 0.2347 | 1.276 | 0.101 | 0.2985 |
| CA4 197 | 1.929 | 0.02688 | 0.1822 | 0.749 | 0.2269 | 0.4691 | -1.058 | 0.855 | 0.9452 |
| Amyg-Exc 174 | 1.918 | 0.02758 | 0.1843 | 2.091 | 0.01825 | 0.1185 | 1.994 | 0.02305 | 0.1497 |
| MGE-IN 258 | 1.906 | 0.0283 | 0.1863 | 2.086 | 0.0185 | 0.1185 | 2.234 | 0.01275 | 0.1251 |
| HiDG 200 | 1.882 | 0.02995 | 0.1937 | 2 | 0.02275 | 0.1263 | 1.897 | 0.02891 | 0.1666 |
| Amyg-Exc 407 | 1.854 | 0.03183 | 0.1937 | 1.328 | 0.09211 | 0.2687 | 1.877 | 0.03027 | 0.1681 |
| Misc 178 | 1.854 | 0.03185 | 0.1937 | 1.643 | 0.05015 | 0.1946 | 0.8312 | 0.2029 | 0.4214 |
| DL-IT 148 | 1.847 | 0.03239 | 0.1937 | 2.48 | 0.006574 | 0.09471 | 0.5124 | 0.3042 | 0.5414 |
| CA1-3 3 | 1.845 | 0.03249 | 0.1937 | 1.197 | 0.1156 | 0.3082 | -0.372 | 0.645 | 0.8376 |
| HiDG 202 | 1.837 | 0.03311 | 0.1937 | 0.6895 | 0.2452 | 0.4916 | 0.9839 | 0.1626 | 0.3735 |
| LLC-IN 269 | 1.836 | 0.03315 | 0.1937 | 1.738 | 0.04115 | 0.1842 | 2.212 | 0.01347 | 0.1293 |
| U RhombLip 308 | 1.833 | 0.03342 | 0.1937 | 1.152 | 0.1246 | 0.3245 | 0.3615 | 0.3589 | 0.6082 |
| DL-NP 92 | 1.831 | 0.03358 | 0.1937 | 2.29 | 0.011 | 0.1045 | -0.6482 | 0.7416 | 0.891 |
| MB-In 444 | 1.83 | 0.03361 | 0.1937 | 1.602 | 0.05455 | 0.2012 | 0.4016 | 0.344 | 0.5906 |
| Splatter 422 | 1.818 | 0.03451 | 0.1964 | -1.183 | 0.8817 | 0.9805 | 0.2293 | 0.4093 | 0.6621 |
| Astr 59 | 1.809 | 0.03519 | 0.1964 | -0.07849 | 0.5313 | 0.803 | -0.9388 | 0.8261 | 0.9266 |
| Splatter 386 | 1.8 | 0.03589 | 0.1964 | -2.199 | 0.9861 | 0.9999 | -1.037 | 0.8502 | 0.9422 |
| eMSN 233 | 1.797 | 0.03619 | 0.1964 | 0.9806 | 0.1634 | 0.3903 | 1.37 | 0.08538 | 0.2696 |
| UL-IT 121 | 1.796 | 0.03621 | 0.1964 | 2.067 | 0.01939 | 0.1185 | 2.594 | 0.004742 | 0.08322 |
| Splatter 428 | 1.785 | 0.0371 | 0.1969 | 0.9698 | 0.1661 | 0.3934 | 2.346 | 0.009492 | 0.1028 |
| UL-IT 135 | 1.785 | 0.03716 | 0.1969 | 2.405 | 0.008088 | 0.1042 | 1.512 | 0.06525 | 0.2387 |
| Misc 176 | 1.769 | 0.03847 | 0.1993 | 0.6228 | 0.2667 | 0.5188 | 0.6844 | 0.2469 | 0.4802 |
| Amyg-Exc 162 | 1.769 | 0.03848 | 0.1993 | 2.158 | 0.01545 | 0.1113 | 3.134 | 0.0008616 | 0.07788 |
| HiDG 201 | 1.747 | 0.04036 | 0.2067 | 1.784 | 0.03723 | 0.1733 | 1.588 | 0.05611 | 0.2182 |
| MSN 209 | 1.74 | 0.04095 | 0.2074 | 1.189 | 0.1171 | 0.3102 | 2.454 | 0.007067 | 0.09522 |
| MGE-IN 240 | 1.72 | 0.04273 | 0.2108 | 2.147 | 0.01588 | 0.1126 | 2.585 | 0.004874 | 0.08322 |
| CGE-IN 292 | 1.716 | 0.0431 | 0.2108 | 0.6252 | 0.2659 | 0.5188 | 2.557 | 0.005284 | 0.087 |

| Cluster (Yao naming) | z (19 cohorts) | p (19 cohorts) | FDR (19 cohorts) | z (MVP diagnostic code) | p (MVP diagnostic code) | FDR (MVP diagnostic code) | z (MVP self-report) | p (MVP self-report) | FDR (MVP self-report) |
|---|---|---|---|---|---|---|---|---|---|
| Amyg-Exc 171 | 1.716 | 0.04313 | 0.2108 | 1.832 | 0.03347 | 0.1634 | 2.868 | 0.002066 | 0.07788 |
| Splatter 364 | 1.709 | 0.04376 | 0.2108 | 0.712 | 0.2382 | 0.4814 | 1.558 | 0.05958 | 0.2233 |
| Misc 177 | 1.707 | 0.0439 | 0.2108 | 2.035 | 0.02094 | 0.1238 | 2.378 | 0.008693 | 0.1028 |
| Mam 327 | 1.661 | 0.04833 | 0.2263 | -0.155 | 0.5616 | 0.8127 | -1.335 | 0.909 | 0.9723 |
| MGE-IN 255 | 1.66 | 0.04848 | 0.2263 | 1.785 | 0.03716 | 0.1733 | 1.221 | 0.1111 | 0.3161 |
| Cer-In 302 | 1.659 | 0.04859 | 0.2263 | 0.678 | 0.2489 | 0.4925 | 1.012 | 0.1558 | 0.3647 |
| Astr 60 | 1.648 | 0.04965 | 0.2274 | -0.1973 | 0.5782 | 0.8204 | -0.5683 | 0.7151 | 0.8767 |
| eMSN 229 | 1.646 | 0.04983 | 0.2274 | 0.8231 | 0.2052 | 0.4474 | 1.212 | 0.1128 | 0.317 |
| Misc 132 | 1.641 | 0.05044 | 0.2275 | 2.402 | 0.008142 | 0.1042 | 1.276 | 0.101 | 0.2985 |
| DL-NP 95 | 1.637 | 0.05084 | 0.2275 | 1.637 | 0.05084 | 0.1946 | 0.5437 | 0.2933 | 0.5303 |
| UL-IT 123 | 1.625 | 0.05205 | 0.2276 | 2.3 | 0.01073 | 0.1045 | 2.833 | 0.002308 | 0.07788 |
| CA1-3 3 | 1.622 | 0.0524 | 0.2276 | 1.081 | 0.1398 | 0.358 | -0.08062 | 0.5321 | 0.7434 |
| MB-In 434 | 1.62 | 0.05259 | 0.2276 | 1.985 | 0.02359 | 0.1295 | 1.493 | 0.06768 | 0.2438 |
| MSN 215 | 1.618 | 0.05283 | 0.2276 | 0.8646 | 0.1936 | 0.4375 | 1.227 | 0.11 | 0.3161 |
| MGE-IN 262 | 1.597 | 0.05513 | 0.2329 | 1.921 | 0.02738 | 0.1434 | 2.508 | 0.006071 | 0.09522 |
| MSN 217 | 1.586 | 0.05642 | 0.2329 | 1.732 | 0.04165 | 0.1846 | 2.001 | 0.0227 | 0.1497 |
| MSN 213 | 1.575 | 0.05764 | 0.2329 | 0.5941 | 0.2762 | 0.5284 | 1.842 | 0.03275 | 0.1737 |
| CGE-IN 289 | 1.574 | 0.05771 | 0.2329 | 1.638 | 0.05071 | 0.1946 | 2.288 | 0.01108 | 0.1135 |
| DL-IT 140 | 1.574 | 0.05776 | 0.2329 | 3.051 | 0.001141 | 0.04784 | 2.086 | 0.01848 | 0.1444 |
| Splatter 237 | 1.572 | 0.05796 | 0.2329 | 1.699 | 0.04466 | 0.1883 | 1.128 | 0.1297 | 0.3329 |
| MSN 207 | 1.572 | 0.05799 | 0.2329 | 1.611 | 0.05358 | 0.1992 | 1.393 | 0.08179 | 0.2637 |
| Mam 328 | 1.571 | 0.0581 | 0.2329 | 0.4073 | 0.3419 | 0.6085 | -1.258 | 0.8958 | 0.9649 |
| DL-CT/6b 109 | 1.566 | 0.0587 | 0.2333 | 2.41 | 0.007974 | 0.1042 | 1.08 | 0.14 | 0.345 |
| MGE-IN 254 | 1.556 | 0.05985 | 0.2358 | 0.8188 | 0.2065 | 0.4474 | 1.658 | 0.04867 | 0.2077 |
| MGE-IN 250 | 1.546 | 0.06105 | 0.2385 | 1.557 | 0.05971 | 0.2101 | 1.628 | 0.05178 | 0.217 |
| Misc 167 | 1.54 | 0.06177 | 0.2393 | 1.353 | 0.08797 | 0.2636 | 1.802 | 0.0358 | 0.1788 |
| Mam 333 | 1.514 | 0.06507 | 0.2493 | -1.457 | 0.9274 | 0.9826 | 0.1321 | 0.4475 | 0.6766 |
| Splatter 429 | 1.511 | 0.06544 | 0.2493 | -0.04823 | 0.5192 | 0.7926 | 0.3703 | 0.3556 | 0.6049 |
| MGE-IN 244 | 1.47 | 0.07081 | 0.2675 | 2.393 | 0.008364 | 0.1042 | 2.653 | 0.003993 | 0.07788 |
| MGE-IN 239 | 1.466 | 0.07138 | 0.2675 | 3.468 | 0.0002617 | 0.03017 | 2.151 | 0.01575 | 0.1378 |
| Splatter 403 | 1.457 | 0.07261 | 0.2688 | 0.8464 | 0.1987 | 0.4421 | 1.952 | 0.02546 | 0.1565 |
| DL-NP 87 | 1.454 | 0.07291 | 0.2688 | 1.576 | 0.0575 | 0.2087 | -0.3341 | 0.6309 | 0.8262 |
| CA1-3 3 | 1.45 | 0.07356 | 0.2688 | 0.3002 | 0.382 | 0.6615 | -1.565 | 0.9412 | 0.9794 |
| MGE-IN 248 | 1.446 | 0.07414 | 0.2688 | 2.075 | 0.01899 | 0.1185 | 0.8726 | 0.1914 | 0.403 |
| CA1-3 3 | 1.442 | 0.07463 | 0.2688 | 0.03088 | 0.4877 | 0.7628 | -0.9897 | 0.8388 | 0.9357 |
| UL-IT 134 | 1.436 | 0.07556 | 0.27 | 2.162 | 0.01531 | 0.1113 | 2.722 | 0.003241 | 0.07788 |
| Splatter 313 | 1.428 | 0.07668 | 0.2719 | 0.1206 | 0.452 | 0.7363 | -1.004 | 0.8424 | 0.9357 |
| Splatter 425 | 1.415 | 0.07852 | 0.2763 | 0.519 | 0.3019 | 0.5544 | 1.384 | 0.08312 | 0.2643 |
| DL-IT 147 | 1.397 | 0.08124 | 0.2806 | 3.317 | 0.0004547 | 0.03557 | 1.296 | 0.09753 | 0.2923 |
| Splatter 410 | 1.394 | 0.08168 | 0.2806 | -0.514 | 0.6964 | 0.9043 | -0.4217 | 0.6634 | 0.8495 |
| CGE-IN 286 | 1.392 | 0.08194 | 0.2806 | 3.128 | 0.0008803 | 0.04509 | 2.971 | 0.001486 | 0.07788 |
| CA4 192 | 1.391 | 0.08217 | 0.2806 | -0.2439 | 0.5963 | 0.838 | -0.792 | 0.7858 | 0.9209 |
| DL-NP 93 | 1.378 | 0.08417 | 0.2838 | 1.593 | 0.05556 | 0.2033 | -0.2414 | 0.5954 | 0.8002 |
| CA1-3 3 | 1.376 | 0.08435 | 0.2838 | 0.7662 | 0.2218 | 0.4667 | -0.01945 | 0.5078 | 0.7361 |
| Astr 58 | 1.365 | 0.0862 | 0.2863 | -0.6376 | 0.7381 | 0.9223 | -0.9772 | 0.8358 | 0.9352 |
| CA1-3 3 | 1.364 | 0.08634 | 0.2863 | 2.111 | 0.01737 | 0.1185 | 1.615 | 0.05311 | 0.2182 |
| CA4 198 | 1.355 | 0.08768 | 0.2873 | 1.134 | 0.1284 | 0.3326 | 0.09461 | 0.4623 | 0.6858 |

| Cluster (Yao naming) | z (19 cohorts) | p (19 cohorts) | FDR (19 cohorts) | z (MVP diagnostic code) | p (MVP diagnostic code) | FDR (MVP diagnostic code) | z (MVP self-report) | p (MVP self-report) | FDR (MVP self-report) |
|---|---|---|---|---|---|---|---|---|---|
| CA1-3 3 | 1.354 | 0.08787 | 0.2873 | 1.863 | 0.03122 | 0.1564 | 0.4851 | 0.3138 | 0.5531 |
| Thal-Exc 459 | 1.305 | 0.09599 | 0.3116 | 1.45 | 0.07346 | 0.2385 | 2.701 | 0.003458 | 0.07788 |
| Splatter 387 | 1.291 | 0.09828 | 0.3148 | -2.402 | 0.9918 | 0.9999 | 0.1554 | 0.4382 | 0.6766 |
| Splatter 396 | 1.288 | 0.09887 | 0.3148 | -0.7205 | 0.7644 | 0.9323 | 1.307 | 0.09566 | 0.2923 |
| Astr 53 | 1.282 | 0.09984 | 0.3148 | -1.373 | 0.9152 | 0.9826 | -0.5408 | 0.7057 | 0.8698 |
| MSN 430 | 1.28 | 0.1003 | 0.3148 | 1.937 | 0.02635 | 0.1413 | 2.368 | 0.008949 | 0.1028 |
| DL-IT 137 | 1.279 | 0.1004 | 0.3148 | 2.844 | 0.002226 | 0.0684 | 2.435 | 0.007439 | 0.09526 |
| Splatter 367 | 1.261 | 0.1036 | 0.3226 | 0.2732 | 0.3924 | 0.6724 | 2.233 | 0.01276 | 0.1251 |
| MGE-IN 245 | 1.253 | 0.1051 | 0.3251 | 2.217 | 0.0133 | 0.1072 | 3.273 | 0.0005314 | 0.07788 |
| LLC-IN 264 | 1.236 | 0.1083 | 0.3279 | 2.248 | 0.01228 | 0.1045 | 2.119 | 0.01704 | 0.1378 |
| LLC-IN 268 | 1.235 | 0.1083 | 0.3279 | 1.678 | 0.04671 | 0.1883 | 1.451 | 0.07342 | 0.2564 |
| Mam 329 | 1.235 | 0.1083 | 0.3279 | 0.05667 | 0.4774 | 0.7615 | -1.531 | 0.9371 | 0.9776 |
| DL-IT 149 | 1.233 | 0.1088 | 0.3279 | 2.586 | 0.004854 | 0.08287 | 1.795 | 0.03632 | 0.1788 |
| DL-IT 151 | 1.224 | 0.1105 | 0.3309 | 2.063 | 0.01954 | 0.1185 | 0.7913 | 0.2144 | 0.4363 |
| eMSN 223 | 1.179 | 0.1192 | 0.3528 | -0.3844 | 0.6496 | 0.8785 | 0.9578 | 0.1691 | 0.3821 |
| DL-NP 91 | 1.178 | 0.1194 | 0.3528 | 1.542 | 0.06156 | 0.215 | 0.6429 | 0.2601 | 0.4904 |
| Splatter 393 | 1.163 | 0.1224 | 0.3593 | 1.624 | 0.05223 | 0.1957 | 1.763 | 0.03898 | 0.1846 |
| Astr 54 | 1.148 | 0.1256 | 0.3642 | -1.703 | 0.9557 | 0.9901 | -0.6452 | 0.7406 | 0.891 |
| DL-NP 96 | 1.147 | 0.1256 | 0.3642 | 1.567 | 0.05859 | 0.2098 | -0.4629 | 0.6783 | 0.8626 |
| DL-IT 141 | 1.129 | 0.1295 | 0.3732 | 3.244 | 0.0005891 | 0.03557 | 3.249 | 0.0005791 | 0.07788 |
| Amyg-Exc 406 | 1.121 | 0.1312 | 0.3755 | 2.027 | 0.02132 | 0.1244 | 2.185 | 0.01443 | 0.133 |
| MGE-IN 252 | 1.104 | 0.1348 | 0.3782 | 1.098 | 0.1361 | 0.3506 | 1.409 | 0.07937 | 0.2592 |
| CA1-3 3 | 1.103 | 0.1349 | 0.3782 | 1.013 | 0.1554 | 0.3808 | -0.5245 | 0.7 | 0.8698 |
| eMSN 227 | 1.101 | 0.1353 | 0.3782 | 0.7562 | 0.2248 | 0.4667 | 1.416 | 0.07844 | 0.2592 |
| Astr 64 | 1.1 | 0.1356 | 0.3782 | -1.45 | 0.9264 | 0.9826 | -1.479 | 0.9304 | 0.9776 |
| MSN 218 | 1.098 | 0.1362 | 0.3782 | -0.513 | 0.696 | 0.9043 | 1.548 | 0.06079 | 0.226 |
| Splatter 395 | 1.087 | 0.1385 | 0.3792 | 0.996 | 0.1596 | 0.3853 | 0.6749 | 0.2499 | 0.482 |
| Mam 332 | 1.083 | 0.1394 | 0.3792 | -0.9539 | 0.8299 | 0.9605 | -0.1581 | 0.5628 | 0.7745 |
| eMSN 224 | 1.082 | 0.1396 | 0.3792 | 0.006928 | 0.4972 | 0.7691 | 0.882 | 0.1889 | 0.403 |
| MSN 210 | 1.081 | 0.1399 | 0.3792 | 0.8441 | 0.1993 | 0.4421 | 1.153 | 0.1245 | 0.3295 |
| Astr 56 | 1.077 | 0.1407 | 0.3792 | -0.5874 | 0.7215 | 0.9209 | -0.2298 | 0.5909 | 0.7965 |
| Misc 117 | 1.063 | 0.144 | 0.3858 | 1.704 | 0.04414 | 0.1883 | 1.252 | 0.1053 | 0.3071 |
| DL-IT 145 | 1.038 | 0.1496 | 0.3986 | 2.76 | 0.002893 | 0.07132 | 0.0253 | 0.4899 | 0.717 |
| Amyg-Exc 419 | 1.024 | 0.153 | 0.4048 | -0.571 | 0.716 | 0.9194 | 1.319 | 0.09366 | 0.2915 |
| UL-IT 129 | 1.02 | 0.1537 | 0.4048 | 3.592 | 0.0001644 | 0.02526 | 2.446 | 0.007229 | 0.09522 |
| UL-IT 125 | 1.017 | 0.1545 | 0.4048 | 1.328 | 0.09204 | 0.2687 | 1.588 | 0.05609 | 0.2182 |
| CGE-IN 296 | 1.003 | 0.158 | 0.4116 | 2.375 | 0.00877 | 0.1045 | 2.189 | 0.01429 | 0.133 |
| Astr 52 | 0.9986 | 0.159 | 0.4118 | -0.7682 | 0.7788 | 0.9341 | -0.2973 | 0.6169 | 0.8149 |
| Astr 63 | 0.9884 | 0.1615 | 0.4159 | -0.7965 | 0.7871 | 0.9341 | -1.292 | 0.9018 | 0.9688 |
| CA4 191 | 0.9728 | 0.1653 | 0.4234 | -0.3955 | 0.6537 | 0.8785 | -1.399 | 0.919 | 0.9776 |
| MGE-IN 247 | 0.9613 | 0.1682 | 0.4264 | 0.9249 | 0.1775 | 0.4092 | 1.062 | 0.1442 | 0.3498 |
| Mam 326 | 0.959 | 0.1688 | 0.4264 | -0.3419 | 0.6338 | 0.867 | -1.862 | 0.9687 | 0.9953 |
| Splatter 392 | 0.9536 | 0.1701 | 0.4264 | -0.6594 | 0.7452 | 0.926 | -0.01633 | 0.5065 | 0.7361 |
| Thal-Exc 448 | 0.9534 | 0.1702 | 0.4264 | -0.2458 | 0.5971 | 0.838 | -0.4903 | 0.688 | 0.8643 |
| DL-CT/6b 103 | 0.9391 | 0.1738 | 0.4332 | 1.447 | 0.07402 | 0.2386 | 1.755 | 0.03961 | 0.1846 |
| CA1-3 3 | 0.9355 | 0.1748 | 0.4332 | 2.681 | 0.003669 | 0.07667 | 0.8922 | 0.1862 | 0.4022 |
| Splatter 340 | 0.9286 | 0.1765 | 0.4352 | 0.1073 | 0.4573 | 0.7422 | 0.592 | 0.2769 | 0.5106 |

| Cluster (Yao naming) | z (19 cohorts) | p (19 cohorts) | FDR (19 cohorts) | z (MVP diagnostic code) | p (MVP diagnostic code) | FDR (MVP diagnostic code) | z (MVP self-report) | p (MVP self-report) | FDR (MVP self-report) |
|---|---|---|---|---|---|---|---|---|---|
| Amyg-Exc 160 | 0.9215 | 0.1784 | 0.4374 | 0.5659 | 0.2857 | 0.537 | 1.04 | 0.1491 | 0.3598 |
| CGE-IN 295 | 0.9154 | 0.18 | 0.439 | 2.111 | 0.01739 | 0.1185 | 3.031 | 0.001217 | 0.07788 |
| Splatter 400 | 0.91 | 0.1814 | 0.4393 | 0.7092 | 0.2391 | 0.4814 | 0.09382 | 0.4626 | 0.6858 |
| Astr 62 | 0.9077 | 0.182 | 0.4393 | -0.4392 | 0.6697 | 0.8898 | -0.6422 | 0.7396 | 0.891 |
| DL-IT 142 | 0.901 | 0.1838 | 0.4413 | 2.944 | 0.001619 | 0.05742 | 1.221 | 0.111 | 0.3161 |
| MSN 427 | 0.8949 | 0.1854 | 0.4429 | -0.3985 | 0.6549 | 0.8785 | 1.802 | 0.03578 | 0.1788 |
| HiDG 199 | 0.8847 | 0.1882 | 0.4457 | 0.5079 | 0.3058 | 0.5551 | 0.8062 | 0.2101 | 0.4343 |
| UL-IT 128 | 0.8833 | 0.1885 | 0.4457 | 1.739 | 0.04099 | 0.1842 | 2.686 | 0.003615 | 0.07788 |
| Splatter 235 | 0.8577 | 0.1955 | 0.4554 | -0.4517 | 0.6743 | 0.8927 | -0.1094 | 0.5435 | 0.7547 |
| Splatter 349 | 0.856 | 0.196 | 0.4554 | -1.361 | 0.9132 | 0.9826 | 0.6286 | 0.2648 | 0.4962 |
| Astr 61 | 0.8535 | 0.1967 | 0.4554 | -1.284 | 0.9004 | 0.9826 | -1.125 | 0.8698 | 0.9489 |
| DL-NP 85 | 0.8506 | 0.1975 | 0.4554 | 2.294 | 0.0109 | 0.1045 | 0.6504 | 0.2577 | 0.4889 |
| LLC-IN 270 | 0.8503 | 0.1976 | 0.4554 | 2.038 | 0.02076 | 0.1238 | 2.355 | 0.009269 | 0.1028 |
| eMSN 426 | 0.8349 | 0.2019 | 0.463 | 1.429 | 0.07645 | 0.2431 | 1.295 | 0.09765 | 0.2923 |
| LLC-IN 271 | 0.8262 | 0.2044 | 0.4635 | 2.67 | 0.003791 | 0.07667 | 1.793 | 0.03645 | 0.1788 |
| Mam 331 | 0.8232 | 0.2052 | 0.4635 | -1.512 | 0.9347 | 0.9826 | -1.44 | 0.925 | 0.9776 |
| CA1-3 3 | 0.8205 | 0.206 | 0.4635 | 0.9127 | 0.1807 | 0.4145 | -0.5169 | 0.6974 | 0.8698 |
| Splatter 421 | 0.8199 | 0.2061 | 0.4635 | -0.06622 | 0.5264 | 0.8009 | 1.963 | 0.02482 | 0.1546 |
| Astr 57 | 0.8012 | 0.2115 | 0.4733 | -1.175 | 0.88 | 0.9805 | -0.3589 | 0.6402 | 0.8337 |
| CGE-IN 290 | 0.7766 | 0.2187 | 0.487 | 1.763 | 0.03891 | 0.1776 | 2.657 | 0.003938 | 0.07788 |
| CGE-IN 276 | 0.7609 | 0.2234 | 0.4932 | 1.017 | 0.1545 | 0.3808 | 0.5673 | 0.2853 | 0.5239 |
| Splatter 341 | 0.7579 | 0.2243 | 0.4932 | -0.2887 | 0.6136 | 0.8506 | 0.2726 | 0.3926 | 0.6457 |
| DL-NP 90 | 0.7566 | 0.2247 | 0.4932 | 2.165 | 0.01519 | 0.1113 | 1.12 | 0.1314 | 0.3329 |
| eMSN 232 | 0.749 | 0.2269 | 0.4958 | 1.303 | 0.09627 | 0.2774 | 1.616 | 0.05308 | 0.2182 |
| Splatter 378 | 0.7438 | 0.2285 | 0.4969 | -0.6001 | 0.7258 | 0.9209 | 0.8929 | 0.186 | 0.4022 |
| LLC-IN 273 | 0.728 | 0.2333 | 0.5014 | 1.434 | 0.0758 | 0.2427 | 0.6954 | 0.2434 | 0.4775 |
| CGE-IN 287 | 0.7278 | 0.2334 | 0.5014 | 2.525 | 0.005788 | 0.08894 | 1.628 | 0.05173 | 0.217 |
| MGE-IN 251 | 0.7262 | 0.2339 | 0.5014 | 1.012 | 0.1557 | 0.3808 | 1.191 | 0.1169 | 0.3171 |
| Cer-In 300 | 0.7214 | 0.2353 | 0.5023 | 1.365 | 0.08608 | 0.2633 | 0.4953 | 0.3102 | 0.55 |
| Splatter 418 | 0.6985 | 0.2424 | 0.515 | 0.06845 | 0.4727 | 0.7567 | 0.2383 | 0.4058 | 0.6611 |
| Splatter 371 | 0.6851 | 0.2466 | 0.5152 | 0.7306 | 0.2325 | 0.4761 | -0.04278 | 0.5171 | 0.7401 |
| L RhombLip 322 | 0.6847 | 0.2468 | 0.5152 | -1.478 | 0.9303 | 0.9826 | -0.6276 | 0.7349 | 0.891 |
| HiDG 205 | 0.6843 | 0.2469 | 0.5152 | 0.03063 | 0.4878 | 0.7628 | 0.2521 | 0.4005 | 0.6547 |
| Thal-Exc 450 | 0.684 | 0.247 | 0.5152 | 1.066 | 0.1432 | 0.3628 | 1.069 | 0.1425 | 0.3477 |
| Thal-Exc 449 | 0.6784 | 0.2487 | 0.5156 | 1.051 | 0.1466 | 0.3692 | 0.6863 | 0.2463 | 0.4802 |
| LLC-IN 267 | 0.6734 | 0.2503 | 0.5156 | 2.639 | 0.004158 | 0.07667 | 2.017 | 0.02184 | 0.1497 |
| U RhombLip 309 | 0.6729 | 0.2505 | 0.5156 | 0.7277 | 0.2334 | 0.4761 | 0.7987 | 0.2122 | 0.4348 |
| Splatter 363 | 0.6621 | 0.254 | 0.5203 | -0.9933 | 0.8397 | 0.9606 | 1.597 | 0.05513 | 0.2182 |
| DL-CT/6b 98 | 0.6567 | 0.2557 | 0.5215 | 2.467 | 0.006804 | 0.09505 | 1.75 | 0.04005 | 0.1846 |
| DL-CT/6b 110 | 0.6431 | 0.2601 | 0.5226 | 1.768 | 0.03856 | 0.1776 | 0.2922 | 0.3851 | 0.6408 |
| Splatter 412 | 0.643 | 0.2601 | 0.5226 | -0.07531 | 0.53 | 0.803 | 0.6694 | 0.2516 | 0.4833 |
| Mam 325 | 0.6422 | 0.2604 | 0.5226 | -0.8003 | 0.7882 | 0.9341 | -1.809 | 0.9648 | 0.995 |
| Misc 116 | 0.641 | 0.2608 | 0.5226 | 1.675 | 0.04697 | 0.1883 | 1.763 | 0.03893 | 0.1846 |
| CA4 190 | 0.6351 | 0.2627 | 0.5242 | 0.7566 | 0.2246 | 0.4667 | 0.1514 | 0.4398 | 0.6766 |
| DL-CT/6b 101 | 0.6296 | 0.2645 | 0.525 | 1.788 | 0.03689 | 0.1733 | 1.177 | 0.1196 | 0.3225 |
| DL-CT/6b 100 | 0.6247 | 0.2661 | 0.525 | 2.65 | 0.004028 | 0.07667 | 1.015 | 0.1552 | 0.3647 |
| DL-NP 88 | 0.6198 | 0.2677 | 0.525 | 1.222 | 0.1109 | 0.306 | 0.2215 | 0.4123 | 0.6646 |

| Cluster (Yao naming) | z (19 cohorts) | p (19 cohorts) | FDR (19 cohorts) | z (MVP diagnostic code) | p (MVP diagnostic code) | FDR (MVP diagnostic code) | z (MVP self-report) | p (MVP self-report) | FDR (MVP self-report) |
|---|---|---|---|---|---|---|---|---|---|
| DL-NP 83 | 0.6174 | 0.2685 | 0.525 | 2.21 | 0.01356 | 0.1072 | 1.124 | 0.1304 | 0.3329 |
| U RhombLip 297 | 0.6166 | 0.2687 | 0.525 | 0.2024 | 0.4198 | 0.7038 | -1.129 | 0.8706 | 0.9489 |
| Cer-In 303 | 0.6024 | 0.2734 | 0.53 | 0.4814 | 0.3151 | 0.5697 | 0.2313 | 0.4086 | 0.6621 |
| DL-NP 86 | 0.6018 | 0.2736 | 0.53 | 1.691 | 0.04543 | 0.1883 | -0.749 | 0.7731 | 0.9138 |
| Splatter 402 | 0.5936 | 0.2764 | 0.5331 | 1.511 | 0.06536 | 0.2232 | 1.994 | 0.02306 | 0.1497 |
| Splatter 375 | 0.5809 | 0.2806 | 0.5391 | -1.591 | 0.9441 | 0.987 | 0.1145 | 0.4544 | 0.6801 |
| Cer-In 299 | 0.5686 | 0.2848 | 0.5419 | -0.7206 | 0.7644 | 0.9323 | 0.182 | 0.4278 | 0.6766 |
| DL-CT/6b 107 | 0.5684 | 0.2849 | 0.5419 | 0.8439 | 0.1994 | 0.4421 | 1.683 | 0.04614 | 0.2039 |
| CA1-3 3 | 0.5662 | 0.2856 | 0.5419 | 1.293 | 0.09795 | 0.2805 | -0.9017 | 0.8164 | 0.9209 |
| Splatter 343 | 0.5602 | 0.2877 | 0.5435 | 0.5217 | 0.301 | 0.5544 | 0.9607 | 0.1684 | 0.3821 |
| Cer-In 304 | 0.5505 | 0.291 | 0.5475 | -0.7758 | 0.7811 | 0.9341 | 0.4252 | 0.3354 | 0.5812 |
| MB-In 433 | 0.5348 | 0.2964 | 0.5555 | -0.1127 | 0.5449 | 0.8125 | -1.446 | 0.9259 | 0.9776 |
| MSN 219 | 0.5249 | 0.2998 | 0.5596 | -0.1893 | 0.5751 | 0.8204 | 1.09 | 0.1379 | 0.3419 |
| CGE-IN 288 | 0.508 | 0.3057 | 0.5683 | 2.754 | 0.00294 | 0.07132 | 1.385 | 0.08306 | 0.2643 |
| UL-IT 130 | 0.484 | 0.3142 | 0.5817 | 2.274 | 0.01149 | 0.1045 | 1.662 | 0.04823 | 0.2077 |
| DL-IT 139 | 0.478 | 0.3163 | 0.5833 | 2.249 | 0.01224 | 0.1045 | 1.81 | 0.03511 | 0.1788 |
| CA1-3 3 | 0.4734 | 0.318 | 0.584 | -0.3398 | 0.633 | 0.867 | -1.629 | 0.9483 | 0.9824 |
| CA1-3 3 | 0.4448 | 0.3282 | 0.6004 | 0.1216 | 0.4516 | 0.7363 | -0.8349 | 0.7981 | 0.9209 |
| DL-NP 89 | 0.4346 | 0.3319 | 0.6033 | 0.6242 | 0.2662 | 0.5188 | -0.5824 | 0.7198 | 0.8802 |
| CGE-IN 277 | 0.4333 | 0.3324 | 0.6033 | 2.367 | 0.008972 | 0.1045 | 1.263 | 0.1034 | 0.3035 |
| Splatter 391 | 0.4295 | 0.3338 | 0.6034 | -1.098 | 0.8639 | 0.9737 | 1.076 | 0.141 | 0.3458 |
| OPC 33 | 0.4126 | 0.3399 | 0.6083 | 1.03 | 0.1515 | 0.3796 | -0.1351 | 0.5537 | 0.7666 |
| Splatter 346 | 0.4104 | 0.3408 | 0.6083 | -0.1053 | 0.5419 | 0.8125 | 1.228 | 0.1097 | 0.3161 |
| MSN 206 | 0.4081 | 0.3416 | 0.6083 | 1.397 | 0.08125 | 0.2531 | 1.883 | 0.02987 | 0.1679 |
| DL-CT/6b 112 | 0.4048 | 0.3428 | 0.6083 | 0.9684 | 0.1664 | 0.3934 | 0.705 | 0.2404 | 0.4757 |
| UL-IT 138 | 0.4041 | 0.3431 | 0.6083 | 3.231 | 0.0006173 | 0.03557 | 2.36 | 0.009148 | 0.1028 |
| CGE-IN 293 | 0.3923 | 0.3474 | 0.6124 | 2.255 | 0.01208 | 0.1045 | 2.12 | 0.01702 | 0.1378 |
| DL-CT/6b 108 | 0.3892 | 0.3486 | 0.6124 | 2.607 | 0.004565 | 0.08094 | 1.013 | 0.1556 | 0.3647 |
| DL-CT/6b 104 | 0.384 | 0.3505 | 0.6124 | 2.572 | 0.00505 | 0.08314 | 0.7663 | 0.2218 | 0.4464 |
| Splatter 423 | 0.3787 | 0.3525 | 0.6124 | 0.3367 | 0.3682 | 0.6453 | 0.8735 | 0.1912 | 0.403 |
| Splatter 366 | 0.3777 | 0.3528 | 0.6124 | -2.84 | 0.9977 | 0.9999 | 0.1507 | 0.4401 | 0.6766 |
| Mam 330 | 0.3742 | 0.3541 | 0.6124 | -0.7927 | 0.786 | 0.9341 | -2.482 | 0.9935 | 0.9993 |
| Splatter 385 | 0.3701 | 0.3557 | 0.6124 | -1.348 | 0.9112 | 0.9826 | 0.201 | 0.4204 | 0.6705 |
| UL-IT 133 | 0.3659 | 0.3572 | 0.6124 | 1.834 | 0.03335 | 0.1634 | 0.4546 | 0.3247 | 0.5692 |
| Misc 114 | 0.3629 | 0.3583 | 0.6124 | 1.93 | 0.0268 | 0.142 | 1.407 | 0.07965 | 0.2592 |
| MGE-IN 256 | 0.3596 | 0.3596 | 0.6124 | 0.6107 | 0.2707 | 0.5221 | 0.9074 | 0.1821 | 0.3997 |
| OPC 32 | 0.3555 | 0.3611 | 0.6124 | -1.208 | 0.8866 | 0.9805 | -0.8291 | 0.7965 | 0.9209 |
| DL-CT/6b 106 | 0.3514 | 0.3626 | 0.6124 | 1.564 | 0.05889 | 0.2098 | 2.79 | 0.002639 | 0.07788 |
| Cer-In 306 | 0.3513 | 0.3627 | 0.6124 | 0.578 | 0.2816 | 0.5365 | 1.595 | 0.05541 | 0.2182 |
| LLC-IN 272 | 0.3417 | 0.3663 | 0.6147 | 2.267 | 0.01171 | 0.1045 | 2.413 | 0.007905 | 0.09849 |
| Splatter 373 | 0.3407 | 0.3667 | 0.6147 | -2.343 | 0.9904 | 0.9999 | 1.025 | 0.1526 | 0.3645 |
| Splatter 334 | 0.3285 | 0.3713 | 0.6202 | -0.1562 | 0.5621 | 0.8127 | -1.096 | 0.8634 | 0.9489 |
| Cer-In 301 | 0.3074 | 0.3793 | 0.6312 | -0.2483 | 0.5981 | 0.838 | -0.8939 | 0.8143 | 0.9209 |
| Splatter 424 | 0.3012 | 0.3816 | 0.6328 | -0.6075 | 0.7283 | 0.9209 | -0.0703 | 0.528 | 0.7434 |
| CA4 193 | 0.2858 | 0.3875 | 0.6403 | 0.5112 | 0.3046 | 0.5551 | -1.532 | 0.9373 | 0.9776 |
| DL-IT 150 | 0.2652 | 0.3954 | 0.6511 | 1.267 | 0.1025 | 0.2882 | 0.8786 | 0.1898 | 0.403 |
| DL-CT/6b 97 | 0.242 | 0.4044 | 0.6634 | 1.803 | 0.0357 | 0.1714 | -0.8188 | 0.7936 | 0.9209 |

| Cluster (Yao naming) | z (19 cohorts) | p (19 cohorts) | FDR (19 cohorts) | z (MVP diagnostic code) | p (MVP diagnostic code) | FDR (MVP diagnostic code) | z (MVP self-report) | p (MVP self-report) | FDR (MVP self-report) |
|---|---|---|---|---|---|---|---|---|---|
| Splatter 368 | 0.2186 | 0.4135 | 0.6759 | -2.086 | 0.9815 | 0.9999 | 0.5169 | 0.3026 | 0.5407 |
| CA4 195 | 0.2115 | 0.4162 | 0.6781 | 0.1925 | 0.4237 | 0.7051 | -1.076 | 0.8591 | 0.9475 |
| Splatter 354 | 0.2063 | 0.4183 | 0.679 | 0.9427 | 0.1729 | 0.4006 | 0.8901 | 0.1867 | 0.4022 |
| Splatter 389 | 0.1983 | 0.4214 | 0.6804 | -1.306 | 0.9042 | 0.9826 | -0.1426 | 0.5567 | 0.7684 |
| MSN 220 | 0.1965 | 0.4221 | 0.6804 | 0.7187 | 0.2362 | 0.4796 | 1.563 | 0.05898 | 0.2229 |
| Splatter 348 | 0.1876 | 0.4256 | 0.6836 | -1.514 | 0.935 | 0.9826 | 0.2026 | 0.4197 | 0.6705 |
| Thal-Exc 457 | 0.1819 | 0.4278 | 0.6841 | 0.2496 | 0.4014 | 0.6804 | 1.101 | 0.1354 | 0.3375 |
| Thal-Exc 372 | 0.1793 | 0.4288 | 0.6841 | -1.582 | 0.9432 | 0.987 | 0.2708 | 0.3933 | 0.6457 |
| eMSN 230 | 0.1437 | 0.4429 | 0.704 | 1.186 | 0.1178 | 0.3102 | 1.753 | 0.03981 | 0.1846 |
| DL-CT/6b 99 | 0.1327 | 0.4472 | 0.7085 | 2.323 | 0.0101 | 0.1045 | 1.462 | 0.07186 | 0.2529 |
| Thal-Exc 452 | 0.1247 | 0.4504 | 0.7111 | 0.3033 | 0.3808 | 0.6615 | -0.8759 | 0.8095 | 0.9209 |
| Cer-In 307 | 0.1017 | 0.4595 | 0.723 | 1.639 | 0.05056 | 0.1946 | 1.417 | 0.07821 | 0.2592 |
| Mam 323 | 0.08994 | 0.4642 | 0.7278 | -0.174 | 0.5691 | 0.8195 | -1.764 | 0.9611 | 0.9934 |
| Thal-Exc 454 | 0.08254 | 0.4671 | 0.73 | 0.66 | 0.2546 | 0.5016 | -0.1782 | 0.5707 | 0.7807 |
| Amyg-Exc 408 | 0.07822 | 0.4688 | 0.7302 | -0.181 | 0.5718 | 0.8195 | -1.232 | 0.8911 | 0.962 |
| Splatter 417 | 0.05616 | 0.4776 | 0.7402 | 0.787 | 0.2156 | 0.4581 | 1.471 | 0.07062 | 0.2524 |
| DL-CT/6b 105 | 0.05344 | 0.4787 | 0.7402 | 0.986 | 0.1621 | 0.3891 | 1.197 | 0.1157 | 0.3171 |
| Splatter 376 | 0.04994 | 0.4801 | 0.7402 | -1.984 | 0.9764 | 0.9999 | 1.2 | 0.1151 | 0.3171 |
| CGE-IN 284 | 0.03973 | 0.4842 | 0.744 | 1.244 | 0.1067 | 0.2964 | 1.467 | 0.0712 | 0.2525 |
| Splatter 382 | 0.03083 | 0.4877 | 0.7469 | -0.6586 | 0.7449 | 0.926 | 0.7297 | 0.2328 | 0.4666 |
| Astr 55 | 0.02175 | 0.4913 | 0.75 | -1.502 | 0.9334 | 0.9826 | -0.06951 | 0.5277 | 0.7434 |
| OPC 34 | 0.007568 | 0.497 | 0.7561 | -1.092 | 0.8627 | 0.9737 | 0.3304 | 0.3705 | 0.6257 |
| B 51 | 0.002375 | 0.4991 | 0.7568 | -0.7527 | 0.7742 | 0.9341 | 0.1585 | 0.437 | 0.6766 |
| CGE-IN 294 | -0.008594 | 0.5034 | 0.7609 | 2.164 | 0.01523 | 0.1113 | 2.041 | 0.02063 | 0.1497 |
| Splatter 415 | -0.02092 | 0.5083 | 0.7658 | 0.7951 | 0.2133 | 0.4552 | 1.583 | 0.05666 | 0.2182 |
| Thal-Exc 456 | -0.02737 | 0.5109 | 0.7672 | -0.1825 | 0.5724 | 0.8195 | 0.1147 | 0.4543 | 0.6801 |
| Splatter 365 | -0.04956 | 0.5198 | 0.7726 | -2.098 | 0.982 | 0.9999 | -0.9117 | 0.819 | 0.9209 |
| Mam 324 | -0.05064 | 0.5202 | 0.7726 | -0.6223 | 0.7331 | 0.9209 | -1.528 | 0.9368 | 0.9776 |
| Splatter 374 | -0.05252 | 0.5209 | 0.7726 | -0.621 | 0.7327 | 0.9209 | 2.142 | 0.0161 | 0.1378 |
| Splatter 383 | -0.05316 | 0.5212 | 0.7726 | -2.468 | 0.9932 | 0.9999 | 0.3081 | 0.379 | 0.6377 |
| Thal-Exc 447 | -0.06484 | 0.5258 | 0.7742 | 0.683 | 0.2473 | 0.4925 | -0.1004 | 0.54 | 0.752 |
| OPC 35 | -0.06966 | 0.5278 | 0.7742 | -0.2704 | 0.6066 | 0.847 | 0.1327 | 0.4472 | 0.6766 |
| Fibr 30 | -0.07394 | 0.5295 | 0.7742 | -0.9802 | 0.8365 | 0.9605 | -0.9059 | 0.8175 | 0.9209 |
| U RhombLip 311 | -0.07513 | 0.5299 | 0.7742 | 0.7981 | 0.2124 | 0.4552 | 0.66 | 0.2546 | 0.4852 |
| Misc 118 | -0.07703 | 0.5307 | 0.7742 | 2.488 | 0.006428 | 0.09471 | 2.106 | 0.01759 | 0.1398 |
| Thal-Exc 460 | -0.09934 | 0.5396 | 0.7833 | -0.9846 | 0.8376 | 0.9605 | -0.6073 | 0.7282 | 0.8881 |
| Splatter 355 | -0.1012 | 0.5403 | 0.7833 | -1.39 | 0.9177 | 0.9826 | 0.176 | 0.4301 | 0.6766 |
| Splatter 394 | -0.1737 | 0.5689 | 0.8209 | -0.01066 | 0.5043 | 0.773 | 1.406 | 0.07984 | 0.2592 |
| CGE-IN 282 | -0.1759 | 0.5698 | 0.8209 | -0.11 | 0.5438 | 0.8125 | 1.755 | 0.0396 | 0.1846 |
| L RhombLip 319 | -0.1848 | 0.5733 | 0.8234 | -1.52 | 0.9357 | 0.9826 | -1.231 | 0.8909 | 0.962 |
| MGE-IN 246 | -0.1928 | 0.5764 | 0.8253 | 0.2363 | 0.4066 | 0.6866 | 0.767 | 0.2215 | 0.4464 |
| Splatter 345 | -0.2053 | 0.5813 | 0.8291 | 0.08783 | 0.465 | 0.7469 | 0.6414 | 0.2606 | 0.4904 |
| Splatter 358 | -0.2089 | 0.5827 | 0.8291 | -1.701 | 0.9556 | 0.9901 | -0.836 | 0.7984 | 0.9209 |
| U RhombLip 310 | -0.2185 | 0.5865 | 0.8319 | 1.371 | 0.08513 | 0.2633 | 0.186 | 0.4262 | 0.6766 |
| Fibr 31 | -0.2277 | 0.5901 | 0.8336 | -1.028 | 0.848 | 0.9653 | -1.591 | 0.9441 | 0.9803 |
| Misc 3 | -0.2368 | 0.5936 | 0.8336 | 0.2321 | 0.4082 | 0.6868 | -0.5392 | 0.7051 | 0.8698 |
| L RhombLip 321 | -0.2426 | 0.5959 | 0.8336 | -1.021 | 0.8464 | 0.9653 | -0.6682 | 0.748 | 0.891 |

| Cluster (Yao naming) | z (19 cohorts) | p (19 cohorts) | FDR (19 cohorts) | z (MVP diagnostic code) | p (MVP diagnostic code) | FDR (MVP diagnostic code) | z (MVP self-report) | p (MVP self-report) | FDR (MVP self-report) |
|---|---|---|---|---|---|---|---|---|---|
| L RhombLip 318 | -0.2432 | 0.5961 | 0.8336 | -0.2745 | 0.6082 | 0.847 | -0.2659 | 0.6049 | 0.8059 |
| UL-IT 131 | -0.2448 | 0.5967 | 0.8336 | 2.288 | 0.01107 | 0.1045 | 1.407 | 0.07964 | 0.2592 |
| Splatter 388 | -0.2587 | 0.6021 | 0.8349 | -1.144 | 0.8736 | 0.9791 | 0.4118 | 0.3402 | 0.5874 |
| Splatter 413 | -0.2617 | 0.6032 | 0.8349 | -1.924 | 0.9728 | 0.9999 | -1.476 | 0.93 | 0.9776 |
| L RhombLip 315 | -0.2658 | 0.6048 | 0.8349 | -0.4685 | 0.6803 | 0.8935 | -0.558 | 0.7116 | 0.8748 |
| Misc 113 | -0.2661 | 0.6049 | 0.8349 | 1.962 | 0.02491 | 0.1351 | 0.7898 | 0.2148 | 0.4363 |
| L RhombLip 317 | -0.2735 | 0.6078 | 0.8364 | -0.1383 | 0.555 | 0.8127 | -0.8961 | 0.8149 | 0.9209 |
| CGE-IN 279 | -0.2924 | 0.615 | 0.8438 | 0.258 | 0.3982 | 0.6774 | 0.8506 | 0.1975 | 0.4138 |
| Splatter 369 | -0.315 | 0.6236 | 0.8531 | -1.087 | 0.8614 | 0.9737 | 0.8797 | 0.1895 | 0.403 |
| MiG 11 | -0.3215 | 0.6261 | 0.8539 | 0.6137 | 0.2697 | 0.5221 | -0.2006 | 0.5795 | 0.788 |
| Cer-In 305 | -0.3402 | 0.6331 | 0.861 | 2.291 | 0.01097 | 0.1045 | 1.409 | 0.07939 | 0.2592 |
| CGE-IN 278 | -0.3468 | 0.6356 | 0.8618 | 0.5582 | 0.2884 | 0.5382 | 0.9829 | 0.1628 | 0.3735 |
| DL-CT/6b 111 | -0.3668 | 0.6431 | 0.8691 | 2.098 | 0.01793 | 0.1185 | -0.226 | 0.5894 | 0.7965 |
| Splatter 432 | -0.3712 | 0.6448 | 0.8691 | -1.15 | 0.875 | 0.9791 | 0.2102 | 0.4167 | 0.6694 |
| Splatter 359 | -0.4088 | 0.6587 | 0.8853 | -0.9695 | 0.8338 | 0.9605 | -0.07917 | 0.5315 | 0.7434 |
| CGE-IN 283 | -0.4382 | 0.6694 | 0.8971 | 1.413 | 0.07883 | 0.2489 | 1.208 | 0.1135 | 0.317 |
| L RhombLip 316 | -0.481 | 0.6847 | 0.909 | -1.34 | 0.9099 | 0.9826 | 0.5223 | 0.3007 | 0.5394 |
| DL-IT 152 | -0.484 | 0.6858 | 0.909 | 1.483 | 0.06906 | 0.2307 | 0.6962 | 0.2432 | 0.4775 |
| Thal-Exc 446 | -0.487 | 0.6869 | 0.909 | -0.1237 | 0.5492 | 0.8127 | -0.2943 | 0.6157 | 0.8149 |
| DL-NP 94 | -0.4981 | 0.6908 | 0.909 | 1.007 | 0.157 | 0.3808 | 0.2847 | 0.3879 | 0.6433 |
| LLC-IN 274 | -0.5002 | 0.6915 | 0.909 | 1.897 | 0.02891 | 0.1481 | 1.848 | 0.03227 | 0.1737 |
| Splatter 238 | -0.5048 | 0.6932 | 0.909 | 1.213 | 0.1126 | 0.308 | 1.335 | 0.09094 | 0.2852 |
| Thal-Exc 453 | -0.5051 | 0.6932 | 0.909 | 0.1376 | 0.4453 | 0.7305 | -0.4361 | 0.6686 | 0.8538 |
| CGE-IN 285 | -0.5152 | 0.6968 | 0.909 | 3.014 | 0.00129 | 0.04957 | 1.15 | 0.1251 | 0.3295 |
| Splatter 338 | -0.5187 | 0.698 | 0.909 | -0.7687 | 0.779 | 0.9341 | -0.3284 | 0.6287 | 0.8257 |
| L RhombLip 320 | -0.5188 | 0.6981 | 0.909 | -0.9694 | 0.8338 | 0.9605 | -1.145 | 0.8738 | 0.949 |
| Splatter 360 | -0.5481 | 0.7082 | 0.9196 | -0.01183 | 0.5047 | 0.773 | -0.4061 | 0.6577 | 0.8469 |
| Splatter 370 | -0.5717 | 0.7163 | 0.9261 | -0.1494 | 0.5594 | 0.8127 | 1.591 | 0.05585 | 0.2182 |
| CGE-IN 281 | -0.5745 | 0.7172 | 0.9261 | 0.5383 | 0.2952 | 0.5487 | 1.163 | 0.1224 | 0.3262 |
| Splatter 431 | -0.5833 | 0.7202 | 0.9274 | -0.1129 | 0.545 | 0.8125 | 0.5997 | 0.2744 | 0.508 |
| Thal-Exc 458 | -0.5945 | 0.7239 | 0.9296 | 0.8434 | 0.1995 | 0.4421 | 0.7995 | 0.212 | 0.4348 |
| Thal-Exc 451 | -0.6024 | 0.7266 | 0.9304 | 1.335 | 0.09098 | 0.2687 | 1.211 | 0.113 | 0.317 |
| Thal-Exc 445 | -0.6432 | 0.7399 | 0.9366 | -0.4049 | 0.6572 | 0.8785 | -0.5075 | 0.6941 | 0.8695 |
| MiG 12 | -0.6441 | 0.7402 | 0.9366 | -0.8792 | 0.8104 | 0.9458 | -0.5362 | 0.7041 | 0.8698 |
| Splatter 399 | -0.646 | 0.7409 | 0.9366 | -0.2277 | 0.5901 | 0.8344 | -0.03413 | 0.5136 | 0.7394 |
| COP 37 | -0.6461 | 0.7409 | 0.9366 | 0.8822 | 0.1888 | 0.4309 | 0.5532 | 0.2901 | 0.5264 |
| Misc 0 | -0.6483 | 0.7416 | 0.9366 | -1.312 | 0.9053 | 0.9826 | -0.8149 | 0.7924 | 0.9209 |
| Splatter 352 | -0.6584 | 0.7449 | 0.9382 | 1.008 | 0.1568 | 0.3808 | 1.298 | 0.09716 | 0.2923 |
| U RhombLip 312 | -0.6912 | 0.7553 | 0.9467 | 0.1865 | 0.426 | 0.7064 | -0.2466 | 0.5974 | 0.8006 |
| Splatter 377 | -0.6967 | 0.757 | 0.9467 | 0.1583 | 0.4371 | 0.7222 | 0.1637 | 0.435 | 0.6766 |
| Splatter 356 | -0.6991 | 0.7577 | 0.9467 | -3.094 | 0.999 | 0.9999 | -0.06675 | 0.5266 | 0.7434 |
| Splatter 357 | -0.7184 | 0.7637 | 0.9487 | -0.8118 | 0.7915 | 0.9356 | -0.481 | 0.6847 | 0.8643 |
| DL-CT/6b 84 | -0.7226 | 0.765 | 0.9487 | 1.208 | 0.1136 | 0.308 | 0.1597 | 0.4365 | 0.6766 |
| LLC-IN 275 | -0.7242 | 0.7655 | 0.9487 | 1.562 | 0.05917 | 0.2098 | 1.436 | 0.07548 | 0.2592 |
| Fibr 24 | -0.7411 | 0.7707 | 0.9525 | -0.8739 | 0.8089 | 0.9458 | -0.9117 | 0.819 | 0.9209 |
| CGE-IN 280 | -0.7622 | 0.777 | 0.9554 | 0.09714 | 0.4613 | 0.7436 | 0.9215 | 0.1784 | 0.3954 |
| Splatter 384 | -0.7626 | 0.7772 | 0.9554 | -0.157 | 0.5624 | 0.8127 | 0.27 | 0.3936 | 0.6457 |

| Cluster (Yao naming) | z (19 cohorts) | p (19 cohorts) | FDR (19 cohorts) | z (MVP diagnostic code) | p (MVP diagnostic code) | FDR (MVP diagnostic code) | z (MVP self-report) | p (MVP self-report) | FDR (MVP self-report) |
|---|---|---|---|---|---|---|---|---|---|
| CGE-IN 291 | -0.7997 | 0.788 | 0.9662 | 1.683 | 0.04622 | 0.1883 | 2.8 | 0.002553 | 0.07788 |
| HiDG 204 | -0.8117 | 0.7915 | 0.9679 | 0.5076 | 0.3059 | 0.5551 | 0.9237 | 0.1778 | 0.3954 |
| Thal-Exc 455 | -0.8317 | 0.7972 | 0.9721 | 1.354 | 0.08791 | 0.2636 | 0.927 | 0.177 | 0.3954 |
| Splatter 347 | -0.8387 | 0.7992 | 0.9721 | -0.852 | 0.8029 | 0.9445 | -0.04659 | 0.5186 | 0.7401 |
| OPC 36 | -0.858 | 0.8045 | 0.976 | -0.4101 | 0.6591 | 0.8785 | 0.1226 | 0.4512 | 0.6798 |
| Splatter 381 | -0.8735 | 0.8088 | 0.9786 | -0.1977 | 0.5783 | 0.8204 | 0.9294 | 0.1763 | 0.3954 |
| Splatter 411 | -0.9441 | 0.8274 | 0.9986 | -0.1266 | 0.5504 | 0.8127 | 1.114 | 0.1327 | 0.3342 |
| Splatter 362 | -0.9654 | 0.8328 | 0.999 | -0.5116 | 0.6955 | 0.9043 | 1.295 | 0.09761 | 0.2923 |
| Splatter 409 | -0.9781 | 0.836 | 0.999 | 0.01636 | 0.4935 | 0.766 | 0.624 | 0.2663 | 0.4971 |
| Splatter 361 | -1.035 | 0.8497 | 0.999 | -0.8768 | 0.8097 | 0.9458 | -0.4752 | 0.6827 | 0.8643 |
| MiG 8 | -1.046 | 0.8523 | 0.999 | -0.3722 | 0.6451 | 0.8773 | -0.6543 | 0.7435 | 0.891 |
| Splatter 350 | -1.074 | 0.8587 | 0.999 | -1.338 | 0.9095 | 0.9826 | 0.4267 | 0.3348 | 0.5812 |
| Fibr 26 | -1.084 | 0.8608 | 0.999 | 0.1394 | 0.4445 | 0.7305 | -1.129 | 0.8706 | 0.9489 |
| MiG 4 | -1.086 | 0.8613 | 0.999 | -1.777 | 0.9622 | 0.9946 | -1.975 | 0.9759 | 0.9953 |
| Splatter 414 | -1.098 | 0.8639 | 0.999 | -0.6917 | 0.7554 | 0.9262 | 0.6153 | 0.2692 | 0.5004 |
| Epend 74 | -1.105 | 0.8655 | 0.999 | -1.21 | 0.8869 | 0.9805 | -2.721 | 0.9967 | 0.9993 |
| MiG 10 | -1.142 | 0.8733 | 0.999 | 0.04973 | 0.4802 | 0.7619 | -0.6498 | 0.7421 | 0.891 |
| Misc 2 | -1.146 | 0.8741 | 0.999 | -0.3644 | 0.6422 | 0.8759 | 0.01514 | 0.494 | 0.7206 |
| Vasc 13 | -1.156 | 0.8761 | 0.999 | 1.364 | 0.08625 | 0.2633 | -0.7021 | 0.7587 | 0.9014 |
| Splatter 380 | -1.161 | 0.8771 | 0.999 | -3.844 | 0.9999 | 0.9999 | -1.129 | 0.8705 | 0.9489 |
| Cer-In 298 | -1.185 | 0.882 | 0.999 | -1.635 | 0.9489 | 0.9875 | -0.4851 | 0.6862 | 0.8643 |
| Epend 68 | -1.186 | 0.8822 | 0.999 | -0.8002 | 0.7882 | 0.9341 | -1.494 | 0.9324 | 0.9776 |
| Splatter 335 | -1.223 | 0.8893 | 0.999 | 0.415 | 0.3391 | 0.6058 | -0.6627 | 0.7462 | 0.891 |
| Vasc 14 | -1.223 | 0.8893 | 0.999 | 0.5241 | 0.3001 | 0.5544 | 0.1376 | 0.4453 | 0.6766 |
| Vasc 18 | -1.235 | 0.8916 | 0.999 | 0.2678 | 0.3944 | 0.6735 | 0.05303 | 0.4789 | 0.703 |
| Splatter 342 | -1.241 | 0.8926 | 0.999 | -0.5957 | 0.7243 | 0.9209 | 0.1443 | 0.4426 | 0.6766 |
| Splatter 416 | -1.243 | 0.8931 | 0.999 | -0.1164 | 0.5463 | 0.8125 | 0.8969 | 0.1849 | 0.4022 |
| Misc 1 | -1.247 | 0.8938 | 0.999 | 1.352 | 0.08816 | 0.2636 | -0.1912 | 0.5758 | 0.7854 |
| COP 38 | -1.266 | 0.8973 | 0.999 | 2.005 | 0.02248 | 0.1263 | 0.91 | 0.1814 | 0.3997 |
| Epend 71 | -1.277 | 0.8992 | 0.999 | -1.227 | 0.8902 | 0.9817 | -2.318 | 0.9898 | 0.9993 |
| CA4 194 | -1.279 | 0.8996 | 0.999 | 0.6794 | 0.2484 | 0.4925 | -0.6577 | 0.7446 | 0.891 |
| MiG 7 | -1.305 | 0.904 | 0.999 | -0.6671 | 0.7477 | 0.9262 | -0.4655 | 0.6792 | 0.8626 |
| Misc 401 | -1.315 | 0.9057 | 0.999 | -1.477 | 0.9301 | 0.9826 | 0.3861 | 0.3497 | 0.5971 |
| Epend 67 | -1.329 | 0.9081 | 0.999 | -1.618 | 0.9471 | 0.9875 | -2.126 | 0.9832 | 0.9984 |
| MiG 5 | -1.332 | 0.9085 | 0.999 | -1.079 | 0.8598 | 0.9737 | -1.15 | 0.8749 | 0.949 |
| Splatter 339 | -1.341 | 0.91 | 0.999 | -0.6378 | 0.7382 | 0.9223 | 0.1316 | 0.4476 | 0.6766 |
| Misc 314 | -1.398 | 0.919 | 0.999 | -0.4781 | 0.6837 | 0.8954 | -0.07562 | 0.5301 | 0.7434 |
| MiG 6 | -1.437 | 0.9246 | 0.999 | -0.5512 | 0.7093 | 0.9157 | -0.8423 | 0.8002 | 0.9209 |
| Splatter 344 | -1.458 | 0.9275 | 0.999 | -0.3312 | 0.6298 | 0.8666 | 1.104 | 0.1348 | 0.3375 |
| COP 39 | -1.463 | 0.9283 | 0.999 | -0.4619 | 0.6779 | 0.8929 | -0.3389 | 0.6327 | 0.8262 |
| Vasc 16 | -1.464 | 0.9283 | 0.999 | 1.476 | 0.06999 | 0.2321 | -0.1667 | 0.5662 | 0.7768 |
| Vasc 15 | -1.465 | 0.9286 | 0.999 | 0.7615 | 0.2232 | 0.4667 | 0.6792 | 0.2485 | 0.4813 |
| COP 42 | -1.474 | 0.9297 | 0.999 | 0.00298 | 0.4988 | 0.7691 | -0.06409 | 0.5256 | 0.7434 |
| Splatter 397 | -1.479 | 0.9305 | 0.999 | 0.3809 | 0.3516 | 0.6235 | 0.3998 | 0.3446 | 0.5906 |
| MiG 9 | -1.491 | 0.932 | 0.999 | -1.368 | 0.9143 | 0.9826 | -2.056 | 0.9801 | 0.9974 |
| Splatter 353 | -1.538 | 0.9379 | 0.999 | 0.04782 | 0.4809 | 0.7619 | 0.7057 | 0.2402 | 0.4757 |
| Splatter 398 | -1.566 | 0.9413 | 0.999 | -0.8529 | 0.8032 | 0.9445 | 0.7207 | 0.2355 | 0.4701 |

| Cluster (Yao naming) | z (19 cohorts) | p (19 cohorts) | FDR (19 cohorts) | z (MVP diagnostic code) | p (MVP diagnostic code) | FDR (MVP diagnostic code) | z (MVP self-report) | p (MVP self-report) | FDR (MVP self-report) |
|---|---|---|---|---|---|---|---|---|---|
| Olig 40 | -1.596 | 0.9448 | 0.999 | -0.5415 | 0.7059 | 0.9141 | -0.3139 | 0.6232 | 0.8208 |
| Olig 46 | -1.612 | 0.9465 | 0.999 | 0.7438 | 0.2285 | 0.4703 | 0.4836 | 0.3143 | 0.5531 |
| Splatter 379 | -1.62 | 0.9474 | 0.999 | -2.042 | 0.9794 | 0.9999 | -0.8882 | 0.8128 | 0.9209 |
| Vasc 20 | -1.65 | 0.9505 | 0.999 | -1.473 | 0.9296 | 0.9826 | -2.472 | 0.9933 | 0.9993 |
| CPlx 78 | -1.678 | 0.9533 | 0.999 | -0.5566 | 0.7111 | 0.9157 | -0.4006 | 0.6557 | 0.8467 |
| Epend 65 | -1.73 | 0.9582 | 0.999 | -1.242 | 0.8929 | 0.9824 | -1.943 | 0.974 | 0.9953 |
| Epend 70 | -1.73 | 0.9582 | 0.999 | -0.7809 | 0.7826 | 0.9341 | -2.733 | 0.9969 | 0.9993 |
| CPlx 80 | -1.766 | 0.9613 | 0.999 | 0.5645 | 0.2862 | 0.537 | 1.192 | 0.1167 | 0.3171 |
| Vasc 23 | -1.824 | 0.966 | 0.999 | 0.0297 | 0.4882 | 0.7628 | -0.8583 | 0.8046 | 0.9209 |
| Epend 72 | -1.86 | 0.9685 | 0.999 | -1.433 | 0.9241 | 0.9826 | -1.497 | 0.9328 | 0.9776 |
| CPlx 82 | -1.917 | 0.9724 | 0.999 | -0.3226 | 0.6265 | 0.8647 | -1.001 | 0.8416 | 0.9357 |
| Splatter 337 | -1.918 | 0.9724 | 0.999 | 0.1946 | 0.4229 | 0.7051 | -0.2202 | 0.5871 | 0.7961 |
| Splatter 351 | -1.928 | 0.9731 | 0.999 | -0.7458 | 0.7721 | 0.9341 | 1.124 | 0.1305 | 0.3329 |
| Olig 48 | -1.937 | 0.9736 | 0.999 | -0.676 | 0.7505 | 0.9262 | -0.4151 | 0.661 | 0.8488 |
| Epend 66 | -1.94 | 0.9738 | 0.999 | -2.305 | 0.9894 | 0.9999 | -2.758 | 0.9971 | 0.9993 |
| Epend 73 | -1.963 | 0.9752 | 0.999 | -2.867 | 0.9979 | 0.9999 | -3.204 | 0.9993 | 0.9993 |
| Olig 47 | -1.973 | 0.9758 | 0.999 | 0.2815 | 0.3892 | 0.6694 | 0.3031 | 0.3809 | 0.6385 |
| CPlx 77 | -1.977 | 0.976 | 0.999 | -0.6114 | 0.7295 | 0.9209 | 0.5359 | 0.296 | 0.533 |
| COP 41 | -1.987 | 0.9765 | 0.999 | 0.8325 | 0.2026 | 0.4468 | 0.1432 | 0.4431 | 0.6766 |
| Fibr 29 | -2.024 | 0.9785 | 0.999 | -1.906 | 0.9717 | 0.9999 | -1.945 | 0.9741 | 0.9953 |
| Fibr 28 | -2.049 | 0.9798 | 0.999 | -0.1454 | 0.5578 | 0.8127 | -0.8768 | 0.8097 | 0.9209 |
| CPlx 79 | -2.099 | 0.9821 | 0.999 | -0.6867 | 0.7539 | 0.9262 | -0.2571 | 0.6015 | 0.8037 |
| COP 75 | -2.146 | 0.984 | 0.999 | -0.5999 | 0.7257 | 0.9209 | -1.43 | 0.9236 | 0.9776 |
| Splatter 390 | -2.164 | 0.9848 | 0.999 | 0.7691 | 0.2209 | 0.4667 | 0.08899 | 0.4645 | 0.6864 |
| Vasc 19 | -2.175 | 0.9852 | 0.999 | 0.03261 | 0.487 | 0.7628 | -0.8044 | 0.7894 | 0.9209 |
| Vasc 17 | -2.21 | 0.9865 | 0.999 | 0.8067 | 0.2099 | 0.4522 | -0.02863 | 0.5114 | 0.7391 |
| CPlx 81 | -2.251 | 0.9878 | 0.999 | -0.3947 | 0.6535 | 0.8785 | -0.5296 | 0.7018 | 0.8698 |
| Fibr 27 | -2.305 | 0.9894 | 0.999 | -0.9428 | 0.8271 | 0.9605 | -0.7124 | 0.7619 | 0.9029 |
| Epend 69 | -2.365 | 0.991 | 0.999 | -1.118 | 0.8682 | 0.9761 | -1.905 | 0.9716 | 0.9953 |
| Olig 49 | -2.431 | 0.9925 | 0.999 | 0.2973 | 0.3831 | 0.6615 | 0.6597 | 0.2547 | 0.4852 |
| Fibr 25 | -2.48 | 0.9934 | 0.999 | -0.456 | 0.6758 | 0.8927 | -0.03719 | 0.5148 | 0.7394 |
| Olig 45 | -2.502 | 0.9938 | 0.999 | 0.3412 | 0.3665 | 0.6448 | 0.133 | 0.4471 | 0.6766 |
| COP 43 | -2.618 | 0.9956 | 0.999 | 0.5635 | 0.2866 | 0.537 | -0.3966 | 0.6542 | 0.8467 |
| Splatter 336 | -2.721 | 0.9967 | 0.999 | -0.2908 | 0.6144 | 0.8506 | 0.2953 | 0.3839 | 0.6408 |
| CPlx 76 | -2.744 | 0.997 | 0.999 | -0.679 | 0.7514 | 0.9262 | 0.5543 | 0.2897 | 0.5264 |
| Vasc 21 | -2.817 | 0.9976 | 0.999 | -0.9146 | 0.8198 | 0.9544 | -1.303 | 0.9037 | 0.9688 |
| Vasc 22 | -2.939 | 0.9984 | 0.999 | -0.7982 | 0.7876 | 0.9341 | -0.7654 | 0.778 | 0.9173 |
| Olig 44 | -2.974 | 0.9985 | 0.999 | 0.3276 | 0.3716 | 0.6489 | 0.09415 | 0.4625 | 0.6858 |
| Olig 50 | -3.105 | 0.999 | 0.999 | -1.204 | 0.8857 | 0.9805 | 0.08325 | 0.4668 | 0.6876 |

#### Supplementary Table S16

Pairwise conditional analysis of the nine clusters reaching FDR < 0.10 in EARGEN

| Cluster tested | Cluster added to the model | Coefficient | Coefficient SE | Conditional coefficient z | Unconditional coefficient z |
|---|---|---|---|---|---|
| Misc_164 | Amex_161 | 5.66e-10 | 3.33e-10 | 1.701 | 3.587 |
| Amex_156 | Amex_161 | 3e-10 | 3.79e-10 | 0.7898 | 3.334 |
| DG_203 | Amex_161 | 3.53e-10 | 3.04e-10 | 1.159 | 3.23 |
| MGE_259 | Amex_161 | 1.06e-09 | 3.43e-10 | 3.076 | 3.2 |
| Amex_155 | Amex_161 | 3.68e-10 | 3.54e-10 | 1.041 | 3.132 |
| Amex_153 | Amex_161 | -7.74e-11 | 3.62e-10 | -0.2139 | 3.033 |
| Amex_159 | Amex_161 | 1.35e-10 | 3.41e-10 | 0.3946 | 2.965 |
| MGE_236 | Amex_161 | 7.76e-10 | 2.95e-10 | 2.631 | 2.927 |
| Amex_161 | Misc_164 | 1.16e-09 | 3.23e-10 | 3.585 | 4.823 |
| Amex_156 | Misc_164 | 8.2e-10 | 3.24e-10 | 2.527 | 3.334 |
| DG_203 | Misc_164 | 6.22e-10 | 2.85e-10 | 2.186 | 3.23 |
| MGE_259 | Misc_164 | 1.03e-09 | 3.4e-10 | 3.021 | 3.2 |
| Amex_155 | Misc_164 | 6.11e-10 | 3.36e-10 | 1.816 | 3.132 |
| Amex_153 | Misc_164 | 5.7e-10 | 3.11e-10 | 1.829 | 3.033 |
| Amex_159 | Misc_164 | 5.04e-10 | 2.98e-10 | 1.691 | 2.965 |
| MGE_236 | Misc_164 | 7.94e-10 | 2.9e-10 | 2.741 | 2.927 |
| Amex_161 | Amex_156 | 1.23e-09 | 3.5e-10 | 3.526 | 4.823 |
| Misc_164 | Amex_156 | 8.71e-10 | 3.11e-10 | 2.8 | 3.587 |
| DG_203 | Amex_156 | 5.78e-10 | 2.93e-10 | 1.97 | 3.23 |
| MGE_259 | Amex_156 | 1.05e-09 | 3.46e-10 | 3.041 | 3.2 |
| Amex_155 | Amex_156 | 6.82e-10 | 3.38e-10 | 2.017 | 3.132 |
| Amex_153 | Amex_156 | 4.25e-10 | 3.58e-10 | 1.186 | 3.033 |
| Amex_159 | Amex_156 | 5.24e-10 | 3.2e-10 | 1.636 | 2.965 |
| MGE_236 | Amex_156 | 8.07e-10 | 2.99e-10 | 2.694 | 2.927 |
| Amex_161 | DG_203 | 1.26e-09 | 3.33e-10 | 3.794 | 4.823 |
| Misc_164 | DG_203 | 9.2e-10 | 3.2e-10 | 2.88 | 3.587 |
| Amex_156 | DG_203 | 8.57e-10 | 3.49e-10 | 2.453 | 3.334 |
| MGE_259 | DG_203 | 1.07e-09 | 3.4e-10 | 3.133 | 3.2 |
| Amex_155 | DG_203 | 7.48e-10 | 3.24e-10 | 2.308 | 3.132 |
| Amex_153 | DG_203 | 6.25e-10 | 3.08e-10 | 2.031 | 3.033 |
| Amex_159 | DG_203 | 6.46e-10 | 2.98e-10 | 2.169 | 2.965 |
| MGE_236 | DG_203 | 8.41e-10 | 2.94e-10 | 2.863 | 2.927 |
| Amex_161 | MGE_259 | 1.37e-09 | 2.88e-10 | 4.764 | 4.823 |
| Misc_164 | MGE_259 | 1.02e-09 | 2.98e-10 | 3.404 | 3.587 |
| Amex_156 | MGE_259 | 1.01e-09 | 3.14e-10 | 3.203 | 3.334 |
| DG_203 | MGE_259 | 8.13e-10 | 2.6e-10 | 3.122 | 3.23 |
| Amex_155 | MGE_259 | 8.82e-10 | 2.99e-10 | 2.947 | 3.132 |
| Amex_153 | MGE_259 | 8.19e-10 | 2.86e-10 | 2.862 | 3.033 |
| Amex_159 | MGE_259 | 7.91e-10 | 2.82e-10 | 2.799 | 2.965 |
| MGE_236 | MGE_259 | 3.53e-10 | 3.64e-10 | 0.9704 | 2.927 |
| Amex_161 | Amex_155 | 1.22e-09 | 3.42e-10 | 3.559 | 4.823 |
| Misc_164 | Amex_155 | 8.08e-10 | 3.42e-10 | 2.361 | 3.587 |
| Amex_156 | Amex_155 | 8.01e-10 | 3.5e-10 | 2.291 | 3.334 |
| DG_203 | Amex_155 | 5.88e-10 | 2.88e-10 | 2.046 | 3.23 |
| MGE_259 | Amex_155 | 1.04e-09 | 3.45e-10 | 3.005 | 3.2 |
| Amex_153 | Amex_155 | 4.22e-10 | 4.06e-10 | 1.04 | 3.033 |
| Amex_159 | Amex_155 | 4.95e-10 | 3.15e-10 | 1.574 | 2.965 |
| MGE_236 | Amex_155 | 7.78e-10 | 2.93e-10 | 2.658 | 2.927 |
| Amex_161 | Amex_153 | 1.45e-09 | 3.72e-10 | 3.89 | 4.823 |
| Misc_164 | Amex_153 | 8.91e-10 | 3.28e-10 | 2.713 | 3.587 |
| Amex_156 | Amex_153 | 8.1e-10 | 3.91e-10 | 2.071 | 3.334 |
| DG_203 | Amex_153 | 6.04e-10 | 2.82e-10 | 2.142 | 3.23 |
| MGE_259 | Amex_153 | 1.06e-09 | 3.46e-10 | 3.047 | 3.2 |
| Amex_155 | Amex_153 | 6.96e-10 | 4.2e-10 | 1.656 | 3.132 |
| Amex_159 | Amex_153 | 5.83e-10 | 3.23e-10 | 1.804 | 2.965 |
| MGE_236 | Amex_153 | 8.18e-10 | 2.99e-10 | 2.738 | 2.927 |
| Amex_161 | Amex_159 | 1.33e-09 | 3.48e-10 | 3.817 | 4.823 |
| Misc_164 | Amex_159 | 8.67e-10 | 3.19e-10 | 2.714 | 3.587 |
| Amex_156 | Amex_159 | 8.24e-10 | 3.56e-10 | 2.315 | 3.334 |
| DG_203 | Amex_159 | 6.4e-10 | 2.8e-10 | 2.286 | 3.23 |

| Cluster tested | Cluster added to the model | Coefficient | Coefficient SE | Conditional coefficient z | Unconditional coefficient z |
|---|---|---|---|---|---|
| MGE_259 | Amex_159 | 1.05e-09 | 3.44e-10 | 3.059 | 3.2 |
| Amex_155 | Amex_159 | 6.95e-10 | 3.34e-10 | 2.083 | 3.132 |
| Amex_153 | Amex_159 | 5.88e-10 | 3.25e-10 | 1.81 | 3.033 |
| MGE_236 | Amex_159 | 8.11e-10 | 2.93e-10 | 2.763 | 2.927 |
| Amex_161 | MGE_236 | 1.35e-09 | 2.87e-10 | 4.693 | 4.823 |
| Misc_164 | MGE_236 | 1.02e-09 | 2.96e-10 | 3.452 | 3.587 |
| Amex_156 | MGE_236 | 9.99e-10 | 3.13e-10 | 3.197 | 3.334 |
| DG_203 | MGE_236 | 8.24e-10 | 2.63e-10 | 3.136 | 3.23 |
| MGE_259 | MGE_236 | 8.4e-10 | 4.25e-10 | 1.975 | 3.2 |
| Amex_155 | MGE_236 | 8.63e-10 | 2.98e-10 | 2.898 | 3.132 |
| Amex_153 | MGE_236 | 8.18e-10 | 2.84e-10 | 2.875 | 3.033 |
| Amex_159 | MGE_236 | 7.86e-10 | 2.81e-10 | 2.795 | 2.965 |
